# Educational attainment, structural brain reserve, and Alzheimer’s disease: a Mendelian randomization analysis

**DOI:** 10.1101/2022.03.19.22272444

**Authors:** Aida Seyedsalehi, Varun Warrier, Richard A. I. Bethlehem, Benjamin I. Perry, Stephen Burgess, Graham K. Murray

## Abstract

Higher educational attainment is observationally associated with lower risk of Alzheimer’s disease. However, the biological mechanisms underpinning this association remain unclear. The protective effect of education on Alzheimer’s disease may be mediated via increased brain reserve. We used two-sample Mendelian randomization to explore putative causal relationships between educational attainment, structural brain reserve as proxied by MRI phenotypes, and Alzheimer’s disease.

Summary statistics were obtained from genome-wide association studies of educational attainment (*n*=1,131,881), late-onset Alzheimer’s disease (35,274 cases, 59,163 controls), and 15 measures of grey or white matter macro- or micro-structure derived from structural or diffusion MRI (*n*_max_=33,211). We conducted univariable Mendelian randomization analyses to investigate bidirectional associations between (i) educational attainment and Alzheimer’s disease, (ii) educational attainment and imaging-derived phenotypes, (iii) imaging-derived phenotypes and Alzheimer’s disease. Multivariable Mendelian randomization was used to assess whether brain structure phenotypes mediated the protective effect of education on Alzheimer’s disease risk.

Genetically-proxied educational attainment was inversely associated with Alzheimer’s disease (odds ratio per standard deviation increase in genetically-predicted years of schooling = 0.70, 95% confidence interval [CI]: 0.60, 0.80). There was also evidence for positive associations between genetically-predicted educational attainment and four cortical macro-structure metrics (surface area: β=0.30, 95% CI: 0.20, 0.40; volume: β=0.29, 95% CI: 0.20, 0.37; intrinsic curvature: β=0.18, 95% CI: 0.11, 0.25; local gyrification index: β=0.21, 95% CI: 0.11, 0.31), as well as inverse associations with cortical intracellular volume fraction (β=-0.09, 95% CI: − 0.15, −0.03) and white matter hyperintensities volume (β=-0.14, 95% CI: −0.23, −0.05).

Genetically-proxied levels of three cortical macro-structure metrics were positively associated with years of education (surface area: β=0.13, 95% CI: 0.10, 0.16; volume: β=0.15, 95% CI: 0.11, 0.19; intrinsic curvature: β=0.12, 95% CI: 0.04, 0.19). We found no evidence of associations between genetically-predicted imaging-derived phenotypes and Alzheimer’s disease. The inverse association of genetically-predicted education with Alzheimer’s disease did not attenuate after adjusting for imaging-derived phenotypes in multivariable analyses.

Our results provide support for a protective causal effect of educational attainment on Alzheimer’s disease risk, as well as bi-directional causal associations between education and several brain macro- and micro-structure metrics. However, we did not find evidence that these structural markers affect risk of Alzheimer’s disease. The protective effect of education on Alzheimer’s disease may be mediated via other measures of brain reserve not included in the present study, or by alternative mechanisms.

## Introduction

Despite a large body of observational epidemiological evidence supporting education as a protective factor for Alzheimer’s disease,^1–8^ the biological mechanisms underpinning this association are not well-established. A potential mechanism by which higher educational attainment protects against risk of Alzheimer’s is through increasing or maintaining the underlying ‘brain reserve’ of the individual.^5^ Brain reserve refers to individual differences in the anatomical and structural characteristics of the brain that enable some individuals to preserve their cognitive and functional status despite neuropathology.^9–11^ The progression of amyloid-β and tau pathology in Alzheimer’s disease is associated with several structural alterations in the brain, including progressive cortical thinning,^12–15^ widespread grey matter atrophy in cortical and sub-cortical regions,^16,17^ and damage to the integrity and organization of white matter tracts.^18–21^ Many of these structural characteristics can be measured in-vivo using structural or diffusion MRI, and thus their premorbid levels may serve as a useful proxy for the structural basis of brain reserve capacity.^22,23^ In observational neuroimaging studies of cognitively intact older adults, higher education levels have been associated with increased whole-brain and regional grey matter volume,^24–26^ cortical thickness,^27–31^ and increased surface areas of sub-regions of the hippocampus and amygdala that are vulnerable to Alzheimer’s disease pathology.^32^ Similar findings have been reported for the association between education levels and white matter micro-structure in cognitively intact elderly, with more highly educated individuals showing increased white matter tract integrity in several brain regions that are characteristic sites of Alzheimer’s pathology.^28,33^

Given the observed associations between education and structural markers of brain reserve, it is plausible to hypothesize that changes in brain structure may mediate the protective effect of education on Alzheimer’s disease risk, through determining the underlying brain reserve of the individual. However, the weight of the evidence derives from conventional observational studies, which are prone to several limitations including residual and/or unmeasured confounding, reverse causation, and measurement error.^34^ Mendelian randomization (MR) is an epidemiological approach that can be used to assess evidence for a causal hypothesis using observational data.^34–37^ In MR, genetic variants that are specifically associated with a putative exposure are used to define population subgroups with different average exposure levels, which can be compared with respect to the outcome of interest.^38,39^ Due to the random assortment of alleles at conception, the distribution of genetic variants that are associated with a particular exposure is largely independent of factors that confound exposure-outcome associations in conventional observational analyses.^40–42^ Furthermore, since the genetic code for an individual is determined at conception and cannot be modified by subsequent disease outcomes, the direction of causation will always be from the genetic variants to the trait of interest, eliminating the potential for reverse causation.^34^

In recent years, a growing number of MR analyses have reported inverse associations of genetically-predicted educational attainment with Alzheimer’s disease risk.^43–47^ However, the mechanisms underlying the protective effect of education remain relatively unexplored. Recent advances in genotyping and multi-modal neuroimaging technologies have facilitated the collection of these data in large-scale prospective cohort studies,^48^ enabling the discovery of genetic variants associated with brain structure using considerably larger sample sizes than was previously possible.^49–52^ The present study aimed to expand on previous work by using MR to investigate putative causal relationships between educational attainment, brain structure as measured by MRI, and risk of Alzheimer’s disease. In addition, we aimed to explore whether there was any evidence for the hypothesis that brain structural alterations lie on the causal pathway from education to Alzheimer’s disease (i.e., the brain reserve hypothesis). We proxied structural brain reserve using imaging markers of grey and white-matter macro- and micro-structure derived from structural or diffusion MRI (see Methods for the description and choice of imaging-derived phenotypes used as markers of brain reserve).Our primary objectives were: (i) to replicate, using the latest GWAS data, the previous MR finding supporting a protective causal effect of educational attainment on Alzheimer’s disease risk; (ii) to assess whether educational attainment has a causal effect on brain macro- and/or micro-structure; (iii) to assess whether brain macro- and/or micro-structure phenotypes causally affect risk of Alzheimer’s disease; and (iv) to assess whether the protective effect of educational attainment on Alzheimer’s disease risk is mediated via changes to brain macro- and/or micro-structure, using multivariable MR (an extension to univariable MR which can be used to investigate mediating relationships).^53,54^ A directed acyclic graph illustrating the putative causal effects explored in this study is presented in Fig. 1.

**Figure 1.**
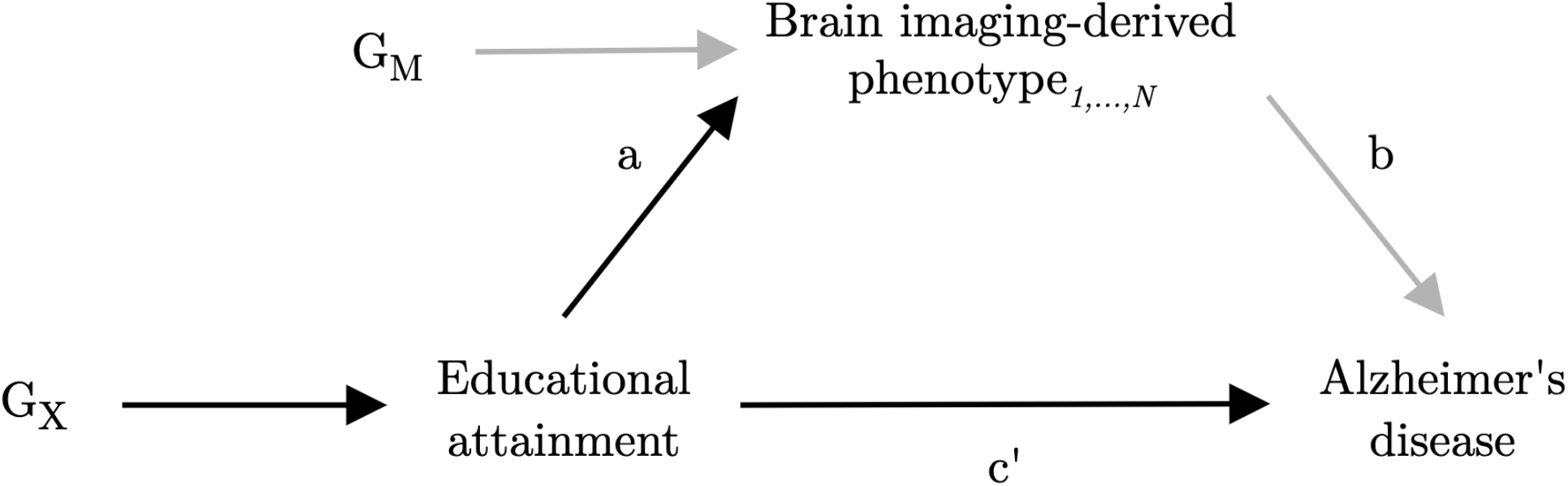
Directed acyclic graph illustrating the putative causal relationships examined in this study. Imaging-derived measures of brain structure are explored as potential mediators of the effect of educational attainment (exposure) on Alzheimer’s disease (outcome). G_X_ represents the set of instrumental variables for educational attainment. G_M_ represents the set of instrumental variables for brain imaging-derived phenotype_*1,…,N*_. Parameter *a* represents the direct causal effect of educational attainment on imaging-derived phenotype_*1,…,N*_. Parameter *b* represents the direct causal effect of imaging-derived phenotype_*1,…,N*_ on Alzheimer’s disease. Parameter *c’* represents the direct causal effect of educational attainment on Alzheimer’s disease. Parameters *a* and *c’* are estimated using the set of variants G_X_, and parameter *b* is estimated using the set of variants G_M_. Confounding variables are omitted from the diagram.

## Materials and methods

### Data Sources

The present study is based on a two-sample MR analysis strategy, an extension of MR in which the effects of the genetic instruments on the exposure and the outcome are obtained from two separate datasets.^55,56^ To minimize the possibility of confounding due to population stratification, all data sources were restricted to individuals of European ancestry. The GWAS used to obtain the summary statistics for each phenotype are listed in Table 1. Detailed description of the data sources is available in Supplementary Methods.

**Table 1.** Data sources used to obtain summary statistics for Mendelian randomization analyses.

| Phenotype | GWAS | n (as exposure) | n (as outcome) |
| --- | --- | --- | --- |
| Educational attainment | Lee *et al.*<sup>57</sup> | 1 131 881 | 766 345 |
| Late-onset Alzheimer's disease | Kunkle *et al.*<sup>58</sup> | 35 274 cases<br>59 163 controls | 21 982 cases<br>41 944 controls |
| **Cortical macro-structure** |  |  |  |
| Surface area | In-house GWAS | 31 779 | 31 779 |
| Volume |  | 31 780 | 31 780 |
| Cortical thickness |  | 31 779 | 31 779 |
| Local gyrification index |  | 31 708 | 31 708 |
| Mean curvature |  | 31 779 | 31 779 |
| Intrinsic curvature |  | 31 748 | 31 748 |
| **Cortical micro-structure** |  |  |  |
| Fractional anisotropy (cortex) | In-house GWAS | 31 782 | 31 782 |
| Mean diffusivity (cortex) |  | 31 714 | 31 714 |
| Intracellular volume fraction |  | 31 768 | 31 768 |
| Orientation dispersion index |  | 31 797 | 31 797 |
| **White matter micro-structure** |  |  |  |
| Fractional anisotropy (white matter tracts) | Zhao *et al.*<sup>52</sup> | 32 565 | 32 565 |
| Mean diffusivity (white matter tracts) |  | 32 343 | 32 343 |
| **Subcortical volume** |  |  |  |
| Volume of left hippocampus | Smith *et al.*<sup>51</sup> | 33 211 | 33 211 |
| Volume of right hippocampus |  | 33 211 | 33 211 |
| **White matter hyperintensities volume** |  |  |  |
| Total volume of white matter hyperintensities | Smith *et al.*<sup>51</sup> | 32 114 | 32 114 |

Note that for some phenotypes there is a difference in sample size between the exposure and outcome datasets. This was either because the summary statistics for the full set of variants were not publicly available for one or more of the meta-analysed cohorts (e.g., 23andMe), or the full replication analysis on which the final list of genome-wide significant variants was based was only conducted on a subset of the variants meeting pre-specified significance thresholds at earlier discovery stages. Where this was the case, all analyses of the phenotype as an exposure were based on genome-wide significant variants from the largest analysed sample (to maximise power for selecting instruments). In the analyses of the phenotype as an outcome, summary statistics were obtained from the dataset for which association estimates were provided for the full set of variants. GWAS = genome-wide association study.

Summary-level genetic association estimates with educational attainment were obtained from a GWAS meta-analysis of approximately 1.1 million individuals by Lee *et al*.^57^ Educational attainment was defined as the number of years of schooling completed, measured at an age of at least 30 years. Genetic variant association estimates with Alzheimer’s disease were taken from the largest available GWAS meta-analysis of clinically diagnosed late-onset Alzheimer’s disease (onset age > 65 years), as conducted by the International Genomics of Alzheimer’s Project (IGAP).^58^ Summary statistics for the imaging-derived phenotypes were obtained from three separate data sources based on the early-2020 release of combined genetic and multi-modal brain imaging data from the UK Biobank.^59^ Although several thousand imaging-derived phenotypes can be generated from UK Biobank MRI data, many are highly inter-correlated, and analysis of all imaging-derived phenotypes could lead to spurious results or challenges in controlling for multiple comparisons. Our strategy was to focus on 15 imaging-derived phenotypes (Fig. 2; Supplementary Methods), most of which reflect global brain measures previously shown to be associated phenotypically with educational attainment and/or Alzheimer’s disease.^12,14,16–19,24–28,31–33^ We also chose to study the total volume of white matter hyperintensities as a metric closely linked to age-related cognitive decline and thus a putative marker of structural brain health.^60–63^ In order to generate genetic variant association estimates with the whole-brain cortical macro-structure (surface area, volume, cortical thickness, intrinsic curvature, mean curvature, local gyrification index) and micro-structure phenotypes (mean diffusivity, fractional anisotropy, intracellular volume fraction, and orientation dispersion index), we conducted an in-house GWAS using the latest UK Biobank data release (Supplementary Methods). Genetic variant association estimates with the white matter micro-structure phenotypes were obtained from a recently published GWAS of diffusion MRI data by Zhao *et al*.^52^ For this study, we used summary statistics for the global fractional anisotropy and mean diffusivity phenotypes, which were averaged across 21 predefined cerebral white matter tracts. For hippocampal volume and the total volume of white matter hyperintensities, we obtained genetic variant association estimates from a recently published GWAS of 3,913 imaging-derived phenotypes by Smith *et al*.^51^

**Figure 2.**
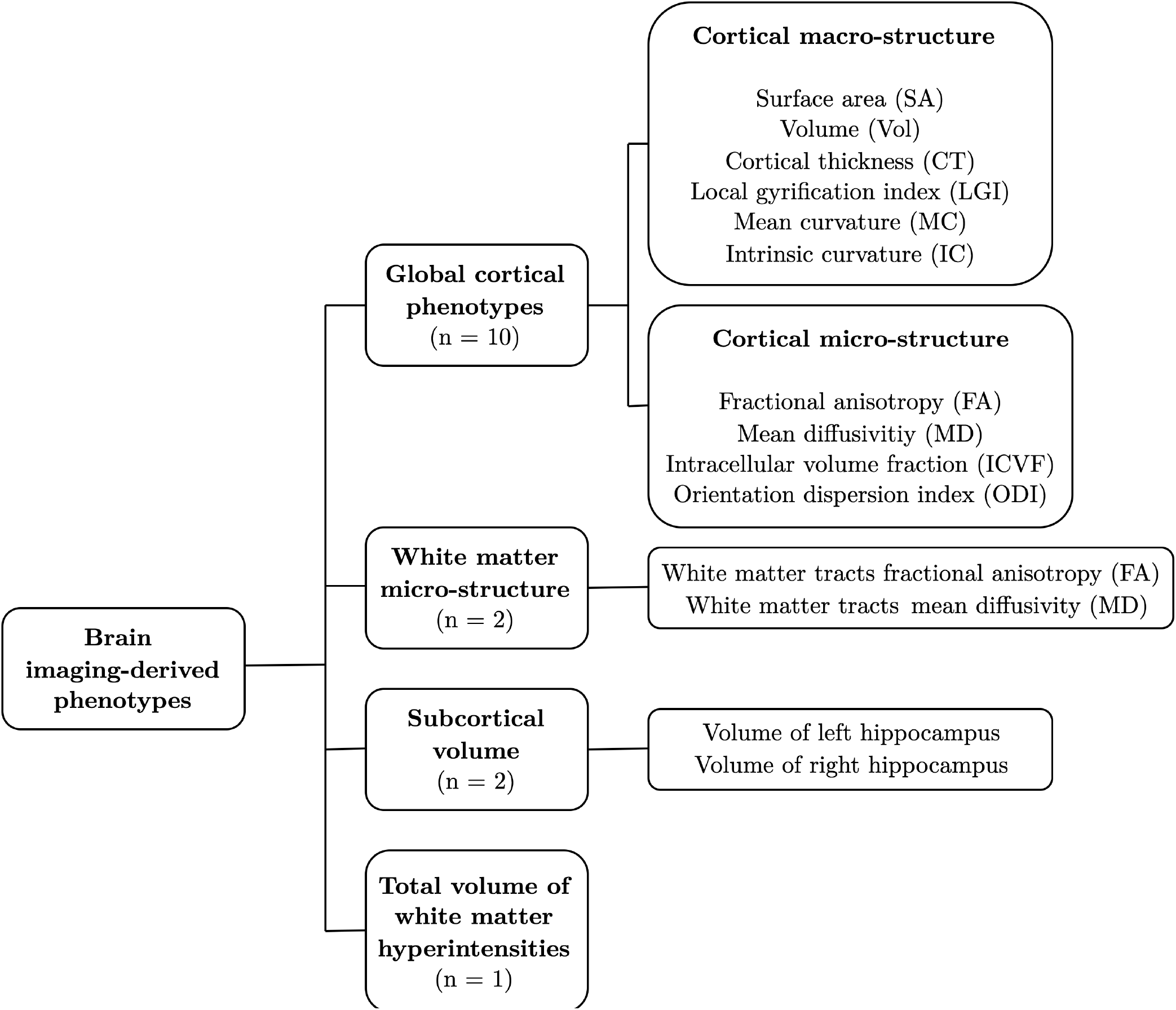
Overview of the brain imaging-derived phenotypes investigated in this study. Structural brain reserve was proxied by 15 brain imaging-derived phenotypes. These included ten global cortical phenotypes (six macro-structural metrics derived from T1-weighted structural MRI and four micro-structural metrics derived from diffusion MRI), two white matter micro-structural phenotypes (measured at 21 major white matter tracts across the whole brain), bilateral hippocampal volumes, and the total volume of white matter hyperintensities in the brain (derived from T2-weighted MRI).

### Selection of genetic instruments and data harmonization

For each MR analysis, we selected all single nucleotide polymorphisms (SNPs) associated with the exposure of interest at a genome-wide significance threshold (*P* < 5 × 10^−8^) as instrumental variables. The resulting instruments were pruned to near-independence using a linkage disequilibrium (LD) threshold of *r*^*2*^ < 0.001 over 10,000 kilobasepairs. We used the LD reference panel for the European super-population in the 1000 Genomes Project reference dataset, which was restricted to bi-allelic SNPs with minor allele frequency (MAF) > 0.01. We extracted the following summary-level data from each exposure and outcome GWAS: SNP rs number, effect and other alleles, effect allele frequency, sample size, number of cases/controls (if applicable), standardized β-coefficients, standard errors, and *p*-values. Where an instrument SNP was not available in the outcome dataset, we used LDlink^64^ to identify a proxy SNP in LD with the target SNP (*r*^*2*^ > 0.8). The exposure and outcome GWAS datasets were then harmonized to ensure that the genetic variant association estimates corresponded to the effect of the same allele. We used allele frequency information to infer the orientation of alleles in the exposure and outcome GWAS. Palindromic SNPs with MAF > 0.42 were dropped from the analysis.^65^ Details of the SNPs used as instruments in each MR analysis (including proxies for SNPs not available in the outcome datasets) are available in the Supplementary Data file. For each analysis, we used the online power calculator by Burgess^66^ (https://sb452.shinyapps.io/power/) to estimate the minimum causal effect that we had 80% power to detect at a multiple testing-corrected significance threshold (see the ‘Correction for multiple testing’ section below). Power calculations are presented in Supplementary Table 1.

### Statistical analyses

#### Bidirectional univariable Mendelian randomization

We performed univariable MR analyses to estimate each of the following total causal effects: (i) educational attainment on Alzheimer’s disease, (ii) educational attainment on each of the 15 imaging-derived phenotypes, and (iii) imaging-derived phenotypes on Alzheimer’s disease. To examine the direction of association and investigate the possibility of reverse causation, we additionally estimated each of the above causal effects in the direction opposite to that initially hypothesized (i.e., bidirectional MR).^67^ As recommended by Burgess *et al*.,^68^ we used inverse-variance weighted (IVW) MR^69^ with multiplicative random-effects as the primary analysis method, as it provides the most efficient combination of the variant-specific ratio estimates,^68– 70^ and accounts for heterogeneity in the causal estimates obtained from individual variants.^68,71^

### Correction for multiple testing

To account for the number of MR analyses performed to test for bidirectional effects of education on brain structure and brain structure on Alzheimer’s disease, we applied the multiple-testing correction approach described by Lord *et al*.^46^ Given the correlation structure between the various MRI metrics considered, applying a simple Bonferroni correction could be overly conservative. We therefore used a principal component analysis approach (https://github.com/hagax8/independent_tests) to estimate the number of independent hypotheses tested from the matrix of squared phenotypic correlations between the imaging-derived phenotypes (Supplementary Methods; Supplementary Fig. 1). We first calculated the number of principal components that explained 99.5% of the variance in the matrix as *N* = 13. The number of individual tests within the squared correlation matrix was then calculated as *T* = (*N*\**N* – *N*)/2 (=78), and the square root of *T* (=8.83) was used to establish the final number of individual imaging-derived phenotypes to correct for. Bidirectional associations between educational attainment and imaging-derived phenotypes and between imaging-derived phenotypes and Alzheimer’s disease were therefore considered significant at a multiple-testing corrected threshold of *P* < 0.006 (=0.05/8.83). For testing bidirectional causal relationships between educational attainment and Alzheimer’s disease, we set statistical significance at *P* < 0.05.

Note that the purpose of the multiple testing correction was to control the family-wise error rate (FWER) within each objective (i-iv), rather than across the total number of univariable MR analyses performed (i.e., 62). Controlling the FWER across all 62 tests would be appropriate if we were interested in the global null hypothesis (i.e., the intersection hypothesis of all null hypotheses of interest).^72^ Given that each of the objectives of interest were distinct (i.e., we were interested in detecting putative causal relationships between education and Alzheimer’s disease, education and brain structure, and brain structure and Alzheimer’s disease), we applied the multiple testing correction to the number of hypotheses within each objective.

#### Sensitivity analyses

For each univariable MR analysis where a causal effect was detected using the IVW method, we used the following four methods to assess the robustness of that finding to potential violations of MR assumptions: the Contamination Mixture method,^73^ Weighted Median MR,^74^ MR-Egger,^75^ and the Mendelian randomization pleiotropy residual sum and outlier (MR-PRESSO) method.^76^ These four robust methods were selected since they each produce a valid estimate of the causal effect of the exposure on the outcome under different assumptions (see Supplementary Methods for a description of the assumptions of each method).^77^

In addition to robust MR methods, we performed a range of other sensitivity analyses where a causal effect was detected using the IVW method. First, we assessed the presence of heterogeneity amongst the variant-specific causal estimates using Cochran’s *Q* statistic.^78,79^ Second, we generated funnel plots of the precision of the variant-specific causal estimates against the estimates themselves. The funnel plot is expected to be symmetrical about the IVW estimate, with more precise estimates having less variability.^80^ The presence of asymmetry in the funnel plot is suggestive of directional pleiotropy and bias in the overall causal estimate.^75,80^ Third, we carried out leave-one-out sensitivity analyses to identify influential data points amongst the instruments for each exposure, and to assess the reliance of the causal effect estimate on individual genetic variants.^81^ Finally, to orient the direction of causality between the exposure and the outcome, we used the Steiger directionality test,^82^ an extension to two-sample MR which detects variants that have a stronger association with the outcome than with the exposure. Where there was evidence from the Steiger test that some genetic instruments had a stronger association with the outcome, we repeated the analysis excluding those variants.^68^

#### Multivariable Mendelian randomization to test for mediation

We performed multivariable MR analyses^53,54^ to investigate the potential mediating role of brain structure in the relationship between educational attainment and Alzheimer’s disease. Multivariable MR allows the genetic variants associated with both the primary exposure and the secondary exposure (mediator) to be included as instruments in the analysis, and can be used to estimate the direct effect of the exposure on the outcome whilst adjusting for the mediator (Supplementary Methods; Supplementary Fig. 2).^53,54,83^ Using this approach, we estimated the direct causal effect of educational attainment on Alzheimer’s disease risk, while adjusting for each of the 15 imaging-derived phenotypes in turn. A difference between the estimate of the total causal effect of educational attainment on Alzheimer’s disease (from univariable MR) and the direct causal effect estimate (from the multivariable MR model including an imaging-derived phenotype) would indicate a mediating role of that brain structure phenotype on the causal pathway from education to Alzheimer’s disease.^83,84^ For each multivariable MR analysis, we selected all SNPs associated with the primary exposure (educational attainment) or the mediator (imaging-derived phenotype of interest) as instrumental variables. The pooled set of SNPs were clumped to pairwise LD (*r*^*2*^ < 0.001) over 10,000 kilobasepairs, based on the lowest *P*-value for association with any of the two traits. The extraction of summary-level data from the Alzheimer’s disease GWAS, harmonization of the exposure and outcome datasets, and identification of proxies for missing SNPs, followed the same procedure as univariable MR, as described above.

All statistical analyses were performed in R software (The R Foundation for Statistical Computing, Vienna, Austria).^85^ We used the “TwoSampleMR” package^65^ (v0.5.6) for data extraction and harmonization, the “ieugwasr” package (https://mrcieu.github.io/ieugwasr/index.html) for clumping, and the “LDlink” package^64^ (v1.1.2.9) for extracting LD proxies. All MR analyses were carried out using the “MendelianRandomization”^86^ (v0.5.1) and “MRPRESSO”^76^ (v1.0) packages.

### Ethical approval

The present study is a secondary analysis of publicly available data. Ethical approval was granted for each of the original GWAS studies, the details of which can be found in the respective publications.

### Data and code availability

Summary statistics from the in-house GWAS of cortical macro- and micro-structure are available on publication. Individual-level imaging and genetic data used for the in-house GWAS analyses may be requested through the UK Biobank (https://www.ukbiobank.ac.uk/), and the code used in generating the imaging phenotypes is available on GitHub (https://github.com/ucam-department-of-psychiatry/UKB). Summary-level genetic association estimates with all other phenotypes were obtained from publicly available published GWAS, and can be accessed from the respective publications. R code for reproducing all MR analyses can be found on https://github.com/as2970/EA_brain_AD_MR.

## Results

### Inverse association of genetically-predicted educational attainment with Alzheimer’s disease risk

There was strong evidence for an inverse association between genetically-proxied educational attainment and Alzheimer’s disease across all MR methods (Fig. 3; Supplementary Fig. 3). The results of the main IVW analysis suggested that each SD increase in genetically-predicted years of schooling (approximately 4.2 years) was associated with a 30% reduction in odds of Alzheimer’s disease (OR = 0.70, 95% CI 0.60, 0.80). The weighted median and MR-PRESSO methods provided similar estimates to the IVW analysis, while larger inverse estimates were observed in the contamination mixture and MR-Egger analyses (Fig. 3).

**Figure 3.**
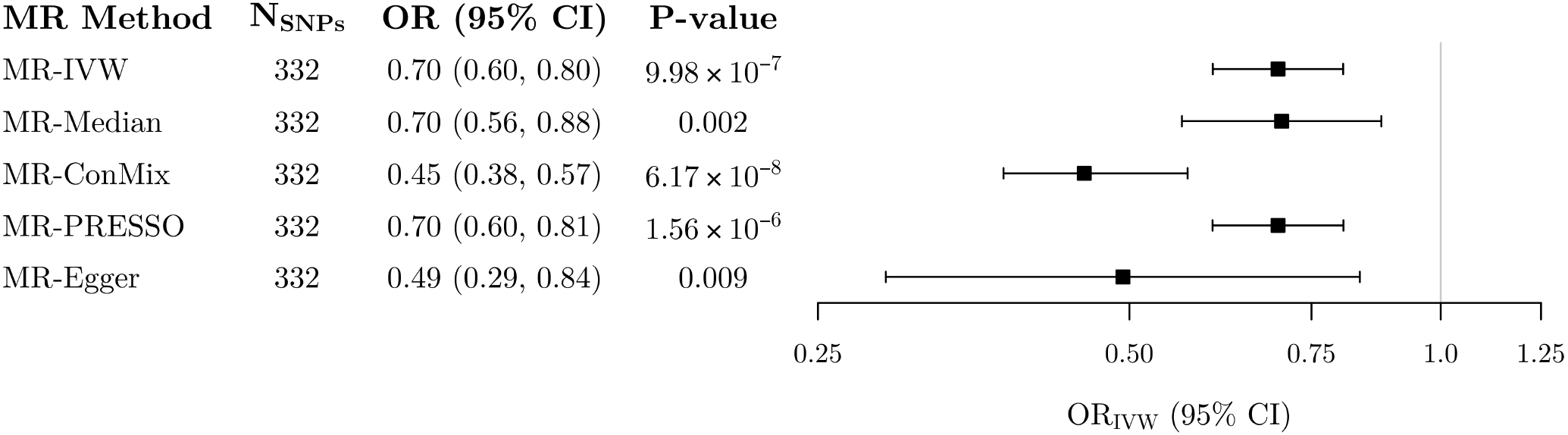
Mendelian randomization estimates of the association between genetically-proxied educational attainment and Alzheimer’s disease. Estimates represent odds ratios (95% confidence intervals) for late-onset Alzheimer’s disease per 1 standard deviation increase in genetically-predicted years of schooling (approximately 4.2 years). MR = Mendelian randomization; SNP = single nucleotide polymorphism; OR = odds ratio; CI = confidence interval; IVW = inverse-variance weighted.

Cochran’s *Q* statistic provided no evidence that there was more heterogeneity in the variant-specific causal estimates than expected due to chance (Supplementary Table 2). There was no evidence of directional pleiotropy using the MR-Egger intercept test (Supplementary Table 3), no evidence of departure from symmetry in the funnel plot (Supplementary Fig. 4), and no distortion in the leave-one-out plot (Supplementary Fig. 5). The Steiger directionality test indicated that 24 (7%) of the 332 education-associated SNPs explained more variance in Alzheimer’s disease than in educational attainment. When these variants were excluded from the analysis, there was still strong evidence for an inverse association of genetically-proxied education with Alzheimer’s disease (OR_IVW_ = 0.72, 95% CI 0.62, 0.83; Supplementary Table 4).

In the opposite direction, there was no association between genetically-predicted Alzheimer’s disease and years of education. The IVW estimate (representing the SD change in years of schooling per doubling the odds of Alzheimer’s disease) was 0.00 (95% CI 0.00, 0.01, *P* = 0.145).

### Association of genetically-predicted educational attainment with brain structure

After correction for multiple testing, genetically-proxied educational attainment was positively associated with four measures of cortical macro-structure (Fig. 4; Supplementary Fig. 6). In IVW analyses, each SD increase in genetically-predicted years of schooling was associated with a 0.30 SD increase in surface area (95% CI: 0.20, 0.40), 0.29 SD increase in cortical volume (95% CI: 0.20, 0.37), 0.18 SD increase in intrinsic curvature (95% CI: 0.11, 0.25), and 0.21 SD increase in local gyrification index (95% CI: 0.11, 0.31). In addition, inverse associations were observed with intracellular volume fraction (β_IVW_ = −0.09, 95% CI: −0.15, − 0.03) and the total volume of white matter hyperintensities (β_IVW_ = −0.14, 95% CI: −0.23, − 0.05). The results of the pleiotropy robust methods were broadly consistent with the IVW analyses (Supplementary Fig. 6). For local gyrification index, intracellular volume fraction, and white matter hyperintensities volume, confidence intervals around the MR-Egger estimate included the null. However, the point estimates were still in the same direction, and all other robust methods were in line with the IVW analysis.

**Figure 4.**
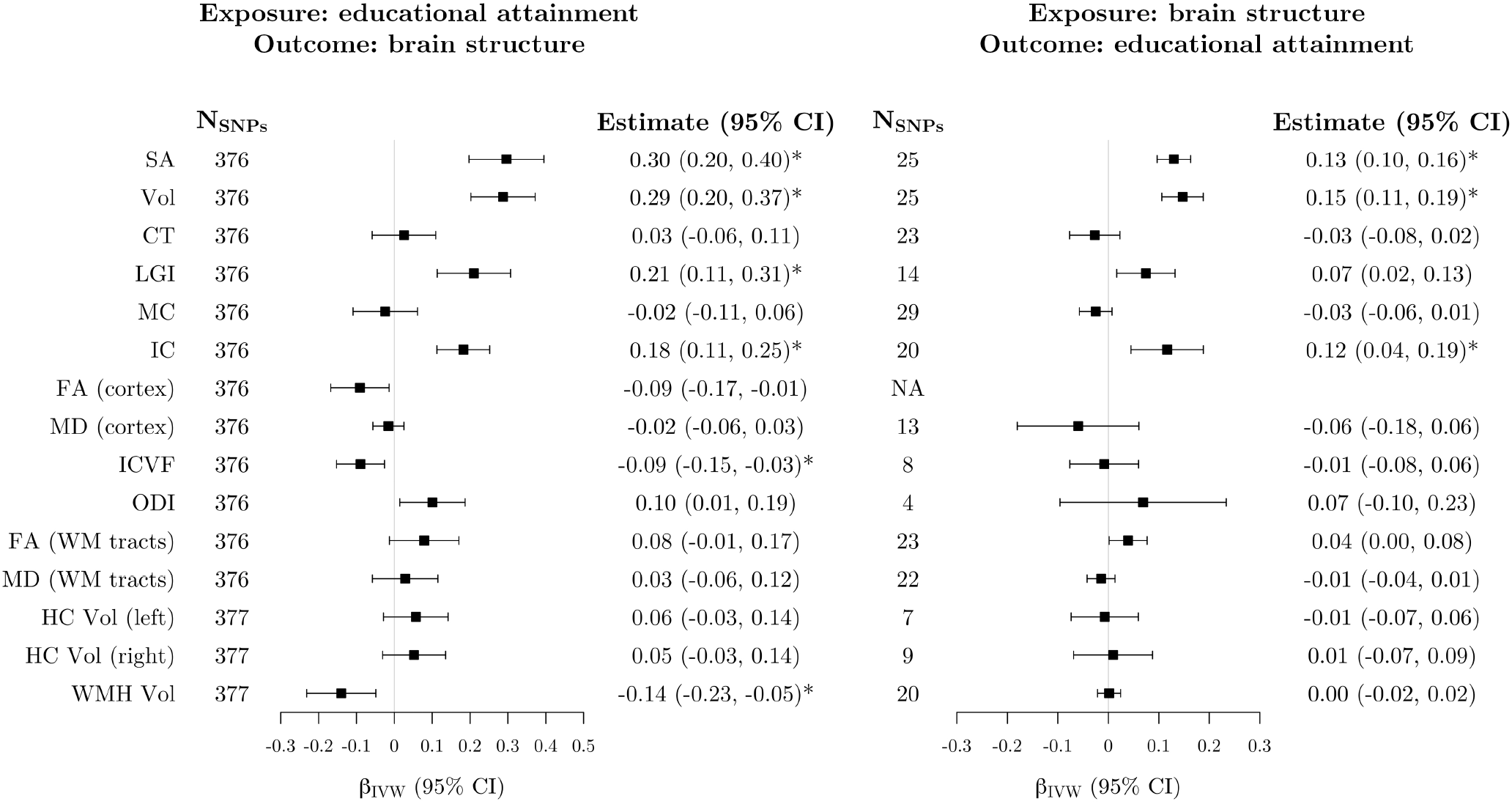
Mendelian randomization estimates of bidirectional associations between educational attainment and brain imaging-derived phenotypes. Left: estimates from inverse-variance weighted Mendelian randomization of the association between genetically-proxied educational attainment and imaging-derived brain structure phenotypes. Estimates represent the standard deviation change in the imaging phenotype per 1 standard deviation increase in genetically-predicted years of schooling (approximately 4.2 years). Right: estimates from inverse-variance weighted Mendelian randomization of the association between genetically-proxied brain structure phenotypes and educational attainment. Estimates represent the standard deviation change in years of schooling per 1 standard deviation increase in genetically-predicted levels of each imaging-derived phenotype. Estimates marked with an asterisk were significant after correction for multiple testing. The SNPs column represents the number of SNPs remaining after clumping for independence and data harmonization. There were no significant clumps for the FA (cortex) phenotype after LD pruning. SNP = single nucleotide polymorphism; CI = confidence interval; SA = surface area; Vol = volume; CT = cortical thickness; LGI = local gyrification index; MC = mean curvature; IC = intrinsic curvature; FA = fractional anisotropy; MD = mean diffusivity; ICVF = intracellular volume fraction; ODI = orientation dispersion index; WM = white matter; HC = hippocampus; WMH = white matter hyperintensities; IVW = inverse-variance weighted; LD = linkage disequilibrium.

All analyses demonstrated significant evidence of heterogeneity in the variant-specific causal estimates (Supplementary Table 2). The funnel plot for cortical surface area demonstrated a slight departure from symmetry, but all other funnel plots were symmetrical about the IVW estimate (Supplementary Figs 7–12). The MR-Egger intercept test did not show evidence of directional pleiotropy in any of the analyses (Supplementary Table 3), and there was no distortion in any of the leave-one-out plots (Supplementary Figs 13-18). The proportion of education-associated SNPs that explained more variance in the imaging phenotype than in educational attainment ranged from 14% for intracellular volume fraction to 30% for white matter hyperintensities volume (Supplementary Table 4). In sensitivity analyses excluding the invalid variants from each analysis, all estimates were attenuated towards the null, and the associations with local gyrification index and white matter hyperintensities volume were no longer statistically significant (Supplementary Table 4).

### Association of genetically-predicted brain structure with educational attainment

After correction for multiple testing, genetically-predicted levels of three cortical macro-structure phenotypes (surface area, volume, and intrinsic curvature) were positively associated with educational attainment (Fig. 4; Supplementary Fig. 19). The IVW estimates, representing the change in years of schooling (SD units) per 1 SD increase in genetically-predicted levels of imaging-derived phenotypes were 0.13 (95% CI 0.10, 0.16) for surface area, 0.15 (95% CI 0.11, 0.19) for volume, and 0.12 (95% CI 0.04, 0.19) for intrinsic curvature. For cortical surface area and volume, all robust methods provided similar estimates to the IVW analysis, but confidence intervals around the MR-Egger estimates were considerably wider than the other methods and included the null (Supplementary Fig. 19). The MR-Egger estimate of the association between genetically-predicted intrinsic curvature and educational attainment was negative, whereas all other estimates of this association were positive. However, the confidence interval for the MR-Egger estimate still overlapped with all other point estimates (Supplementary Fig. 19).

Using Cochran’s *Q* statistic, we found evidence of heterogeneity in all three analyses (Supplementary Table 2). However, the funnel plots showed little evidence of departure from symmetry (Supplementary Figs 20–22), and the MR-Egger intercept term did not significantly differ from zero in any analysis (Supplementary Table 3). In leave-one-out sensitivity analyses, the overall IVW estimates did not change upon exclusion of any variant (Supplementary Figs 23–25). The Steiger directionality test did not identify any SNPs that explained more variance in educational attainment than in the imaging phenotype for any analysis (Supplementary Table 4).

### Association of genetically-predicted Alzheimer’s disease with brain structure

Using the primary IVW analysis method, we did not find evidence for an association between genetically-predicted levels of any imaging-derived phenotype and Alzheimer’s disease (Fig. 5). However, genetically-predicted Alzheimer’s disease was associated with four measures of brain structure (Fig. 5; Supplementary Fig. 26), namely, reduced cortical orientation dispersion index (β_IVW_ = −0.02 [SD units per doubling the odds of genetically-proxied Alzheimer’s disease], 95% CI −0.03, −0.01), increased white matter mean diffusivity (β_IVW_ = 0.02, 95% CI 0.01, 0.04), and reduced hippocampal volume in both hemispheres (left hemisphere: β_IVW_ = −0.02, 95% CI −0.03, −0.01; right hemisphere: β_IVW_ = −0.02, 95% CI −0.04, −0.01). For left and right hippocampal volume, all robust methods provided similar estimates to the IVW analysis (Supplementary Fig. 26). For orientation dispersion index, the 95% CI for the contamination mixture estimate included the null, but the point estimate was identical to the other methods (Supplementary Fig. 26). The contamination mixture and MR-PRESSO methods both suggested a null association of genetically-predicted Alzheimer’s disease with white matter mean diffusivity (Supplementary Fig. 26).

**Figure 5.**
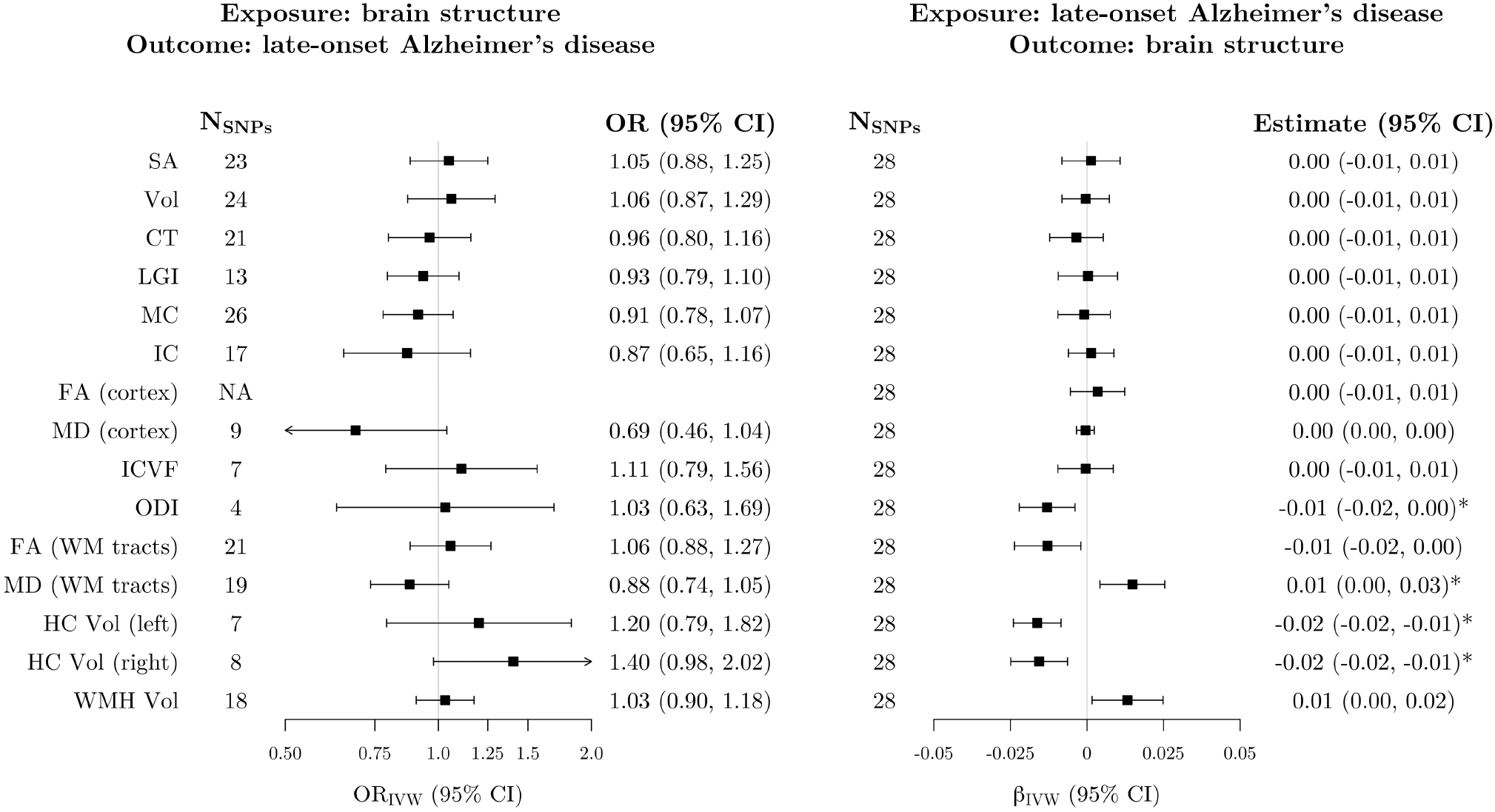
Mendelian randomization estimates of bidirectional associations between brain imaging-derived phenotypes and Alzheimer’s disease. Left: estimates from inverse-variance weighted Mendelian randomization of the association between genetically-proxied imaging-derived phenotypes and Alzheimer’s disease. Estimates represent the odds ratio for late-onset Alzheimer’s disease per standard deviation increase in genetically-predicted levels of each imaging-derived phenotype. Right: estimates from inverse-variance weighted Mendelian randomization of the association between genetically-proxied Alzheimer’s disease and brain structure phenotypes. Estimates represent the average change in each imaging-derived phenotype (standard deviation units) per doubling (2-fold increase) in the odds of genetically-predicted late-onset Alzheimer’s disease. Estimates marked with an asterisk were significant after correction for multiple testing. The SNPs column represents the number of SNPs remaining after clumping for independence and data harmonization. There were no significant clumps for the FA (cortex) phenotype after LD pruning. SNP = single nucleotide polymorphism; OR = odds ratio; CI = confidence interval; SA = surface area; Vol = volume; CT = cortical thickness; LGI = local gyrification index; MC = mean curvature; IC = intrinsic curvature; FA = fractional anisotropy; MD = mean diffusivity; ICVF = intracellular volume fraction; ODI = orientation dispersion index; WM = white matter; HC = hippocampus; WMH = white matter hyperintensities; IVW = inverse-variance weighted; LD = linkage disequilibrium.

We found evidence of heterogeneity in the variant-specific causal estimates for the association of genetically-proxied Alzheimer’s disease with white matter mean diffusivity (Supplementary Table 2). In addition, there was evidence of potential horizontal pleiotropy in this analysis using the MR-Egger intercept test (Supplementary Table 3). There was little evidence of departure from symmetry in the funnel plots (Supplementary Figs 27–30). In leave-one-out sensitivity analyses, the association of genetically-predicted Alzheimer’s disease with all four imaging-derived phenotypes appeared to be driven entirely by one genetic variant (rs12721046). In each case, exclusion of this variant from the analysis attenuated the IVW estimate towards the null (Supplementary Fig. 31). To further investigate the reliance of the IVW estimates on this variant, we conducted post-hoc single-SNP MR analyses and visualized the results using forest plots (Supplementary Fig. 32). Although the rs12721046 variant did not have an outlying MR estimate for any imaging-derived phenotype, the precision of its estimates was unusually high, causing this variant to dominate the wider polygenic signal in each case (Supplementary Fig. 32). In Steiger filtering sensitivity analyses, all instruments explained more variance in Alzheimer’s disease than in the imaging phenotypes (Supplementary Table 4).

### Mediation analysis

In multivariable MR analyses, there was little change in the association of genetically-predicted years of schooling with Alzheimer’s disease after adjusting for genetically-predicted levels of each of the 15 imaging-derived phenotypes (Table 2).

**Table 2.** Mendelian randomization estimates of the association between genetically-proxied educational attainment and Alzheimer’s disease risk, estimated with no adjustment and after adjustment for genetically-proxied levels of brain imaging-derived phenotypes.

| **Adjustment** | **OR<sub>IVW</sub> (95% CI)** | **P-value** |
| --- | --- | --- |
| None (univariable analysis) | 0.70 (0.60, 0.80) | $9.98 \times 10^{-7}$ |
| **Cortical macro-structure** |  |  |
| Surface area | 0.71 (0.62, 0.82) | $3.68 \times 10^{-6}$ |
| Volume | 0.72 (0.62, 0.83) | $6.12 \times 10^{-6}$ |
| Cortical thickness | 0.69 (0.60, 0.80) | $3.62 \times 10^{-7}$ |
| Local gyrification index | 0.71 (0.61, 0.81) | $1.39 \times 10^{-6}$ |
| Mean curvature | 0.69 (0.60, 0.79) | $1.80 \times 10^{-7}$ |
| Intrinsic curvature | 0.71 (0.62, 0.82) | $2.74 \times 10^{-6}$ |
| **Cortical micro-structure** |  |  |
| Mean diffusivity (cortex) | 0.70 (0.61, 0.81) | $9.35 \times 10^{-7}$ |
| Intracellular volume fraction | 0.71 (0.62, 0.82) | $2.72 \times 10^{-6}$ |
| Orientation dispersion index | 0.70 (0.61, 0.81) | $8.65 \times 10^{-7}$ |
| **White matter micro-structure** |  |  |
| Fractional anisotropy (white matter tracts) | 0.71 (0.61, 0.81) | $1.53 \times 10^{-6}$ |
| Mean diffusivity (white matter tracts) | 0.71 (0.61, 0.82) | $2.42 \times 10^{-6}$ |
| **Subcortical volume** |  |  |
| Volume of left hippocampus | 0.70 (0.61, 0.80) | $5.66 \times 10^{-7}$ |
| Volume of right hippocampus | 0.70 (0.61, 0.81) | $7.74 \times 10^{-7}$ |
| **White matter hyperintensities volume** |  |  |
| Total volume of white matter hyperintensities | 0.71 (0.61, 0.81) | $1.43 \times 10^{-6}$ |

Odds ratios (95% confidence intervals) for Alzheimer's disease are estimated per 1 SD increase in genetically-predicted years of schooling (approximately 4.2 years), estimated with no adjustment (univariable IVW analysis) and after adjustment for genetically-predicted levels of each brain imaging-derived phenotype in turn (multivariable IVW analyses). OR = odds ratio; IVW = inverse-variance weighted; CI = confidence interval.

## Discussion

We conducted bidirectional univariable and multivariable Mendelian randomization analyses using large genome-wide association datasets to examine putative causal relationships between educational attainment, brain macro- and micro-structure as measured by MRI, and risk of Alzheimer’s disease. We found strong evidence in support of a protective causal effect of educational attainment on Alzheimer’s disease risk, as well as evidence supporting bidirectional causal relationships between education and several measures of cortical macro- and micro-structure. We did not find any evidence in support of a causal effect of brain structure on Alzheimer’s disease, or for the hypothesis that the protective effect of increased education on Alzheimer’s risk is mediated via changes to brain structure. Although we found evidence for an association of genetically-predicted Alzheimer’s disease with hippocampal volume, as well as cortical and white-matter micro-structure, these effects appeared to be driven entirely by one variant in the *APOC1* gene region, and may therefore represent horizontally pleiotropic effects of this variant, rather than a causal effect of Alzheimer’s disease on brain structure.

Our finding of a protective causal effect of education on Alzheimer’s disease risk is consistent with the large body of observational epidemiological literature.^1–5,7^ In the current study, the IVW estimate suggested that every additional 4.2 years of schooling reduced the odds of Alzheimer’s disease by approximately 30%. This effect size is broadly consistent with the findings of a dose-response meta-analysis, in which every additional year of schooling was associated with a 7% reduction in risk of all-cause dementia.^87^ Our results are also consistent with previous MR evidence supporting a causally protective effect of education on Alzheimer’s disease risk.^43–47^ Evidence from three recent multivariable MR studies suggests that the protective effect of educational attainment on Alzheimer’s disease is largely mediated via increased intelligence levels, rather than an independent protective effect of years of schooling.^43,46,88^ This finding is consistent with evidence from a large meta-analysis of quasi-experimental studies,^89^ in which every additional year of education was estimated to be associated with an increase of approximately 1-5 standardized IQ points, a result which was consistent across the lifespan and across broad categories of cognitive ability. Nevertheless, even if the protective effect of educational attainment on Alzheimer’s disease risk is entirely mediated via intelligence, increasing the number of years of schooling could still be an effective public health strategy for reducing the incidence of Alzheimer’s disease, as education has been suggested to be the most durable, robust, and consistent method for increasing intelligence levels.^89^ Findings from a recent quasi-experimental study suggest that increasing the school-leaving age in the UK in 1972 has led to improvements in IQ as well as a range of health and well-being outcomes, including a reduced risk of diabetes and all-cause mortality.^90^ Our results suggest that education policy reforms that target the length of compulsory schooling are also likely to be effective for the primary prevention of Alzheimer’s disease at the population level.^43,47^

We found evidence for bidirectional causal relationships between educational attainment and three global macro-structural markers of cortical morphology, namely, volume, surface area, and intrinsic curvature. These findings are in agreement with the existing observational neuroimaging literature on the association between educational attainment and brain morphology,^24,26,32^ as well as evidence for positive genetic correlations between education and MRI markers of cortical macro-structure.^49,91^ To investigate the direction of causation in these associations, we performed sensitivity analyses using Steiger filtering where we excluded any SNPs that explained more variance in the outcome than in the exposure. Although the magnitude of the effect size attenuated in all analyses following the removal of these SNPs, all causal effect estimates remained statistically significant, suggesting that the relationship between educational attainment and changes in macro-structural markers is indeed likely to be bidirectional. We also found evidence for causal effects of educational attainment on local gyrification index and on the total volume of white matter hyperintensities. However, when we removed variants that explained more variance in the outcome than in the exposure, these effects were no longer statistically significant, suggesting that they may be explained by reverse causation or horizontal pleiotropy. As white matter hyperintensities mainly develop in elderly individuals,^63^ reverse causation is unlikely in this case, and the association with educational attainment likely reflects horizontal pleiotropy.

Across the 15 imaging-derived phenotypes examined, we did not find any evidence of a causal effect of brain structure on Alzheimer’s disease risk. In addition, our results provide no support for the hypothesis that the protective effect of education on Alzheimer’s disease is mediated via structural alterations in the brain that increase tolerance to Alzheimer’s pathology (i.e., the brain reserve hypothesis).^10,11,22^ It is possible that capturing the neural basis of brain reserve requires more granular and specific MRI metrics than the ones used in the current study. We reasoned that any putative protective effects of education on Alzheimer’s risk would be unlikely to be manifest only in a highly regionally-specific manner, and hence decided to primarily study global rather than localized imaging phenotypes. Nevertheless, the use of global imaging metrics in the current study may have masked any significant regionally-specific causal effects of brain macro- or micro-structure on Alzheimer’s disease risk.^32^ It is also plausible that the protective effect of education on Alzheimer’s disease is better explained by the related concept of ‘cognitive reserve’, which refers to the capacity of the brain to cope with pathology through more efficient use of pre-existing cognitive networks or via recruitment of compensatory neural pathways.^9,22,92^ Cognitive reserve is a dynamic and active process of adaptation, and is typically measured using behavioral testing or functional neuroimaging modalities (e.g., resting state or task-related networks of brain activation that moderate the effect of Alzheimer’s pathology on cognition).^9,93^Although the concepts of cognitive and brain reserve are not mutually exclusive and are hypothesized to operate synergistically in moderating the effect of Alzheimer’s disease pathology on clinical outcome,^9,22^ it is possible that the protective effect of education on Alzheimer’s disease risk is largely mediated via increased cognitive reserve.^93^ Therefore, the use of functional, rather than structural, MRI phenotypes might be better suited to capturing such mediating effects.

In the reverse direction, we found evidence for an association of genetically-predicted Alzheimer’s disease with cortical orientation dispersion index, mean diffusivity in the white matter tracts, and bilateral hippocampal volumes. However, these results were entirely driven by a single intron variant in the *APOC1* gene region (rs12721046), which has been associated with cerebral amyloid deposition as measured by PET imaging in a recent GWAS.^94^ The *APOC1* gene encodes a member of the apolipoprotein C1 family, which plays a central role in the regulation of lipid levels and metabolism.^95^ It is located in a cluster on chromosome 19, approximately 5 kilobasepairs downstream from the *APOE* gene,^95^ where common genetic variation is known to have a relatively large effect on Alzheimer’s disease risk.^96,97^ A recent fine-mapping study of the *APOE* and surrounding regions using whole-genome sequencing data identified a cluster of risk variants (including rs12721046) in the *APOC1* region as potential causal variants for Alzheimer’s disease.^98^ This cluster of variants were found to confer increased risk of Alzheimer’s independent of the *APOE*-ε4 genotype.^98^ In all four analyses, the precision of the variant-specific causal estimate for rs12721046 was substantially higher than the remaining instruments, causing this variant to dominate the IVW estimate. When this variant was removed from the analysis, the overall causal effect estimate was attenuated towards the null and was no longer significant. Since all causal effects were only evidenced by the rs12721046 variant, they may represent horizontally pleiotropic pathways from this variant to brain structure. Therefore, we cannot draw definitive conclusions regarding the effect of genetic liability to Alzheimer’s disease on brain structure.

Our findings should be interpreted in the context of some limitations. First, although we have used imaging-derived phenotypes as markers of structural brain reserve, we cannot determine the extent to which these phenotypes capture pre-morbid brain reserve, as opposed to neuropathological changes secondary to disease (e.g., atrophy secondary to Alzheimer’s disease).^22^ Given the age range of the UK Biobank imaging sample (45–82 years),^51^ the possibility that some individuals in the imaging GWAS harbor pre-clinical Alzheimer’s disease cannot be excluded. Using both self-reported diagnoses and International Classification of Diseases^99^ codes, we found that only 0.6% of participants in the UK Biobank have a current Alzheimer’s diagnosis. Therefore, it is unlikely that a large proportion of the imaging GWAS sample had current or imminent Alzheimer’s disease at the time of scanning. Nevertheless, with increasing duration of follow-up, a substantial proportion of these participants may go on to develop Alzheimer’s or other dementias. Second, causal effect estimates in our analyses might be affected by Winner’s curse bias, which occurs when the same dataset is used to select the genetic variants as instrumental variables and to estimate their association with the exposure.^100^ Although this bias could be avoided by using three non-overlapping datasets from the same underlying population for genetic discovery and the estimation of variant-exposure and variant-outcome associations, restricting our analyses to non-overlapping datasets for the phenotypes examined would have substantially reduced the sample sizes. Therefore, a compromise was necessary to balance the risk of bias against imprecision of the causal estimates.^101^ Third, although there were no overlapping samples in the analyses of education and brain structure on Alzheimer’s disease, 39% of participants in the educational attainment GWAS and all of those in the brain imaging GWAS were recruited from the UK Biobank. Overlap between the exposure and outcome datasets in two-sample MR can exacerbate bias due to weak instruments,^68^ with the magnitude of bias being a linear function of the degree of overlap between the two samples.^102^ However, brain imaging is a recent addition to the UK Biobank data collection protocol, and less than 10% of the full UK Biobank cohort contribute to the imaging sample used in the present study. Even if all participants from the UK Biobank imaging sample were included in the educational attainment GWAS, the percentage overlap between the exposure and outcome datasets in the current study would range from 2.8% to 2.9% for the education → brain structure analyses, and from 4.1% to 4.3% for the brain structure → education analyses, suggesting that the extent of sample overlap in the current study is minimal. Therefore, it is unlikely that the use of completely non-overlapping samples would have changed the direction or magnitude of our results. Fourth, to minimize the possibility of confounding due to population stratification, we limited our analyses to participants of European ancestry. Therefore, it remains unclear whether our findings are applicable to other populations. Finally, the GWAS data sources used for brain imaging phenotypes had relatively small sample sizes. Although larger GWAS of brain structure are available,^49,103^ these are meta-analyses of MRI data from different imaging cohorts. The use of uniform genotyping and neuroimaging protocols in the UK Biobank and the application of consistent genetic and phenotypic quality control pipelines to the data used in the current study is likely to have resulted in lower measurement error and enhanced power, despite the slightly smaller sample size. Nevertheless, statistical power calculations indicated that our analyses may have been underpowered to detect particularly subtle causal effects of brain structure on Alzheimer’s risk (Supplementary Table 1). The imaging sample on which the present study is based represents only 40% of the eventual UK Biobank imaging cohort size.^51^ As data from more participants become available, replication of these findings using larger and better-powered GWAS of imaging-derived phenotypes will enable us to draw more definitive conclusions regarding putative causal associations between education, brain structure, and Alzheimer’s disease.

In conclusion, our findings add to the extensive body of observational epidemiological literature, as well as evidence from a growing number of MR investigations, providing support for a causally protective role of increased educational attainment on risk of Alzheimer’s disease. In addition, our results support bidirectional causal associations between education and several aspects of cortical macro- and micro-structure. However, we found limited evidence to suggest that these structural alterations have down-stream effects on Alzheimer’s disease risk. Our findings therefore provide little support for the hypothesis that changes in brain structure mediate the protective effect of education on Alzheimer’s risk through determining the underlying brain reserve of the individual.

## Supporting information

STROBE-MR Checklist

## Data Availability

Summary statistics from the in-house GWAS of cortical macro- and micro-structure are available on publication. Individual-level imaging and genetic data used for the in-house GWAS analyses may be requested through the UK Biobank (https://www.ukbiobank.ac.uk/), and the code used in generating the imaging phenotypes is available on GitHub (https://github.com/ucam-department-of-psychiatry/UKB). Summary-level genetic association estimates with all other phenotypes were obtained from publicly available published GWAS, and can be accessed from the respective publications as cited in the article. R code for reproducing all MR analyses can be found on https://github.com/as2970/EA_brain_AD_MR.

## Abbreviations

CI: confidence interval
FWER: family-wise error rate
GWAS: genome-wide association study
IGAP: International Genomics of Alzheimer’s Project
IQ: intelligence quotient
IVW: inverse-variance weighted
LD: linkage disequilibrium
MAF: minor allele frequency
MR: Mendelian randomization
MR-PRESSO: Mendelian randomization pleiotropy residual sum and outlier
OR: odds ratio
SD: standard deviation
SNP: single nucleotide polymorphism

## Supplementary material

Supplementary material is available at *Brain* online.

## Author contributions

AS: Conceptualization, Methodology, Formal Analysis, Writing – Original Draft, Review & Editing. VW: Conceptualization, Methodology, Formal analysis, Writing – Review & Editing. RAIB: Conceptualization, Methodology, Formal analysis, Writing – Review & Editing. BIP: Conceptualization, Methodology, Writing – Review & Editing. SB: Conceptualization, Methodology, Writing – Review & Editing, Supervision. GKM: Conceptualization, Methodology, Writing – Original Draft, Review & Editing, Supervision.

