## Supplementary Data for "Educational attainment, structural brain reserve, and Alzheimer’s disease: a Mendelian randomization analysis"

**List of proxies for imaging-derived phenotypes instruments**

| **Exposure** | **N_SNPs_ (after clumping)** | **Outcome** | | | |
| --- | --- | --- | --- | --- | --- |
|  |  | **Late-onset Alzheimer's disease** | | **Educational attainment** | |
|  |  | **SNP missing from outcome GWAS** | **Proxy SNP** | **SNP missing from outcome GWAS** | **Proxy SNP** |
| Surface area | 26 | rs1712512 | rs1712509 | rs9266618 | NA |
|  |  |  |  | rs1712512 | rs1712509 |
|  |  |  |  | rs56269136 | rs113856644 |
| Volume | 26 | – | – | rs73100947 | rs7659195 |
|  |  |  |  | rs3130346 | rs3130342 |
|  |  |  |  | rs2529418 | rs2244480 |
|  |  |  |  | rs62057121 | rs79501144 |
| Cortical thickness | 24 | – | – | rs7219015 | rs7218235 |
|  |  |  |  | rs12150230 | rs62059005 |
| Intrinsic curvature | 21 | rs201872292 | rs72714147 | rs2362111 | rs147431626 |
|  |  |  |  | rs2362111 | rs72714147 |
|  |  |  |  | rs147431626 | rs55686102 |
| Local gyrification index | 14 | – | – | rs147431626 | rs55686102 |
| Mean curvature | 29 | – | – | rs4652758 | rs4652757 |
|  |  |  |  | rs10197724 | rs13399586 |
|  |  |  |  | rs55976523 | rs7592770 |
|  |  |  |  | rs7219015 | rs7218235 |
| Intracellular volume fraction | 11 | – | – | rs9272456 | rs9272461 |
| Orientation dispersion index | 4 | rs200261326 | rs183388514 | rs7219015 | rs7218235 |
|  |  |  |  | rs79172804 | rs146746174 |
|  |  |  |  | rs200261326 | rs61218414 |
| Fractional anisotropy (cortical) | No significant clumps | – | – | – | – |
| Mean diffusivity (cortical) | 13 | rs192856660 | rs79523636 | rs192856660 | rs55686102 |
| Fractional anisotropy (white matter) | 26 | rs28376477 | NA | rs186399184 | rs147361301 |
|  |  | rs28361101 | rs28361098 | rs3129787 | NA |
|  |  |  |  | rs28361101 | rs28361098 |
| Mean diffusivity (white matter) | 26 | rs9692176 | NA | rs3116836 | NA |
|  |  | rs372295105 | rs10103291 | rs372295105 | rs10103291 |
|  |  | rs201911236 | NA | rs201911236 | NA |
|  |  |  |  | rs62071596 | rs1378358 |
| Volume of left hippocampus | 8 | – | – | rs150434736 | rs62288104 |
|  |  |  |  | rs188402627 | rs7137149 |
|  |  |  |  | rs593626 | NA |
| Volume of right hippocampus | 9 | – | – | rs150434736 | rs62288104 |
|  |  |  |  | rs2732706 | rs55686102 |
| White matter hyperintensities volume | 20 | rs554632245 | rs1983386 | rs554632245 | rs1983386 |
|  |  |  |  | rs10445411 | rs9899649 |

**List of proxies for educational attainment instruments**

| **Exposure** | **N_SNPs_ (after clumping)** | **Outcome** | **SNP missing from outcome GWAS** | **Proxy SNP** |
| --- | --- | --- | --- | --- |
| Educational attainment  Educational attainment  Educational attainment | 393  393  393 | Late-onset Alzheimer's disease | rs72486027 | rs968454 |
|  |  | Surface area | rs11678980 | NA |
|  |  |  | rs73581580 | NA |
|  |  |  | rs72622559 | rs72622558 |
|  |  |  | rs72486027 | rs968454 |
|  |  | Volume | rs11678980 | NA |
|  |  |  | rs73581580 | NA |
|  |  |  | rs72622559 | rs72622558 |
|  |  |  | rs72486027 | rs968454 |
|  |  | Cortical thickness | rs11678980 | NA |
|  |  |  | rs73581580 | NA |
|  |  |  | rs72622559 | rs72622558 |
|  |  |  | rs72486027 | rs968454 |
|  |  | Intrinsic curvature | rs11678980 | NA |
|  |  |  | rs73581580 | NA |
|  |  |  | rs72622559 | rs72622558 |
|  |  |  | rs72486027 | rs968454 |
|  |  | Local gyrification index | rs11678980 | NA |
|  |  |  | rs73581580 | NA |
|  |  |  | rs72622559 | rs72622558 |
|  |  |  | rs72486027 | rs968454 |
|  |  | Mean curvature | rs11678980 | NA |
|  |  |  | rs73581580 | NA |
|  |  |  | rs72622559 | rs72622558 |
|  |  |  | rs72486027 | rs968454 |
|  |  | Intracellular volume fraction  Intracellular volume fraction | rs11678980 | NA |
|  |  |  | rs73581580 | NA |
|  |  |  | rs72622559 | rs72622558 |
|  |  |  | rs72486027 | rs968454 |
|  |  | Orientation dispersion index | rs11678980 | NA |
|  |  |  | rs73581580 | NA |
|  |  |  | rs72622559 | rs72622558 |
|  |  |  | rs72486027 | rs968454 |
|  |  | Fractional anisotropy (cortical) | rs11678980 | NA |
|  |  |  | rs73581580 | NA |
|  |  |  | rs72622559 | rs72622558 |
|  |  |  | rs72486027 | rs968454 |
|  |  | mean diffusivity (cortical) | rs11678980 | NA |
|  |  |  | rs73581580 | NA |
|  |  |  | rs72622559 | rs72622558 |
|  |  |  | rs72486027 | rs968454 |
|  |  | Fractional anisotropy (white matter) | rs11678980 | NA |
|  |  |  | rs73581580 | NA |
|  |  |  | rs72622559 | rs72622558 |
|  |  |  | rs72486027 | rs968454 |
|  |  | Mean diffusivity (white matter) | rs11678980 | NA |
|  |  |  | rs73581580 | NA |
|  |  |  | rs72622559 | rs72622558 |
|  |  |  | rs72486027 | rs968454 |
|  |  | Volume of left hippocampus | rs73581580 | NA |
|  |  |  | rs72486027 | rs968454 |
|  |  | Volume of right hippocampus | rs73581580 | NA |
|  |  |  | rs72486027 | rs968454 |
|  |  | White matter hyperintensities volume | rs73581580 | NA |
|  |  |  | rs72486027 | rs968454 |

**List of proxies for Alzheimer's disease instruments**

| **Exposure** | **N_SNPs_ (after clumping)** | **Outcome** | **SNP missing from outcome GWAS** | **Proxy SNP** |
| --- | --- | --- | --- | --- |
| Late-onset Alzheimer's disease | 29 | Late-onset Alzheimer's disease | rs34665982 | NA |
|  |  | Surface area | – | – |
|  |  | Volume | – | – |
|  |  | Cortical thickness | – | – |
|  |  | Intrinsic curvature | – | – |
|  |  | Local gyrification index | – | – |
|  |  | Mean curvature | – | – |
|  |  | Intracellular volume fraction | – | – |
|  |  | Orientation dispersion index | – | – |
|  |  | Fractional anisotropy (cortical) | – | – |
|  |  | mean diffusivity (cortical) | – | – |
|  |  | Fractional anisotropy (white matter) | – | – |
|  |  | Mean diffusivity (white matter) | – | – |
|  |  | Volume of left hippocampus | – | – |
|  |  | Volume of right hippocampus | – | – |
|  |  | White matter hyperintensities volume | – | – |

**SNPs used as instrumental variables in the MR analysis of educational attainment on late-onset Alzheimer's disease**

| **SNP** | **Effect allele** | **Other allele** | **Effect allele frequency** | **beta (exposure)** | **SE (exposure)** | ***P* (exposure)** | **beta (outcome)** | **SE (outcome)** | ***P* (outcome)** | ***F*-statistic** |
| --- | --- | --- | --- | --- | --- | --- | --- | --- | --- | --- |
| rs10006235 | T | C | 0.27 | -0.009 | 0.002 | 4.48E-09 | 0.015 | 0.016 | 0.347 | 33.79 |
| rs10021733 | T | C | 0.15 | -0.016 | 0.003 | 9.45E-10 | -0.001 | 0.02 | 0.968 | 36.89 |
| rs10046069 | A | G | 0.12 | 0.013 | 0.002 | 4.40E-09 | -0.027 | 0.022 | 0.22 | 33.32 |
| rs10060023 | T | C | 0.33 | 0.011 | 0.002 | 1.88E-13 | -0.022 | 0.016 | 0.157 | 53.78 |
| rs10080647 | A | C | 0.14 | 0.012 | 0.002 | 7.05E-10 | -0.01 | 0.02 | 0.629 | 38.44 |
| rs1008078 | T | C | 0.4 | -0.018 | 0.001 | 4.50E-37 | 0.02 | 0.015 | 0.168 | 169 |
| rs10189857 | A | G | 0.57 | 0.016 | 0.001 | 4.69E-29 | -0.017 | 0.015 | 0.24 | 127.37 |
| rs10215082 | A | G | 0.47 | -0.015 | 0.001 | 7.03E-26 | -0.016 | 0.015 | 0.275 | 113.27 |
| rs10460095 | A | G | 0.57 | -0.012 | 0.001 | 2.61E-18 | 0.011 | 0.015 | 0.435 | 78.45 |
| rs10773208 | T | C | 0.25 | -0.011 | 0.002 | 8.09E-12 | -0.02 | 0.016 | 0.217 | 47.27 |
| rs10798888 | T | G | 0.17 | -0.014 | 0.002 | 5.15E-14 | 0.042 | 0.02 | 0.031 | 54.29 |
| rs10799615 | A | G | 0.25 | -0.009 | 0.002 | 4.11E-08 | -0.012 | 0.016 | 0.481 | 30.94 |
| rs10805383 | A | G | 0.49 | -0.01 | 0.001 | 2.22E-13 | -0.018 | 0.014 | 0.2 | 54.13 |
| rs10830858 | T | C | 0.51 | 0.009 | 0.001 | 5.31E-11 | 0.016 | 0.014 | 0.274 | 43.18 |
| rs10844179 | A | G | 0.23 | 0.01 | 0.002 | 5.03E-10 | 0.028 | 0.017 | 0.104 | 37.43 |
| rs10892807 | T | C | 0.57 | 0.011 | 0.001 | 1.09E-14 | 0.015 | 0.015 | 0.333 | 60.62 |
| rs10940921 | T | G | 0.42 | 0.011 | 0.001 | 1.31E-14 | 0.006 | 0.015 | 0.701 | 62.86 |
| rs10951590 | T | C | 0.33 | -0.011 | 0.002 | 3.62E-13 | -0.007 | 0.015 | 0.656 | 52.8 |
| rs10979613 | T | C | 0.64 | -0.012 | 0.002 | 1.25E-15 | 0.002 | 0.015 | 0.902 | 60.84 |
| rs11003463 | T | G | 0.61 | 0.009 | 0.002 | 5.58E-10 | -0.015 | 0.016 | 0.33 | 36.8 |
| rs11023764 | A | G | 0.64 | 0.009 | 0.002 | 5.32E-10 | -0.016 | 0.016 | 0.302 | 36.8 |
| rs11076962 | T | C | 0.72 | 0.009 | 0.002 | 4.29E-08 | -0.01 | 0.016 | 0.521 | 28.89 |
| rs11081529 | T | C | 0.71 | 0.013 | 0.002 | 5.22E-16 | -0.032 | 0.016 | 0.049 | 70.56 |
| rs11082011 | T | C | 0.67 | 0.019 | 0.002 | 2.06E-38 | 0.007 | 0.016 | 0.641 | 167.27 |
| rs11121177 | A | G | 0.18 | 0.015 | 0.002 | 1.44E-16 | 0.003 | 0.018 | 0.858 | 70.37 |
| rs11130380 | T | G | 0.61 | 0.009 | 0.001 | 4.17E-10 | 0.017 | 0.015 | 0.243 | 41.33 |
| rs11138947 | T | C | 0.72 | 0.011 | 0.002 | 2.36E-11 | 0.012 | 0.016 | 0.447 | 43.07 |
| rs11157931 | A | C | 0.39 | -0.014 | 0.001 | 3.86E-22 | 0.014 | 0.015 | 0.337 | 98.58 |
| rs111852224 | T | C | 0.12 | 0.016 | 0.002 | 4.85E-13 | -0.044 | 0.023 | 0.057 | 50.28 |
| rs11211123 | A | G | 0.22 | 0.01 | 0.002 | 7.62E-10 | 0.007 | 0.017 | 0.687 | 37.43 |
| rs11213482 | A | G | 0.84 | -0.011 | 0.002 | 2.68E-08 | 0.011 | 0.022 | 0.622 | 31.12 |
| rs112603734 | A | C | 0.77 | -0.013 | 0.002 | 2.72E-09 | 0.004 | 0.017 | 0.787 | 35.46 |
| rs1128956 | T | G | 0.83 | -0.014 | 0.002 | 5.34E-14 | 0.005 | 0.02 | 0.803 | 54.29 |
| rs113520408 | A | G | 0.28 | 0.016 | 0.002 | 1.03E-22 | 0.024 | 0.016 | 0.131 | 93.85 |
| rs113615161 | T | C | 0.13 | -0.012 | 0.002 | 2.98E-09 | 0.02 | 0.021 | 0.34 | 34.31 |
| rs11542663 | A | C | 0.69 | 0.011 | 0.002 | 2.44E-13 | -0.026 | 0.016 | 0.093 | 55.75 |
| rs11588857 | A | G | 0.21 | 0.02 | 0.002 | 4.45E-31 | 0.013 | 0.017 | 0.461 | 137.03 |
| rs11598765 | A | G | 0.2 | -0.011 | 0.002 | 1.73E-09 | -0.002 | 0.018 | 0.93 | 34.68 |
| rs11620355 | A | G | 0.09 | 0.018 | 0.002 | 1.09E-13 | -0.009 | 0.024 | 0.716 | 53 |
| rs11657342 | A | G | 0.37 | 0.014 | 0.002 | 2.32E-13 | -0.018 | 0.018 | 0.318 | 54.29 |
| rs11657979 | A | G | 0.24 | -0.009 | 0.002 | 4.57E-08 | 0.009 | 0.018 | 0.614 | 28.65 |
| rs11663602 | A | C | 0.28 | -0.013 | 0.002 | 9.27E-17 | -0.027 | 0.016 | 0.092 | 66.02 |
| rs1167827 | A | G | 0.43 | 0.01 | 0.001 | 2.93E-13 | 0.007 | 0.015 | 0.627 | 55.18 |
| rs11678980 | A | G | 0.45 | -0.017 | 0.001 | 1.60E-31 | -0.027 | 0.016 | 0.089 | 140.59 |
| rs11724690 | T | G | 0.29 | 0.009 | 0.002 | 2.08E-09 | -0.024 | 0.016 | 0.123 | 38.44 |
| rs118134876 | T | C | 0.06 | -0.022 | 0.003 | 1.50E-13 | -0.039 | 0.032 | 0.228 | 55.25 |
| rs11871429 | A | G | 0.77 | 0.013 | 0.002 | 2.52E-14 | -0.026 | 0.017 | 0.131 | 56.69 |
| rs11894424 | A | C | 0.95 | 0.021 | 0.003 | 2.14E-10 | -0.038 | 0.032 | 0.24 | 40.11 |
| rs12113634 | T | C | 0.65 | 0.009 | 0.002 | 2.50E-09 | -0.011 | 0.015 | 0.45 | 34.42 |
| rs12151248 | T | C | 0.11 | -0.015 | 0.002 | 3.81E-11 | 0.038 | 0.025 | 0.136 | 41.97 |
| rs1220779 | A | G | 0.47 | -0.008 | 0.001 | 1.58E-08 | -0.009 | 0.014 | 0.515 | 31.84 |
| rs12273435 | A | G | 0.21 | -0.013 | 0.002 | 4.72E-14 | -0.004 | 0.018 | 0.826 | 60.29 |
| rs12375949 | T | C | 0.43 | -0.014 | 0.001 | 1.61E-22 | 0.017 | 0.015 | 0.242 | 97.16 |
| rs12438177 | A | G | 0.37 | 0.009 | 0.002 | 1.48E-10 | 0.004 | 0.015 | 0.798 | 38.44 |
| rs12468040 | T | G | 0.38 | 0.014 | 0.001 | 3.12E-21 | -0.018 | 0.015 | 0.217 | 95.76 |
| rs12503522 | T | C | 0.28 | -0.011 | 0.002 | 3.93E-13 | -0.011 | 0.016 | 0.505 | 49.88 |
| rs12506221 | T | G | 0.44 | -0.013 | 0.001 | 4.23E-20 | 0.005 | 0.014 | 0.75 | 86.22 |
| rs12519073 | T | C | 0.23 | -0.011 | 0.002 | 1.16E-10 | 0.008 | 0.017 | 0.649 | 39.62 |
| rs12524795 | T | C | 0.44 | 0.01 | 0.001 | 5.28E-13 | 0.018 | 0.015 | 0.227 | 53.08 |
| rs12571549 | A | G | 0.14 | 0.015 | 0.002 | 1.45E-13 | -0.016 | 0.02 | 0.418 | 57 |
| rs12574281 | A | C | 0.63 | -0.008 | 0.002 | 3.04E-08 | -0.012 | 0.015 | 0.432 | 28.44 |
| rs12591647 | T | C | 0.82 | -0.015 | 0.002 | 7.78E-17 | 0.023 | 0.019 | 0.225 | 71.31 |
| rs12602286 | T | G | 0.87 | 0.016 | 0.002 | 4.62E-14 | 0.029 | 0.022 | 0.189 | 58.05 |
| rs12614263 | A | G | 0.5 | 0.008 | 0.001 | 3.38E-09 | 0.005 | 0.014 | 0.732 | 35.15 |
| rs12638072 | A | G | 0.3 | -0.011 | 0.002 | 3.13E-12 | -0.002 | 0.016 | 0.918 | 50.88 |
| rs12643771 | T | C | 0.31 | 0.013 | 0.002 | 1.32E-17 | -0.014 | 0.016 | 0.398 | 75.11 |
| rs12646523 | T | C | 0.25 | -0.013 | 0.002 | 3.79E-16 | 0.029 | 0.016 | 0.074 | 68.06 |
| rs12670376 | A | G | 0.45 | 0.009 | 0.001 | 5.99E-11 | -0.004 | 0.014 | 0.76 | 43.18 |
| rs12761761 | T | C | 0.24 | 0.015 | 0.002 | 4.79E-20 | 0.002 | 0.018 | 0.891 | 81 |
| rs12789313 | T | C | 0.49 | 0.009 | 0.001 | 6.79E-11 | 0.025 | 0.014 | 0.084 | 43.18 |
| rs12875339 | A | C | 0.64 | -0.011 | 0.002 | 1.71E-13 | 0.021 | 0.015 | 0.145 | 51.84 |
| rs12888615 | T | C | 0.2 | 0.012 | 0.002 | 9.42E-12 | 0.002 | 0.018 | 0.91 | 44.44 |
| rs12957463 | A | G | 0.8 | 0.015 | 0.002 | 9.20E-17 | 0.004 | 0.018 | 0.819 | 65.79 |
| rs12967010 | T | C | 0.77 | 0.011 | 0.002 | 1.33E-11 | -0.015 | 0.018 | 0.399 | 44.97 |
| rs12981405 | T | C | 0.17 | -0.011 | 0.002 | 2.75E-09 | 0.029 | 0.02 | 0.132 | 34.75 |
| rs1301838 | T | C | 0.32 | -0.01 | 0.002 | 1.29E-10 | 0.007 | 0.015 | 0.641 | 40.96 |
| rs13018640 | T | C | 0.6 | -0.021 | 0.001 | 1.05E-50 | 0.027 | 0.015 | 0.066 | 235.84 |
| rs13050131 | A | C | 0.66 | -0.008 | 0.002 | 4.40E-08 | 0.018 | 0.015 | 0.231 | 29.16 |
| rs13133213 | A | G | 0.5 | 0.01 | 0.001 | 7.57E-12 | 0.002 | 0.014 | 0.914 | 47.02 |
| rs13145650 | T | C | 0.92 | -0.017 | 0.002 | 8.99E-12 | 0.022 | 0.026 | 0.398 | 47.89 |
| rs13163845 | T | C | 0.85 | -0.015 | 0.002 | 4.66E-14 | 0.014 | 0.021 | 0.501 | 55.5 |
| rs13197257 | T | G | 0.28 | 0.011 | 0.002 | 2.83E-12 | -0.029 | 0.016 | 0.08 | 46.41 |
| rs13212041 | T | C | 0.8 | 0.01 | 0.002 | 7.42E-09 | -0.008 | 0.018 | 0.655 | 31.48 |
| rs13240401 | T | C | 0.78 | 0.018 | 0.002 | 2.36E-24 | -0.002 | 0.017 | 0.907 | 107.18 |
| rs1329125 | T | C | 0.33 | -0.01 | 0.002 | 8.37E-11 | 0.02 | 0.015 | 0.183 | 41.82 |
| rs13327482 | A | G | 0.82 | -0.012 | 0.002 | 4.47E-11 | 0.005 | 0.019 | 0.776 | 45.19 |
| rs1334297 | A | G | 0.74 | 0.026 | 0.002 | 2.53E-58 | 0.014 | 0.016 | 0.383 | 258 |
| rs13422673 | T | C | 0.47 | -0.011 | 0.001 | 6.48E-15 | 0.005 | 0.014 | 0.704 | 61.73 |
| rs137079 | T | C | 0.14 | 0.013 | 0.002 | 3.06E-10 | -0.008 | 0.021 | 0.708 | 40.96 |
| rs1426619 | T | C | 0.43 | 0.01 | 0.001 | 1.40E-11 | -0.022 | 0.014 | 0.131 | 47.02 |
| rs1434630 | T | G | 0.15 | -0.014 | 0.002 | 4.63E-12 | 0.003 | 0.02 | 0.885 | 46.92 |
| rs143743568 | A | G | 0.15 | 0.012 | 0.002 | 4.99E-09 | 0.007 | 0.022 | 0.752 | 34.81 |
| rs150537577 | A | G | 0.92 | -0.015 | 0.003 | 9.50E-09 | 0.007 | 0.028 | 0.798 | 33.73 |
| rs1542354 | A | G | 0.48 | -0.008 | 0.001 | 2.72E-09 | 0.007 | 0.015 | 0.629 | 36 |
| rs1544 | A | G | 0.26 | -0.01 | 0.002 | 1.76E-09 | -0.018 | 0.016 | 0.263 | 36 |
| rs1564347 | T | G | 0.34 | 0.009 | 0.002 | 2.39E-09 | 0.003 | 0.015 | 0.827 | 34.42 |
| rs1603460 | T | G | 0.42 | 0.009 | 0.001 | 2.20E-10 | 0.014 | 0.014 | 0.34 | 41.33 |
| rs162445 | A | G | 0.08 | 0.015 | 0.003 | 5.72E-09 | -0.004 | 0.026 | 0.871 | 34.18 |
| rs17048855 | A | G | 0.34 | 0.011 | 0.002 | 3.10E-13 | -0.02 | 0.015 | 0.178 | 51.84 |
| rs17110109 | T | C | 0.62 | -0.009 | 0.001 | 1.47E-10 | 0.038 | 0.015 | 0.011 | 44.13 |
| rs17144467 | A | C | 0.69 | 0.01 | 0.002 | 8.74E-11 | 0.037 | 0.015 | 0.015 | 42.68 |
| rs17148998 | A | G | 0.2 | 0.01 | 0.002 | 6.98E-09 | -0.016 | 0.018 | 0.379 | 35.3 |
| rs1717204 | A | C | 0.18 | -0.012 | 0.002 | 1.20E-10 | -0.002 | 0.02 | 0.918 | 42.25 |
| rs17248751 | A | G | 0.78 | -0.014 | 0.002 | 1.28E-15 | -0.001 | 0.018 | 0.969 | 64 |
| rs173003 | A | C | 0.49 | -0.008 | 0.001 | 5.50E-09 | 0.012 | 0.014 | 0.397 | 34.31 |
| rs17411339 | A | G | 0.56 | 0.014 | 0.001 | 5.10E-23 | 0.014 | 0.014 | 0.318 | 100 |
| rs1747714 | T | C | 0.53 | 0.011 | 0.001 | 3.82E-14 | -0.002 | 0.014 | 0.889 | 57.33 |
| rs1747817 | T | C | 0.76 | -0.009 | 0.002 | 2.29E-08 | 0.022 | 0.017 | 0.193 | 33.06 |
| rs17489649 | A | G | 0.68 | 0.013 | 0.002 | 2.77E-17 | -0.014 | 0.016 | 0.372 | 71.68 |
| rs17502934 | T | G | 0.15 | -0.017 | 0.002 | 2.12E-17 | 0.038 | 0.02 | 0.06 | 69.72 |
| rs17551064 | A | G | 0.84 | 0.014 | 0.002 | 1.53E-13 | -0.022 | 0.02 | 0.271 | 55.07 |
| rs17563464 | A | C | 0.22 | -0.015 | 0.002 | 1.74E-17 | 0.033 | 0.019 | 0.085 | 74.77 |
| rs17565975 | A | G | 0.56 | -0.011 | 0.001 | 5.54E-14 | -0.01 | 0.014 | 0.48 | 57.33 |
| rs17598675 | T | C | 0.51 | -0.012 | 0.001 | 1.37E-16 | 0.011 | 0.014 | 0.449 | 68.65 |
| rs17604349 | A | G | 0.19 | 0.013 | 0.002 | 2.36E-12 | -0.047 | 0.018 | 0.01 | 49.78 |
| rs176218 | T | G | 0.2 | 0.02 | 0.002 | 1.10E-29 | -0.005 | 0.018 | 0.766 | 123.46 |
| rs17732878 | T | C | 0.79 | -0.01 | 0.002 | 4.06E-08 | -0.002 | 0.018 | 0.888 | 31.23 |
| rs178183 | T | C | 0.25 | 0.015 | 0.002 | 4.81E-19 | -0.007 | 0.016 | 0.659 | 82.13 |
| rs17882802 | A | G | 0.43 | 0.008 | 0.001 | 3.00E-09 | -0.014 | 0.014 | 0.33 | 36 |
| rs182902112 | A | C | 0.02 | -0.03 | 0.005 | 1.17E-08 | -0.015 | 0.051 | 0.764 | 32.04 |
| rs1842713 | A | G | 0.79 | -0.012 | 0.002 | 9.67E-12 | -0.002 | 0.017 | 0.923 | 47.37 |
| rs1861786 | A | G | 0.38 | -0.008 | 0.001 | 2.39E-08 | -0.02 | 0.015 | 0.182 | 33.47 |
| rs1865955 | T | C | 0.82 | 0.012 | 0.002 | 4.06E-11 | -0.019 | 0.019 | 0.301 | 41.23 |
| rs1866823 | A | G | 0.54 | 0.011 | 0.001 | 3.09E-14 | -0.013 | 0.014 | 0.352 | 58.41 |
| rs1890132 | T | C | 0.71 | -0.009 | 0.002 | 6.56E-09 | -0.006 | 0.015 | 0.714 | 35.2 |
| rs192436652 | T | C | 0.03 | -0.032 | 0.004 | 2.06E-12 | -0.066 | 0.059 | 0.262 | 50.25 |
| rs1933264 | T | C | 0.75 | 0.01 | 0.002 | 5.85E-10 | 0.009 | 0.016 | 0.599 | 39.06 |
| rs1955250 | A | C | 0.91 | -0.014 | 0.002 | 2.26E-08 | 0.013 | 0.025 | 0.611 | 31.36 |
| rs1991585 | T | C | 0.71 | -0.01 | 0.002 | 6.26E-11 | -0.018 | 0.016 | 0.26 | 45.34 |
| rs2007655 | T | G | 0.5 | 0.009 | 0.001 | 8.64E-10 | 0.004 | 0.014 | 0.765 | 37.73 |
| rs2011603 | A | G | 0.74 | -0.009 | 0.002 | 1.12E-08 | 0.02 | 0.017 | 0.228 | 32.35 |
| rs2034631 | T | C | 0.65 | 0.009 | 0.002 | 2.64E-09 | 0.009 | 0.015 | 0.556 | 35.2 |
| rs2052285 | A | G | 0.6 | 0.011 | 0.001 | 7.25E-15 | -0.023 | 0.015 | 0.138 | 64 |
| rs2055940 | A | G | 0.32 | 0.008 | 0.002 | 4.95E-08 | -0.01 | 0.016 | 0.518 | 29.88 |
| rs2131167 | A | G | 0.45 | 0.008 | 0.001 | 1.52E-08 | -0.009 | 0.014 | 0.528 | 32.65 |
| rs2179152 | T | C | 0.37 | -0.013 | 0.002 | 1.68E-19 | 0.023 | 0.015 | 0.123 | 76.27 |
| rs2183271 | T | C | 0.64 | 0.008 | 0.002 | 1.91E-08 | 0.006 | 0.015 | 0.694 | 31.36 |
| rs2199409 | T | C | 0.8 | 0.01 | 0.002 | 3.14E-09 | 0.017 | 0.018 | 0.338 | 36.71 |
| rs2212430 | T | C | 0.3 | -0.01 | 0.002 | 1.32E-10 | -0.011 | 0.016 | 0.466 | 39.06 |
| rs2216144 | T | C | 0.53 | 0.01 | 0.001 | 2.58E-12 | 0 | 0.014 | 0.988 | 49 |
| rs2256965 | A | G | 0.42 | 0.011 | 0.001 | 2.40E-13 | 0.023 | 0.015 | 0.114 | 57.33 |
| rs2287838 | A | G | 0.55 | -0.01 | 0.001 | 1.27E-13 | 0.012 | 0.014 | 0.401 | 55.18 |
| rs2290601 | T | C | 0.77 | -0.015 | 0.002 | 4.49E-19 | 0.032 | 0.017 | 0.067 | 76.82 |
| rs232496 | T | C | 0.37 | 0.009 | 0.002 | 7.14E-10 | -0.013 | 0.015 | 0.394 | 36 |
| rs2364544 | A | G | 0.39 | -0.011 | 0.001 | 2.91E-13 | 0.025 | 0.015 | 0.09 | 56.25 |
| rs2434672 | A | C | 0.53 | 0.009 | 0.001 | 1.06E-10 | 0.002 | 0.015 | 0.869 | 42.25 |
| rs2436760 | T | C | 0.97 | 0.023 | 0.004 | 4.52E-08 | -0.056 | 0.044 | 0.211 | 30.51 |
| rs2447097 | T | G | 0.46 | 0.01 | 0.001 | 1.10E-11 | 0.013 | 0.015 | 0.367 | 47.02 |
| rs2496482 | T | C | 0.42 | 0.012 | 0.002 | 9.20E-17 | 0.004 | 0.015 | 0.764 | 65.07 |
| rs252991 | A | G | 0.37 | 0.01 | 0.002 | 6.30E-11 | -0.004 | 0.015 | 0.799 | 40.11 |
| rs2545795 | A | C | 0.44 | 0.013 | 0.001 | 1.03E-18 | -0.004 | 0.014 | 0.755 | 79.72 |
| rs2554835 | A | G | 0.4 | 0.009 | 0.001 | 8.95E-10 | -0.016 | 0.015 | 0.276 | 39.51 |
| rs2570497 | T | C | 0.64 | -0.012 | 0.002 | 1.27E-16 | -0.015 | 0.015 | 0.304 | 65.07 |
| rs2725370 | T | C | 0.3 | -0.014 | 0.002 | 4.96E-20 | 0.013 | 0.016 | 0.395 | 88.36 |
| rs2764684 | T | C | 0.83 | 0.014 | 0.002 | 9.99E-15 | -0.01 | 0.019 | 0.62 | 56.65 |
| rs2838006 | T | C | 0.36 | 0.013 | 0.002 | 1.38E-19 | 0.014 | 0.015 | 0.359 | 77.44 |
| rs28513882 | A | G | 0.18 | -0.011 | 0.002 | 2.03E-09 | 0.038 | 0.019 | 0.039 | 37.35 |
| rs2885198 | A | G | 0.53 | 0.009 | 0.001 | 4.63E-10 | 0.001 | 0.015 | 0.949 | 39.51 |
| rs2923424 | A | G | 0.61 | 0.012 | 0.001 | 6.31E-17 | 0.044 | 0.015 | 0.003 | 73.47 |
| rs2929032 | A | G | 0.47 | -0.008 | 0.001 | 8.40E-09 | 0.007 | 0.014 | 0.608 | 33.47 |
| rs2964199 | T | C | 0.31 | -0.01 | 0.002 | 4.61E-11 | -0.01 | 0.016 | 0.523 | 43.56 |
| rs2964255 | A | G | 0.31 | 0.009 | 0.002 | 6.34E-09 | 0.003 | 0.015 | 0.844 | 34.42 |
| rs2998299 | T | G | 0.21 | -0.014 | 0.002 | 1.51E-15 | -0.001 | 0.017 | 0.943 | 64 |
| rs3026996 | A | C | 0.76 | 0.014 | 0.002 | 5.42E-17 | 0.015 | 0.017 | 0.378 | 74.39 |
| rs303752 | A | G | 0.41 | -0.011 | 0.001 | 1.47E-14 | -0.004 | 0.015 | 0.792 | 61.73 |
| rs3111251 | T | C | 0.43 | -0.009 | 0.001 | 3.88E-10 | 0.006 | 0.015 | 0.675 | 41.33 |
| rs34067381 | T | G | 0.36 | -0.01 | 0.002 | 9.77E-13 | 0.009 | 0.015 | 0.523 | 48.07 |
| rs34098770 | A | G | 0.15 | -0.016 | 0.002 | 2.95E-16 | -0.001 | 0.02 | 0.962 | 64 |
| rs34305371 | A | G | 0.1 | 0.031 | 0.002 | 1.10E-39 | -0.033 | 0.026 | 0.197 | 171.17 |
| rs34394051 | A | G | 0.84 | -0.012 | 0.002 | 2.30E-09 | -0.016 | 0.02 | 0.429 | 34.22 |
| rs34720381 | T | C | 0.09 | -0.018 | 0.002 | 1.90E-13 | 0.018 | 0.026 | 0.494 | 56.25 |
| rs34807077 | A | C | 0.16 | 0.013 | 0.002 | 3.29E-11 | 0.04 | 0.02 | 0.05 | 45.39 |
| rs35417702 | T | C | 0.52 | -0.015 | 0.001 | 2.40E-27 | 0.007 | 0.014 | 0.604 | 117.88 |
| rs35606437 | A | G | 0.27 | 0.011 | 0.002 | 3.98E-11 | -0.003 | 0.017 | 0.855 | 43.07 |
| rs35929923 | A | G | 0.25 | -0.011 | 0.002 | 1.77E-12 | 0.013 | 0.016 | 0.433 | 50.77 |
| rs363096 | T | C | 0.42 | -0.014 | 0.001 | 5.77E-24 | 0.006 | 0.014 | 0.652 | 104.33 |
| rs3751331 | A | G | 0.38 | -0.01 | 0.001 | 1.75E-11 | 0.007 | 0.015 | 0.647 | 48.01 |
| rs3809634 | A | G | 0.68 | -0.011 | 0.002 | 6.82E-13 | 0.012 | 0.015 | 0.435 | 52.8 |
| rs382196 | T | G | 0.38 | -0.008 | 0.001 | 1.47E-08 | 0 | 0.015 | 0.992 | 34.31 |
| rs3859523 | T | C | 0.82 | 0.012 | 0.002 | 9.92E-12 | -0.033 | 0.019 | 0.074 | 47.46 |
| rs3897821 | A | G | 0.67 | 0.015 | 0.002 | 3.58E-25 | -0.026 | 0.015 | 0.084 | 105.4 |
| rs3948495 | T | G | 0.41 | -0.009 | 0.001 | 3.95E-10 | 0.005 | 0.015 | 0.708 | 40.41 |
| rs401526 | T | C | 0.53 | -0.01 | 0.001 | 1.15E-13 | 0.022 | 0.014 | 0.12 | 55.18 |
| rs42302 | A | G | 0.34 | 0.009 | 0.002 | 5.96E-09 | 0.009 | 0.015 | 0.532 | 32.87 |
| rs4298514 | T | C | 0.7 | 0.01 | 0.002 | 1.07E-11 | 0.012 | 0.016 | 0.437 | 48.07 |
| rs4320563 | A | G | 0.51 | 0.011 | 0.001 | 2.24E-14 | 0 | 0.014 | 0.977 | 60.62 |
| rs4328757 | T | C | 0.61 | 0.01 | 0.001 | 2.95E-11 | -0.013 | 0.015 | 0.398 | 47.02 |
| rs4352658 | T | C | 0.08 | -0.019 | 0.003 | 1.71E-13 | 0.006 | 0.027 | 0.81 | 53.4 |
| rs4358081 | A | C | 0.53 | -0.01 | 0.001 | 1.28E-11 | -0.001 | 0.014 | 0.94 | 46.05 |
| rs4369924 | A | G | 0.16 | 0.013 | 0.002 | 5.58E-11 | 0.002 | 0.02 | 0.92 | 43.28 |
| rs4384309 | A | G | 0.46 | 0.01 | 0.001 | 4.89E-13 | -0.022 | 0.015 | 0.144 | 53.08 |
| rs4467547 | T | G | 0.41 | 0.012 | 0.001 | 4.88E-17 | 0.001 | 0.015 | 0.928 | 74.7 |
| rs4500930 | T | C | 0.34 | -0.011 | 0.002 | 1.67E-13 | 0.005 | 0.015 | 0.76 | 52.8 |
| rs4673840 | T | C | 0.84 | -0.014 | 0.002 | 5.86E-13 | -0.003 | 0.019 | 0.884 | 52.75 |
| rs4675248 | A | G | 0.4 | -0.009 | 0.001 | 4.50E-11 | 0.002 | 0.015 | 0.878 | 45.08 |
| rs4719944 | T | C | 0.55 | -0.012 | 0.001 | 9.25E-18 | 0.016 | 0.014 | 0.251 | 74.7 |
| rs4726070 | A | G | 0.6 | 0.012 | 0.001 | 1.36E-17 | -0.006 | 0.015 | 0.685 | 75.94 |
| rs4778058 | T | C | 0.5 | -0.009 | 0.001 | 4.92E-11 | -0.005 | 0.015 | 0.755 | 43.18 |
| rs4787457 | A | G | 0.63 | 0.016 | 0.002 | 1.23E-28 | 0.002 | 0.015 | 0.893 | 116.64 |
| rs4793090 | A | G | 0.66 | 0.008 | 0.002 | 1.67E-08 | -0.023 | 0.015 | 0.133 | 31.36 |
| rs4810894 | A | G | 0.37 | -0.009 | 0.002 | 4.38E-09 | 0.016 | 0.015 | 0.272 | 32.11 |
| rs4839155 | T | G | 0.77 | 0.012 | 0.002 | 4.66E-12 | -0.001 | 0.017 | 0.972 | 45.76 |
| rs4846724 | A | G | 0.53 | 0.01 | 0.001 | 2.89E-12 | -0.009 | 0.014 | 0.54 | 49 |
| rs4848924 | A | C | 0.71 | -0.012 | 0.002 | 3.56E-14 | 0.013 | 0.016 | 0.409 | 60.84 |
| rs4895650 | T | C | 0.55 | 0.009 | 0.001 | 5.31E-11 | 0.022 | 0.015 | 0.139 | 44.13 |
| rs4904523 | A | G | 0.52 | -0.008 | 0.001 | 6.27E-09 | 0.006 | 0.014 | 0.685 | 34.31 |
| rs4915735 | A | G | 0.14 | 0.012 | 0.002 | 9.56E-09 | -0.042 | 0.022 | 0.054 | 33.64 |
| rs4977885 | A | G | 0.4 | -0.008 | 0.001 | 6.78E-09 | 0.027 | 0.016 | 0.088 | 36 |
| rs4984541 | A | G | 0.77 | -0.014 | 0.002 | 3.40E-16 | 0.01 | 0.018 | 0.589 | 65.9 |
| rs56099375 | T | C | 0.24 | 0.012 | 0.002 | 3.10E-13 | 0.015 | 0.018 | 0.401 | 49.83 |
| rs56319902 | T | C | 0.22 | -0.021 | 0.002 | 5.81E-33 | -0.039 | 0.018 | 0.029 | 146.84 |
| rs56391344 | A | G | 0.25 | 0.015 | 0.002 | 7.49E-20 | -0.01 | 0.017 | 0.543 | 85.56 |
| rs56794817 | A | G | 0.16 | 0.012 | 0.002 | 1.69E-10 | -0.063 | 0.02 | 0.001 | 41.23 |
| rs57349798 | A | G | 0.41 | 0.009 | 0.001 | 8.63E-11 | -0.021 | 0.015 | 0.164 | 44.13 |
| rs575113 | A | G | 0.29 | 0.013 | 0.002 | 7.52E-18 | -0.014 | 0.016 | 0.379 | 78.62 |
| rs5754753 | T | C | 0.72 | -0.013 | 0.002 | 6.15E-16 | 0.024 | 0.016 | 0.14 | 62.02 |
| rs57661533 | T | C | 0.13 | -0.011 | 0.002 | 3.24E-08 | -0.006 | 0.021 | 0.767 | 29.47 |
| rs580652 | T | C | 0.9 | -0.015 | 0.002 | 4.65E-10 | -0.024 | 0.024 | 0.308 | 37.52 |
| rs590013 | T | C | 0.68 | 0.01 | 0.002 | 2.47E-11 | -0.01 | 0.015 | 0.518 | 44.44 |
| rs6043521 | T | C | 0.6 | -0.008 | 0.001 | 9.72E-09 | -0.017 | 0.015 | 0.276 | 35.15 |
| rs6065080 | T | C | 0.36 | -0.013 | 0.002 | 7.49E-19 | -0.015 | 0.015 | 0.301 | 75.11 |
| rs61104616 | A | G | 0.52 | -0.017 | 0.001 | 1.56E-34 | 0.037 | 0.015 | 0.011 | 150.94 |
| rs6123924 | A | G | 0.84 | 0.013 | 0.002 | 3.20E-11 | -0.025 | 0.02 | 0.216 | 46.1 |
| rs61527214 | A | G | 0.41 | 0.011 | 0.001 | 1.13E-14 | 0.002 | 0.015 | 0.88 | 61.73 |
| rs61997667 | T | C | 0.15 | -0.014 | 0.002 | 2.24E-13 | -0.039 | 0.02 | 0.054 | 51.84 |
| rs62155350 | A | G | 0.02 | 0.034 | 0.005 | 1.69E-10 | 0.04 | 0.056 | 0.481 | 40.67 |
| rs62155873 | T | C | 0.12 | -0.015 | 0.002 | 1.65E-12 | -0.012 | 0.022 | 0.598 | 51.7 |
| rs62174974 | A | G | 0.19 | -0.011 | 0.002 | 2.76E-09 | 0.017 | 0.018 | 0.339 | 34.03 |
| rs62179650 | A | G | 0.3 | 0.011 | 0.002 | 5.66E-13 | 0.006 | 0.016 | 0.7 | 49 |
| rs62182994 | T | C | 0.69 | 0.014 | 0.002 | 2.55E-19 | -0.016 | 0.016 | 0.315 | 83.42 |
| rs62379838 | T | C | 0.7 | 0.012 | 0.002 | 1.77E-15 | 0.004 | 0.016 | 0.803 | 65.07 |
| rs628993 | A | G | 0.11 | 0.013 | 0.002 | 3.83E-08 | -0.009 | 0.025 | 0.709 | 30.97 |
| rs6449503 | A | G | 0.5 | 0.018 | 0.001 | 1.84E-39 | -0.028 | 0.015 | 0.051 | 174.62 |
| rs6490618 | T | C | 0.33 | 0.009 | 0.002 | 3.48E-09 | -0.01 | 0.015 | 0.5 | 34.42 |
| rs6493265 | T | C | 0.39 | -0.012 | 0.001 | 2.26E-17 | -0.002 | 0.015 | 0.91 | 75.94 |
| rs6557171 | T | C | 0.32 | -0.016 | 0.002 | 5.48E-25 | 0.011 | 0.015 | 0.476 | 106.78 |
| rs660001 | A | G | 0.21 | -0.016 | 0.002 | 1.66E-21 | 0.002 | 0.017 | 0.893 | 93.07 |
| rs66568921 | T | G | 0.64 | -0.018 | 0.002 | 7.60E-34 | 0.004 | 0.015 | 0.761 | 145.6 |
| rs66721975 | A | G | 0.29 | -0.009 | 0.002 | 1.51E-08 | 0.013 | 0.016 | 0.418 | 33.64 |
| rs6690195 | T | C | 0.49 | -0.011 | 0.001 | 7.89E-16 | -0.024 | 0.014 | 0.099 | 65.15 |
| rs6697584 | T | C | 0.79 | -0.013 | 0.002 | 1.43E-13 | 0.026 | 0.018 | 0.147 | 54.93 |
| rs6704768 | A | G | 0.56 | -0.012 | 0.001 | 1.84E-16 | 0.018 | 0.014 | 0.22 | 68.65 |
| rs6731373 | A | G | 0.34 | -0.012 | 0.002 | 1.39E-14 | 0.001 | 0.016 | 0.97 | 58.78 |
| rs6812533 | T | C | 0.74 | 0.011 | 0.002 | 4.85E-11 | 0.001 | 0.017 | 0.95 | 43.89 |
| rs68145588 | T | G | 0.14 | -0.013 | 0.002 | 7.53E-10 | -0.03 | 0.021 | 0.163 | 36.57 |
| rs6917154 | T | C | 0.12 | 0.013 | 0.002 | 2.55E-09 | 0.032 | 0.022 | 0.147 | 36.57 |
| rs6924023 | A | G | 0.75 | 0.011 | 0.002 | 5.17E-11 | 0.002 | 0.017 | 0.916 | 44.72 |
| rs6977237 | T | C | 0.47 | 0.008 | 0.001 | 5.59E-09 | 0.018 | 0.014 | 0.218 | 34.31 |
| rs6994287 | A | G | 0.41 | 0.01 | 0.001 | 1.11E-12 | 0.005 | 0.015 | 0.711 | 53.08 |
| rs7012546 | T | C | 0.42 | 0.009 | 0.001 | 1.25E-10 | 0.002 | 0.014 | 0.868 | 43.18 |
| rs7029718 | A | G | 0.41 | 0.024 | 0.001 | 7.82E-65 | -0.03 | 0.016 | 0.057 | 303.76 |
| rs7041702 | A | G | 0.74 | -0.011 | 0.002 | 4.53E-12 | -0.007 | 0.016 | 0.648 | 48.13 |
| rs7167688 | T | C | 0.49 | -0.009 | 0.001 | 1.71E-11 | 0.005 | 0.014 | 0.73 | 45.08 |
| rs7171405 | A | G | 0.24 | -0.01 | 0.002 | 7.53E-10 | 0.015 | 0.017 | 0.384 | 39.85 |
| rs717996 | T | C | 0.6 | -0.012 | 0.001 | 1.89E-17 | -0.029 | 0.015 | 0.052 | 75.94 |
| rs7226824 | T | C | 0.53 | -0.008 | 0.001 | 2.72E-08 | -0.007 | 0.015 | 0.624 | 31.04 |
| rs72486027 | T | C | 0.25 | -0.01 | 0.002 | 1.61E-10 | -0.04 | 0.017 | 0.016 | 42.25 |
| rs7254263 | T | C | 0.28 | -0.012 | 0.002 | 1.84E-14 | -0.018 | 0.016 | 0.25 | 55.32 |
| rs72622559 | T | C | 0.23 | -0.012 | 0.002 | 5.50E-13 | 0.03 | 0.017 | 0.073 | 49.83 |
| rs72709560 | A | G | 0.69 | 0.009 | 0.002 | 2.03E-08 | 0.005 | 0.017 | 0.743 | 32.11 |
| rs72828517 | T | C | 0.83 | -0.017 | 0.002 | 1.38E-19 | 0.013 | 0.019 | 0.495 | 78.18 |
| rs72829857 | A | G | 0.76 | -0.015 | 0.002 | 1.52E-19 | 0.03 | 0.016 | 0.071 | 86.72 |
| rs72917504 | T | C | 0.06 | -0.018 | 0.003 | 2.91E-09 | 0.043 | 0.03 | 0.154 | 35.2 |
| rs72944064 | T | C | 0.74 | -0.01 | 0.002 | 9.39E-10 | 0.002 | 0.017 | 0.918 | 37.52 |
| rs72962169 | T | C | 0.16 | -0.017 | 0.002 | 1.26E-19 | -0.007 | 0.02 | 0.72 | 83.87 |
| rs72993796 | T | C | 0.88 | 0.013 | 0.002 | 1.12E-08 | -0.009 | 0.025 | 0.705 | 33.32 |
| rs730384 | A | G | 0.44 | 0.01 | 0.001 | 9.38E-13 | 0.006 | 0.014 | 0.689 | 52.05 |
| rs73055556 | A | G | 0.14 | 0.011 | 0.002 | 4.95E-08 | -0.041 | 0.022 | 0.057 | 30.8 |
| rs73191311 | A | G | 0.34 | -0.009 | 0.002 | 2.03E-10 | 0.001 | 0.015 | 0.932 | 39.27 |
| rs7321274 | A | G | 0.8 | 0.011 | 0.002 | 8.21E-11 | 0.008 | 0.018 | 0.671 | 40.11 |
| rs7326331 | A | G | 0.28 | -0.013 | 0.002 | 3.42E-16 | 0.006 | 0.016 | 0.687 | 64 |
| rs73344830 | A | G | 0.42 | 0.017 | 0.001 | 1.00E-32 | -0.005 | 0.015 | 0.721 | 147.45 |
| rs73581580 | A | G | 0.12 | -0.018 | 0.002 | 5.78E-16 | -0.001 | 0.035 | 0.976 | 67.69 |
| rs736281 | T | C | 0.39 | 0.009 | 0.001 | 2.49E-09 | -0.007 | 0.015 | 0.652 | 37.73 |
| rs73648455 | T | C | 0.08 | -0.018 | 0.003 | 1.99E-12 | -0.012 | 0.028 | 0.678 | 49.54 |
| rs743316 | T | C | 0.79 | 0.011 | 0.002 | 8.21E-11 | 0.011 | 0.018 | 0.546 | 43.4 |
| rs74415461 | T | C | 0.08 | 0.018 | 0.003 | 6.25E-12 | -0.023 | 0.028 | 0.401 | 45.82 |
| rs7449561 | A | G | 0.23 | 0.01 | 0.002 | 9.73E-10 | -0.002 | 0.017 | 0.924 | 36 |
| rs7460106 | T | C | 0.76 | -0.012 | 0.002 | 2.06E-13 | 0 | 0.018 | 0.985 | 52.35 |
| rs74787922 | A | C | 0.93 | 0.018 | 0.003 | 8.97E-11 | -0.042 | 0.029 | 0.143 | 42.25 |
| rs75177132 | T | C | 0.05 | 0.03 | 0.004 | 1.01E-16 | -0.048 | 0.054 | 0.374 | 68.52 |
| rs7575637 | A | G | 0.45 | 0.011 | 0.001 | 3.43E-14 | 0.015 | 0.014 | 0.309 | 58.41 |
| rs7603132 | A | G | 0.19 | 0.014 | 0.002 | 1.20E-14 | -0.012 | 0.018 | 0.499 | 57.93 |
| rs76076331 | T | C | 0.13 | 0.021 | 0.002 | 1.71E-23 | -0.015 | 0.022 | 0.5 | 96.23 |
| rs7617204 | A | G | 0.52 | -0.01 | 0.001 | 4.33E-12 | 0 | 0.014 | 0.978 | 48.01 |
| rs76235882 | A | G | 0.97 | 0.024 | 0.004 | 4.52E-10 | -0.034 | 0.04 | 0.393 | 38.19 |
| rs76246107 | A | G | 0.09 | -0.016 | 0.003 | 6.52E-10 | -0.009 | 0.033 | 0.79 | 38.82 |
| rs7650602 | T | C | 0.56 | -0.009 | 0.001 | 2.77E-10 | 0.015 | 0.015 | 0.304 | 40.41 |
| rs7672622 | A | G | 0.75 | -0.009 | 0.002 | 4.72E-08 | -0.01 | 0.017 | 0.544 | 30.25 |
| rs7683416 | T | C | 0.46 | 0.014 | 0.001 | 6.05E-23 | -0.029 | 0.014 | 0.044 | 98.58 |
| rs7692359 | T | C | 0.78 | -0.011 | 0.002 | 1.40E-10 | -0.034 | 0.017 | 0.047 | 41.11 |
| rs77025239 | A | G | 0.16 | -0.014 | 0.002 | 4.65E-13 | 0.019 | 0.02 | 0.328 | 54.29 |
| rs77609760 | A | G | 0.93 | 0.024 | 0.003 | 7.71E-17 | 0.011 | 0.03 | 0.724 | 71.04 |
| rs7849487 | T | G | 0.66 | -0.017 | 0.002 | 1.25E-29 | -0.002 | 0.016 | 0.883 | 123.95 |
| rs78648104 | T | C | 0.91 | -0.014 | 0.002 | 1.70E-08 | 0.035 | 0.026 | 0.187 | 31.81 |
| rs7875078 | A | C | 0.46 | -0.008 | 0.001 | 2.72E-08 | 0 | 0.015 | 0.981 | 31.04 |
| rs7894722 | T | C | 0.37 | 0.009 | 0.001 | 5.41E-09 | 0.004 | 0.015 | 0.796 | 36.86 |
| rs790647 | A | C | 0.23 | -0.015 | 0.002 | 3.81E-18 | 0.018 | 0.017 | 0.297 | 72.75 |
| rs7910403 | T | G | 0.2 | 0.012 | 0.002 | 5.08E-12 | 0.013 | 0.018 | 0.467 | 45.94 |
| rs7924036 | T | G | 0.51 | 0.013 | 0.001 | 2.55E-21 | -0.007 | 0.014 | 0.624 | 90.25 |
| rs79265434 | A | G | 0.88 | -0.02 | 0.002 | 1.98E-19 | 0.003 | 0.023 | 0.889 | 80.18 |
| rs7972246 | T | C | 0.35 | 0.011 | 0.002 | 1.01E-13 | -0.001 | 0.015 | 0.939 | 53.78 |
| rs79728014 | A | G | 0.86 | -0.013 | 0.002 | 2.06E-11 | 0.004 | 0.021 | 0.858 | 44.89 |
| rs7974852 | A | C | 0.52 | 0.012 | 0.001 | 1.14E-16 | -0.002 | 0.014 | 0.877 | 69.84 |
| rs7977614 | A | G | 0.71 | -0.013 | 0.002 | 6.84E-17 | -0.008 | 0.017 | 0.634 | 70.14 |
| rs79994730 | T | C | 0.04 | 0.024 | 0.004 | 5.88E-12 | 0.006 | 0.038 | 0.879 | 47.02 |
| rs8008382 | T | C | 0.3 | -0.011 | 0.002 | 4.07E-12 | 0.045 | 0.016 | 0.005 | 49.94 |
| rs8024 | A | C | 0.33 | -0.012 | 0.002 | 1.13E-14 | -0.001 | 0.015 | 0.953 | 58.78 |
| rs80257979 | T | G | 0.97 | -0.025 | 0.004 | 3.31E-10 | 0.063 | 0.043 | 0.141 | 39.69 |
| rs8030487 | A | G | 0.69 | 0.009 | 0.002 | 2.52E-08 | 0 | 0.016 | 0.977 | 32.11 |
| rs8097125 | T | C | 0.38 | 0.009 | 0.001 | 2.31E-09 | -0.014 | 0.015 | 0.343 | 37.73 |
| rs912883 | T | C | 0.68 | 0.009 | 0.002 | 6.20E-09 | -0.01 | 0.015 | 0.51 | 33.64 |
| rs9289300 | T | C | 0.84 | -0.016 | 0.002 | 1.00E-17 | 0.009 | 0.019 | 0.632 | 74.5 |
| rs929511 | T | C | 0.13 | -0.016 | 0.002 | 3.91E-14 | 0.005 | 0.024 | 0.845 | 58.05 |
| rs9359939 | A | C | 0.24 | -0.011 | 0.002 | 1.51E-10 | 0.009 | 0.017 | 0.602 | 43.07 |
| rs936496 | A | G | 0.62 | -0.008 | 0.001 | 4.38E-08 | -0.011 | 0.015 | 0.462 | 31.84 |
| rs9373363 | A | G | 0.75 | -0.011 | 0.002 | 5.54E-12 | 0.01 | 0.017 | 0.568 | 48.13 |
| rs9375188 | T | C | 0.48 | 0.021 | 0.001 | 8.78E-52 | -0.029 | 0.014 | 0.041 | 229.31 |
| rs9411331 | A | C | 0.68 | 0.015 | 0.002 | 7.63E-22 | 0.006 | 0.015 | 0.714 | 93.44 |
| rs9529146 | T | C | 0.23 | 0.013 | 0.002 | 3.48E-14 | -0.024 | 0.017 | 0.163 | 55.81 |
| rs9556958 | T | C | 0.52 | -0.011 | 0.001 | 5.96E-15 | 0.004 | 0.014 | 0.768 | 61.73 |
| rs9616947 | T | C | 0.38 | -0.013 | 0.002 | 3.58E-20 | 0.002 | 0.015 | 0.917 | 79.8 |
| rs9633970 | T | C | 0.76 | 0.009 | 0.002 | 2.81E-08 | -0.02 | 0.017 | 0.242 | 32.35 |
| rs9655780 | A | G | 0.83 | -0.015 | 0.002 | 3.70E-16 | 0.024 | 0.019 | 0.202 | 64.84 |
| rs9853928 | T | C | 0.21 | -0.013 | 0.002 | 1.59E-13 | 0.016 | 0.017 | 0.348 | 55.81 |
| rs9859556 | T | G | 0.31 | 0.029 | 0.002 | 4.61E-82 | -0.007 | 0.015 | 0.665 | 373.78 |
| rs9882532 | T | C | 0.64 | 0.012 | 0.002 | 2.15E-17 | 0.022 | 0.015 | 0.143 | 68.34 |
| rs9927137 | A | G | 0.53 | 0.008 | 0.001 | 5.11E-09 | 0.006 | 0.015 | 0.671 | 34.31 |
| rs9927842 | T | C | 0.15 | -0.013 | 0.002 | 6.22E-11 | 0.003 | 0.021 | 0.897 | 42.25 |
| rs9929556 | T | G | 0.57 | -0.01 | 0.001 | 2.31E-12 | -0.012 | 0.015 | 0.394 | 50.01 |
| rs9929762 | A | G | 0.56 | 0.01 | 0.001 | 1.03E-11 | -0.009 | 0.014 | 0.549 | 47.02 |
| rs9933256 | A | G | 0.56 | 0.012 | 0.001 | 6.06E-17 | -0.016 | 0.015 | 0.276 | 72.25 |

**SNPs used as instrumental variables in the MR analysis of late-onset Alzheimer's disease on educational attainment**

| **SNP** | **Effect allele** | **Other allele** | **Effect allele frequency** | **beta (exposure)** | **SE (exposure)** | ***P* (exposure)** | **beta (outcome)** | **SE (outcome)** | ***P* (outcome)** | ***F*-statistic** |
| --- | --- | --- | --- | --- | --- | --- | --- | --- | --- | --- |
| rs10808026 | A | C | 0.2 | -0.101 | 0.016 | 1.30E-10 | -0.001 | 0.002 | 0.616 | 41.06 |
| rs10933431 | G | C | 0.22 | -0.094 | 0.016 | 3.42E-09 | -0.002 | 0.002 | 0.362 | 34.88 |
| rs11218343 | C | T | 0.04 | -0.22 | 0.032 | 2.88E-12 | 0.009 | 0.005 | 0.051 | 48.91 |
| rs114812713 | C | G | 0.03 | 0.278 | 0.039 | 2.10E-13 | -0.001 | 0.005 | 0.828 | 51.4 |
| rs12539172 | T | C | 0.3 | -0.082 | 0.013 | 9.26E-10 | 0.003 | 0.002 | 0.157 | 37.72 |
| rs12721046 | A | G | NA | 1.006 | 0.02 | 1.00E-200 | 0.004 | 0.002 | 0.067 | 2606.18 |
| rs12881735 | C | T | 0.22 | -0.086 | 0.015 | 7.35E-09 | 0.006 | 0.002 | 0.004 | 33.39 |
| rs138190086 | A | G | 0.02 | 0.262 | 0.045 | 5.30E-09 | -0.01 | 0.006 | 0.117 | 33.88 |
| rs17125924 | G | A | 0.09 | 0.128 | 0.021 | 1.42E-09 | 0.006 | 0.003 | 0.044 | 36.74 |
| rs2830500 | A | C | 0.31 | -0.073 | 0.014 | 2.60E-08 | -0.003 | 0.002 | 0.09 | 28.29 |
| rs35912475 | T | C | NA | -0.104 | 0.016 | 1.72E-10 | -0.001 | 0.002 | 0.687 | 40.63 |
| rs3740688 | G | T | 0.45 | -0.089 | 0.012 | 5.46E-13 | 0.002 | 0.002 | 0.263 | 52 |
| rs3752246 | G | C | 0.18 | 0.136 | 0.017 | 3.06E-16 | 0.001 | 0.002 | 0.746 | 66.61 |
| rs3851179 | T | C | 0.36 | -0.13 | 0.013 | 6.03E-25 | 0.006 | 0.002 | 9.74E-04 | 107.11 |
| rs4844610 | A | C | 0.19 | 0.157 | 0.016 | 3.60E-24 | 0.001 | 0.002 | 0.727 | 102.47 |
| rs57537848 | T | G | NA | 0.207 | 0.015 | 2.11E-42 | 0.001 | 0.002 | 0.706 | 187.2 |
| rs593742 | G | A | 0.3 | -0.073 | 0.011 | 6.80E-09 | 0.001 | 0.002 | 0.471 | 43.73 |
| rs6024870 | A | G | 0.09 | -0.123 | 0.022 | 3.46E-08 | -0.001 | 0.003 | 0.656 | 30.37 |
| rs62039712 | A | G | 0.12 | 0.148 | 0.028 | 3.70E-08 | 0.002 | 0.003 | 0.494 | 27.13 |
| rs6733839 | T | C | 0.41 | 0.181 | 0.013 | 2.05E-44 | -0.001 | 0.002 | 0.726 | 194.71 |
| rs7185636 | C | T | 0.18 | -0.083 | 0.017 | 2.40E-08 | 0.001 | 0.002 | 0.714 | 25.1 |
| rs73223431 | T | C | 0.37 | 0.096 | 0.013 | 6.30E-14 | 0.004 | 0.002 | 0.03 | 56.48 |
| rs77301115 | A | G | NA | 0.949 | 0.043 | 2.25E-109 | 0.011 | 0.006 | 0.044 | 493.53 |
| rs7920721 | G | A | 0.39 | 0.077 | 0.012 | 1.80E-11 | 0.002 | 0.002 | 0.364 | 42.84 |
| rs7933202 | C | A | 0.39 | -0.113 | 0.013 | 1.91E-19 | -0.002 | 0.002 | 0.316 | 81.72 |
| rs9331896 | C | T | 0.39 | -0.132 | 0.013 | 4.63E-24 | -0.001 | 0.002 | 0.483 | 103.1 |
| rs9473117 | C | A | 0.28 | 0.087 | 0.014 | 1.16E-10 | 0.003 | 0.002 | 0.067 | 41.82 |

**SNPs used as instrumental variables in the MR analysis of educational attainment on surface area**

| **SNP** | **Effect allele** | **Other allele** | **Effect allele frequency** | **beta (exposure)** | **SE (exposure)** | ***P* (exposure)** | **beta (outcome)** | **SE (outcome)** | ***P* (outcome)** | ***F*-statistic** |
| --- | --- | --- | --- | --- | --- | --- | --- | --- | --- | --- |
| rs10006235 | T | C | 0.27 | -0.009 | 0.002 | 4.48E-09 | -0.005 | 0.007 | 0.472 | 33.79 |
| rs10021733 | T | C | 0.15 | -0.016 | 0.003 | 9.45E-10 | 0.003 | 0.009 | 0.753 | 36.89 |
| rs10046069 | A | G | 0.12 | 0.013 | 0.002 | 4.40E-09 | -0.003 | 0.01 | 0.801 | 33.32 |
| rs10060023 | T | C | 0.33 | 0.011 | 0.002 | 1.88E-13 | 0.001 | 0.007 | 0.885 | 53.78 |
| rs10080647 | A | C | 0.14 | 0.012 | 0.002 | 7.05E-10 | -0.004 | 0.009 | 0.664 | 38.44 |
| rs1008078 | T | C | 0.4 | -0.018 | 0.001 | 4.50E-37 | -0.003 | 0.007 | 0.664 | 169 |
| rs10189857 | A | G | 0.57 | 0.016 | 0.001 | 4.69E-29 | 0.007 | 0.006 | 0.265 | 127.37 |
| rs10193498 | A | T | 0.76 | 0.01 | 0.002 | 3.10E-09 | 0.004 | 0.008 | 0.57 | 36.75 |
| rs10215082 | A | G | 0.47 | -0.015 | 0.001 | 7.03E-26 | -0.028 | 0.006 | 8.78E-06 | 113.27 |
| rs10460095 | A | G | 0.57 | -0.012 | 0.001 | 2.61E-18 | -0.007 | 0.006 | 0.249 | 78.45 |
| rs10761251 | A | T | 0.66 | 0.011 | 0.002 | 2.52E-14 | 0.008 | 0.007 | 0.211 | 56.75 |
| rs10772644 | C | G | 0.89 | 0.014 | 0.002 | 1.69E-10 | 0.012 | 0.01 | 0.264 | 41.66 |
| rs10773002 | A | T | 0.25 | 0.018 | 0.002 | 2.50E-30 | 0.031 | 0.007 | 2.18E-05 | 133.69 |
| rs10773208 | T | C | 0.25 | -0.011 | 0.002 | 8.09E-12 | 0.019 | 0.007 | 0.008 | 47.27 |
| rs10798888 | T | G | 0.17 | -0.014 | 0.002 | 5.15E-14 | -0.006 | 0.008 | 0.474 | 54.29 |
| rs10799615 | A | G | 0.25 | -0.009 | 0.002 | 4.11E-08 | -0.012 | 0.007 | 0.095 | 30.94 |
| rs10805383 | A | G | 0.49 | -0.01 | 0.001 | 2.22E-13 | -0.01 | 0.006 | 0.115 | 54.13 |
| rs10830858 | T | C | 0.51 | 0.009 | 0.001 | 5.31E-11 | 0.001 | 0.006 | 0.84 | 43.18 |
| rs10844179 | A | G | 0.23 | 0.01 | 0.002 | 5.03E-10 | -0.007 | 0.008 | 0.335 | 37.43 |
| rs10875121 | C | G | 0.84 | 0.017 | 0.002 | 4.46E-20 | 0.005 | 0.009 | 0.568 | 83.87 |
| rs10892807 | T | C | 0.57 | 0.011 | 0.001 | 1.09E-14 | -0.002 | 0.006 | 0.791 | 60.62 |
| rs10940921 | T | G | 0.42 | 0.011 | 0.001 | 1.31E-14 | -0.004 | 0.006 | 0.556 | 62.86 |
| rs10951590 | T | C | 0.33 | -0.011 | 0.002 | 3.62E-13 | 0 | 0.007 | 0.97 | 52.8 |
| rs10979613 | T | C | 0.64 | -0.012 | 0.002 | 1.25E-15 | 0 | 0.007 | 0.956 | 60.84 |
| rs10996167 | C | G | 0.65 | 0.011 | 0.002 | 7.47E-09 | 0.01 | 0.007 | 0.134 | 31.92 |
| rs11003463 | T | G | 0.61 | 0.009 | 0.002 | 5.58E-10 | 0.006 | 0.007 | 0.344 | 36.8 |
| rs11023764 | A | G | 0.64 | 0.009 | 0.002 | 5.32E-10 | 0.011 | 0.007 | 0.099 | 36.8 |
| rs11030102 | C | G | 0.75 | 0.01 | 0.002 | 4.60E-10 | 0.01 | 0.007 | 0.177 | 39.85 |
| rs11076962 | T | C | 0.72 | 0.009 | 0.002 | 4.29E-08 | 0.004 | 0.007 | 0.604 | 28.89 |
| rs11081529 | T | C | 0.71 | 0.013 | 0.002 | 5.22E-16 | 0.004 | 0.007 | 0.588 | 70.56 |
| rs11082011 | T | C | 0.67 | 0.019 | 0.002 | 2.06E-38 | 0.008 | 0.007 | 0.227 | 167.27 |
| rs11121177 | A | G | 0.18 | 0.015 | 0.002 | 1.44E-16 | -0.001 | 0.008 | 0.916 | 70.37 |
| rs11130380 | T | G | 0.61 | 0.009 | 0.001 | 4.17E-10 | 0.005 | 0.006 | 0.443 | 41.33 |
| rs11138947 | T | C | 0.72 | 0.011 | 0.002 | 2.36E-11 | -0.017 | 0.007 | 0.018 | 43.07 |
| rs11157931 | A | C | 0.39 | -0.014 | 0.001 | 3.86E-22 | -0.04 | 0.007 | 1.11E-09 | 98.58 |
| rs111852224 | T | C | 0.12 | 0.016 | 0.002 | 4.85E-13 | 0.012 | 0.01 | 0.208 | 50.28 |
| rs11211123 | A | G | 0.22 | 0.01 | 0.002 | 7.62E-10 | -0.019 | 0.008 | 0.013 | 37.43 |
| rs11213482 | A | G | 0.84 | -0.011 | 0.002 | 2.68E-08 | 0.012 | 0.008 | 0.17 | 31.12 |
| rs112603734 | A | C | 0.77 | -0.013 | 0.002 | 2.72E-09 | 0.009 | 0.007 | 0.218 | 35.46 |
| rs112806496 | C | G | 0.91 | -0.016 | 0.002 | 1.72E-10 | -0.004 | 0.011 | 0.739 | 40.96 |
| rs1128956 | T | G | 0.83 | -0.014 | 0.002 | 5.34E-14 | -0.003 | 0.008 | 0.747 | 54.29 |
| rs113520408 | A | G | 0.28 | 0.016 | 0.002 | 1.03E-22 | 0.001 | 0.007 | 0.922 | 93.85 |
| rs113615161 | T | C | 0.13 | -0.012 | 0.002 | 2.98E-09 | -0.004 | 0.009 | 0.692 | 34.31 |
| rs114593137 | A | T | 0.79 | -0.012 | 0.002 | 1.65E-12 | -0.013 | 0.008 | 0.085 | 51.5 |
| rs115000530 | A | T | 0.95 | -0.025 | 0.003 | 1.09E-15 | -0.026 | 0.014 | 0.068 | 65.04 |
| rs11542663 | A | C | 0.69 | 0.011 | 0.002 | 2.44E-13 | 0.008 | 0.007 | 0.228 | 55.75 |
| rs11588857 | A | G | 0.21 | 0.02 | 0.002 | 4.45E-31 | 0.021 | 0.008 | 0.006 | 137.03 |
| rs11598765 | A | G | 0.2 | -0.011 | 0.002 | 1.73E-09 | -0.002 | 0.008 | 0.82 | 34.68 |
| rs11620355 | A | G | 0.09 | 0.018 | 0.002 | 1.09E-13 | 0 | 0.011 | 0.973 | 53 |
| rs11657342 | A | G | 0.37 | 0.014 | 0.002 | 2.32E-13 | 0.012 | 0.007 | 0.066 | 54.29 |
| rs11657979 | A | G | 0.24 | -0.009 | 0.002 | 4.57E-08 | 0 | 0.008 | 0.982 | 28.65 |
| rs11663602 | A | C | 0.28 | -0.013 | 0.002 | 9.27E-17 | -0.007 | 0.007 | 0.326 | 66.02 |
| rs1167827 | A | G | 0.43 | 0.01 | 0.001 | 2.93E-13 | -0.006 | 0.006 | 0.344 | 55.18 |
| rs11724690 | T | G | 0.29 | 0.009 | 0.002 | 2.08E-09 | 0.001 | 0.007 | 0.94 | 38.44 |
| rs117398064 | C | G | 0.09 | -0.014 | 0.002 | 1.16E-08 | 0.012 | 0.011 | 0.288 | 31.81 |
| rs118134876 | T | C | 0.06 | -0.022 | 0.003 | 1.50E-13 | 0.001 | 0.013 | 0.944 | 55.25 |
| rs11871429 | A | G | 0.77 | 0.013 | 0.002 | 2.52E-14 | 0.001 | 0.008 | 0.85 | 56.69 |
| rs11894424 | A | C | 0.95 | 0.021 | 0.003 | 2.14E-10 | 0.018 | 0.015 | 0.242 | 40.11 |
| rs12054166 | C | G | 0.74 | 0.01 | 0.002 | 2.30E-09 | 0.009 | 0.007 | 0.24 | 36 |
| rs12076635 | C | G | 0.78 | 0.021 | 0.002 | 3.33E-34 | 0.025 | 0.008 | 0.002 | 148.27 |
| rs12113634 | T | C | 0.65 | 0.009 | 0.002 | 2.50E-09 | 0.01 | 0.007 | 0.129 | 34.42 |
| rs12151248 | T | C | 0.11 | -0.015 | 0.002 | 3.81E-11 | 0.023 | 0.01 | 0.021 | 41.97 |
| rs1220779 | A | G | 0.47 | -0.008 | 0.001 | 1.58E-08 | 0.008 | 0.006 | 0.226 | 31.84 |
| rs12273435 | A | G | 0.21 | -0.013 | 0.002 | 4.72E-14 | 0 | 0.008 | 0.993 | 60.29 |
| rs12332731 | A | T | 0.19 | 0.014 | 0.002 | 5.96E-14 | 0.006 | 0.008 | 0.469 | 56.25 |
| rs12375949 | T | C | 0.43 | -0.014 | 0.001 | 1.61E-22 | -0.005 | 0.006 | 0.47 | 97.16 |
| rs12438177 | A | G | 0.37 | 0.009 | 0.002 | 1.48E-10 | 0.019 | 0.007 | 0.004 | 38.44 |
| rs12468040 | T | G | 0.38 | 0.014 | 0.001 | 3.12E-21 | -0.002 | 0.007 | 0.777 | 95.76 |
| rs12503522 | T | C | 0.28 | -0.011 | 0.002 | 3.93E-13 | 0.003 | 0.007 | 0.63 | 49.88 |
| rs12506221 | T | G | 0.44 | -0.013 | 0.001 | 4.23E-20 | 0.004 | 0.006 | 0.545 | 86.22 |
| rs12519073 | T | C | 0.23 | -0.011 | 0.002 | 1.16E-10 | 0.006 | 0.008 | 0.443 | 39.62 |
| rs12524795 | T | C | 0.44 | 0.01 | 0.001 | 5.28E-13 | -0.007 | 0.006 | 0.301 | 53.08 |
| rs12571549 | A | G | 0.14 | 0.015 | 0.002 | 1.45E-13 | -0.009 | 0.009 | 0.34 | 57 |
| rs12574281 | A | C | 0.63 | -0.008 | 0.002 | 3.04E-08 | 0.008 | 0.007 | 0.244 | 28.44 |
| rs12591647 | T | C | 0.82 | -0.015 | 0.002 | 7.78E-17 | -0.029 | 0.008 | 4.36E-04 | 71.31 |
| rs12602286 | T | G | 0.87 | 0.016 | 0.002 | 4.62E-14 | -0.02 | 0.01 | 0.035 | 58.05 |
| rs12614263 | A | G | 0.5 | 0.008 | 0.001 | 3.38E-09 | 0.003 | 0.006 | 0.595 | 35.15 |
| rs12638072 | A | G | 0.3 | -0.011 | 0.002 | 3.13E-12 | -0.008 | 0.007 | 0.278 | 50.88 |
| rs12643771 | T | C | 0.31 | 0.013 | 0.002 | 1.32E-17 | 0.002 | 0.007 | 0.719 | 75.11 |
| rs12646523 | T | C | 0.25 | -0.013 | 0.002 | 3.79E-16 | -0.012 | 0.007 | 0.116 | 68.06 |
| rs12670376 | A | G | 0.45 | 0.009 | 0.001 | 5.99E-11 | 0.005 | 0.006 | 0.391 | 43.18 |
| rs12761761 | T | C | 0.24 | 0.015 | 0.002 | 4.79E-20 | -0.004 | 0.007 | 0.605 | 81 |
| rs12765185 | A | T | 0.28 | -0.009 | 0.002 | 2.96E-08 | -0.011 | 0.007 | 0.111 | 30.25 |
| rs12789313 | T | C | 0.49 | 0.009 | 0.001 | 6.79E-11 | 0.004 | 0.006 | 0.561 | 43.18 |
| rs12875339 | A | C | 0.64 | -0.011 | 0.002 | 1.71E-13 | 0.001 | 0.007 | 0.85 | 51.84 |
| rs12888615 | T | C | 0.2 | 0.012 | 0.002 | 9.42E-12 | -0.007 | 0.008 | 0.354 | 44.44 |
| rs12957463 | A | G | 0.8 | 0.015 | 0.002 | 9.20E-17 | 0.022 | 0.008 | 0.006 | 65.79 |
| rs12967010 | T | C | 0.77 | 0.011 | 0.002 | 1.33E-11 | 0.016 | 0.008 | 0.041 | 44.97 |
| rs12981405 | T | C | 0.17 | -0.011 | 0.002 | 2.75E-09 | -0.008 | 0.009 | 0.354 | 34.75 |
| rs1301838 | T | C | 0.32 | -0.01 | 0.002 | 1.29E-10 | 0.005 | 0.007 | 0.444 | 40.96 |
| rs13018640 | T | C | 0.6 | -0.021 | 0.001 | 1.05E-50 | 0.012 | 0.006 | 0.064 | 235.84 |
| rs13050131 | A | C | 0.66 | -0.008 | 0.002 | 4.40E-08 | 0 | 0.007 | 0.987 | 29.16 |
| rs13133213 | A | G | 0.5 | 0.01 | 0.001 | 7.57E-12 | -0.002 | 0.006 | 0.77 | 47.02 |
| rs13145650 | T | C | 0.92 | -0.017 | 0.002 | 8.99E-12 | 0.012 | 0.011 | 0.283 | 47.89 |
| rs13163845 | T | C | 0.85 | -0.015 | 0.002 | 4.66E-14 | -0.003 | 0.009 | 0.755 | 55.5 |
| rs13197257 | T | G | 0.28 | 0.011 | 0.002 | 2.83E-12 | -0.015 | 0.007 | 0.034 | 46.41 |
| rs13212041 | T | C | 0.8 | 0.01 | 0.002 | 7.42E-09 | 0.008 | 0.008 | 0.321 | 31.48 |
| rs13240401 | T | C | 0.78 | 0.018 | 0.002 | 2.36E-24 | 0.007 | 0.008 | 0.384 | 107.18 |
| rs13261773 | C | G | 0.16 | 0.012 | 0.002 | 3.12E-10 | 0.005 | 0.009 | 0.555 | 41.23 |
| rs13266287 | A | T | 0.67 | -0.009 | 0.002 | 1.75E-08 | -0.011 | 0.007 | 0.107 | 32.11 |
| rs1329125 | T | C | 0.33 | -0.01 | 0.002 | 8.37E-11 | -0.009 | 0.007 | 0.202 | 41.82 |
| rs13327482 | A | G | 0.82 | -0.012 | 0.002 | 4.47E-11 | 0.029 | 0.008 | 5.61E-04 | 45.19 |
| rs1334297 | A | G | 0.74 | 0.026 | 0.002 | 2.53E-58 | 0.001 | 0.007 | 0.85 | 258 |
| rs13422673 | T | C | 0.47 | -0.011 | 0.001 | 6.48E-15 | -0.003 | 0.006 | 0.591 | 61.73 |
| rs137079 | T | C | 0.14 | 0.013 | 0.002 | 3.06E-10 | 0.013 | 0.009 | 0.143 | 40.96 |
| rs1426619 | T | C | 0.43 | 0.01 | 0.001 | 1.40E-11 | 0.008 | 0.006 | 0.198 | 47.02 |
| rs1434630 | T | G | 0.15 | -0.014 | 0.002 | 4.63E-12 | -0.006 | 0.009 | 0.534 | 46.92 |
| rs143743568 | A | G | 0.15 | 0.012 | 0.002 | 4.99E-09 | 0.014 | 0.009 | 0.121 | 34.81 |
| rs150537577 | A | G | 0.92 | -0.015 | 0.003 | 9.50E-09 | -0.005 | 0.012 | 0.701 | 33.73 |
| rs1505676 | C | G | 0.63 | 0.008 | 0.002 | 6.67E-09 | -0.002 | 0.007 | 0.735 | 31.36 |
| rs1542354 | A | G | 0.48 | -0.008 | 0.001 | 2.72E-09 | 0.012 | 0.006 | 0.066 | 36 |
| rs1544 | A | G | 0.26 | -0.01 | 0.002 | 1.76E-09 | -0.029 | 0.007 | 4.41E-05 | 36 |
| rs1564347 | T | G | 0.34 | 0.009 | 0.002 | 2.39E-09 | -0.002 | 0.007 | 0.799 | 34.42 |
| rs1603460 | T | G | 0.42 | 0.009 | 0.001 | 2.20E-10 | 0.001 | 0.006 | 0.827 | 41.33 |
| rs162445 | A | G | 0.08 | 0.015 | 0.003 | 5.72E-09 | 0.021 | 0.012 | 0.084 | 34.18 |
| rs1689510 | C | G | 0.33 | 0.019 | 0.002 | 1.35E-38 | 0.034 | 0.007 | 3.57E-07 | 167.27 |
| rs17048855 | A | G | 0.34 | 0.011 | 0.002 | 3.10E-13 | 0.008 | 0.007 | 0.205 | 51.84 |
| rs17110109 | T | C | 0.62 | -0.009 | 0.001 | 1.47E-10 | -0.006 | 0.006 | 0.385 | 44.13 |
| rs17144467 | A | C | 0.69 | 0.01 | 0.002 | 8.74E-11 | -0.004 | 0.007 | 0.522 | 42.68 |
| rs17148998 | A | G | 0.2 | 0.01 | 0.002 | 6.98E-09 | 0.016 | 0.008 | 0.041 | 35.3 |
| rs1717204 | A | C | 0.18 | -0.012 | 0.002 | 1.20E-10 | 0.007 | 0.008 | 0.417 | 42.25 |
| rs17248751 | A | G | 0.78 | -0.014 | 0.002 | 1.28E-15 | -0.004 | 0.008 | 0.587 | 64 |
| rs173003 | A | C | 0.49 | -0.008 | 0.001 | 5.50E-09 | 0.013 | 0.006 | 0.042 | 34.31 |
| rs1738050 | C | G | 0.62 | -0.009 | 0.001 | 1.42E-10 | -0.009 | 0.007 | 0.156 | 44.13 |
| rs17411339 | A | G | 0.56 | 0.014 | 0.001 | 5.10E-23 | -0.007 | 0.006 | 0.298 | 100 |
| rs17428076 | C | G | 0.76 | 0.014 | 0.002 | 6.41E-17 | 0.006 | 0.007 | 0.381 | 73.32 |
| rs1747714 | T | C | 0.53 | 0.011 | 0.001 | 3.82E-14 | -0.005 | 0.006 | 0.401 | 57.33 |
| rs1747817 | T | C | 0.76 | -0.009 | 0.002 | 2.29E-08 | 0.001 | 0.007 | 0.915 | 33.06 |
| rs17489649 | A | G | 0.68 | 0.013 | 0.002 | 2.77E-17 | 0.002 | 0.007 | 0.82 | 71.68 |
| rs17502934 | T | G | 0.15 | -0.017 | 0.002 | 2.12E-17 | -0.005 | 0.009 | 0.608 | 69.72 |
| rs175325 | A | T | 0.6 | -0.01 | 0.001 | 2.24E-12 | 0 | 0.006 | 0.974 | 51.02 |
| rs17551064 | A | G | 0.84 | 0.014 | 0.002 | 1.53E-13 | 0.006 | 0.009 | 0.506 | 55.07 |
| rs17563464 | A | C | 0.22 | -0.015 | 0.002 | 1.74E-17 | -0.015 | 0.008 | 0.054 | 74.77 |
| rs17565975 | A | G | 0.56 | -0.011 | 0.001 | 5.54E-14 | -0.006 | 0.006 | 0.343 | 57.33 |
| rs17598675 | T | C | 0.51 | -0.012 | 0.001 | 1.37E-16 | -0.006 | 0.006 | 0.368 | 68.65 |
| rs17604349 | A | G | 0.19 | 0.013 | 0.002 | 2.36E-12 | -0.012 | 0.008 | 0.162 | 49.78 |
| rs176218 | T | G | 0.2 | 0.02 | 0.002 | 1.10E-29 | -0.01 | 0.008 | 0.203 | 123.46 |
| rs17732878 | T | C | 0.79 | -0.01 | 0.002 | 4.06E-08 | -0.007 | 0.008 | 0.342 | 31.23 |
| rs178183 | T | C | 0.25 | 0.015 | 0.002 | 4.81E-19 | 0.005 | 0.007 | 0.452 | 82.13 |
| rs17882802 | A | G | 0.43 | 0.008 | 0.001 | 3.00E-09 | 0.009 | 0.006 | 0.171 | 36 |
| rs182902112 | A | C | 0.02 | -0.03 | 0.005 | 1.17E-08 | -0.028 | 0.027 | 0.302 | 32.04 |
| rs1842713 | A | G | 0.79 | -0.012 | 0.002 | 9.67E-12 | -0.003 | 0.008 | 0.708 | 47.37 |
| rs1861786 | A | G | 0.38 | -0.008 | 0.001 | 2.39E-08 | -0.002 | 0.007 | 0.738 | 33.47 |
| rs1865955 | T | C | 0.82 | 0.012 | 0.002 | 4.06E-11 | -0.009 | 0.008 | 0.309 | 41.23 |
| rs1866823 | A | G | 0.54 | 0.011 | 0.001 | 3.09E-14 | -0.012 | 0.007 | 0.066 | 58.41 |
| rs1890132 | T | C | 0.71 | -0.009 | 0.002 | 6.56E-09 | -0.001 | 0.007 | 0.931 | 35.2 |
| rs192436652 | T | C | 0.03 | -0.032 | 0.004 | 2.06E-12 | -0.005 | 0.02 | 0.797 | 50.25 |
| rs1933264 | T | C | 0.75 | 0.01 | 0.002 | 5.85E-10 | 0.007 | 0.007 | 0.373 | 39.06 |
| rs1955250 | A | C | 0.91 | -0.014 | 0.002 | 2.26E-08 | 0.006 | 0.012 | 0.617 | 31.36 |
| rs1991585 | T | C | 0.71 | -0.01 | 0.002 | 6.26E-11 | -0.003 | 0.007 | 0.717 | 45.34 |
| rs2007655 | T | G | 0.5 | 0.009 | 0.001 | 8.64E-10 | 0.004 | 0.006 | 0.526 | 37.73 |
| rs2011603 | A | G | 0.74 | -0.009 | 0.002 | 1.12E-08 | -0.018 | 0.007 | 0.014 | 32.35 |
| rs2034631 | T | C | 0.65 | 0.009 | 0.002 | 2.64E-09 | 0 | 0.007 | 0.952 | 35.2 |
| rs2052285 | A | G | 0.6 | 0.011 | 0.001 | 7.25E-15 | 0.024 | 0.007 | 2.50E-04 | 64 |
| rs2055940 | A | G | 0.32 | 0.008 | 0.002 | 4.95E-08 | 0.012 | 0.007 | 0.071 | 29.88 |
| rs2081652 | A | T | 0.66 | 0.012 | 0.002 | 1.09E-16 | 0.024 | 0.007 | 3.56E-04 | 67.24 |
| rs2131167 | A | G | 0.45 | 0.008 | 0.001 | 1.52E-08 | 0.006 | 0.006 | 0.351 | 32.65 |
| rs2179152 | T | C | 0.37 | -0.013 | 0.002 | 1.68E-19 | 0.01 | 0.007 | 0.127 | 76.27 |
| rs2183271 | T | C | 0.64 | 0.008 | 0.002 | 1.91E-08 | 0.031 | 0.007 | 3.67E-06 | 31.36 |
| rs2199409 | T | C | 0.8 | 0.01 | 0.002 | 3.14E-09 | 0.003 | 0.008 | 0.674 | 36.71 |
| rs2212430 | T | C | 0.3 | -0.01 | 0.002 | 1.32E-10 | 0.006 | 0.007 | 0.358 | 39.06 |
| rs2216144 | T | C | 0.53 | 0.01 | 0.001 | 2.58E-12 | -0.008 | 0.006 | 0.218 | 49 |
| rs2256965 | A | G | 0.42 | 0.011 | 0.001 | 2.40E-13 | 0.022 | 0.006 | 6.07E-04 | 57.33 |
| rs2287838 | A | G | 0.55 | -0.01 | 0.001 | 1.27E-13 | 0.003 | 0.006 | 0.585 | 55.18 |
| rs2290601 | T | C | 0.77 | -0.015 | 0.002 | 4.49E-19 | 0.002 | 0.008 | 0.787 | 76.82 |
| rs2297293 | C | G | 0.31 | 0.01 | 0.002 | 8.80E-11 | 0.009 | 0.007 | 0.176 | 43.56 |
| rs232496 | T | C | 0.37 | 0.009 | 0.002 | 7.14E-10 | 0.001 | 0.007 | 0.933 | 36 |
| rs2364544 | A | G | 0.39 | -0.011 | 0.001 | 2.91E-13 | -0.009 | 0.007 | 0.19 | 56.25 |
| rs2434672 | A | C | 0.53 | 0.009 | 0.001 | 1.06E-10 | 0.007 | 0.007 | 0.269 | 42.25 |
| rs2436760 | T | C | 0.97 | 0.023 | 0.004 | 4.52E-08 | 0.007 | 0.019 | 0.711 | 30.51 |
| rs2447097 | T | G | 0.46 | 0.01 | 0.001 | 1.10E-11 | 0.01 | 0.006 | 0.121 | 47.02 |
| rs2469226 | A | T | 0.22 | 0.01 | 0.002 | 1.28E-08 | -0.011 | 0.008 | 0.139 | 31.89 |
| rs2496482 | T | C | 0.42 | 0.012 | 0.002 | 9.20E-17 | -0.029 | 0.007 | 7.46E-06 | 65.07 |
| rs252991 | A | G | 0.37 | 0.01 | 0.002 | 6.30E-11 | -0.004 | 0.007 | 0.522 | 40.11 |
| rs2545795 | A | C | 0.44 | 0.013 | 0.001 | 1.03E-18 | 0.008 | 0.006 | 0.209 | 79.72 |
| rs2554835 | A | G | 0.4 | 0.009 | 0.001 | 8.95E-10 | -0.006 | 0.007 | 0.389 | 39.51 |
| rs2570497 | T | C | 0.64 | -0.012 | 0.002 | 1.27E-16 | 0.013 | 0.007 | 0.062 | 65.07 |
| rs2725370 | T | C | 0.3 | -0.014 | 0.002 | 4.96E-20 | -0.007 | 0.007 | 0.337 | 88.36 |
| rs2764684 | T | C | 0.83 | 0.014 | 0.002 | 9.99E-15 | 0.018 | 0.008 | 0.029 | 56.65 |
| rs2838006 | T | C | 0.36 | 0.013 | 0.002 | 1.38E-19 | -0.019 | 0.007 | 0.005 | 77.44 |
| rs28513882 | A | G | 0.18 | -0.011 | 0.002 | 2.03E-09 | -0.007 | 0.008 | 0.375 | 37.35 |
| rs2885198 | A | G | 0.53 | 0.009 | 0.001 | 4.63E-10 | 0.012 | 0.006 | 0.061 | 39.51 |
| rs2923424 | A | G | 0.61 | 0.012 | 0.001 | 6.31E-17 | -0.011 | 0.007 | 0.098 | 73.47 |
| rs2929032 | A | G | 0.47 | -0.008 | 0.001 | 8.40E-09 | 0.013 | 0.006 | 0.04 | 33.47 |
| rs2958182 | A | T | 0.34 | 0.008 | 0.002 | 3.46E-08 | -0.003 | 0.007 | 0.708 | 29.88 |
| rs2964199 | T | C | 0.31 | -0.01 | 0.002 | 4.61E-11 | -0.008 | 0.007 | 0.236 | 43.56 |
| rs2964255 | A | G | 0.31 | 0.009 | 0.002 | 6.34E-09 | -0.006 | 0.007 | 0.386 | 34.42 |
| rs2998299 | T | G | 0.21 | -0.014 | 0.002 | 1.51E-15 | 0.001 | 0.008 | 0.922 | 64 |
| rs3026996 | A | C | 0.76 | 0.014 | 0.002 | 5.42E-17 | 0.003 | 0.007 | 0.643 | 74.39 |
| rs303752 | A | G | 0.41 | -0.011 | 0.001 | 1.47E-14 | -0.003 | 0.007 | 0.628 | 61.73 |
| rs3111251 | T | C | 0.43 | -0.009 | 0.001 | 3.88E-10 | -0.005 | 0.006 | 0.469 | 41.33 |
| rs34067381 | T | G | 0.36 | -0.01 | 0.002 | 9.77E-13 | -0.01 | 0.007 | 0.118 | 48.07 |
| rs34098770 | A | G | 0.15 | -0.016 | 0.002 | 2.95E-16 | -0.014 | 0.009 | 0.124 | 64 |
| rs34305371 | A | G | 0.1 | 0.031 | 0.002 | 1.10E-39 | -0.026 | 0.011 | 0.013 | 171.17 |
| rs34394051 | A | G | 0.84 | -0.012 | 0.002 | 2.30E-09 | -0.024 | 0.009 | 0.007 | 34.22 |
| rs34720381 | T | C | 0.09 | -0.018 | 0.002 | 1.90E-13 | 0.025 | 0.011 | 0.022 | 56.25 |
| rs34807077 | A | C | 0.16 | 0.013 | 0.002 | 3.29E-11 | -0.013 | 0.009 | 0.14 | 45.39 |
| rs35417702 | T | C | 0.52 | -0.015 | 0.001 | 2.40E-27 | -0.005 | 0.006 | 0.478 | 117.88 |
| rs35606437 | A | G | 0.27 | 0.011 | 0.002 | 3.98E-11 | 0.015 | 0.007 | 0.036 | 43.07 |
| rs35929923 | A | G | 0.25 | -0.011 | 0.002 | 1.77E-12 | -0.017 | 0.007 | 0.024 | 50.77 |
| rs363096 | T | C | 0.42 | -0.014 | 0.001 | 5.77E-24 | 0.004 | 0.006 | 0.559 | 104.33 |
| rs3751331 | A | G | 0.38 | -0.01 | 0.001 | 1.75E-11 | -0.006 | 0.007 | 0.397 | 48.01 |
| rs3809634 | A | G | 0.68 | -0.011 | 0.002 | 6.82E-13 | -0.006 | 0.007 | 0.367 | 52.8 |
| rs382196 | T | G | 0.38 | -0.008 | 0.001 | 1.47E-08 | 0.004 | 0.007 | 0.52 | 34.31 |
| rs3859523 | T | C | 0.82 | 0.012 | 0.002 | 9.92E-12 | 0.006 | 0.008 | 0.454 | 47.46 |
| rs3897821 | A | G | 0.67 | 0.015 | 0.002 | 3.58E-25 | 0.001 | 0.007 | 0.916 | 105.4 |
| rs3948495 | T | G | 0.41 | -0.009 | 0.001 | 3.95E-10 | 0.003 | 0.006 | 0.657 | 40.41 |
| rs401526 | T | C | 0.53 | -0.01 | 0.001 | 1.15E-13 | -0.028 | 0.006 | 1.08E-05 | 55.18 |
| rs401966 | C | G | 0.39 | -0.01 | 0.001 | 1.72E-11 | 0.001 | 0.007 | 0.876 | 48.01 |
| rs42210 | C | G | 0.71 | -0.01 | 0.002 | 2.74E-10 | 0.012 | 0.007 | 0.101 | 37.52 |
| rs42302 | A | G | 0.34 | 0.009 | 0.002 | 5.96E-09 | 0.017 | 0.007 | 0.013 | 32.87 |
| rs4298514 | T | C | 0.7 | 0.01 | 0.002 | 1.07E-11 | 0.013 | 0.007 | 0.065 | 48.07 |
| rs4320563 | A | G | 0.51 | 0.011 | 0.001 | 2.24E-14 | -0.005 | 0.006 | 0.41 | 60.62 |
| rs4328757 | T | C | 0.61 | 0.01 | 0.001 | 2.95E-11 | -0.006 | 0.007 | 0.39 | 47.02 |
| rs4352658 | T | C | 0.08 | -0.019 | 0.003 | 1.71E-13 | -0.016 | 0.012 | 0.158 | 53.4 |
| rs4358081 | A | C | 0.53 | -0.01 | 0.001 | 1.28E-11 | -0.006 | 0.006 | 0.355 | 46.05 |
| rs4369924 | A | G | 0.16 | 0.013 | 0.002 | 5.58E-11 | 0.009 | 0.009 | 0.329 | 43.28 |
| rs4384309 | A | G | 0.46 | 0.01 | 0.001 | 4.89E-13 | -0.003 | 0.006 | 0.64 | 53.08 |
| rs4467547 | T | G | 0.41 | 0.012 | 0.001 | 4.88E-17 | 0.006 | 0.006 | 0.322 | 74.7 |
| rs4500930 | T | C | 0.34 | -0.011 | 0.002 | 1.67E-13 | 0.004 | 0.007 | 0.575 | 52.8 |
| rs4663617 | A | T | 0.23 | 0.01 | 0.002 | 4.12E-10 | 0.007 | 0.008 | 0.352 | 37.43 |
| rs4673840 | T | C | 0.84 | -0.014 | 0.002 | 5.86E-13 | -0.008 | 0.009 | 0.331 | 52.75 |
| rs4675248 | A | G | 0.4 | -0.009 | 0.001 | 4.50E-11 | 0.004 | 0.007 | 0.53 | 45.08 |
| rs4719944 | T | C | 0.55 | -0.012 | 0.001 | 9.25E-18 | 0.008 | 0.006 | 0.232 | 74.7 |
| rs4726070 | A | G | 0.6 | 0.012 | 0.001 | 1.36E-17 | 0.012 | 0.007 | 0.06 | 75.94 |
| rs4757957 | C | G | 0.69 | 0.013 | 0.002 | 1.79E-18 | 0.022 | 0.007 | 0.001 | 78.62 |
| rs4766424 | C | G | 0.12 | 0.014 | 0.002 | 2.47E-11 | 0.006 | 0.01 | 0.561 | 45.72 |
| rs4778058 | T | C | 0.5 | -0.009 | 0.001 | 4.92E-11 | -0.017 | 0.007 | 0.01 | 43.18 |
| rs4787457 | A | G | 0.63 | 0.016 | 0.002 | 1.23E-28 | 0.02 | 0.007 | 0.002 | 116.64 |
| rs4793090 | A | G | 0.66 | 0.008 | 0.002 | 1.67E-08 | 0.001 | 0.007 | 0.872 | 31.36 |
| rs4810894 | A | G | 0.37 | -0.009 | 0.002 | 4.38E-09 | -0.003 | 0.007 | 0.61 | 32.11 |
| rs4839155 | T | G | 0.77 | 0.012 | 0.002 | 4.66E-12 | -0.002 | 0.008 | 0.786 | 45.76 |
| rs4846724 | A | G | 0.53 | 0.01 | 0.001 | 2.89E-12 | 0.001 | 0.006 | 0.84 | 49 |
| rs4848924 | A | C | 0.71 | -0.012 | 0.002 | 3.56E-14 | -0.007 | 0.007 | 0.306 | 60.84 |
| rs488476 | C | G | 0.62 | -0.011 | 0.002 | 8.29E-15 | -0.002 | 0.007 | 0.803 | 56.75 |
| rs4895650 | T | C | 0.55 | 0.009 | 0.001 | 5.31E-11 | -0.009 | 0.007 | 0.167 | 44.13 |
| rs4899012 | C | G | 0.6 | -0.01 | 0.001 | 8.06E-13 | -0.01 | 0.006 | 0.116 | 54.13 |
| rs4904523 | A | G | 0.52 | -0.008 | 0.001 | 6.27E-09 | -0.003 | 0.006 | 0.679 | 34.31 |
| rs4915735 | A | G | 0.14 | 0.012 | 0.002 | 9.56E-09 | -0.007 | 0.009 | 0.455 | 33.64 |
| rs4977885 | A | G | 0.4 | -0.008 | 0.001 | 6.78E-09 | -0.005 | 0.006 | 0.417 | 36 |
| rs4984541 | A | G | 0.77 | -0.014 | 0.002 | 3.40E-16 | -0.003 | 0.008 | 0.685 | 65.9 |
| rs55736314 | C | G | 0.6 | -0.015 | 0.001 | 4.59E-27 | -0.031 | 0.006 | 1.99E-06 | 121 |
| rs56099375 | T | C | 0.24 | 0.012 | 0.002 | 3.10E-13 | -0.011 | 0.008 | 0.133 | 49.83 |
| rs56319902 | T | C | 0.22 | -0.021 | 0.002 | 5.81E-33 | -0.112 | 0.008 | 1.21E-48 | 146.84 |
| rs56391344 | A | G | 0.25 | 0.015 | 0.002 | 7.49E-20 | -0.008 | 0.007 | 0.256 | 85.56 |
| rs56794817 | A | G | 0.16 | 0.012 | 0.002 | 1.69E-10 | 0.005 | 0.009 | 0.558 | 41.23 |
| rs57349798 | A | G | 0.41 | 0.009 | 0.001 | 8.63E-11 | 0.002 | 0.007 | 0.805 | 44.13 |
| rs57352738 | A | T | 0.2 | -0.015 | 0.002 | 2.51E-18 | 0.007 | 0.008 | 0.383 | 79.94 |
| rs575113 | A | G | 0.29 | 0.013 | 0.002 | 7.52E-18 | 0.012 | 0.007 | 0.101 | 78.62 |
| rs5754753 | T | C | 0.72 | -0.013 | 0.002 | 6.15E-16 | -0.005 | 0.007 | 0.479 | 62.02 |
| rs57661533 | T | C | 0.13 | -0.011 | 0.002 | 3.24E-08 | 0.006 | 0.009 | 0.527 | 29.47 |
| rs580652 | T | C | 0.9 | -0.015 | 0.002 | 4.65E-10 | 0.004 | 0.011 | 0.732 | 37.52 |
| rs590013 | T | C | 0.68 | 0.01 | 0.002 | 2.47E-11 | -0.001 | 0.007 | 0.838 | 44.44 |
| rs6043521 | T | C | 0.6 | -0.008 | 0.001 | 9.72E-09 | 0.005 | 0.007 | 0.427 | 35.15 |
| rs6065080 | T | C | 0.36 | -0.013 | 0.002 | 7.49E-19 | -0.018 | 0.007 | 0.007 | 75.11 |
| rs6065784 | C | G | 0.7 | 0.011 | 0.002 | 5.78E-13 | 0.01 | 0.007 | 0.162 | 53.78 |
| rs61104616 | A | G | 0.52 | -0.017 | 0.001 | 1.56E-34 | 0.03 | 0.006 | 3.40E-06 | 150.94 |
| rs6123924 | A | G | 0.84 | 0.013 | 0.002 | 3.20E-11 | 0.015 | 0.009 | 0.096 | 46.1 |
| rs61527214 | A | G | 0.41 | 0.011 | 0.001 | 1.13E-14 | 0.004 | 0.006 | 0.568 | 61.73 |
| rs61997667 | T | C | 0.15 | -0.014 | 0.002 | 2.24E-13 | -0.004 | 0.009 | 0.665 | 51.84 |
| rs62155350 | A | G | 0.02 | 0.034 | 0.005 | 1.69E-10 | 0.019 | 0.023 | 0.4 | 40.67 |
| rs62155873 | T | C | 0.12 | -0.015 | 0.002 | 1.65E-12 | -0.013 | 0.01 | 0.183 | 51.7 |
| rs62174974 | A | G | 0.19 | -0.011 | 0.002 | 2.76E-09 | -0.005 | 0.008 | 0.564 | 34.03 |
| rs62179650 | A | G | 0.3 | 0.011 | 0.002 | 5.66E-13 | 0.011 | 0.007 | 0.116 | 49 |
| rs62182994 | T | C | 0.69 | 0.014 | 0.002 | 2.55E-19 | -0.001 | 0.007 | 0.914 | 83.42 |
| rs62379838 | T | C | 0.7 | 0.012 | 0.002 | 1.77E-15 | 0.001 | 0.007 | 0.856 | 65.07 |
| rs628993 | A | G | 0.11 | 0.013 | 0.002 | 3.83E-08 | -0.004 | 0.011 | 0.721 | 30.97 |
| rs6449503 | A | G | 0.5 | 0.018 | 0.001 | 1.84E-39 | 0.015 | 0.006 | 0.019 | 174.62 |
| rs6490618 | T | C | 0.33 | 0.009 | 0.002 | 3.48E-09 | -0.004 | 0.007 | 0.571 | 34.42 |
| rs6493265 | T | C | 0.39 | -0.012 | 0.001 | 2.26E-17 | -0.006 | 0.007 | 0.36 | 75.94 |
| rs6557171 | T | C | 0.32 | -0.016 | 0.002 | 5.48E-25 | -0.006 | 0.007 | 0.348 | 106.78 |
| rs660001 | A | G | 0.21 | -0.016 | 0.002 | 1.66E-21 | 0.009 | 0.008 | 0.254 | 93.07 |
| rs66568921 | T | G | 0.64 | -0.018 | 0.002 | 7.60E-34 | -0.004 | 0.007 | 0.587 | 145.6 |
| rs66721975 | A | G | 0.29 | -0.009 | 0.002 | 1.51E-08 | 0.019 | 0.007 | 0.008 | 33.64 |
| rs6690195 | T | C | 0.49 | -0.011 | 0.001 | 7.89E-16 | 0.011 | 0.006 | 0.096 | 65.15 |
| rs6697584 | T | C | 0.79 | -0.013 | 0.002 | 1.43E-13 | 0.003 | 0.008 | 0.711 | 54.93 |
| rs6704768 | A | G | 0.56 | -0.012 | 0.001 | 1.84E-16 | -0.003 | 0.006 | 0.65 | 68.65 |
| rs6731373 | A | G | 0.34 | -0.012 | 0.002 | 1.39E-14 | -0.009 | 0.007 | 0.159 | 58.78 |
| rs6812533 | T | C | 0.74 | 0.011 | 0.002 | 4.85E-11 | -0.007 | 0.007 | 0.33 | 43.89 |
| rs68145588 | T | G | 0.14 | -0.013 | 0.002 | 7.53E-10 | 0.028 | 0.009 | 0.002 | 36.57 |
| rs6917154 | T | C | 0.12 | 0.013 | 0.002 | 2.55E-09 | -0.005 | 0.01 | 0.614 | 36.57 |
| rs6924023 | A | G | 0.75 | 0.011 | 0.002 | 5.17E-11 | -0.02 | 0.008 | 0.008 | 44.72 |
| rs6977237 | T | C | 0.47 | 0.008 | 0.001 | 5.59E-09 | 0.002 | 0.006 | 0.729 | 34.31 |
| rs6994287 | A | G | 0.41 | 0.01 | 0.001 | 1.11E-12 | -0.002 | 0.006 | 0.765 | 53.08 |
| rs7012546 | T | C | 0.42 | 0.009 | 0.001 | 1.25E-10 | -0.005 | 0.006 | 0.417 | 43.18 |
| rs7029718 | A | G | 0.41 | 0.024 | 0.001 | 7.82E-65 | 0.007 | 0.006 | 0.296 | 303.76 |
| rs7041702 | A | G | 0.74 | -0.011 | 0.002 | 4.53E-12 | 0.006 | 0.007 | 0.38 | 48.13 |
| rs7167688 | T | C | 0.49 | -0.009 | 0.001 | 1.71E-11 | 0.001 | 0.006 | 0.879 | 45.08 |
| rs7171405 | A | G | 0.24 | -0.01 | 0.002 | 7.53E-10 | -0.003 | 0.008 | 0.653 | 39.85 |
| rs717996 | T | C | 0.6 | -0.012 | 0.001 | 1.89E-17 | -0.002 | 0.006 | 0.794 | 75.94 |
| rs7226824 | T | C | 0.53 | -0.008 | 0.001 | 2.72E-08 | 0.006 | 0.006 | 0.327 | 31.04 |
| rs72486027 | T | C | 0.25 | -0.01 | 0.002 | 1.61E-10 | 0.006 | 0.007 | 0.432 | 42.25 |
| rs7254263 | T | C | 0.28 | -0.012 | 0.002 | 1.84E-14 | -0.013 | 0.007 | 0.074 | 55.32 |
| rs72622559 | T | C | 0.23 | -0.012 | 0.002 | 5.50E-13 | 0 | 0.007 | 0.973 | 49.83 |
| rs72672052 | A | T | 0.18 | -0.01 | 0.002 | 4.87E-08 | -0.016 | 0.008 | 0.053 | 31.48 |
| rs72709560 | A | G | 0.69 | 0.009 | 0.002 | 2.03E-08 | 0.007 | 0.007 | 0.293 | 32.11 |
| rs72828517 | T | C | 0.83 | -0.017 | 0.002 | 1.38E-19 | -0.012 | 0.008 | 0.143 | 78.18 |
| rs72829857 | A | G | 0.76 | -0.015 | 0.002 | 1.52E-19 | 0.002 | 0.008 | 0.828 | 86.72 |
| rs72917504 | T | C | 0.06 | -0.018 | 0.003 | 2.91E-09 | 0.004 | 0.014 | 0.749 | 35.2 |
| rs72944064 | T | C | 0.74 | -0.01 | 0.002 | 9.39E-10 | -0.009 | 0.007 | 0.205 | 37.52 |
| rs72962169 | T | C | 0.16 | -0.017 | 0.002 | 1.26E-19 | -0.004 | 0.009 | 0.639 | 83.87 |
| rs72993796 | T | C | 0.88 | 0.013 | 0.002 | 1.12E-08 | 0.016 | 0.01 | 0.107 | 33.32 |
| rs730384 | A | G | 0.44 | 0.01 | 0.001 | 9.38E-13 | -0.007 | 0.006 | 0.284 | 52.05 |
| rs73055556 | A | G | 0.14 | 0.011 | 0.002 | 4.95E-08 | 0.045 | 0.009 | 1.36E-06 | 30.8 |
| rs73191311 | A | G | 0.34 | -0.009 | 0.002 | 2.03E-10 | 0.004 | 0.007 | 0.561 | 39.27 |
| rs7321274 | A | G | 0.8 | 0.011 | 0.002 | 8.21E-11 | -0.006 | 0.008 | 0.416 | 40.11 |
| rs7326331 | A | G | 0.28 | -0.013 | 0.002 | 3.42E-16 | 0.001 | 0.007 | 0.883 | 64 |
| rs73344830 | A | G | 0.42 | 0.017 | 0.001 | 1.00E-32 | -0.012 | 0.006 | 0.063 | 147.45 |
| rs736281 | T | C | 0.39 | 0.009 | 0.001 | 2.49E-09 | -0.002 | 0.006 | 0.73 | 37.73 |
| rs73648455 | T | C | 0.08 | -0.018 | 0.003 | 1.99E-12 | 0.006 | 0.012 | 0.609 | 49.54 |
| rs743316 | T | C | 0.79 | 0.011 | 0.002 | 8.21E-11 | 0.007 | 0.008 | 0.396 | 43.4 |
| rs74415461 | T | C | 0.08 | 0.018 | 0.003 | 6.25E-12 | 0.007 | 0.011 | 0.548 | 45.82 |
| rs7449561 | A | G | 0.23 | 0.01 | 0.002 | 9.73E-10 | -0.001 | 0.008 | 0.854 | 36 |
| rs7460106 | T | C | 0.76 | -0.012 | 0.002 | 2.06E-13 | -0.005 | 0.008 | 0.52 | 52.35 |
| rs746839 | C | G | 0.63 | 0.014 | 0.002 | 1.19E-20 | -0.004 | 0.007 | 0.569 | 81 |
| rs74787922 | A | C | 0.93 | 0.018 | 0.003 | 8.97E-11 | 0.003 | 0.013 | 0.811 | 42.25 |
| rs75033012 | C | G | 0.04 | -0.026 | 0.004 | 1.38E-11 | 0.005 | 0.016 | 0.771 | 46.1 |
| rs75177132 | T | C | 0.05 | 0.03 | 0.004 | 1.01E-16 | 0.023 | 0.015 | 0.121 | 68.52 |
| rs7575637 | A | G | 0.45 | 0.011 | 0.001 | 3.43E-14 | 0.003 | 0.006 | 0.602 | 58.41 |
| rs7603132 | A | G | 0.19 | 0.014 | 0.002 | 1.20E-14 | 0.005 | 0.008 | 0.529 | 57.93 |
| rs76076331 | T | C | 0.13 | 0.021 | 0.002 | 1.71E-23 | -0.006 | 0.01 | 0.511 | 96.23 |
| rs7617204 | A | G | 0.52 | -0.01 | 0.001 | 4.33E-12 | -0.007 | 0.006 | 0.241 | 48.01 |
| rs76235882 | A | G | 0.97 | 0.024 | 0.004 | 4.52E-10 | -0.01 | 0.019 | 0.589 | 38.19 |
| rs76246107 | A | G | 0.09 | -0.016 | 0.003 | 6.52E-10 | -0.044 | 0.012 | 3.37E-04 | 38.82 |
| rs7650602 | T | C | 0.56 | -0.009 | 0.001 | 2.77E-10 | -0.024 | 0.006 | 1.72E-04 | 40.41 |
| rs76577427 | C | G | 0.9 | 0.02 | 0.002 | 7.78E-18 | -0.005 | 0.01 | 0.628 | 71.54 |
| rs7672622 | A | G | 0.75 | -0.009 | 0.002 | 4.72E-08 | -0.012 | 0.007 | 0.098 | 30.25 |
| rs7683416 | T | C | 0.46 | 0.014 | 0.001 | 6.05E-23 | 0.023 | 0.006 | 3.19E-04 | 98.58 |
| rs76878669 | C | G | 0.76 | 0.014 | 0.002 | 3.10E-17 | 0.009 | 0.008 | 0.225 | 68.79 |
| rs7692359 | T | C | 0.78 | -0.011 | 0.002 | 1.40E-10 | -0.003 | 0.008 | 0.673 | 41.11 |
| rs77025239 | A | G | 0.16 | -0.014 | 0.002 | 4.65E-13 | -0.009 | 0.009 | 0.332 | 54.29 |
| rs77609760 | A | G | 0.93 | 0.024 | 0.003 | 7.71E-17 | -0.002 | 0.013 | 0.888 | 71.04 |
| rs77719387 | A | T | 0.02 | -0.04 | 0.006 | 3.78E-12 | -0.031 | 0.027 | 0.256 | 48.28 |
| rs7849487 | T | G | 0.66 | -0.017 | 0.002 | 1.25E-29 | 0.009 | 0.007 | 0.179 | 123.95 |
| rs78648104 | T | C | 0.91 | -0.014 | 0.002 | 1.70E-08 | -0.025 | 0.011 | 0.029 | 31.81 |
| rs7875078 | A | C | 0.46 | -0.008 | 0.001 | 2.72E-08 | 0.004 | 0.006 | 0.548 | 31.04 |
| rs7894722 | T | C | 0.37 | 0.009 | 0.001 | 5.41E-09 | -0.007 | 0.007 | 0.293 | 36.86 |
| rs790647 | A | C | 0.23 | -0.015 | 0.002 | 3.81E-18 | -0.006 | 0.008 | 0.422 | 72.75 |
| rs7910403 | T | G | 0.2 | 0.012 | 0.002 | 5.08E-12 | -0.001 | 0.008 | 0.939 | 45.94 |
| rs7924036 | T | G | 0.51 | 0.013 | 0.001 | 2.55E-21 | 0.016 | 0.006 | 0.012 | 90.25 |
| rs79265434 | A | G | 0.88 | -0.02 | 0.002 | 1.98E-19 | -0.015 | 0.01 | 0.141 | 80.18 |
| rs7972246 | T | C | 0.35 | 0.011 | 0.002 | 1.01E-13 | 0 | 0.007 | 0.959 | 53.78 |
| rs79728014 | A | G | 0.86 | -0.013 | 0.002 | 2.06E-11 | -0.023 | 0.009 | 0.01 | 44.89 |
| rs7974852 | A | C | 0.52 | 0.012 | 0.001 | 1.14E-16 | 0.001 | 0.006 | 0.904 | 69.84 |
| rs7977614 | A | G | 0.71 | -0.013 | 0.002 | 6.84E-17 | -0.011 | 0.007 | 0.11 | 70.14 |
| rs79994730 | T | C | 0.04 | 0.024 | 0.004 | 5.88E-12 | -0.041 | 0.015 | 0.005 | 47.02 |
| rs8008382 | T | C | 0.3 | -0.011 | 0.002 | 4.07E-12 | -0.01 | 0.007 | 0.146 | 49.94 |
| rs8024 | A | C | 0.33 | -0.012 | 0.002 | 1.13E-14 | 0.009 | 0.007 | 0.182 | 58.78 |
| rs80257979 | T | G | 0.97 | -0.025 | 0.004 | 3.31E-10 | -0.042 | 0.019 | 0.025 | 39.69 |
| rs8030487 | A | G | 0.69 | 0.009 | 0.002 | 2.52E-08 | -0.004 | 0.007 | 0.518 | 32.11 |
| rs806816 | A | T | 0.75 | 0.01 | 0.002 | 4.80E-10 | 0.016 | 0.007 | 0.026 | 39.85 |
| rs8097125 | T | C | 0.38 | 0.009 | 0.001 | 2.31E-09 | 0.007 | 0.007 | 0.303 | 37.73 |
| rs912883 | T | C | 0.68 | 0.009 | 0.002 | 6.20E-09 | -0.001 | 0.007 | 0.919 | 33.64 |
| rs9289300 | T | C | 0.84 | -0.016 | 0.002 | 1.00E-17 | 0.003 | 0.009 | 0.691 | 74.5 |
| rs929511 | T | C | 0.13 | -0.016 | 0.002 | 3.91E-14 | 0.015 | 0.01 | 0.127 | 58.05 |
| rs9359939 | A | C | 0.24 | -0.011 | 0.002 | 1.51E-10 | 0.011 | 0.008 | 0.161 | 43.07 |
| rs936496 | A | G | 0.62 | -0.008 | 0.001 | 4.38E-08 | 0.001 | 0.007 | 0.881 | 31.84 |
| rs9373363 | A | G | 0.75 | -0.011 | 0.002 | 5.54E-12 | -0.006 | 0.007 | 0.406 | 48.13 |
| rs9375188 | T | C | 0.48 | 0.021 | 0.001 | 8.78E-52 | 0.016 | 0.006 | 0.015 | 229.31 |
| rs9411331 | A | C | 0.68 | 0.015 | 0.002 | 7.63E-22 | -0.012 | 0.007 | 0.07 | 93.44 |
| rs9492774 | C | G | 0.37 | 0.008 | 0.002 | 4.33E-08 | 0.006 | 0.007 | 0.367 | 28.44 |
| rs9529146 | T | C | 0.23 | 0.013 | 0.002 | 3.48E-14 | 0.015 | 0.008 | 0.043 | 55.81 |
| rs9556958 | T | C | 0.52 | -0.011 | 0.001 | 5.96E-15 | -0.008 | 0.006 | 0.225 | 61.73 |
| rs9616947 | T | C | 0.38 | -0.013 | 0.002 | 3.58E-20 | 0.001 | 0.007 | 0.938 | 79.8 |
| rs9633970 | T | C | 0.76 | 0.009 | 0.002 | 2.81E-08 | -0.009 | 0.008 | 0.267 | 32.35 |
| rs9655780 | A | G | 0.83 | -0.015 | 0.002 | 3.70E-16 | 0.017 | 0.009 | 0.056 | 64.84 |
| rs969512 | A | T | 0.66 | -0.011 | 0.002 | 9.84E-13 | 0 | 0.007 | 0.953 | 49.94 |
| rs9853928 | T | C | 0.21 | -0.013 | 0.002 | 1.59E-13 | 0.004 | 0.008 | 0.61 | 55.81 |
| rs9859556 | T | G | 0.31 | 0.029 | 0.002 | 4.61E-82 | 0.023 | 0.007 | 7.00E-04 | 373.78 |
| rs9882532 | T | C | 0.64 | 0.012 | 0.002 | 2.15E-17 | 0.006 | 0.007 | 0.342 | 68.34 |
| rs9886703 | A | T | 0.17 | -0.014 | 0.002 | 4.46E-13 | -0.004 | 0.009 | 0.604 | 51.24 |
| rs9927137 | A | G | 0.53 | 0.008 | 0.001 | 5.11E-09 | 0.015 | 0.006 | 0.016 | 34.31 |
| rs9927842 | T | C | 0.15 | -0.013 | 0.002 | 6.22E-11 | 0.001 | 0.009 | 0.943 | 42.25 |
| rs9929556 | T | G | 0.57 | -0.01 | 0.001 | 2.31E-12 | 0 | 0.006 | 0.962 | 50.01 |
| rs9929762 | A | G | 0.56 | 0.01 | 0.001 | 1.03E-11 | 0.005 | 0.006 | 0.46 | 47.02 |
| rs9933256 | A | G | 0.56 | 0.012 | 0.001 | 6.06E-17 | 0.002 | 0.006 | 0.731 | 72.25 |

**SNPs used as instrumental variables in the MR analysis of educational attainment on volume**

| **SNP** | **Effect allele** | **Other allele** | **Effect allele frequency** | **beta (exposure)** | **SE (exposure)** | ***P* (exposure)** | **beta (outcome)** | **SE (outcome)** | ***P* (outcome)** | ***F*-statistic** |
| --- | --- | --- | --- | --- | --- | --- | --- | --- | --- | --- |
| rs10006235 | T | C | 0.27 | -0.009 | 0.002 | 4.48E-09 | -0.003 | 0.007 | 0.706 | 33.79 |
| rs10021733 | T | C | 0.15 | -0.016 | 0.003 | 9.45E-10 | 0.007 | 0.008 | 0.412 | 36.89 |
| rs10046069 | A | G | 0.12 | 0.013 | 0.002 | 4.40E-09 | 0.006 | 0.01 | 0.536 | 33.32 |
| rs10060023 | T | C | 0.33 | 0.011 | 0.002 | 1.88E-13 | 0.003 | 0.007 | 0.698 | 53.78 |
| rs10080647 | A | C | 0.14 | 0.012 | 0.002 | 7.05E-10 | 0 | 0.009 | 0.956 | 38.44 |
| rs1008078 | T | C | 0.4 | -0.018 | 0.001 | 4.50E-37 | 0 | 0.006 | 0.946 | 169 |
| rs10189857 | A | G | 0.57 | 0.016 | 0.001 | 4.69E-29 | 0.007 | 0.006 | 0.298 | 127.37 |
| rs10193498 | A | T | 0.76 | 0.01 | 0.002 | 3.10E-09 | 0.002 | 0.007 | 0.746 | 36.75 |
| rs10215082 | A | G | 0.47 | -0.015 | 0.001 | 7.03E-26 | -0.037 | 0.006 | 2.68E-09 | 113.27 |
| rs10460095 | A | G | 0.57 | -0.012 | 0.001 | 2.61E-18 | -0.006 | 0.006 | 0.352 | 78.45 |
| rs10761251 | A | T | 0.66 | 0.011 | 0.002 | 2.52E-14 | 0.002 | 0.007 | 0.759 | 56.75 |
| rs10772644 | C | G | 0.89 | 0.014 | 0.002 | 1.69E-10 | 0.025 | 0.01 | 0.015 | 41.66 |
| rs10773002 | A | T | 0.25 | 0.018 | 0.002 | 2.50E-30 | 0.034 | 0.007 | 2.95E-06 | 133.69 |
| rs10773208 | T | C | 0.25 | -0.011 | 0.002 | 8.09E-12 | 0.017 | 0.007 | 0.017 | 47.27 |
| rs10798888 | T | G | 0.17 | -0.014 | 0.002 | 5.15E-14 | -0.012 | 0.008 | 0.132 | 54.29 |
| rs10799615 | A | G | 0.25 | -0.009 | 0.002 | 4.11E-08 | -0.009 | 0.007 | 0.218 | 30.94 |
| rs10805383 | A | G | 0.49 | -0.01 | 0.001 | 2.22E-13 | -0.005 | 0.006 | 0.39 | 54.13 |
| rs10830858 | T | C | 0.51 | 0.009 | 0.001 | 5.31E-11 | 0.005 | 0.006 | 0.411 | 43.18 |
| rs10844179 | A | G | 0.23 | 0.01 | 0.002 | 5.03E-10 | 0 | 0.007 | 0.977 | 37.43 |
| rs10875121 | C | G | 0.84 | 0.017 | 0.002 | 4.46E-20 | 0.007 | 0.008 | 0.433 | 83.87 |
| rs10892807 | T | C | 0.57 | 0.011 | 0.001 | 1.09E-14 | 0.006 | 0.006 | 0.364 | 60.62 |
| rs10940921 | T | G | 0.42 | 0.011 | 0.001 | 1.31E-14 | -0.005 | 0.006 | 0.395 | 62.86 |
| rs10951590 | T | C | 0.33 | -0.011 | 0.002 | 3.62E-13 | -0.007 | 0.007 | 0.26 | 52.8 |
| rs10979613 | T | C | 0.64 | -0.012 | 0.002 | 1.25E-15 | 0.005 | 0.006 | 0.415 | 60.84 |
| rs10996167 | C | G | 0.65 | 0.011 | 0.002 | 7.47E-09 | 0.008 | 0.007 | 0.21 | 31.92 |
| rs11003463 | T | G | 0.61 | 0.009 | 0.002 | 5.58E-10 | 0.01 | 0.006 | 0.111 | 36.8 |
| rs11023764 | A | G | 0.64 | 0.009 | 0.002 | 5.32E-10 | 0.004 | 0.007 | 0.519 | 36.8 |
| rs11030102 | C | G | 0.75 | 0.01 | 0.002 | 4.60E-10 | -0.001 | 0.007 | 0.909 | 39.85 |
| rs11076962 | T | C | 0.72 | 0.009 | 0.002 | 4.29E-08 | 0 | 0.007 | 0.979 | 28.89 |
| rs11081529 | T | C | 0.71 | 0.013 | 0.002 | 5.22E-16 | 0.002 | 0.007 | 0.754 | 70.56 |
| rs11082011 | T | C | 0.67 | 0.019 | 0.002 | 2.06E-38 | 0.011 | 0.007 | 0.102 | 167.27 |
| rs11121177 | A | G | 0.18 | 0.015 | 0.002 | 1.44E-16 | 0 | 0.008 | 0.991 | 70.37 |
| rs11130380 | T | G | 0.61 | 0.009 | 0.001 | 4.17E-10 | 0.004 | 0.006 | 0.505 | 41.33 |
| rs11138947 | T | C | 0.72 | 0.011 | 0.002 | 2.36E-11 | -0.011 | 0.007 | 0.103 | 43.07 |
| rs11157931 | A | C | 0.39 | -0.014 | 0.001 | 3.86E-22 | -0.027 | 0.006 | 2.89E-05 | 98.58 |
| rs111852224 | T | C | 0.12 | 0.016 | 0.002 | 4.85E-13 | 0.014 | 0.009 | 0.15 | 50.28 |
| rs11211123 | A | G | 0.22 | 0.01 | 0.002 | 7.62E-10 | -0.009 | 0.007 | 0.216 | 37.43 |
| rs11213482 | A | G | 0.84 | -0.011 | 0.002 | 2.68E-08 | 0.007 | 0.008 | 0.423 | 31.12 |
| rs112603734 | A | C | 0.77 | -0.013 | 0.002 | 2.72E-09 | 0.005 | 0.007 | 0.474 | 35.46 |
| rs112806496 | C | G | 0.91 | -0.016 | 0.002 | 1.72E-10 | -0.002 | 0.011 | 0.82 | 40.96 |
| rs1128956 | T | G | 0.83 | -0.014 | 0.002 | 5.34E-14 | -0.009 | 0.008 | 0.27 | 54.29 |
| rs113520408 | A | G | 0.28 | 0.016 | 0.002 | 1.03E-22 | 0.001 | 0.007 | 0.855 | 93.85 |
| rs113615161 | T | C | 0.13 | -0.012 | 0.002 | 2.98E-09 | -0.01 | 0.009 | 0.279 | 34.31 |
| rs114593137 | A | T | 0.79 | -0.012 | 0.002 | 1.65E-12 | -0.014 | 0.008 | 0.067 | 51.5 |
| rs115000530 | A | T | 0.95 | -0.025 | 0.003 | 1.09E-15 | -0.009 | 0.014 | 0.493 | 65.04 |
| rs11542663 | A | C | 0.69 | 0.011 | 0.002 | 2.44E-13 | 0.01 | 0.007 | 0.149 | 55.75 |
| rs11588857 | A | G | 0.21 | 0.02 | 0.002 | 4.45E-31 | 0.03 | 0.008 | 8.83E-05 | 137.03 |
| rs11598765 | A | G | 0.2 | -0.011 | 0.002 | 1.73E-09 | -0.006 | 0.008 | 0.473 | 34.68 |
| rs11620355 | A | G | 0.09 | 0.018 | 0.002 | 1.09E-13 | -0.008 | 0.011 | 0.491 | 53 |
| rs11657342 | A | G | 0.37 | 0.014 | 0.002 | 2.32E-13 | 0.007 | 0.006 | 0.253 | 54.29 |
| rs11657979 | A | G | 0.24 | -0.009 | 0.002 | 4.57E-08 | -0.01 | 0.008 | 0.191 | 28.65 |
| rs11663602 | A | C | 0.28 | -0.013 | 0.002 | 9.27E-17 | -0.007 | 0.007 | 0.299 | 66.02 |
| rs1167827 | A | G | 0.43 | 0.01 | 0.001 | 2.93E-13 | 0.011 | 0.006 | 0.09 | 55.18 |
| rs11724690 | T | G | 0.29 | 0.009 | 0.002 | 2.08E-09 | 0.004 | 0.007 | 0.527 | 38.44 |
| rs117398064 | C | G | 0.09 | -0.014 | 0.002 | 1.16E-08 | 0.011 | 0.011 | 0.315 | 31.81 |
| rs118134876 | T | C | 0.06 | -0.022 | 0.003 | 1.50E-13 | 0.014 | 0.013 | 0.282 | 55.25 |
| rs11871429 | A | G | 0.77 | 0.013 | 0.002 | 2.52E-14 | 0.003 | 0.007 | 0.728 | 56.69 |
| rs11894424 | A | C | 0.95 | 0.021 | 0.003 | 2.14E-10 | 0.014 | 0.015 | 0.363 | 40.11 |
| rs12054166 | C | G | 0.74 | 0.01 | 0.002 | 2.30E-09 | 0.01 | 0.007 | 0.18 | 36 |
| rs12076635 | C | G | 0.78 | 0.021 | 0.002 | 3.33E-34 | 0.027 | 0.008 | 2.74E-04 | 148.27 |
| rs12113634 | T | C | 0.65 | 0.009 | 0.002 | 2.50E-09 | 0.006 | 0.007 | 0.331 | 34.42 |
| rs12151248 | T | C | 0.11 | -0.015 | 0.002 | 3.81E-11 | 0.022 | 0.01 | 0.021 | 41.97 |
| rs1220779 | A | G | 0.47 | -0.008 | 0.001 | 1.58E-08 | 0.01 | 0.006 | 0.122 | 31.84 |
| rs12273435 | A | G | 0.21 | -0.013 | 0.002 | 4.72E-14 | 0.002 | 0.008 | 0.808 | 60.29 |
| rs12332731 | A | T | 0.19 | 0.014 | 0.002 | 5.96E-14 | 0 | 0.008 | 0.971 | 56.25 |
| rs12375949 | T | C | 0.43 | -0.014 | 0.001 | 1.61E-22 | 0.003 | 0.006 | 0.615 | 97.16 |
| rs12438177 | A | G | 0.37 | 0.009 | 0.002 | 1.48E-10 | 0.014 | 0.006 | 0.028 | 38.44 |
| rs12468040 | T | G | 0.38 | 0.014 | 0.001 | 3.12E-21 | -0.006 | 0.006 | 0.375 | 95.76 |
| rs12503522 | T | C | 0.28 | -0.011 | 0.002 | 3.93E-13 | -0.002 | 0.007 | 0.72 | 49.88 |
| rs12506221 | T | G | 0.44 | -0.013 | 0.001 | 4.23E-20 | 0.004 | 0.006 | 0.483 | 86.22 |
| rs12519073 | T | C | 0.23 | -0.011 | 0.002 | 1.16E-10 | 0.003 | 0.007 | 0.712 | 39.62 |
| rs12524795 | T | C | 0.44 | 0.01 | 0.001 | 5.28E-13 | 0 | 0.006 | 0.964 | 53.08 |
| rs12571549 | A | G | 0.14 | 0.015 | 0.002 | 1.45E-13 | -0.002 | 0.009 | 0.857 | 57 |
| rs12574281 | A | C | 0.63 | -0.008 | 0.002 | 3.04E-08 | 0.01 | 0.006 | 0.117 | 28.44 |
| rs12591647 | T | C | 0.82 | -0.015 | 0.002 | 7.78E-17 | -0.024 | 0.008 | 0.003 | 71.31 |
| rs12602286 | T | G | 0.87 | 0.016 | 0.002 | 4.62E-14 | -0.008 | 0.009 | 0.412 | 58.05 |
| rs12614263 | A | G | 0.5 | 0.008 | 0.001 | 3.38E-09 | 0.005 | 0.006 | 0.412 | 35.15 |
| rs12638072 | A | G | 0.3 | -0.011 | 0.002 | 3.13E-12 | -0.012 | 0.007 | 0.092 | 50.88 |
| rs12643771 | T | C | 0.31 | 0.013 | 0.002 | 1.32E-17 | 0.002 | 0.007 | 0.744 | 75.11 |
| rs12646523 | T | C | 0.25 | -0.013 | 0.002 | 3.79E-16 | -0.019 | 0.007 | 0.007 | 68.06 |
| rs12670376 | A | G | 0.45 | 0.009 | 0.001 | 5.99E-11 | 0.009 | 0.006 | 0.133 | 43.18 |
| rs12761761 | T | C | 0.24 | 0.015 | 0.002 | 4.79E-20 | -0.007 | 0.007 | 0.324 | 81 |
| rs12765185 | A | T | 0.28 | -0.009 | 0.002 | 2.96E-08 | -0.006 | 0.007 | 0.427 | 30.25 |
| rs12789313 | T | C | 0.49 | 0.009 | 0.001 | 6.79E-11 | 0 | 0.006 | 0.956 | 43.18 |
| rs12875339 | A | C | 0.64 | -0.011 | 0.002 | 1.71E-13 | -0.003 | 0.006 | 0.666 | 51.84 |
| rs12888615 | T | C | 0.2 | 0.012 | 0.002 | 9.42E-12 | -0.008 | 0.008 | 0.334 | 44.44 |
| rs12957463 | A | G | 0.8 | 0.015 | 0.002 | 9.20E-17 | 0.012 | 0.008 | 0.106 | 65.79 |
| rs12967010 | T | C | 0.77 | 0.011 | 0.002 | 1.33E-11 | 0.015 | 0.008 | 0.048 | 44.97 |
| rs12981405 | T | C | 0.17 | -0.011 | 0.002 | 2.75E-09 | -0.019 | 0.008 | 0.023 | 34.75 |
| rs1301838 | T | C | 0.32 | -0.01 | 0.002 | 1.29E-10 | 0.009 | 0.007 | 0.161 | 40.96 |
| rs13018640 | T | C | 0.6 | -0.021 | 0.001 | 1.05E-50 | 0.006 | 0.006 | 0.383 | 235.84 |
| rs13050131 | A | C | 0.66 | -0.008 | 0.002 | 4.40E-08 | 0.008 | 0.007 | 0.241 | 29.16 |
| rs13133213 | A | G | 0.5 | 0.01 | 0.001 | 7.57E-12 | -0.006 | 0.006 | 0.305 | 47.02 |
| rs13145650 | T | C | 0.92 | -0.017 | 0.002 | 8.99E-12 | -0.002 | 0.011 | 0.891 | 47.89 |
| rs13163845 | T | C | 0.85 | -0.015 | 0.002 | 4.66E-14 | -0.011 | 0.009 | 0.222 | 55.5 |
| rs13197257 | T | G | 0.28 | 0.011 | 0.002 | 2.83E-12 | -0.015 | 0.007 | 0.03 | 46.41 |
| rs13212041 | T | C | 0.8 | 0.01 | 0.002 | 7.42E-09 | 0.003 | 0.008 | 0.707 | 31.48 |
| rs13240401 | T | C | 0.78 | 0.018 | 0.002 | 2.36E-24 | 0.01 | 0.007 | 0.163 | 107.18 |
| rs13261773 | C | G | 0.16 | 0.012 | 0.002 | 3.12E-10 | -0.004 | 0.009 | 0.674 | 41.23 |
| rs13266287 | A | T | 0.67 | -0.009 | 0.002 | 1.75E-08 | -0.009 | 0.007 | 0.196 | 32.11 |
| rs1329125 | T | C | 0.33 | -0.01 | 0.002 | 8.37E-11 | -0.011 | 0.007 | 0.095 | 41.82 |
| rs13327482 | A | G | 0.82 | -0.012 | 0.002 | 4.47E-11 | 0.026 | 0.008 | 0.001 | 45.19 |
| rs1334297 | A | G | 0.74 | 0.026 | 0.002 | 2.53E-58 | -0.002 | 0.007 | 0.736 | 258 |
| rs13422673 | T | C | 0.47 | -0.011 | 0.001 | 6.48E-15 | 0.005 | 0.006 | 0.429 | 61.73 |
| rs137079 | T | C | 0.14 | 0.013 | 0.002 | 3.06E-10 | 0.005 | 0.009 | 0.536 | 40.96 |
| rs1426619 | T | C | 0.43 | 0.01 | 0.001 | 1.40E-11 | 0.009 | 0.006 | 0.133 | 47.02 |
| rs1434630 | T | G | 0.15 | -0.014 | 0.002 | 4.63E-12 | -0.004 | 0.009 | 0.623 | 46.92 |
| rs143743568 | A | G | 0.15 | 0.012 | 0.002 | 4.99E-09 | 0.007 | 0.009 | 0.401 | 34.81 |
| rs150537577 | A | G | 0.92 | -0.015 | 0.003 | 9.50E-09 | -0.005 | 0.012 | 0.648 | 33.73 |
| rs1505676 | C | G | 0.63 | 0.008 | 0.002 | 6.67E-09 | -0.003 | 0.006 | 0.644 | 31.36 |
| rs1542354 | A | G | 0.48 | -0.008 | 0.001 | 2.72E-09 | 0.015 | 0.006 | 0.019 | 36 |
| rs1544 | A | G | 0.26 | -0.01 | 0.002 | 1.76E-09 | -0.031 | 0.007 | 1.23E-05 | 36 |
| rs1564347 | T | G | 0.34 | 0.009 | 0.002 | 2.39E-09 | 0.002 | 0.007 | 0.733 | 34.42 |
| rs1603460 | T | G | 0.42 | 0.009 | 0.001 | 2.20E-10 | 0.006 | 0.006 | 0.321 | 41.33 |
| rs162445 | A | G | 0.08 | 0.015 | 0.003 | 5.72E-09 | 0.011 | 0.012 | 0.338 | 34.18 |
| rs1689510 | C | G | 0.33 | 0.019 | 0.002 | 1.35E-38 | 0.026 | 0.007 | 5.18E-05 | 167.27 |
| rs17048855 | A | G | 0.34 | 0.011 | 0.002 | 3.10E-13 | 0.004 | 0.006 | 0.562 | 51.84 |
| rs17110109 | T | C | 0.62 | -0.009 | 0.001 | 1.47E-10 | -0.01 | 0.006 | 0.099 | 44.13 |
| rs17144467 | A | C | 0.69 | 0.01 | 0.002 | 8.74E-11 | 0.001 | 0.007 | 0.893 | 42.68 |
| rs17148998 | A | G | 0.2 | 0.01 | 0.002 | 6.98E-09 | 0.009 | 0.008 | 0.236 | 35.3 |
| rs1717204 | A | C | 0.18 | -0.012 | 0.002 | 1.20E-10 | 0.007 | 0.008 | 0.413 | 42.25 |
| rs17248751 | A | G | 0.78 | -0.014 | 0.002 | 1.28E-15 | -0.004 | 0.007 | 0.635 | 64 |
| rs173003 | A | C | 0.49 | -0.008 | 0.001 | 5.50E-09 | -0.006 | 0.006 | 0.333 | 34.31 |
| rs1738050 | C | G | 0.62 | -0.009 | 0.001 | 1.42E-10 | -0.01 | 0.006 | 0.11 | 44.13 |
| rs17411339 | A | G | 0.56 | 0.014 | 0.001 | 5.10E-23 | -0.008 | 0.006 | 0.226 | 100 |
| rs17428076 | C | G | 0.76 | 0.014 | 0.002 | 6.41E-17 | 0.003 | 0.007 | 0.698 | 73.32 |
| rs1747714 | T | C | 0.53 | 0.011 | 0.001 | 3.82E-14 | -0.007 | 0.006 | 0.294 | 57.33 |
| rs1747817 | T | C | 0.76 | -0.009 | 0.002 | 2.29E-08 | 0 | 0.007 | 0.963 | 33.06 |
| rs17489649 | A | G | 0.68 | 0.013 | 0.002 | 2.77E-17 | 0.002 | 0.007 | 0.738 | 71.68 |
| rs17502934 | T | G | 0.15 | -0.017 | 0.002 | 2.12E-17 | -0.005 | 0.009 | 0.608 | 69.72 |
| rs175325 | A | T | 0.6 | -0.01 | 0.001 | 2.24E-12 | -0.004 | 0.006 | 0.518 | 51.02 |
| rs17551064 | A | G | 0.84 | 0.014 | 0.002 | 1.53E-13 | 0.006 | 0.008 | 0.498 | 55.07 |
| rs17563464 | A | C | 0.22 | -0.015 | 0.002 | 1.74E-17 | -0.012 | 0.008 | 0.11 | 74.77 |
| rs17565975 | A | G | 0.56 | -0.011 | 0.001 | 5.54E-14 | 0.001 | 0.006 | 0.833 | 57.33 |
| rs17598675 | T | C | 0.51 | -0.012 | 0.001 | 1.37E-16 | -0.006 | 0.006 | 0.343 | 68.65 |
| rs17604349 | A | G | 0.19 | 0.013 | 0.002 | 2.36E-12 | -0.002 | 0.008 | 0.845 | 49.78 |
| rs176218 | T | G | 0.2 | 0.02 | 0.002 | 1.10E-29 | 0 | 0.008 | 0.979 | 123.46 |
| rs17732878 | T | C | 0.79 | -0.01 | 0.002 | 4.06E-08 | -0.013 | 0.008 | 0.097 | 31.23 |
| rs178183 | T | C | 0.25 | 0.015 | 0.002 | 4.81E-19 | 0.003 | 0.007 | 0.642 | 82.13 |
| rs17882802 | A | G | 0.43 | 0.008 | 0.001 | 3.00E-09 | 0.014 | 0.006 | 0.029 | 36 |
| rs182902112 | A | C | 0.02 | -0.03 | 0.005 | 1.17E-08 | -0.055 | 0.026 | 0.038 | 32.04 |
| rs1842713 | A | G | 0.79 | -0.012 | 0.002 | 9.67E-12 | -0.002 | 0.008 | 0.79 | 47.37 |
| rs1861786 | A | G | 0.38 | -0.008 | 0.001 | 2.39E-08 | 0.002 | 0.006 | 0.744 | 33.47 |
| rs1865955 | T | C | 0.82 | 0.012 | 0.002 | 4.06E-11 | 0 | 0.008 | 0.957 | 41.23 |
| rs1866823 | A | G | 0.54 | 0.011 | 0.001 | 3.09E-14 | -0.007 | 0.006 | 0.259 | 58.41 |
| rs1890132 | T | C | 0.71 | -0.009 | 0.002 | 6.56E-09 | 0.001 | 0.007 | 0.934 | 35.2 |
| rs192436652 | T | C | 0.03 | -0.032 | 0.004 | 2.06E-12 | -0.006 | 0.019 | 0.774 | 50.25 |
| rs1933264 | T | C | 0.75 | 0.01 | 0.002 | 5.85E-10 | 0.017 | 0.007 | 0.019 | 39.06 |
| rs1955250 | A | C | 0.91 | -0.014 | 0.002 | 2.26E-08 | 0.002 | 0.011 | 0.84 | 31.36 |
| rs1991585 | T | C | 0.71 | -0.01 | 0.002 | 6.26E-11 | -0.001 | 0.007 | 0.896 | 45.34 |
| rs2007655 | T | G | 0.5 | 0.009 | 0.001 | 8.64E-10 | -0.001 | 0.006 | 0.932 | 37.73 |
| rs2011603 | A | G | 0.74 | -0.009 | 0.002 | 1.12E-08 | -0.012 | 0.007 | 0.089 | 32.35 |
| rs2034631 | T | C | 0.65 | 0.009 | 0.002 | 2.64E-09 | -0.001 | 0.007 | 0.896 | 35.2 |
| rs2052285 | A | G | 0.6 | 0.011 | 0.001 | 7.25E-15 | 0.018 | 0.006 | 0.004 | 64 |
| rs2055940 | A | G | 0.32 | 0.008 | 0.002 | 4.95E-08 | 0.007 | 0.007 | 0.274 | 29.88 |
| rs2081652 | A | T | 0.66 | 0.012 | 0.002 | 1.09E-16 | 0.01 | 0.007 | 0.125 | 67.24 |
| rs2131167 | A | G | 0.45 | 0.008 | 0.001 | 1.52E-08 | 0.013 | 0.006 | 0.043 | 32.65 |
| rs2179152 | T | C | 0.37 | -0.013 | 0.002 | 1.68E-19 | 0.01 | 0.006 | 0.112 | 76.27 |
| rs2183271 | T | C | 0.64 | 0.008 | 0.002 | 1.91E-08 | 0.025 | 0.006 | 1.18E-04 | 31.36 |
| rs2199409 | T | C | 0.8 | 0.01 | 0.002 | 3.14E-09 | 0.002 | 0.008 | 0.808 | 36.71 |
| rs2212430 | T | C | 0.3 | -0.01 | 0.002 | 1.32E-10 | 0.002 | 0.007 | 0.748 | 39.06 |
| rs2216144 | T | C | 0.53 | 0.01 | 0.001 | 2.58E-12 | -0.002 | 0.006 | 0.691 | 49 |
| rs2256965 | A | G | 0.42 | 0.011 | 0.001 | 2.40E-13 | 0.016 | 0.006 | 0.01 | 57.33 |
| rs2287838 | A | G | 0.55 | -0.01 | 0.001 | 1.27E-13 | 0.003 | 0.006 | 0.676 | 55.18 |
| rs2290601 | T | C | 0.77 | -0.015 | 0.002 | 4.49E-19 | 0.007 | 0.007 | 0.329 | 76.82 |
| rs2297293 | C | G | 0.31 | 0.01 | 0.002 | 8.80E-11 | 0.009 | 0.007 | 0.205 | 43.56 |
| rs232496 | T | C | 0.37 | 0.009 | 0.002 | 7.14E-10 | 0.003 | 0.006 | 0.689 | 36 |
| rs2364544 | A | G | 0.39 | -0.011 | 0.001 | 2.91E-13 | -0.003 | 0.006 | 0.673 | 56.25 |
| rs2434672 | A | C | 0.53 | 0.009 | 0.001 | 1.06E-10 | 0.006 | 0.006 | 0.386 | 42.25 |
| rs2436760 | T | C | 0.97 | 0.023 | 0.004 | 4.52E-08 | 0.015 | 0.018 | 0.404 | 30.51 |
| rs2447097 | T | G | 0.46 | 0.01 | 0.001 | 1.10E-11 | 0.009 | 0.006 | 0.174 | 47.02 |
| rs2469226 | A | T | 0.22 | 0.01 | 0.002 | 1.28E-08 | -0.012 | 0.007 | 0.101 | 31.89 |
| rs2496482 | T | C | 0.42 | 0.012 | 0.002 | 9.20E-17 | -0.024 | 0.006 | 1.86E-04 | 65.07 |
| rs252991 | A | G | 0.37 | 0.01 | 0.002 | 6.30E-11 | -0.005 | 0.006 | 0.434 | 40.11 |
| rs2545795 | A | C | 0.44 | 0.013 | 0.001 | 1.03E-18 | 0.002 | 0.006 | 0.703 | 79.72 |
| rs2554835 | A | G | 0.4 | 0.009 | 0.001 | 8.95E-10 | 0.002 | 0.006 | 0.748 | 39.51 |
| rs2570497 | T | C | 0.64 | -0.012 | 0.002 | 1.27E-16 | 0 | 0.007 | 0.942 | 65.07 |
| rs2725370 | T | C | 0.3 | -0.014 | 0.002 | 4.96E-20 | -0.011 | 0.007 | 0.096 | 88.36 |
| rs2764684 | T | C | 0.83 | 0.014 | 0.002 | 9.99E-15 | -0.002 | 0.008 | 0.785 | 56.65 |
| rs2838006 | T | C | 0.36 | 0.013 | 0.002 | 1.38E-19 | -0.014 | 0.007 | 0.029 | 77.44 |
| rs28513882 | A | G | 0.18 | -0.011 | 0.002 | 2.03E-09 | -0.006 | 0.008 | 0.476 | 37.35 |
| rs2885198 | A | G | 0.53 | 0.009 | 0.001 | 4.63E-10 | 0.008 | 0.006 | 0.201 | 39.51 |
| rs2923424 | A | G | 0.61 | 0.012 | 0.001 | 6.31E-17 | -0.008 | 0.006 | 0.21 | 73.47 |
| rs2929032 | A | G | 0.47 | -0.008 | 0.001 | 8.40E-09 | 0.011 | 0.006 | 0.079 | 33.47 |
| rs2958182 | A | T | 0.34 | 0.008 | 0.002 | 3.46E-08 | 0.005 | 0.007 | 0.468 | 29.88 |
| rs2964199 | T | C | 0.31 | -0.01 | 0.002 | 4.61E-11 | -0.007 | 0.007 | 0.32 | 43.56 |
| rs2964255 | A | G | 0.31 | 0.009 | 0.002 | 6.34E-09 | 0.006 | 0.007 | 0.401 | 34.42 |
| rs2998299 | T | G | 0.21 | -0.014 | 0.002 | 1.51E-15 | -0.005 | 0.008 | 0.519 | 64 |
| rs3026996 | A | C | 0.76 | 0.014 | 0.002 | 5.42E-17 | -0.001 | 0.007 | 0.911 | 74.39 |
| rs303752 | A | G | 0.41 | -0.011 | 0.001 | 1.47E-14 | 0.007 | 0.006 | 0.306 | 61.73 |
| rs3111251 | T | C | 0.43 | -0.009 | 0.001 | 3.88E-10 | -0.008 | 0.006 | 0.212 | 41.33 |
| rs34067381 | T | G | 0.36 | -0.01 | 0.002 | 9.77E-13 | -0.007 | 0.007 | 0.263 | 48.07 |
| rs34098770 | A | G | 0.15 | -0.016 | 0.002 | 2.95E-16 | -0.006 | 0.009 | 0.511 | 64 |
| rs34305371 | A | G | 0.1 | 0.031 | 0.002 | 1.10E-39 | -0.011 | 0.01 | 0.284 | 171.17 |
| rs34394051 | A | G | 0.84 | -0.012 | 0.002 | 2.30E-09 | -0.021 | 0.009 | 0.014 | 34.22 |
| rs34720381 | T | C | 0.09 | -0.018 | 0.002 | 1.90E-13 | 0.015 | 0.011 | 0.157 | 56.25 |
| rs34807077 | A | C | 0.16 | 0.013 | 0.002 | 3.29E-11 | -0.015 | 0.008 | 0.08 | 45.39 |
| rs35417702 | T | C | 0.52 | -0.015 | 0.001 | 2.40E-27 | -0.004 | 0.006 | 0.488 | 117.88 |
| rs35606437 | A | G | 0.27 | 0.011 | 0.002 | 3.98E-11 | 0.006 | 0.007 | 0.408 | 43.07 |
| rs35929923 | A | G | 0.25 | -0.011 | 0.002 | 1.77E-12 | -0.011 | 0.007 | 0.14 | 50.77 |
| rs363096 | T | C | 0.42 | -0.014 | 0.001 | 5.77E-24 | 0 | 0.006 | 0.971 | 104.33 |
| rs3751331 | A | G | 0.38 | -0.01 | 0.001 | 1.75E-11 | -0.004 | 0.006 | 0.514 | 48.01 |
| rs3809634 | A | G | 0.68 | -0.011 | 0.002 | 6.82E-13 | 0.003 | 0.007 | 0.665 | 52.8 |
| rs382196 | T | G | 0.38 | -0.008 | 0.001 | 1.47E-08 | 0.005 | 0.006 | 0.438 | 34.31 |
| rs3859523 | T | C | 0.82 | 0.012 | 0.002 | 9.92E-12 | -0.003 | 0.008 | 0.73 | 47.46 |
| rs3897821 | A | G | 0.67 | 0.015 | 0.002 | 3.58E-25 | 0.002 | 0.007 | 0.814 | 105.4 |
| rs3948495 | T | G | 0.41 | -0.009 | 0.001 | 3.95E-10 | 0.01 | 0.006 | 0.124 | 40.41 |
| rs401526 | T | C | 0.53 | -0.01 | 0.001 | 1.15E-13 | -0.038 | 0.006 | 8.50E-10 | 55.18 |
| rs401966 | C | G | 0.39 | -0.01 | 0.001 | 1.72E-11 | 0.005 | 0.006 | 0.464 | 48.01 |
| rs42210 | C | G | 0.71 | -0.01 | 0.002 | 2.74E-10 | 0.018 | 0.007 | 0.009 | 37.52 |
| rs42302 | A | G | 0.34 | 0.009 | 0.002 | 5.96E-09 | 0.013 | 0.007 | 0.057 | 32.87 |
| rs4298514 | T | C | 0.7 | 0.01 | 0.002 | 1.07E-11 | 0.012 | 0.007 | 0.082 | 48.07 |
| rs4320563 | A | G | 0.51 | 0.011 | 0.001 | 2.24E-14 | -0.005 | 0.006 | 0.422 | 60.62 |
| rs4328757 | T | C | 0.61 | 0.01 | 0.001 | 2.95E-11 | -0.004 | 0.006 | 0.557 | 47.02 |
| rs4352658 | T | C | 0.08 | -0.019 | 0.003 | 1.71E-13 | -0.013 | 0.011 | 0.257 | 53.4 |
| rs4358081 | A | C | 0.53 | -0.01 | 0.001 | 1.28E-11 | 0.003 | 0.006 | 0.605 | 46.05 |
| rs4369924 | A | G | 0.16 | 0.013 | 0.002 | 5.58E-11 | 0.013 | 0.009 | 0.127 | 43.28 |
| rs4384309 | A | G | 0.46 | 0.01 | 0.001 | 4.89E-13 | -0.004 | 0.006 | 0.498 | 53.08 |
| rs4467547 | T | G | 0.41 | 0.012 | 0.001 | 4.88E-17 | 0.007 | 0.006 | 0.299 | 74.7 |
| rs4500930 | T | C | 0.34 | -0.011 | 0.002 | 1.67E-13 | 0.003 | 0.007 | 0.607 | 52.8 |
| rs4663617 | A | T | 0.23 | 0.01 | 0.002 | 4.12E-10 | 0.007 | 0.008 | 0.379 | 37.43 |
| rs4673840 | T | C | 0.84 | -0.014 | 0.002 | 5.86E-13 | -0.005 | 0.008 | 0.566 | 52.75 |
| rs4675248 | A | G | 0.4 | -0.009 | 0.001 | 4.50E-11 | -0.004 | 0.006 | 0.541 | 45.08 |
| rs4719944 | T | C | 0.55 | -0.012 | 0.001 | 9.25E-18 | 0.006 | 0.006 | 0.307 | 74.7 |
| rs4726070 | A | G | 0.6 | 0.012 | 0.001 | 1.36E-17 | 0.009 | 0.006 | 0.141 | 75.94 |
| rs4757957 | C | G | 0.69 | 0.013 | 0.002 | 1.79E-18 | 0.017 | 0.007 | 0.009 | 78.62 |
| rs4766424 | C | G | 0.12 | 0.014 | 0.002 | 2.47E-11 | 0.007 | 0.009 | 0.473 | 45.72 |
| rs4778058 | T | C | 0.5 | -0.009 | 0.001 | 4.92E-11 | -0.013 | 0.006 | 0.044 | 43.18 |
| rs4787457 | A | G | 0.63 | 0.016 | 0.002 | 1.23E-28 | 0.021 | 0.006 | 0.001 | 116.64 |
| rs4793090 | A | G | 0.66 | 0.008 | 0.002 | 1.67E-08 | -0.001 | 0.007 | 0.825 | 31.36 |
| rs4810894 | A | G | 0.37 | -0.009 | 0.002 | 4.38E-09 | 0.002 | 0.006 | 0.728 | 32.11 |
| rs4839155 | T | G | 0.77 | 0.012 | 0.002 | 4.66E-12 | 0.003 | 0.007 | 0.728 | 45.76 |
| rs4846724 | A | G | 0.53 | 0.01 | 0.001 | 2.89E-12 | 0.002 | 0.006 | 0.716 | 49 |
| rs4848924 | A | C | 0.71 | -0.012 | 0.002 | 3.56E-14 | -0.006 | 0.007 | 0.383 | 60.84 |
| rs488476 | C | G | 0.62 | -0.011 | 0.002 | 8.29E-15 | -0.011 | 0.006 | 0.098 | 56.75 |
| rs4895650 | T | C | 0.55 | 0.009 | 0.001 | 5.31E-11 | -0.002 | 0.006 | 0.697 | 44.13 |
| rs4899012 | C | G | 0.6 | -0.01 | 0.001 | 8.06E-13 | -0.012 | 0.006 | 0.061 | 54.13 |
| rs4904523 | A | G | 0.52 | -0.008 | 0.001 | 6.27E-09 | -0.004 | 0.006 | 0.485 | 34.31 |
| rs4915735 | A | G | 0.14 | 0.012 | 0.002 | 9.56E-09 | -0.009 | 0.009 | 0.29 | 33.64 |
| rs4977885 | A | G | 0.4 | -0.008 | 0.001 | 6.78E-09 | -0.001 | 0.006 | 0.89 | 36 |
| rs4984541 | A | G | 0.77 | -0.014 | 0.002 | 3.40E-16 | -0.001 | 0.007 | 0.917 | 65.9 |
| rs55736314 | C | G | 0.6 | -0.015 | 0.001 | 4.59E-27 | -0.016 | 0.006 | 0.009 | 121 |
| rs56099375 | T | C | 0.24 | 0.012 | 0.002 | 3.10E-13 | -0.007 | 0.007 | 0.353 | 49.83 |
| rs56319902 | T | C | 0.22 | -0.021 | 0.002 | 5.81E-33 | -0.068 | 0.007 | 7.78E-20 | 146.84 |
| rs56391344 | A | G | 0.25 | 0.015 | 0.002 | 7.49E-20 | -0.016 | 0.007 | 0.029 | 85.56 |
| rs56794817 | A | G | 0.16 | 0.012 | 0.002 | 1.69E-10 | 0.009 | 0.009 | 0.317 | 41.23 |
| rs57349798 | A | G | 0.41 | 0.009 | 0.001 | 8.63E-11 | 0 | 0.006 | 0.972 | 44.13 |
| rs57352738 | A | T | 0.2 | -0.015 | 0.002 | 2.51E-18 | 0.008 | 0.008 | 0.286 | 79.94 |
| rs575113 | A | G | 0.29 | 0.013 | 0.002 | 7.52E-18 | 0.01 | 0.007 | 0.163 | 78.62 |
| rs5754753 | T | C | 0.72 | -0.013 | 0.002 | 6.15E-16 | -0.01 | 0.007 | 0.133 | 62.02 |
| rs57661533 | T | C | 0.13 | -0.011 | 0.002 | 3.24E-08 | 0.013 | 0.009 | 0.148 | 29.47 |
| rs580652 | T | C | 0.9 | -0.015 | 0.002 | 4.65E-10 | 0.002 | 0.011 | 0.846 | 37.52 |
| rs590013 | T | C | 0.68 | 0.01 | 0.002 | 2.47E-11 | -0.005 | 0.007 | 0.481 | 44.44 |
| rs6043521 | T | C | 0.6 | -0.008 | 0.001 | 9.72E-09 | 0.013 | 0.006 | 0.045 | 35.15 |
| rs6065080 | T | C | 0.36 | -0.013 | 0.002 | 7.49E-19 | -0.009 | 0.006 | 0.151 | 75.11 |
| rs6065784 | C | G | 0.7 | 0.011 | 0.002 | 5.78E-13 | 0.007 | 0.007 | 0.283 | 53.78 |
| rs61104616 | A | G | 0.52 | -0.017 | 0.001 | 1.56E-34 | 0.021 | 0.006 | 9.69E-04 | 150.94 |
| rs6123924 | A | G | 0.84 | 0.013 | 0.002 | 3.20E-11 | 0.024 | 0.009 | 0.006 | 46.1 |
| rs61527214 | A | G | 0.41 | 0.011 | 0.001 | 1.13E-14 | 0.002 | 0.006 | 0.788 | 61.73 |
| rs61997667 | T | C | 0.15 | -0.014 | 0.002 | 2.24E-13 | -0.003 | 0.009 | 0.748 | 51.84 |
| rs62155350 | A | G | 0.02 | 0.034 | 0.005 | 1.69E-10 | 0.019 | 0.022 | 0.388 | 40.67 |
| rs62155873 | T | C | 0.12 | -0.015 | 0.002 | 1.65E-12 | -0.002 | 0.009 | 0.83 | 51.7 |
| rs62174974 | A | G | 0.19 | -0.011 | 0.002 | 2.76E-09 | -0.005 | 0.008 | 0.56 | 34.03 |
| rs62179650 | A | G | 0.3 | 0.011 | 0.002 | 5.66E-13 | 0.011 | 0.007 | 0.089 | 49 |
| rs62182994 | T | C | 0.69 | 0.014 | 0.002 | 2.55E-19 | 0 | 0.007 | 0.951 | 83.42 |
| rs62379838 | T | C | 0.7 | 0.012 | 0.002 | 1.77E-15 | -0.002 | 0.007 | 0.747 | 65.07 |
| rs628993 | A | G | 0.11 | 0.013 | 0.002 | 3.83E-08 | 0.003 | 0.011 | 0.812 | 30.97 |
| rs6449503 | A | G | 0.5 | 0.018 | 0.001 | 1.84E-39 | 0.015 | 0.006 | 0.018 | 174.62 |
| rs6490618 | T | C | 0.33 | 0.009 | 0.002 | 3.48E-09 | 0.005 | 0.007 | 0.47 | 34.42 |
| rs6493265 | T | C | 0.39 | -0.012 | 0.001 | 2.26E-17 | -0.005 | 0.006 | 0.459 | 75.94 |
| rs6557171 | T | C | 0.32 | -0.016 | 0.002 | 5.48E-25 | -0.005 | 0.007 | 0.464 | 106.78 |
| rs660001 | A | G | 0.21 | -0.016 | 0.002 | 1.66E-21 | 0.001 | 0.008 | 0.862 | 93.07 |
| rs66568921 | T | G | 0.64 | -0.018 | 0.002 | 7.60E-34 | -0.005 | 0.007 | 0.448 | 145.6 |
| rs66721975 | A | G | 0.29 | -0.009 | 0.002 | 1.51E-08 | 0.014 | 0.007 | 0.052 | 33.64 |
| rs6690195 | T | C | 0.49 | -0.011 | 0.001 | 7.89E-16 | 0.008 | 0.006 | 0.228 | 65.15 |
| rs6697584 | T | C | 0.79 | -0.013 | 0.002 | 1.43E-13 | 0.006 | 0.008 | 0.434 | 54.93 |
| rs6704768 | A | G | 0.56 | -0.012 | 0.001 | 1.84E-16 | -0.006 | 0.006 | 0.368 | 68.65 |
| rs6731373 | A | G | 0.34 | -0.012 | 0.002 | 1.39E-14 | -0.008 | 0.006 | 0.223 | 58.78 |
| rs6812533 | T | C | 0.74 | 0.011 | 0.002 | 4.85E-11 | -0.011 | 0.007 | 0.114 | 43.89 |
| rs68145588 | T | G | 0.14 | -0.013 | 0.002 | 7.53E-10 | 0.024 | 0.009 | 0.008 | 36.57 |
| rs6917154 | T | C | 0.12 | 0.013 | 0.002 | 2.55E-09 | -0.003 | 0.009 | 0.738 | 36.57 |
| rs6924023 | A | G | 0.75 | 0.011 | 0.002 | 5.17E-11 | -0.015 | 0.007 | 0.042 | 44.72 |
| rs6977237 | T | C | 0.47 | 0.008 | 0.001 | 5.59E-09 | 0.005 | 0.006 | 0.43 | 34.31 |
| rs6994287 | A | G | 0.41 | 0.01 | 0.001 | 1.11E-12 | -0.002 | 0.006 | 0.757 | 53.08 |
| rs7012546 | T | C | 0.42 | 0.009 | 0.001 | 1.25E-10 | -0.005 | 0.006 | 0.402 | 43.18 |
| rs7029718 | A | G | 0.41 | 0.024 | 0.001 | 7.82E-65 | 0.007 | 0.006 | 0.301 | 303.76 |
| rs7041702 | A | G | 0.74 | -0.011 | 0.002 | 4.53E-12 | -0.003 | 0.007 | 0.652 | 48.13 |
| rs7167688 | T | C | 0.49 | -0.009 | 0.001 | 1.71E-11 | 0.001 | 0.006 | 0.904 | 45.08 |
| rs7171405 | A | G | 0.24 | -0.01 | 0.002 | 7.53E-10 | -0.008 | 0.007 | 0.261 | 39.85 |
| rs717996 | T | C | 0.6 | -0.012 | 0.001 | 1.89E-17 | -0.003 | 0.006 | 0.648 | 75.94 |
| rs7226824 | T | C | 0.53 | -0.008 | 0.001 | 2.72E-08 | 0 | 0.006 | 0.938 | 31.04 |
| rs72486027 | T | C | 0.25 | -0.01 | 0.002 | 1.61E-10 | 0.008 | 0.007 | 0.261 | 42.25 |
| rs7254263 | T | C | 0.28 | -0.012 | 0.002 | 1.84E-14 | -0.009 | 0.007 | 0.193 | 55.32 |
| rs72622559 | T | C | 0.23 | -0.012 | 0.002 | 5.50E-13 | -0.005 | 0.007 | 0.456 | 49.83 |
| rs72672052 | A | T | 0.18 | -0.01 | 0.002 | 4.87E-08 | -0.013 | 0.008 | 0.116 | 31.48 |
| rs72709560 | A | G | 0.69 | 0.009 | 0.002 | 2.03E-08 | 0.009 | 0.007 | 0.189 | 32.11 |
| rs72828517 | T | C | 0.83 | -0.017 | 0.002 | 1.38E-19 | -0.013 | 0.008 | 0.119 | 78.18 |
| rs72829857 | A | G | 0.76 | -0.015 | 0.002 | 1.52E-19 | -0.002 | 0.007 | 0.839 | 86.72 |
| rs72917504 | T | C | 0.06 | -0.018 | 0.003 | 2.91E-09 | -0.003 | 0.013 | 0.814 | 35.2 |
| rs72944064 | T | C | 0.74 | -0.01 | 0.002 | 9.39E-10 | -0.003 | 0.007 | 0.655 | 37.52 |
| rs72962169 | T | C | 0.16 | -0.017 | 0.002 | 1.26E-19 | -0.013 | 0.009 | 0.119 | 83.87 |
| rs72993796 | T | C | 0.88 | 0.013 | 0.002 | 1.12E-08 | 0.018 | 0.01 | 0.057 | 33.32 |
| rs730384 | A | G | 0.44 | 0.01 | 0.001 | 9.38E-13 | -0.009 | 0.006 | 0.152 | 52.05 |
| rs73055556 | A | G | 0.14 | 0.011 | 0.002 | 4.95E-08 | 0.037 | 0.009 | 4.26E-05 | 30.8 |
| rs73191311 | A | G | 0.34 | -0.009 | 0.002 | 2.03E-10 | 0.003 | 0.007 | 0.611 | 39.27 |
| rs7321274 | A | G | 0.8 | 0.011 | 0.002 | 8.21E-11 | -0.012 | 0.008 | 0.11 | 40.11 |
| rs7326331 | A | G | 0.28 | -0.013 | 0.002 | 3.42E-16 | 0 | 0.007 | 0.998 | 64 |
| rs73344830 | A | G | 0.42 | 0.017 | 0.001 | 1.00E-32 | -0.008 | 0.006 | 0.2 | 147.45 |
| rs736281 | T | C | 0.39 | 0.009 | 0.001 | 2.49E-09 | 0 | 0.006 | 0.977 | 37.73 |
| rs73648455 | T | C | 0.08 | -0.018 | 0.003 | 1.99E-12 | 0.004 | 0.012 | 0.762 | 49.54 |
| rs743316 | T | C | 0.79 | 0.011 | 0.002 | 8.21E-11 | -0.001 | 0.008 | 0.848 | 43.4 |
| rs74415461 | T | C | 0.08 | 0.018 | 0.003 | 6.25E-12 | 0.017 | 0.011 | 0.127 | 45.82 |
| rs7449561 | A | G | 0.23 | 0.01 | 0.002 | 9.73E-10 | -0.002 | 0.007 | 0.767 | 36 |
| rs7460106 | T | C | 0.76 | -0.012 | 0.002 | 2.06E-13 | -0.005 | 0.007 | 0.478 | 52.35 |
| rs746839 | C | G | 0.63 | 0.014 | 0.002 | 1.19E-20 | -0.004 | 0.007 | 0.544 | 81 |
| rs74787922 | A | C | 0.93 | 0.018 | 0.003 | 8.97E-11 | 0.013 | 0.012 | 0.299 | 42.25 |
| rs75033012 | C | G | 0.04 | -0.026 | 0.004 | 1.38E-11 | 0.008 | 0.016 | 0.621 | 46.1 |
| rs75177132 | T | C | 0.05 | 0.03 | 0.004 | 1.01E-16 | 0.021 | 0.014 | 0.149 | 68.52 |
| rs7575637 | A | G | 0.45 | 0.011 | 0.001 | 3.43E-14 | 0.006 | 0.006 | 0.297 | 58.41 |
| rs7603132 | A | G | 0.19 | 0.014 | 0.002 | 1.20E-14 | 0.004 | 0.008 | 0.585 | 57.93 |
| rs76076331 | T | C | 0.13 | 0.021 | 0.002 | 1.71E-23 | -0.016 | 0.009 | 0.093 | 96.23 |
| rs7617204 | A | G | 0.52 | -0.01 | 0.001 | 4.33E-12 | -0.005 | 0.006 | 0.443 | 48.01 |
| rs76235882 | A | G | 0.97 | 0.024 | 0.004 | 4.52E-10 | -0.02 | 0.019 | 0.283 | 38.19 |
| rs76246107 | A | G | 0.09 | -0.016 | 0.003 | 6.52E-10 | -0.044 | 0.012 | 2.84E-04 | 38.82 |
| rs7650602 | T | C | 0.56 | -0.009 | 0.001 | 2.77E-10 | -0.016 | 0.006 | 0.01 | 40.41 |
| rs76577427 | C | G | 0.9 | 0.02 | 0.002 | 7.78E-18 | -0.003 | 0.01 | 0.789 | 71.54 |
| rs7672622 | A | G | 0.75 | -0.009 | 0.002 | 4.72E-08 | -0.012 | 0.007 | 0.088 | 30.25 |
| rs7683416 | T | C | 0.46 | 0.014 | 0.001 | 6.05E-23 | 0.02 | 0.006 | 0.001 | 98.58 |
| rs76878669 | C | G | 0.76 | 0.014 | 0.002 | 3.10E-17 | 0.011 | 0.007 | 0.157 | 68.79 |
| rs7692359 | T | C | 0.78 | -0.011 | 0.002 | 1.40E-10 | -0.016 | 0.008 | 0.03 | 41.11 |
| rs77025239 | A | G | 0.16 | -0.014 | 0.002 | 4.65E-13 | -0.017 | 0.009 | 0.056 | 54.29 |
| rs77609760 | A | G | 0.93 | 0.024 | 0.003 | 7.71E-17 | -0.007 | 0.012 | 0.598 | 71.04 |
| rs77719387 | A | T | 0.02 | -0.04 | 0.006 | 3.78E-12 | -0.039 | 0.026 | 0.14 | 48.28 |
| rs7849487 | T | G | 0.66 | -0.017 | 0.002 | 1.25E-29 | 0.011 | 0.007 | 0.085 | 123.95 |
| rs78648104 | T | C | 0.91 | -0.014 | 0.002 | 1.70E-08 | -0.026 | 0.011 | 0.02 | 31.81 |
| rs7875078 | A | C | 0.46 | -0.008 | 0.001 | 2.72E-08 | 0.005 | 0.006 | 0.458 | 31.04 |
| rs7894722 | T | C | 0.37 | 0.009 | 0.001 | 5.41E-09 | -0.005 | 0.006 | 0.416 | 36.86 |
| rs790647 | A | C | 0.23 | -0.015 | 0.002 | 3.81E-18 | -0.006 | 0.007 | 0.442 | 72.75 |
| rs7910403 | T | G | 0.2 | 0.012 | 0.002 | 5.08E-12 | -0.006 | 0.008 | 0.44 | 45.94 |
| rs7924036 | T | G | 0.51 | 0.013 | 0.001 | 2.55E-21 | 0.001 | 0.006 | 0.876 | 90.25 |
| rs79265434 | A | G | 0.88 | -0.02 | 0.002 | 1.98E-19 | -0.004 | 0.01 | 0.69 | 80.18 |
| rs7972246 | T | C | 0.35 | 0.011 | 0.002 | 1.01E-13 | 0.013 | 0.007 | 0.054 | 53.78 |
| rs79728014 | A | G | 0.86 | -0.013 | 0.002 | 2.06E-11 | -0.023 | 0.009 | 0.01 | 44.89 |
| rs7974852 | A | C | 0.52 | 0.012 | 0.001 | 1.14E-16 | -0.002 | 0.006 | 0.785 | 69.84 |
| rs7977614 | A | G | 0.71 | -0.013 | 0.002 | 6.84E-17 | -0.006 | 0.007 | 0.369 | 70.14 |
| rs79994730 | T | C | 0.04 | 0.024 | 0.004 | 5.88E-12 | -0.032 | 0.014 | 0.024 | 47.02 |
| rs8008382 | T | C | 0.3 | -0.011 | 0.002 | 4.07E-12 | -0.018 | 0.007 | 0.01 | 49.94 |
| rs8024 | A | C | 0.33 | -0.012 | 0.002 | 1.13E-14 | 0.009 | 0.007 | 0.184 | 58.78 |
| rs80257979 | T | G | 0.97 | -0.025 | 0.004 | 3.31E-10 | -0.031 | 0.018 | 0.087 | 39.69 |
| rs8030487 | A | G | 0.69 | 0.009 | 0.002 | 2.52E-08 | -0.002 | 0.007 | 0.775 | 32.11 |
| rs806816 | A | T | 0.75 | 0.01 | 0.002 | 4.80E-10 | 0.004 | 0.007 | 0.619 | 39.85 |
| rs8097125 | T | C | 0.38 | 0.009 | 0.001 | 2.31E-09 | 0.006 | 0.006 | 0.332 | 37.73 |
| rs912883 | T | C | 0.68 | 0.009 | 0.002 | 6.20E-09 | 0.006 | 0.007 | 0.388 | 33.64 |
| rs9289300 | T | C | 0.84 | -0.016 | 0.002 | 1.00E-17 | 0.004 | 0.009 | 0.615 | 74.5 |
| rs929511 | T | C | 0.13 | -0.016 | 0.002 | 3.91E-14 | 0.013 | 0.01 | 0.178 | 58.05 |
| rs9359939 | A | C | 0.24 | -0.011 | 0.002 | 1.51E-10 | 0.007 | 0.007 | 0.362 | 43.07 |
| rs936496 | A | G | 0.62 | -0.008 | 0.001 | 4.38E-08 | -0.005 | 0.006 | 0.437 | 31.84 |
| rs9373363 | A | G | 0.75 | -0.011 | 0.002 | 5.54E-12 | -0.003 | 0.007 | 0.661 | 48.13 |
| rs9375188 | T | C | 0.48 | 0.021 | 0.001 | 8.78E-52 | 0.015 | 0.006 | 0.019 | 229.31 |
| rs9411331 | A | C | 0.68 | 0.015 | 0.002 | 7.63E-22 | -0.006 | 0.007 | 0.367 | 93.44 |
| rs9492774 | C | G | 0.37 | 0.008 | 0.002 | 4.33E-08 | 0.003 | 0.006 | 0.623 | 28.44 |
| rs9529146 | T | C | 0.23 | 0.013 | 0.002 | 3.48E-14 | 0.012 | 0.007 | 0.09 | 55.81 |
| rs9556958 | T | C | 0.52 | -0.011 | 0.001 | 5.96E-15 | -0.009 | 0.006 | 0.154 | 61.73 |
| rs9616947 | T | C | 0.38 | -0.013 | 0.002 | 3.58E-20 | -0.002 | 0.006 | 0.794 | 79.8 |
| rs9633970 | T | C | 0.76 | 0.009 | 0.002 | 2.81E-08 | -0.013 | 0.008 | 0.092 | 32.35 |
| rs9655780 | A | G | 0.83 | -0.015 | 0.002 | 3.70E-16 | 0.015 | 0.008 | 0.085 | 64.84 |
| rs969512 | A | T | 0.66 | -0.011 | 0.002 | 9.84E-13 | -0.003 | 0.007 | 0.605 | 49.94 |
| rs9853928 | T | C | 0.21 | -0.013 | 0.002 | 1.59E-13 | -0.001 | 0.008 | 0.897 | 55.81 |
| rs9859556 | T | G | 0.31 | 0.029 | 0.002 | 4.61E-82 | 0.027 | 0.007 | 5.66E-05 | 373.78 |
| rs9882532 | T | C | 0.64 | 0.012 | 0.002 | 2.15E-17 | 0.005 | 0.007 | 0.424 | 68.34 |
| rs9886703 | A | T | 0.17 | -0.014 | 0.002 | 4.46E-13 | -0.004 | 0.008 | 0.604 | 51.24 |
| rs9927137 | A | G | 0.53 | 0.008 | 0.001 | 5.11E-09 | 0.013 | 0.006 | 0.038 | 34.31 |
| rs9927842 | T | C | 0.15 | -0.013 | 0.002 | 6.22E-11 | 0.011 | 0.009 | 0.218 | 42.25 |
| rs9929556 | T | G | 0.57 | -0.01 | 0.001 | 2.31E-12 | 0.001 | 0.006 | 0.877 | 50.01 |
| rs9929762 | A | G | 0.56 | 0.01 | 0.001 | 1.03E-11 | 0.007 | 0.006 | 0.273 | 47.02 |
| rs9933256 | A | G | 0.56 | 0.012 | 0.001 | 6.06E-17 | 0 | 0.006 | 0.962 | 72.25 |

**SNPs used as instrumental variables in the MR analysis of educational attainment on cortical thickness**

| **SNP** | **Effect allele** | **Other allele** | **Effect allele frequency** | **beta (exposure)** | **SE (exposure)** | ***P* (exposure)** | **beta (outcome)** | **SE (outcome)** | ***P* (outcome)** | ***F*-statistic** |
| --- | --- | --- | --- | --- | --- | --- | --- | --- | --- | --- |
| rs10006235 | T | C | 0.27 | -0.009 | 0.002 | 4.48E-09 | 0.002 | 0.007 | 0.754 | 33.79 |
| rs10021733 | T | C | 0.15 | -0.016 | 0.003 | 9.45E-10 | 0.003 | 0.009 | 0.736 | 36.89 |
| rs10046069 | A | G | 0.12 | 0.013 | 0.002 | 4.40E-09 | 0.016 | 0.01 | 0.108 | 33.32 |
| rs10060023 | T | C | 0.33 | 0.011 | 0.002 | 1.88E-13 | 0.007 | 0.007 | 0.273 | 53.78 |
| rs10080647 | A | C | 0.14 | 0.012 | 0.002 | 7.05E-10 | 0.009 | 0.009 | 0.316 | 38.44 |
| rs1008078 | T | C | 0.4 | -0.018 | 0.001 | 4.50E-37 | 0.002 | 0.007 | 0.755 | 169 |
| rs10189857 | A | G | 0.57 | 0.016 | 0.001 | 4.69E-29 | -0.003 | 0.007 | 0.624 | 127.37 |
| rs10193498 | A | T | 0.76 | 0.01 | 0.002 | 3.10E-09 | 0.001 | 0.008 | 0.942 | 36.75 |
| rs10215082 | A | G | 0.47 | -0.015 | 0.001 | 7.03E-26 | -0.015 | 0.006 | 0.017 | 113.27 |
| rs10460095 | A | G | 0.57 | -0.012 | 0.001 | 2.61E-18 | 0.002 | 0.007 | 0.71 | 78.45 |
| rs10761251 | A | T | 0.66 | 0.011 | 0.002 | 2.52E-14 | -0.008 | 0.007 | 0.222 | 56.75 |
| rs10772644 | C | G | 0.89 | 0.014 | 0.002 | 1.69E-10 | 0.023 | 0.01 | 0.026 | 41.66 |
| rs10773002 | A | T | 0.25 | 0.018 | 0.002 | 2.50E-30 | -0.003 | 0.007 | 0.716 | 133.69 |
| rs10773208 | T | C | 0.25 | -0.011 | 0.002 | 8.09E-12 | 0.001 | 0.007 | 0.932 | 47.27 |
| rs10798888 | T | G | 0.17 | -0.014 | 0.002 | 5.15E-14 | -0.008 | 0.009 | 0.324 | 54.29 |
| rs10799615 | A | G | 0.25 | -0.009 | 0.002 | 4.11E-08 | 0.003 | 0.007 | 0.719 | 30.94 |
| rs10805383 | A | G | 0.49 | -0.01 | 0.001 | 2.22E-13 | 0.009 | 0.006 | 0.178 | 54.13 |
| rs10830858 | T | C | 0.51 | 0.009 | 0.001 | 5.31E-11 | 0.006 | 0.006 | 0.386 | 43.18 |
| rs10844179 | A | G | 0.23 | 0.01 | 0.002 | 5.03E-10 | 0.014 | 0.008 | 0.071 | 37.43 |
| rs10875121 | C | G | 0.84 | 0.017 | 0.002 | 4.46E-20 | 0.002 | 0.009 | 0.848 | 83.87 |
| rs10892807 | T | C | 0.57 | 0.011 | 0.001 | 1.09E-14 | 0.013 | 0.007 | 0.054 | 60.62 |
| rs10940921 | T | G | 0.42 | 0.011 | 0.001 | 1.31E-14 | 0 | 0.007 | 0.962 | 62.86 |
| rs10951590 | T | C | 0.33 | -0.011 | 0.002 | 3.62E-13 | -0.013 | 0.007 | 0.052 | 52.8 |
| rs10979613 | T | C | 0.64 | -0.012 | 0.002 | 1.25E-15 | 0.009 | 0.007 | 0.169 | 60.84 |
| rs10996167 | C | G | 0.65 | 0.011 | 0.002 | 7.47E-09 | -0.001 | 0.007 | 0.844 | 31.92 |
| rs11003463 | T | G | 0.61 | 0.009 | 0.002 | 5.58E-10 | 0.012 | 0.007 | 0.062 | 36.8 |
| rs11023764 | A | G | 0.64 | 0.009 | 0.002 | 5.32E-10 | -0.009 | 0.007 | 0.168 | 36.8 |
| rs11030102 | C | G | 0.75 | 0.01 | 0.002 | 4.60E-10 | -0.02 | 0.007 | 0.007 | 39.85 |
| rs11076962 | T | C | 0.72 | 0.009 | 0.002 | 4.29E-08 | -0.005 | 0.007 | 0.517 | 28.89 |
| rs11081529 | T | C | 0.71 | 0.013 | 0.002 | 5.22E-16 | -0.003 | 0.007 | 0.623 | 70.56 |
| rs11082011 | T | C | 0.67 | 0.019 | 0.002 | 2.06E-38 | 0.008 | 0.007 | 0.234 | 167.27 |
| rs11121177 | A | G | 0.18 | 0.015 | 0.002 | 1.44E-16 | 0.005 | 0.008 | 0.548 | 70.37 |
| rs11130380 | T | G | 0.61 | 0.009 | 0.001 | 4.17E-10 | -0.003 | 0.007 | 0.614 | 41.33 |
| rs11138947 | T | C | 0.72 | 0.011 | 0.002 | 2.36E-11 | 0.008 | 0.007 | 0.241 | 43.07 |
| rs11157931 | A | C | 0.39 | -0.014 | 0.001 | 3.86E-22 | 0.017 | 0.007 | 0.012 | 98.58 |
| rs111852224 | T | C | 0.12 | 0.016 | 0.002 | 4.85E-13 | 0.004 | 0.01 | 0.678 | 50.28 |
| rs11211123 | A | G | 0.22 | 0.01 | 0.002 | 7.62E-10 | 0.009 | 0.008 | 0.241 | 37.43 |
| rs11213482 | A | G | 0.84 | -0.011 | 0.002 | 2.68E-08 | -0.01 | 0.008 | 0.258 | 31.12 |
| rs112603734 | A | C | 0.77 | -0.013 | 0.002 | 2.72E-09 | -0.011 | 0.007 | 0.16 | 35.46 |
| rs112806496 | C | G | 0.91 | -0.016 | 0.002 | 1.72E-10 | 0.001 | 0.011 | 0.935 | 40.96 |
| rs1128956 | T | G | 0.83 | -0.014 | 0.002 | 5.34E-14 | -0.013 | 0.008 | 0.112 | 54.29 |
| rs113520408 | A | G | 0.28 | 0.016 | 0.002 | 1.03E-22 | 0 | 0.007 | 0.972 | 93.85 |
| rs113615161 | T | C | 0.13 | -0.012 | 0.002 | 2.98E-09 | -0.015 | 0.01 | 0.117 | 34.31 |
| rs114593137 | A | T | 0.79 | -0.012 | 0.002 | 1.65E-12 | -0.003 | 0.008 | 0.733 | 51.5 |
| rs115000530 | A | T | 0.95 | -0.025 | 0.003 | 1.09E-15 | 0.022 | 0.014 | 0.132 | 65.04 |
| rs11542663 | A | C | 0.69 | 0.011 | 0.002 | 2.44E-13 | 0.001 | 0.007 | 0.907 | 55.75 |
| rs11588857 | A | G | 0.21 | 0.02 | 0.002 | 4.45E-31 | 0.017 | 0.008 | 0.033 | 137.03 |
| rs11598765 | A | G | 0.2 | -0.011 | 0.002 | 1.73E-09 | -0.013 | 0.008 | 0.114 | 34.68 |
| rs11620355 | A | G | 0.09 | 0.018 | 0.002 | 1.09E-13 | -0.013 | 0.011 | 0.268 | 53 |
| rs11657342 | A | G | 0.37 | 0.014 | 0.002 | 2.32E-13 | -0.007 | 0.007 | 0.319 | 54.29 |
| rs11657979 | A | G | 0.24 | -0.009 | 0.002 | 4.57E-08 | -0.02 | 0.008 | 0.012 | 28.65 |
| rs11663602 | A | C | 0.28 | -0.013 | 0.002 | 9.27E-17 | 0 | 0.007 | 0.994 | 66.02 |
| rs1167827 | A | G | 0.43 | 0.01 | 0.001 | 2.93E-13 | 0.031 | 0.006 | 1.63E-06 | 55.18 |
| rs11724690 | T | G | 0.29 | 0.009 | 0.002 | 2.08E-09 | 0.003 | 0.007 | 0.718 | 38.44 |
| rs117398064 | C | G | 0.09 | -0.014 | 0.002 | 1.16E-08 | -0.004 | 0.011 | 0.729 | 31.81 |
| rs118134876 | T | C | 0.06 | -0.022 | 0.003 | 1.50E-13 | 0.027 | 0.013 | 0.042 | 55.25 |
| rs11871429 | A | G | 0.77 | 0.013 | 0.002 | 2.52E-14 | 0.002 | 0.008 | 0.773 | 56.69 |
| rs11894424 | A | C | 0.95 | 0.021 | 0.003 | 2.14E-10 | -0.01 | 0.016 | 0.536 | 40.11 |
| rs12054166 | C | G | 0.74 | 0.01 | 0.002 | 2.30E-09 | 0.006 | 0.007 | 0.451 | 36 |
| rs12076635 | C | G | 0.78 | 0.021 | 0.002 | 3.33E-34 | 0.01 | 0.008 | 0.218 | 148.27 |
| rs12113634 | T | C | 0.65 | 0.009 | 0.002 | 2.50E-09 | -0.002 | 0.007 | 0.772 | 34.42 |
| rs12151248 | T | C | 0.11 | -0.015 | 0.002 | 3.81E-11 | -0.003 | 0.01 | 0.753 | 41.97 |
| rs1220779 | A | G | 0.47 | -0.008 | 0.001 | 1.58E-08 | 0.006 | 0.006 | 0.362 | 31.84 |
| rs12273435 | A | G | 0.21 | -0.013 | 0.002 | 4.72E-14 | 0.002 | 0.008 | 0.852 | 60.29 |
| rs12332731 | A | T | 0.19 | 0.014 | 0.002 | 5.96E-14 | -0.013 | 0.008 | 0.122 | 56.25 |
| rs12375949 | T | C | 0.43 | -0.014 | 0.001 | 1.61E-22 | 0.011 | 0.006 | 0.094 | 97.16 |
| rs12438177 | A | G | 0.37 | 0.009 | 0.002 | 1.48E-10 | -0.005 | 0.007 | 0.419 | 38.44 |
| rs12468040 | T | G | 0.38 | 0.014 | 0.001 | 3.12E-21 | -0.006 | 0.007 | 0.362 | 95.76 |
| rs12503522 | T | C | 0.28 | -0.011 | 0.002 | 3.93E-13 | -0.011 | 0.007 | 0.131 | 49.88 |
| rs12506221 | T | G | 0.44 | -0.013 | 0.001 | 4.23E-20 | 0.001 | 0.007 | 0.906 | 86.22 |
| rs12519073 | T | C | 0.23 | -0.011 | 0.002 | 1.16E-10 | -0.006 | 0.008 | 0.403 | 39.62 |
| rs12524795 | T | C | 0.44 | 0.01 | 0.001 | 5.28E-13 | 0.016 | 0.006 | 0.013 | 53.08 |
| rs12571549 | A | G | 0.14 | 0.015 | 0.002 | 1.45E-13 | 0.009 | 0.009 | 0.344 | 57 |
| rs12574281 | A | C | 0.63 | -0.008 | 0.002 | 3.04E-08 | 0.006 | 0.007 | 0.386 | 28.44 |
| rs12591647 | T | C | 0.82 | -0.015 | 0.002 | 7.78E-17 | 0.003 | 0.008 | 0.696 | 71.31 |
| rs12602286 | T | G | 0.87 | 0.016 | 0.002 | 4.62E-14 | 0.019 | 0.01 | 0.052 | 58.05 |
| rs12614263 | A | G | 0.5 | 0.008 | 0.001 | 3.38E-09 | 0.002 | 0.006 | 0.706 | 35.15 |
| rs12638072 | A | G | 0.3 | -0.011 | 0.002 | 3.13E-12 | -0.007 | 0.007 | 0.345 | 50.88 |
| rs12643771 | T | C | 0.31 | 0.013 | 0.002 | 1.32E-17 | 0.002 | 0.007 | 0.724 | 75.11 |
| rs12646523 | T | C | 0.25 | -0.013 | 0.002 | 3.79E-16 | -0.014 | 0.007 | 0.051 | 68.06 |
| rs12670376 | A | G | 0.45 | 0.009 | 0.001 | 5.99E-11 | 0.007 | 0.006 | 0.302 | 43.18 |
| rs12761761 | T | C | 0.24 | 0.015 | 0.002 | 4.79E-20 | -0.005 | 0.007 | 0.538 | 81 |
| rs12765185 | A | T | 0.28 | -0.009 | 0.002 | 2.96E-08 | 0.01 | 0.007 | 0.159 | 30.25 |
| rs12789313 | T | C | 0.49 | 0.009 | 0.001 | 6.79E-11 | -0.006 | 0.006 | 0.363 | 43.18 |
| rs12875339 | A | C | 0.64 | -0.011 | 0.002 | 1.71E-13 | -0.007 | 0.007 | 0.324 | 51.84 |
| rs12888615 | T | C | 0.2 | 0.012 | 0.002 | 9.42E-12 | 0.002 | 0.008 | 0.757 | 44.44 |
| rs12957463 | A | G | 0.8 | 0.015 | 0.002 | 9.20E-17 | -0.02 | 0.008 | 0.011 | 65.79 |
| rs12967010 | T | C | 0.77 | 0.011 | 0.002 | 1.33E-11 | 0.003 | 0.008 | 0.681 | 44.97 |
| rs12981405 | T | C | 0.17 | -0.011 | 0.002 | 2.75E-09 | -0.021 | 0.009 | 0.015 | 34.75 |
| rs1301838 | T | C | 0.32 | -0.01 | 0.002 | 1.29E-10 | 0.009 | 0.007 | 0.201 | 40.96 |
| rs13018640 | T | C | 0.6 | -0.021 | 0.001 | 1.05E-50 | -0.013 | 0.007 | 0.05 | 235.84 |
| rs13050131 | A | C | 0.66 | -0.008 | 0.002 | 4.40E-08 | 0.013 | 0.007 | 0.067 | 29.16 |
| rs13133213 | A | G | 0.5 | 0.01 | 0.001 | 7.57E-12 | -0.003 | 0.006 | 0.599 | 47.02 |
| rs13145650 | T | C | 0.92 | -0.017 | 0.002 | 8.99E-12 | -0.021 | 0.011 | 0.063 | 47.89 |
| rs13163845 | T | C | 0.85 | -0.015 | 0.002 | 4.66E-14 | -0.016 | 0.009 | 0.079 | 55.5 |
| rs13197257 | T | G | 0.28 | 0.011 | 0.002 | 2.83E-12 | -0.003 | 0.007 | 0.674 | 46.41 |
| rs13212041 | T | C | 0.8 | 0.01 | 0.002 | 7.42E-09 | -0.008 | 0.008 | 0.29 | 31.48 |
| rs13240401 | T | C | 0.78 | 0.018 | 0.002 | 2.36E-24 | 0.009 | 0.008 | 0.268 | 107.18 |
| rs13261773 | C | G | 0.16 | 0.012 | 0.002 | 3.12E-10 | -0.019 | 0.009 | 0.033 | 41.23 |
| rs13266287 | A | T | 0.67 | -0.009 | 0.002 | 1.75E-08 | 0.005 | 0.007 | 0.475 | 32.11 |
| rs1329125 | T | C | 0.33 | -0.01 | 0.002 | 8.37E-11 | -0.006 | 0.007 | 0.396 | 41.82 |
| rs13327482 | A | G | 0.82 | -0.012 | 0.002 | 4.47E-11 | -0.002 | 0.008 | 0.774 | 45.19 |
| rs1334297 | A | G | 0.74 | 0.026 | 0.002 | 2.53E-58 | -0.001 | 0.007 | 0.915 | 258 |
| rs13422673 | T | C | 0.47 | -0.011 | 0.001 | 6.48E-15 | 0.015 | 0.006 | 0.018 | 61.73 |
| rs137079 | T | C | 0.14 | 0.013 | 0.002 | 3.06E-10 | -0.018 | 0.009 | 0.043 | 40.96 |
| rs1426619 | T | C | 0.43 | 0.01 | 0.001 | 1.40E-11 | 0.004 | 0.006 | 0.522 | 47.02 |
| rs1434630 | T | G | 0.15 | -0.014 | 0.002 | 4.63E-12 | 0.001 | 0.009 | 0.897 | 46.92 |
| rs143743568 | A | G | 0.15 | 0.012 | 0.002 | 4.99E-09 | -0.014 | 0.009 | 0.105 | 34.81 |
| rs150537577 | A | G | 0.92 | -0.015 | 0.003 | 9.50E-09 | -0.002 | 0.012 | 0.843 | 33.73 |
| rs1505676 | C | G | 0.63 | 0.008 | 0.002 | 6.67E-09 | -0.002 | 0.007 | 0.799 | 31.36 |
| rs1542354 | A | G | 0.48 | -0.008 | 0.001 | 2.72E-09 | 0.007 | 0.007 | 0.286 | 36 |
| rs1544 | A | G | 0.26 | -0.01 | 0.002 | 1.76E-09 | -0.005 | 0.007 | 0.475 | 36 |
| rs1564347 | T | G | 0.34 | 0.009 | 0.002 | 2.39E-09 | 0.007 | 0.007 | 0.323 | 34.42 |
| rs1603460 | T | G | 0.42 | 0.009 | 0.001 | 2.20E-10 | 0.009 | 0.007 | 0.151 | 41.33 |
| rs162445 | A | G | 0.08 | 0.015 | 0.003 | 5.72E-09 | -0.014 | 0.012 | 0.249 | 34.18 |
| rs1689510 | C | G | 0.33 | 0.019 | 0.002 | 1.35E-38 | -0.01 | 0.007 | 0.136 | 167.27 |
| rs17048855 | A | G | 0.34 | 0.011 | 0.002 | 3.10E-13 | -0.007 | 0.007 | 0.305 | 51.84 |
| rs17110109 | T | C | 0.62 | -0.009 | 0.001 | 1.47E-10 | -0.012 | 0.007 | 0.073 | 44.13 |
| rs17144467 | A | C | 0.69 | 0.01 | 0.002 | 8.74E-11 | 0.009 | 0.007 | 0.186 | 42.68 |
| rs17148998 | A | G | 0.2 | 0.01 | 0.002 | 6.98E-09 | -0.011 | 0.008 | 0.174 | 35.3 |
| rs1717204 | A | C | 0.18 | -0.012 | 0.002 | 1.20E-10 | 0.001 | 0.008 | 0.896 | 42.25 |
| rs17248751 | A | G | 0.78 | -0.014 | 0.002 | 1.28E-15 | 0.002 | 0.008 | 0.78 | 64 |
| rs173003 | A | C | 0.49 | -0.008 | 0.001 | 5.50E-09 | -0.031 | 0.006 | 1.78E-06 | 34.31 |
| rs1738050 | C | G | 0.62 | -0.009 | 0.001 | 1.42E-10 | -0.003 | 0.007 | 0.651 | 44.13 |
| rs17411339 | A | G | 0.56 | 0.014 | 0.001 | 5.10E-23 | -0.003 | 0.007 | 0.644 | 100 |
| rs17428076 | C | G | 0.76 | 0.014 | 0.002 | 6.41E-17 | -0.011 | 0.007 | 0.142 | 73.32 |
| rs1747714 | T | C | 0.53 | 0.011 | 0.001 | 3.82E-14 | 0.002 | 0.006 | 0.754 | 57.33 |
| rs1747817 | T | C | 0.76 | -0.009 | 0.002 | 2.29E-08 | -0.002 | 0.008 | 0.776 | 33.06 |
| rs17489649 | A | G | 0.68 | 0.013 | 0.002 | 2.77E-17 | -0.003 | 0.007 | 0.702 | 71.68 |
| rs17502934 | T | G | 0.15 | -0.017 | 0.002 | 2.12E-17 | 0.003 | 0.009 | 0.737 | 69.72 |
| rs175325 | A | T | 0.6 | -0.01 | 0.001 | 2.24E-12 | -0.006 | 0.007 | 0.368 | 51.02 |
| rs17551064 | A | G | 0.84 | 0.014 | 0.002 | 1.53E-13 | 0 | 0.009 | 0.988 | 55.07 |
| rs17563464 | A | C | 0.22 | -0.015 | 0.002 | 1.74E-17 | 0.001 | 0.008 | 0.906 | 74.77 |
| rs17565975 | A | G | 0.56 | -0.011 | 0.001 | 5.54E-14 | 0.012 | 0.007 | 0.059 | 57.33 |
| rs17598675 | T | C | 0.51 | -0.012 | 0.001 | 1.37E-16 | -0.002 | 0.006 | 0.809 | 68.65 |
| rs17604349 | A | G | 0.19 | 0.013 | 0.002 | 2.36E-12 | 0.018 | 0.008 | 0.034 | 49.78 |
| rs176218 | T | G | 0.2 | 0.02 | 0.002 | 1.10E-29 | 0.016 | 0.008 | 0.046 | 123.46 |
| rs17732878 | T | C | 0.79 | -0.01 | 0.002 | 4.06E-08 | -0.013 | 0.008 | 0.108 | 31.23 |
| rs178183 | T | C | 0.25 | 0.015 | 0.002 | 4.81E-19 | -0.004 | 0.007 | 0.549 | 82.13 |
| rs17882802 | A | G | 0.43 | 0.008 | 0.001 | 3.00E-09 | 0.007 | 0.006 | 0.302 | 36 |
| rs182902112 | A | C | 0.02 | -0.03 | 0.005 | 1.17E-08 | -0.058 | 0.027 | 0.035 | 32.04 |
| rs1842713 | A | G | 0.79 | -0.012 | 0.002 | 9.67E-12 | 0.008 | 0.008 | 0.297 | 47.37 |
| rs1861786 | A | G | 0.38 | -0.008 | 0.001 | 2.39E-08 | 0.009 | 0.007 | 0.192 | 33.47 |
| rs1865955 | T | C | 0.82 | 0.012 | 0.002 | 4.06E-11 | 0.019 | 0.008 | 0.028 | 41.23 |
| rs1866823 | A | G | 0.54 | 0.011 | 0.001 | 3.09E-14 | 0.006 | 0.007 | 0.356 | 58.41 |
| rs1890132 | T | C | 0.71 | -0.009 | 0.002 | 6.56E-09 | 0.003 | 0.007 | 0.649 | 35.2 |
| rs192436652 | T | C | 0.03 | -0.032 | 0.004 | 2.06E-12 | 0.002 | 0.02 | 0.937 | 50.25 |
| rs1933264 | T | C | 0.75 | 0.01 | 0.002 | 5.85E-10 | 0.019 | 0.007 | 0.012 | 39.06 |
| rs1955250 | A | C | 0.91 | -0.014 | 0.002 | 2.26E-08 | -0.015 | 0.012 | 0.211 | 31.36 |
| rs1991585 | T | C | 0.71 | -0.01 | 0.002 | 6.26E-11 | 0.003 | 0.007 | 0.654 | 45.34 |
| rs2007655 | T | G | 0.5 | 0.009 | 0.001 | 8.64E-10 | -0.008 | 0.006 | 0.19 | 37.73 |
| rs2011603 | A | G | 0.74 | -0.009 | 0.002 | 1.12E-08 | 0.013 | 0.007 | 0.074 | 32.35 |
| rs2034631 | T | C | 0.65 | 0.009 | 0.002 | 2.64E-09 | -0.004 | 0.007 | 0.563 | 35.2 |
| rs2052285 | A | G | 0.6 | 0.011 | 0.001 | 7.25E-15 | -0.006 | 0.007 | 0.358 | 64 |
| rs2055940 | A | G | 0.32 | 0.008 | 0.002 | 4.95E-08 | -0.008 | 0.007 | 0.224 | 29.88 |
| rs2081652 | A | T | 0.66 | 0.012 | 0.002 | 1.09E-16 | -0.024 | 0.007 | 4.19E-04 | 67.24 |
| rs2131167 | A | G | 0.45 | 0.008 | 0.001 | 1.52E-08 | 0.012 | 0.006 | 0.063 | 32.65 |
| rs2179152 | T | C | 0.37 | -0.013 | 0.002 | 1.68E-19 | -0.004 | 0.007 | 0.56 | 76.27 |
| rs2183271 | T | C | 0.64 | 0.008 | 0.002 | 1.91E-08 | -0.01 | 0.007 | 0.148 | 31.36 |
| rs2199409 | T | C | 0.8 | 0.01 | 0.002 | 3.14E-09 | -0.004 | 0.008 | 0.651 | 36.71 |
| rs2212430 | T | C | 0.3 | -0.01 | 0.002 | 1.32E-10 | -0.009 | 0.007 | 0.225 | 39.06 |
| rs2216144 | T | C | 0.53 | 0.01 | 0.001 | 2.58E-12 | 0.01 | 0.006 | 0.135 | 49 |
| rs2256965 | A | G | 0.42 | 0.011 | 0.001 | 2.40E-13 | -0.006 | 0.007 | 0.392 | 57.33 |
| rs2287838 | A | G | 0.55 | -0.01 | 0.001 | 1.27E-13 | 0 | 0.006 | 0.998 | 55.18 |
| rs2290601 | T | C | 0.77 | -0.015 | 0.002 | 4.49E-19 | 0.011 | 0.008 | 0.152 | 76.82 |
| rs2297293 | C | G | 0.31 | 0.01 | 0.002 | 8.80E-11 | 0.001 | 0.007 | 0.927 | 43.56 |
| rs232496 | T | C | 0.37 | 0.009 | 0.002 | 7.14E-10 | 0.003 | 0.007 | 0.609 | 36 |
| rs2364544 | A | G | 0.39 | -0.011 | 0.001 | 2.91E-13 | 0.01 | 0.007 | 0.146 | 56.25 |
| rs2434672 | A | C | 0.53 | 0.009 | 0.001 | 1.06E-10 | -0.004 | 0.007 | 0.536 | 42.25 |
| rs2436760 | T | C | 0.97 | 0.023 | 0.004 | 4.52E-08 | 0.022 | 0.019 | 0.243 | 30.51 |
| rs2447097 | T | G | 0.46 | 0.01 | 0.001 | 1.10E-11 | 0 | 0.007 | 0.964 | 47.02 |
| rs2469226 | A | T | 0.22 | 0.01 | 0.002 | 1.28E-08 | -0.005 | 0.008 | 0.533 | 31.89 |
| rs2496482 | T | C | 0.42 | 0.012 | 0.002 | 9.20E-17 | 0.01 | 0.007 | 0.124 | 65.07 |
| rs252991 | A | G | 0.37 | 0.01 | 0.002 | 6.30E-11 | -0.002 | 0.007 | 0.755 | 40.11 |
| rs2545795 | A | C | 0.44 | 0.013 | 0.001 | 1.03E-18 | -0.009 | 0.006 | 0.148 | 79.72 |
| rs2554835 | A | G | 0.4 | 0.009 | 0.001 | 8.95E-10 | 0.014 | 0.007 | 0.04 | 39.51 |
| rs2570497 | T | C | 0.64 | -0.012 | 0.002 | 1.27E-16 | -0.02 | 0.007 | 0.003 | 65.07 |
| rs2725370 | T | C | 0.3 | -0.014 | 0.002 | 4.96E-20 | -0.008 | 0.007 | 0.265 | 88.36 |
| rs2764684 | T | C | 0.83 | 0.014 | 0.002 | 9.99E-15 | -0.039 | 0.009 | 5.44E-06 | 56.65 |
| rs2838006 | T | C | 0.36 | 0.013 | 0.002 | 1.38E-19 | 0.006 | 0.007 | 0.383 | 77.44 |
| rs28513882 | A | G | 0.18 | -0.011 | 0.002 | 2.03E-09 | 0.004 | 0.008 | 0.637 | 37.35 |
| rs2885198 | A | G | 0.53 | 0.009 | 0.001 | 4.63E-10 | -0.01 | 0.007 | 0.133 | 39.51 |
| rs2923424 | A | G | 0.61 | 0.012 | 0.001 | 6.31E-17 | 0.004 | 0.007 | 0.564 | 73.47 |
| rs2929032 | A | G | 0.47 | -0.008 | 0.001 | 8.40E-09 | -0.005 | 0.006 | 0.459 | 33.47 |
| rs2958182 | A | T | 0.34 | 0.008 | 0.002 | 3.46E-08 | 0.012 | 0.007 | 0.076 | 29.88 |
| rs2964199 | T | C | 0.31 | -0.01 | 0.002 | 4.61E-11 | 0.001 | 0.007 | 0.897 | 43.56 |
| rs2964255 | A | G | 0.31 | 0.009 | 0.002 | 6.34E-09 | 0.02 | 0.007 | 0.004 | 34.42 |
| rs2998299 | T | G | 0.21 | -0.014 | 0.002 | 1.51E-15 | -0.007 | 0.008 | 0.37 | 64 |
| rs3026996 | A | C | 0.76 | 0.014 | 0.002 | 5.42E-17 | -0.007 | 0.007 | 0.362 | 74.39 |
| rs303752 | A | G | 0.41 | -0.011 | 0.001 | 1.47E-14 | 0.015 | 0.007 | 0.022 | 61.73 |
| rs3111251 | T | C | 0.43 | -0.009 | 0.001 | 3.88E-10 | -0.006 | 0.007 | 0.395 | 41.33 |
| rs34067381 | T | G | 0.36 | -0.01 | 0.002 | 9.77E-13 | 0.004 | 0.007 | 0.52 | 48.07 |
| rs34098770 | A | G | 0.15 | -0.016 | 0.002 | 2.95E-16 | 0.014 | 0.009 | 0.118 | 64 |
| rs34305371 | A | G | 0.1 | 0.031 | 0.002 | 1.10E-39 | 0.027 | 0.011 | 0.013 | 171.17 |
| rs34394051 | A | G | 0.84 | -0.012 | 0.002 | 2.30E-09 | -0.002 | 0.009 | 0.799 | 34.22 |
| rs34720381 | T | C | 0.09 | -0.018 | 0.002 | 1.90E-13 | -0.018 | 0.011 | 0.11 | 56.25 |
| rs34807077 | A | C | 0.16 | 0.013 | 0.002 | 3.29E-11 | -0.004 | 0.009 | 0.665 | 45.39 |
| rs35417702 | T | C | 0.52 | -0.015 | 0.001 | 2.40E-27 | -0.003 | 0.006 | 0.624 | 117.88 |
| rs35606437 | A | G | 0.27 | 0.011 | 0.002 | 3.98E-11 | -0.017 | 0.007 | 0.021 | 43.07 |
| rs35929923 | A | G | 0.25 | -0.011 | 0.002 | 1.77E-12 | 0.011 | 0.007 | 0.139 | 50.77 |
| rs363096 | T | C | 0.42 | -0.014 | 0.001 | 5.77E-24 | -0.007 | 0.007 | 0.264 | 104.33 |
| rs3751331 | A | G | 0.38 | -0.01 | 0.001 | 1.75E-11 | 0.002 | 0.007 | 0.8 | 48.01 |
| rs3809634 | A | G | 0.68 | -0.011 | 0.002 | 6.82E-13 | 0.017 | 0.007 | 0.018 | 52.8 |
| rs382196 | T | G | 0.38 | -0.008 | 0.001 | 1.47E-08 | -0.001 | 0.007 | 0.846 | 34.31 |
| rs3859523 | T | C | 0.82 | 0.012 | 0.002 | 9.92E-12 | -0.016 | 0.008 | 0.057 | 47.46 |
| rs3897821 | A | G | 0.67 | 0.015 | 0.002 | 3.58E-25 | 0.002 | 0.007 | 0.763 | 105.4 |
| rs3948495 | T | G | 0.41 | -0.009 | 0.001 | 3.95E-10 | 0.014 | 0.007 | 0.033 | 40.41 |
| rs401526 | T | C | 0.53 | -0.01 | 0.001 | 1.15E-13 | -0.02 | 0.006 | 0.002 | 55.18 |
| rs401966 | C | G | 0.39 | -0.01 | 0.001 | 1.72E-11 | 0.008 | 0.007 | 0.261 | 48.01 |
| rs42210 | C | G | 0.71 | -0.01 | 0.002 | 2.74E-10 | 0.01 | 0.007 | 0.181 | 37.52 |
| rs42302 | A | G | 0.34 | 0.009 | 0.002 | 5.96E-09 | -0.001 | 0.007 | 0.844 | 32.87 |
| rs4298514 | T | C | 0.7 | 0.01 | 0.002 | 1.07E-11 | -0.001 | 0.007 | 0.894 | 48.07 |
| rs4320563 | A | G | 0.51 | 0.011 | 0.001 | 2.24E-14 | 0.001 | 0.006 | 0.873 | 60.62 |
| rs4328757 | T | C | 0.61 | 0.01 | 0.001 | 2.95E-11 | 0.004 | 0.007 | 0.569 | 47.02 |
| rs4352658 | T | C | 0.08 | -0.019 | 0.003 | 1.71E-13 | 0.012 | 0.012 | 0.308 | 53.4 |
| rs4358081 | A | C | 0.53 | -0.01 | 0.001 | 1.28E-11 | 0.015 | 0.006 | 0.021 | 46.05 |
| rs4369924 | A | G | 0.16 | 0.013 | 0.002 | 5.58E-11 | 0.008 | 0.009 | 0.39 | 43.28 |
| rs4384309 | A | G | 0.46 | 0.01 | 0.001 | 4.89E-13 | -0.005 | 0.007 | 0.463 | 53.08 |
| rs4467547 | T | G | 0.41 | 0.012 | 0.001 | 4.88E-17 | 0.004 | 0.007 | 0.547 | 74.7 |
| rs4500930 | T | C | 0.34 | -0.011 | 0.002 | 1.67E-13 | 0.001 | 0.007 | 0.941 | 52.8 |
| rs4663617 | A | T | 0.23 | 0.01 | 0.002 | 4.12E-10 | 0.001 | 0.008 | 0.916 | 37.43 |
| rs4673840 | T | C | 0.84 | -0.014 | 0.002 | 5.86E-13 | 0.008 | 0.009 | 0.353 | 52.75 |
| rs4675248 | A | G | 0.4 | -0.009 | 0.001 | 4.50E-11 | -0.017 | 0.007 | 0.009 | 45.08 |
| rs4719944 | T | C | 0.55 | -0.012 | 0.001 | 9.25E-18 | 0.001 | 0.006 | 0.861 | 74.7 |
| rs4726070 | A | G | 0.6 | 0.012 | 0.001 | 1.36E-17 | -0.009 | 0.007 | 0.193 | 75.94 |
| rs4757957 | C | G | 0.69 | 0.013 | 0.002 | 1.79E-18 | -0.008 | 0.007 | 0.241 | 78.62 |
| rs4766424 | C | G | 0.12 | 0.014 | 0.002 | 2.47E-11 | 0.006 | 0.01 | 0.566 | 45.72 |
| rs4778058 | T | C | 0.5 | -0.009 | 0.001 | 4.92E-11 | 0.01 | 0.007 | 0.14 | 43.18 |
| rs4787457 | A | G | 0.63 | 0.016 | 0.002 | 1.23E-28 | 0.007 | 0.007 | 0.263 | 116.64 |
| rs4793090 | A | G | 0.66 | 0.008 | 0.002 | 1.67E-08 | 0.003 | 0.007 | 0.621 | 31.36 |
| rs4810894 | A | G | 0.37 | -0.009 | 0.002 | 4.38E-09 | 0.008 | 0.007 | 0.242 | 32.11 |
| rs4839155 | T | G | 0.77 | 0.012 | 0.002 | 4.66E-12 | 0.003 | 0.008 | 0.666 | 45.76 |
| rs4846724 | A | G | 0.53 | 0.01 | 0.001 | 2.89E-12 | 0 | 0.006 | 0.973 | 49 |
| rs4848924 | A | C | 0.71 | -0.012 | 0.002 | 3.56E-14 | -0.003 | 0.007 | 0.679 | 60.84 |
| rs488476 | C | G | 0.62 | -0.011 | 0.002 | 8.29E-15 | -0.021 | 0.007 | 0.002 | 56.75 |
| rs4895650 | T | C | 0.55 | 0.009 | 0.001 | 5.31E-11 | 0.013 | 0.007 | 0.044 | 44.13 |
| rs4899012 | C | G | 0.6 | -0.01 | 0.001 | 8.06E-13 | 0 | 0.007 | 0.942 | 54.13 |
| rs4904523 | A | G | 0.52 | -0.008 | 0.001 | 6.27E-09 | -0.002 | 0.006 | 0.787 | 34.31 |
| rs4915735 | A | G | 0.14 | 0.012 | 0.002 | 9.56E-09 | -0.008 | 0.009 | 0.377 | 33.64 |
| rs4977885 | A | G | 0.4 | -0.008 | 0.001 | 6.78E-09 | 0.006 | 0.007 | 0.368 | 36 |
| rs4984541 | A | G | 0.77 | -0.014 | 0.002 | 3.40E-16 | 0.001 | 0.008 | 0.925 | 65.9 |
| rs55736314 | C | G | 0.6 | -0.015 | 0.001 | 4.59E-27 | 0.025 | 0.007 | 9.66E-05 | 121 |
| rs56099375 | T | C | 0.24 | 0.012 | 0.002 | 3.10E-13 | 0.01 | 0.008 | 0.169 | 49.83 |
| rs56319902 | T | C | 0.22 | -0.021 | 0.002 | 5.81E-33 | 0.063 | 0.008 | 4.93E-16 | 146.84 |
| rs56391344 | A | G | 0.25 | 0.015 | 0.002 | 7.49E-20 | -0.014 | 0.007 | 0.068 | 85.56 |
| rs56794817 | A | G | 0.16 | 0.012 | 0.002 | 1.69E-10 | 0.007 | 0.009 | 0.409 | 41.23 |
| rs57349798 | A | G | 0.41 | 0.009 | 0.001 | 8.63E-11 | -0.001 | 0.007 | 0.825 | 44.13 |
| rs57352738 | A | T | 0.2 | -0.015 | 0.002 | 2.51E-18 | 0.001 | 0.008 | 0.924 | 79.94 |
| rs575113 | A | G | 0.29 | 0.013 | 0.002 | 7.52E-18 | -0.003 | 0.007 | 0.663 | 78.62 |
| rs5754753 | T | C | 0.72 | -0.013 | 0.002 | 6.15E-16 | -0.011 | 0.007 | 0.124 | 62.02 |
| rs57661533 | T | C | 0.13 | -0.011 | 0.002 | 3.24E-08 | 0.015 | 0.01 | 0.13 | 29.47 |
| rs580652 | T | C | 0.9 | -0.015 | 0.002 | 4.65E-10 | -0.003 | 0.011 | 0.768 | 37.52 |
| rs590013 | T | C | 0.68 | 0.01 | 0.002 | 2.47E-11 | -0.009 | 0.007 | 0.214 | 44.44 |
| rs6043521 | T | C | 0.6 | -0.008 | 0.001 | 9.72E-09 | 0.012 | 0.007 | 0.077 | 35.15 |
| rs6065080 | T | C | 0.36 | -0.013 | 0.002 | 7.49E-19 | 0.013 | 0.007 | 0.05 | 75.11 |
| rs6065784 | C | G | 0.7 | 0.011 | 0.002 | 5.78E-13 | -0.003 | 0.007 | 0.643 | 53.78 |
| rs61104616 | A | G | 0.52 | -0.017 | 0.001 | 1.56E-34 | -0.015 | 0.006 | 0.023 | 150.94 |
| rs6123924 | A | G | 0.84 | 0.013 | 0.002 | 3.20E-11 | 0.015 | 0.009 | 0.086 | 46.1 |
| rs61527214 | A | G | 0.41 | 0.011 | 0.001 | 1.13E-14 | 0.001 | 0.007 | 0.902 | 61.73 |
| rs61997667 | T | C | 0.15 | -0.014 | 0.002 | 2.24E-13 | 0.002 | 0.009 | 0.838 | 51.84 |
| rs62155350 | A | G | 0.02 | 0.034 | 0.005 | 1.69E-10 | -0.007 | 0.023 | 0.754 | 40.67 |
| rs62155873 | T | C | 0.12 | -0.015 | 0.002 | 1.65E-12 | 0.014 | 0.01 | 0.145 | 51.7 |
| rs62174974 | A | G | 0.19 | -0.011 | 0.002 | 2.76E-09 | 0.002 | 0.008 | 0.828 | 34.03 |
| rs62179650 | A | G | 0.3 | 0.011 | 0.002 | 5.66E-13 | 0.001 | 0.007 | 0.899 | 49 |
| rs62182994 | T | C | 0.69 | 0.014 | 0.002 | 2.55E-19 | 0.002 | 0.007 | 0.789 | 83.42 |
| rs62379838 | T | C | 0.7 | 0.012 | 0.002 | 1.77E-15 | -0.009 | 0.007 | 0.181 | 65.07 |
| rs628993 | A | G | 0.11 | 0.013 | 0.002 | 3.83E-08 | 0.009 | 0.011 | 0.422 | 30.97 |
| rs6449503 | A | G | 0.5 | 0.018 | 0.001 | 1.84E-39 | -0.006 | 0.007 | 0.379 | 174.62 |
| rs6490618 | T | C | 0.33 | 0.009 | 0.002 | 3.48E-09 | 0.015 | 0.007 | 0.033 | 34.42 |
| rs6493265 | T | C | 0.39 | -0.012 | 0.001 | 2.26E-17 | 0 | 0.007 | 0.963 | 75.94 |
| rs6557171 | T | C | 0.32 | -0.016 | 0.002 | 5.48E-25 | 0.001 | 0.007 | 0.845 | 106.78 |
| rs660001 | A | G | 0.21 | -0.016 | 0.002 | 1.66E-21 | -0.013 | 0.008 | 0.088 | 93.07 |
| rs66568921 | T | G | 0.64 | -0.018 | 0.002 | 7.60E-34 | -0.003 | 0.007 | 0.692 | 145.6 |
| rs66721975 | A | G | 0.29 | -0.009 | 0.002 | 1.51E-08 | -0.007 | 0.007 | 0.314 | 33.64 |
| rs6690195 | T | C | 0.49 | -0.011 | 0.001 | 7.89E-16 | -0.005 | 0.006 | 0.437 | 65.15 |
| rs6697584 | T | C | 0.79 | -0.013 | 0.002 | 1.43E-13 | 0.003 | 0.008 | 0.661 | 54.93 |
| rs6704768 | A | G | 0.56 | -0.012 | 0.001 | 1.84E-16 | -0.001 | 0.006 | 0.933 | 68.65 |
| rs6731373 | A | G | 0.34 | -0.012 | 0.002 | 1.39E-14 | 0.004 | 0.007 | 0.536 | 58.78 |
| rs6812533 | T | C | 0.74 | 0.011 | 0.002 | 4.85E-11 | -0.009 | 0.007 | 0.213 | 43.89 |
| rs68145588 | T | G | 0.14 | -0.013 | 0.002 | 7.53E-10 | -0.01 | 0.009 | 0.293 | 36.57 |
| rs6917154 | T | C | 0.12 | 0.013 | 0.002 | 2.55E-09 | 0.003 | 0.01 | 0.795 | 36.57 |
| rs6924023 | A | G | 0.75 | 0.011 | 0.002 | 5.17E-11 | 0.009 | 0.008 | 0.253 | 44.72 |
| rs6977237 | T | C | 0.47 | 0.008 | 0.001 | 5.59E-09 | 0.002 | 0.006 | 0.705 | 34.31 |
| rs6994287 | A | G | 0.41 | 0.01 | 0.001 | 1.11E-12 | -0.002 | 0.007 | 0.708 | 53.08 |
| rs7012546 | T | C | 0.42 | 0.009 | 0.001 | 1.25E-10 | 0.002 | 0.007 | 0.788 | 43.18 |
| rs7029718 | A | G | 0.41 | 0.024 | 0.001 | 7.82E-65 | 0 | 0.007 | 0.947 | 303.76 |
| rs7041702 | A | G | 0.74 | -0.011 | 0.002 | 4.53E-12 | -0.015 | 0.007 | 0.037 | 48.13 |
| rs7167688 | T | C | 0.49 | -0.009 | 0.001 | 1.71E-11 | 0.002 | 0.006 | 0.717 | 45.08 |
| rs7171405 | A | G | 0.24 | -0.01 | 0.002 | 7.53E-10 | -0.007 | 0.008 | 0.334 | 39.85 |
| rs717996 | T | C | 0.6 | -0.012 | 0.001 | 1.89E-17 | -0.003 | 0.007 | 0.606 | 75.94 |
| rs7226824 | T | C | 0.53 | -0.008 | 0.001 | 2.72E-08 | -0.01 | 0.006 | 0.128 | 31.04 |
| rs72486027 | T | C | 0.25 | -0.01 | 0.002 | 1.61E-10 | 0.007 | 0.008 | 0.37 | 42.25 |
| rs7254263 | T | C | 0.28 | -0.012 | 0.002 | 1.84E-14 | 0.005 | 0.007 | 0.51 | 55.32 |
| rs72622559 | T | C | 0.23 | -0.012 | 0.002 | 5.50E-13 | -0.011 | 0.008 | 0.159 | 49.83 |
| rs72672052 | A | T | 0.18 | -0.01 | 0.002 | 4.87E-08 | 0.004 | 0.008 | 0.677 | 31.48 |
| rs72709560 | A | G | 0.69 | 0.009 | 0.002 | 2.03E-08 | 0.005 | 0.007 | 0.486 | 32.11 |
| rs72828517 | T | C | 0.83 | -0.017 | 0.002 | 1.38E-19 | -0.005 | 0.008 | 0.546 | 78.18 |
| rs72829857 | A | G | 0.76 | -0.015 | 0.002 | 1.52E-19 | -0.006 | 0.008 | 0.444 | 86.72 |
| rs72917504 | T | C | 0.06 | -0.018 | 0.003 | 2.91E-09 | -0.011 | 0.014 | 0.435 | 35.2 |
| rs72944064 | T | C | 0.74 | -0.01 | 0.002 | 9.39E-10 | 0.013 | 0.007 | 0.074 | 37.52 |
| rs72962169 | T | C | 0.16 | -0.017 | 0.002 | 1.26E-19 | -0.02 | 0.009 | 0.027 | 83.87 |
| rs72993796 | T | C | 0.88 | 0.013 | 0.002 | 1.12E-08 | 0.01 | 0.01 | 0.331 | 33.32 |
| rs730384 | A | G | 0.44 | 0.01 | 0.001 | 9.38E-13 | -0.002 | 0.007 | 0.705 | 52.05 |
| rs73055556 | A | G | 0.14 | 0.011 | 0.002 | 4.95E-08 | -0.008 | 0.009 | 0.405 | 30.8 |
| rs73191311 | A | G | 0.34 | -0.009 | 0.002 | 2.03E-10 | -0.001 | 0.007 | 0.88 | 39.27 |
| rs7321274 | A | G | 0.8 | 0.011 | 0.002 | 8.21E-11 | -0.01 | 0.008 | 0.196 | 40.11 |
| rs7326331 | A | G | 0.28 | -0.013 | 0.002 | 3.42E-16 | -0.004 | 0.007 | 0.573 | 64 |
| rs73344830 | A | G | 0.42 | 0.017 | 0.001 | 1.00E-32 | 0.006 | 0.007 | 0.346 | 147.45 |
| rs736281 | T | C | 0.39 | 0.009 | 0.001 | 2.49E-09 | 0.006 | 0.007 | 0.389 | 37.73 |
| rs73648455 | T | C | 0.08 | -0.018 | 0.003 | 1.99E-12 | -0.002 | 0.012 | 0.859 | 49.54 |
| rs743316 | T | C | 0.79 | 0.011 | 0.002 | 8.21E-11 | -0.014 | 0.008 | 0.073 | 43.4 |
| rs74415461 | T | C | 0.08 | 0.018 | 0.003 | 6.25E-12 | 0.021 | 0.011 | 0.066 | 45.82 |
| rs7449561 | A | G | 0.23 | 0.01 | 0.002 | 9.73E-10 | -0.004 | 0.008 | 0.623 | 36 |
| rs7460106 | T | C | 0.76 | -0.012 | 0.002 | 2.06E-13 | 0.001 | 0.008 | 0.919 | 52.35 |
| rs746839 | C | G | 0.63 | 0.014 | 0.002 | 1.19E-20 | 0.002 | 0.007 | 0.764 | 81 |
| rs74787922 | A | C | 0.93 | 0.018 | 0.003 | 8.97E-11 | 0.017 | 0.013 | 0.18 | 42.25 |
| rs75033012 | C | G | 0.04 | -0.026 | 0.004 | 1.38E-11 | 0.015 | 0.016 | 0.358 | 46.1 |
| rs75177132 | T | C | 0.05 | 0.03 | 0.004 | 1.01E-16 | -0.001 | 0.015 | 0.97 | 68.52 |
| rs7575637 | A | G | 0.45 | 0.011 | 0.001 | 3.43E-14 | 0.005 | 0.006 | 0.396 | 58.41 |
| rs7603132 | A | G | 0.19 | 0.014 | 0.002 | 1.20E-14 | -0.005 | 0.008 | 0.568 | 57.93 |
| rs76076331 | T | C | 0.13 | 0.021 | 0.002 | 1.71E-23 | -0.014 | 0.01 | 0.153 | 96.23 |
| rs7617204 | A | G | 0.52 | -0.01 | 0.001 | 4.33E-12 | 0.007 | 0.006 | 0.266 | 48.01 |
| rs76235882 | A | G | 0.97 | 0.024 | 0.004 | 4.52E-10 | -0.015 | 0.02 | 0.444 | 38.19 |
| rs76246107 | A | G | 0.09 | -0.016 | 0.003 | 6.52E-10 | 0.003 | 0.012 | 0.83 | 38.82 |
| rs7650602 | T | C | 0.56 | -0.009 | 0.001 | 2.77E-10 | 0.013 | 0.007 | 0.044 | 40.41 |
| rs76577427 | C | G | 0.9 | 0.02 | 0.002 | 7.78E-18 | 0.004 | 0.011 | 0.671 | 71.54 |
| rs7672622 | A | G | 0.75 | -0.009 | 0.002 | 4.72E-08 | 0.003 | 0.007 | 0.663 | 30.25 |
| rs7683416 | T | C | 0.46 | 0.014 | 0.001 | 6.05E-23 | -0.001 | 0.006 | 0.899 | 98.58 |
| rs76878669 | C | G | 0.76 | 0.014 | 0.002 | 3.10E-17 | 0.002 | 0.008 | 0.8 | 68.79 |
| rs7692359 | T | C | 0.78 | -0.011 | 0.002 | 1.40E-10 | -0.022 | 0.008 | 0.004 | 41.11 |
| rs77025239 | A | G | 0.16 | -0.014 | 0.002 | 4.65E-13 | -0.013 | 0.009 | 0.139 | 54.29 |
| rs77609760 | A | G | 0.93 | 0.024 | 0.003 | 7.71E-17 | -0.013 | 0.013 | 0.316 | 71.04 |
| rs77719387 | A | T | 0.02 | -0.04 | 0.006 | 3.78E-12 | -0.015 | 0.027 | 0.585 | 48.28 |
| rs7849487 | T | G | 0.66 | -0.017 | 0.002 | 1.25E-29 | 0.007 | 0.007 | 0.285 | 123.95 |
| rs78648104 | T | C | 0.91 | -0.014 | 0.002 | 1.70E-08 | -0.01 | 0.011 | 0.388 | 31.81 |
| rs7875078 | A | C | 0.46 | -0.008 | 0.001 | 2.72E-08 | 0.001 | 0.007 | 0.848 | 31.04 |
| rs7894722 | T | C | 0.37 | 0.009 | 0.001 | 5.41E-09 | 0.004 | 0.007 | 0.527 | 36.86 |
| rs790647 | A | C | 0.23 | -0.015 | 0.002 | 3.81E-18 | -0.002 | 0.008 | 0.809 | 72.75 |
| rs7910403 | T | G | 0.2 | 0.012 | 0.002 | 5.08E-12 | -0.013 | 0.008 | 0.109 | 45.94 |
| rs7924036 | T | G | 0.51 | 0.013 | 0.001 | 2.55E-21 | -0.026 | 0.006 | 3.97E-05 | 90.25 |
| rs79265434 | A | G | 0.88 | -0.02 | 0.002 | 1.98E-19 | 0.018 | 0.01 | 0.07 | 80.18 |
| rs7972246 | T | C | 0.35 | 0.011 | 0.002 | 1.01E-13 | 0.017 | 0.007 | 0.012 | 53.78 |
| rs79728014 | A | G | 0.86 | -0.013 | 0.002 | 2.06E-11 | 0 | 0.009 | 0.988 | 44.89 |
| rs7974852 | A | C | 0.52 | 0.012 | 0.001 | 1.14E-16 | -0.007 | 0.006 | 0.286 | 69.84 |
| rs7977614 | A | G | 0.71 | -0.013 | 0.002 | 6.84E-17 | 0.007 | 0.007 | 0.3 | 70.14 |
| rs79994730 | T | C | 0.04 | 0.024 | 0.004 | 5.88E-12 | 0.016 | 0.015 | 0.273 | 47.02 |
| rs8008382 | T | C | 0.3 | -0.011 | 0.002 | 4.07E-12 | -0.013 | 0.007 | 0.065 | 49.94 |
| rs8024 | A | C | 0.33 | -0.012 | 0.002 | 1.13E-14 | -0.001 | 0.007 | 0.856 | 58.78 |
| rs80257979 | T | G | 0.97 | -0.025 | 0.004 | 3.31E-10 | 0.026 | 0.019 | 0.167 | 39.69 |
| rs8030487 | A | G | 0.69 | 0.009 | 0.002 | 2.52E-08 | 0.01 | 0.007 | 0.147 | 32.11 |
| rs806816 | A | T | 0.75 | 0.01 | 0.002 | 4.80E-10 | -0.022 | 0.007 | 0.003 | 39.85 |
| rs8097125 | T | C | 0.38 | 0.009 | 0.001 | 2.31E-09 | 0 | 0.007 | 0.97 | 37.73 |
| rs912883 | T | C | 0.68 | 0.009 | 0.002 | 6.20E-09 | 0.007 | 0.007 | 0.288 | 33.64 |
| rs9289300 | T | C | 0.84 | -0.016 | 0.002 | 1.00E-17 | 0.004 | 0.009 | 0.661 | 74.5 |
| rs929511 | T | C | 0.13 | -0.016 | 0.002 | 3.91E-14 | -0.008 | 0.01 | 0.403 | 58.05 |
| rs9359939 | A | C | 0.24 | -0.011 | 0.002 | 1.51E-10 | -0.005 | 0.008 | 0.48 | 43.07 |
| rs936496 | A | G | 0.62 | -0.008 | 0.001 | 4.38E-08 | -0.012 | 0.007 | 0.07 | 31.84 |
| rs9373363 | A | G | 0.75 | -0.011 | 0.002 | 5.54E-12 | 0.003 | 0.007 | 0.736 | 48.13 |
| rs9375188 | T | C | 0.48 | 0.021 | 0.001 | 8.78E-52 | 0.002 | 0.006 | 0.711 | 229.31 |
| rs9411331 | A | C | 0.68 | 0.015 | 0.002 | 7.63E-22 | 0.011 | 0.007 | 0.106 | 93.44 |
| rs9492774 | C | G | 0.37 | 0.008 | 0.002 | 4.33E-08 | -0.004 | 0.007 | 0.527 | 28.44 |
| rs9529146 | T | C | 0.23 | 0.013 | 0.002 | 3.48E-14 | -0.005 | 0.008 | 0.484 | 55.81 |
| rs9556958 | T | C | 0.52 | -0.011 | 0.001 | 5.96E-15 | -0.003 | 0.006 | 0.606 | 61.73 |
| rs9616947 | T | C | 0.38 | -0.013 | 0.002 | 3.58E-20 | 0 | 0.007 | 0.973 | 79.8 |
| rs9633970 | T | C | 0.76 | 0.009 | 0.002 | 2.81E-08 | -0.002 | 0.008 | 0.771 | 32.35 |
| rs9655780 | A | G | 0.83 | -0.015 | 0.002 | 3.70E-16 | -0.005 | 0.009 | 0.583 | 64.84 |
| rs969512 | A | T | 0.66 | -0.011 | 0.002 | 9.84E-13 | -0.007 | 0.007 | 0.327 | 49.94 |
| rs9853928 | T | C | 0.21 | -0.013 | 0.002 | 1.59E-13 | -0.01 | 0.008 | 0.205 | 55.81 |
| rs9859556 | T | G | 0.31 | 0.029 | 0.002 | 4.61E-82 | 0.011 | 0.007 | 0.123 | 373.78 |
| rs9882532 | T | C | 0.64 | 0.012 | 0.002 | 2.15E-17 | 0 | 0.007 | 0.997 | 68.34 |
| rs9886703 | A | T | 0.17 | -0.014 | 0.002 | 4.46E-13 | 0 | 0.009 | 0.976 | 51.24 |
| rs9927137 | A | G | 0.53 | 0.008 | 0.001 | 5.11E-09 | -0.004 | 0.006 | 0.556 | 34.31 |
| rs9927842 | T | C | 0.15 | -0.013 | 0.002 | 6.22E-11 | 0.016 | 0.009 | 0.069 | 42.25 |
| rs9929556 | T | G | 0.57 | -0.01 | 0.001 | 2.31E-12 | 0.004 | 0.007 | 0.573 | 50.01 |
| rs9929762 | A | G | 0.56 | 0.01 | 0.001 | 1.03E-11 | 0.004 | 0.006 | 0.589 | 47.02 |
| rs9933256 | A | G | 0.56 | 0.012 | 0.001 | 6.06E-17 | -0.002 | 0.006 | 0.714 | 72.25 |

**SNPs used as instrumental variables in the MR analysis of educational attainment on local gyrification index**

| **SNP** | **Effect allele** | **Other allele** | **Effect allele frequency** | **beta (exposure)** | **SE (exposure)** | ***P* (exposure)** | **beta (outcome)** | **SE (outcome)** | ***P* (outcome)** | ***F*-statistic** |
| --- | --- | --- | --- | --- | --- | --- | --- | --- | --- | --- |
| rs10006235 | T | C | 0.27 | -0.009 | 0.002 | 4.48E-09 | -0.012 | 0.008 | 0.161 | 33.79 |
| rs10021733 | T | C | 0.15 | -0.016 | 0.003 | 9.45E-10 | 0.011 | 0.01 | 0.268 | 36.89 |
| rs10046069 | A | G | 0.12 | 0.013 | 0.002 | 4.40E-09 | 0.001 | 0.012 | 0.958 | 33.32 |
| rs10060023 | T | C | 0.33 | 0.011 | 0.002 | 1.88E-13 | 0.001 | 0.008 | 0.906 | 53.78 |
| rs10080647 | A | C | 0.14 | 0.012 | 0.002 | 7.05E-10 | -0.001 | 0.011 | 0.944 | 38.44 |
| rs1008078 | T | C | 0.4 | -0.018 | 0.001 | 4.50E-37 | 0.001 | 0.008 | 0.872 | 169 |
| rs10189857 | A | G | 0.57 | 0.016 | 0.001 | 4.69E-29 | 0.011 | 0.008 | 0.145 | 127.37 |
| rs10193498 | A | T | 0.76 | 0.01 | 0.002 | 3.10E-09 | 0.005 | 0.009 | 0.565 | 36.75 |
| rs10215082 | A | G | 0.47 | -0.015 | 0.001 | 7.03E-26 | -0.023 | 0.007 | 0.002 | 113.27 |
| rs10460095 | A | G | 0.57 | -0.012 | 0.001 | 2.61E-18 | -0.007 | 0.008 | 0.378 | 78.45 |
| rs10761251 | A | T | 0.66 | 0.011 | 0.002 | 2.52E-14 | 0.006 | 0.008 | 0.422 | 56.75 |
| rs10772644 | C | G | 0.89 | 0.014 | 0.002 | 1.69E-10 | -0.015 | 0.012 | 0.232 | 41.66 |
| rs10773002 | A | T | 0.25 | 0.018 | 0.002 | 2.50E-30 | 0.028 | 0.009 | 0.001 | 133.69 |
| rs10773208 | T | C | 0.25 | -0.011 | 0.002 | 8.09E-12 | 0.013 | 0.009 | 0.128 | 47.27 |
| rs10798888 | T | G | 0.17 | -0.014 | 0.002 | 5.15E-14 | -0.023 | 0.01 | 0.018 | 54.29 |
| rs10799615 | A | G | 0.25 | -0.009 | 0.002 | 4.11E-08 | -0.016 | 0.009 | 0.069 | 30.94 |
| rs10805383 | A | G | 0.49 | -0.01 | 0.001 | 2.22E-13 | -0.007 | 0.008 | 0.379 | 54.13 |
| rs10830858 | T | C | 0.51 | 0.009 | 0.001 | 5.31E-11 | 0.005 | 0.008 | 0.512 | 43.18 |
| rs10844179 | A | G | 0.23 | 0.01 | 0.002 | 5.03E-10 | -0.001 | 0.009 | 0.893 | 37.43 |
| rs10875121 | C | G | 0.84 | 0.017 | 0.002 | 4.46E-20 | 0.012 | 0.01 | 0.232 | 83.87 |
| rs10892807 | T | C | 0.57 | 0.011 | 0.001 | 1.09E-14 | 0.012 | 0.008 | 0.109 | 60.62 |
| rs10940921 | T | G | 0.42 | 0.011 | 0.001 | 1.31E-14 | -0.008 | 0.008 | 0.307 | 62.86 |
| rs10951590 | T | C | 0.33 | -0.011 | 0.002 | 3.62E-13 | -0.004 | 0.008 | 0.626 | 52.8 |
| rs10979613 | T | C | 0.64 | -0.012 | 0.002 | 1.25E-15 | 0.006 | 0.008 | 0.475 | 60.84 |
| rs10996167 | C | G | 0.65 | 0.011 | 0.002 | 7.47E-09 | 0.013 | 0.008 | 0.102 | 31.92 |
| rs11003463 | T | G | 0.61 | 0.009 | 0.002 | 5.58E-10 | -0.004 | 0.008 | 0.608 | 36.8 |
| rs11023764 | A | G | 0.64 | 0.009 | 0.002 | 5.32E-10 | -0.008 | 0.008 | 0.306 | 36.8 |
| rs11030102 | C | G | 0.75 | 0.01 | 0.002 | 4.60E-10 | 0.005 | 0.009 | 0.544 | 39.85 |
| rs11076962 | T | C | 0.72 | 0.009 | 0.002 | 4.29E-08 | 0.002 | 0.008 | 0.785 | 28.89 |
| rs11081529 | T | C | 0.71 | 0.013 | 0.002 | 5.22E-16 | 0.004 | 0.008 | 0.617 | 70.56 |
| rs11082011 | T | C | 0.67 | 0.019 | 0.002 | 2.06E-38 | -0.003 | 0.008 | 0.718 | 167.27 |
| rs11121177 | A | G | 0.18 | 0.015 | 0.002 | 1.44E-16 | -0.012 | 0.01 | 0.231 | 70.37 |
| rs11130380 | T | G | 0.61 | 0.009 | 0.001 | 4.17E-10 | 0.016 | 0.008 | 0.033 | 41.33 |
| rs11138947 | T | C | 0.72 | 0.011 | 0.002 | 2.36E-11 | -0.004 | 0.008 | 0.609 | 43.07 |
| rs11157931 | A | C | 0.39 | -0.014 | 0.001 | 3.86E-22 | -0.015 | 0.008 | 0.053 | 98.58 |
| rs111852224 | T | C | 0.12 | 0.016 | 0.002 | 4.85E-13 | 0 | 0.011 | 0.993 | 50.28 |
| rs11211123 | A | G | 0.22 | 0.01 | 0.002 | 7.62E-10 | -0.002 | 0.009 | 0.781 | 37.43 |
| rs11213482 | A | G | 0.84 | -0.011 | 0.002 | 2.68E-08 | 0.017 | 0.01 | 0.094 | 31.12 |
| rs112603734 | A | C | 0.77 | -0.013 | 0.002 | 2.72E-09 | 0.007 | 0.009 | 0.419 | 35.46 |
| rs112806496 | C | G | 0.91 | -0.016 | 0.002 | 1.72E-10 | -0.02 | 0.013 | 0.128 | 40.96 |
| rs1128956 | T | G | 0.83 | -0.014 | 0.002 | 5.34E-14 | -0.003 | 0.01 | 0.788 | 54.29 |
| rs113520408 | A | G | 0.28 | 0.016 | 0.002 | 1.03E-22 | -0.002 | 0.008 | 0.782 | 93.85 |
| rs113615161 | T | C | 0.13 | -0.012 | 0.002 | 2.98E-09 | -0.002 | 0.011 | 0.836 | 34.31 |
| rs114593137 | A | T | 0.79 | -0.012 | 0.002 | 1.65E-12 | -0.004 | 0.009 | 0.627 | 51.5 |
| rs115000530 | A | T | 0.95 | -0.025 | 0.003 | 1.09E-15 | -0.025 | 0.017 | 0.13 | 65.04 |
| rs11542663 | A | C | 0.69 | 0.011 | 0.002 | 2.44E-13 | -0.008 | 0.008 | 0.363 | 55.75 |
| rs11588857 | A | G | 0.21 | 0.02 | 0.002 | 4.45E-31 | 0.016 | 0.009 | 0.09 | 137.03 |
| rs11598765 | A | G | 0.2 | -0.011 | 0.002 | 1.73E-09 | 0.006 | 0.01 | 0.52 | 34.68 |
| rs11620355 | A | G | 0.09 | 0.018 | 0.002 | 1.09E-13 | -0.01 | 0.013 | 0.45 | 53 |
| rs11657342 | A | G | 0.37 | 0.014 | 0.002 | 2.32E-13 | 0.009 | 0.008 | 0.235 | 54.29 |
| rs11657979 | A | G | 0.24 | -0.009 | 0.002 | 4.57E-08 | -0.002 | 0.009 | 0.813 | 28.65 |
| rs11663602 | A | C | 0.28 | -0.013 | 0.002 | 9.27E-17 | -0.009 | 0.008 | 0.26 | 66.02 |
| rs1167827 | A | G | 0.43 | 0.01 | 0.001 | 2.93E-13 | -0.021 | 0.008 | 0.005 | 55.18 |
| rs11724690 | T | G | 0.29 | 0.009 | 0.002 | 2.08E-09 | 0.004 | 0.008 | 0.613 | 38.44 |
| rs117398064 | C | G | 0.09 | -0.014 | 0.002 | 1.16E-08 | -0.002 | 0.013 | 0.85 | 31.81 |
| rs118134876 | T | C | 0.06 | -0.022 | 0.003 | 1.50E-13 | 0.003 | 0.016 | 0.858 | 55.25 |
| rs11871429 | A | G | 0.77 | 0.013 | 0.002 | 2.52E-14 | -0.003 | 0.009 | 0.721 | 56.69 |
| rs11894424 | A | C | 0.95 | 0.021 | 0.003 | 2.14E-10 | 0.015 | 0.018 | 0.399 | 40.11 |
| rs12054166 | C | G | 0.74 | 0.01 | 0.002 | 2.30E-09 | 0.012 | 0.009 | 0.174 | 36 |
| rs12076635 | C | G | 0.78 | 0.021 | 0.002 | 3.33E-34 | 0.02 | 0.009 | 0.025 | 148.27 |
| rs12113634 | T | C | 0.65 | 0.009 | 0.002 | 2.50E-09 | 0.022 | 0.008 | 0.006 | 34.42 |
| rs12151248 | T | C | 0.11 | -0.015 | 0.002 | 3.81E-11 | 0.022 | 0.012 | 0.065 | 41.97 |
| rs1220779 | A | G | 0.47 | -0.008 | 0.001 | 1.58E-08 | 0.009 | 0.007 | 0.207 | 31.84 |
| rs12273435 | A | G | 0.21 | -0.013 | 0.002 | 4.72E-14 | -0.007 | 0.009 | 0.488 | 60.29 |
| rs12332731 | A | T | 0.19 | 0.014 | 0.002 | 5.96E-14 | 0.006 | 0.009 | 0.517 | 56.25 |
| rs12375949 | T | C | 0.43 | -0.014 | 0.001 | 1.61E-22 | 0.008 | 0.008 | 0.268 | 97.16 |
| rs12438177 | A | G | 0.37 | 0.009 | 0.002 | 1.48E-10 | 0.01 | 0.008 | 0.197 | 38.44 |
| rs12468040 | T | G | 0.38 | 0.014 | 0.001 | 3.12E-21 | -0.014 | 0.008 | 0.076 | 95.76 |
| rs12503522 | T | C | 0.28 | -0.011 | 0.002 | 3.93E-13 | -0.003 | 0.008 | 0.719 | 49.88 |
| rs12506221 | T | G | 0.44 | -0.013 | 0.001 | 4.23E-20 | -0.002 | 0.008 | 0.835 | 86.22 |
| rs12519073 | T | C | 0.23 | -0.011 | 0.002 | 1.16E-10 | 0.009 | 0.009 | 0.34 | 39.62 |
| rs12524795 | T | C | 0.44 | 0.01 | 0.001 | 5.28E-13 | -0.015 | 0.008 | 0.051 | 53.08 |
| rs12571549 | A | G | 0.14 | 0.015 | 0.002 | 1.45E-13 | 0.024 | 0.011 | 0.025 | 57 |
| rs12574281 | A | C | 0.63 | -0.008 | 0.002 | 3.04E-08 | 0.007 | 0.008 | 0.349 | 28.44 |
| rs12591647 | T | C | 0.82 | -0.015 | 0.002 | 7.78E-17 | -0.023 | 0.01 | 0.017 | 71.31 |
| rs12602286 | T | G | 0.87 | 0.016 | 0.002 | 4.62E-14 | -0.008 | 0.011 | 0.504 | 58.05 |
| rs12614263 | A | G | 0.5 | 0.008 | 0.001 | 3.38E-09 | 0.015 | 0.008 | 0.044 | 35.15 |
| rs12638072 | A | G | 0.3 | -0.011 | 0.002 | 3.13E-12 | -0.008 | 0.008 | 0.319 | 50.88 |
| rs12643771 | T | C | 0.31 | 0.013 | 0.002 | 1.32E-17 | 0.006 | 0.008 | 0.438 | 75.11 |
| rs12646523 | T | C | 0.25 | -0.013 | 0.002 | 3.79E-16 | -0.006 | 0.009 | 0.524 | 68.06 |
| rs12670376 | A | G | 0.45 | 0.009 | 0.001 | 5.99E-11 | -0.002 | 0.008 | 0.777 | 43.18 |
| rs12761761 | T | C | 0.24 | 0.015 | 0.002 | 4.79E-20 | -0.004 | 0.009 | 0.65 | 81 |
| rs12765185 | A | T | 0.28 | -0.009 | 0.002 | 2.96E-08 | -0.017 | 0.008 | 0.044 | 30.25 |
| rs12789313 | T | C | 0.49 | 0.009 | 0.001 | 6.79E-11 | 0.008 | 0.007 | 0.3 | 43.18 |
| rs12875339 | A | C | 0.64 | -0.011 | 0.002 | 1.71E-13 | -0.001 | 0.008 | 0.854 | 51.84 |
| rs12888615 | T | C | 0.2 | 0.012 | 0.002 | 9.42E-12 | -0.01 | 0.009 | 0.296 | 44.44 |
| rs12957463 | A | G | 0.8 | 0.015 | 0.002 | 9.20E-17 | 0.016 | 0.009 | 0.094 | 65.79 |
| rs12967010 | T | C | 0.77 | 0.011 | 0.002 | 1.33E-11 | -0.004 | 0.009 | 0.685 | 44.97 |
| rs12981405 | T | C | 0.17 | -0.011 | 0.002 | 2.75E-09 | 0.009 | 0.01 | 0.367 | 34.75 |
| rs1301838 | T | C | 0.32 | -0.01 | 0.002 | 1.29E-10 | -0.004 | 0.008 | 0.603 | 40.96 |
| rs13018640 | T | C | 0.6 | -0.021 | 0.001 | 1.05E-50 | 0.014 | 0.008 | 0.068 | 235.84 |
| rs13050131 | A | C | 0.66 | -0.008 | 0.002 | 4.40E-08 | 0.01 | 0.008 | 0.204 | 29.16 |
| rs13133213 | A | G | 0.5 | 0.01 | 0.001 | 7.57E-12 | 0.001 | 0.007 | 0.9 | 47.02 |
| rs13145650 | T | C | 0.92 | -0.017 | 0.002 | 8.99E-12 | -0.002 | 0.013 | 0.86 | 47.89 |
| rs13163845 | T | C | 0.85 | -0.015 | 0.002 | 4.66E-14 | 0.009 | 0.011 | 0.419 | 55.5 |
| rs13197257 | T | G | 0.28 | 0.011 | 0.002 | 2.83E-12 | -0.008 | 0.008 | 0.329 | 46.41 |
| rs13212041 | T | C | 0.8 | 0.01 | 0.002 | 7.42E-09 | -0.001 | 0.009 | 0.951 | 31.48 |
| rs13240401 | T | C | 0.78 | 0.018 | 0.002 | 2.36E-24 | 0.009 | 0.009 | 0.314 | 107.18 |
| rs13261773 | C | G | 0.16 | 0.012 | 0.002 | 3.12E-10 | 0.014 | 0.01 | 0.179 | 41.23 |
| rs13266287 | A | T | 0.67 | -0.009 | 0.002 | 1.75E-08 | -0.026 | 0.008 | 0.001 | 32.11 |
| rs1329125 | T | C | 0.33 | -0.01 | 0.002 | 8.37E-11 | -0.008 | 0.008 | 0.314 | 41.82 |
| rs13327482 | A | G | 0.82 | -0.012 | 0.002 | 4.47E-11 | 0.023 | 0.01 | 0.018 | 45.19 |
| rs1334297 | A | G | 0.74 | 0.026 | 0.002 | 2.53E-58 | 0.001 | 0.009 | 0.867 | 258 |
| rs13422673 | T | C | 0.47 | -0.011 | 0.001 | 6.48E-15 | -0.002 | 0.008 | 0.753 | 61.73 |
| rs137079 | T | C | 0.14 | 0.013 | 0.002 | 3.06E-10 | 0.013 | 0.011 | 0.226 | 40.96 |
| rs1426619 | T | C | 0.43 | 0.01 | 0.001 | 1.40E-11 | 0.019 | 0.008 | 0.013 | 47.02 |
| rs1434630 | T | G | 0.15 | -0.014 | 0.002 | 4.63E-12 | -0.014 | 0.011 | 0.171 | 46.92 |
| rs143743568 | A | G | 0.15 | 0.012 | 0.002 | 4.99E-09 | 0.005 | 0.01 | 0.615 | 34.81 |
| rs150537577 | A | G | 0.92 | -0.015 | 0.003 | 9.50E-09 | 0.027 | 0.014 | 0.062 | 33.73 |
| rs1505676 | C | G | 0.63 | 0.008 | 0.002 | 6.67E-09 | 0 | 0.008 | 0.988 | 31.36 |
| rs1542354 | A | G | 0.48 | -0.008 | 0.001 | 2.72E-09 | 0.017 | 0.008 | 0.023 | 36 |
| rs1544 | A | G | 0.26 | -0.01 | 0.002 | 1.76E-09 | -0.02 | 0.008 | 0.021 | 36 |
| rs1564347 | T | G | 0.34 | 0.009 | 0.002 | 2.39E-09 | -0.008 | 0.008 | 0.289 | 34.42 |
| rs1603460 | T | G | 0.42 | 0.009 | 0.001 | 2.20E-10 | 0 | 0.008 | 0.97 | 41.33 |
| rs162445 | A | G | 0.08 | 0.015 | 0.003 | 5.72E-09 | 0.012 | 0.014 | 0.403 | 34.18 |
| rs1689510 | C | G | 0.33 | 0.019 | 0.002 | 1.35E-38 | 0.019 | 0.008 | 0.015 | 167.27 |
| rs17048855 | A | G | 0.34 | 0.011 | 0.002 | 3.10E-13 | 0 | 0.008 | 0.981 | 51.84 |
| rs17110109 | T | C | 0.62 | -0.009 | 0.001 | 1.47E-10 | -0.001 | 0.008 | 0.892 | 44.13 |
| rs17144467 | A | C | 0.69 | 0.01 | 0.002 | 8.74E-11 | 0.003 | 0.008 | 0.74 | 42.68 |
| rs17148998 | A | G | 0.2 | 0.01 | 0.002 | 6.98E-09 | -0.001 | 0.009 | 0.904 | 35.3 |
| rs1717204 | A | C | 0.18 | -0.012 | 0.002 | 1.20E-10 | 0.012 | 0.01 | 0.2 | 42.25 |
| rs17248751 | A | G | 0.78 | -0.014 | 0.002 | 1.28E-15 | -0.006 | 0.009 | 0.483 | 64 |
| rs173003 | A | C | 0.49 | -0.008 | 0.001 | 5.50E-09 | -0.018 | 0.007 | 0.017 | 34.31 |
| rs1738050 | C | G | 0.62 | -0.009 | 0.001 | 1.42E-10 | 0 | 0.008 | 0.999 | 44.13 |
| rs17411339 | A | G | 0.56 | 0.014 | 0.001 | 5.10E-23 | 0 | 0.008 | 0.966 | 100 |
| rs17428076 | C | G | 0.76 | 0.014 | 0.002 | 6.41E-17 | -0.004 | 0.009 | 0.653 | 73.32 |
| rs1747714 | T | C | 0.53 | 0.011 | 0.001 | 3.82E-14 | 0.004 | 0.008 | 0.598 | 57.33 |
| rs1747817 | T | C | 0.76 | -0.009 | 0.002 | 2.29E-08 | -0.001 | 0.009 | 0.938 | 33.06 |
| rs17489649 | A | G | 0.68 | 0.013 | 0.002 | 2.77E-17 | 0.015 | 0.008 | 0.058 | 71.68 |
| rs17502934 | T | G | 0.15 | -0.017 | 0.002 | 2.12E-17 | -0.008 | 0.011 | 0.437 | 69.72 |
| rs175325 | A | T | 0.6 | -0.01 | 0.001 | 2.24E-12 | -0.003 | 0.008 | 0.653 | 51.02 |
| rs17551064 | A | G | 0.84 | 0.014 | 0.002 | 1.53E-13 | 0.018 | 0.01 | 0.081 | 55.07 |
| rs17563464 | A | C | 0.22 | -0.015 | 0.002 | 1.74E-17 | -0.007 | 0.009 | 0.456 | 74.77 |
| rs17565975 | A | G | 0.56 | -0.011 | 0.001 | 5.54E-14 | -0.005 | 0.008 | 0.539 | 57.33 |
| rs17598675 | T | C | 0.51 | -0.012 | 0.001 | 1.37E-16 | 0.007 | 0.008 | 0.368 | 68.65 |
| rs17604349 | A | G | 0.19 | 0.013 | 0.002 | 2.36E-12 | 0.004 | 0.01 | 0.681 | 49.78 |
| rs176218 | T | G | 0.2 | 0.02 | 0.002 | 1.10E-29 | -0.006 | 0.01 | 0.548 | 123.46 |
| rs17732878 | T | C | 0.79 | -0.01 | 0.002 | 4.06E-08 | -0.001 | 0.009 | 0.88 | 31.23 |
| rs178183 | T | C | 0.25 | 0.015 | 0.002 | 4.81E-19 | 0.006 | 0.009 | 0.508 | 82.13 |
| rs17882802 | A | G | 0.43 | 0.008 | 0.001 | 3.00E-09 | -0.004 | 0.007 | 0.601 | 36 |
| rs182902112 | A | C | 0.02 | -0.03 | 0.005 | 1.17E-08 | 0.01 | 0.032 | 0.76 | 32.04 |
| rs1842713 | A | G | 0.79 | -0.012 | 0.002 | 9.67E-12 | -0.001 | 0.009 | 0.914 | 47.37 |
| rs1861786 | A | G | 0.38 | -0.008 | 0.001 | 2.39E-08 | -0.009 | 0.008 | 0.261 | 33.47 |
| rs1865955 | T | C | 0.82 | 0.012 | 0.002 | 4.06E-11 | 0.005 | 0.01 | 0.645 | 41.23 |
| rs1866823 | A | G | 0.54 | 0.011 | 0.001 | 3.09E-14 | -0.007 | 0.008 | 0.343 | 58.41 |
| rs1890132 | T | C | 0.71 | -0.009 | 0.002 | 6.56E-09 | 0.015 | 0.008 | 0.078 | 35.2 |
| rs192436652 | T | C | 0.03 | -0.032 | 0.004 | 2.06E-12 | 0.003 | 0.023 | 0.914 | 50.25 |
| rs1933264 | T | C | 0.75 | 0.01 | 0.002 | 5.85E-10 | 0.013 | 0.009 | 0.129 | 39.06 |
| rs1955250 | A | C | 0.91 | -0.014 | 0.002 | 2.26E-08 | -0.008 | 0.014 | 0.537 | 31.36 |
| rs1991585 | T | C | 0.71 | -0.01 | 0.002 | 6.26E-11 | -0.004 | 0.008 | 0.631 | 45.34 |
| rs2007655 | T | G | 0.5 | 0.009 | 0.001 | 8.64E-10 | 0.007 | 0.008 | 0.377 | 37.73 |
| rs2011603 | A | G | 0.74 | -0.009 | 0.002 | 1.12E-08 | -0.019 | 0.008 | 0.024 | 32.35 |
| rs2034631 | T | C | 0.65 | 0.009 | 0.002 | 2.64E-09 | -0.001 | 0.008 | 0.884 | 35.2 |
| rs2052285 | A | G | 0.6 | 0.011 | 0.001 | 7.25E-15 | 0.012 | 0.008 | 0.13 | 64 |
| rs2055940 | A | G | 0.32 | 0.008 | 0.002 | 4.95E-08 | 0.017 | 0.008 | 0.035 | 29.88 |
| rs2081652 | A | T | 0.66 | 0.012 | 0.002 | 1.09E-16 | 0.014 | 0.008 | 0.088 | 67.24 |
| rs2131167 | A | G | 0.45 | 0.008 | 0.001 | 1.52E-08 | 0.004 | 0.008 | 0.561 | 32.65 |
| rs2179152 | T | C | 0.37 | -0.013 | 0.002 | 1.68E-19 | 0.011 | 0.008 | 0.151 | 76.27 |
| rs2183271 | T | C | 0.64 | 0.008 | 0.002 | 1.91E-08 | 0.032 | 0.008 | 5.07E-05 | 31.36 |
| rs2199409 | T | C | 0.8 | 0.01 | 0.002 | 3.14E-09 | -0.001 | 0.009 | 0.884 | 36.71 |
| rs2212430 | T | C | 0.3 | -0.01 | 0.002 | 1.32E-10 | 0.002 | 0.008 | 0.803 | 39.06 |
| rs2216144 | T | C | 0.53 | 0.01 | 0.001 | 2.58E-12 | -0.001 | 0.008 | 0.846 | 49 |
| rs2256965 | A | G | 0.42 | 0.011 | 0.001 | 2.40E-13 | 0.004 | 0.008 | 0.589 | 57.33 |
| rs2287838 | A | G | 0.55 | -0.01 | 0.001 | 1.27E-13 | -0.003 | 0.008 | 0.738 | 55.18 |
| rs2290601 | T | C | 0.77 | -0.015 | 0.002 | 4.49E-19 | 0 | 0.009 | 0.985 | 76.82 |
| rs2297293 | C | G | 0.31 | 0.01 | 0.002 | 8.80E-11 | 0.01 | 0.008 | 0.21 | 43.56 |
| rs232496 | T | C | 0.37 | 0.009 | 0.002 | 7.14E-10 | -0.01 | 0.008 | 0.2 | 36 |
| rs2364544 | A | G | 0.39 | -0.011 | 0.001 | 2.91E-13 | -0.012 | 0.008 | 0.121 | 56.25 |
| rs2434672 | A | C | 0.53 | 0.009 | 0.001 | 1.06E-10 | 0.006 | 0.008 | 0.421 | 42.25 |
| rs2436760 | T | C | 0.97 | 0.023 | 0.004 | 4.52E-08 | 0.012 | 0.022 | 0.582 | 30.51 |
| rs2447097 | T | G | 0.46 | 0.01 | 0.001 | 1.10E-11 | 0.029 | 0.008 | 1.14E-04 | 47.02 |
| rs2469226 | A | T | 0.22 | 0.01 | 0.002 | 1.28E-08 | -0.015 | 0.009 | 0.086 | 31.89 |
| rs2496482 | T | C | 0.42 | 0.012 | 0.002 | 9.20E-17 | -0.042 | 0.008 | 5.03E-08 | 65.07 |
| rs252991 | A | G | 0.37 | 0.01 | 0.002 | 6.30E-11 | -0.001 | 0.008 | 0.896 | 40.11 |
| rs2545795 | A | C | 0.44 | 0.013 | 0.001 | 1.03E-18 | 0.012 | 0.008 | 0.102 | 79.72 |
| rs2554835 | A | G | 0.4 | 0.009 | 0.001 | 8.95E-10 | -0.016 | 0.008 | 0.042 | 39.51 |
| rs2570497 | T | C | 0.64 | -0.012 | 0.002 | 1.27E-16 | -0.001 | 0.008 | 0.916 | 65.07 |
| rs2725370 | T | C | 0.3 | -0.014 | 0.002 | 4.96E-20 | -0.003 | 0.008 | 0.709 | 88.36 |
| rs2764684 | T | C | 0.83 | 0.014 | 0.002 | 9.99E-15 | 0.05 | 0.01 | 5.94E-07 | 56.65 |
| rs2838006 | T | C | 0.36 | 0.013 | 0.002 | 1.38E-19 | -0.015 | 0.008 | 0.053 | 77.44 |
| rs28513882 | A | G | 0.18 | -0.011 | 0.002 | 2.03E-09 | -0.022 | 0.01 | 0.026 | 37.35 |
| rs2885198 | A | G | 0.53 | 0.009 | 0.001 | 4.63E-10 | 0.012 | 0.008 | 0.114 | 39.51 |
| rs2923424 | A | G | 0.61 | 0.012 | 0.001 | 6.31E-17 | 0 | 0.008 | 0.99 | 73.47 |
| rs2929032 | A | G | 0.47 | -0.008 | 0.001 | 8.40E-09 | 0.012 | 0.007 | 0.113 | 33.47 |
| rs2958182 | A | T | 0.34 | 0.008 | 0.002 | 3.46E-08 | -0.006 | 0.008 | 0.47 | 29.88 |
| rs2964199 | T | C | 0.31 | -0.01 | 0.002 | 4.61E-11 | -0.001 | 0.008 | 0.92 | 43.56 |
| rs2964255 | A | G | 0.31 | 0.009 | 0.002 | 6.34E-09 | -0.002 | 0.008 | 0.811 | 34.42 |
| rs2998299 | T | G | 0.21 | -0.014 | 0.002 | 1.51E-15 | -0.002 | 0.009 | 0.817 | 64 |
| rs3026996 | A | C | 0.76 | 0.014 | 0.002 | 5.42E-17 | 0.008 | 0.009 | 0.389 | 74.39 |
| rs303752 | A | G | 0.41 | -0.011 | 0.001 | 1.47E-14 | 0.006 | 0.008 | 0.437 | 61.73 |
| rs3111251 | T | C | 0.43 | -0.009 | 0.001 | 3.88E-10 | -0.003 | 0.008 | 0.718 | 41.33 |
| rs34067381 | T | G | 0.36 | -0.01 | 0.002 | 9.77E-13 | 0 | 0.008 | 0.981 | 48.07 |
| rs34098770 | A | G | 0.15 | -0.016 | 0.002 | 2.95E-16 | -0.015 | 0.011 | 0.142 | 64 |
| rs34305371 | A | G | 0.1 | 0.031 | 0.002 | 1.10E-39 | -0.025 | 0.012 | 0.046 | 171.17 |
| rs34394051 | A | G | 0.84 | -0.012 | 0.002 | 2.30E-09 | -0.006 | 0.01 | 0.529 | 34.22 |
| rs34720381 | T | C | 0.09 | -0.018 | 0.002 | 1.90E-13 | 0.03 | 0.013 | 0.02 | 56.25 |
| rs34807077 | A | C | 0.16 | 0.013 | 0.002 | 3.29E-11 | -0.009 | 0.01 | 0.396 | 45.39 |
| rs35417702 | T | C | 0.52 | -0.015 | 0.001 | 2.40E-27 | -0.007 | 0.007 | 0.322 | 117.88 |
| rs35606437 | A | G | 0.27 | 0.011 | 0.002 | 3.98E-11 | -0.004 | 0.009 | 0.626 | 43.07 |
| rs35929923 | A | G | 0.25 | -0.011 | 0.002 | 1.77E-12 | -0.019 | 0.009 | 0.028 | 50.77 |
| rs363096 | T | C | 0.42 | -0.014 | 0.001 | 5.77E-24 | 0.008 | 0.008 | 0.305 | 104.33 |
| rs3751331 | A | G | 0.38 | -0.01 | 0.001 | 1.75E-11 | 0.005 | 0.008 | 0.493 | 48.01 |
| rs3809634 | A | G | 0.68 | -0.011 | 0.002 | 6.82E-13 | -0.006 | 0.008 | 0.441 | 52.8 |
| rs382196 | T | G | 0.38 | -0.008 | 0.001 | 1.47E-08 | 0.005 | 0.008 | 0.487 | 34.31 |
| rs3859523 | T | C | 0.82 | 0.012 | 0.002 | 9.92E-12 | 0.023 | 0.01 | 0.019 | 47.46 |
| rs3897821 | A | G | 0.67 | 0.015 | 0.002 | 3.58E-25 | 0.023 | 0.008 | 0.005 | 105.4 |
| rs3948495 | T | G | 0.41 | -0.009 | 0.001 | 3.95E-10 | -0.008 | 0.008 | 0.296 | 40.41 |
| rs401526 | T | C | 0.53 | -0.01 | 0.001 | 1.15E-13 | -0.02 | 0.007 | 0.008 | 55.18 |
| rs401966 | C | G | 0.39 | -0.01 | 0.001 | 1.72E-11 | 0.002 | 0.008 | 0.828 | 48.01 |
| rs42210 | C | G | 0.71 | -0.01 | 0.002 | 2.74E-10 | 0.015 | 0.008 | 0.072 | 37.52 |
| rs42302 | A | G | 0.34 | 0.009 | 0.002 | 5.96E-09 | 0.008 | 0.008 | 0.284 | 32.87 |
| rs4298514 | T | C | 0.7 | 0.01 | 0.002 | 1.07E-11 | 0.017 | 0.008 | 0.035 | 48.07 |
| rs4320563 | A | G | 0.51 | 0.011 | 0.001 | 2.24E-14 | -0.004 | 0.007 | 0.572 | 60.62 |
| rs4328757 | T | C | 0.61 | 0.01 | 0.001 | 2.95E-11 | -0.009 | 0.008 | 0.253 | 47.02 |
| rs4352658 | T | C | 0.08 | -0.019 | 0.003 | 1.71E-13 | -0.013 | 0.014 | 0.339 | 53.4 |
| rs4358081 | A | C | 0.53 | -0.01 | 0.001 | 1.28E-11 | 0.002 | 0.008 | 0.824 | 46.05 |
| rs4369924 | A | G | 0.16 | 0.013 | 0.002 | 5.58E-11 | 0.009 | 0.01 | 0.368 | 43.28 |
| rs4384309 | A | G | 0.46 | 0.01 | 0.001 | 4.89E-13 | -0.003 | 0.008 | 0.685 | 53.08 |
| rs4467547 | T | G | 0.41 | 0.012 | 0.001 | 4.88E-17 | 0.009 | 0.008 | 0.248 | 74.7 |
| rs4500930 | T | C | 0.34 | -0.011 | 0.002 | 1.67E-13 | 0.008 | 0.008 | 0.324 | 52.8 |
| rs4663617 | A | T | 0.23 | 0.01 | 0.002 | 4.12E-10 | 0.005 | 0.009 | 0.584 | 37.43 |
| rs4673840 | T | C | 0.84 | -0.014 | 0.002 | 5.86E-13 | -0.009 | 0.01 | 0.38 | 52.75 |
| rs4675248 | A | G | 0.4 | -0.009 | 0.001 | 4.50E-11 | 0.006 | 0.008 | 0.423 | 45.08 |
| rs4719944 | T | C | 0.55 | -0.012 | 0.001 | 9.25E-18 | -0.002 | 0.008 | 0.808 | 74.7 |
| rs4726070 | A | G | 0.6 | 0.012 | 0.001 | 1.36E-17 | 0.014 | 0.008 | 0.076 | 75.94 |
| rs4757957 | C | G | 0.69 | 0.013 | 0.002 | 1.79E-18 | 0.01 | 0.008 | 0.238 | 78.62 |
| rs4766424 | C | G | 0.12 | 0.014 | 0.002 | 2.47E-11 | 0.004 | 0.011 | 0.729 | 45.72 |
| rs4778058 | T | C | 0.5 | -0.009 | 0.001 | 4.92E-11 | -0.018 | 0.008 | 0.02 | 43.18 |
| rs4787457 | A | G | 0.63 | 0.016 | 0.002 | 1.23E-28 | 0.005 | 0.008 | 0.52 | 116.64 |
| rs4793090 | A | G | 0.66 | 0.008 | 0.002 | 1.67E-08 | -0.013 | 0.008 | 0.112 | 31.36 |
| rs4810894 | A | G | 0.37 | -0.009 | 0.002 | 4.38E-09 | -0.017 | 0.008 | 0.025 | 32.11 |
| rs4839155 | T | G | 0.77 | 0.012 | 0.002 | 4.66E-12 | -0.004 | 0.009 | 0.617 | 45.76 |
| rs4846724 | A | G | 0.53 | 0.01 | 0.001 | 2.89E-12 | 0.013 | 0.008 | 0.091 | 49 |
| rs4848924 | A | C | 0.71 | -0.012 | 0.002 | 3.56E-14 | -0.005 | 0.008 | 0.539 | 60.84 |
| rs488476 | C | G | 0.62 | -0.011 | 0.002 | 8.29E-15 | 0.009 | 0.008 | 0.262 | 56.75 |
| rs4895650 | T | C | 0.55 | 0.009 | 0.001 | 5.31E-11 | -0.01 | 0.008 | 0.185 | 44.13 |
| rs4899012 | C | G | 0.6 | -0.01 | 0.001 | 8.06E-13 | -0.011 | 0.008 | 0.138 | 54.13 |
| rs4904523 | A | G | 0.52 | -0.008 | 0.001 | 6.27E-09 | -0.001 | 0.007 | 0.934 | 34.31 |
| rs4915735 | A | G | 0.14 | 0.012 | 0.002 | 9.56E-09 | -0.002 | 0.011 | 0.835 | 33.64 |
| rs4977885 | A | G | 0.4 | -0.008 | 0.001 | 6.78E-09 | -0.001 | 0.008 | 0.929 | 36 |
| rs4984541 | A | G | 0.77 | -0.014 | 0.002 | 3.40E-16 | -0.009 | 0.009 | 0.309 | 65.9 |
| rs55736314 | C | G | 0.6 | -0.015 | 0.001 | 4.59E-27 | -0.013 | 0.008 | 0.099 | 121 |
| rs56099375 | T | C | 0.24 | 0.012 | 0.002 | 3.10E-13 | -0.031 | 0.009 | 4.24E-04 | 49.83 |
| rs56319902 | T | C | 0.22 | -0.021 | 0.002 | 5.81E-33 | -0.079 | 0.009 | 2.24E-18 | 146.84 |
| rs56391344 | A | G | 0.25 | 0.015 | 0.002 | 7.49E-20 | -0.008 | 0.009 | 0.379 | 85.56 |
| rs56794817 | A | G | 0.16 | 0.012 | 0.002 | 1.69E-10 | -0.007 | 0.01 | 0.502 | 41.23 |
| rs57349798 | A | G | 0.41 | 0.009 | 0.001 | 8.63E-11 | 0.01 | 0.008 | 0.191 | 44.13 |
| rs57352738 | A | T | 0.2 | -0.015 | 0.002 | 2.51E-18 | 0.009 | 0.009 | 0.328 | 79.94 |
| rs575113 | A | G | 0.29 | 0.013 | 0.002 | 7.52E-18 | 0.006 | 0.008 | 0.447 | 78.62 |
| rs5754753 | T | C | 0.72 | -0.013 | 0.002 | 6.15E-16 | 0.005 | 0.008 | 0.515 | 62.02 |
| rs57661533 | T | C | 0.13 | -0.011 | 0.002 | 3.24E-08 | -0.001 | 0.011 | 0.96 | 29.47 |
| rs580652 | T | C | 0.9 | -0.015 | 0.002 | 4.65E-10 | 0.01 | 0.013 | 0.411 | 37.52 |
| rs590013 | T | C | 0.68 | 0.01 | 0.002 | 2.47E-11 | -0.004 | 0.008 | 0.607 | 44.44 |
| rs6043521 | T | C | 0.6 | -0.008 | 0.001 | 9.72E-09 | 0.007 | 0.008 | 0.368 | 35.15 |
| rs6065080 | T | C | 0.36 | -0.013 | 0.002 | 7.49E-19 | -0.013 | 0.008 | 0.09 | 75.11 |
| rs6065784 | C | G | 0.7 | 0.011 | 0.002 | 5.78E-13 | 0.005 | 0.008 | 0.545 | 53.78 |
| rs61104616 | A | G | 0.52 | -0.017 | 0.001 | 1.56E-34 | 0.025 | 0.008 | 9.66E-04 | 150.94 |
| rs6123924 | A | G | 0.84 | 0.013 | 0.002 | 3.20E-11 | 0.009 | 0.01 | 0.39 | 46.1 |
| rs61527214 | A | G | 0.41 | 0.011 | 0.001 | 1.13E-14 | 0.006 | 0.008 | 0.433 | 61.73 |
| rs61997667 | T | C | 0.15 | -0.014 | 0.002 | 2.24E-13 | 0.003 | 0.01 | 0.768 | 51.84 |
| rs62155350 | A | G | 0.02 | 0.034 | 0.005 | 1.69E-10 | -0.022 | 0.027 | 0.405 | 40.67 |
| rs62155873 | T | C | 0.12 | -0.015 | 0.002 | 1.65E-12 | -0.011 | 0.011 | 0.322 | 51.7 |
| rs62174974 | A | G | 0.19 | -0.011 | 0.002 | 2.76E-09 | 0.002 | 0.01 | 0.844 | 34.03 |
| rs62179650 | A | G | 0.3 | 0.011 | 0.002 | 5.66E-13 | 0.007 | 0.008 | 0.369 | 49 |
| rs62182994 | T | C | 0.69 | 0.014 | 0.002 | 2.55E-19 | -0.005 | 0.008 | 0.562 | 83.42 |
| rs62379838 | T | C | 0.7 | 0.012 | 0.002 | 1.77E-15 | 0.01 | 0.008 | 0.234 | 65.07 |
| rs628993 | A | G | 0.11 | 0.013 | 0.002 | 3.83E-08 | -0.002 | 0.013 | 0.9 | 30.97 |
| rs6449503 | A | G | 0.5 | 0.018 | 0.001 | 1.84E-39 | 0.014 | 0.008 | 0.064 | 174.62 |
| rs6490618 | T | C | 0.33 | 0.009 | 0.002 | 3.48E-09 | -0.006 | 0.008 | 0.448 | 34.42 |
| rs6493265 | T | C | 0.39 | -0.012 | 0.001 | 2.26E-17 | -0.007 | 0.008 | 0.343 | 75.94 |
| rs6557171 | T | C | 0.32 | -0.016 | 0.002 | 5.48E-25 | -0.004 | 0.008 | 0.575 | 106.78 |
| rs660001 | A | G | 0.21 | -0.016 | 0.002 | 1.66E-21 | 0.005 | 0.009 | 0.594 | 93.07 |
| rs66568921 | T | G | 0.64 | -0.018 | 0.002 | 7.60E-34 | -0.004 | 0.008 | 0.62 | 145.6 |
| rs66721975 | A | G | 0.29 | -0.009 | 0.002 | 1.51E-08 | 0.014 | 0.008 | 0.108 | 33.64 |
| rs6690195 | T | C | 0.49 | -0.011 | 0.001 | 7.89E-16 | 0.015 | 0.008 | 0.047 | 65.15 |
| rs6697584 | T | C | 0.79 | -0.013 | 0.002 | 1.43E-13 | 0.018 | 0.009 | 0.052 | 54.93 |
| rs6704768 | A | G | 0.56 | -0.012 | 0.001 | 1.84E-16 | -0.001 | 0.008 | 0.942 | 68.65 |
| rs6731373 | A | G | 0.34 | -0.012 | 0.002 | 1.39E-14 | 0.002 | 0.008 | 0.784 | 58.78 |
| rs6812533 | T | C | 0.74 | 0.011 | 0.002 | 4.85E-11 | -0.003 | 0.009 | 0.773 | 43.89 |
| rs68145588 | T | G | 0.14 | -0.013 | 0.002 | 7.53E-10 | 0.024 | 0.011 | 0.023 | 36.57 |
| rs6917154 | T | C | 0.12 | 0.013 | 0.002 | 2.55E-09 | -0.003 | 0.011 | 0.815 | 36.57 |
| rs6924023 | A | G | 0.75 | 0.011 | 0.002 | 5.17E-11 | -0.01 | 0.009 | 0.249 | 44.72 |
| rs6977237 | T | C | 0.47 | 0.008 | 0.001 | 5.59E-09 | 0.005 | 0.007 | 0.539 | 34.31 |
| rs6994287 | A | G | 0.41 | 0.01 | 0.001 | 1.11E-12 | -0.001 | 0.008 | 0.903 | 53.08 |
| rs7012546 | T | C | 0.42 | 0.009 | 0.001 | 1.25E-10 | 0 | 0.008 | 0.966 | 43.18 |
| rs7029718 | A | G | 0.41 | 0.024 | 0.001 | 7.82E-65 | 0 | 0.008 | 0.98 | 303.76 |
| rs7041702 | A | G | 0.74 | -0.011 | 0.002 | 4.53E-12 | -0.005 | 0.009 | 0.535 | 48.13 |
| rs7167688 | T | C | 0.49 | -0.009 | 0.001 | 1.71E-11 | 0.007 | 0.007 | 0.338 | 45.08 |
| rs7171405 | A | G | 0.24 | -0.01 | 0.002 | 7.53E-10 | -0.004 | 0.009 | 0.663 | 39.85 |
| rs717996 | T | C | 0.6 | -0.012 | 0.001 | 1.89E-17 | -0.012 | 0.008 | 0.121 | 75.94 |
| rs7226824 | T | C | 0.53 | -0.008 | 0.001 | 2.72E-08 | -0.003 | 0.008 | 0.7 | 31.04 |
| rs72486027 | T | C | 0.25 | -0.01 | 0.002 | 1.61E-10 | -0.008 | 0.009 | 0.339 | 42.25 |
| rs7254263 | T | C | 0.28 | -0.012 | 0.002 | 1.84E-14 | -0.013 | 0.008 | 0.114 | 55.32 |
| rs72622559 | T | C | 0.23 | -0.012 | 0.002 | 5.50E-13 | -0.002 | 0.009 | 0.804 | 49.83 |
| rs72672052 | A | T | 0.18 | -0.01 | 0.002 | 4.87E-08 | -0.007 | 0.01 | 0.504 | 31.48 |
| rs72709560 | A | G | 0.69 | 0.009 | 0.002 | 2.03E-08 | 0.011 | 0.008 | 0.154 | 32.11 |
| rs72828517 | T | C | 0.83 | -0.017 | 0.002 | 1.38E-19 | 0.006 | 0.01 | 0.559 | 78.18 |
| rs72829857 | A | G | 0.76 | -0.015 | 0.002 | 1.52E-19 | 0.006 | 0.009 | 0.481 | 86.72 |
| rs72917504 | T | C | 0.06 | -0.018 | 0.003 | 2.91E-09 | -0.007 | 0.016 | 0.651 | 35.2 |
| rs72944064 | T | C | 0.74 | -0.01 | 0.002 | 9.39E-10 | -0.003 | 0.009 | 0.73 | 37.52 |
| rs72962169 | T | C | 0.16 | -0.017 | 0.002 | 1.26E-19 | 0.005 | 0.01 | 0.65 | 83.87 |
| rs72993796 | T | C | 0.88 | 0.013 | 0.002 | 1.12E-08 | 0.017 | 0.012 | 0.153 | 33.32 |
| rs730384 | A | G | 0.44 | 0.01 | 0.001 | 9.38E-13 | 0.005 | 0.008 | 0.544 | 52.05 |
| rs73055556 | A | G | 0.14 | 0.011 | 0.002 | 4.95E-08 | 0.039 | 0.011 | 3.01E-04 | 30.8 |
| rs73191311 | A | G | 0.34 | -0.009 | 0.002 | 2.03E-10 | -0.001 | 0.008 | 0.883 | 39.27 |
| rs7321274 | A | G | 0.8 | 0.011 | 0.002 | 8.21E-11 | -0.005 | 0.009 | 0.617 | 40.11 |
| rs7326331 | A | G | 0.28 | -0.013 | 0.002 | 3.42E-16 | 0 | 0.008 | 0.959 | 64 |
| rs73344830 | A | G | 0.42 | 0.017 | 0.001 | 1.00E-32 | -0.005 | 0.008 | 0.543 | 147.45 |
| rs736281 | T | C | 0.39 | 0.009 | 0.001 | 2.49E-09 | -0.003 | 0.008 | 0.667 | 37.73 |
| rs73648455 | T | C | 0.08 | -0.018 | 0.003 | 1.99E-12 | -0.014 | 0.014 | 0.324 | 49.54 |
| rs743316 | T | C | 0.79 | 0.011 | 0.002 | 8.21E-11 | -0.007 | 0.009 | 0.446 | 43.4 |
| rs74415461 | T | C | 0.08 | 0.018 | 0.003 | 6.25E-12 | -0.004 | 0.013 | 0.785 | 45.82 |
| rs7449561 | A | G | 0.23 | 0.01 | 0.002 | 9.73E-10 | 0.005 | 0.009 | 0.592 | 36 |
| rs7460106 | T | C | 0.76 | -0.012 | 0.002 | 2.06E-13 | -0.001 | 0.009 | 0.894 | 52.35 |
| rs746839 | C | G | 0.63 | 0.014 | 0.002 | 1.19E-20 | -0.003 | 0.008 | 0.683 | 81 |
| rs74787922 | A | C | 0.93 | 0.018 | 0.003 | 8.97E-11 | 0.009 | 0.015 | 0.547 | 42.25 |
| rs75033012 | C | G | 0.04 | -0.026 | 0.004 | 1.38E-11 | 0.001 | 0.019 | 0.953 | 46.1 |
| rs75177132 | T | C | 0.05 | 0.03 | 0.004 | 1.01E-16 | 0.008 | 0.017 | 0.635 | 68.52 |
| rs7575637 | A | G | 0.45 | 0.011 | 0.001 | 3.43E-14 | 0.007 | 0.008 | 0.365 | 58.41 |
| rs7603132 | A | G | 0.19 | 0.014 | 0.002 | 1.20E-14 | 0 | 0.009 | 0.973 | 57.93 |
| rs76076331 | T | C | 0.13 | 0.021 | 0.002 | 1.71E-23 | -0.009 | 0.011 | 0.427 | 96.23 |
| rs7617204 | A | G | 0.52 | -0.01 | 0.001 | 4.33E-12 | -0.001 | 0.007 | 0.875 | 48.01 |
| rs76235882 | A | G | 0.97 | 0.024 | 0.004 | 4.52E-10 | -0.004 | 0.023 | 0.859 | 38.19 |
| rs76246107 | A | G | 0.09 | -0.016 | 0.003 | 6.52E-10 | -0.036 | 0.014 | 0.012 | 38.82 |
| rs7650602 | T | C | 0.56 | -0.009 | 0.001 | 2.77E-10 | -0.028 | 0.008 | 2.53E-04 | 40.41 |
| rs76577427 | C | G | 0.9 | 0.02 | 0.002 | 7.78E-18 | 0 | 0.012 | 0.984 | 71.54 |
| rs7672622 | A | G | 0.75 | -0.009 | 0.002 | 4.72E-08 | 0.003 | 0.009 | 0.706 | 30.25 |
| rs7683416 | T | C | 0.46 | 0.014 | 0.001 | 6.05E-23 | 0.018 | 0.008 | 0.017 | 98.58 |
| rs76878669 | C | G | 0.76 | 0.014 | 0.002 | 3.10E-17 | 0.015 | 0.009 | 0.104 | 68.79 |
| rs7692359 | T | C | 0.78 | -0.011 | 0.002 | 1.40E-10 | 0.003 | 0.009 | 0.763 | 41.11 |
| rs77025239 | A | G | 0.16 | -0.014 | 0.002 | 4.65E-13 | -0.013 | 0.01 | 0.199 | 54.29 |
| rs77609760 | A | G | 0.93 | 0.024 | 0.003 | 7.71E-17 | 0.005 | 0.015 | 0.721 | 71.04 |
| rs77719387 | A | T | 0.02 | -0.04 | 0.006 | 3.78E-12 | -0.035 | 0.032 | 0.27 | 48.28 |
| rs7849487 | T | G | 0.66 | -0.017 | 0.002 | 1.25E-29 | 0.01 | 0.008 | 0.213 | 123.95 |
| rs78648104 | T | C | 0.91 | -0.014 | 0.002 | 1.70E-08 | -0.017 | 0.013 | 0.21 | 31.81 |
| rs7875078 | A | C | 0.46 | -0.008 | 0.001 | 2.72E-08 | 0.001 | 0.008 | 0.847 | 31.04 |
| rs7894722 | T | C | 0.37 | 0.009 | 0.001 | 5.41E-09 | -0.007 | 0.008 | 0.386 | 36.86 |
| rs790647 | A | C | 0.23 | -0.015 | 0.002 | 3.81E-18 | 0.004 | 0.009 | 0.643 | 72.75 |
| rs7910403 | T | G | 0.2 | 0.012 | 0.002 | 5.08E-12 | 0.009 | 0.009 | 0.349 | 45.94 |
| rs7924036 | T | G | 0.51 | 0.013 | 0.001 | 2.55E-21 | 0.016 | 0.008 | 0.035 | 90.25 |
| rs79265434 | A | G | 0.88 | -0.02 | 0.002 | 1.98E-19 | -0.005 | 0.012 | 0.692 | 80.18 |
| rs7972246 | T | C | 0.35 | 0.011 | 0.002 | 1.01E-13 | -0.004 | 0.008 | 0.636 | 53.78 |
| rs79728014 | A | G | 0.86 | -0.013 | 0.002 | 2.06E-11 | -0.005 | 0.011 | 0.632 | 44.89 |
| rs7974852 | A | C | 0.52 | 0.012 | 0.001 | 1.14E-16 | 0.013 | 0.008 | 0.085 | 69.84 |
| rs7977614 | A | G | 0.71 | -0.013 | 0.002 | 6.84E-17 | -0.004 | 0.008 | 0.668 | 70.14 |
| rs79994730 | T | C | 0.04 | 0.024 | 0.004 | 5.88E-12 | -0.061 | 0.017 | 4.18E-04 | 47.02 |
| rs8008382 | T | C | 0.3 | -0.011 | 0.002 | 4.07E-12 | 0.003 | 0.008 | 0.674 | 49.94 |
| rs8024 | A | C | 0.33 | -0.012 | 0.002 | 1.13E-14 | 0.017 | 0.008 | 0.03 | 58.78 |
| rs80257979 | T | G | 0.97 | -0.025 | 0.004 | 3.31E-10 | -0.052 | 0.022 | 0.018 | 39.69 |
| rs8030487 | A | G | 0.69 | 0.009 | 0.002 | 2.52E-08 | -0.003 | 0.008 | 0.744 | 32.11 |
| rs806816 | A | T | 0.75 | 0.01 | 0.002 | 4.80E-10 | 0.013 | 0.009 | 0.126 | 39.85 |
| rs8097125 | T | C | 0.38 | 0.009 | 0.001 | 2.31E-09 | -0.005 | 0.008 | 0.545 | 37.73 |
| rs912883 | T | C | 0.68 | 0.009 | 0.002 | 6.20E-09 | -0.001 | 0.008 | 0.929 | 33.64 |
| rs9289300 | T | C | 0.84 | -0.016 | 0.002 | 1.00E-17 | 0.007 | 0.01 | 0.47 | 74.5 |
| rs929511 | T | C | 0.13 | -0.016 | 0.002 | 3.91E-14 | 0.017 | 0.012 | 0.137 | 58.05 |
| rs9359939 | A | C | 0.24 | -0.011 | 0.002 | 1.51E-10 | 0.002 | 0.009 | 0.792 | 43.07 |
| rs936496 | A | G | 0.62 | -0.008 | 0.001 | 4.38E-08 | 0.001 | 0.008 | 0.874 | 31.84 |
| rs9373363 | A | G | 0.75 | -0.011 | 0.002 | 5.54E-12 | -0.006 | 0.009 | 0.485 | 48.13 |
| rs9375188 | T | C | 0.48 | 0.021 | 0.001 | 8.78E-52 | 0 | 0.008 | 0.973 | 229.31 |
| rs9411331 | A | C | 0.68 | 0.015 | 0.002 | 7.63E-22 | -0.016 | 0.008 | 0.046 | 93.44 |
| rs9492774 | C | G | 0.37 | 0.008 | 0.002 | 4.33E-08 | 0.018 | 0.008 | 0.023 | 28.44 |
| rs9529146 | T | C | 0.23 | 0.013 | 0.002 | 3.48E-14 | 0.001 | 0.009 | 0.88 | 55.81 |
| rs9556958 | T | C | 0.52 | -0.011 | 0.001 | 5.96E-15 | -0.003 | 0.008 | 0.693 | 61.73 |
| rs9616947 | T | C | 0.38 | -0.013 | 0.002 | 3.58E-20 | -0.011 | 0.008 | 0.14 | 79.8 |
| rs9633970 | T | C | 0.76 | 0.009 | 0.002 | 2.81E-08 | -0.009 | 0.009 | 0.333 | 32.35 |
| rs9655780 | A | G | 0.83 | -0.015 | 0.002 | 3.70E-16 | 0.017 | 0.01 | 0.093 | 64.84 |
| rs969512 | A | T | 0.66 | -0.011 | 0.002 | 9.84E-13 | -0.012 | 0.008 | 0.147 | 49.94 |
| rs9853928 | T | C | 0.21 | -0.013 | 0.002 | 1.59E-13 | -0.001 | 0.009 | 0.882 | 55.81 |
| rs9859556 | T | G | 0.31 | 0.029 | 0.002 | 4.61E-82 | 0.022 | 0.008 | 0.008 | 373.78 |
| rs9882532 | T | C | 0.64 | 0.012 | 0.002 | 2.15E-17 | 0.002 | 0.008 | 0.847 | 68.34 |
| rs9886703 | A | T | 0.17 | -0.014 | 0.002 | 4.46E-13 | 0.009 | 0.01 | 0.387 | 51.24 |
| rs9927137 | A | G | 0.53 | 0.008 | 0.001 | 5.11E-09 | 0.017 | 0.008 | 0.02 | 34.31 |
| rs9927842 | T | C | 0.15 | -0.013 | 0.002 | 6.22E-11 | -0.01 | 0.01 | 0.314 | 42.25 |
| rs9929556 | T | G | 0.57 | -0.01 | 0.001 | 2.31E-12 | 0.007 | 0.008 | 0.381 | 50.01 |
| rs9929762 | A | G | 0.56 | 0.01 | 0.001 | 1.03E-11 | 0.001 | 0.008 | 0.927 | 47.02 |
| rs9933256 | A | G | 0.56 | 0.012 | 0.001 | 6.06E-17 | 0.014 | 0.008 | 0.063 | 72.25 |

**SNPs used as instrumental variables in the MR analysis of educational attainment on mean curvature**

| **SNP** | **Effect allele** | **Other allele** | **Effect allele frequency** | **beta (exposure)** | **SE (exposure)** | ***P* (exposure)** | **beta (outcome)** | **SE (outcome)** | ***P* (outcome)** | ***F*-statistic** |
| --- | --- | --- | --- | --- | --- | --- | --- | --- | --- | --- |
| rs10006235 | T | C | 0.27 | -0.009 | 0.002 | 4.48E-09 | -0.015 | 0.008 | 0.049 | 33.79 |
| rs10021733 | T | C | 0.15 | -0.016 | 0.003 | 9.45E-10 | -0.008 | 0.009 | 0.402 | 36.89 |
| rs10046069 | A | G | 0.12 | 0.013 | 0.002 | 4.40E-09 | -0.002 | 0.011 | 0.866 | 33.32 |
| rs10060023 | T | C | 0.33 | 0.011 | 0.002 | 1.88E-13 | 0.001 | 0.007 | 0.878 | 53.78 |
| rs10080647 | A | C | 0.14 | 0.012 | 0.002 | 7.05E-10 | -0.001 | 0.01 | 0.894 | 38.44 |
| rs1008078 | T | C | 0.4 | -0.018 | 0.001 | 4.50E-37 | -0.007 | 0.007 | 0.307 | 169 |
| rs10189857 | A | G | 0.57 | 0.016 | 0.001 | 4.69E-29 | -0.012 | 0.007 | 0.088 | 127.37 |
| rs10193498 | A | T | 0.76 | 0.01 | 0.002 | 3.10E-09 | 0.001 | 0.008 | 0.886 | 36.75 |
| rs10215082 | A | G | 0.47 | -0.015 | 0.001 | 7.03E-26 | 0.011 | 0.007 | 0.108 | 113.27 |
| rs10460095 | A | G | 0.57 | -0.012 | 0.001 | 2.61E-18 | 0.001 | 0.007 | 0.857 | 78.45 |
| rs10761251 | A | T | 0.66 | 0.011 | 0.002 | 2.52E-14 | 0.003 | 0.007 | 0.642 | 56.75 |
| rs10772644 | C | G | 0.89 | 0.014 | 0.002 | 1.69E-10 | -0.015 | 0.011 | 0.2 | 41.66 |
| rs10773002 | A | T | 0.25 | 0.018 | 0.002 | 2.50E-30 | -0.019 | 0.008 | 0.022 | 133.69 |
| rs10773208 | T | C | 0.25 | -0.011 | 0.002 | 8.09E-12 | -0.005 | 0.008 | 0.571 | 47.27 |
| rs10798888 | T | G | 0.17 | -0.014 | 0.002 | 5.15E-14 | 0.007 | 0.009 | 0.43 | 54.29 |
| rs10799615 | A | G | 0.25 | -0.009 | 0.002 | 4.11E-08 | -0.009 | 0.008 | 0.279 | 30.94 |
| rs10805383 | A | G | 0.49 | -0.01 | 0.001 | 2.22E-13 | -0.003 | 0.007 | 0.636 | 54.13 |
| rs10830858 | T | C | 0.51 | 0.009 | 0.001 | 5.31E-11 | 0.004 | 0.007 | 0.533 | 43.18 |
| rs10844179 | A | G | 0.23 | 0.01 | 0.002 | 5.03E-10 | -0.024 | 0.008 | 0.005 | 37.43 |
| rs10875121 | C | G | 0.84 | 0.017 | 0.002 | 4.46E-20 | -0.019 | 0.01 | 0.051 | 83.87 |
| rs10892807 | T | C | 0.57 | 0.011 | 0.001 | 1.09E-14 | -0.007 | 0.007 | 0.295 | 60.62 |
| rs10940921 | T | G | 0.42 | 0.011 | 0.001 | 1.31E-14 | -0.001 | 0.007 | 0.909 | 62.86 |
| rs10951590 | T | C | 0.33 | -0.011 | 0.002 | 3.62E-13 | 0.01 | 0.007 | 0.173 | 52.8 |
| rs10979613 | T | C | 0.64 | -0.012 | 0.002 | 1.25E-15 | -0.027 | 0.007 | 1.79E-04 | 60.84 |
| rs10996167 | C | G | 0.65 | 0.011 | 0.002 | 7.47E-09 | -0.004 | 0.007 | 0.603 | 31.92 |
| rs11003463 | T | G | 0.61 | 0.009 | 0.002 | 5.58E-10 | 0.005 | 0.007 | 0.476 | 36.8 |
| rs11023764 | A | G | 0.64 | 0.009 | 0.002 | 5.32E-10 | -0.004 | 0.007 | 0.587 | 36.8 |
| rs11030102 | C | G | 0.75 | 0.01 | 0.002 | 4.60E-10 | 0.017 | 0.008 | 0.028 | 39.85 |
| rs11076962 | T | C | 0.72 | 0.009 | 0.002 | 4.29E-08 | -0.008 | 0.008 | 0.315 | 28.89 |
| rs11081529 | T | C | 0.71 | 0.013 | 0.002 | 5.22E-16 | 0 | 0.008 | 0.953 | 70.56 |
| rs11082011 | T | C | 0.67 | 0.019 | 0.002 | 2.06E-38 | 0.007 | 0.007 | 0.352 | 167.27 |
| rs11121177 | A | G | 0.18 | 0.015 | 0.002 | 1.44E-16 | 0.014 | 0.009 | 0.131 | 70.37 |
| rs11130380 | T | G | 0.61 | 0.009 | 0.001 | 4.17E-10 | 0.015 | 0.007 | 0.033 | 41.33 |
| rs11138947 | T | C | 0.72 | 0.011 | 0.002 | 2.36E-11 | 0.003 | 0.008 | 0.674 | 43.07 |
| rs11157931 | A | C | 0.39 | -0.014 | 0.001 | 3.86E-22 | 0.007 | 0.007 | 0.342 | 98.58 |
| rs111852224 | T | C | 0.12 | 0.016 | 0.002 | 4.85E-13 | -0.013 | 0.011 | 0.225 | 50.28 |
| rs11211123 | A | G | 0.22 | 0.01 | 0.002 | 7.62E-10 | -0.022 | 0.008 | 0.008 | 37.43 |
| rs11213482 | A | G | 0.84 | -0.011 | 0.002 | 2.68E-08 | -0.016 | 0.009 | 0.086 | 31.12 |
| rs112603734 | A | C | 0.77 | -0.013 | 0.002 | 2.72E-09 | -0.004 | 0.008 | 0.621 | 35.46 |
| rs112806496 | C | G | 0.91 | -0.016 | 0.002 | 1.72E-10 | 0 | 0.012 | 0.992 | 40.96 |
| rs1128956 | T | G | 0.83 | -0.014 | 0.002 | 5.34E-14 | 0.006 | 0.009 | 0.481 | 54.29 |
| rs113520408 | A | G | 0.28 | 0.016 | 0.002 | 1.03E-22 | 0.002 | 0.008 | 0.78 | 93.85 |
| rs113615161 | T | C | 0.13 | -0.012 | 0.002 | 2.98E-09 | -0.003 | 0.01 | 0.807 | 34.31 |
| rs114593137 | A | T | 0.79 | -0.012 | 0.002 | 1.65E-12 | -0.005 | 0.009 | 0.523 | 51.5 |
| rs115000530 | A | T | 0.95 | -0.025 | 0.003 | 1.09E-15 | -0.029 | 0.016 | 0.063 | 65.04 |
| rs11542663 | A | C | 0.69 | 0.011 | 0.002 | 2.44E-13 | -0.003 | 0.008 | 0.667 | 55.75 |
| rs11588857 | A | G | 0.21 | 0.02 | 0.002 | 4.45E-31 | -0.017 | 0.009 | 0.044 | 137.03 |
| rs11598765 | A | G | 0.2 | -0.011 | 0.002 | 1.73E-09 | 0.008 | 0.009 | 0.354 | 34.68 |
| rs11620355 | A | G | 0.09 | 0.018 | 0.002 | 1.09E-13 | -0.002 | 0.012 | 0.864 | 53 |
| rs11657342 | A | G | 0.37 | 0.014 | 0.002 | 2.32E-13 | 0.004 | 0.007 | 0.579 | 54.29 |
| rs11657979 | A | G | 0.24 | -0.009 | 0.002 | 4.57E-08 | 0.006 | 0.009 | 0.448 | 28.65 |
| rs11663602 | A | C | 0.28 | -0.013 | 0.002 | 9.27E-17 | 0.003 | 0.008 | 0.692 | 66.02 |
| rs1167827 | A | G | 0.43 | 0.01 | 0.001 | 2.93E-13 | -0.017 | 0.007 | 0.013 | 55.18 |
| rs11724690 | T | G | 0.29 | 0.009 | 0.002 | 2.08E-09 | -0.006 | 0.008 | 0.459 | 38.44 |
| rs117398064 | C | G | 0.09 | -0.014 | 0.002 | 1.16E-08 | -0.013 | 0.012 | 0.288 | 31.81 |
| rs118134876 | T | C | 0.06 | -0.022 | 0.003 | 1.50E-13 | -0.02 | 0.015 | 0.161 | 55.25 |
| rs11871429 | A | G | 0.77 | 0.013 | 0.002 | 2.52E-14 | 0.031 | 0.008 | 2.52E-04 | 56.69 |
| rs11894424 | A | C | 0.95 | 0.021 | 0.003 | 2.14E-10 | -0.009 | 0.017 | 0.607 | 40.11 |
| rs12054166 | C | G | 0.74 | 0.01 | 0.002 | 2.30E-09 | 0.012 | 0.008 | 0.146 | 36 |
| rs12076635 | C | G | 0.78 | 0.021 | 0.002 | 3.33E-34 | -0.018 | 0.008 | 0.036 | 148.27 |
| rs12113634 | T | C | 0.65 | 0.009 | 0.002 | 2.50E-09 | 0.011 | 0.007 | 0.12 | 34.42 |
| rs12151248 | T | C | 0.11 | -0.015 | 0.002 | 3.81E-11 | 0.007 | 0.011 | 0.508 | 41.97 |
| rs1220779 | A | G | 0.47 | -0.008 | 0.001 | 1.58E-08 | -0.005 | 0.007 | 0.508 | 31.84 |
| rs12273435 | A | G | 0.21 | -0.013 | 0.002 | 4.72E-14 | -0.005 | 0.009 | 0.599 | 60.29 |
| rs12332731 | A | T | 0.19 | 0.014 | 0.002 | 5.96E-14 | 0.024 | 0.009 | 0.006 | 56.25 |
| rs12375949 | T | C | 0.43 | -0.014 | 0.001 | 1.61E-22 | 0.002 | 0.007 | 0.793 | 97.16 |
| rs12438177 | A | G | 0.37 | 0.009 | 0.002 | 1.48E-10 | 0.001 | 0.007 | 0.849 | 38.44 |
| rs12468040 | T | G | 0.38 | 0.014 | 0.001 | 3.12E-21 | 0.007 | 0.007 | 0.307 | 95.76 |
| rs12503522 | T | C | 0.28 | -0.011 | 0.002 | 3.93E-13 | 0.002 | 0.008 | 0.798 | 49.88 |
| rs12506221 | T | G | 0.44 | -0.013 | 0.001 | 4.23E-20 | -0.002 | 0.007 | 0.752 | 86.22 |
| rs12519073 | T | C | 0.23 | -0.011 | 0.002 | 1.16E-10 | 0.01 | 0.008 | 0.253 | 39.62 |
| rs12524795 | T | C | 0.44 | 0.01 | 0.001 | 5.28E-13 | -0.009 | 0.007 | 0.22 | 53.08 |
| rs12571549 | A | G | 0.14 | 0.015 | 0.002 | 1.45E-13 | -0.015 | 0.01 | 0.132 | 57 |
| rs12574281 | A | C | 0.63 | -0.008 | 0.002 | 3.04E-08 | -0.007 | 0.007 | 0.309 | 28.44 |
| rs12591647 | T | C | 0.82 | -0.015 | 0.002 | 7.78E-17 | 0.003 | 0.009 | 0.706 | 71.31 |
| rs12602286 | T | G | 0.87 | 0.016 | 0.002 | 4.62E-14 | -0.008 | 0.011 | 0.452 | 58.05 |
| rs12614263 | A | G | 0.5 | 0.008 | 0.001 | 3.38E-09 | 0.005 | 0.007 | 0.454 | 35.15 |
| rs12638072 | A | G | 0.3 | -0.011 | 0.002 | 3.13E-12 | 0.003 | 0.008 | 0.746 | 50.88 |
| rs12643771 | T | C | 0.31 | 0.013 | 0.002 | 1.32E-17 | 0.009 | 0.008 | 0.25 | 75.11 |
| rs12646523 | T | C | 0.25 | -0.013 | 0.002 | 3.79E-16 | 0.02 | 0.008 | 0.015 | 68.06 |
| rs12670376 | A | G | 0.45 | 0.009 | 0.001 | 5.99E-11 | 0.002 | 0.007 | 0.809 | 43.18 |
| rs12761761 | T | C | 0.24 | 0.015 | 0.002 | 4.79E-20 | 0.024 | 0.008 | 0.003 | 81 |
| rs12765185 | A | T | 0.28 | -0.009 | 0.002 | 2.96E-08 | -0.015 | 0.008 | 0.054 | 30.25 |
| rs12789313 | T | C | 0.49 | 0.009 | 0.001 | 6.79E-11 | 0.007 | 0.007 | 0.35 | 43.18 |
| rs12875339 | A | C | 0.64 | -0.011 | 0.002 | 1.71E-13 | 0.003 | 0.007 | 0.637 | 51.84 |
| rs12888615 | T | C | 0.2 | 0.012 | 0.002 | 9.42E-12 | 0.013 | 0.009 | 0.135 | 44.44 |
| rs12957463 | A | G | 0.8 | 0.015 | 0.002 | 9.20E-17 | 0.01 | 0.009 | 0.272 | 65.79 |
| rs12967010 | T | C | 0.77 | 0.011 | 0.002 | 1.33E-11 | -0.012 | 0.008 | 0.162 | 44.97 |
| rs12981405 | T | C | 0.17 | -0.011 | 0.002 | 2.75E-09 | 0.001 | 0.009 | 0.903 | 34.75 |
| rs1301838 | T | C | 0.32 | -0.01 | 0.002 | 1.29E-10 | -0.003 | 0.007 | 0.668 | 40.96 |
| rs13018640 | T | C | 0.6 | -0.021 | 0.001 | 1.05E-50 | 0.004 | 0.007 | 0.619 | 235.84 |
| rs13050131 | A | C | 0.66 | -0.008 | 0.002 | 4.40E-08 | -0.01 | 0.007 | 0.171 | 29.16 |
| rs13133213 | A | G | 0.5 | 0.01 | 0.001 | 7.57E-12 | 0 | 0.007 | 0.95 | 47.02 |
| rs13145650 | T | C | 0.92 | -0.017 | 0.002 | 8.99E-12 | 0.003 | 0.012 | 0.823 | 47.89 |
| rs13163845 | T | C | 0.85 | -0.015 | 0.002 | 4.66E-14 | 0.002 | 0.01 | 0.842 | 55.5 |
| rs13197257 | T | G | 0.28 | 0.011 | 0.002 | 2.83E-12 | 0.028 | 0.008 | 3.72E-04 | 46.41 |
| rs13212041 | T | C | 0.8 | 0.01 | 0.002 | 7.42E-09 | 0.012 | 0.009 | 0.148 | 31.48 |
| rs13240401 | T | C | 0.78 | 0.018 | 0.002 | 2.36E-24 | -0.006 | 0.008 | 0.488 | 107.18 |
| rs13261773 | C | G | 0.16 | 0.012 | 0.002 | 3.12E-10 | 0.017 | 0.01 | 0.08 | 41.23 |
| rs13266287 | A | T | 0.67 | -0.009 | 0.002 | 1.75E-08 | -0.016 | 0.007 | 0.032 | 32.11 |
| rs1329125 | T | C | 0.33 | -0.01 | 0.002 | 8.37E-11 | 0.002 | 0.008 | 0.806 | 41.82 |
| rs13327482 | A | G | 0.82 | -0.012 | 0.002 | 4.47E-11 | 0.01 | 0.009 | 0.256 | 45.19 |
| rs1334297 | A | G | 0.74 | 0.026 | 0.002 | 2.53E-58 | 0.006 | 0.008 | 0.457 | 258 |
| rs13422673 | T | C | 0.47 | -0.011 | 0.001 | 6.48E-15 | -0.009 | 0.007 | 0.206 | 61.73 |
| rs137079 | T | C | 0.14 | 0.013 | 0.002 | 3.06E-10 | 0.008 | 0.01 | 0.43 | 40.96 |
| rs1426619 | T | C | 0.43 | 0.01 | 0.001 | 1.40E-11 | 0.004 | 0.007 | 0.53 | 47.02 |
| rs1434630 | T | G | 0.15 | -0.014 | 0.002 | 4.63E-12 | 0.007 | 0.01 | 0.447 | 46.92 |
| rs143743568 | A | G | 0.15 | 0.012 | 0.002 | 4.99E-09 | 0.014 | 0.01 | 0.154 | 34.81 |
| rs150537577 | A | G | 0.92 | -0.015 | 0.003 | 9.50E-09 | 0.009 | 0.013 | 0.499 | 33.73 |
| rs1505676 | C | G | 0.63 | 0.008 | 0.002 | 6.67E-09 | -0.003 | 0.007 | 0.679 | 31.36 |
| rs1542354 | A | G | 0.48 | -0.008 | 0.001 | 2.72E-09 | 0.009 | 0.007 | 0.203 | 36 |
| rs1544 | A | G | 0.26 | -0.01 | 0.002 | 1.76E-09 | 0.019 | 0.008 | 0.016 | 36 |
| rs1564347 | T | G | 0.34 | 0.009 | 0.002 | 2.39E-09 | -0.017 | 0.007 | 0.02 | 34.42 |
| rs1603460 | T | G | 0.42 | 0.009 | 0.001 | 2.20E-10 | -0.005 | 0.007 | 0.471 | 41.33 |
| rs162445 | A | G | 0.08 | 0.015 | 0.003 | 5.72E-09 | 0.009 | 0.013 | 0.509 | 34.18 |
| rs1689510 | C | G | 0.33 | 0.019 | 0.002 | 1.35E-38 | -0.017 | 0.007 | 0.019 | 167.27 |
| rs17048855 | A | G | 0.34 | 0.011 | 0.002 | 3.10E-13 | 0.003 | 0.007 | 0.701 | 51.84 |
| rs17110109 | T | C | 0.62 | -0.009 | 0.001 | 1.47E-10 | -0.008 | 0.007 | 0.264 | 44.13 |
| rs17144467 | A | C | 0.69 | 0.01 | 0.002 | 8.74E-11 | -0.012 | 0.008 | 0.11 | 42.68 |
| rs17148998 | A | G | 0.2 | 0.01 | 0.002 | 6.98E-09 | 0.022 | 0.009 | 0.01 | 35.3 |
| rs1717204 | A | C | 0.18 | -0.012 | 0.002 | 1.20E-10 | 0.009 | 0.009 | 0.319 | 42.25 |
| rs17248751 | A | G | 0.78 | -0.014 | 0.002 | 1.28E-15 | -0.008 | 0.008 | 0.325 | 64 |
| rs173003 | A | C | 0.49 | -0.008 | 0.001 | 5.50E-09 | -0.006 | 0.007 | 0.368 | 34.31 |
| rs1738050 | C | G | 0.62 | -0.009 | 0.001 | 1.42E-10 | 0.009 | 0.007 | 0.198 | 44.13 |
| rs17411339 | A | G | 0.56 | 0.014 | 0.001 | 5.10E-23 | 0.01 | 0.007 | 0.139 | 100 |
| rs17428076 | C | G | 0.76 | 0.014 | 0.002 | 6.41E-17 | -0.007 | 0.008 | 0.42 | 73.32 |
| rs1747714 | T | C | 0.53 | 0.011 | 0.001 | 3.82E-14 | 0.006 | 0.007 | 0.407 | 57.33 |
| rs1747817 | T | C | 0.76 | -0.009 | 0.002 | 2.29E-08 | -0.004 | 0.008 | 0.658 | 33.06 |
| rs17489649 | A | G | 0.68 | 0.013 | 0.002 | 2.77E-17 | 0.007 | 0.007 | 0.339 | 71.68 |
| rs17502934 | T | G | 0.15 | -0.017 | 0.002 | 2.12E-17 | -0.005 | 0.01 | 0.651 | 69.72 |
| rs175325 | A | T | 0.6 | -0.01 | 0.001 | 2.24E-12 | 0.015 | 0.007 | 0.04 | 51.02 |
| rs17551064 | A | G | 0.84 | 0.014 | 0.002 | 1.53E-13 | 0.007 | 0.01 | 0.477 | 55.07 |
| rs17563464 | A | C | 0.22 | -0.015 | 0.002 | 1.74E-17 | 0.01 | 0.009 | 0.265 | 74.77 |
| rs17565975 | A | G | 0.56 | -0.011 | 0.001 | 5.54E-14 | -0.027 | 0.007 | 1.84E-04 | 57.33 |
| rs17598675 | T | C | 0.51 | -0.012 | 0.001 | 1.37E-16 | 0.01 | 0.007 | 0.161 | 68.65 |
| rs17604349 | A | G | 0.19 | 0.013 | 0.002 | 2.36E-12 | -0.003 | 0.009 | 0.719 | 49.78 |
| rs176218 | T | G | 0.2 | 0.02 | 0.002 | 1.10E-29 | -0.012 | 0.009 | 0.183 | 123.46 |
| rs17732878 | T | C | 0.79 | -0.01 | 0.002 | 4.06E-08 | -0.003 | 0.009 | 0.73 | 31.23 |
| rs178183 | T | C | 0.25 | 0.015 | 0.002 | 4.81E-19 | -0.002 | 0.008 | 0.785 | 82.13 |
| rs17882802 | A | G | 0.43 | 0.008 | 0.001 | 3.00E-09 | -0.021 | 0.007 | 0.003 | 36 |
| rs182902112 | A | C | 0.02 | -0.03 | 0.005 | 1.17E-08 | 0.008 | 0.03 | 0.778 | 32.04 |
| rs1842713 | A | G | 0.79 | -0.012 | 0.002 | 9.67E-12 | 0.003 | 0.009 | 0.763 | 47.37 |
| rs1861786 | A | G | 0.38 | -0.008 | 0.001 | 2.39E-08 | -0.007 | 0.007 | 0.344 | 33.47 |
| rs1865955 | T | C | 0.82 | 0.012 | 0.002 | 4.06E-11 | -0.006 | 0.009 | 0.484 | 41.23 |
| rs1866823 | A | G | 0.54 | 0.011 | 0.001 | 3.09E-14 | 0 | 0.007 | 0.995 | 58.41 |
| rs1890132 | T | C | 0.71 | -0.009 | 0.002 | 6.56E-09 | 0.008 | 0.008 | 0.308 | 35.2 |
| rs192436652 | T | C | 0.03 | -0.032 | 0.004 | 2.06E-12 | 0.022 | 0.022 | 0.32 | 50.25 |
| rs1933264 | T | C | 0.75 | 0.01 | 0.002 | 5.85E-10 | -0.008 | 0.008 | 0.303 | 39.06 |
| rs1955250 | A | C | 0.91 | -0.014 | 0.002 | 2.26E-08 | -0.018 | 0.013 | 0.147 | 31.36 |
| rs1991585 | T | C | 0.71 | -0.01 | 0.002 | 6.26E-11 | -0.002 | 0.008 | 0.785 | 45.34 |
| rs2007655 | T | G | 0.5 | 0.009 | 0.001 | 8.64E-10 | 0.007 | 0.007 | 0.335 | 37.73 |
| rs2011603 | A | G | 0.74 | -0.009 | 0.002 | 1.12E-08 | 0 | 0.008 | 0.983 | 32.35 |
| rs2034631 | T | C | 0.65 | 0.009 | 0.002 | 2.64E-09 | -0.003 | 0.007 | 0.684 | 35.2 |
| rs2052285 | A | G | 0.6 | 0.011 | 0.001 | 7.25E-15 | 0.005 | 0.007 | 0.485 | 64 |
| rs2055940 | A | G | 0.32 | 0.008 | 0.002 | 4.95E-08 | 0.004 | 0.007 | 0.6 | 29.88 |
| rs2081652 | A | T | 0.66 | 0.012 | 0.002 | 1.09E-16 | 0.001 | 0.007 | 0.878 | 67.24 |
| rs2131167 | A | G | 0.45 | 0.008 | 0.001 | 1.52E-08 | -0.007 | 0.007 | 0.351 | 32.65 |
| rs2179152 | T | C | 0.37 | -0.013 | 0.002 | 1.68E-19 | -0.003 | 0.007 | 0.685 | 76.27 |
| rs2183271 | T | C | 0.64 | 0.008 | 0.002 | 1.91E-08 | 0.012 | 0.007 | 0.095 | 31.36 |
| rs2199409 | T | C | 0.8 | 0.01 | 0.002 | 3.14E-09 | 0.008 | 0.009 | 0.389 | 36.71 |
| rs2212430 | T | C | 0.3 | -0.01 | 0.002 | 1.32E-10 | 0.009 | 0.008 | 0.263 | 39.06 |
| rs2216144 | T | C | 0.53 | 0.01 | 0.001 | 2.58E-12 | -0.003 | 0.007 | 0.625 | 49 |
| rs2256965 | A | G | 0.42 | 0.011 | 0.001 | 2.40E-13 | -0.016 | 0.007 | 0.021 | 57.33 |
| rs2287838 | A | G | 0.55 | -0.01 | 0.001 | 1.27E-13 | 0 | 0.007 | 0.989 | 55.18 |
| rs2290601 | T | C | 0.77 | -0.015 | 0.002 | 4.49E-19 | 0 | 0.008 | 0.986 | 76.82 |
| rs2297293 | C | G | 0.31 | 0.01 | 0.002 | 8.80E-11 | 0.003 | 0.008 | 0.685 | 43.56 |
| rs232496 | T | C | 0.37 | 0.009 | 0.002 | 7.14E-10 | -0.005 | 0.007 | 0.481 | 36 |
| rs2364544 | A | G | 0.39 | -0.011 | 0.001 | 2.91E-13 | 0 | 0.007 | 0.973 | 56.25 |
| rs2434672 | A | C | 0.53 | 0.009 | 0.001 | 1.06E-10 | -0.005 | 0.007 | 0.464 | 42.25 |
| rs2436760 | T | C | 0.97 | 0.023 | 0.004 | 4.52E-08 | -0.025 | 0.021 | 0.219 | 30.51 |
| rs2447097 | T | G | 0.46 | 0.01 | 0.001 | 1.10E-11 | 0.021 | 0.007 | 0.003 | 47.02 |
| rs2469226 | A | T | 0.22 | 0.01 | 0.002 | 1.28E-08 | -0.004 | 0.008 | 0.596 | 31.89 |
| rs2496482 | T | C | 0.42 | 0.012 | 0.002 | 9.20E-17 | -0.007 | 0.007 | 0.31 | 65.07 |
| rs252991 | A | G | 0.37 | 0.01 | 0.002 | 6.30E-11 | 0 | 0.007 | 0.964 | 40.11 |
| rs2545795 | A | C | 0.44 | 0.013 | 0.001 | 1.03E-18 | 0.01 | 0.007 | 0.165 | 79.72 |
| rs2554835 | A | G | 0.4 | 0.009 | 0.001 | 8.95E-10 | -0.009 | 0.007 | 0.204 | 39.51 |
| rs2570497 | T | C | 0.64 | -0.012 | 0.002 | 1.27E-16 | 0.005 | 0.007 | 0.518 | 65.07 |
| rs2725370 | T | C | 0.3 | -0.014 | 0.002 | 4.96E-20 | -0.007 | 0.008 | 0.384 | 88.36 |
| rs2764684 | T | C | 0.83 | 0.014 | 0.002 | 9.99E-15 | 0.056 | 0.009 | 1.39E-09 | 56.65 |
| rs2838006 | T | C | 0.36 | 0.013 | 0.002 | 1.38E-19 | 0.006 | 0.007 | 0.405 | 77.44 |
| rs28513882 | A | G | 0.18 | -0.011 | 0.002 | 2.03E-09 | -0.02 | 0.009 | 0.026 | 37.35 |
| rs2885198 | A | G | 0.53 | 0.009 | 0.001 | 4.63E-10 | 0.001 | 0.007 | 0.882 | 39.51 |
| rs2923424 | A | G | 0.61 | 0.012 | 0.001 | 6.31E-17 | 0.011 | 0.007 | 0.113 | 73.47 |
| rs2929032 | A | G | 0.47 | -0.008 | 0.001 | 8.40E-09 | -0.004 | 0.007 | 0.554 | 33.47 |
| rs2958182 | A | T | 0.34 | 0.008 | 0.002 | 3.46E-08 | -0.011 | 0.007 | 0.128 | 29.88 |
| rs2964199 | T | C | 0.31 | -0.01 | 0.002 | 4.61E-11 | -0.008 | 0.008 | 0.313 | 43.56 |
| rs2964255 | A | G | 0.31 | 0.009 | 0.002 | 6.34E-09 | 0.004 | 0.008 | 0.622 | 34.42 |
| rs2998299 | T | G | 0.21 | -0.014 | 0.002 | 1.51E-15 | 0.006 | 0.009 | 0.453 | 64 |
| rs3026996 | A | C | 0.76 | 0.014 | 0.002 | 5.42E-17 | 0.011 | 0.008 | 0.161 | 74.39 |
| rs303752 | A | G | 0.41 | -0.011 | 0.001 | 1.47E-14 | -0.006 | 0.007 | 0.424 | 61.73 |
| rs3111251 | T | C | 0.43 | -0.009 | 0.001 | 3.88E-10 | 0.006 | 0.007 | 0.375 | 41.33 |
| rs34067381 | T | G | 0.36 | -0.01 | 0.002 | 9.77E-13 | -0.001 | 0.007 | 0.863 | 48.07 |
| rs34098770 | A | G | 0.15 | -0.016 | 0.002 | 2.95E-16 | -0.005 | 0.01 | 0.604 | 64 |
| rs34305371 | A | G | 0.1 | 0.031 | 0.002 | 1.10E-39 | -0.011 | 0.012 | 0.357 | 171.17 |
| rs34394051 | A | G | 0.84 | -0.012 | 0.002 | 2.30E-09 | 0.015 | 0.01 | 0.128 | 34.22 |
| rs34720381 | T | C | 0.09 | -0.018 | 0.002 | 1.90E-13 | 0.022 | 0.012 | 0.064 | 56.25 |
| rs34807077 | A | C | 0.16 | 0.013 | 0.002 | 3.29E-11 | 0.019 | 0.01 | 0.049 | 45.39 |
| rs35417702 | T | C | 0.52 | -0.015 | 0.001 | 2.40E-27 | -0.001 | 0.007 | 0.862 | 117.88 |
| rs35606437 | A | G | 0.27 | 0.011 | 0.002 | 3.98E-11 | 0.006 | 0.008 | 0.437 | 43.07 |
| rs35929923 | A | G | 0.25 | -0.011 | 0.002 | 1.77E-12 | -0.008 | 0.008 | 0.318 | 50.77 |
| rs363096 | T | C | 0.42 | -0.014 | 0.001 | 5.77E-24 | 0.005 | 0.007 | 0.44 | 104.33 |
| rs3751331 | A | G | 0.38 | -0.01 | 0.001 | 1.75E-11 | 0.001 | 0.007 | 0.908 | 48.01 |
| rs3809634 | A | G | 0.68 | -0.011 | 0.002 | 6.82E-13 | -0.023 | 0.008 | 0.003 | 52.8 |
| rs382196 | T | G | 0.38 | -0.008 | 0.001 | 1.47E-08 | -0.008 | 0.007 | 0.24 | 34.31 |
| rs3859523 | T | C | 0.82 | 0.012 | 0.002 | 9.92E-12 | 0.017 | 0.009 | 0.07 | 47.46 |
| rs3897821 | A | G | 0.67 | 0.015 | 0.002 | 3.58E-25 | -0.009 | 0.007 | 0.248 | 105.4 |
| rs3948495 | T | G | 0.41 | -0.009 | 0.001 | 3.95E-10 | -0.024 | 0.007 | 9.21E-04 | 40.41 |
| rs401526 | T | C | 0.53 | -0.01 | 0.001 | 1.15E-13 | 0.017 | 0.007 | 0.015 | 55.18 |
| rs401966 | C | G | 0.39 | -0.01 | 0.001 | 1.72E-11 | -0.009 | 0.007 | 0.234 | 48.01 |
| rs42210 | C | G | 0.71 | -0.01 | 0.002 | 2.74E-10 | -0.011 | 0.008 | 0.166 | 37.52 |
| rs42302 | A | G | 0.34 | 0.009 | 0.002 | 5.96E-09 | -0.008 | 0.007 | 0.279 | 32.87 |
| rs4298514 | T | C | 0.7 | 0.01 | 0.002 | 1.07E-11 | -0.005 | 0.008 | 0.538 | 48.07 |
| rs4320563 | A | G | 0.51 | 0.011 | 0.001 | 2.24E-14 | -0.014 | 0.007 | 0.039 | 60.62 |
| rs4328757 | T | C | 0.61 | 0.01 | 0.001 | 2.95E-11 | 0 | 0.007 | 0.982 | 47.02 |
| rs4352658 | T | C | 0.08 | -0.019 | 0.003 | 1.71E-13 | 0.004 | 0.013 | 0.745 | 53.4 |
| rs4358081 | A | C | 0.53 | -0.01 | 0.001 | 1.28E-11 | -0.014 | 0.007 | 0.048 | 46.05 |
| rs4369924 | A | G | 0.16 | 0.013 | 0.002 | 5.58E-11 | -0.001 | 0.01 | 0.956 | 43.28 |
| rs4384309 | A | G | 0.46 | 0.01 | 0.001 | 4.89E-13 | 0.004 | 0.007 | 0.554 | 53.08 |
| rs4467547 | T | G | 0.41 | 0.012 | 0.001 | 4.88E-17 | 0.003 | 0.007 | 0.722 | 74.7 |
| rs4500930 | T | C | 0.34 | -0.011 | 0.002 | 1.67E-13 | -0.013 | 0.007 | 0.08 | 52.8 |
| rs4663617 | A | T | 0.23 | 0.01 | 0.002 | 4.12E-10 | 0.001 | 0.008 | 0.911 | 37.43 |
| rs4673840 | T | C | 0.84 | -0.014 | 0.002 | 5.86E-13 | -0.001 | 0.01 | 0.926 | 52.75 |
| rs4675248 | A | G | 0.4 | -0.009 | 0.001 | 4.50E-11 | 0.009 | 0.007 | 0.206 | 45.08 |
| rs4719944 | T | C | 0.55 | -0.012 | 0.001 | 9.25E-18 | -0.008 | 0.007 | 0.282 | 74.7 |
| rs4726070 | A | G | 0.6 | 0.012 | 0.001 | 1.36E-17 | -0.017 | 0.007 | 0.015 | 75.94 |
| rs4757957 | C | G | 0.69 | 0.013 | 0.002 | 1.79E-18 | -0.006 | 0.008 | 0.411 | 78.62 |
| rs4766424 | C | G | 0.12 | 0.014 | 0.002 | 2.47E-11 | -0.009 | 0.01 | 0.405 | 45.72 |
| rs4778058 | T | C | 0.5 | -0.009 | 0.001 | 4.92E-11 | -0.005 | 0.007 | 0.529 | 43.18 |
| rs4787457 | A | G | 0.63 | 0.016 | 0.002 | 1.23E-28 | 0.008 | 0.007 | 0.285 | 116.64 |
| rs4793090 | A | G | 0.66 | 0.008 | 0.002 | 1.67E-08 | -0.008 | 0.007 | 0.26 | 31.36 |
| rs4810894 | A | G | 0.37 | -0.009 | 0.002 | 4.38E-09 | -0.02 | 0.007 | 0.006 | 32.11 |
| rs4839155 | T | G | 0.77 | 0.012 | 0.002 | 4.66E-12 | -0.003 | 0.008 | 0.675 | 45.76 |
| rs4846724 | A | G | 0.53 | 0.01 | 0.001 | 2.89E-12 | 0.01 | 0.007 | 0.161 | 49 |
| rs4848924 | A | C | 0.71 | -0.012 | 0.002 | 3.56E-14 | -0.001 | 0.008 | 0.882 | 60.84 |
| rs488476 | C | G | 0.62 | -0.011 | 0.002 | 8.29E-15 | 0.018 | 0.007 | 0.012 | 56.75 |
| rs4895650 | T | C | 0.55 | 0.009 | 0.001 | 5.31E-11 | -0.01 | 0.007 | 0.177 | 44.13 |
| rs4899012 | C | G | 0.6 | -0.01 | 0.001 | 8.06E-13 | -0.001 | 0.007 | 0.924 | 54.13 |
| rs4904523 | A | G | 0.52 | -0.008 | 0.001 | 6.27E-09 | 0.017 | 0.007 | 0.015 | 34.31 |
| rs4915735 | A | G | 0.14 | 0.012 | 0.002 | 9.56E-09 | 0.006 | 0.01 | 0.572 | 33.64 |
| rs4977885 | A | G | 0.4 | -0.008 | 0.001 | 6.78E-09 | -0.001 | 0.007 | 0.888 | 36 |
| rs4984541 | A | G | 0.77 | -0.014 | 0.002 | 3.40E-16 | -0.003 | 0.008 | 0.718 | 65.9 |
| rs55736314 | C | G | 0.6 | -0.015 | 0.001 | 4.59E-27 | -0.003 | 0.007 | 0.709 | 121 |
| rs56099375 | T | C | 0.24 | 0.012 | 0.002 | 3.10E-13 | -0.008 | 0.008 | 0.317 | 49.83 |
| rs56319902 | T | C | 0.22 | -0.021 | 0.002 | 5.81E-33 | 0.004 | 0.008 | 0.664 | 146.84 |
| rs56391344 | A | G | 0.25 | 0.015 | 0.002 | 7.49E-20 | 0.016 | 0.008 | 0.048 | 85.56 |
| rs56794817 | A | G | 0.16 | 0.012 | 0.002 | 1.69E-10 | -0.011 | 0.01 | 0.265 | 41.23 |
| rs57349798 | A | G | 0.41 | 0.009 | 0.001 | 8.63E-11 | 0.003 | 0.007 | 0.707 | 44.13 |
| rs57352738 | A | T | 0.2 | -0.015 | 0.002 | 2.51E-18 | -0.014 | 0.009 | 0.098 | 79.94 |
| rs575113 | A | G | 0.29 | 0.013 | 0.002 | 7.52E-18 | 0.012 | 0.008 | 0.11 | 78.62 |
| rs5754753 | T | C | 0.72 | -0.013 | 0.002 | 6.15E-16 | 0.011 | 0.008 | 0.136 | 62.02 |
| rs57661533 | T | C | 0.13 | -0.011 | 0.002 | 3.24E-08 | -0.009 | 0.01 | 0.405 | 29.47 |
| rs580652 | T | C | 0.9 | -0.015 | 0.002 | 4.65E-10 | -0.01 | 0.012 | 0.41 | 37.52 |
| rs590013 | T | C | 0.68 | 0.01 | 0.002 | 2.47E-11 | 0.009 | 0.007 | 0.251 | 44.44 |
| rs6043521 | T | C | 0.6 | -0.008 | 0.001 | 9.72E-09 | -0.006 | 0.007 | 0.39 | 35.15 |
| rs6065080 | T | C | 0.36 | -0.013 | 0.002 | 7.49E-19 | -0.006 | 0.007 | 0.399 | 75.11 |
| rs6065784 | C | G | 0.7 | 0.011 | 0.002 | 5.78E-13 | 0.004 | 0.008 | 0.563 | 53.78 |
| rs61104616 | A | G | 0.52 | -0.017 | 0.001 | 1.56E-34 | -0.002 | 0.007 | 0.815 | 150.94 |
| rs6123924 | A | G | 0.84 | 0.013 | 0.002 | 3.20E-11 | -0.018 | 0.01 | 0.068 | 46.1 |
| rs61527214 | A | G | 0.41 | 0.011 | 0.001 | 1.13E-14 | 0.011 | 0.007 | 0.134 | 61.73 |
| rs61997667 | T | C | 0.15 | -0.014 | 0.002 | 2.24E-13 | 0.02 | 0.01 | 0.037 | 51.84 |
| rs62155350 | A | G | 0.02 | 0.034 | 0.005 | 1.69E-10 | -0.021 | 0.025 | 0.397 | 40.67 |
| rs62155873 | T | C | 0.12 | -0.015 | 0.002 | 1.65E-12 | -0.026 | 0.011 | 0.014 | 51.7 |
| rs62174974 | A | G | 0.19 | -0.011 | 0.002 | 2.76E-09 | 0.001 | 0.009 | 0.893 | 34.03 |
| rs62179650 | A | G | 0.3 | 0.011 | 0.002 | 5.66E-13 | -0.005 | 0.008 | 0.485 | 49 |
| rs62182994 | T | C | 0.69 | 0.014 | 0.002 | 2.55E-19 | -0.002 | 0.007 | 0.75 | 83.42 |
| rs62379838 | T | C | 0.7 | 0.012 | 0.002 | 1.77E-15 | 0.003 | 0.008 | 0.703 | 65.07 |
| rs628993 | A | G | 0.11 | 0.013 | 0.002 | 3.83E-08 | 0.002 | 0.012 | 0.898 | 30.97 |
| rs6449503 | A | G | 0.5 | 0.018 | 0.001 | 1.84E-39 | 0.014 | 0.007 | 0.053 | 174.62 |
| rs6490618 | T | C | 0.33 | 0.009 | 0.002 | 3.48E-09 | -0.004 | 0.007 | 0.636 | 34.42 |
| rs6493265 | T | C | 0.39 | -0.012 | 0.001 | 2.26E-17 | -0.01 | 0.007 | 0.174 | 75.94 |
| rs6557171 | T | C | 0.32 | -0.016 | 0.002 | 5.48E-25 | 0.009 | 0.007 | 0.224 | 106.78 |
| rs660001 | A | G | 0.21 | -0.016 | 0.002 | 1.66E-21 | 0.003 | 0.009 | 0.743 | 93.07 |
| rs66568921 | T | G | 0.64 | -0.018 | 0.002 | 7.60E-34 | 0.014 | 0.007 | 0.053 | 145.6 |
| rs66721975 | A | G | 0.29 | -0.009 | 0.002 | 1.51E-08 | 0.002 | 0.008 | 0.836 | 33.64 |
| rs6690195 | T | C | 0.49 | -0.011 | 0.001 | 7.89E-16 | -0.001 | 0.007 | 0.938 | 65.15 |
| rs6697584 | T | C | 0.79 | -0.013 | 0.002 | 1.43E-13 | 0.005 | 0.008 | 0.528 | 54.93 |
| rs6704768 | A | G | 0.56 | -0.012 | 0.001 | 1.84E-16 | -0.001 | 0.007 | 0.833 | 68.65 |
| rs6731373 | A | G | 0.34 | -0.012 | 0.002 | 1.39E-14 | 0.002 | 0.007 | 0.744 | 58.78 |
| rs6812533 | T | C | 0.74 | 0.011 | 0.002 | 4.85E-11 | 0.019 | 0.008 | 0.016 | 43.89 |
| rs68145588 | T | G | 0.14 | -0.013 | 0.002 | 7.53E-10 | -0.004 | 0.01 | 0.714 | 36.57 |
| rs6917154 | T | C | 0.12 | 0.013 | 0.002 | 2.55E-09 | -0.017 | 0.01 | 0.094 | 36.57 |
| rs6924023 | A | G | 0.75 | 0.011 | 0.002 | 5.17E-11 | -0.013 | 0.008 | 0.111 | 44.72 |
| rs6977237 | T | C | 0.47 | 0.008 | 0.001 | 5.59E-09 | 0.002 | 0.007 | 0.778 | 34.31 |
| rs6994287 | A | G | 0.41 | 0.01 | 0.001 | 1.11E-12 | 0.001 | 0.007 | 0.892 | 53.08 |
| rs7012546 | T | C | 0.42 | 0.009 | 0.001 | 1.25E-10 | 0.01 | 0.007 | 0.153 | 43.18 |
| rs7029718 | A | G | 0.41 | 0.024 | 0.001 | 7.82E-65 | -0.001 | 0.007 | 0.903 | 303.76 |
| rs7041702 | A | G | 0.74 | -0.011 | 0.002 | 4.53E-12 | -0.007 | 0.008 | 0.354 | 48.13 |
| rs7167688 | T | C | 0.49 | -0.009 | 0.001 | 1.71E-11 | 0.003 | 0.007 | 0.643 | 45.08 |
| rs7171405 | A | G | 0.24 | -0.01 | 0.002 | 7.53E-10 | 0.011 | 0.008 | 0.167 | 39.85 |
| rs717996 | T | C | 0.6 | -0.012 | 0.001 | 1.89E-17 | -0.003 | 0.007 | 0.632 | 75.94 |
| rs7226824 | T | C | 0.53 | -0.008 | 0.001 | 2.72E-08 | 0.013 | 0.007 | 0.063 | 31.04 |
| rs72486027 | T | C | 0.25 | -0.01 | 0.002 | 1.61E-10 | 0.002 | 0.008 | 0.846 | 42.25 |
| rs7254263 | T | C | 0.28 | -0.012 | 0.002 | 1.84E-14 | 0.003 | 0.008 | 0.727 | 55.32 |
| rs72622559 | T | C | 0.23 | -0.012 | 0.002 | 5.50E-13 | 0.011 | 0.008 | 0.17 | 49.83 |
| rs72672052 | A | T | 0.18 | -0.01 | 0.002 | 4.87E-08 | -0.004 | 0.009 | 0.632 | 31.48 |
| rs72709560 | A | G | 0.69 | 0.009 | 0.002 | 2.03E-08 | -0.004 | 0.007 | 0.613 | 32.11 |
| rs72828517 | T | C | 0.83 | -0.017 | 0.002 | 1.38E-19 | 0.008 | 0.009 | 0.363 | 78.18 |
| rs72829857 | A | G | 0.76 | -0.015 | 0.002 | 1.52E-19 | 0.011 | 0.008 | 0.186 | 86.72 |
| rs72917504 | T | C | 0.06 | -0.018 | 0.003 | 2.91E-09 | 0 | 0.015 | 0.978 | 35.2 |
| rs72944064 | T | C | 0.74 | -0.01 | 0.002 | 9.39E-10 | -0.009 | 0.008 | 0.272 | 37.52 |
| rs72962169 | T | C | 0.16 | -0.017 | 0.002 | 1.26E-19 | 0.017 | 0.01 | 0.073 | 83.87 |
| rs72993796 | T | C | 0.88 | 0.013 | 0.002 | 1.12E-08 | -0.005 | 0.011 | 0.671 | 33.32 |
| rs730384 | A | G | 0.44 | 0.01 | 0.001 | 9.38E-13 | 0.009 | 0.007 | 0.193 | 52.05 |
| rs73055556 | A | G | 0.14 | 0.011 | 0.002 | 4.95E-08 | 0.02 | 0.01 | 0.046 | 30.8 |
| rs73191311 | A | G | 0.34 | -0.009 | 0.002 | 2.03E-10 | -0.001 | 0.007 | 0.848 | 39.27 |
| rs7321274 | A | G | 0.8 | 0.011 | 0.002 | 8.21E-11 | 0.017 | 0.009 | 0.048 | 40.11 |
| rs7326331 | A | G | 0.28 | -0.013 | 0.002 | 3.42E-16 | 0.011 | 0.008 | 0.15 | 64 |
| rs73344830 | A | G | 0.42 | 0.017 | 0.001 | 1.00E-32 | 0.004 | 0.007 | 0.596 | 147.45 |
| rs736281 | T | C | 0.39 | 0.009 | 0.001 | 2.49E-09 | -0.004 | 0.007 | 0.573 | 37.73 |
| rs73648455 | T | C | 0.08 | -0.018 | 0.003 | 1.99E-12 | -0.01 | 0.013 | 0.445 | 49.54 |
| rs743316 | T | C | 0.79 | 0.011 | 0.002 | 8.21E-11 | 0 | 0.009 | 0.967 | 43.4 |
| rs74415461 | T | C | 0.08 | 0.018 | 0.003 | 6.25E-12 | -0.037 | 0.012 | 0.003 | 45.82 |
| rs7449561 | A | G | 0.23 | 0.01 | 0.002 | 9.73E-10 | 0.002 | 0.008 | 0.775 | 36 |
| rs7460106 | T | C | 0.76 | -0.012 | 0.002 | 2.06E-13 | 0.009 | 0.008 | 0.295 | 52.35 |
| rs746839 | C | G | 0.63 | 0.014 | 0.002 | 1.19E-20 | -0.008 | 0.007 | 0.282 | 81 |
| rs74787922 | A | C | 0.93 | 0.018 | 0.003 | 8.97E-11 | -0.002 | 0.014 | 0.868 | 42.25 |
| rs75033012 | C | G | 0.04 | -0.026 | 0.004 | 1.38E-11 | -0.002 | 0.018 | 0.9 | 46.1 |
| rs75177132 | T | C | 0.05 | 0.03 | 0.004 | 1.01E-16 | -0.01 | 0.016 | 0.544 | 68.52 |
| rs7575637 | A | G | 0.45 | 0.011 | 0.001 | 3.43E-14 | -0.007 | 0.007 | 0.295 | 58.41 |
| rs7603132 | A | G | 0.19 | 0.014 | 0.002 | 1.20E-14 | -0.008 | 0.009 | 0.353 | 57.93 |
| rs76076331 | T | C | 0.13 | 0.021 | 0.002 | 1.71E-23 | 0.02 | 0.011 | 0.056 | 96.23 |
| rs7617204 | A | G | 0.52 | -0.01 | 0.001 | 4.33E-12 | 0.013 | 0.007 | 0.065 | 48.01 |
| rs76235882 | A | G | 0.97 | 0.024 | 0.004 | 4.52E-10 | -0.016 | 0.021 | 0.463 | 38.19 |
| rs76246107 | A | G | 0.09 | -0.016 | 0.003 | 6.52E-10 | 0.018 | 0.014 | 0.177 | 38.82 |
| rs7650602 | T | C | 0.56 | -0.009 | 0.001 | 2.77E-10 | -0.007 | 0.007 | 0.351 | 40.41 |
| rs76577427 | C | G | 0.9 | 0.02 | 0.002 | 7.78E-18 | -0.012 | 0.011 | 0.303 | 71.54 |
| rs7672622 | A | G | 0.75 | -0.009 | 0.002 | 4.72E-08 | 0.002 | 0.008 | 0.801 | 30.25 |
| rs7683416 | T | C | 0.46 | 0.014 | 0.001 | 6.05E-23 | -0.003 | 0.007 | 0.689 | 98.58 |
| rs76878669 | C | G | 0.76 | 0.014 | 0.002 | 3.10E-17 | 0.003 | 0.008 | 0.755 | 68.79 |
| rs7692359 | T | C | 0.78 | -0.011 | 0.002 | 1.40E-10 | 0.03 | 0.009 | 4.84E-04 | 41.11 |
| rs77025239 | A | G | 0.16 | -0.014 | 0.002 | 4.65E-13 | 0.01 | 0.01 | 0.301 | 54.29 |
| rs77609760 | A | G | 0.93 | 0.024 | 0.003 | 7.71E-17 | 0.003 | 0.014 | 0.848 | 71.04 |
| rs77719387 | A | T | 0.02 | -0.04 | 0.006 | 3.78E-12 | 0.009 | 0.03 | 0.765 | 48.28 |
| rs7849487 | T | G | 0.66 | -0.017 | 0.002 | 1.25E-29 | 0.005 | 0.007 | 0.528 | 123.95 |
| rs78648104 | T | C | 0.91 | -0.014 | 0.002 | 1.70E-08 | -0.006 | 0.012 | 0.606 | 31.81 |
| rs7875078 | A | C | 0.46 | -0.008 | 0.001 | 2.72E-08 | -0.009 | 0.007 | 0.206 | 31.04 |
| rs7894722 | T | C | 0.37 | 0.009 | 0.001 | 5.41E-09 | -0.005 | 0.007 | 0.46 | 36.86 |
| rs790647 | A | C | 0.23 | -0.015 | 0.002 | 3.81E-18 | 0.014 | 0.008 | 0.083 | 72.75 |
| rs7910403 | T | G | 0.2 | 0.012 | 0.002 | 5.08E-12 | 0.01 | 0.009 | 0.27 | 45.94 |
| rs7924036 | T | G | 0.51 | 0.013 | 0.001 | 2.55E-21 | -0.011 | 0.007 | 0.129 | 90.25 |
| rs79265434 | A | G | 0.88 | -0.02 | 0.002 | 1.98E-19 | -0.006 | 0.011 | 0.591 | 80.18 |
| rs7972246 | T | C | 0.35 | 0.011 | 0.002 | 1.01E-13 | -0.017 | 0.007 | 0.021 | 53.78 |
| rs79728014 | A | G | 0.86 | -0.013 | 0.002 | 2.06E-11 | 0.014 | 0.01 | 0.166 | 44.89 |
| rs7974852 | A | C | 0.52 | 0.012 | 0.001 | 1.14E-16 | 0.004 | 0.007 | 0.559 | 69.84 |
| rs7977614 | A | G | 0.71 | -0.013 | 0.002 | 6.84E-17 | -0.005 | 0.008 | 0.506 | 70.14 |
| rs79994730 | T | C | 0.04 | 0.024 | 0.004 | 5.88E-12 | -0.013 | 0.016 | 0.414 | 47.02 |
| rs8008382 | T | C | 0.3 | -0.011 | 0.002 | 4.07E-12 | 0.019 | 0.008 | 0.013 | 49.94 |
| rs8024 | A | C | 0.33 | -0.012 | 0.002 | 1.13E-14 | 0.01 | 0.007 | 0.16 | 58.78 |
| rs80257979 | T | G | 0.97 | -0.025 | 0.004 | 3.31E-10 | 0.006 | 0.02 | 0.751 | 39.69 |
| rs8030487 | A | G | 0.69 | 0.009 | 0.002 | 2.52E-08 | -0.011 | 0.008 | 0.147 | 32.11 |
| rs806816 | A | T | 0.75 | 0.01 | 0.002 | 4.80E-10 | 0.009 | 0.008 | 0.24 | 39.85 |
| rs8097125 | T | C | 0.38 | 0.009 | 0.001 | 2.31E-09 | -0.016 | 0.007 | 0.026 | 37.73 |
| rs912883 | T | C | 0.68 | 0.009 | 0.002 | 6.20E-09 | 0.007 | 0.007 | 0.367 | 33.64 |
| rs9289300 | T | C | 0.84 | -0.016 | 0.002 | 1.00E-17 | 0.006 | 0.01 | 0.503 | 74.5 |
| rs929511 | T | C | 0.13 | -0.016 | 0.002 | 3.91E-14 | 0.003 | 0.011 | 0.806 | 58.05 |
| rs9359939 | A | C | 0.24 | -0.011 | 0.002 | 1.51E-10 | -0.002 | 0.008 | 0.824 | 43.07 |
| rs936496 | A | G | 0.62 | -0.008 | 0.001 | 4.38E-08 | -0.005 | 0.007 | 0.467 | 31.84 |
| rs9373363 | A | G | 0.75 | -0.011 | 0.002 | 5.54E-12 | -0.006 | 0.008 | 0.484 | 48.13 |
| rs9375188 | T | C | 0.48 | 0.021 | 0.001 | 8.78E-52 | -0.005 | 0.007 | 0.468 | 229.31 |
| rs9411331 | A | C | 0.68 | 0.015 | 0.002 | 7.63E-22 | -0.006 | 0.007 | 0.458 | 93.44 |
| rs9492774 | C | G | 0.37 | 0.008 | 0.002 | 4.33E-08 | 0.005 | 0.007 | 0.533 | 28.44 |
| rs9529146 | T | C | 0.23 | 0.013 | 0.002 | 3.48E-14 | -0.003 | 0.008 | 0.716 | 55.81 |
| rs9556958 | T | C | 0.52 | -0.011 | 0.001 | 5.96E-15 | 0.002 | 0.007 | 0.724 | 61.73 |
| rs9616947 | T | C | 0.38 | -0.013 | 0.002 | 3.58E-20 | 0.004 | 0.007 | 0.597 | 79.8 |
| rs9633970 | T | C | 0.76 | 0.009 | 0.002 | 2.81E-08 | 0.003 | 0.009 | 0.692 | 32.35 |
| rs9655780 | A | G | 0.83 | -0.015 | 0.002 | 3.70E-16 | -0.006 | 0.009 | 0.505 | 64.84 |
| rs969512 | A | T | 0.66 | -0.011 | 0.002 | 9.84E-13 | 0 | 0.007 | 0.994 | 49.94 |
| rs9853928 | T | C | 0.21 | -0.013 | 0.002 | 1.59E-13 | 0.009 | 0.009 | 0.294 | 55.81 |
| rs9859556 | T | G | 0.31 | 0.029 | 0.002 | 4.61E-82 | -0.012 | 0.008 | 0.103 | 373.78 |
| rs9882532 | T | C | 0.64 | 0.012 | 0.002 | 2.15E-17 | 0.001 | 0.007 | 0.945 | 68.34 |
| rs9886703 | A | T | 0.17 | -0.014 | 0.002 | 4.46E-13 | -0.001 | 0.009 | 0.887 | 51.24 |
| rs9927137 | A | G | 0.53 | 0.008 | 0.001 | 5.11E-09 | 0.01 | 0.007 | 0.167 | 34.31 |
| rs9927842 | T | C | 0.15 | -0.013 | 0.002 | 6.22E-11 | 0.003 | 0.01 | 0.732 | 42.25 |
| rs9929556 | T | G | 0.57 | -0.01 | 0.001 | 2.31E-12 | 0.002 | 0.007 | 0.777 | 50.01 |
| rs9929762 | A | G | 0.56 | 0.01 | 0.001 | 1.03E-11 | -0.009 | 0.007 | 0.213 | 47.02 |
| rs9933256 | A | G | 0.56 | 0.012 | 0.001 | 6.06E-17 | 0 | 0.007 | 0.992 | 72.25 |

**SNPs used as instrumental variables in the MR analysis of educational attainment on intrinsic curvature**

| **SNP** | **Effect allele** | **Other allele** | **Effect allele frequency** | **beta (exposure)** | **SE (exposure)** | ***P* (exposure)** | **beta (outcome)** | **SE (outcome)** | ***P* (outcome)** | ***F*-statistic** |
| --- | --- | --- | --- | --- | --- | --- | --- | --- | --- | --- |
| rs10006235 | T | C | 0.27 | -0.009 | 0.002 | 4.48E-09 | -0.008 | 0.005 | 0.123 | 33.79 |
| rs10021733 | T | C | 0.15 | -0.016 | 0.003 | 9.45E-10 | -0.004 | 0.007 | 0.5 | 36.89 |
| rs10046069 | A | G | 0.12 | 0.013 | 0.002 | 4.40E-09 | -0.003 | 0.008 | 0.716 | 33.32 |
| rs10060023 | T | C | 0.33 | 0.011 | 0.002 | 1.88E-13 | 0.001 | 0.005 | 0.842 | 53.78 |
| rs10080647 | A | C | 0.14 | 0.012 | 0.002 | 7.05E-10 | -0.001 | 0.007 | 0.881 | 38.44 |
| rs1008078 | T | C | 0.4 | -0.018 | 0.001 | 4.50E-37 | -0.005 | 0.005 | 0.349 | 169 |
| rs10189857 | A | G | 0.57 | 0.016 | 0.001 | 4.69E-29 | 0.002 | 0.005 | 0.645 | 127.37 |
| rs10193498 | A | T | 0.76 | 0.01 | 0.002 | 3.10E-09 | 0.001 | 0.006 | 0.853 | 36.75 |
| rs10215082 | A | G | 0.47 | -0.015 | 0.001 | 7.03E-26 | -0.017 | 0.005 | 5.71E-04 | 113.27 |
| rs10460095 | A | G | 0.57 | -0.012 | 0.001 | 2.61E-18 | -0.004 | 0.005 | 0.451 | 78.45 |
| rs10761251 | A | T | 0.66 | 0.011 | 0.002 | 2.52E-14 | 0.011 | 0.005 | 0.028 | 56.75 |
| rs10772644 | C | G | 0.89 | 0.014 | 0.002 | 1.69E-10 | 0.005 | 0.008 | 0.511 | 41.66 |
| rs10773002 | A | T | 0.25 | 0.018 | 0.002 | 2.50E-30 | 0.01 | 0.006 | 0.073 | 133.69 |
| rs10773208 | T | C | 0.25 | -0.011 | 0.002 | 8.09E-12 | 0.011 | 0.006 | 0.056 | 47.27 |
| rs10798888 | T | G | 0.17 | -0.014 | 0.002 | 5.15E-14 | 0.005 | 0.006 | 0.444 | 54.29 |
| rs10799615 | A | G | 0.25 | -0.009 | 0.002 | 4.11E-08 | -0.012 | 0.006 | 0.028 | 30.94 |
| rs10805383 | A | G | 0.49 | -0.01 | 0.001 | 2.22E-13 | -0.008 | 0.005 | 0.089 | 54.13 |
| rs10830858 | T | C | 0.51 | 0.009 | 0.001 | 5.31E-11 | 0.003 | 0.005 | 0.514 | 43.18 |
| rs10844179 | A | G | 0.23 | 0.01 | 0.002 | 5.03E-10 | -0.012 | 0.006 | 0.046 | 37.43 |
| rs10875121 | C | G | 0.84 | 0.017 | 0.002 | 4.46E-20 | -0.007 | 0.007 | 0.318 | 83.87 |
| rs10892807 | T | C | 0.57 | 0.011 | 0.001 | 1.09E-14 | -0.005 | 0.005 | 0.336 | 60.62 |
| rs10940921 | T | G | 0.42 | 0.011 | 0.001 | 1.31E-14 | 0.001 | 0.005 | 0.813 | 62.86 |
| rs10951590 | T | C | 0.33 | -0.011 | 0.002 | 3.62E-13 | 0.004 | 0.005 | 0.492 | 52.8 |
| rs10979613 | T | C | 0.64 | -0.012 | 0.002 | 1.25E-15 | -0.004 | 0.005 | 0.413 | 60.84 |
| rs10996167 | C | G | 0.65 | 0.011 | 0.002 | 7.47E-09 | 0.005 | 0.005 | 0.337 | 31.92 |
| rs11003463 | T | G | 0.61 | 0.009 | 0.002 | 5.58E-10 | 0.006 | 0.005 | 0.2 | 36.8 |
| rs11023764 | A | G | 0.64 | 0.009 | 0.002 | 5.32E-10 | 0.006 | 0.005 | 0.261 | 36.8 |
| rs11030102 | C | G | 0.75 | 0.01 | 0.002 | 4.60E-10 | 0.011 | 0.006 | 0.038 | 39.85 |
| rs11076962 | T | C | 0.72 | 0.009 | 0.002 | 4.29E-08 | 0.002 | 0.005 | 0.694 | 28.89 |
| rs11081529 | T | C | 0.71 | 0.013 | 0.002 | 5.22E-16 | 0.007 | 0.005 | 0.188 | 70.56 |
| rs11082011 | T | C | 0.67 | 0.019 | 0.002 | 2.06E-38 | 0.009 | 0.005 | 0.078 | 167.27 |
| rs11121177 | A | G | 0.18 | 0.015 | 0.002 | 1.44E-16 | 0.002 | 0.006 | 0.773 | 70.37 |
| rs11130380 | T | G | 0.61 | 0.009 | 0.001 | 4.17E-10 | 0.007 | 0.005 | 0.146 | 41.33 |
| rs11138947 | T | C | 0.72 | 0.011 | 0.002 | 2.36E-11 | -0.007 | 0.005 | 0.198 | 43.07 |
| rs11157931 | A | C | 0.39 | -0.014 | 0.001 | 3.86E-22 | -0.028 | 0.005 | 1.31E-08 | 98.58 |
| rs111852224 | T | C | 0.12 | 0.016 | 0.002 | 4.85E-13 | 0.001 | 0.007 | 0.938 | 50.28 |
| rs11211123 | A | G | 0.22 | 0.01 | 0.002 | 7.62E-10 | -0.011 | 0.006 | 0.053 | 37.43 |
| rs11213482 | A | G | 0.84 | -0.011 | 0.002 | 2.68E-08 | 0.003 | 0.006 | 0.673 | 31.12 |
| rs112603734 | A | C | 0.77 | -0.013 | 0.002 | 2.72E-09 | 0.005 | 0.006 | 0.4 | 35.46 |
| rs112806496 | C | G | 0.91 | -0.016 | 0.002 | 1.72E-10 | -0.01 | 0.008 | 0.251 | 40.96 |
| rs1128956 | T | G | 0.83 | -0.014 | 0.002 | 5.34E-14 | 0.002 | 0.006 | 0.746 | 54.29 |
| rs113520408 | A | G | 0.28 | 0.016 | 0.002 | 1.03E-22 | 0.001 | 0.005 | 0.868 | 93.85 |
| rs113615161 | T | C | 0.13 | -0.012 | 0.002 | 2.98E-09 | -0.001 | 0.007 | 0.927 | 34.31 |
| rs114593137 | A | T | 0.79 | -0.012 | 0.002 | 1.65E-12 | -0.013 | 0.006 | 0.025 | 51.5 |
| rs115000530 | A | T | 0.95 | -0.025 | 0.003 | 1.09E-15 | -0.01 | 0.011 | 0.353 | 65.04 |
| rs11542663 | A | C | 0.69 | 0.011 | 0.002 | 2.44E-13 | 0.003 | 0.005 | 0.553 | 55.75 |
| rs11588857 | A | G | 0.21 | 0.02 | 0.002 | 4.45E-31 | 0.015 | 0.006 | 0.012 | 137.03 |
| rs11598765 | A | G | 0.2 | -0.011 | 0.002 | 1.73E-09 | 0 | 0.006 | 0.947 | 34.68 |
| rs11620355 | A | G | 0.09 | 0.018 | 0.002 | 1.09E-13 | 0.006 | 0.009 | 0.497 | 53 |
| rs11657342 | A | G | 0.37 | 0.014 | 0.002 | 2.32E-13 | 0.008 | 0.005 | 0.091 | 54.29 |
| rs11657979 | A | G | 0.24 | -0.009 | 0.002 | 4.57E-08 | 0.002 | 0.006 | 0.775 | 28.65 |
| rs11663602 | A | C | 0.28 | -0.013 | 0.002 | 9.27E-17 | -0.003 | 0.005 | 0.616 | 66.02 |
| rs1167827 | A | G | 0.43 | 0.01 | 0.001 | 2.93E-13 | -0.008 | 0.005 | 0.111 | 55.18 |
| rs11724690 | T | G | 0.29 | 0.009 | 0.002 | 2.08E-09 | 0.001 | 0.005 | 0.923 | 38.44 |
| rs117398064 | C | G | 0.09 | -0.014 | 0.002 | 1.16E-08 | 0.005 | 0.008 | 0.544 | 31.81 |
| rs118134876 | T | C | 0.06 | -0.022 | 0.003 | 1.50E-13 | -0.009 | 0.01 | 0.368 | 55.25 |
| rs11871429 | A | G | 0.77 | 0.013 | 0.002 | 2.52E-14 | 0.012 | 0.006 | 0.046 | 56.69 |
| rs11894424 | A | C | 0.95 | 0.021 | 0.003 | 2.14E-10 | 0.005 | 0.012 | 0.652 | 40.11 |
| rs12054166 | C | G | 0.74 | 0.01 | 0.002 | 2.30E-09 | 0.009 | 0.006 | 0.099 | 36 |
| rs12076635 | C | G | 0.78 | 0.021 | 0.002 | 3.33E-34 | 0.01 | 0.006 | 0.1 | 148.27 |
| rs12113634 | T | C | 0.65 | 0.009 | 0.002 | 2.50E-09 | 0.006 | 0.005 | 0.255 | 34.42 |
| rs12151248 | T | C | 0.11 | -0.015 | 0.002 | 3.81E-11 | 0.018 | 0.008 | 0.016 | 41.97 |
| rs1220779 | A | G | 0.47 | -0.008 | 0.001 | 1.58E-08 | 0.005 | 0.005 | 0.295 | 31.84 |
| rs12273435 | A | G | 0.21 | -0.013 | 0.002 | 4.72E-14 | -0.004 | 0.006 | 0.494 | 60.29 |
| rs12332731 | A | T | 0.19 | 0.014 | 0.002 | 5.96E-14 | 0.012 | 0.006 | 0.06 | 56.25 |
| rs12375949 | T | C | 0.43 | -0.014 | 0.001 | 1.61E-22 | 0 | 0.005 | 0.93 | 97.16 |
| rs12438177 | A | G | 0.37 | 0.009 | 0.002 | 1.48E-10 | 0.016 | 0.005 | 0.002 | 38.44 |
| rs12468040 | T | G | 0.38 | 0.014 | 0.001 | 3.12E-21 | 0.007 | 0.005 | 0.162 | 95.76 |
| rs12503522 | T | C | 0.28 | -0.011 | 0.002 | 3.93E-13 | 0 | 0.005 | 0.98 | 49.88 |
| rs12506221 | T | G | 0.44 | -0.013 | 0.001 | 4.23E-20 | -0.002 | 0.005 | 0.674 | 86.22 |
| rs12519073 | T | C | 0.23 | -0.011 | 0.002 | 1.16E-10 | 0.01 | 0.006 | 0.101 | 39.62 |
| rs12524795 | T | C | 0.44 | 0.01 | 0.001 | 5.28E-13 | -0.01 | 0.005 | 0.034 | 53.08 |
| rs12571549 | A | G | 0.14 | 0.015 | 0.002 | 1.45E-13 | -0.007 | 0.007 | 0.291 | 57 |
| rs12574281 | A | C | 0.63 | -0.008 | 0.002 | 3.04E-08 | 0.004 | 0.005 | 0.399 | 28.44 |
| rs12591647 | T | C | 0.82 | -0.015 | 0.002 | 7.78E-17 | -0.02 | 0.006 | 0.001 | 71.31 |
| rs12602286 | T | G | 0.87 | 0.016 | 0.002 | 4.62E-14 | -0.022 | 0.007 | 0.003 | 58.05 |
| rs12614263 | A | G | 0.5 | 0.008 | 0.001 | 3.38E-09 | 0.009 | 0.005 | 0.054 | 35.15 |
| rs12638072 | A | G | 0.3 | -0.011 | 0.002 | 3.13E-12 | -0.007 | 0.005 | 0.217 | 50.88 |
| rs12643771 | T | C | 0.31 | 0.013 | 0.002 | 1.32E-17 | 0.013 | 0.005 | 0.016 | 75.11 |
| rs12646523 | T | C | 0.25 | -0.013 | 0.002 | 3.79E-16 | -0.004 | 0.006 | 0.435 | 68.06 |
| rs12670376 | A | G | 0.45 | 0.009 | 0.001 | 5.99E-11 | 0.001 | 0.005 | 0.767 | 43.18 |
| rs12761761 | T | C | 0.24 | 0.015 | 0.002 | 4.79E-20 | 0 | 0.006 | 0.947 | 81 |
| rs12765185 | A | T | 0.28 | -0.009 | 0.002 | 2.96E-08 | -0.011 | 0.005 | 0.038 | 30.25 |
| rs12789313 | T | C | 0.49 | 0.009 | 0.001 | 6.79E-11 | 0.003 | 0.005 | 0.558 | 43.18 |
| rs12875339 | A | C | 0.64 | -0.011 | 0.002 | 1.71E-13 | 0.005 | 0.005 | 0.333 | 51.84 |
| rs12888615 | T | C | 0.2 | 0.012 | 0.002 | 9.42E-12 | 0.001 | 0.006 | 0.898 | 44.44 |
| rs12957463 | A | G | 0.8 | 0.015 | 0.002 | 9.20E-17 | 0.013 | 0.006 | 0.028 | 65.79 |
| rs12967010 | T | C | 0.77 | 0.011 | 0.002 | 1.33E-11 | 0.007 | 0.006 | 0.257 | 44.97 |
| rs12981405 | T | C | 0.17 | -0.011 | 0.002 | 2.75E-09 | -0.008 | 0.007 | 0.241 | 34.75 |
| rs1301838 | T | C | 0.32 | -0.01 | 0.002 | 1.29E-10 | 0.003 | 0.005 | 0.619 | 40.96 |
| rs13018640 | T | C | 0.6 | -0.021 | 0.001 | 1.05E-50 | 0.007 | 0.005 | 0.166 | 235.84 |
| rs13050131 | A | C | 0.66 | -0.008 | 0.002 | 4.40E-08 | -0.003 | 0.005 | 0.548 | 29.16 |
| rs13133213 | A | G | 0.5 | 0.01 | 0.001 | 7.57E-12 | 0.002 | 0.005 | 0.638 | 47.02 |
| rs13145650 | T | C | 0.92 | -0.017 | 0.002 | 8.99E-12 | 0.014 | 0.009 | 0.112 | 47.89 |
| rs13163845 | T | C | 0.85 | -0.015 | 0.002 | 4.66E-14 | 0.003 | 0.007 | 0.686 | 55.5 |
| rs13197257 | T | G | 0.28 | 0.011 | 0.002 | 2.83E-12 | -0.005 | 0.005 | 0.339 | 46.41 |
| rs13212041 | T | C | 0.8 | 0.01 | 0.002 | 7.42E-09 | 0.01 | 0.006 | 0.103 | 31.48 |
| rs13240401 | T | C | 0.78 | 0.018 | 0.002 | 2.36E-24 | 0.003 | 0.006 | 0.604 | 107.18 |
| rs13261773 | C | G | 0.16 | 0.012 | 0.002 | 3.12E-10 | 0.008 | 0.007 | 0.227 | 41.23 |
| rs13266287 | A | T | 0.67 | -0.009 | 0.002 | 1.75E-08 | -0.005 | 0.005 | 0.322 | 32.11 |
| rs1329125 | T | C | 0.33 | -0.01 | 0.002 | 8.37E-11 | -0.004 | 0.005 | 0.402 | 41.82 |
| rs13327482 | A | G | 0.82 | -0.012 | 0.002 | 4.47E-11 | 0.018 | 0.006 | 0.004 | 45.19 |
| rs1334297 | A | G | 0.74 | 0.026 | 0.002 | 2.53E-58 | 0.004 | 0.006 | 0.522 | 258 |
| rs13422673 | T | C | 0.47 | -0.011 | 0.001 | 6.48E-15 | -0.004 | 0.005 | 0.431 | 61.73 |
| rs137079 | T | C | 0.14 | 0.013 | 0.002 | 3.06E-10 | 0.008 | 0.007 | 0.226 | 40.96 |
| rs1426619 | T | C | 0.43 | 0.01 | 0.001 | 1.40E-11 | 0.006 | 0.005 | 0.188 | 47.02 |
| rs1434630 | T | G | 0.15 | -0.014 | 0.002 | 4.63E-12 | -0.007 | 0.007 | 0.34 | 46.92 |
| rs143743568 | A | G | 0.15 | 0.012 | 0.002 | 4.99E-09 | 0.019 | 0.007 | 0.006 | 34.81 |
| rs150537577 | A | G | 0.92 | -0.015 | 0.003 | 9.50E-09 | 0.001 | 0.009 | 0.937 | 33.73 |
| rs1505676 | C | G | 0.63 | 0.008 | 0.002 | 6.67E-09 | -0.006 | 0.005 | 0.265 | 31.36 |
| rs1542354 | A | G | 0.48 | -0.008 | 0.001 | 2.72E-09 | 0.012 | 0.005 | 0.017 | 36 |
| rs1544 | A | G | 0.26 | -0.01 | 0.002 | 1.76E-09 | -0.01 | 0.005 | 0.066 | 36 |
| rs1564347 | T | G | 0.34 | 0.009 | 0.002 | 2.39E-09 | -0.003 | 0.005 | 0.523 | 34.42 |
| rs1603460 | T | G | 0.42 | 0.009 | 0.001 | 2.20E-10 | 0.001 | 0.005 | 0.817 | 41.33 |
| rs162445 | A | G | 0.08 | 0.015 | 0.003 | 5.72E-09 | 0.02 | 0.009 | 0.031 | 34.18 |
| rs1689510 | C | G | 0.33 | 0.019 | 0.002 | 1.35E-38 | 0.014 | 0.005 | 0.005 | 167.27 |
| rs17048855 | A | G | 0.34 | 0.011 | 0.002 | 3.10E-13 | 0.01 | 0.005 | 0.039 | 51.84 |
| rs17110109 | T | C | 0.62 | -0.009 | 0.001 | 1.47E-10 | -0.01 | 0.005 | 0.051 | 44.13 |
| rs17144467 | A | C | 0.69 | 0.01 | 0.002 | 8.74E-11 | -0.005 | 0.005 | 0.392 | 42.68 |
| rs17148998 | A | G | 0.2 | 0.01 | 0.002 | 6.98E-09 | 0.016 | 0.006 | 0.01 | 35.3 |
| rs1717204 | A | C | 0.18 | -0.012 | 0.002 | 1.20E-10 | 0.004 | 0.006 | 0.51 | 42.25 |
| rs17248751 | A | G | 0.78 | -0.014 | 0.002 | 1.28E-15 | 0 | 0.006 | 0.949 | 64 |
| rs173003 | A | C | 0.49 | -0.008 | 0.001 | 5.50E-09 | 0 | 0.005 | 0.962 | 34.31 |
| rs1738050 | C | G | 0.62 | -0.009 | 0.001 | 1.42E-10 | -0.006 | 0.005 | 0.202 | 44.13 |
| rs17411339 | A | G | 0.56 | 0.014 | 0.001 | 5.10E-23 | -0.001 | 0.005 | 0.839 | 100 |
| rs17428076 | C | G | 0.76 | 0.014 | 0.002 | 6.41E-17 | 0.004 | 0.006 | 0.523 | 73.32 |
| rs1747714 | T | C | 0.53 | 0.011 | 0.001 | 3.82E-14 | -0.001 | 0.005 | 0.896 | 57.33 |
| rs1747817 | T | C | 0.76 | -0.009 | 0.002 | 2.29E-08 | 0.003 | 0.006 | 0.581 | 33.06 |
| rs17489649 | A | G | 0.68 | 0.013 | 0.002 | 2.77E-17 | 0 | 0.005 | 0.955 | 71.68 |
| rs17502934 | T | G | 0.15 | -0.017 | 0.002 | 2.12E-17 | -0.002 | 0.007 | 0.829 | 69.72 |
| rs175325 | A | T | 0.6 | -0.01 | 0.001 | 2.24E-12 | 0.008 | 0.005 | 0.115 | 51.02 |
| rs17551064 | A | G | 0.84 | 0.014 | 0.002 | 1.53E-13 | 0.007 | 0.007 | 0.327 | 55.07 |
| rs17563464 | A | C | 0.22 | -0.015 | 0.002 | 1.74E-17 | -0.008 | 0.006 | 0.203 | 74.77 |
| rs17565975 | A | G | 0.56 | -0.011 | 0.001 | 5.54E-14 | -0.013 | 0.005 | 0.007 | 57.33 |
| rs17598675 | T | C | 0.51 | -0.012 | 0.001 | 1.37E-16 | -0.002 | 0.005 | 0.757 | 68.65 |
| rs17604349 | A | G | 0.19 | 0.013 | 0.002 | 2.36E-12 | -0.013 | 0.006 | 0.046 | 49.78 |
| rs176218 | T | G | 0.2 | 0.02 | 0.002 | 1.10E-29 | -0.013 | 0.006 | 0.031 | 123.46 |
| rs17732878 | T | C | 0.79 | -0.01 | 0.002 | 4.06E-08 | -0.015 | 0.006 | 0.014 | 31.23 |
| rs178183 | T | C | 0.25 | 0.015 | 0.002 | 4.81E-19 | 0.005 | 0.006 | 0.361 | 82.13 |
| rs17882802 | A | G | 0.43 | 0.008 | 0.001 | 3.00E-09 | -0.005 | 0.005 | 0.284 | 36 |
| rs182902112 | A | C | 0.02 | -0.03 | 0.005 | 1.17E-08 | -0.025 | 0.021 | 0.232 | 32.04 |
| rs1842713 | A | G | 0.79 | -0.012 | 0.002 | 9.67E-12 | 0.005 | 0.006 | 0.386 | 47.37 |
| rs1861786 | A | G | 0.38 | -0.008 | 0.001 | 2.39E-08 | -0.002 | 0.005 | 0.7 | 33.47 |
| rs1865955 | T | C | 0.82 | 0.012 | 0.002 | 4.06E-11 | -0.007 | 0.006 | 0.28 | 41.23 |
| rs1866823 | A | G | 0.54 | 0.011 | 0.001 | 3.09E-14 | -0.004 | 0.005 | 0.465 | 58.41 |
| rs1890132 | T | C | 0.71 | -0.009 | 0.002 | 6.56E-09 | 0.001 | 0.005 | 0.854 | 35.2 |
| rs192436652 | T | C | 0.03 | -0.032 | 0.004 | 2.06E-12 | -0.001 | 0.015 | 0.962 | 50.25 |
| rs1933264 | T | C | 0.75 | 0.01 | 0.002 | 5.85E-10 | 0.002 | 0.006 | 0.749 | 39.06 |
| rs1955250 | A | C | 0.91 | -0.014 | 0.002 | 2.26E-08 | -0.002 | 0.009 | 0.853 | 31.36 |
| rs1991585 | T | C | 0.71 | -0.01 | 0.002 | 6.26E-11 | -0.006 | 0.005 | 0.249 | 45.34 |
| rs2007655 | T | G | 0.5 | 0.009 | 0.001 | 8.64E-10 | 0.005 | 0.005 | 0.263 | 37.73 |
| rs2011603 | A | G | 0.74 | -0.009 | 0.002 | 1.12E-08 | -0.016 | 0.006 | 0.003 | 32.35 |
| rs2034631 | T | C | 0.65 | 0.009 | 0.002 | 2.64E-09 | -0.002 | 0.005 | 0.706 | 35.2 |
| rs2052285 | A | G | 0.6 | 0.011 | 0.001 | 7.25E-15 | 0.017 | 0.005 | 6.57E-04 | 64 |
| rs2055940 | A | G | 0.32 | 0.008 | 0.002 | 4.95E-08 | 0.006 | 0.005 | 0.252 | 29.88 |
| rs2081652 | A | T | 0.66 | 0.012 | 0.002 | 1.09E-16 | 0.012 | 0.005 | 0.018 | 67.24 |
| rs2131167 | A | G | 0.45 | 0.008 | 0.001 | 1.52E-08 | -0.002 | 0.005 | 0.62 | 32.65 |
| rs2179152 | T | C | 0.37 | -0.013 | 0.002 | 1.68E-19 | 0.007 | 0.005 | 0.142 | 76.27 |
| rs2183271 | T | C | 0.64 | 0.008 | 0.002 | 1.91E-08 | 0.026 | 0.005 | 3.38E-07 | 31.36 |
| rs2199409 | T | C | 0.8 | 0.01 | 0.002 | 3.14E-09 | 0.002 | 0.006 | 0.737 | 36.71 |
| rs2212430 | T | C | 0.3 | -0.01 | 0.002 | 1.32E-10 | 0.009 | 0.005 | 0.093 | 39.06 |
| rs2216144 | T | C | 0.53 | 0.01 | 0.001 | 2.58E-12 | -0.01 | 0.005 | 0.045 | 49 |
| rs2256965 | A | G | 0.42 | 0.011 | 0.001 | 2.40E-13 | 0.012 | 0.005 | 0.019 | 57.33 |
| rs2287838 | A | G | 0.55 | -0.01 | 0.001 | 1.27E-13 | 0.003 | 0.005 | 0.487 | 55.18 |
| rs2290601 | T | C | 0.77 | -0.015 | 0.002 | 4.49E-19 | -0.005 | 0.006 | 0.362 | 76.82 |
| rs2297293 | C | G | 0.31 | 0.01 | 0.002 | 8.80E-11 | 0.007 | 0.005 | 0.178 | 43.56 |
| rs232496 | T | C | 0.37 | 0.009 | 0.002 | 7.14E-10 | -0.006 | 0.005 | 0.227 | 36 |
| rs2364544 | A | G | 0.39 | -0.011 | 0.001 | 2.91E-13 | -0.009 | 0.005 | 0.06 | 56.25 |
| rs2434672 | A | C | 0.53 | 0.009 | 0.001 | 1.06E-10 | 0.005 | 0.005 | 0.296 | 42.25 |
| rs2436760 | T | C | 0.97 | 0.023 | 0.004 | 4.52E-08 | 0.013 | 0.014 | 0.36 | 30.51 |
| rs2447097 | T | G | 0.46 | 0.01 | 0.001 | 1.10E-11 | 0.005 | 0.005 | 0.336 | 47.02 |
| rs2469226 | A | T | 0.22 | 0.01 | 0.002 | 1.28E-08 | -0.009 | 0.006 | 0.124 | 31.89 |
| rs2496482 | T | C | 0.42 | 0.012 | 0.002 | 9.20E-17 | -0.021 | 0.005 | 1.80E-05 | 65.07 |
| rs252991 | A | G | 0.37 | 0.01 | 0.002 | 6.30E-11 | 0 | 0.005 | 0.971 | 40.11 |
| rs2545795 | A | C | 0.44 | 0.013 | 0.001 | 1.03E-18 | 0.011 | 0.005 | 0.026 | 79.72 |
| rs2554835 | A | G | 0.4 | 0.009 | 0.001 | 8.95E-10 | -0.009 | 0.005 | 0.074 | 39.51 |
| rs2570497 | T | C | 0.64 | -0.012 | 0.002 | 1.27E-16 | 0.012 | 0.005 | 0.021 | 65.07 |
| rs2725370 | T | C | 0.3 | -0.014 | 0.002 | 4.96E-20 | -0.003 | 0.005 | 0.604 | 88.36 |
| rs2764684 | T | C | 0.83 | 0.014 | 0.002 | 9.99E-15 | 0.02 | 0.006 | 0.002 | 56.65 |
| rs2838006 | T | C | 0.36 | 0.013 | 0.002 | 1.38E-19 | -0.007 | 0.005 | 0.196 | 77.44 |
| rs28513882 | A | G | 0.18 | -0.011 | 0.002 | 2.03E-09 | -0.002 | 0.006 | 0.775 | 37.35 |
| rs2885198 | A | G | 0.53 | 0.009 | 0.001 | 4.63E-10 | 0.009 | 0.005 | 0.06 | 39.51 |
| rs2923424 | A | G | 0.61 | 0.012 | 0.001 | 6.31E-17 | -0.01 | 0.005 | 0.036 | 73.47 |
| rs2929032 | A | G | 0.47 | -0.008 | 0.001 | 8.40E-09 | 0.006 | 0.005 | 0.244 | 33.47 |
| rs2958182 | A | T | 0.34 | 0.008 | 0.002 | 3.46E-08 | -0.006 | 0.005 | 0.217 | 29.88 |
| rs2964199 | T | C | 0.31 | -0.01 | 0.002 | 4.61E-11 | -0.004 | 0.005 | 0.411 | 43.56 |
| rs2964255 | A | G | 0.31 | 0.009 | 0.002 | 6.34E-09 | -0.001 | 0.005 | 0.921 | 34.42 |
| rs2998299 | T | G | 0.21 | -0.014 | 0.002 | 1.51E-15 | 0.004 | 0.006 | 0.545 | 64 |
| rs3026996 | A | C | 0.76 | 0.014 | 0.002 | 5.42E-17 | 0.003 | 0.006 | 0.629 | 74.39 |
| rs303752 | A | G | 0.41 | -0.011 | 0.001 | 1.47E-14 | 0.001 | 0.005 | 0.903 | 61.73 |
| rs3111251 | T | C | 0.43 | -0.009 | 0.001 | 3.88E-10 | -0.001 | 0.005 | 0.815 | 41.33 |
| rs34067381 | T | G | 0.36 | -0.01 | 0.002 | 9.77E-13 | -0.006 | 0.005 | 0.23 | 48.07 |
| rs34098770 | A | G | 0.15 | -0.016 | 0.002 | 2.95E-16 | -0.012 | 0.007 | 0.08 | 64 |
| rs34305371 | A | G | 0.1 | 0.031 | 0.002 | 1.10E-39 | -0.014 | 0.008 | 0.091 | 171.17 |
| rs34394051 | A | G | 0.84 | -0.012 | 0.002 | 2.30E-09 | -0.01 | 0.007 | 0.13 | 34.22 |
| rs34720381 | T | C | 0.09 | -0.018 | 0.002 | 1.90E-13 | 0.014 | 0.008 | 0.096 | 56.25 |
| rs34807077 | A | C | 0.16 | 0.013 | 0.002 | 3.29E-11 | -0.006 | 0.007 | 0.398 | 45.39 |
| rs35417702 | T | C | 0.52 | -0.015 | 0.001 | 2.40E-27 | -0.001 | 0.005 | 0.838 | 117.88 |
| rs35606437 | A | G | 0.27 | 0.011 | 0.002 | 3.98E-11 | 0.018 | 0.006 | 0.001 | 43.07 |
| rs35929923 | A | G | 0.25 | -0.011 | 0.002 | 1.77E-12 | -0.013 | 0.006 | 0.017 | 50.77 |
| rs363096 | T | C | 0.42 | -0.014 | 0.001 | 5.77E-24 | -0.002 | 0.005 | 0.642 | 104.33 |
| rs3751331 | A | G | 0.38 | -0.01 | 0.001 | 1.75E-11 | -0.004 | 0.005 | 0.443 | 48.01 |
| rs3809634 | A | G | 0.68 | -0.011 | 0.002 | 6.82E-13 | -0.006 | 0.005 | 0.278 | 52.8 |
| rs382196 | T | G | 0.38 | -0.008 | 0.001 | 1.47E-08 | -0.004 | 0.005 | 0.392 | 34.31 |
| rs3859523 | T | C | 0.82 | 0.012 | 0.002 | 9.92E-12 | 0.006 | 0.006 | 0.309 | 47.46 |
| rs3897821 | A | G | 0.67 | 0.015 | 0.002 | 3.58E-25 | 0.005 | 0.005 | 0.352 | 105.4 |
| rs3948495 | T | G | 0.41 | -0.009 | 0.001 | 3.95E-10 | -0.008 | 0.005 | 0.119 | 40.41 |
| rs401526 | T | C | 0.53 | -0.01 | 0.001 | 1.15E-13 | -0.012 | 0.005 | 0.014 | 55.18 |
| rs401966 | C | G | 0.39 | -0.01 | 0.001 | 1.72E-11 | -0.005 | 0.005 | 0.287 | 48.01 |
| rs42210 | C | G | 0.71 | -0.01 | 0.002 | 2.74E-10 | -0.001 | 0.005 | 0.847 | 37.52 |
| rs42302 | A | G | 0.34 | 0.009 | 0.002 | 5.96E-09 | 0.005 | 0.005 | 0.331 | 32.87 |
| rs4298514 | T | C | 0.7 | 0.01 | 0.002 | 1.07E-11 | 0.002 | 0.005 | 0.67 | 48.07 |
| rs4320563 | A | G | 0.51 | 0.011 | 0.001 | 2.24E-14 | -0.01 | 0.005 | 0.047 | 60.62 |
| rs4328757 | T | C | 0.61 | 0.01 | 0.001 | 2.95E-11 | -0.006 | 0.005 | 0.199 | 47.02 |
| rs4352658 | T | C | 0.08 | -0.019 | 0.003 | 1.71E-13 | -0.011 | 0.009 | 0.228 | 53.4 |
| rs4358081 | A | C | 0.53 | -0.01 | 0.001 | 1.28E-11 | -0.006 | 0.005 | 0.24 | 46.05 |
| rs4369924 | A | G | 0.16 | 0.013 | 0.002 | 5.58E-11 | 0.009 | 0.007 | 0.203 | 43.28 |
| rs4384309 | A | G | 0.46 | 0.01 | 0.001 | 4.89E-13 | -0.002 | 0.005 | 0.699 | 53.08 |
| rs4467547 | T | G | 0.41 | 0.012 | 0.001 | 4.88E-17 | 0.01 | 0.005 | 0.046 | 74.7 |
| rs4500930 | T | C | 0.34 | -0.011 | 0.002 | 1.67E-13 | -0.001 | 0.005 | 0.805 | 52.8 |
| rs4663617 | A | T | 0.23 | 0.01 | 0.002 | 4.12E-10 | 0.008 | 0.006 | 0.18 | 37.43 |
| rs4673840 | T | C | 0.84 | -0.014 | 0.002 | 5.86E-13 | -0.002 | 0.007 | 0.731 | 52.75 |
| rs4675248 | A | G | 0.4 | -0.009 | 0.001 | 4.50E-11 | 0.009 | 0.005 | 0.064 | 45.08 |
| rs4719944 | T | C | 0.55 | -0.012 | 0.001 | 9.25E-18 | 0.002 | 0.005 | 0.708 | 74.7 |
| rs4726070 | A | G | 0.6 | 0.012 | 0.001 | 1.36E-17 | 0.002 | 0.005 | 0.74 | 75.94 |
| rs4757957 | C | G | 0.69 | 0.013 | 0.002 | 1.79E-18 | 0.011 | 0.005 | 0.029 | 78.62 |
| rs4766424 | C | G | 0.12 | 0.014 | 0.002 | 2.47E-11 | 0.007 | 0.007 | 0.344 | 45.72 |
| rs4778058 | T | C | 0.5 | -0.009 | 0.001 | 4.92E-11 | -0.01 | 0.005 | 0.053 | 43.18 |
| rs4787457 | A | G | 0.63 | 0.016 | 0.002 | 1.23E-28 | 0.017 | 0.005 | 8.04E-04 | 116.64 |
| rs4793090 | A | G | 0.66 | 0.008 | 0.002 | 1.67E-08 | 0.004 | 0.005 | 0.484 | 31.36 |
| rs4810894 | A | G | 0.37 | -0.009 | 0.002 | 4.38E-09 | -0.008 | 0.005 | 0.094 | 32.11 |
| rs4839155 | T | G | 0.77 | 0.012 | 0.002 | 4.66E-12 | -0.007 | 0.006 | 0.233 | 45.76 |
| rs4846724 | A | G | 0.53 | 0.01 | 0.001 | 2.89E-12 | 0 | 0.005 | 0.921 | 49 |
| rs4848924 | A | C | 0.71 | -0.012 | 0.002 | 3.56E-14 | -0.006 | 0.005 | 0.275 | 60.84 |
| rs488476 | C | G | 0.62 | -0.011 | 0.002 | 8.29E-15 | 0.005 | 0.005 | 0.303 | 56.75 |
| rs4895650 | T | C | 0.55 | 0.009 | 0.001 | 5.31E-11 | -0.007 | 0.005 | 0.146 | 44.13 |
| rs4899012 | C | G | 0.6 | -0.01 | 0.001 | 8.06E-13 | -0.012 | 0.005 | 0.015 | 54.13 |
| rs4904523 | A | G | 0.52 | -0.008 | 0.001 | 6.27E-09 | 0.004 | 0.005 | 0.47 | 34.31 |
| rs4915735 | A | G | 0.14 | 0.012 | 0.002 | 9.56E-09 | 0 | 0.007 | 0.988 | 33.64 |
| rs4977885 | A | G | 0.4 | -0.008 | 0.001 | 6.78E-09 | -0.005 | 0.005 | 0.357 | 36 |
| rs4984541 | A | G | 0.77 | -0.014 | 0.002 | 3.40E-16 | -0.001 | 0.006 | 0.864 | 65.9 |
| rs55736314 | C | G | 0.6 | -0.015 | 0.001 | 4.59E-27 | -0.02 | 0.005 | 3.82E-05 | 121 |
| rs56099375 | T | C | 0.24 | 0.012 | 0.002 | 3.10E-13 | -0.008 | 0.006 | 0.153 | 49.83 |
| rs56319902 | T | C | 0.22 | -0.021 | 0.002 | 5.81E-33 | -0.055 | 0.006 | 7.11E-21 | 146.84 |
| rs56391344 | A | G | 0.25 | 0.015 | 0.002 | 7.49E-20 | 0 | 0.006 | 0.957 | 85.56 |
| rs56794817 | A | G | 0.16 | 0.012 | 0.002 | 1.69E-10 | -0.002 | 0.007 | 0.742 | 41.23 |
| rs57349798 | A | G | 0.41 | 0.009 | 0.001 | 8.63E-11 | 0 | 0.005 | 0.955 | 44.13 |
| rs57352738 | A | T | 0.2 | -0.015 | 0.002 | 2.51E-18 | -0.001 | 0.006 | 0.885 | 79.94 |
| rs575113 | A | G | 0.29 | 0.013 | 0.002 | 7.52E-18 | 0.006 | 0.005 | 0.271 | 78.62 |
| rs5754753 | T | C | 0.72 | -0.013 | 0.002 | 6.15E-16 | -0.001 | 0.005 | 0.783 | 62.02 |
| rs57661533 | T | C | 0.13 | -0.011 | 0.002 | 3.24E-08 | 0.007 | 0.007 | 0.317 | 29.47 |
| rs580652 | T | C | 0.9 | -0.015 | 0.002 | 4.65E-10 | -0.004 | 0.008 | 0.664 | 37.52 |
| rs590013 | T | C | 0.68 | 0.01 | 0.002 | 2.47E-11 | 0.003 | 0.005 | 0.504 | 44.44 |
| rs6043521 | T | C | 0.6 | -0.008 | 0.001 | 9.72E-09 | -0.002 | 0.005 | 0.762 | 35.15 |
| rs6065080 | T | C | 0.36 | -0.013 | 0.002 | 7.49E-19 | -0.01 | 0.005 | 0.054 | 75.11 |
| rs6065784 | C | G | 0.7 | 0.011 | 0.002 | 5.78E-13 | 0.009 | 0.005 | 0.095 | 53.78 |
| rs61104616 | A | G | 0.52 | -0.017 | 0.001 | 1.56E-34 | 0.028 | 0.005 | 1.27E-08 | 150.94 |
| rs6123924 | A | G | 0.84 | 0.013 | 0.002 | 3.20E-11 | 0.009 | 0.007 | 0.194 | 46.1 |
| rs61527214 | A | G | 0.41 | 0.011 | 0.001 | 1.13E-14 | 0.009 | 0.005 | 0.076 | 61.73 |
| rs61997667 | T | C | 0.15 | -0.014 | 0.002 | 2.24E-13 | 0.005 | 0.007 | 0.448 | 51.84 |
| rs62155350 | A | G | 0.02 | 0.034 | 0.005 | 1.69E-10 | 0.007 | 0.017 | 0.683 | 40.67 |
| rs62155873 | T | C | 0.12 | -0.015 | 0.002 | 1.65E-12 | -0.019 | 0.007 | 0.011 | 51.7 |
| rs62174974 | A | G | 0.19 | -0.011 | 0.002 | 2.76E-09 | -0.002 | 0.006 | 0.772 | 34.03 |
| rs62179650 | A | G | 0.3 | 0.011 | 0.002 | 5.66E-13 | 0.009 | 0.005 | 0.085 | 49 |
| rs62182994 | T | C | 0.69 | 0.014 | 0.002 | 2.55E-19 | -0.001 | 0.005 | 0.871 | 83.42 |
| rs62379838 | T | C | 0.7 | 0.012 | 0.002 | 1.77E-15 | 0 | 0.005 | 0.945 | 65.07 |
| rs628993 | A | G | 0.11 | 0.013 | 0.002 | 3.83E-08 | 0 | 0.009 | 0.983 | 30.97 |
| rs6449503 | A | G | 0.5 | 0.018 | 0.001 | 1.84E-39 | 0.013 | 0.005 | 0.007 | 174.62 |
| rs6490618 | T | C | 0.33 | 0.009 | 0.002 | 3.48E-09 | -0.001 | 0.005 | 0.864 | 34.42 |
| rs6493265 | T | C | 0.39 | -0.012 | 0.001 | 2.26E-17 | -0.008 | 0.005 | 0.112 | 75.94 |
| rs6557171 | T | C | 0.32 | -0.016 | 0.002 | 5.48E-25 | -0.002 | 0.005 | 0.747 | 106.78 |
| rs660001 | A | G | 0.21 | -0.016 | 0.002 | 1.66E-21 | 0.002 | 0.006 | 0.687 | 93.07 |
| rs66568921 | T | G | 0.64 | -0.018 | 0.002 | 7.60E-34 | -0.004 | 0.005 | 0.486 | 145.6 |
| rs66721975 | A | G | 0.29 | -0.009 | 0.002 | 1.51E-08 | 0.017 | 0.005 | 0.002 | 33.64 |
| rs6690195 | T | C | 0.49 | -0.011 | 0.001 | 7.89E-16 | 0.01 | 0.005 | 0.04 | 65.15 |
| rs6697584 | T | C | 0.79 | -0.013 | 0.002 | 1.43E-13 | 0.003 | 0.006 | 0.635 | 54.93 |
| rs6704768 | A | G | 0.56 | -0.012 | 0.001 | 1.84E-16 | 0 | 0.005 | 0.996 | 68.65 |
| rs6731373 | A | G | 0.34 | -0.012 | 0.002 | 1.39E-14 | -0.005 | 0.005 | 0.307 | 58.78 |
| rs6812533 | T | C | 0.74 | 0.011 | 0.002 | 4.85E-11 | 0 | 0.006 | 0.985 | 43.89 |
| rs68145588 | T | G | 0.14 | -0.013 | 0.002 | 7.53E-10 | 0.014 | 0.007 | 0.043 | 36.57 |
| rs6917154 | T | C | 0.12 | 0.013 | 0.002 | 2.55E-09 | -0.008 | 0.007 | 0.278 | 36.57 |
| rs6924023 | A | G | 0.75 | 0.011 | 0.002 | 5.17E-11 | -0.015 | 0.006 | 0.008 | 44.72 |
| rs6977237 | T | C | 0.47 | 0.008 | 0.001 | 5.59E-09 | 0.002 | 0.005 | 0.679 | 34.31 |
| rs6994287 | A | G | 0.41 | 0.01 | 0.001 | 1.11E-12 | -0.004 | 0.005 | 0.478 | 53.08 |
| rs7012546 | T | C | 0.42 | 0.009 | 0.001 | 1.25E-10 | 0.001 | 0.005 | 0.766 | 43.18 |
| rs7029718 | A | G | 0.41 | 0.024 | 0.001 | 7.82E-65 | 0.005 | 0.005 | 0.319 | 303.76 |
| rs7041702 | A | G | 0.74 | -0.011 | 0.002 | 4.53E-12 | 0.001 | 0.006 | 0.822 | 48.13 |
| rs7167688 | T | C | 0.49 | -0.009 | 0.001 | 1.71E-11 | 0 | 0.005 | 0.959 | 45.08 |
| rs7171405 | A | G | 0.24 | -0.01 | 0.002 | 7.53E-10 | 0.004 | 0.006 | 0.481 | 39.85 |
| rs717996 | T | C | 0.6 | -0.012 | 0.001 | 1.89E-17 | 0.001 | 0.005 | 0.779 | 75.94 |
| rs7226824 | T | C | 0.53 | -0.008 | 0.001 | 2.72E-08 | 0.007 | 0.005 | 0.134 | 31.04 |
| rs72486027 | T | C | 0.25 | -0.01 | 0.002 | 1.61E-10 | 0.006 | 0.006 | 0.269 | 42.25 |
| rs7254263 | T | C | 0.28 | -0.012 | 0.002 | 1.84E-14 | -0.002 | 0.005 | 0.732 | 55.32 |
| rs72622559 | T | C | 0.23 | -0.012 | 0.002 | 5.50E-13 | -0.001 | 0.006 | 0.894 | 49.83 |
| rs72672052 | A | T | 0.18 | -0.01 | 0.002 | 4.87E-08 | -0.008 | 0.006 | 0.194 | 31.48 |
| rs72709560 | A | G | 0.69 | 0.009 | 0.002 | 2.03E-08 | 0.003 | 0.005 | 0.578 | 32.11 |
| rs72828517 | T | C | 0.83 | -0.017 | 0.002 | 1.38E-19 | -0.004 | 0.006 | 0.476 | 78.18 |
| rs72829857 | A | G | 0.76 | -0.015 | 0.002 | 1.52E-19 | 0.005 | 0.006 | 0.422 | 86.72 |
| rs72917504 | T | C | 0.06 | -0.018 | 0.003 | 2.91E-09 | 0.02 | 0.01 | 0.057 | 35.2 |
| rs72944064 | T | C | 0.74 | -0.01 | 0.002 | 9.39E-10 | -0.006 | 0.006 | 0.265 | 37.52 |
| rs72962169 | T | C | 0.16 | -0.017 | 0.002 | 1.26E-19 | 0.002 | 0.007 | 0.75 | 83.87 |
| rs72993796 | T | C | 0.88 | 0.013 | 0.002 | 1.12E-08 | 0.011 | 0.008 | 0.136 | 33.32 |
| rs730384 | A | G | 0.44 | 0.01 | 0.001 | 9.38E-13 | -0.002 | 0.005 | 0.639 | 52.05 |
| rs73055556 | A | G | 0.14 | 0.011 | 0.002 | 4.95E-08 | 0.031 | 0.007 | 8.52E-06 | 30.8 |
| rs73191311 | A | G | 0.34 | -0.009 | 0.002 | 2.03E-10 | 0.003 | 0.005 | 0.568 | 39.27 |
| rs7321274 | A | G | 0.8 | 0.011 | 0.002 | 8.21E-11 | 0.004 | 0.006 | 0.51 | 40.11 |
| rs7326331 | A | G | 0.28 | -0.013 | 0.002 | 3.42E-16 | 0.007 | 0.005 | 0.2 | 64 |
| rs73344830 | A | G | 0.42 | 0.017 | 0.001 | 1.00E-32 | -0.01 | 0.005 | 0.035 | 147.45 |
| rs736281 | T | C | 0.39 | 0.009 | 0.001 | 2.49E-09 | 0.003 | 0.005 | 0.536 | 37.73 |
| rs73648455 | T | C | 0.08 | -0.018 | 0.003 | 1.99E-12 | 0.003 | 0.009 | 0.78 | 49.54 |
| rs743316 | T | C | 0.79 | 0.011 | 0.002 | 8.21E-11 | 0.007 | 0.006 | 0.26 | 43.4 |
| rs74415461 | T | C | 0.08 | 0.018 | 0.003 | 6.25E-12 | -0.011 | 0.009 | 0.193 | 45.82 |
| rs7449561 | A | G | 0.23 | 0.01 | 0.002 | 9.73E-10 | -0.001 | 0.006 | 0.917 | 36 |
| rs7460106 | T | C | 0.76 | -0.012 | 0.002 | 2.06E-13 | 0.006 | 0.006 | 0.324 | 52.35 |
| rs746839 | C | G | 0.63 | 0.014 | 0.002 | 1.19E-20 | -0.007 | 0.005 | 0.196 | 81 |
| rs74787922 | A | C | 0.93 | 0.018 | 0.003 | 8.97E-11 | 0.002 | 0.01 | 0.845 | 42.25 |
| rs75033012 | C | G | 0.04 | -0.026 | 0.004 | 1.38E-11 | 0.011 | 0.012 | 0.386 | 46.1 |
| rs75177132 | T | C | 0.05 | 0.03 | 0.004 | 1.01E-16 | 0.018 | 0.011 | 0.114 | 68.52 |
| rs7575637 | A | G | 0.45 | 0.011 | 0.001 | 3.43E-14 | 0.003 | 0.005 | 0.476 | 58.41 |
| rs7603132 | A | G | 0.19 | 0.014 | 0.002 | 1.20E-14 | -0.002 | 0.006 | 0.725 | 57.93 |
| rs76076331 | T | C | 0.13 | 0.021 | 0.002 | 1.71E-23 | -0.001 | 0.007 | 0.852 | 96.23 |
| rs7617204 | A | G | 0.52 | -0.01 | 0.001 | 4.33E-12 | -0.002 | 0.005 | 0.642 | 48.01 |
| rs76235882 | A | G | 0.97 | 0.024 | 0.004 | 4.52E-10 | -0.01 | 0.015 | 0.479 | 38.19 |
| rs76246107 | A | G | 0.09 | -0.016 | 0.003 | 6.52E-10 | -0.026 | 0.009 | 0.006 | 38.82 |
| rs7650602 | T | C | 0.56 | -0.009 | 0.001 | 2.77E-10 | -0.016 | 0.005 | 0.001 | 40.41 |
| rs76577427 | C | G | 0.9 | 0.02 | 0.002 | 7.78E-18 | -0.007 | 0.008 | 0.358 | 71.54 |
| rs7672622 | A | G | 0.75 | -0.009 | 0.002 | 4.72E-08 | -0.007 | 0.006 | 0.216 | 30.25 |
| rs7683416 | T | C | 0.46 | 0.014 | 0.001 | 6.05E-23 | 0.012 | 0.005 | 0.011 | 98.58 |
| rs76878669 | C | G | 0.76 | 0.014 | 0.002 | 3.10E-17 | 0.005 | 0.006 | 0.431 | 68.79 |
| rs7692359 | T | C | 0.78 | -0.011 | 0.002 | 1.40E-10 | -0.004 | 0.006 | 0.477 | 41.11 |
| rs77025239 | A | G | 0.16 | -0.014 | 0.002 | 4.65E-13 | 0.006 | 0.007 | 0.384 | 54.29 |
| rs77609760 | A | G | 0.93 | 0.024 | 0.003 | 7.71E-17 | 0 | 0.01 | 0.963 | 71.04 |
| rs77719387 | A | T | 0.02 | -0.04 | 0.006 | 3.78E-12 | -0.038 | 0.021 | 0.068 | 48.28 |
| rs7849487 | T | G | 0.66 | -0.017 | 0.002 | 1.25E-29 | 0.009 | 0.005 | 0.068 | 123.95 |
| rs78648104 | T | C | 0.91 | -0.014 | 0.002 | 1.70E-08 | -0.014 | 0.009 | 0.115 | 31.81 |
| rs7875078 | A | C | 0.46 | -0.008 | 0.001 | 2.72E-08 | -0.001 | 0.005 | 0.914 | 31.04 |
| rs7894722 | T | C | 0.37 | 0.009 | 0.001 | 5.41E-09 | -0.01 | 0.005 | 0.055 | 36.86 |
| rs790647 | A | C | 0.23 | -0.015 | 0.002 | 3.81E-18 | 0.005 | 0.006 | 0.379 | 72.75 |
| rs7910403 | T | G | 0.2 | 0.012 | 0.002 | 5.08E-12 | 0.005 | 0.006 | 0.411 | 45.94 |
| rs7924036 | T | G | 0.51 | 0.013 | 0.001 | 2.55E-21 | 0.016 | 0.005 | 0.001 | 90.25 |
| rs79265434 | A | G | 0.88 | -0.02 | 0.002 | 1.98E-19 | -0.012 | 0.008 | 0.11 | 80.18 |
| rs7972246 | T | C | 0.35 | 0.011 | 0.002 | 1.01E-13 | -0.006 | 0.005 | 0.204 | 53.78 |
| rs79728014 | A | G | 0.86 | -0.013 | 0.002 | 2.06E-11 | -0.011 | 0.007 | 0.1 | 44.89 |
| rs7974852 | A | C | 0.52 | 0.012 | 0.001 | 1.14E-16 | -0.005 | 0.005 | 0.354 | 69.84 |
| rs7977614 | A | G | 0.71 | -0.013 | 0.002 | 6.84E-17 | -0.006 | 0.005 | 0.292 | 70.14 |
| rs79994730 | T | C | 0.04 | 0.024 | 0.004 | 5.88E-12 | -0.033 | 0.011 | 0.003 | 47.02 |
| rs8008382 | T | C | 0.3 | -0.011 | 0.002 | 4.07E-12 | -0.004 | 0.005 | 0.426 | 49.94 |
| rs8024 | A | C | 0.33 | -0.012 | 0.002 | 1.13E-14 | 0.005 | 0.005 | 0.312 | 58.78 |
| rs80257979 | T | G | 0.97 | -0.025 | 0.004 | 3.31E-10 | -0.029 | 0.014 | 0.039 | 39.69 |
| rs8030487 | A | G | 0.69 | 0.009 | 0.002 | 2.52E-08 | -0.008 | 0.005 | 0.145 | 32.11 |
| rs806816 | A | T | 0.75 | 0.01 | 0.002 | 4.80E-10 | 0.011 | 0.006 | 0.041 | 39.85 |
| rs8097125 | T | C | 0.38 | 0.009 | 0.001 | 2.31E-09 | 0.001 | 0.005 | 0.898 | 37.73 |
| rs912883 | T | C | 0.68 | 0.009 | 0.002 | 6.20E-09 | -0.004 | 0.005 | 0.411 | 33.64 |
| rs9289300 | T | C | 0.84 | -0.016 | 0.002 | 1.00E-17 | 0.003 | 0.007 | 0.697 | 74.5 |
| rs929511 | T | C | 0.13 | -0.016 | 0.002 | 3.91E-14 | 0.009 | 0.008 | 0.226 | 58.05 |
| rs9359939 | A | C | 0.24 | -0.011 | 0.002 | 1.51E-10 | 0.007 | 0.006 | 0.257 | 43.07 |
| rs936496 | A | G | 0.62 | -0.008 | 0.001 | 4.38E-08 | 0.002 | 0.005 | 0.674 | 31.84 |
| rs9373363 | A | G | 0.75 | -0.011 | 0.002 | 5.54E-12 | -0.006 | 0.006 | 0.314 | 48.13 |
| rs9375188 | T | C | 0.48 | 0.021 | 0.001 | 8.78E-52 | 0.012 | 0.005 | 0.011 | 229.31 |
| rs9411331 | A | C | 0.68 | 0.015 | 0.002 | 7.63E-22 | -0.009 | 0.005 | 0.072 | 93.44 |
| rs9492774 | C | G | 0.37 | 0.008 | 0.002 | 4.33E-08 | 0.004 | 0.005 | 0.448 | 28.44 |
| rs9529146 | T | C | 0.23 | 0.013 | 0.002 | 3.48E-14 | 0.01 | 0.006 | 0.095 | 55.81 |
| rs9556958 | T | C | 0.52 | -0.011 | 0.001 | 5.96E-15 | -0.008 | 0.005 | 0.117 | 61.73 |
| rs9616947 | T | C | 0.38 | -0.013 | 0.002 | 3.58E-20 | 0.002 | 0.005 | 0.622 | 79.8 |
| rs9633970 | T | C | 0.76 | 0.009 | 0.002 | 2.81E-08 | -0.007 | 0.006 | 0.264 | 32.35 |
| rs9655780 | A | G | 0.83 | -0.015 | 0.002 | 3.70E-16 | 0.005 | 0.007 | 0.448 | 64.84 |
| rs969512 | A | T | 0.66 | -0.011 | 0.002 | 9.84E-13 | 0.003 | 0.005 | 0.536 | 49.94 |
| rs9853928 | T | C | 0.21 | -0.013 | 0.002 | 1.59E-13 | 0.006 | 0.006 | 0.322 | 55.81 |
| rs9859556 | T | G | 0.31 | 0.029 | 0.002 | 4.61E-82 | 0.015 | 0.005 | 0.005 | 373.78 |
| rs9882532 | T | C | 0.64 | 0.012 | 0.002 | 2.15E-17 | 0.002 | 0.005 | 0.627 | 68.34 |
| rs9886703 | A | T | 0.17 | -0.014 | 0.002 | 4.46E-13 | -0.012 | 0.007 | 0.072 | 51.24 |
| rs9927137 | A | G | 0.53 | 0.008 | 0.001 | 5.11E-09 | 0.006 | 0.005 | 0.187 | 34.31 |
| rs9927842 | T | C | 0.15 | -0.013 | 0.002 | 6.22E-11 | 0.002 | 0.007 | 0.773 | 42.25 |
| rs9929556 | T | G | 0.57 | -0.01 | 0.001 | 2.31E-12 | 0.003 | 0.005 | 0.524 | 50.01 |
| rs9929762 | A | G | 0.56 | 0.01 | 0.001 | 1.03E-11 | 0.005 | 0.005 | 0.345 | 47.02 |
| rs9933256 | A | G | 0.56 | 0.012 | 0.001 | 6.06E-17 | -0.002 | 0.005 | 0.712 | 72.25 |

**SNPs used as instrumental variables in the MR analysis of educational attainment on fractional anisotropy (cortical)**

| **SNP** | **Effect allele** | **Other allele** | **Effect allele frequency** | **beta (exposure)** | **SE (exposure)** | ***P* (exposure)** | **beta (outcome)** | **SE (outcome)** | ***P* (outcome)** | ***F*-statistic** |
| --- | --- | --- | --- | --- | --- | --- | --- | --- | --- | --- |
| rs10006235 | T | C | 0.27 | -0.009 | 0.002 | 4.48E-09 | -0.009 | 0.009 | 0.28 | 33.79 |
| rs10021733 | T | C | 0.15 | -0.016 | 0.003 | 9.45E-10 | -0.011 | 0.01 | 0.3 | 36.89 |
| rs10046069 | A | G | 0.12 | 0.013 | 0.002 | 4.40E-09 | 0.008 | 0.012 | 0.509 | 33.32 |
| rs10060023 | T | C | 0.33 | 0.011 | 0.002 | 1.88E-13 | -0.004 | 0.008 | 0.576 | 53.78 |
| rs10080647 | A | C | 0.14 | 0.012 | 0.002 | 7.05E-10 | 0.01 | 0.011 | 0.34 | 38.44 |
| rs1008078 | T | C | 0.4 | -0.018 | 0.001 | 4.50E-37 | -0.008 | 0.008 | 0.295 | 169 |
| rs10189857 | A | G | 0.57 | 0.016 | 0.001 | 4.69E-29 | -0.01 | 0.008 | 0.211 | 127.37 |
| rs10193498 | A | T | 0.76 | 0.01 | 0.002 | 3.10E-09 | 0.001 | 0.009 | 0.925 | 36.75 |
| rs10215082 | A | G | 0.47 | -0.015 | 0.001 | 7.03E-26 | -0.014 | 0.008 | 0.072 | 113.27 |
| rs10460095 | A | G | 0.57 | -0.012 | 0.001 | 2.61E-18 | 0.007 | 0.008 | 0.374 | 78.45 |
| rs10761251 | A | T | 0.66 | 0.011 | 0.002 | 2.52E-14 | -0.005 | 0.008 | 0.549 | 56.75 |
| rs10772644 | C | G | 0.89 | 0.014 | 0.002 | 1.69E-10 | 0.011 | 0.012 | 0.369 | 41.66 |
| rs10773002 | A | T | 0.25 | 0.018 | 0.002 | 2.50E-30 | 0.004 | 0.009 | 0.69 | 133.69 |
| rs10773208 | T | C | 0.25 | -0.011 | 0.002 | 8.09E-12 | -0.012 | 0.009 | 0.182 | 47.27 |
| rs10798888 | T | G | 0.17 | -0.014 | 0.002 | 5.15E-14 | -0.028 | 0.01 | 0.006 | 54.29 |
| rs10799615 | A | G | 0.25 | -0.009 | 0.002 | 4.11E-08 | -0.003 | 0.009 | 0.766 | 30.94 |
| rs10805383 | A | G | 0.49 | -0.01 | 0.001 | 2.22E-13 | 0 | 0.008 | 0.962 | 54.13 |
| rs10830858 | T | C | 0.51 | 0.009 | 0.001 | 5.31E-11 | -0.002 | 0.008 | 0.795 | 43.18 |
| rs10844179 | A | G | 0.23 | 0.01 | 0.002 | 5.03E-10 | 0.012 | 0.009 | 0.207 | 37.43 |
| rs10875121 | C | G | 0.84 | 0.017 | 0.002 | 4.46E-20 | -0.015 | 0.01 | 0.162 | 83.87 |
| rs10892807 | T | C | 0.57 | 0.011 | 0.001 | 1.09E-14 | -0.006 | 0.008 | 0.41 | 60.62 |
| rs10940921 | T | G | 0.42 | 0.011 | 0.001 | 1.31E-14 | 0.002 | 0.008 | 0.812 | 62.86 |
| rs10951590 | T | C | 0.33 | -0.011 | 0.002 | 3.62E-13 | 0.011 | 0.008 | 0.192 | 52.8 |
| rs10979613 | T | C | 0.64 | -0.012 | 0.002 | 1.25E-15 | 0.012 | 0.008 | 0.136 | 60.84 |
| rs10996167 | C | G | 0.65 | 0.011 | 0.002 | 7.47E-09 | -0.012 | 0.008 | 0.151 | 31.92 |
| rs11003463 | T | G | 0.61 | 0.009 | 0.002 | 5.58E-10 | 0.003 | 0.008 | 0.698 | 36.8 |
| rs11023764 | A | G | 0.64 | 0.009 | 0.002 | 5.32E-10 | 0.008 | 0.008 | 0.329 | 36.8 |
| rs11030102 | C | G | 0.75 | 0.01 | 0.002 | 4.60E-10 | 0 | 0.009 | 0.954 | 39.85 |
| rs11076962 | T | C | 0.72 | 0.009 | 0.002 | 4.29E-08 | -0.011 | 0.009 | 0.205 | 28.89 |
| rs11081529 | T | C | 0.71 | 0.013 | 0.002 | 5.22E-16 | 0.001 | 0.008 | 0.924 | 70.56 |
| rs11082011 | T | C | 0.67 | 0.019 | 0.002 | 2.06E-38 | -0.007 | 0.008 | 0.394 | 167.27 |
| rs11121177 | A | G | 0.18 | 0.015 | 0.002 | 1.44E-16 | -0.003 | 0.01 | 0.801 | 70.37 |
| rs11130380 | T | G | 0.61 | 0.009 | 0.001 | 4.17E-10 | 0.006 | 0.008 | 0.43 | 41.33 |
| rs11138947 | T | C | 0.72 | 0.011 | 0.002 | 2.36E-11 | 0.006 | 0.009 | 0.46 | 43.07 |
| rs11157931 | A | C | 0.39 | -0.014 | 0.001 | 3.86E-22 | -0.005 | 0.008 | 0.487 | 98.58 |
| rs111852224 | T | C | 0.12 | 0.016 | 0.002 | 4.85E-13 | 0.004 | 0.012 | 0.715 | 50.28 |
| rs11211123 | A | G | 0.22 | 0.01 | 0.002 | 7.62E-10 | 0.011 | 0.009 | 0.231 | 37.43 |
| rs11213482 | A | G | 0.84 | -0.011 | 0.002 | 2.68E-08 | -0.002 | 0.01 | 0.848 | 31.12 |
| rs112603734 | A | C | 0.77 | -0.013 | 0.002 | 2.72E-09 | 0 | 0.009 | 0.976 | 35.46 |
| rs112806496 | C | G | 0.91 | -0.016 | 0.002 | 1.72E-10 | -0.003 | 0.013 | 0.818 | 40.96 |
| rs1128956 | T | G | 0.83 | -0.014 | 0.002 | 5.34E-14 | 0.009 | 0.01 | 0.366 | 54.29 |
| rs113520408 | A | G | 0.28 | 0.016 | 0.002 | 1.03E-22 | 0.006 | 0.009 | 0.454 | 93.85 |
| rs113615161 | T | C | 0.13 | -0.012 | 0.002 | 2.98E-09 | 0.009 | 0.011 | 0.432 | 34.31 |
| rs114593137 | A | T | 0.79 | -0.012 | 0.002 | 1.65E-12 | 0.021 | 0.009 | 0.028 | 51.5 |
| rs115000530 | A | T | 0.95 | -0.025 | 0.003 | 1.09E-15 | 0.004 | 0.017 | 0.815 | 65.04 |
| rs11542663 | A | C | 0.69 | 0.011 | 0.002 | 2.44E-13 | -0.003 | 0.008 | 0.763 | 55.75 |
| rs11588857 | A | G | 0.21 | 0.02 | 0.002 | 4.45E-31 | 0.013 | 0.009 | 0.164 | 137.03 |
| rs11598765 | A | G | 0.2 | -0.011 | 0.002 | 1.73E-09 | 0.004 | 0.01 | 0.673 | 34.68 |
| rs11620355 | A | G | 0.09 | 0.018 | 0.002 | 1.09E-13 | -0.019 | 0.014 | 0.169 | 53 |
| rs11657342 | A | G | 0.37 | 0.014 | 0.002 | 2.32E-13 | -0.001 | 0.008 | 0.915 | 54.29 |
| rs11657979 | A | G | 0.24 | -0.009 | 0.002 | 4.57E-08 | -0.006 | 0.009 | 0.548 | 28.65 |
| rs11663602 | A | C | 0.28 | -0.013 | 0.002 | 9.27E-17 | 0.004 | 0.009 | 0.642 | 66.02 |
| rs1167827 | A | G | 0.43 | 0.01 | 0.001 | 2.93E-13 | -0.021 | 0.008 | 0.007 | 55.18 |
| rs11724690 | T | G | 0.29 | 0.009 | 0.002 | 2.08E-09 | 0 | 0.009 | 0.975 | 38.44 |
| rs117398064 | C | G | 0.09 | -0.014 | 0.002 | 1.16E-08 | -0.008 | 0.013 | 0.559 | 31.81 |
| rs118134876 | T | C | 0.06 | -0.022 | 0.003 | 1.50E-13 | -0.009 | 0.016 | 0.595 | 55.25 |
| rs11871429 | A | G | 0.77 | 0.013 | 0.002 | 2.52E-14 | 0.002 | 0.009 | 0.808 | 56.69 |
| rs11894424 | A | C | 0.95 | 0.021 | 0.003 | 2.14E-10 | -0.002 | 0.019 | 0.909 | 40.11 |
| rs12054166 | C | G | 0.74 | 0.01 | 0.002 | 2.30E-09 | -0.017 | 0.009 | 0.058 | 36 |
| rs12076635 | C | G | 0.78 | 0.021 | 0.002 | 3.33E-34 | 0.022 | 0.009 | 0.019 | 148.27 |
| rs12113634 | T | C | 0.65 | 0.009 | 0.002 | 2.50E-09 | 0 | 0.008 | 0.995 | 34.42 |
| rs12151248 | T | C | 0.11 | -0.015 | 0.002 | 3.81E-11 | -0.004 | 0.012 | 0.724 | 41.97 |
| rs1220779 | A | G | 0.47 | -0.008 | 0.001 | 1.58E-08 | -0.003 | 0.008 | 0.677 | 31.84 |
| rs12273435 | A | G | 0.21 | -0.013 | 0.002 | 4.72E-14 | -0.001 | 0.01 | 0.909 | 60.29 |
| rs12332731 | A | T | 0.19 | 0.014 | 0.002 | 5.96E-14 | -0.006 | 0.01 | 0.517 | 56.25 |
| rs12375949 | T | C | 0.43 | -0.014 | 0.001 | 1.61E-22 | 0.012 | 0.008 | 0.107 | 97.16 |
| rs12438177 | A | G | 0.37 | 0.009 | 0.002 | 1.48E-10 | 0 | 0.008 | 0.985 | 38.44 |
| rs12468040 | T | G | 0.38 | 0.014 | 0.001 | 3.12E-21 | 0.01 | 0.008 | 0.216 | 95.76 |
| rs12503522 | T | C | 0.28 | -0.011 | 0.002 | 3.93E-13 | 0.006 | 0.008 | 0.476 | 49.88 |
| rs12506221 | T | G | 0.44 | -0.013 | 0.001 | 4.23E-20 | 0.005 | 0.008 | 0.489 | 86.22 |
| rs12519073 | T | C | 0.23 | -0.011 | 0.002 | 1.16E-10 | 0.001 | 0.009 | 0.883 | 39.62 |
| rs12524795 | T | C | 0.44 | 0.01 | 0.001 | 5.28E-13 | 0 | 0.008 | 0.972 | 53.08 |
| rs12571549 | A | G | 0.14 | 0.015 | 0.002 | 1.45E-13 | -0.002 | 0.011 | 0.84 | 57 |
| rs12574281 | A | C | 0.63 | -0.008 | 0.002 | 3.04E-08 | 0.004 | 0.008 | 0.575 | 28.44 |
| rs12591647 | T | C | 0.82 | -0.015 | 0.002 | 7.78E-17 | -0.007 | 0.01 | 0.484 | 71.31 |
| rs12602286 | T | G | 0.87 | 0.016 | 0.002 | 4.62E-14 | 0.01 | 0.012 | 0.386 | 58.05 |
| rs12614263 | A | G | 0.5 | 0.008 | 0.001 | 3.38E-09 | 0.007 | 0.008 | 0.34 | 35.15 |
| rs12638072 | A | G | 0.3 | -0.011 | 0.002 | 3.13E-12 | -0.015 | 0.009 | 0.084 | 50.88 |
| rs12643771 | T | C | 0.31 | 0.013 | 0.002 | 1.32E-17 | -0.012 | 0.008 | 0.142 | 75.11 |
| rs12646523 | T | C | 0.25 | -0.013 | 0.002 | 3.79E-16 | 0.002 | 0.009 | 0.854 | 68.06 |
| rs12670376 | A | G | 0.45 | 0.009 | 0.001 | 5.99E-11 | 0.007 | 0.008 | 0.364 | 43.18 |
| rs12761761 | T | C | 0.24 | 0.015 | 0.002 | 4.79E-20 | -0.014 | 0.009 | 0.113 | 81 |
| rs12765185 | A | T | 0.28 | -0.009 | 0.002 | 2.96E-08 | -0.004 | 0.009 | 0.631 | 30.25 |
| rs12789313 | T | C | 0.49 | 0.009 | 0.001 | 6.79E-11 | 0.001 | 0.008 | 0.911 | 43.18 |
| rs12875339 | A | C | 0.64 | -0.011 | 0.002 | 1.71E-13 | -0.004 | 0.008 | 0.642 | 51.84 |
| rs12888615 | T | C | 0.2 | 0.012 | 0.002 | 9.42E-12 | -0.003 | 0.01 | 0.791 | 44.44 |
| rs12957463 | A | G | 0.8 | 0.015 | 0.002 | 9.20E-17 | -0.004 | 0.009 | 0.681 | 65.79 |
| rs12967010 | T | C | 0.77 | 0.011 | 0.002 | 1.33E-11 | -0.005 | 0.009 | 0.583 | 44.97 |
| rs12981405 | T | C | 0.17 | -0.011 | 0.002 | 2.75E-09 | -0.017 | 0.01 | 0.108 | 34.75 |
| rs1301838 | T | C | 0.32 | -0.01 | 0.002 | 1.29E-10 | 0.005 | 0.008 | 0.506 | 40.96 |
| rs13018640 | T | C | 0.6 | -0.021 | 0.001 | 1.05E-50 | -0.005 | 0.008 | 0.5 | 235.84 |
| rs13050131 | A | C | 0.66 | -0.008 | 0.002 | 4.40E-08 | 0.003 | 0.008 | 0.715 | 29.16 |
| rs13133213 | A | G | 0.5 | 0.01 | 0.001 | 7.57E-12 | -0.012 | 0.008 | 0.123 | 47.02 |
| rs13145650 | T | C | 0.92 | -0.017 | 0.002 | 8.99E-12 | -0.007 | 0.014 | 0.599 | 47.89 |
| rs13163845 | T | C | 0.85 | -0.015 | 0.002 | 4.66E-14 | 0.002 | 0.011 | 0.881 | 55.5 |
| rs13197257 | T | G | 0.28 | 0.011 | 0.002 | 2.83E-12 | -0.021 | 0.009 | 0.013 | 46.41 |
| rs13212041 | T | C | 0.8 | 0.01 | 0.002 | 7.42E-09 | 0 | 0.009 | 0.991 | 31.48 |
| rs13240401 | T | C | 0.78 | 0.018 | 0.002 | 2.36E-24 | -0.007 | 0.009 | 0.459 | 107.18 |
| rs13261773 | C | G | 0.16 | 0.012 | 0.002 | 3.12E-10 | -0.001 | 0.011 | 0.959 | 41.23 |
| rs13266287 | A | T | 0.67 | -0.009 | 0.002 | 1.75E-08 | -0.008 | 0.008 | 0.307 | 32.11 |
| rs1329125 | T | C | 0.33 | -0.01 | 0.002 | 8.37E-11 | -0.01 | 0.008 | 0.223 | 41.82 |
| rs13327482 | A | G | 0.82 | -0.012 | 0.002 | 4.47E-11 | -0.018 | 0.01 | 0.076 | 45.19 |
| rs1334297 | A | G | 0.74 | 0.026 | 0.002 | 2.53E-58 | -0.013 | 0.009 | 0.136 | 258 |
| rs13422673 | T | C | 0.47 | -0.011 | 0.001 | 6.48E-15 | 0.001 | 0.008 | 0.871 | 61.73 |
| rs137079 | T | C | 0.14 | 0.013 | 0.002 | 3.06E-10 | 0.015 | 0.011 | 0.173 | 40.96 |
| rs1426619 | T | C | 0.43 | 0.01 | 0.001 | 1.40E-11 | -0.002 | 0.008 | 0.759 | 47.02 |
| rs1434630 | T | G | 0.15 | -0.014 | 0.002 | 4.63E-12 | 0.006 | 0.011 | 0.569 | 46.92 |
| rs143743568 | A | G | 0.15 | 0.012 | 0.002 | 4.99E-09 | 0.002 | 0.011 | 0.816 | 34.81 |
| rs150537577 | A | G | 0.92 | -0.015 | 0.003 | 9.50E-09 | 0.008 | 0.015 | 0.605 | 33.73 |
| rs1505676 | C | G | 0.63 | 0.008 | 0.002 | 6.67E-09 | 0.002 | 0.008 | 0.81 | 31.36 |
| rs1542354 | A | G | 0.48 | -0.008 | 0.001 | 2.72E-09 | 0.003 | 0.008 | 0.727 | 36 |
| rs1544 | A | G | 0.26 | -0.01 | 0.002 | 1.76E-09 | -0.014 | 0.009 | 0.096 | 36 |
| rs1564347 | T | G | 0.34 | 0.009 | 0.002 | 2.39E-09 | -0.005 | 0.008 | 0.565 | 34.42 |
| rs1603460 | T | G | 0.42 | 0.009 | 0.001 | 2.20E-10 | 0.004 | 0.008 | 0.643 | 41.33 |
| rs162445 | A | G | 0.08 | 0.015 | 0.003 | 5.72E-09 | 0.007 | 0.014 | 0.643 | 34.18 |
| rs1689510 | C | G | 0.33 | 0.019 | 0.002 | 1.35E-38 | -0.019 | 0.008 | 0.017 | 167.27 |
| rs17048855 | A | G | 0.34 | 0.011 | 0.002 | 3.10E-13 | -0.005 | 0.008 | 0.515 | 51.84 |
| rs17110109 | T | C | 0.62 | -0.009 | 0.001 | 1.47E-10 | 0.006 | 0.008 | 0.42 | 44.13 |
| rs17144467 | A | C | 0.69 | 0.01 | 0.002 | 8.74E-11 | -0.003 | 0.008 | 0.696 | 42.68 |
| rs17148998 | A | G | 0.2 | 0.01 | 0.002 | 6.98E-09 | -0.01 | 0.009 | 0.284 | 35.3 |
| rs1717204 | A | C | 0.18 | -0.012 | 0.002 | 1.20E-10 | 0.017 | 0.01 | 0.09 | 42.25 |
| rs17248751 | A | G | 0.78 | -0.014 | 0.002 | 1.28E-15 | 0 | 0.009 | 0.993 | 64 |
| rs173003 | A | C | 0.49 | -0.008 | 0.001 | 5.50E-09 | -0.034 | 0.008 | 6.12E-06 | 34.31 |
| rs1738050 | C | G | 0.62 | -0.009 | 0.001 | 1.42E-10 | 0.01 | 0.008 | 0.213 | 44.13 |
| rs17411339 | A | G | 0.56 | 0.014 | 0.001 | 5.10E-23 | -0.001 | 0.008 | 0.942 | 100 |
| rs17428076 | C | G | 0.76 | 0.014 | 0.002 | 6.41E-17 | -0.001 | 0.009 | 0.925 | 73.32 |
| rs1747714 | T | C | 0.53 | 0.011 | 0.001 | 3.82E-14 | -0.015 | 0.008 | 0.055 | 57.33 |
| rs1747817 | T | C | 0.76 | -0.009 | 0.002 | 2.29E-08 | 0.01 | 0.009 | 0.257 | 33.06 |
| rs17489649 | A | G | 0.68 | 0.013 | 0.002 | 2.77E-17 | 0.003 | 0.008 | 0.678 | 71.68 |
| rs17502934 | T | G | 0.15 | -0.017 | 0.002 | 2.12E-17 | 0.003 | 0.011 | 0.768 | 69.72 |
| rs175325 | A | T | 0.6 | -0.01 | 0.001 | 2.24E-12 | -0.006 | 0.008 | 0.415 | 51.02 |
| rs17551064 | A | G | 0.84 | 0.014 | 0.002 | 1.53E-13 | 0.003 | 0.01 | 0.799 | 55.07 |
| rs17563464 | A | C | 0.22 | -0.015 | 0.002 | 1.74E-17 | -0.013 | 0.01 | 0.185 | 74.77 |
| rs17565975 | A | G | 0.56 | -0.011 | 0.001 | 5.54E-14 | 0.012 | 0.008 | 0.125 | 57.33 |
| rs17598675 | T | C | 0.51 | -0.012 | 0.001 | 1.37E-16 | -0.004 | 0.008 | 0.584 | 68.65 |
| rs17604349 | A | G | 0.19 | 0.013 | 0.002 | 2.36E-12 | -0.009 | 0.01 | 0.375 | 49.78 |
| rs176218 | T | G | 0.2 | 0.02 | 0.002 | 1.10E-29 | -0.002 | 0.01 | 0.839 | 123.46 |
| rs17732878 | T | C | 0.79 | -0.01 | 0.002 | 4.06E-08 | 0.002 | 0.009 | 0.811 | 31.23 |
| rs178183 | T | C | 0.25 | 0.015 | 0.002 | 4.81E-19 | 0.001 | 0.009 | 0.897 | 82.13 |
| rs17882802 | A | G | 0.43 | 0.008 | 0.001 | 3.00E-09 | 0.01 | 0.008 | 0.195 | 36 |
| rs182902112 | A | C | 0.02 | -0.03 | 0.005 | 1.17E-08 | -0.043 | 0.033 | 0.186 | 32.04 |
| rs1842713 | A | G | 0.79 | -0.012 | 0.002 | 9.67E-12 | -0.003 | 0.009 | 0.766 | 47.37 |
| rs1861786 | A | G | 0.38 | -0.008 | 0.001 | 2.39E-08 | 0.011 | 0.008 | 0.165 | 33.47 |
| rs1865955 | T | C | 0.82 | 0.012 | 0.002 | 4.06E-11 | -0.021 | 0.01 | 0.04 | 41.23 |
| rs1866823 | A | G | 0.54 | 0.011 | 0.001 | 3.09E-14 | 0.013 | 0.008 | 0.093 | 58.41 |
| rs1890132 | T | C | 0.71 | -0.009 | 0.002 | 6.56E-09 | 0.001 | 0.008 | 0.899 | 35.2 |
| rs192436652 | T | C | 0.03 | -0.032 | 0.004 | 2.06E-12 | -0.002 | 0.024 | 0.922 | 50.25 |
| rs1933264 | T | C | 0.75 | 0.01 | 0.002 | 5.85E-10 | 0.014 | 0.009 | 0.113 | 39.06 |
| rs1955250 | A | C | 0.91 | -0.014 | 0.002 | 2.26E-08 | -0.009 | 0.014 | 0.495 | 31.36 |
| rs1991585 | T | C | 0.71 | -0.01 | 0.002 | 6.26E-11 | 0.008 | 0.008 | 0.336 | 45.34 |
| rs2007655 | T | G | 0.5 | 0.009 | 0.001 | 8.64E-10 | 0.001 | 0.008 | 0.914 | 37.73 |
| rs2011603 | A | G | 0.74 | -0.009 | 0.002 | 1.12E-08 | -0.004 | 0.009 | 0.614 | 32.35 |
| rs2034631 | T | C | 0.65 | 0.009 | 0.002 | 2.64E-09 | -0.007 | 0.008 | 0.355 | 35.2 |
| rs2052285 | A | G | 0.6 | 0.011 | 0.001 | 7.25E-15 | -0.004 | 0.008 | 0.593 | 64 |
| rs2055940 | A | G | 0.32 | 0.008 | 0.002 | 4.95E-08 | 0 | 0.008 | 0.968 | 29.88 |
| rs2081652 | A | T | 0.66 | 0.012 | 0.002 | 1.09E-16 | -0.005 | 0.008 | 0.537 | 67.24 |
| rs2131167 | A | G | 0.45 | 0.008 | 0.001 | 1.52E-08 | -0.004 | 0.008 | 0.605 | 32.65 |
| rs2179152 | T | C | 0.37 | -0.013 | 0.002 | 1.68E-19 | 0.01 | 0.008 | 0.197 | 76.27 |
| rs2183271 | T | C | 0.64 | 0.008 | 0.002 | 1.91E-08 | 0.003 | 0.008 | 0.735 | 31.36 |
| rs2199409 | T | C | 0.8 | 0.01 | 0.002 | 3.14E-09 | -0.003 | 0.01 | 0.789 | 36.71 |
| rs2212430 | T | C | 0.3 | -0.01 | 0.002 | 1.32E-10 | 0.005 | 0.008 | 0.551 | 39.06 |
| rs2216144 | T | C | 0.53 | 0.01 | 0.001 | 2.58E-12 | 0.012 | 0.008 | 0.104 | 49 |
| rs2256965 | A | G | 0.42 | 0.011 | 0.001 | 2.40E-13 | -0.016 | 0.008 | 0.04 | 57.33 |
| rs2287838 | A | G | 0.55 | -0.01 | 0.001 | 1.27E-13 | -0.018 | 0.008 | 0.021 | 55.18 |
| rs2290601 | T | C | 0.77 | -0.015 | 0.002 | 4.49E-19 | -0.008 | 0.009 | 0.387 | 76.82 |
| rs2297293 | C | G | 0.31 | 0.01 | 0.002 | 8.80E-11 | -0.001 | 0.008 | 0.919 | 43.56 |
| rs232496 | T | C | 0.37 | 0.009 | 0.002 | 7.14E-10 | 0.006 | 0.008 | 0.442 | 36 |
| rs2364544 | A | G | 0.39 | -0.011 | 0.001 | 2.91E-13 | 0.004 | 0.008 | 0.658 | 56.25 |
| rs2434672 | A | C | 0.53 | 0.009 | 0.001 | 1.06E-10 | -0.001 | 0.008 | 0.904 | 42.25 |
| rs2436760 | T | C | 0.97 | 0.023 | 0.004 | 4.52E-08 | 0.017 | 0.023 | 0.447 | 30.51 |
| rs2447097 | T | G | 0.46 | 0.01 | 0.001 | 1.10E-11 | -0.008 | 0.008 | 0.323 | 47.02 |
| rs2469226 | A | T | 0.22 | 0.01 | 0.002 | 1.28E-08 | -0.004 | 0.009 | 0.631 | 31.89 |
| rs2496482 | T | C | 0.42 | 0.012 | 0.002 | 9.20E-17 | -0.006 | 0.008 | 0.463 | 65.07 |
| rs252991 | A | G | 0.37 | 0.01 | 0.002 | 6.30E-11 | 0.006 | 0.008 | 0.451 | 40.11 |
| rs2545795 | A | C | 0.44 | 0.013 | 0.001 | 1.03E-18 | 0.002 | 0.008 | 0.808 | 79.72 |
| rs2554835 | A | G | 0.4 | 0.009 | 0.001 | 8.95E-10 | -0.009 | 0.008 | 0.252 | 39.51 |
| rs2570497 | T | C | 0.64 | -0.012 | 0.002 | 1.27E-16 | -0.004 | 0.008 | 0.59 | 65.07 |
| rs2725370 | T | C | 0.3 | -0.014 | 0.002 | 4.96E-20 | 0.007 | 0.008 | 0.392 | 88.36 |
| rs2764684 | T | C | 0.83 | 0.014 | 0.002 | 9.99E-15 | -0.016 | 0.01 | 0.124 | 56.65 |
| rs2838006 | T | C | 0.36 | 0.013 | 0.002 | 1.38E-19 | 0.015 | 0.008 | 0.065 | 77.44 |
| rs28513882 | A | G | 0.18 | -0.011 | 0.002 | 2.03E-09 | -0.006 | 0.01 | 0.534 | 37.35 |
| rs2885198 | A | G | 0.53 | 0.009 | 0.001 | 4.63E-10 | -0.002 | 0.008 | 0.752 | 39.51 |
| rs2923424 | A | G | 0.61 | 0.012 | 0.001 | 6.31E-17 | -0.001 | 0.008 | 0.849 | 73.47 |
| rs2929032 | A | G | 0.47 | -0.008 | 0.001 | 8.40E-09 | 0.004 | 0.008 | 0.587 | 33.47 |
| rs2958182 | A | T | 0.34 | 0.008 | 0.002 | 3.46E-08 | -0.011 | 0.008 | 0.181 | 29.88 |
| rs2964199 | T | C | 0.31 | -0.01 | 0.002 | 4.61E-11 | -0.005 | 0.008 | 0.523 | 43.56 |
| rs2964255 | A | G | 0.31 | 0.009 | 0.002 | 6.34E-09 | 0.012 | 0.008 | 0.166 | 34.42 |
| rs2998299 | T | G | 0.21 | -0.014 | 0.002 | 1.51E-15 | 0.004 | 0.009 | 0.676 | 64 |
| rs3026996 | A | C | 0.76 | 0.014 | 0.002 | 5.42E-17 | -0.001 | 0.009 | 0.922 | 74.39 |
| rs303752 | A | G | 0.41 | -0.011 | 0.001 | 1.47E-14 | -0.007 | 0.008 | 0.409 | 61.73 |
| rs3111251 | T | C | 0.43 | -0.009 | 0.001 | 3.88E-10 | 0.003 | 0.008 | 0.71 | 41.33 |
| rs34067381 | T | G | 0.36 | -0.01 | 0.002 | 9.77E-13 | -0.004 | 0.008 | 0.624 | 48.07 |
| rs34098770 | A | G | 0.15 | -0.016 | 0.002 | 2.95E-16 | 0.004 | 0.011 | 0.735 | 64 |
| rs34305371 | A | G | 0.1 | 0.031 | 0.002 | 1.10E-39 | -0.015 | 0.013 | 0.222 | 171.17 |
| rs34394051 | A | G | 0.84 | -0.012 | 0.002 | 2.30E-09 | -0.024 | 0.01 | 0.022 | 34.22 |
| rs34720381 | T | C | 0.09 | -0.018 | 0.002 | 1.90E-13 | 0.018 | 0.013 | 0.187 | 56.25 |
| rs34807077 | A | C | 0.16 | 0.013 | 0.002 | 3.29E-11 | -0.007 | 0.01 | 0.478 | 45.39 |
| rs35417702 | T | C | 0.52 | -0.015 | 0.001 | 2.40E-27 | 0 | 0.008 | 0.993 | 117.88 |
| rs35606437 | A | G | 0.27 | 0.011 | 0.002 | 3.98E-11 | 0.007 | 0.009 | 0.417 | 43.07 |
| rs35929923 | A | G | 0.25 | -0.011 | 0.002 | 1.77E-12 | 0.007 | 0.009 | 0.439 | 50.77 |
| rs363096 | T | C | 0.42 | -0.014 | 0.001 | 5.77E-24 | -0.009 | 0.008 | 0.234 | 104.33 |
| rs3751331 | A | G | 0.38 | -0.01 | 0.001 | 1.75E-11 | 0.009 | 0.008 | 0.238 | 48.01 |
| rs3809634 | A | G | 0.68 | -0.011 | 0.002 | 6.82E-13 | 0.004 | 0.008 | 0.628 | 52.8 |
| rs382196 | T | G | 0.38 | -0.008 | 0.001 | 1.47E-08 | 0.001 | 0.008 | 0.918 | 34.31 |
| rs3859523 | T | C | 0.82 | 0.012 | 0.002 | 9.92E-12 | -0.003 | 0.01 | 0.781 | 47.46 |
| rs3897821 | A | G | 0.67 | 0.015 | 0.002 | 3.58E-25 | -0.003 | 0.008 | 0.726 | 105.4 |
| rs3948495 | T | G | 0.41 | -0.009 | 0.001 | 3.95E-10 | -0.008 | 0.008 | 0.318 | 40.41 |
| rs401526 | T | C | 0.53 | -0.01 | 0.001 | 1.15E-13 | -0.01 | 0.008 | 0.197 | 55.18 |
| rs401966 | C | G | 0.39 | -0.01 | 0.001 | 1.72E-11 | 0.022 | 0.008 | 0.006 | 48.01 |
| rs42210 | C | G | 0.71 | -0.01 | 0.002 | 2.74E-10 | 0 | 0.008 | 0.975 | 37.52 |
| rs42302 | A | G | 0.34 | 0.009 | 0.002 | 5.96E-09 | -0.011 | 0.008 | 0.165 | 32.87 |
| rs4298514 | T | C | 0.7 | 0.01 | 0.002 | 1.07E-11 | 0.013 | 0.008 | 0.115 | 48.07 |
| rs4320563 | A | G | 0.51 | 0.011 | 0.001 | 2.24E-14 | -0.006 | 0.008 | 0.443 | 60.62 |
| rs4328757 | T | C | 0.61 | 0.01 | 0.001 | 2.95E-11 | -0.015 | 0.008 | 0.061 | 47.02 |
| rs4352658 | T | C | 0.08 | -0.019 | 0.003 | 1.71E-13 | 0 | 0.014 | 0.975 | 53.4 |
| rs4358081 | A | C | 0.53 | -0.01 | 0.001 | 1.28E-11 | 0.015 | 0.008 | 0.051 | 46.05 |
| rs4369924 | A | G | 0.16 | 0.013 | 0.002 | 5.58E-11 | 0.012 | 0.01 | 0.269 | 43.28 |
| rs4384309 | A | G | 0.46 | 0.01 | 0.001 | 4.89E-13 | -0.008 | 0.008 | 0.291 | 53.08 |
| rs4467547 | T | G | 0.41 | 0.012 | 0.001 | 4.88E-17 | 0.006 | 0.008 | 0.468 | 74.7 |
| rs4500930 | T | C | 0.34 | -0.011 | 0.002 | 1.67E-13 | 0.004 | 0.008 | 0.631 | 52.8 |
| rs4663617 | A | T | 0.23 | 0.01 | 0.002 | 4.12E-10 | -0.008 | 0.009 | 0.365 | 37.43 |
| rs4673840 | T | C | 0.84 | -0.014 | 0.002 | 5.86E-13 | -0.01 | 0.01 | 0.336 | 52.75 |
| rs4675248 | A | G | 0.4 | -0.009 | 0.001 | 4.50E-11 | -0.005 | 0.008 | 0.566 | 45.08 |
| rs4719944 | T | C | 0.55 | -0.012 | 0.001 | 9.25E-18 | 0.012 | 0.008 | 0.124 | 74.7 |
| rs4726070 | A | G | 0.6 | 0.012 | 0.001 | 1.36E-17 | -0.013 | 0.008 | 0.096 | 75.94 |
| rs4757957 | C | G | 0.69 | 0.013 | 0.002 | 1.79E-18 | -0.006 | 0.008 | 0.473 | 78.62 |
| rs4766424 | C | G | 0.12 | 0.014 | 0.002 | 2.47E-11 | 0.005 | 0.011 | 0.638 | 45.72 |
| rs4778058 | T | C | 0.5 | -0.009 | 0.001 | 4.92E-11 | 0.003 | 0.008 | 0.69 | 43.18 |
| rs4787457 | A | G | 0.63 | 0.016 | 0.002 | 1.23E-28 | 0.014 | 0.008 | 0.075 | 116.64 |
| rs4793090 | A | G | 0.66 | 0.008 | 0.002 | 1.67E-08 | -0.004 | 0.008 | 0.605 | 31.36 |
| rs4810894 | A | G | 0.37 | -0.009 | 0.002 | 4.38E-09 | -0.005 | 0.008 | 0.529 | 32.11 |
| rs4839155 | T | G | 0.77 | 0.012 | 0.002 | 4.66E-12 | 0 | 0.009 | 0.994 | 45.76 |
| rs4846724 | A | G | 0.53 | 0.01 | 0.001 | 2.89E-12 | -0.006 | 0.008 | 0.466 | 49 |
| rs4848924 | A | C | 0.71 | -0.012 | 0.002 | 3.56E-14 | 0.011 | 0.009 | 0.179 | 60.84 |
| rs488476 | C | G | 0.62 | -0.011 | 0.002 | 8.29E-15 | -0.001 | 0.008 | 0.897 | 56.75 |
| rs4895650 | T | C | 0.55 | 0.009 | 0.001 | 5.31E-11 | -0.003 | 0.008 | 0.681 | 44.13 |
| rs4899012 | C | G | 0.6 | -0.01 | 0.001 | 8.06E-13 | -0.004 | 0.008 | 0.614 | 54.13 |
| rs4904523 | A | G | 0.52 | -0.008 | 0.001 | 6.27E-09 | -0.008 | 0.008 | 0.302 | 34.31 |
| rs4915735 | A | G | 0.14 | 0.012 | 0.002 | 9.56E-09 | -0.014 | 0.011 | 0.195 | 33.64 |
| rs4977885 | A | G | 0.4 | -0.008 | 0.001 | 6.78E-09 | 0.008 | 0.008 | 0.317 | 36 |
| rs4984541 | A | G | 0.77 | -0.014 | 0.002 | 3.40E-16 | 0.012 | 0.009 | 0.2 | 65.9 |
| rs55736314 | C | G | 0.6 | -0.015 | 0.001 | 4.59E-27 | 0.007 | 0.008 | 0.361 | 121 |
| rs56099375 | T | C | 0.24 | 0.012 | 0.002 | 3.10E-13 | -0.008 | 0.009 | 0.36 | 49.83 |
| rs56319902 | T | C | 0.22 | -0.021 | 0.002 | 5.81E-33 | 0.018 | 0.009 | 0.056 | 146.84 |
| rs56391344 | A | G | 0.25 | 0.015 | 0.002 | 7.49E-20 | -0.016 | 0.009 | 0.074 | 85.56 |
| rs56794817 | A | G | 0.16 | 0.012 | 0.002 | 1.69E-10 | 0.011 | 0.01 | 0.289 | 41.23 |
| rs57349798 | A | G | 0.41 | 0.009 | 0.001 | 8.63E-11 | 0.001 | 0.008 | 0.939 | 44.13 |
| rs57352738 | A | T | 0.2 | -0.015 | 0.002 | 2.51E-18 | 0.013 | 0.009 | 0.159 | 79.94 |
| rs575113 | A | G | 0.29 | 0.013 | 0.002 | 7.52E-18 | -0.008 | 0.008 | 0.322 | 78.62 |
| rs5754753 | T | C | 0.72 | -0.013 | 0.002 | 6.15E-16 | 0.007 | 0.008 | 0.439 | 62.02 |
| rs57661533 | T | C | 0.13 | -0.011 | 0.002 | 3.24E-08 | 0.024 | 0.011 | 0.036 | 29.47 |
| rs580652 | T | C | 0.9 | -0.015 | 0.002 | 4.65E-10 | -0.013 | 0.013 | 0.317 | 37.52 |
| rs590013 | T | C | 0.68 | 0.01 | 0.002 | 2.47E-11 | -0.006 | 0.008 | 0.468 | 44.44 |
| rs6043521 | T | C | 0.6 | -0.008 | 0.001 | 9.72E-09 | 0.007 | 0.008 | 0.386 | 35.15 |
| rs6065080 | T | C | 0.36 | -0.013 | 0.002 | 7.49E-19 | -0.007 | 0.008 | 0.369 | 75.11 |
| rs6065784 | C | G | 0.7 | 0.011 | 0.002 | 5.78E-13 | 0.014 | 0.008 | 0.109 | 53.78 |
| rs61104616 | A | G | 0.52 | -0.017 | 0.001 | 1.56E-34 | 0.003 | 0.008 | 0.649 | 150.94 |
| rs6123924 | A | G | 0.84 | 0.013 | 0.002 | 3.20E-11 | 0.014 | 0.011 | 0.201 | 46.1 |
| rs61527214 | A | G | 0.41 | 0.011 | 0.001 | 1.13E-14 | 0.001 | 0.008 | 0.861 | 61.73 |
| rs61997667 | T | C | 0.15 | -0.014 | 0.002 | 2.24E-13 | 0.002 | 0.011 | 0.836 | 51.84 |
| rs62155350 | A | G | 0.02 | 0.034 | 0.005 | 1.69E-10 | -0.014 | 0.027 | 0.601 | 40.67 |
| rs62155873 | T | C | 0.12 | -0.015 | 0.002 | 1.65E-12 | 0.016 | 0.012 | 0.182 | 51.7 |
| rs62174974 | A | G | 0.19 | -0.011 | 0.002 | 2.76E-09 | -0.009 | 0.01 | 0.368 | 34.03 |
| rs62179650 | A | G | 0.3 | 0.011 | 0.002 | 5.66E-13 | 0.001 | 0.008 | 0.871 | 49 |
| rs62182994 | T | C | 0.69 | 0.014 | 0.002 | 2.55E-19 | -0.008 | 0.008 | 0.317 | 83.42 |
| rs62379838 | T | C | 0.7 | 0.012 | 0.002 | 1.77E-15 | -0.012 | 0.008 | 0.164 | 65.07 |
| rs628993 | A | G | 0.11 | 0.013 | 0.002 | 3.83E-08 | -0.01 | 0.013 | 0.457 | 30.97 |
| rs6449503 | A | G | 0.5 | 0.018 | 0.001 | 1.84E-39 | 0.002 | 0.008 | 0.799 | 174.62 |
| rs6490618 | T | C | 0.33 | 0.009 | 0.002 | 3.48E-09 | -0.007 | 0.008 | 0.397 | 34.42 |
| rs6493265 | T | C | 0.39 | -0.012 | 0.001 | 2.26E-17 | -0.007 | 0.008 | 0.361 | 75.94 |
| rs6557171 | T | C | 0.32 | -0.016 | 0.002 | 5.48E-25 | -0.004 | 0.008 | 0.583 | 106.78 |
| rs660001 | A | G | 0.21 | -0.016 | 0.002 | 1.66E-21 | 0 | 0.009 | 0.999 | 93.07 |
| rs66568921 | T | G | 0.64 | -0.018 | 0.002 | 7.60E-34 | 0.005 | 0.008 | 0.535 | 145.6 |
| rs66721975 | A | G | 0.29 | -0.009 | 0.002 | 1.51E-08 | 0.01 | 0.009 | 0.266 | 33.64 |
| rs6690195 | T | C | 0.49 | -0.011 | 0.001 | 7.89E-16 | -0.002 | 0.008 | 0.762 | 65.15 |
| rs6697584 | T | C | 0.79 | -0.013 | 0.002 | 1.43E-13 | 0.008 | 0.009 | 0.361 | 54.93 |
| rs6704768 | A | G | 0.56 | -0.012 | 0.001 | 1.84E-16 | 0 | 0.008 | 0.962 | 68.65 |
| rs6731373 | A | G | 0.34 | -0.012 | 0.002 | 1.39E-14 | -0.017 | 0.008 | 0.033 | 58.78 |
| rs6812533 | T | C | 0.74 | 0.011 | 0.002 | 4.85E-11 | 0.003 | 0.009 | 0.693 | 43.89 |
| rs68145588 | T | G | 0.14 | -0.013 | 0.002 | 7.53E-10 | 0 | 0.011 | 0.98 | 36.57 |
| rs6917154 | T | C | 0.12 | 0.013 | 0.002 | 2.55E-09 | 0.005 | 0.011 | 0.646 | 36.57 |
| rs6924023 | A | G | 0.75 | 0.011 | 0.002 | 5.17E-11 | 0 | 0.009 | 0.989 | 44.72 |
| rs6977237 | T | C | 0.47 | 0.008 | 0.001 | 5.59E-09 | 0.015 | 0.008 | 0.052 | 34.31 |
| rs6994287 | A | G | 0.41 | 0.01 | 0.001 | 1.11E-12 | -0.005 | 0.008 | 0.541 | 53.08 |
| rs7012546 | T | C | 0.42 | 0.009 | 0.001 | 1.25E-10 | -0.008 | 0.008 | 0.31 | 43.18 |
| rs7029718 | A | G | 0.41 | 0.024 | 0.001 | 7.82E-65 | -0.001 | 0.008 | 0.929 | 303.76 |
| rs7041702 | A | G | 0.74 | -0.011 | 0.002 | 4.53E-12 | -0.012 | 0.009 | 0.177 | 48.13 |
| rs7167688 | T | C | 0.49 | -0.009 | 0.001 | 1.71E-11 | 0.019 | 0.008 | 0.013 | 45.08 |
| rs7171405 | A | G | 0.24 | -0.01 | 0.002 | 7.53E-10 | 0.009 | 0.009 | 0.305 | 39.85 |
| rs717996 | T | C | 0.6 | -0.012 | 0.001 | 1.89E-17 | -0.01 | 0.008 | 0.206 | 75.94 |
| rs7226824 | T | C | 0.53 | -0.008 | 0.001 | 2.72E-08 | 0.003 | 0.008 | 0.677 | 31.04 |
| rs72486027 | T | C | 0.25 | -0.01 | 0.002 | 1.61E-10 | -0.002 | 0.009 | 0.81 | 42.25 |
| rs7254263 | T | C | 0.28 | -0.012 | 0.002 | 1.84E-14 | 0.016 | 0.008 | 0.064 | 55.32 |
| rs72622559 | T | C | 0.23 | -0.012 | 0.002 | 5.50E-13 | -0.004 | 0.009 | 0.633 | 49.83 |
| rs72672052 | A | T | 0.18 | -0.01 | 0.002 | 4.87E-08 | -0.003 | 0.01 | 0.802 | 31.48 |
| rs72709560 | A | G | 0.69 | 0.009 | 0.002 | 2.03E-08 | 0.01 | 0.008 | 0.229 | 32.11 |
| rs72828517 | T | C | 0.83 | -0.017 | 0.002 | 1.38E-19 | -0.005 | 0.01 | 0.604 | 78.18 |
| rs72829857 | A | G | 0.76 | -0.015 | 0.002 | 1.52E-19 | 0.002 | 0.009 | 0.81 | 86.72 |
| rs72917504 | T | C | 0.06 | -0.018 | 0.003 | 2.91E-09 | -0.023 | 0.016 | 0.161 | 35.2 |
| rs72944064 | T | C | 0.74 | -0.01 | 0.002 | 9.39E-10 | -0.009 | 0.009 | 0.305 | 37.52 |
| rs72962169 | T | C | 0.16 | -0.017 | 0.002 | 1.26E-19 | -0.004 | 0.011 | 0.707 | 83.87 |
| rs72993796 | T | C | 0.88 | 0.013 | 0.002 | 1.12E-08 | 0.019 | 0.012 | 0.121 | 33.32 |
| rs730384 | A | G | 0.44 | 0.01 | 0.001 | 9.38E-13 | -0.013 | 0.008 | 0.096 | 52.05 |
| rs73055556 | A | G | 0.14 | 0.011 | 0.002 | 4.95E-08 | -0.021 | 0.011 | 0.055 | 30.8 |
| rs73191311 | A | G | 0.34 | -0.009 | 0.002 | 2.03E-10 | -0.002 | 0.008 | 0.794 | 39.27 |
| rs7321274 | A | G | 0.8 | 0.011 | 0.002 | 8.21E-11 | -0.001 | 0.01 | 0.903 | 40.11 |
| rs7326331 | A | G | 0.28 | -0.013 | 0.002 | 3.42E-16 | 0.005 | 0.009 | 0.535 | 64 |
| rs73344830 | A | G | 0.42 | 0.017 | 0.001 | 1.00E-32 | 0 | 0.008 | 0.958 | 147.45 |
| rs736281 | T | C | 0.39 | 0.009 | 0.001 | 2.49E-09 | -0.011 | 0.008 | 0.17 | 37.73 |
| rs73648455 | T | C | 0.08 | -0.018 | 0.003 | 1.99E-12 | -0.006 | 0.015 | 0.674 | 49.54 |
| rs743316 | T | C | 0.79 | 0.011 | 0.002 | 8.21E-11 | -0.004 | 0.009 | 0.649 | 43.4 |
| rs74415461 | T | C | 0.08 | 0.018 | 0.003 | 6.25E-12 | 0.027 | 0.013 | 0.042 | 45.82 |
| rs7449561 | A | G | 0.23 | 0.01 | 0.002 | 9.73E-10 | -0.021 | 0.009 | 0.021 | 36 |
| rs7460106 | T | C | 0.76 | -0.012 | 0.002 | 2.06E-13 | 0.009 | 0.009 | 0.337 | 52.35 |
| rs746839 | C | G | 0.63 | 0.014 | 0.002 | 1.19E-20 | 0.001 | 0.008 | 0.898 | 81 |
| rs74787922 | A | C | 0.93 | 0.018 | 0.003 | 8.97E-11 | 0.007 | 0.015 | 0.635 | 42.25 |
| rs75033012 | C | G | 0.04 | -0.026 | 0.004 | 1.38E-11 | -0.015 | 0.02 | 0.447 | 46.1 |
| rs75177132 | T | C | 0.05 | 0.03 | 0.004 | 1.01E-16 | -0.006 | 0.018 | 0.744 | 68.52 |
| rs7575637 | A | G | 0.45 | 0.011 | 0.001 | 3.43E-14 | -0.005 | 0.008 | 0.48 | 58.41 |
| rs7603132 | A | G | 0.19 | 0.014 | 0.002 | 1.20E-14 | -0.004 | 0.01 | 0.703 | 57.93 |
| rs76076331 | T | C | 0.13 | 0.021 | 0.002 | 1.71E-23 | -0.011 | 0.012 | 0.33 | 96.23 |
| rs7617204 | A | G | 0.52 | -0.01 | 0.001 | 4.33E-12 | -0.029 | 0.008 | 1.84E-04 | 48.01 |
| rs76235882 | A | G | 0.97 | 0.024 | 0.004 | 4.52E-10 | -0.031 | 0.023 | 0.186 | 38.19 |
| rs76246107 | A | G | 0.09 | -0.016 | 0.003 | 6.52E-10 | -0.027 | 0.015 | 0.07 | 38.82 |
| rs7650602 | T | C | 0.56 | -0.009 | 0.001 | 2.77E-10 | -0.001 | 0.008 | 0.895 | 40.41 |
| rs76577427 | C | G | 0.9 | 0.02 | 0.002 | 7.78E-18 | -0.001 | 0.013 | 0.918 | 71.54 |
| rs7672622 | A | G | 0.75 | -0.009 | 0.002 | 4.72E-08 | -0.001 | 0.009 | 0.94 | 30.25 |
| rs7683416 | T | C | 0.46 | 0.014 | 0.001 | 6.05E-23 | -0.01 | 0.008 | 0.197 | 98.58 |
| rs76878669 | C | G | 0.76 | 0.014 | 0.002 | 3.10E-17 | -0.008 | 0.009 | 0.37 | 68.79 |
| rs7692359 | T | C | 0.78 | -0.011 | 0.002 | 1.40E-10 | -0.007 | 0.009 | 0.46 | 41.11 |
| rs77025239 | A | G | 0.16 | -0.014 | 0.002 | 4.65E-13 | -0.02 | 0.011 | 0.061 | 54.29 |
| rs77609760 | A | G | 0.93 | 0.024 | 0.003 | 7.71E-17 | 0.014 | 0.015 | 0.351 | 71.04 |
| rs77719387 | A | T | 0.02 | -0.04 | 0.006 | 3.78E-12 | -0.014 | 0.032 | 0.659 | 48.28 |
| rs7849487 | T | G | 0.66 | -0.017 | 0.002 | 1.25E-29 | -0.003 | 0.008 | 0.694 | 123.95 |
| rs78648104 | T | C | 0.91 | -0.014 | 0.002 | 1.70E-08 | 0.032 | 0.014 | 0.018 | 31.81 |
| rs7875078 | A | C | 0.46 | -0.008 | 0.001 | 2.72E-08 | -0.001 | 0.008 | 0.89 | 31.04 |
| rs7894722 | T | C | 0.37 | 0.009 | 0.001 | 5.41E-09 | -0.009 | 0.008 | 0.275 | 36.86 |
| rs790647 | A | C | 0.23 | -0.015 | 0.002 | 3.81E-18 | 0.014 | 0.009 | 0.122 | 72.75 |
| rs7910403 | T | G | 0.2 | 0.012 | 0.002 | 5.08E-12 | 0.004 | 0.01 | 0.696 | 45.94 |
| rs7924036 | T | G | 0.51 | 0.013 | 0.001 | 2.55E-21 | -0.004 | 0.008 | 0.584 | 90.25 |
| rs79265434 | A | G | 0.88 | -0.02 | 0.002 | 1.98E-19 | 0.004 | 0.012 | 0.718 | 80.18 |
| rs7972246 | T | C | 0.35 | 0.011 | 0.002 | 1.01E-13 | 0.004 | 0.008 | 0.653 | 53.78 |
| rs79728014 | A | G | 0.86 | -0.013 | 0.002 | 2.06E-11 | -0.005 | 0.011 | 0.668 | 44.89 |
| rs7974852 | A | C | 0.52 | 0.012 | 0.001 | 1.14E-16 | -0.023 | 0.008 | 0.003 | 69.84 |
| rs7977614 | A | G | 0.71 | -0.013 | 0.002 | 6.84E-17 | 0.001 | 0.008 | 0.874 | 70.14 |
| rs79994730 | T | C | 0.04 | 0.024 | 0.004 | 5.88E-12 | -0.03 | 0.018 | 0.084 | 47.02 |
| rs8008382 | T | C | 0.3 | -0.011 | 0.002 | 4.07E-12 | -0.01 | 0.008 | 0.246 | 49.94 |
| rs8024 | A | C | 0.33 | -0.012 | 0.002 | 1.13E-14 | 0 | 0.008 | 0.952 | 58.78 |
| rs80257979 | T | G | 0.97 | -0.025 | 0.004 | 3.31E-10 | 0.002 | 0.022 | 0.913 | 39.69 |
| rs8030487 | A | G | 0.69 | 0.009 | 0.002 | 2.52E-08 | -0.01 | 0.008 | 0.233 | 32.11 |
| rs806816 | A | T | 0.75 | 0.01 | 0.002 | 4.80E-10 | -0.001 | 0.009 | 0.928 | 39.85 |
| rs8097125 | T | C | 0.38 | 0.009 | 0.001 | 2.31E-09 | -0.005 | 0.008 | 0.499 | 37.73 |
| rs912883 | T | C | 0.68 | 0.009 | 0.002 | 6.20E-09 | 0.005 | 0.008 | 0.578 | 33.64 |
| rs9289300 | T | C | 0.84 | -0.016 | 0.002 | 1.00E-17 | 0.002 | 0.011 | 0.88 | 74.5 |
| rs929511 | T | C | 0.13 | -0.016 | 0.002 | 3.91E-14 | 0.017 | 0.012 | 0.144 | 58.05 |
| rs9359939 | A | C | 0.24 | -0.011 | 0.002 | 1.51E-10 | -0.002 | 0.009 | 0.818 | 43.07 |
| rs936496 | A | G | 0.62 | -0.008 | 0.001 | 4.38E-08 | 0.009 | 0.008 | 0.242 | 31.84 |
| rs9373363 | A | G | 0.75 | -0.011 | 0.002 | 5.54E-12 | 0.001 | 0.009 | 0.936 | 48.13 |
| rs9375188 | T | C | 0.48 | 0.021 | 0.001 | 8.78E-52 | 0.001 | 0.008 | 0.872 | 229.31 |
| rs9411331 | A | C | 0.68 | 0.015 | 0.002 | 7.63E-22 | 0.003 | 0.008 | 0.715 | 93.44 |
| rs9492774 | C | G | 0.37 | 0.008 | 0.002 | 4.33E-08 | 0 | 0.008 | 0.986 | 28.44 |
| rs9529146 | T | C | 0.23 | 0.013 | 0.002 | 3.48E-14 | -0.028 | 0.009 | 0.002 | 55.81 |
| rs9556958 | T | C | 0.52 | -0.011 | 0.001 | 5.96E-15 | 0.009 | 0.008 | 0.225 | 61.73 |
| rs9616947 | T | C | 0.38 | -0.013 | 0.002 | 3.58E-20 | 0.011 | 0.008 | 0.15 | 79.8 |
| rs9633970 | T | C | 0.76 | 0.009 | 0.002 | 2.81E-08 | -0.021 | 0.009 | 0.023 | 32.35 |
| rs9655780 | A | G | 0.83 | -0.015 | 0.002 | 3.70E-16 | 0.004 | 0.01 | 0.723 | 64.84 |
| rs969512 | A | T | 0.66 | -0.011 | 0.002 | 9.84E-13 | 0.001 | 0.008 | 0.888 | 49.94 |
| rs9853928 | T | C | 0.21 | -0.013 | 0.002 | 1.59E-13 | -0.004 | 0.01 | 0.707 | 55.81 |
| rs9859556 | T | G | 0.31 | 0.029 | 0.002 | 4.61E-82 | -0.002 | 0.008 | 0.797 | 373.78 |
| rs9882532 | T | C | 0.64 | 0.012 | 0.002 | 2.15E-17 | 0.003 | 0.008 | 0.711 | 68.34 |
| rs9886703 | A | T | 0.17 | -0.014 | 0.002 | 4.46E-13 | -0.002 | 0.01 | 0.851 | 51.24 |
| rs9927137 | A | G | 0.53 | 0.008 | 0.001 | 5.11E-09 | -0.004 | 0.008 | 0.558 | 34.31 |
| rs9927842 | T | C | 0.15 | -0.013 | 0.002 | 6.22E-11 | 0.019 | 0.011 | 0.066 | 42.25 |
| rs9929556 | T | G | 0.57 | -0.01 | 0.001 | 2.31E-12 | -0.005 | 0.008 | 0.492 | 50.01 |
| rs9929762 | A | G | 0.56 | 0.01 | 0.001 | 1.03E-11 | 0.01 | 0.008 | 0.203 | 47.02 |
| rs9933256 | A | G | 0.56 | 0.012 | 0.001 | 6.06E-17 | -0.006 | 0.008 | 0.446 | 72.25 |

**SNPs used as instrumental variables in the MR analysis of educational attainment on mean diffusivity (cortical)**

| **SNP** | **Effect allele** | **Other allele** | **Effect allele frequency** | **beta (exposure)** | **SE (exposure)** | ***P* (exposure)** | **beta (outcome)** | **SE (outcome)** | ***P* (outcome)** | ***F*-statistic** |
| --- | --- | --- | --- | --- | --- | --- | --- | --- | --- | --- |
| rs10006235 | T | C | 0.27 | -0.009 | 0.002 | 4.48E-09 | 0.002 | 0.004 | 0.611 | 33.79 |
| rs10021733 | T | C | 0.15 | -0.016 | 0.003 | 9.45E-10 | 0.001 | 0.004 | 0.79 | 36.89 |
| rs10046069 | A | G | 0.12 | 0.013 | 0.002 | 4.40E-09 | -0.002 | 0.004 | 0.649 | 33.32 |
| rs10060023 | T | C | 0.33 | 0.011 | 0.002 | 1.88E-13 | 0.001 | 0.004 | 0.822 | 53.78 |
| rs10080647 | A | C | 0.14 | 0.012 | 0.002 | 7.05E-10 | 0.003 | 0.004 | 0.452 | 38.44 |
| rs1008078 | T | C | 0.4 | -0.018 | 0.001 | 4.50E-37 | 0.006 | 0.004 | 0.189 | 169 |
| rs10189857 | A | G | 0.57 | 0.016 | 0.001 | 4.69E-29 | 0.013 | 0.004 | 0.003 | 127.37 |
| rs10193498 | A | T | 0.76 | 0.01 | 0.002 | 3.10E-09 | -0.010 | 0.004 | 0.025 | 36.75 |
| rs10215082 | A | G | 0.47 | -0.015 | 0.001 | 7.03E-26 | 0.002 | 0.004 | 0.566 | 113.27 |
| rs10460095 | A | G | 0.57 | -0.012 | 0.001 | 2.61E-18 | 0.006 | 0.004 | 0.195 | 78.45 |
| rs10761251 | A | T | 0.66 | 0.011 | 0.002 | 2.52E-14 | -0.005 | 0.004 | 0.253 | 56.75 |
| rs10772644 | C | G | 0.89 | 0.014 | 0.002 | 1.69E-10 | -0.004 | 0.004 | 0.371 | 41.66 |
| rs10773002 | A | T | 0.25 | 0.018 | 0.002 | 2.50E-30 | 0.017 | 0.004 | 8.37E-05 | 133.69 |
| rs10773208 | T | C | 0.25 | -0.011 | 0.002 | 8.09E-12 | -0.002 | 0.004 | 0.68 | 47.27 |
| rs10798888 | T | G | 0.17 | -0.014 | 0.002 | 5.15E-14 | 0.004 | 0.004 | 0.412 | 54.29 |
| rs10799615 | A | G | 0.25 | -0.009 | 0.002 | 4.11E-08 | 0.002 | 0.004 | 0.616 | 30.94 |
| rs10805383 | A | G | 0.49 | -0.01 | 0.001 | 2.22E-13 | 0.004 | 0.004 | 0.313 | 54.13 |
| rs10830858 | T | C | 0.51 | 0.009 | 0.001 | 5.31E-11 | 0.003 | 0.004 | 0.554 | 43.18 |
| rs10844179 | A | G | 0.23 | 0.01 | 0.002 | 5.03E-10 | 0.002 | 0.004 | 0.665 | 37.43 |
| rs10875121 | C | G | 0.84 | 0.017 | 0.002 | 4.46E-20 | 0.001 | 0.004 | 0.753 | 83.87 |
| rs10892807 | T | C | 0.57 | 0.011 | 0.001 | 1.09E-14 | -0.004 | 0.004 | 0.312 | 60.62 |
| rs10940921 | T | G | 0.42 | 0.011 | 0.001 | 1.31E-14 | 0.000 | 0.004 | 0.928 | 62.86 |
| rs10951590 | T | C | 0.33 | -0.011 | 0.002 | 3.62E-13 | 0.003 | 0.004 | 0.505 | 52.8 |
| rs10979613 | T | C | 0.64 | -0.012 | 0.002 | 1.25E-15 | 0.000 | 0.004 | 0.991 | 60.84 |
| rs10996167 | C | G | 0.65 | 0.011 | 0.002 | 7.47E-09 | 0.002 | 0.004 | 0.637 | 31.92 |
| rs11003463 | T | G | 0.61 | 0.009 | 0.002 | 5.58E-10 | 0.000 | 0.004 | 0.983 | 36.8 |
| rs11023764 | A | G | 0.64 | 0.009 | 0.002 | 5.32E-10 | 0.002 | 0.004 | 0.649 | 36.8 |
| rs11030102 | C | G | 0.75 | 0.01 | 0.002 | 4.60E-10 | 0.001 | 0.004 | 0.883 | 39.85 |
| rs11076962 | T | C | 0.72 | 0.009 | 0.002 | 4.29E-08 | -0.003 | 0.004 | 0.553 | 28.89 |
| rs11081529 | T | C | 0.71 | 0.013 | 0.002 | 5.22E-16 | 0.001 | 0.004 | 0.815 | 70.56 |
| rs11082011 | T | C | 0.67 | 0.019 | 0.002 | 2.06E-38 | 0.000 | 0.004 | 0.937 | 167.27 |
| rs11121177 | A | G | 0.18 | 0.015 | 0.002 | 1.44E-16 | -0.001 | 0.004 | 0.76 | 70.37 |
| rs11130380 | T | G | 0.61 | 0.009 | 0.001 | 4.17E-10 | 0.002 | 0.004 | 0.712 | 41.33 |
| rs11138947 | T | C | 0.72 | 0.011 | 0.002 | 2.36E-11 | -0.006 | 0.004 | 0.17 | 43.07 |
| rs11157931 | A | C | 0.39 | -0.014 | 0.001 | 3.86E-22 | -0.012 | 0.004 | 0.007 | 98.58 |
| rs111852224 | T | C | 0.12 | 0.016 | 0.002 | 4.85E-13 | 0.000 | 0.004 | 0.966 | 50.28 |
| rs11211123 | A | G | 0.22 | 0.01 | 0.002 | 7.62E-10 | 0.004 | 0.004 | 0.337 | 37.43 |
| rs11213482 | A | G | 0.84 | -0.011 | 0.002 | 2.68E-08 | 0.001 | 0.004 | 0.775 | 31.12 |
| rs112603734 | A | C | 0.77 | -0.013 | 0.002 | 2.72E-09 | 0.002 | 0.004 | 0.72 | 35.46 |
| rs112806496 | C | G | 0.91 | -0.016 | 0.002 | 1.72E-10 | -0.004 | 0.004 | 0.383 | 40.96 |
| rs1128956 | T | G | 0.83 | -0.014 | 0.002 | 5.34E-14 | -0.002 | 0.004 | 0.602 | 54.29 |
| rs113520408 | A | G | 0.28 | 0.016 | 0.002 | 1.03E-22 | 0.002 | 0.004 | 0.624 | 93.85 |
| rs113615161 | T | C | 0.13 | -0.012 | 0.002 | 2.98E-09 | 0.001 | 0.004 | 0.872 | 34.31 |
| rs114593137 | A | T | 0.79 | -0.012 | 0.002 | 1.65E-12 | -0.002 | 0.004 | 0.641 | 51.5 |
| rs115000530 | A | T | 0.95 | -0.025 | 0.003 | 1.09E-15 | 0.012 | 0.004 | 0.005 | 65.04 |
| rs11542663 | A | C | 0.69 | 0.011 | 0.002 | 2.44E-13 | 0.008 | 0.004 | 0.081 | 55.75 |
| rs11588857 | A | G | 0.21 | 0.02 | 0.002 | 4.45E-31 | -0.001 | 0.004 | 0.755 | 137.03 |
| rs11598765 | A | G | 0.2 | -0.011 | 0.002 | 1.73E-09 | 0.008 | 0.004 | 0.063 | 34.68 |
| rs11620355 | A | G | 0.09 | 0.018 | 0.002 | 1.09E-13 | 0.010 | 0.004 | 0.029 | 53 |
| rs11657342 | A | G | 0.37 | 0.014 | 0.002 | 2.32E-13 | -0.004 | 0.004 | 0.412 | 54.29 |
| rs11657979 | A | G | 0.24 | -0.009 | 0.002 | 4.57E-08 | 0.001 | 0.005 | 0.905 | 28.65 |
| rs11663602 | A | C | 0.28 | -0.013 | 0.002 | 9.27E-17 | 0.002 | 0.004 | 0.587 | 66.02 |
| rs1167827 | A | G | 0.43 | 0.01 | 0.001 | 2.93E-13 | -0.001 | 0.004 | 0.783 | 55.18 |
| rs11724690 | T | G | 0.29 | 0.009 | 0.002 | 2.08E-09 | -0.002 | 0.004 | 0.572 | 38.44 |
| rs117398064 | C | G | 0.09 | -0.014 | 0.002 | 1.16E-08 | 0.000 | 0.004 | 0.986 | 31.81 |
| rs118134876 | T | C | 0.06 | -0.022 | 0.003 | 1.50E-13 | 0.002 | 0.004 | 0.598 | 55.25 |
| rs11871429 | A | G | 0.77 | 0.013 | 0.002 | 2.52E-14 | -0.001 | 0.004 | 0.776 | 56.69 |
| rs11894424 | A | C | 0.95 | 0.021 | 0.003 | 2.14E-10 | -0.004 | 0.004 | 0.322 | 40.11 |
| rs12054166 | C | G | 0.74 | 0.01 | 0.002 | 2.30E-09 | 0.002 | 0.004 | 0.656 | 36 |
| rs12076635 | C | G | 0.78 | 0.021 | 0.002 | 3.33E-34 | -0.004 | 0.004 | 0.362 | 148.27 |
| rs12113634 | T | C | 0.65 | 0.009 | 0.002 | 2.50E-09 | -0.005 | 0.004 | 0.296 | 34.42 |
| rs12151248 | T | C | 0.11 | -0.015 | 0.002 | 3.81E-11 | 0.005 | 0.004 | 0.27 | 41.97 |
| rs1220779 | A | G | 0.47 | -0.008 | 0.001 | 1.58E-08 | -0.002 | 0.004 | 0.584 | 31.84 |
| rs12273435 | A | G | 0.21 | -0.013 | 0.002 | 4.72E-14 | -0.006 | 0.004 | 0.154 | 60.29 |
| rs12332731 | A | T | 0.19 | 0.014 | 0.002 | 5.96E-14 | 0.007 | 0.004 | 0.101 | 56.25 |
| rs12375949 | T | C | 0.43 | -0.014 | 0.001 | 1.61E-22 | -0.001 | 0.004 | 0.792 | 97.16 |
| rs12438177 | A | G | 0.37 | 0.009 | 0.002 | 1.48E-10 | -0.005 | 0.004 | 0.205 | 38.44 |
| rs12468040 | T | G | 0.38 | 0.014 | 0.001 | 3.12E-21 | -0.002 | 0.004 | 0.716 | 95.76 |
| rs12503522 | T | C | 0.28 | -0.011 | 0.002 | 3.93E-13 | 0.003 | 0.004 | 0.433 | 49.88 |
| rs12506221 | T | G | 0.44 | -0.013 | 0.001 | 4.23E-20 | -0.006 | 0.004 | 0.137 | 86.22 |
| rs12519073 | T | C | 0.23 | -0.011 | 0.002 | 1.16E-10 | 0.003 | 0.004 | 0.544 | 39.62 |
| rs12524795 | T | C | 0.44 | 0.01 | 0.001 | 5.28E-13 | -0.009 | 0.004 | 0.04 | 53.08 |
| rs12571549 | A | G | 0.14 | 0.015 | 0.002 | 1.45E-13 | -0.005 | 0.004 | 0.232 | 57 |
| rs12574281 | A | C | 0.63 | -0.008 | 0.002 | 3.04E-08 | 0.002 | 0.004 | 0.609 | 28.44 |
| rs12591647 | T | C | 0.82 | -0.015 | 0.002 | 7.78E-17 | 0.001 | 0.004 | 0.738 | 71.31 |
| rs12602286 | T | G | 0.87 | 0.016 | 0.002 | 4.62E-14 | 0.009 | 0.004 | 0.034 | 58.05 |
| rs12614263 | A | G | 0.5 | 0.008 | 0.001 | 3.38E-09 | 0.005 | 0.004 | 0.213 | 35.15 |
| rs12638072 | A | G | 0.3 | -0.011 | 0.002 | 3.13E-12 | 0.009 | 0.004 | 0.045 | 50.88 |
| rs12643771 | T | C | 0.31 | 0.013 | 0.002 | 1.32E-17 | -0.001 | 0.004 | 0.8 | 75.11 |
| rs12646523 | T | C | 0.25 | -0.013 | 0.002 | 3.79E-16 | -0.009 | 0.004 | 0.034 | 68.06 |
| rs12670376 | A | G | 0.45 | 0.009 | 0.001 | 5.99E-11 | -0.007 | 0.004 | 0.115 | 43.18 |
| rs12761761 | T | C | 0.24 | 0.015 | 0.002 | 4.79E-20 | 0.002 | 0.004 | 0.697 | 81 |
| rs12765185 | A | T | 0.28 | -0.009 | 0.002 | 2.96E-08 | 0.002 | 0.004 | 0.605 | 30.25 |
| rs12789313 | T | C | 0.49 | 0.009 | 0.001 | 6.79E-11 | -0.011 | 0.004 | 0.015 | 43.18 |
| rs12875339 | A | C | 0.64 | -0.011 | 0.002 | 1.71E-13 | 0.005 | 0.004 | 0.283 | 51.84 |
| rs12888615 | T | C | 0.2 | 0.012 | 0.002 | 9.42E-12 | 0.008 | 0.004 | 0.065 | 44.44 |
| rs12957463 | A | G | 0.8 | 0.015 | 0.002 | 9.20E-17 | -0.006 | 0.004 | 0.202 | 65.79 |
| rs12967010 | T | C | 0.77 | 0.011 | 0.002 | 1.33E-11 | -0.005 | 0.004 | 0.235 | 44.97 |
| rs12981405 | T | C | 0.17 | -0.011 | 0.002 | 2.75E-09 | 0.002 | 0.004 | 0.625 | 34.75 |
| rs1301838 | T | C | 0.32 | -0.01 | 0.002 | 1.29E-10 | -0.002 | 0.004 | 0.664 | 40.96 |
| rs13018640 | T | C | 0.6 | -0.021 | 0.001 | 1.05E-50 | -0.003 | 0.004 | 0.472 | 235.84 |
| rs13050131 | A | C | 0.66 | -0.008 | 0.002 | 4.40E-08 | 0.006 | 0.004 | 0.199 | 29.16 |
| rs13133213 | A | G | 0.5 | 0.01 | 0.001 | 7.57E-12 | -0.012 | 0.004 | 0.004 | 47.02 |
| rs13145650 | T | C | 0.92 | -0.017 | 0.002 | 8.99E-12 | -0.002 | 0.004 | 0.591 | 47.89 |
| rs13163845 | T | C | 0.85 | -0.015 | 0.002 | 4.66E-14 | 0.015 | 0.004 | 6.37E-04 | 55.5 |
| rs13197257 | T | G | 0.28 | 0.011 | 0.002 | 2.83E-12 | 0.013 | 0.004 | 0.004 | 46.41 |
| rs13212041 | T | C | 0.8 | 0.01 | 0.002 | 7.42E-09 | -0.008 | 0.004 | 0.08 | 31.48 |
| rs13240401 | T | C | 0.78 | 0.018 | 0.002 | 2.36E-24 | 0.002 | 0.004 | 0.567 | 107.18 |
| rs13261773 | C | G | 0.16 | 0.012 | 0.002 | 3.12E-10 | -0.001 | 0.004 | 0.827 | 41.23 |
| rs13266287 | A | T | 0.67 | -0.009 | 0.002 | 1.75E-08 | -0.003 | 0.004 | 0.502 | 32.11 |
| rs1329125 | T | C | 0.33 | -0.01 | 0.002 | 8.37E-11 | 0.003 | 0.004 | 0.513 | 41.82 |
| rs13327482 | A | G | 0.82 | -0.012 | 0.002 | 4.47E-11 | -0.003 | 0.004 | 0.498 | 45.19 |
| rs1334297 | A | G | 0.74 | 0.026 | 0.002 | 2.53E-58 | 0.001 | 0.004 | 0.791 | 258 |
| rs13422673 | T | C | 0.47 | -0.011 | 0.001 | 6.48E-15 | 0.002 | 0.004 | 0.666 | 61.73 |
| rs137079 | T | C | 0.14 | 0.013 | 0.002 | 3.06E-10 | 0.003 | 0.004 | 0.473 | 40.96 |
| rs1426619 | T | C | 0.43 | 0.01 | 0.001 | 1.40E-11 | -0.003 | 0.004 | 0.523 | 47.02 |
| rs1434630 | T | G | 0.15 | -0.014 | 0.002 | 4.63E-12 | -0.007 | 0.004 | 0.102 | 46.92 |
| rs143743568 | A | G | 0.15 | 0.012 | 0.002 | 4.99E-09 | 0.003 | 0.004 | 0.493 | 34.81 |
| rs150537577 | A | G | 0.92 | -0.015 | 0.003 | 9.50E-09 | -0.011 | 0.004 | 0.015 | 33.73 |
| rs1505676 | C | G | 0.63 | 0.008 | 0.002 | 6.67E-09 | -0.001 | 0.004 | 0.745 | 31.36 |
| rs1542354 | A | G | 0.48 | -0.008 | 0.001 | 2.72E-09 | -0.009 | 0.004 | 0.031 | 36 |
| rs1544 | A | G | 0.26 | -0.01 | 0.002 | 1.76E-09 | 0.001 | 0.004 | 0.853 | 36 |
| rs1564347 | T | G | 0.34 | 0.009 | 0.002 | 2.39E-09 | 0.003 | 0.004 | 0.525 | 34.42 |
| rs1603460 | T | G | 0.42 | 0.009 | 0.001 | 2.20E-10 | 0.001 | 0.004 | 0.805 | 41.33 |
| rs162445 | A | G | 0.08 | 0.015 | 0.003 | 5.72E-09 | -0.006 | 0.004 | 0.156 | 34.18 |
| rs1689510 | C | G | 0.33 | 0.019 | 0.002 | 1.35E-38 | -0.002 | 0.004 | 0.672 | 167.27 |
| rs17048855 | A | G | 0.34 | 0.011 | 0.002 | 3.10E-13 | 0.003 | 0.004 | 0.476 | 51.84 |
| rs17110109 | T | C | 0.62 | -0.009 | 0.001 | 1.47E-10 | 0.004 | 0.004 | 0.326 | 44.13 |
| rs17144467 | A | C | 0.69 | 0.01 | 0.002 | 8.74E-11 | -0.002 | 0.004 | 0.626 | 42.68 |
| rs17148998 | A | G | 0.2 | 0.01 | 0.002 | 6.98E-09 | -0.005 | 0.004 | 0.279 | 35.3 |
| rs1717204 | A | C | 0.18 | -0.012 | 0.002 | 1.20E-10 | -0.007 | 0.004 | 0.117 | 42.25 |
| rs17248751 | A | G | 0.78 | -0.014 | 0.002 | 1.28E-15 | 0.007 | 0.004 | 0.11 | 64 |
| rs173003 | A | C | 0.49 | -0.008 | 0.001 | 5.50E-09 | 0.011 | 0.004 | 0.012 | 34.31 |
| rs1738050 | C | G | 0.62 | -0.009 | 0.001 | 1.42E-10 | -0.003 | 0.004 | 0.453 | 44.13 |
| rs17411339 | A | G | 0.56 | 0.014 | 0.001 | 5.10E-23 | -0.002 | 0.004 | 0.597 | 100 |
| rs17428076 | C | G | 0.76 | 0.014 | 0.002 | 6.41E-17 | 0.004 | 0.004 | 0.297 | 73.32 |
| rs1747714 | T | C | 0.53 | 0.011 | 0.001 | 3.82E-14 | -0.003 | 0.004 | 0.56 | 57.33 |
| rs1747817 | T | C | 0.76 | -0.009 | 0.002 | 2.29E-08 | 0.003 | 0.004 | 0.492 | 33.06 |
| rs17489649 | A | G | 0.68 | 0.013 | 0.002 | 2.77E-17 | 0.002 | 0.004 | 0.643 | 71.68 |
| rs17502934 | T | G | 0.15 | -0.017 | 0.002 | 2.12E-17 | 0.000 | 0.004 | 0.92 | 69.72 |
| rs175325 | A | T | 0.6 | -0.01 | 0.001 | 2.24E-12 | 0.008 | 0.004 | 0.061 | 51.02 |
| rs17551064 | A | G | 0.84 | 0.014 | 0.002 | 1.53E-13 | -0.007 | 0.004 | 0.128 | 55.07 |
| rs17563464 | A | C | 0.22 | -0.015 | 0.002 | 1.74E-17 | 0.003 | 0.004 | 0.446 | 74.77 |
| rs17565975 | A | G | 0.56 | -0.011 | 0.001 | 5.54E-14 | -0.006 | 0.004 | 0.183 | 57.33 |
| rs17598675 | T | C | 0.51 | -0.012 | 0.001 | 1.37E-16 | -0.010 | 0.004 | 0.027 | 68.65 |
| rs17604349 | A | G | 0.19 | 0.013 | 0.002 | 2.36E-12 | 0.003 | 0.004 | 0.514 | 49.78 |
| rs176218 | T | G | 0.2 | 0.02 | 0.002 | 1.10E-29 | 0.008 | 0.004 | 0.056 | 123.46 |
| rs17732878 | T | C | 0.79 | -0.01 | 0.002 | 4.06E-08 | -0.003 | 0.004 | 0.465 | 31.23 |
| rs178183 | T | C | 0.25 | 0.015 | 0.002 | 4.81E-19 | 0.004 | 0.004 | 0.336 | 82.13 |
| rs17882802 | A | G | 0.43 | 0.008 | 0.001 | 3.00E-09 | -0.003 | 0.004 | 0.54 | 36 |
| rs182902112 | A | C | 0.02 | -0.03 | 0.005 | 1.17E-08 | 0.005 | 0.004 | 0.238 | 32.04 |
| rs1842713 | A | G | 0.79 | -0.012 | 0.002 | 9.67E-12 | -0.002 | 0.004 | 0.72 | 47.37 |
| rs1861786 | A | G | 0.38 | -0.008 | 0.001 | 2.39E-08 | -0.002 | 0.004 | 0.576 | 33.47 |
| rs1865955 | T | C | 0.82 | 0.012 | 0.002 | 4.06E-11 | 0.005 | 0.004 | 0.238 | 41.23 |
| rs1866823 | A | G | 0.54 | 0.011 | 0.001 | 3.09E-14 | -0.002 | 0.004 | 0.609 | 58.41 |
| rs1890132 | T | C | 0.71 | -0.009 | 0.002 | 6.56E-09 | -0.003 | 0.004 | 0.522 | 35.2 |
| rs192436652 | T | C | 0.03 | -0.032 | 0.004 | 2.06E-12 | -0.001 | 0.004 | 0.792 | 50.25 |
| rs1933264 | T | C | 0.75 | 0.01 | 0.002 | 5.85E-10 | -0.012 | 0.004 | 0.005 | 39.06 |
| rs1955250 | A | C | 0.91 | -0.014 | 0.002 | 2.26E-08 | 0.008 | 0.004 | 0.053 | 31.36 |
| rs1991585 | T | C | 0.71 | -0.01 | 0.002 | 6.26E-11 | -0.003 | 0.004 | 0.425 | 45.34 |
| rs2007655 | T | G | 0.5 | 0.009 | 0.001 | 8.64E-10 | 0.011 | 0.004 | 0.01 | 37.73 |
| rs2011603 | A | G | 0.74 | -0.009 | 0.002 | 1.12E-08 | -0.003 | 0.004 | 0.512 | 32.35 |
| rs2034631 | T | C | 0.65 | 0.009 | 0.002 | 2.64E-09 | 0.002 | 0.004 | 0.693 | 35.2 |
| rs2052285 | A | G | 0.6 | 0.011 | 0.001 | 7.25E-15 | -0.005 | 0.004 | 0.291 | 64 |
| rs2055940 | A | G | 0.32 | 0.008 | 0.002 | 4.95E-08 | 0.001 | 0.004 | 0.847 | 29.88 |
| rs2081652 | A | T | 0.66 | 0.012 | 0.002 | 1.09E-16 | 0.002 | 0.004 | 0.709 | 67.24 |
| rs2131167 | A | G | 0.45 | 0.008 | 0.001 | 1.52E-08 | 0.002 | 0.004 | 0.607 | 32.65 |
| rs2179152 | T | C | 0.37 | -0.013 | 0.002 | 1.68E-19 | 0.010 | 0.004 | 0.021 | 76.27 |
| rs2183271 | T | C | 0.64 | 0.008 | 0.002 | 1.91E-08 | -0.011 | 0.004 | 0.011 | 31.36 |
| rs2199409 | T | C | 0.8 | 0.01 | 0.002 | 3.14E-09 | -0.011 | 0.004 | 0.014 | 36.71 |
| rs2212430 | T | C | 0.3 | -0.01 | 0.002 | 1.32E-10 | 0.004 | 0.004 | 0.407 | 39.06 |
| rs2216144 | T | C | 0.53 | 0.01 | 0.001 | 2.58E-12 | 0.005 | 0.004 | 0.256 | 49 |
| rs2256965 | A | G | 0.42 | 0.011 | 0.001 | 2.40E-13 | 0.004 | 0.004 | 0.373 | 57.33 |
| rs2287838 | A | G | 0.55 | -0.01 | 0.001 | 1.27E-13 | 0.006 | 0.004 | 0.203 | 55.18 |
| rs2290601 | T | C | 0.77 | -0.015 | 0.002 | 4.49E-19 | 0.002 | 0.004 | 0.659 | 76.82 |
| rs2297293 | C | G | 0.31 | 0.01 | 0.002 | 8.80E-11 | -0.007 | 0.004 | 0.119 | 43.56 |
| rs232496 | T | C | 0.37 | 0.009 | 0.002 | 7.14E-10 | 0.002 | 0.004 | 0.612 | 36 |
| rs2364544 | A | G | 0.39 | -0.011 | 0.001 | 2.91E-13 | -0.001 | 0.004 | 0.898 | 56.25 |
| rs2434672 | A | C | 0.53 | 0.009 | 0.001 | 1.06E-10 | 0.004 | 0.004 | 0.334 | 42.25 |
| rs2436760 | T | C | 0.97 | 0.023 | 0.004 | 4.52E-08 | 0.004 | 0.004 | 0.419 | 30.51 |
| rs2447097 | T | G | 0.46 | 0.01 | 0.001 | 1.10E-11 | -0.004 | 0.004 | 0.414 | 47.02 |
| rs2469226 | A | T | 0.22 | 0.01 | 0.002 | 1.28E-08 | 0.006 | 0.004 | 0.205 | 31.89 |
| rs2496482 | T | C | 0.42 | 0.012 | 0.002 | 9.20E-17 | 0.009 | 0.004 | 0.05 | 65.07 |
| rs252991 | A | G | 0.37 | 0.01 | 0.002 | 6.30E-11 | -0.012 | 0.004 | 0.007 | 40.11 |
| rs2545795 | A | C | 0.44 | 0.013 | 0.001 | 1.03E-18 | 0.000 | 0.004 | 0.968 | 79.72 |
| rs2554835 | A | G | 0.4 | 0.009 | 0.001 | 8.95E-10 | 0.004 | 0.004 | 0.315 | 39.51 |
| rs2570497 | T | C | 0.64 | -0.012 | 0.002 | 1.27E-16 | -0.006 | 0.004 | 0.204 | 65.07 |
| rs2725370 | T | C | 0.3 | -0.014 | 0.002 | 4.96E-20 | -0.005 | 0.004 | 0.219 | 88.36 |
| rs2764684 | T | C | 0.83 | 0.014 | 0.002 | 9.99E-15 | -0.001 | 0.004 | 0.889 | 56.65 |
| rs2838006 | T | C | 0.36 | 0.013 | 0.002 | 1.38E-19 | 0.001 | 0.004 | 0.777 | 77.44 |
| rs28513882 | A | G | 0.18 | -0.011 | 0.002 | 2.03E-09 | 0.001 | 0.004 | 0.903 | 37.35 |
| rs2885198 | A | G | 0.53 | 0.009 | 0.001 | 4.63E-10 | 0.013 | 0.004 | 0.002 | 39.51 |
| rs2923424 | A | G | 0.61 | 0.012 | 0.001 | 6.31E-17 | -0.002 | 0.004 | 0.728 | 73.47 |
| rs2929032 | A | G | 0.47 | -0.008 | 0.001 | 8.40E-09 | -0.004 | 0.004 | 0.382 | 33.47 |
| rs2958182 | A | T | 0.34 | 0.008 | 0.002 | 3.46E-08 | 0.004 | 0.004 | 0.38 | 29.88 |
| rs2964199 | T | C | 0.31 | -0.01 | 0.002 | 4.61E-11 | 0.008 | 0.004 | 0.062 | 43.56 |
| rs2964255 | A | G | 0.31 | 0.009 | 0.002 | 6.34E-09 | 0.002 | 0.004 | 0.626 | 34.42 |
| rs2998299 | T | G | 0.21 | -0.014 | 0.002 | 1.51E-15 | -0.003 | 0.004 | 0.552 | 64 |
| rs3026996 | A | C | 0.76 | 0.014 | 0.002 | 5.42E-17 | -0.005 | 0.004 | 0.219 | 74.39 |
| rs303752 | A | G | 0.41 | -0.011 | 0.001 | 1.47E-14 | -0.008 | 0.004 | 0.086 | 61.73 |
| rs3111251 | T | C | 0.43 | -0.009 | 0.001 | 3.88E-10 | -0.008 | 0.004 | 0.053 | 41.33 |
| rs34067381 | T | G | 0.36 | -0.01 | 0.002 | 9.77E-13 | 0.006 | 0.004 | 0.198 | 48.07 |
| rs34098770 | A | G | 0.15 | -0.016 | 0.002 | 2.95E-16 | -0.006 | 0.004 | 0.184 | 64 |
| rs34305371 | A | G | 0.1 | 0.031 | 0.002 | 1.10E-39 | -0.008 | 0.004 | 0.075 | 171.17 |
| rs34394051 | A | G | 0.84 | -0.012 | 0.002 | 2.30E-09 | 0.024 | 0.004 | 3.50E-08 | 34.22 |
| rs34720381 | T | C | 0.09 | -0.018 | 0.002 | 1.90E-13 | -0.004 | 0.004 | 0.315 | 56.25 |
| rs34807077 | A | C | 0.16 | 0.013 | 0.002 | 3.29E-11 | -0.007 | 0.004 | 0.09 | 45.39 |
| rs35417702 | T | C | 0.52 | -0.015 | 0.001 | 2.40E-27 | 0.000 | 0.004 | 0.982 | 117.88 |
| rs35606437 | A | G | 0.27 | 0.011 | 0.002 | 3.98E-11 | 0.007 | 0.004 | 0.096 | 43.07 |
| rs35929923 | A | G | 0.25 | -0.011 | 0.002 | 1.77E-12 | -0.001 | 0.004 | 0.737 | 50.77 |
| rs363096 | T | C | 0.42 | -0.014 | 0.001 | 5.77E-24 | 0.004 | 0.004 | 0.347 | 104.33 |
| rs3751331 | A | G | 0.38 | -0.01 | 0.001 | 1.75E-11 | -0.003 | 0.004 | 0.569 | 48.01 |
| rs3809634 | A | G | 0.68 | -0.011 | 0.002 | 6.82E-13 | 0.005 | 0.004 | 0.229 | 52.8 |
| rs382196 | T | G | 0.38 | -0.008 | 0.001 | 1.47E-08 | 0.003 | 0.004 | 0.461 | 34.31 |
| rs3859523 | T | C | 0.82 | 0.012 | 0.002 | 9.92E-12 | 0.002 | 0.004 | 0.59 | 47.46 |
| rs3897821 | A | G | 0.67 | 0.015 | 0.002 | 3.58E-25 | 0.001 | 0.004 | 0.845 | 105.4 |
| rs3948495 | T | G | 0.41 | -0.009 | 0.001 | 3.95E-10 | 0.000 | 0.004 | 0.959 | 40.41 |
| rs401526 | T | C | 0.53 | -0.01 | 0.001 | 1.15E-13 | 0.008 | 0.004 | 0.072 | 55.18 |
| rs401966 | C | G | 0.39 | -0.01 | 0.001 | 1.72E-11 | -0.001 | 0.004 | 0.794 | 48.01 |
| rs42210 | C | G | 0.71 | -0.01 | 0.002 | 2.74E-10 | -0.006 | 0.004 | 0.167 | 37.52 |
| rs42302 | A | G | 0.34 | 0.009 | 0.002 | 5.96E-09 | 0.002 | 0.004 | 0.669 | 32.87 |
| rs4298514 | T | C | 0.7 | 0.01 | 0.002 | 1.07E-11 | -0.001 | 0.004 | 0.884 | 48.07 |
| rs4320563 | A | G | 0.51 | 0.011 | 0.001 | 2.24E-14 | 0.007 | 0.004 | 0.105 | 60.62 |
| rs4328757 | T | C | 0.61 | 0.01 | 0.001 | 2.95E-11 | -0.003 | 0.004 | 0.457 | 47.02 |
| rs4352658 | T | C | 0.08 | -0.019 | 0.003 | 1.71E-13 | 0.000 | 0.004 | 0.985 | 53.4 |
| rs4358081 | A | C | 0.53 | -0.01 | 0.001 | 1.28E-11 | 0.006 | 0.004 | 0.201 | 46.05 |
| rs4369924 | A | G | 0.16 | 0.013 | 0.002 | 5.58E-11 | 0.003 | 0.004 | 0.453 | 43.28 |
| rs4384309 | A | G | 0.46 | 0.01 | 0.001 | 4.89E-13 | 0.004 | 0.004 | 0.41 | 53.08 |
| rs4467547 | T | G | 0.41 | 0.012 | 0.001 | 4.88E-17 | -0.003 | 0.004 | 0.516 | 74.7 |
| rs4500930 | T | C | 0.34 | -0.011 | 0.002 | 1.67E-13 | -0.006 | 0.004 | 0.165 | 52.8 |
| rs4663617 | A | T | 0.23 | 0.01 | 0.002 | 4.12E-10 | -0.002 | 0.004 | 0.698 | 37.43 |
| rs4673840 | T | C | 0.84 | -0.014 | 0.002 | 5.86E-13 | -0.001 | 0.004 | 0.796 | 52.75 |
| rs4675248 | A | G | 0.4 | -0.009 | 0.001 | 4.50E-11 | 0.005 | 0.004 | 0.271 | 45.08 |
| rs4719944 | T | C | 0.55 | -0.012 | 0.001 | 9.25E-18 | 0.002 | 0.004 | 0.68 | 74.7 |
| rs4726070 | A | G | 0.6 | 0.012 | 0.001 | 1.36E-17 | 0.001 | 0.004 | 0.793 | 75.94 |
| rs4757957 | C | G | 0.69 | 0.013 | 0.002 | 1.79E-18 | 0.003 | 0.004 | 0.434 | 78.62 |
| rs4766424 | C | G | 0.12 | 0.014 | 0.002 | 2.47E-11 | -0.002 | 0.004 | 0.658 | 45.72 |
| rs4778058 | T | C | 0.5 | -0.009 | 0.001 | 4.92E-11 | 0.007 | 0.004 | 0.146 | 43.18 |
| rs4787457 | A | G | 0.63 | 0.016 | 0.002 | 1.23E-28 | -0.009 | 0.004 | 0.032 | 116.64 |
| rs4793090 | A | G | 0.66 | 0.008 | 0.002 | 1.67E-08 | -0.011 | 0.004 | 0.014 | 31.36 |
| rs4810894 | A | G | 0.37 | -0.009 | 0.002 | 4.38E-09 | -0.001 | 0.004 | 0.777 | 32.11 |
| rs4839155 | T | G | 0.77 | 0.012 | 0.002 | 4.66E-12 | 0.006 | 0.004 | 0.174 | 45.76 |
| rs4846724 | A | G | 0.53 | 0.01 | 0.001 | 2.89E-12 | -0.005 | 0.004 | 0.239 | 49 |
| rs4848924 | A | C | 0.71 | -0.012 | 0.002 | 3.56E-14 | 0.000 | 0.004 | 0.911 | 60.84 |
| rs488476 | C | G | 0.62 | -0.011 | 0.002 | 8.29E-15 | 0.000 | 0.004 | 0.922 | 56.75 |
| rs4895650 | T | C | 0.55 | 0.009 | 0.001 | 5.31E-11 | -0.003 | 0.004 | 0.529 | 44.13 |
| rs4899012 | C | G | 0.6 | -0.01 | 0.001 | 8.06E-13 | -0.002 | 0.004 | 0.658 | 54.13 |
| rs4904523 | A | G | 0.52 | -0.008 | 0.001 | 6.27E-09 | -0.003 | 0.004 | 0.532 | 34.31 |
| rs4915735 | A | G | 0.14 | 0.012 | 0.002 | 9.56E-09 | 0.012 | 0.004 | 0.007 | 33.64 |
| rs4977885 | A | G | 0.4 | -0.008 | 0.001 | 6.78E-09 | 0.000 | 0.004 | 0.964 | 36 |
| rs4984541 | A | G | 0.77 | -0.014 | 0.002 | 3.40E-16 | 0.005 | 0.004 | 0.277 | 65.9 |
| rs55736314 | C | G | 0.6 | -0.015 | 0.001 | 4.59E-27 | -0.003 | 0.004 | 0.475 | 121 |
| rs56099375 | T | C | 0.24 | 0.012 | 0.002 | 3.10E-13 | -0.002 | 0.004 | 0.617 | 49.83 |
| rs56319902 | T | C | 0.22 | -0.021 | 0.002 | 5.81E-33 | 0.029 | 0.004 | 4.67E-11 | 146.84 |
| rs56391344 | A | G | 0.25 | 0.015 | 0.002 | 7.49E-20 | 0.007 | 0.004 | 0.124 | 85.56 |
| rs56794817 | A | G | 0.16 | 0.012 | 0.002 | 1.69E-10 | -0.007 | 0.004 | 0.112 | 41.23 |
| rs57349798 | A | G | 0.41 | 0.009 | 0.001 | 8.63E-11 | 0.002 | 0.004 | 0.682 | 44.13 |
| rs57352738 | A | T | 0.2 | -0.015 | 0.002 | 2.51E-18 | 0.003 | 0.004 | 0.432 | 79.94 |
| rs575113 | A | G | 0.29 | 0.013 | 0.002 | 7.52E-18 | -0.003 | 0.004 | 0.522 | 78.62 |
| rs5754753 | T | C | 0.72 | -0.013 | 0.002 | 6.15E-16 | 0.005 | 0.004 | 0.264 | 62.02 |
| rs57661533 | T | C | 0.13 | -0.011 | 0.002 | 3.24E-08 | -0.003 | 0.004 | 0.557 | 29.47 |
| rs580652 | T | C | 0.9 | -0.015 | 0.002 | 4.65E-10 | -0.002 | 0.004 | 0.675 | 37.52 |
| rs590013 | T | C | 0.68 | 0.01 | 0.002 | 2.47E-11 | -0.004 | 0.004 | 0.404 | 44.44 |
| rs6043521 | T | C | 0.6 | -0.008 | 0.001 | 9.72E-09 | -0.001 | 0.004 | 0.806 | 35.15 |
| rs6065080 | T | C | 0.36 | -0.013 | 0.002 | 7.49E-19 | 0.001 | 0.004 | 0.832 | 75.11 |
| rs6065784 | C | G | 0.7 | 0.011 | 0.002 | 5.78E-13 | 0.000 | 0.004 | 0.962 | 53.78 |
| rs61104616 | A | G | 0.52 | -0.017 | 0.001 | 1.56E-34 | -0.004 | 0.004 | 0.407 | 150.94 |
| rs6123924 | A | G | 0.84 | 0.013 | 0.002 | 3.20E-11 | 0.000 | 0.004 | 0.973 | 46.1 |
| rs61527214 | A | G | 0.41 | 0.011 | 0.001 | 1.13E-14 | -0.002 | 0.004 | 0.697 | 61.73 |
| rs61997667 | T | C | 0.15 | -0.014 | 0.002 | 2.24E-13 | -0.004 | 0.004 | 0.335 | 51.84 |
| rs62155350 | A | G | 0.02 | 0.034 | 0.005 | 1.69E-10 | 0.005 | 0.004 | 0.248 | 40.67 |
| rs62155873 | T | C | 0.12 | -0.015 | 0.002 | 1.65E-12 | 0.000 | 0.004 | 0.91 | 51.7 |
| rs62174974 | A | G | 0.19 | -0.011 | 0.002 | 2.76E-09 | 0.002 | 0.004 | 0.587 | 34.03 |
| rs62179650 | A | G | 0.3 | 0.011 | 0.002 | 5.66E-13 | 0.004 | 0.004 | 0.4 | 49 |
| rs62182994 | T | C | 0.69 | 0.014 | 0.002 | 2.55E-19 | 0.006 | 0.004 | 0.154 | 83.42 |
| rs62379838 | T | C | 0.7 | 0.012 | 0.002 | 1.77E-15 | 0.004 | 0.004 | 0.316 | 65.07 |
| rs628993 | A | G | 0.11 | 0.013 | 0.002 | 3.83E-08 | 0.004 | 0.004 | 0.354 | 30.97 |
| rs6449503 | A | G | 0.5 | 0.018 | 0.001 | 1.84E-39 | 0.001 | 0.004 | 0.834 | 174.62 |
| rs6490618 | T | C | 0.33 | 0.009 | 0.002 | 3.48E-09 | 0.002 | 0.004 | 0.6 | 34.42 |
| rs6493265 | T | C | 0.39 | -0.012 | 0.001 | 2.26E-17 | 0.007 | 0.004 | 0.088 | 75.94 |
| rs6557171 | T | C | 0.32 | -0.016 | 0.002 | 5.48E-25 | 0.006 | 0.004 | 0.199 | 106.78 |
| rs660001 | A | G | 0.21 | -0.016 | 0.002 | 1.66E-21 | 0.001 | 0.004 | 0.737 | 93.07 |
| rs66568921 | T | G | 0.64 | -0.018 | 0.002 | 7.60E-34 | -0.009 | 0.004 | 0.048 | 145.6 |
| rs66721975 | A | G | 0.29 | -0.009 | 0.002 | 1.51E-08 | -0.004 | 0.004 | 0.362 | 33.64 |
| rs6690195 | T | C | 0.49 | -0.011 | 0.001 | 7.89E-16 | 0.007 | 0.004 | 0.103 | 65.15 |
| rs6697584 | T | C | 0.79 | -0.013 | 0.002 | 1.43E-13 | 0.000 | 0.004 | 0.916 | 54.93 |
| rs6704768 | A | G | 0.56 | -0.012 | 0.001 | 1.84E-16 | -0.010 | 0.004 | 0.016 | 68.65 |
| rs6731373 | A | G | 0.34 | -0.012 | 0.002 | 1.39E-14 | 0.003 | 0.004 | 0.438 | 58.78 |
| rs6812533 | T | C | 0.74 | 0.011 | 0.002 | 4.85E-11 | 0.007 | 0.004 | 0.088 | 43.89 |
| rs68145588 | T | G | 0.14 | -0.013 | 0.002 | 7.53E-10 | -0.003 | 0.004 | 0.45 | 36.57 |
| rs6917154 | T | C | 0.12 | 0.013 | 0.002 | 2.55E-09 | -0.001 | 0.004 | 0.819 | 36.57 |
| rs6924023 | A | G | 0.75 | 0.011 | 0.002 | 5.17E-11 | -0.004 | 0.004 | 0.317 | 44.72 |
| rs6977237 | T | C | 0.47 | 0.008 | 0.001 | 5.59E-09 | 0.004 | 0.004 | 0.316 | 34.31 |
| rs6994287 | A | G | 0.41 | 0.01 | 0.001 | 1.11E-12 | 0.009 | 0.004 | 0.039 | 53.08 |
| rs7012546 | T | C | 0.42 | 0.009 | 0.001 | 1.25E-10 | 0.003 | 0.004 | 0.447 | 43.18 |
| rs7029718 | A | G | 0.41 | 0.024 | 0.001 | 7.82E-65 | 0.003 | 0.004 | 0.547 | 303.76 |
| rs7041702 | A | G | 0.74 | -0.011 | 0.002 | 4.53E-12 | 0.001 | 0.004 | 0.848 | 48.13 |
| rs7167688 | T | C | 0.49 | -0.009 | 0.001 | 1.71E-11 | 0.001 | 0.004 | 0.905 | 45.08 |
| rs7171405 | A | G | 0.24 | -0.01 | 0.002 | 7.53E-10 | 0.000 | 0.004 | 0.987 | 39.85 |
| rs717996 | T | C | 0.6 | -0.012 | 0.001 | 1.89E-17 | 0.002 | 0.004 | 0.616 | 75.94 |
| rs7226824 | T | C | 0.53 | -0.008 | 0.001 | 2.72E-08 | 0.001 | 0.004 | 0.897 | 31.04 |
| rs72486027 | T | C | 0.25 | -0.01 | 0.002 | 1.61E-10 | 0.003 | 0.004 | 0.513 | 42.25 |
| rs7254263 | T | C | 0.28 | -0.012 | 0.002 | 1.84E-14 | 0.001 | 0.004 | 0.767 | 55.32 |
| rs72622559 | T | C | 0.23 | -0.012 | 0.002 | 5.50E-13 | -0.004 | 0.004 | 0.354 | 49.83 |
| rs72672052 | A | T | 0.18 | -0.01 | 0.002 | 4.87E-08 | 0.002 | 0.004 | 0.65 | 31.48 |
| rs72709560 | A | G | 0.69 | 0.009 | 0.002 | 2.03E-08 | -0.003 | 0.004 | 0.496 | 32.11 |
| rs72828517 | T | C | 0.83 | -0.017 | 0.002 | 1.38E-19 | 0.009 | 0.004 | 0.03 | 78.18 |
| rs72829857 | A | G | 0.76 | -0.015 | 0.002 | 1.52E-19 | -0.007 | 0.004 | 0.098 | 86.72 |
| rs72917504 | T | C | 0.06 | -0.018 | 0.003 | 2.91E-09 | 0.004 | 0.004 | 0.408 | 35.2 |
| rs72944064 | T | C | 0.74 | -0.01 | 0.002 | 9.39E-10 | -0.003 | 0.004 | 0.486 | 37.52 |
| rs72962169 | T | C | 0.16 | -0.017 | 0.002 | 1.26E-19 | 0.007 | 0.004 | 0.128 | 83.87 |
| rs72993796 | T | C | 0.88 | 0.013 | 0.002 | 1.12E-08 | -0.007 | 0.004 | 0.115 | 33.32 |
| rs730384 | A | G | 0.44 | 0.01 | 0.001 | 9.38E-13 | -0.008 | 0.004 | 0.072 | 52.05 |
| rs73055556 | A | G | 0.14 | 0.011 | 0.002 | 4.95E-08 | -0.002 | 0.004 | 0.652 | 30.8 |
| rs73191311 | A | G | 0.34 | -0.009 | 0.002 | 2.03E-10 | -0.002 | 0.004 | 0.658 | 39.27 |
| rs7321274 | A | G | 0.8 | 0.011 | 0.002 | 8.21E-11 | 0.002 | 0.004 | 0.588 | 40.11 |
| rs7326331 | A | G | 0.28 | -0.013 | 0.002 | 3.42E-16 | -0.001 | 0.004 | 0.746 | 64 |
| rs73344830 | A | G | 0.42 | 0.017 | 0.001 | 1.00E-32 | -0.006 | 0.004 | 0.159 | 147.45 |
| rs736281 | T | C | 0.39 | 0.009 | 0.001 | 2.49E-09 | 0.000 | 0.004 | 0.975 | 37.73 |
| rs73648455 | T | C | 0.08 | -0.018 | 0.003 | 1.99E-12 | 0.004 | 0.004 | 0.304 | 49.54 |
| rs743316 | T | C | 0.79 | 0.011 | 0.002 | 8.21E-11 | 0.001 | 0.004 | 0.775 | 43.4 |
| rs74415461 | T | C | 0.08 | 0.018 | 0.003 | 6.25E-12 | -0.017 | 0.004 | 6.49E-05 | 45.82 |
| rs7449561 | A | G | 0.23 | 0.01 | 0.002 | 9.73E-10 | -0.003 | 0.004 | 0.489 | 36 |
| rs7460106 | T | C | 0.76 | -0.012 | 0.002 | 2.06E-13 | -0.002 | 0.004 | 0.659 | 52.35 |
| rs746839 | C | G | 0.63 | 0.014 | 0.002 | 1.19E-20 | 0.001 | 0.004 | 0.793 | 81 |
| rs74787922 | A | C | 0.93 | 0.018 | 0.003 | 8.97E-11 | -0.002 | 0.004 | 0.61 | 42.25 |
| rs75033012 | C | G | 0.04 | -0.026 | 0.004 | 1.38E-11 | 0.002 | 0.004 | 0.598 | 46.1 |
| rs75177132 | T | C | 0.05 | 0.03 | 0.004 | 1.01E-16 | 0.005 | 0.004 | 0.22 | 68.52 |
| rs7575637 | A | G | 0.45 | 0.011 | 0.001 | 3.43E-14 | 0.006 | 0.004 | 0.168 | 58.41 |
| rs7603132 | A | G | 0.19 | 0.014 | 0.002 | 1.20E-14 | 0.004 | 0.004 | 0.313 | 57.93 |
| rs76076331 | T | C | 0.13 | 0.021 | 0.002 | 1.71E-23 | -0.001 | 0.004 | 0.771 | 96.23 |
| rs7617204 | A | G | 0.52 | -0.01 | 0.001 | 4.33E-12 | 0.004 | 0.004 | 0.333 | 48.01 |
| rs76235882 | A | G | 0.97 | 0.024 | 0.004 | 4.52E-10 | 0.004 | 0.004 | 0.36 | 38.19 |
| rs76246107 | A | G | 0.09 | -0.016 | 0.003 | 6.52E-10 | -0.001 | 0.004 | 0.773 | 38.82 |
| rs7650602 | T | C | 0.56 | -0.009 | 0.001 | 2.77E-10 | -0.002 | 0.004 | 0.63 | 40.41 |
| rs76577427 | C | G | 0.9 | 0.02 | 0.002 | 7.78E-18 | 0.005 | 0.004 | 0.224 | 71.54 |
| rs7672622 | A | G | 0.75 | -0.009 | 0.002 | 4.72E-08 | 0.001 | 0.004 | 0.849 | 30.25 |
| rs7683416 | T | C | 0.46 | 0.014 | 0.001 | 6.05E-23 | -0.003 | 0.004 | 0.484 | 98.58 |
| rs76878669 | C | G | 0.76 | 0.014 | 0.002 | 3.10E-17 | -0.002 | 0.004 | 0.614 | 68.79 |
| rs7692359 | T | C | 0.78 | -0.011 | 0.002 | 1.40E-10 | -0.008 | 0.004 | 0.053 | 41.11 |
| rs77025239 | A | G | 0.16 | -0.014 | 0.002 | 4.65E-13 | 0.006 | 0.004 | 0.187 | 54.29 |
| rs77609760 | A | G | 0.93 | 0.024 | 0.003 | 7.71E-17 | 0.002 | 0.004 | 0.683 | 71.04 |
| rs77719387 | A | T | 0.02 | -0.04 | 0.006 | 3.78E-12 | 0.002 | 0.004 | 0.625 | 48.28 |
| rs7849487 | T | G | 0.66 | -0.017 | 0.002 | 1.25E-29 | 0.005 | 0.004 | 0.224 | 123.95 |
| rs78648104 | T | C | 0.91 | -0.014 | 0.002 | 1.70E-08 | 0.006 | 0.004 | 0.172 | 31.81 |
| rs7875078 | A | C | 0.46 | -0.008 | 0.001 | 2.72E-08 | -0.003 | 0.004 | 0.522 | 31.04 |
| rs7894722 | T | C | 0.37 | 0.009 | 0.001 | 5.41E-09 | -0.002 | 0.004 | 0.58 | 36.86 |
| rs790647 | A | C | 0.23 | -0.015 | 0.002 | 3.81E-18 | -0.004 | 0.004 | 0.414 | 72.75 |
| rs7910403 | T | G | 0.2 | 0.012 | 0.002 | 5.08E-12 | 0.003 | 0.004 | 0.488 | 45.94 |
| rs7924036 | T | G | 0.51 | 0.013 | 0.001 | 2.55E-21 | -0.006 | 0.004 | 0.159 | 90.25 |
| rs79265434 | A | G | 0.88 | -0.02 | 0.002 | 1.98E-19 | 0.004 | 0.004 | 0.365 | 80.18 |
| rs7972246 | T | C | 0.35 | 0.011 | 0.002 | 1.01E-13 | -0.001 | 0.004 | 0.846 | 53.78 |
| rs79728014 | A | G | 0.86 | -0.013 | 0.002 | 2.06E-11 | -0.006 | 0.004 | 0.202 | 44.89 |
| rs7974852 | A | C | 0.52 | 0.012 | 0.001 | 1.14E-16 | 0.012 | 0.004 | 0.008 | 69.84 |
| rs7977614 | A | G | 0.71 | -0.013 | 0.002 | 6.84E-17 | 0.006 | 0.004 | 0.173 | 70.14 |
| rs79994730 | T | C | 0.04 | 0.024 | 0.004 | 5.88E-12 | 0.003 | 0.004 | 0.508 | 47.02 |
| rs8008382 | T | C | 0.3 | -0.011 | 0.002 | 4.07E-12 | 0.007 | 0.004 | 0.102 | 49.94 |
| rs8024 | A | C | 0.33 | -0.012 | 0.002 | 1.13E-14 | 0.003 | 0.004 | 0.48 | 58.78 |
| rs80257979 | T | G | 0.97 | -0.025 | 0.004 | 3.31E-10 | -0.007 | 0.004 | 0.099 | 39.69 |
| rs8030487 | A | G | 0.69 | 0.009 | 0.002 | 2.52E-08 | 0.002 | 0.004 | 0.639 | 32.11 |
| rs806816 | A | T | 0.75 | 0.01 | 0.002 | 4.80E-10 | -0.005 | 0.004 | 0.222 | 39.85 |
| rs8097125 | T | C | 0.38 | 0.009 | 0.001 | 2.31E-09 | 0.002 | 0.004 | 0.616 | 37.73 |
| rs912883 | T | C | 0.68 | 0.009 | 0.002 | 6.20E-09 | -0.004 | 0.004 | 0.309 | 33.64 |
| rs9289300 | T | C | 0.84 | -0.016 | 0.002 | 1.00E-17 | 0.001 | 0.004 | 0.886 | 74.5 |
| rs929511 | T | C | 0.13 | -0.016 | 0.002 | 3.91E-14 | -0.010 | 0.004 | 0.031 | 58.05 |
| rs9359939 | A | C | 0.24 | -0.011 | 0.002 | 1.51E-10 | -0.010 | 0.004 | 0.02 | 43.07 |
| rs936496 | A | G | 0.62 | -0.008 | 0.001 | 4.38E-08 | -0.003 | 0.004 | 0.5 | 31.84 |
| rs9373363 | A | G | 0.75 | -0.011 | 0.002 | 5.54E-12 | 0.004 | 0.004 | 0.33 | 48.13 |
| rs9375188 | T | C | 0.48 | 0.021 | 0.001 | 8.78E-52 | 0.006 | 0.004 | 0.185 | 229.31 |
| rs9411331 | A | C | 0.68 | 0.015 | 0.002 | 7.63E-22 | 0.000 | 0.004 | 0.962 | 93.44 |
| rs9492774 | C | G | 0.37 | 0.008 | 0.002 | 4.33E-08 | -0.002 | 0.004 | 0.586 | 28.44 |
| rs9529146 | T | C | 0.23 | 0.013 | 0.002 | 3.48E-14 | 0.005 | 0.004 | 0.253 | 55.81 |
| rs9556958 | T | C | 0.52 | -0.011 | 0.001 | 5.96E-15 | 0.003 | 0.004 | 0.538 | 61.73 |
| rs9616947 | T | C | 0.38 | -0.013 | 0.002 | 3.58E-20 | 0.000 | 0.004 | 0.93 | 79.8 |
| rs9633970 | T | C | 0.76 | 0.009 | 0.002 | 2.81E-08 | 0.000 | 0.004 | 0.969 | 32.35 |
| rs9655780 | A | G | 0.83 | -0.015 | 0.002 | 3.70E-16 | -0.002 | 0.004 | 0.724 | 64.84 |
| rs969512 | A | T | 0.66 | -0.011 | 0.002 | 9.84E-13 | 0.000 | 0.004 | 0.96 | 49.94 |
| rs9853928 | T | C | 0.21 | -0.013 | 0.002 | 1.59E-13 | -0.002 | 0.004 | 0.611 | 55.81 |
| rs9859556 | T | G | 0.31 | 0.029 | 0.002 | 4.61E-82 | -0.005 | 0.004 | 0.231 | 373.78 |
| rs9882532 | T | C | 0.64 | 0.012 | 0.002 | 2.15E-17 | -0.001 | 0.004 | 0.79 | 68.34 |
| rs9886703 | A | T | 0.17 | -0.014 | 0.002 | 4.46E-13 | -0.005 | 0.004 | 0.287 | 51.24 |
| rs9927137 | A | G | 0.53 | 0.008 | 0.001 | 5.11E-09 | 0.000 | 0.004 | 0.926 | 34.31 |
| rs9927842 | T | C | 0.15 | -0.013 | 0.002 | 6.22E-11 | 0.001 | 0.004 | 0.753 | 42.25 |
| rs9929556 | T | G | 0.57 | -0.01 | 0.001 | 2.31E-12 | 0.000 | 0.004 | 0.999 | 50.01 |
| rs9929762 | A | G | 0.56 | 0.01 | 0.001 | 1.03E-11 | -0.002 | 0.004 | 0.616 | 47.02 |
| rs9933256 | A | G | 0.56 | 0.012 | 0.001 | 6.06E-17 | 0.003 | 0.004 | 0.491 | 72.25 |

**SNPs used as instrumental variables in the MR analysis of educational attainment on intracellular volume fraction**

| **SNP** | **Effect allele** | **Other allele** | **Effect allele frequency** | **beta (exposure)** | **SE (exposure)** | ***P* (exposure)** | **beta (outcome)** | **SE (outcome)** | ***P* (outcome)** | ***F*-statistic** |
| --- | --- | --- | --- | --- | --- | --- | --- | --- | --- | --- |
| rs10006235 | T | C | 0.27 | -0.009 | 0.002 | 4.48E-09 | 0.003 | 0.007 | 0.643 | 33.79 |
| rs10021733 | T | C | 0.15 | -0.016 | 0.003 | 9.45E-10 | -0.006 | 0.008 | 0.431 | 36.89 |
| rs10046069 | A | G | 0.12 | 0.013 | 0.002 | 4.40E-09 | -0.002 | 0.009 | 0.848 | 33.32 |
| rs10060023 | T | C | 0.33 | 0.011 | 0.002 | 1.88E-13 | -0.002 | 0.006 | 0.706 | 53.78 |
| rs10080647 | A | C | 0.14 | 0.012 | 0.002 | 7.05E-10 | 0.004 | 0.009 | 0.655 | 38.44 |
| rs1008078 | T | C | 0.4 | -0.018 | 0.001 | 4.50E-37 | -0.008 | 0.006 | 0.2 | 169 |
| rs10189857 | A | G | 0.57 | 0.016 | 0.001 | 4.69E-29 | 0.003 | 0.006 | 0.658 | 127.37 |
| rs10193498 | A | T | 0.76 | 0.01 | 0.002 | 3.10E-09 | -0.004 | 0.007 | 0.606 | 36.75 |
| rs10215082 | A | G | 0.47 | -0.015 | 0.001 | 7.03E-26 | -0.011 | 0.006 | 0.068 | 113.27 |
| rs10460095 | A | G | 0.57 | -0.012 | 0.001 | 2.61E-18 | 0.001 | 0.006 | 0.859 | 78.45 |
| rs10761251 | A | T | 0.66 | 0.011 | 0.002 | 2.52E-14 | -0.007 | 0.006 | 0.313 | 56.75 |
| rs10772644 | C | G | 0.89 | 0.014 | 0.002 | 1.69E-10 | 0.006 | 0.01 | 0.57 | 41.66 |
| rs10773002 | A | T | 0.25 | 0.018 | 0.002 | 2.50E-30 | 0.013 | 0.007 | 0.056 | 133.69 |
| rs10773208 | T | C | 0.25 | -0.011 | 0.002 | 8.09E-12 | -0.009 | 0.007 | 0.176 | 47.27 |
| rs10798888 | T | G | 0.17 | -0.014 | 0.002 | 5.15E-14 | -0.016 | 0.008 | 0.051 | 54.29 |
| rs10799615 | A | G | 0.25 | -0.009 | 0.002 | 4.11E-08 | -0.003 | 0.007 | 0.671 | 30.94 |
| rs10805383 | A | G | 0.49 | -0.01 | 0.001 | 2.22E-13 | -0.004 | 0.006 | 0.532 | 54.13 |
| rs10830858 | T | C | 0.51 | 0.009 | 0.001 | 5.31E-11 | -0.004 | 0.006 | 0.472 | 43.18 |
| rs10844179 | A | G | 0.23 | 0.01 | 0.002 | 5.03E-10 | 0.006 | 0.007 | 0.442 | 37.43 |
| rs10875121 | C | G | 0.84 | 0.017 | 0.002 | 4.46E-20 | -0.014 | 0.008 | 0.097 | 83.87 |
| rs10892807 | T | C | 0.57 | 0.011 | 0.001 | 1.09E-14 | -0.01 | 0.006 | 0.089 | 60.62 |
| rs10940921 | T | G | 0.42 | 0.011 | 0.001 | 1.31E-14 | -0.003 | 0.006 | 0.669 | 62.86 |
| rs10951590 | T | C | 0.33 | -0.011 | 0.002 | 3.62E-13 | 0.012 | 0.006 | 0.069 | 52.8 |
| rs10979613 | T | C | 0.64 | -0.012 | 0.002 | 1.25E-15 | -0.008 | 0.006 | 0.205 | 60.84 |
| rs10996167 | C | G | 0.65 | 0.011 | 0.002 | 7.47E-09 | -0.008 | 0.006 | 0.194 | 31.92 |
| rs11003463 | T | G | 0.61 | 0.009 | 0.002 | 5.58E-10 | 0.004 | 0.006 | 0.558 | 36.8 |
| rs11023764 | A | G | 0.64 | 0.009 | 0.002 | 5.32E-10 | 0.008 | 0.006 | 0.238 | 36.8 |
| rs11030102 | C | G | 0.75 | 0.01 | 0.002 | 4.60E-10 | 0.004 | 0.007 | 0.601 | 39.85 |
| rs11076962 | T | C | 0.72 | 0.009 | 0.002 | 4.29E-08 | -0.006 | 0.007 | 0.373 | 28.89 |
| rs11081529 | T | C | 0.71 | 0.013 | 0.002 | 5.22E-16 | 0.005 | 0.007 | 0.459 | 70.56 |
| rs11082011 | T | C | 0.67 | 0.019 | 0.002 | 2.06E-38 | -0.004 | 0.006 | 0.582 | 167.27 |
| rs11121177 | A | G | 0.18 | 0.015 | 0.002 | 1.44E-16 | 0.003 | 0.008 | 0.747 | 70.37 |
| rs11130380 | T | G | 0.61 | 0.009 | 0.001 | 4.17E-10 | 0.002 | 0.006 | 0.753 | 41.33 |
| rs11138947 | T | C | 0.72 | 0.011 | 0.002 | 2.36E-11 | 0.002 | 0.007 | 0.823 | 43.07 |
| rs11157931 | A | C | 0.39 | -0.014 | 0.001 | 3.86E-22 | -0.002 | 0.006 | 0.745 | 98.58 |
| rs111852224 | T | C | 0.12 | 0.016 | 0.002 | 4.85E-13 | -0.004 | 0.009 | 0.691 | 50.28 |
| rs11211123 | A | G | 0.22 | 0.01 | 0.002 | 7.62E-10 | 0.003 | 0.007 | 0.674 | 37.43 |
| rs11213482 | A | G | 0.84 | -0.011 | 0.002 | 2.68E-08 | 0.006 | 0.008 | 0.488 | 31.12 |
| rs112603734 | A | C | 0.77 | -0.013 | 0.002 | 2.72E-09 | 0.006 | 0.007 | 0.375 | 35.46 |
| rs112806496 | C | G | 0.91 | -0.016 | 0.002 | 1.72E-10 | -0.01 | 0.011 | 0.338 | 40.96 |
| rs1128956 | T | G | 0.83 | -0.014 | 0.002 | 5.34E-14 | 0.002 | 0.008 | 0.785 | 54.29 |
| rs113520408 | A | G | 0.28 | 0.016 | 0.002 | 1.03E-22 | 0.002 | 0.007 | 0.749 | 93.85 |
| rs113615161 | T | C | 0.13 | -0.012 | 0.002 | 2.98E-09 | 0.014 | 0.009 | 0.108 | 34.31 |
| rs114593137 | A | T | 0.79 | -0.012 | 0.002 | 1.65E-12 | 0.018 | 0.007 | 0.014 | 51.5 |
| rs115000530 | A | T | 0.95 | -0.025 | 0.003 | 1.09E-15 | -0.001 | 0.013 | 0.937 | 65.04 |
| rs11542663 | A | C | 0.69 | 0.011 | 0.002 | 2.44E-13 | -0.004 | 0.007 | 0.526 | 55.75 |
| rs11588857 | A | G | 0.21 | 0.02 | 0.002 | 4.45E-31 | 0.007 | 0.007 | 0.326 | 137.03 |
| rs11598765 | A | G | 0.2 | -0.011 | 0.002 | 1.73E-09 | 0 | 0.008 | 0.99 | 34.68 |
| rs11620355 | A | G | 0.09 | 0.018 | 0.002 | 1.09E-13 | -0.017 | 0.011 | 0.108 | 53 |
| rs11657342 | A | G | 0.37 | 0.014 | 0.002 | 2.32E-13 | 0.001 | 0.006 | 0.865 | 54.29 |
| rs11657979 | A | G | 0.24 | -0.009 | 0.002 | 4.57E-08 | -0.006 | 0.007 | 0.412 | 28.65 |
| rs11663602 | A | C | 0.28 | -0.013 | 0.002 | 9.27E-17 | 0.008 | 0.007 | 0.26 | 66.02 |
| rs1167827 | A | G | 0.43 | 0.01 | 0.001 | 2.93E-13 | -0.017 | 0.006 | 0.006 | 55.18 |
| rs11724690 | T | G | 0.29 | 0.009 | 0.002 | 2.08E-09 | -0.003 | 0.007 | 0.631 | 38.44 |
| rs117398064 | C | G | 0.09 | -0.014 | 0.002 | 1.16E-08 | -0.002 | 0.01 | 0.834 | 31.81 |
| rs118134876 | T | C | 0.06 | -0.022 | 0.003 | 1.50E-13 | -0.009 | 0.013 | 0.455 | 55.25 |
| rs11871429 | A | G | 0.77 | 0.013 | 0.002 | 2.52E-14 | 0.01 | 0.007 | 0.186 | 56.69 |
| rs11894424 | A | C | 0.95 | 0.021 | 0.003 | 2.14E-10 | 0.006 | 0.015 | 0.674 | 40.11 |
| rs12054166 | C | G | 0.74 | 0.01 | 0.002 | 2.30E-09 | -0.019 | 0.007 | 0.006 | 36 |
| rs12076635 | C | G | 0.78 | 0.021 | 0.002 | 3.33E-34 | 0.007 | 0.007 | 0.314 | 148.27 |
| rs12113634 | T | C | 0.65 | 0.009 | 0.002 | 2.50E-09 | -0.003 | 0.006 | 0.601 | 34.42 |
| rs12151248 | T | C | 0.11 | -0.015 | 0.002 | 3.81E-11 | 0.001 | 0.009 | 0.926 | 41.97 |
| rs1220779 | A | G | 0.47 | -0.008 | 0.001 | 1.58E-08 | -0.003 | 0.006 | 0.564 | 31.84 |
| rs12273435 | A | G | 0.21 | -0.013 | 0.002 | 4.72E-14 | -0.002 | 0.008 | 0.825 | 60.29 |
| rs12332731 | A | T | 0.19 | 0.014 | 0.002 | 5.96E-14 | -0.006 | 0.008 | 0.409 | 56.25 |
| rs12375949 | T | C | 0.43 | -0.014 | 0.001 | 1.61E-22 | 0.016 | 0.006 | 0.009 | 97.16 |
| rs12438177 | A | G | 0.37 | 0.009 | 0.002 | 1.48E-10 | -0.007 | 0.006 | 0.274 | 38.44 |
| rs12468040 | T | G | 0.38 | 0.014 | 0.001 | 3.12E-21 | 0.001 | 0.006 | 0.863 | 95.76 |
| rs12503522 | T | C | 0.28 | -0.011 | 0.002 | 3.93E-13 | 0.006 | 0.007 | 0.345 | 49.88 |
| rs12506221 | T | G | 0.44 | -0.013 | 0.001 | 4.23E-20 | 0.01 | 0.006 | 0.098 | 86.22 |
| rs12519073 | T | C | 0.23 | -0.011 | 0.002 | 1.16E-10 | 0 | 0.007 | 0.947 | 39.62 |
| rs12524795 | T | C | 0.44 | 0.01 | 0.001 | 5.28E-13 | -0.001 | 0.006 | 0.823 | 53.08 |
| rs12571549 | A | G | 0.14 | 0.015 | 0.002 | 1.45E-13 | -0.002 | 0.009 | 0.838 | 57 |
| rs12574281 | A | C | 0.63 | -0.008 | 0.002 | 3.04E-08 | 0.002 | 0.006 | 0.787 | 28.44 |
| rs12591647 | T | C | 0.82 | -0.015 | 0.002 | 7.78E-17 | -0.004 | 0.008 | 0.58 | 71.31 |
| rs12602286 | T | G | 0.87 | 0.016 | 0.002 | 4.62E-14 | 0.001 | 0.009 | 0.948 | 58.05 |
| rs12614263 | A | G | 0.5 | 0.008 | 0.001 | 3.38E-09 | 0.001 | 0.006 | 0.826 | 35.15 |
| rs12638072 | A | G | 0.3 | -0.011 | 0.002 | 3.13E-12 | -0.012 | 0.007 | 0.072 | 50.88 |
| rs12643771 | T | C | 0.31 | 0.013 | 0.002 | 1.32E-17 | -0.01 | 0.007 | 0.144 | 75.11 |
| rs12646523 | T | C | 0.25 | -0.013 | 0.002 | 3.79E-16 | 0.007 | 0.007 | 0.284 | 68.06 |
| rs12670376 | A | G | 0.45 | 0.009 | 0.001 | 5.99E-11 | 0.004 | 0.006 | 0.479 | 43.18 |
| rs12761761 | T | C | 0.24 | 0.015 | 0.002 | 4.79E-20 | -0.017 | 0.007 | 0.018 | 81 |
| rs12765185 | A | T | 0.28 | -0.009 | 0.002 | 2.96E-08 | -0.004 | 0.007 | 0.581 | 30.25 |
| rs12789313 | T | C | 0.49 | 0.009 | 0.001 | 6.79E-11 | 0.005 | 0.006 | 0.364 | 43.18 |
| rs12875339 | A | C | 0.64 | -0.011 | 0.002 | 1.71E-13 | 0.005 | 0.006 | 0.428 | 51.84 |
| rs12888615 | T | C | 0.2 | 0.012 | 0.002 | 9.42E-12 | -0.001 | 0.008 | 0.898 | 44.44 |
| rs12957463 | A | G | 0.8 | 0.015 | 0.002 | 9.20E-17 | -0.005 | 0.008 | 0.542 | 65.79 |
| rs12967010 | T | C | 0.77 | 0.011 | 0.002 | 1.33E-11 | -0.004 | 0.007 | 0.542 | 44.97 |
| rs12981405 | T | C | 0.17 | -0.011 | 0.002 | 2.75E-09 | -0.001 | 0.008 | 0.914 | 34.75 |
| rs1301838 | T | C | 0.32 | -0.01 | 0.002 | 1.29E-10 | 0.008 | 0.006 | 0.209 | 40.96 |
| rs13018640 | T | C | 0.6 | -0.021 | 0.001 | 1.05E-50 | -0.006 | 0.006 | 0.304 | 235.84 |
| rs13050131 | A | C | 0.66 | -0.008 | 0.002 | 4.40E-08 | -0.003 | 0.006 | 0.609 | 29.16 |
| rs13133213 | A | G | 0.5 | 0.01 | 0.001 | 7.57E-12 | 0.002 | 0.006 | 0.708 | 47.02 |
| rs13145650 | T | C | 0.92 | -0.017 | 0.002 | 8.99E-12 | -0.001 | 0.011 | 0.893 | 47.89 |
| rs13163845 | T | C | 0.85 | -0.015 | 0.002 | 4.66E-14 | -0.005 | 0.009 | 0.535 | 55.5 |
| rs13197257 | T | G | 0.28 | 0.011 | 0.002 | 2.83E-12 | -0.023 | 0.007 | 8.17E-04 | 46.41 |
| rs13212041 | T | C | 0.8 | 0.01 | 0.002 | 7.42E-09 | 0 | 0.007 | 0.985 | 31.48 |
| rs13240401 | T | C | 0.78 | 0.018 | 0.002 | 2.36E-24 | -0.002 | 0.007 | 0.804 | 107.18 |
| rs13261773 | C | G | 0.16 | 0.012 | 0.002 | 3.12E-10 | 0 | 0.008 | 0.989 | 41.23 |
| rs13266287 | A | T | 0.67 | -0.009 | 0.002 | 1.75E-08 | -0.015 | 0.006 | 0.016 | 32.11 |
| rs1329125 | T | C | 0.33 | -0.01 | 0.002 | 8.37E-11 | 0.008 | 0.007 | 0.251 | 41.82 |
| rs13327482 | A | G | 0.82 | -0.012 | 0.002 | 4.47E-11 | -0.023 | 0.008 | 0.005 | 45.19 |
| rs1334297 | A | G | 0.74 | 0.026 | 0.002 | 2.53E-58 | -0.006 | 0.007 | 0.37 | 258 |
| rs13422673 | T | C | 0.47 | -0.011 | 0.001 | 6.48E-15 | -0.004 | 0.006 | 0.516 | 61.73 |
| rs137079 | T | C | 0.14 | 0.013 | 0.002 | 3.06E-10 | -0.005 | 0.009 | 0.548 | 40.96 |
| rs1426619 | T | C | 0.43 | 0.01 | 0.001 | 1.40E-11 | -0.001 | 0.006 | 0.927 | 47.02 |
| rs1434630 | T | G | 0.15 | -0.014 | 0.002 | 4.63E-12 | 0.01 | 0.008 | 0.227 | 46.92 |
| rs143743568 | A | G | 0.15 | 0.012 | 0.002 | 4.99E-09 | 0.001 | 0.008 | 0.946 | 34.81 |
| rs150537577 | A | G | 0.92 | -0.015 | 0.003 | 9.50E-09 | 0.009 | 0.012 | 0.453 | 33.73 |
| rs1505676 | C | G | 0.63 | 0.008 | 0.002 | 6.67E-09 | 0 | 0.006 | 0.938 | 31.36 |
| rs1542354 | A | G | 0.48 | -0.008 | 0.001 | 2.72E-09 | 0 | 0.006 | 0.947 | 36 |
| rs1544 | A | G | 0.26 | -0.01 | 0.002 | 1.76E-09 | -0.003 | 0.007 | 0.669 | 36 |
| rs1564347 | T | G | 0.34 | 0.009 | 0.002 | 2.39E-09 | -0.002 | 0.006 | 0.752 | 34.42 |
| rs1603460 | T | G | 0.42 | 0.009 | 0.001 | 2.20E-10 | -0.003 | 0.006 | 0.586 | 41.33 |
| rs162445 | A | G | 0.08 | 0.015 | 0.003 | 5.72E-09 | 0.011 | 0.011 | 0.353 | 34.18 |
| rs1689510 | C | G | 0.33 | 0.019 | 0.002 | 1.35E-38 | -0.028 | 0.006 | 1.23E-05 | 167.27 |
| rs17048855 | A | G | 0.34 | 0.011 | 0.002 | 3.10E-13 | -0.003 | 0.006 | 0.579 | 51.84 |
| rs17110109 | T | C | 0.62 | -0.009 | 0.001 | 1.47E-10 | 0.003 | 0.006 | 0.659 | 44.13 |
| rs17144467 | A | C | 0.69 | 0.01 | 0.002 | 8.74E-11 | 0.002 | 0.007 | 0.711 | 42.68 |
| rs17148998 | A | G | 0.2 | 0.01 | 0.002 | 6.98E-09 | -0.019 | 0.007 | 0.013 | 35.3 |
| rs1717204 | A | C | 0.18 | -0.012 | 0.002 | 1.20E-10 | 0.01 | 0.008 | 0.189 | 42.25 |
| rs17248751 | A | G | 0.78 | -0.014 | 0.002 | 1.28E-15 | -0.005 | 0.007 | 0.494 | 64 |
| rs173003 | A | C | 0.49 | -0.008 | 0.001 | 5.50E-09 | -0.013 | 0.006 | 0.025 | 34.31 |
| rs1738050 | C | G | 0.62 | -0.009 | 0.001 | 1.42E-10 | 0.011 | 0.006 | 0.08 | 44.13 |
| rs17411339 | A | G | 0.56 | 0.014 | 0.001 | 5.10E-23 | 0.002 | 0.006 | 0.782 | 100 |
| rs17428076 | C | G | 0.76 | 0.014 | 0.002 | 6.41E-17 | -0.007 | 0.007 | 0.318 | 73.32 |
| rs1747714 | T | C | 0.53 | 0.011 | 0.001 | 3.82E-14 | -0.008 | 0.006 | 0.162 | 57.33 |
| rs1747817 | T | C | 0.76 | -0.009 | 0.002 | 2.29E-08 | 0.005 | 0.007 | 0.515 | 33.06 |
| rs17489649 | A | G | 0.68 | 0.013 | 0.002 | 2.77E-17 | 0 | 0.006 | 0.996 | 71.68 |
| rs17502934 | T | G | 0.15 | -0.017 | 0.002 | 2.12E-17 | 0.011 | 0.009 | 0.196 | 69.72 |
| rs175325 | A | T | 0.6 | -0.01 | 0.001 | 2.24E-12 | 0.001 | 0.006 | 0.877 | 51.02 |
| rs17551064 | A | G | 0.84 | 0.014 | 0.002 | 1.53E-13 | 0.003 | 0.008 | 0.7 | 55.07 |
| rs17563464 | A | C | 0.22 | -0.015 | 0.002 | 1.74E-17 | -0.011 | 0.008 | 0.15 | 74.77 |
| rs17565975 | A | G | 0.56 | -0.011 | 0.001 | 5.54E-14 | 0.008 | 0.006 | 0.21 | 57.33 |
| rs17598675 | T | C | 0.51 | -0.012 | 0.001 | 1.37E-16 | -0.004 | 0.006 | 0.512 | 68.65 |
| rs17604349 | A | G | 0.19 | 0.013 | 0.002 | 2.36E-12 | -0.013 | 0.008 | 0.094 | 49.78 |
| rs176218 | T | G | 0.2 | 0.02 | 0.002 | 1.10E-29 | -0.013 | 0.008 | 0.1 | 123.46 |
| rs17732878 | T | C | 0.79 | -0.01 | 0.002 | 4.06E-08 | -0.003 | 0.007 | 0.711 | 31.23 |
| rs178183 | T | C | 0.25 | 0.015 | 0.002 | 4.81E-19 | -0.007 | 0.007 | 0.31 | 82.13 |
| rs17882802 | A | G | 0.43 | 0.008 | 0.001 | 3.00E-09 | 0.01 | 0.006 | 0.115 | 36 |
| rs182902112 | A | C | 0.02 | -0.03 | 0.005 | 1.17E-08 | -0.009 | 0.026 | 0.738 | 32.04 |
| rs1842713 | A | G | 0.79 | -0.012 | 0.002 | 9.67E-12 | 0.008 | 0.007 | 0.29 | 47.37 |
| rs1861786 | A | G | 0.38 | -0.008 | 0.001 | 2.39E-08 | 0.004 | 0.006 | 0.559 | 33.47 |
| rs1865955 | T | C | 0.82 | 0.012 | 0.002 | 4.06E-11 | -0.015 | 0.008 | 0.053 | 41.23 |
| rs1866823 | A | G | 0.54 | 0.011 | 0.001 | 3.09E-14 | 0.009 | 0.006 | 0.161 | 58.41 |
| rs1890132 | T | C | 0.71 | -0.009 | 0.002 | 6.56E-09 | 0 | 0.007 | 0.951 | 35.2 |
| rs192436652 | T | C | 0.03 | -0.032 | 0.004 | 2.06E-12 | 0.017 | 0.019 | 0.369 | 50.25 |
| rs1933264 | T | C | 0.75 | 0.01 | 0.002 | 5.85E-10 | 0.011 | 0.007 | 0.129 | 39.06 |
| rs1955250 | A | C | 0.91 | -0.014 | 0.002 | 2.26E-08 | 0.004 | 0.011 | 0.69 | 31.36 |
| rs1991585 | T | C | 0.71 | -0.01 | 0.002 | 6.26E-11 | -0.002 | 0.007 | 0.746 | 45.34 |
| rs2007655 | T | G | 0.5 | 0.009 | 0.001 | 8.64E-10 | 0.003 | 0.006 | 0.67 | 37.73 |
| rs2011603 | A | G | 0.74 | -0.009 | 0.002 | 1.12E-08 | -0.004 | 0.007 | 0.583 | 32.35 |
| rs2034631 | T | C | 0.65 | 0.009 | 0.002 | 2.64E-09 | 0 | 0.006 | 0.995 | 35.2 |
| rs2052285 | A | G | 0.6 | 0.011 | 0.001 | 7.25E-15 | -0.008 | 0.006 | 0.176 | 64 |
| rs2055940 | A | G | 0.32 | 0.008 | 0.002 | 4.95E-08 | 0.004 | 0.006 | 0.515 | 29.88 |
| rs2081652 | A | T | 0.66 | 0.012 | 0.002 | 1.09E-16 | -0.005 | 0.006 | 0.422 | 67.24 |
| rs2131167 | A | G | 0.45 | 0.008 | 0.001 | 1.52E-08 | -0.001 | 0.006 | 0.925 | 32.65 |
| rs2179152 | T | C | 0.37 | -0.013 | 0.002 | 1.68E-19 | 0.009 | 0.006 | 0.168 | 76.27 |
| rs2183271 | T | C | 0.64 | 0.008 | 0.002 | 1.91E-08 | -0.011 | 0.006 | 0.083 | 31.36 |
| rs2199409 | T | C | 0.8 | 0.01 | 0.002 | 3.14E-09 | 0.001 | 0.008 | 0.921 | 36.71 |
| rs2212430 | T | C | 0.3 | -0.01 | 0.002 | 1.32E-10 | -0.002 | 0.007 | 0.803 | 39.06 |
| rs2216144 | T | C | 0.53 | 0.01 | 0.001 | 2.58E-12 | 0.007 | 0.006 | 0.266 | 49 |
| rs2256965 | A | G | 0.42 | 0.011 | 0.001 | 2.40E-13 | -0.006 | 0.006 | 0.342 | 57.33 |
| rs2287838 | A | G | 0.55 | -0.01 | 0.001 | 1.27E-13 | -0.015 | 0.006 | 0.017 | 55.18 |
| rs2290601 | T | C | 0.77 | -0.015 | 0.002 | 4.49E-19 | -0.007 | 0.007 | 0.319 | 76.82 |
| rs2297293 | C | G | 0.31 | 0.01 | 0.002 | 8.80E-11 | 0.002 | 0.007 | 0.73 | 43.56 |
| rs232496 | T | C | 0.37 | 0.009 | 0.002 | 7.14E-10 | 0.007 | 0.006 | 0.24 | 36 |
| rs2364544 | A | G | 0.39 | -0.011 | 0.001 | 2.91E-13 | -0.004 | 0.006 | 0.546 | 56.25 |
| rs2434672 | A | C | 0.53 | 0.009 | 0.001 | 1.06E-10 | -0.013 | 0.006 | 0.043 | 42.25 |
| rs2436760 | T | C | 0.97 | 0.023 | 0.004 | 4.52E-08 | 0.011 | 0.018 | 0.523 | 30.51 |
| rs2447097 | T | G | 0.46 | 0.01 | 0.001 | 1.10E-11 | -0.012 | 0.006 | 0.058 | 47.02 |
| rs2469226 | A | T | 0.22 | 0.01 | 0.002 | 1.28E-08 | -0.011 | 0.007 | 0.13 | 31.89 |
| rs2496482 | T | C | 0.42 | 0.012 | 0.002 | 9.20E-17 | -0.002 | 0.006 | 0.761 | 65.07 |
| rs252991 | A | G | 0.37 | 0.01 | 0.002 | 6.30E-11 | 0.006 | 0.006 | 0.379 | 40.11 |
| rs2545795 | A | C | 0.44 | 0.013 | 0.001 | 1.03E-18 | 0.004 | 0.006 | 0.524 | 79.72 |
| rs2554835 | A | G | 0.4 | 0.009 | 0.001 | 8.95E-10 | -0.015 | 0.006 | 0.013 | 39.51 |
| rs2570497 | T | C | 0.64 | -0.012 | 0.002 | 1.27E-16 | 0.001 | 0.006 | 0.86 | 65.07 |
| rs2725370 | T | C | 0.3 | -0.014 | 0.002 | 4.96E-20 | 0.006 | 0.007 | 0.334 | 88.36 |
| rs2764684 | T | C | 0.83 | 0.014 | 0.002 | 9.99E-15 | -0.003 | 0.008 | 0.69 | 56.65 |
| rs2838006 | T | C | 0.36 | 0.013 | 0.002 | 1.38E-19 | 0.011 | 0.006 | 0.084 | 77.44 |
| rs28513882 | A | G | 0.18 | -0.011 | 0.002 | 2.03E-09 | -0.007 | 0.008 | 0.395 | 37.35 |
| rs2885198 | A | G | 0.53 | 0.009 | 0.001 | 4.63E-10 | -0.011 | 0.006 | 0.062 | 39.51 |
| rs2923424 | A | G | 0.61 | 0.012 | 0.001 | 6.31E-17 | 0 | 0.006 | 0.998 | 73.47 |
| rs2929032 | A | G | 0.47 | -0.008 | 0.001 | 8.40E-09 | 0.001 | 0.006 | 0.908 | 33.47 |
| rs2958182 | A | T | 0.34 | 0.008 | 0.002 | 3.46E-08 | -0.008 | 0.006 | 0.194 | 29.88 |
| rs2964199 | T | C | 0.31 | -0.01 | 0.002 | 4.61E-11 | 0 | 0.007 | 0.942 | 43.56 |
| rs2964255 | A | G | 0.31 | 0.009 | 0.002 | 6.34E-09 | 0.005 | 0.007 | 0.459 | 34.42 |
| rs2998299 | T | G | 0.21 | -0.014 | 0.002 | 1.51E-15 | 0.007 | 0.007 | 0.313 | 64 |
| rs3026996 | A | C | 0.76 | 0.014 | 0.002 | 5.42E-17 | -0.002 | 0.007 | 0.756 | 74.39 |
| rs303752 | A | G | 0.41 | -0.011 | 0.001 | 1.47E-14 | -0.005 | 0.006 | 0.433 | 61.73 |
| rs3111251 | T | C | 0.43 | -0.009 | 0.001 | 3.88E-10 | 0.007 | 0.006 | 0.26 | 41.33 |
| rs34067381 | T | G | 0.36 | -0.01 | 0.002 | 9.77E-13 | -0.002 | 0.006 | 0.749 | 48.07 |
| rs34098770 | A | G | 0.15 | -0.016 | 0.002 | 2.95E-16 | 0.004 | 0.008 | 0.649 | 64 |
| rs34305371 | A | G | 0.1 | 0.031 | 0.002 | 1.10E-39 | -0.002 | 0.01 | 0.807 | 171.17 |
| rs34394051 | A | G | 0.84 | -0.012 | 0.002 | 2.30E-09 | -0.032 | 0.008 | 9.73E-05 | 34.22 |
| rs34720381 | T | C | 0.09 | -0.018 | 0.002 | 1.90E-13 | 0.015 | 0.011 | 0.155 | 56.25 |
| rs34807077 | A | C | 0.16 | 0.013 | 0.002 | 3.29E-11 | 0.001 | 0.008 | 0.879 | 45.39 |
| rs35417702 | T | C | 0.52 | -0.015 | 0.001 | 2.40E-27 | 0.002 | 0.006 | 0.699 | 117.88 |
| rs35606437 | A | G | 0.27 | 0.011 | 0.002 | 3.98E-11 | 0.01 | 0.007 | 0.128 | 43.07 |
| rs35929923 | A | G | 0.25 | -0.011 | 0.002 | 1.77E-12 | 0 | 0.007 | 0.968 | 50.77 |
| rs363096 | T | C | 0.42 | -0.014 | 0.001 | 5.77E-24 | -0.009 | 0.006 | 0.134 | 104.33 |
| rs3751331 | A | G | 0.38 | -0.01 | 0.001 | 1.75E-11 | 0.005 | 0.006 | 0.466 | 48.01 |
| rs3809634 | A | G | 0.68 | -0.011 | 0.002 | 6.82E-13 | 0.004 | 0.007 | 0.584 | 52.8 |
| rs382196 | T | G | 0.38 | -0.008 | 0.001 | 1.47E-08 | 0.006 | 0.006 | 0.366 | 34.31 |
| rs3859523 | T | C | 0.82 | 0.012 | 0.002 | 9.92E-12 | 0 | 0.008 | 0.95 | 47.46 |
| rs3897821 | A | G | 0.67 | 0.015 | 0.002 | 3.58E-25 | -0.001 | 0.006 | 0.864 | 105.4 |
| rs3948495 | T | G | 0.41 | -0.009 | 0.001 | 3.95E-10 | -0.001 | 0.006 | 0.916 | 40.41 |
| rs401526 | T | C | 0.53 | -0.01 | 0.001 | 1.15E-13 | -0.009 | 0.006 | 0.147 | 55.18 |
| rs401966 | C | G | 0.39 | -0.01 | 0.001 | 1.72E-11 | 0.008 | 0.006 | 0.192 | 48.01 |
| rs42210 | C | G | 0.71 | -0.01 | 0.002 | 2.74E-10 | 0.003 | 0.007 | 0.687 | 37.52 |
| rs42302 | A | G | 0.34 | 0.009 | 0.002 | 5.96E-09 | -0.016 | 0.006 | 0.015 | 32.87 |
| rs4298514 | T | C | 0.7 | 0.01 | 0.002 | 1.07E-11 | 0.015 | 0.007 | 0.021 | 48.07 |
| rs4320563 | A | G | 0.51 | 0.011 | 0.001 | 2.24E-14 | -0.01 | 0.006 | 0.091 | 60.62 |
| rs4328757 | T | C | 0.61 | 0.01 | 0.001 | 2.95E-11 | -0.009 | 0.006 | 0.165 | 47.02 |
| rs4352658 | T | C | 0.08 | -0.019 | 0.003 | 1.71E-13 | -0.004 | 0.011 | 0.733 | 53.4 |
| rs4358081 | A | C | 0.53 | -0.01 | 0.001 | 1.28E-11 | 0.01 | 0.006 | 0.092 | 46.05 |
| rs4369924 | A | G | 0.16 | 0.013 | 0.002 | 5.58E-11 | 0.003 | 0.008 | 0.746 | 43.28 |
| rs4384309 | A | G | 0.46 | 0.01 | 0.001 | 4.89E-13 | -0.009 | 0.006 | 0.163 | 53.08 |
| rs4467547 | T | G | 0.41 | 0.012 | 0.001 | 4.88E-17 | -0.001 | 0.006 | 0.855 | 74.7 |
| rs4500930 | T | C | 0.34 | -0.011 | 0.002 | 1.67E-13 | 0.005 | 0.006 | 0.415 | 52.8 |
| rs4663617 | A | T | 0.23 | 0.01 | 0.002 | 4.12E-10 | -0.004 | 0.007 | 0.607 | 37.43 |
| rs4673840 | T | C | 0.84 | -0.014 | 0.002 | 5.86E-13 | -0.002 | 0.008 | 0.792 | 52.75 |
| rs4675248 | A | G | 0.4 | -0.009 | 0.001 | 4.50E-11 | -0.001 | 0.006 | 0.874 | 45.08 |
| rs4719944 | T | C | 0.55 | -0.012 | 0.001 | 9.25E-18 | 0.012 | 0.006 | 0.052 | 74.7 |
| rs4726070 | A | G | 0.6 | 0.012 | 0.001 | 1.36E-17 | -0.007 | 0.006 | 0.258 | 75.94 |
| rs4757957 | C | G | 0.69 | 0.013 | 0.002 | 1.79E-18 | -0.01 | 0.007 | 0.142 | 78.62 |
| rs4766424 | C | G | 0.12 | 0.014 | 0.002 | 2.47E-11 | 0 | 0.009 | 1 | 45.72 |
| rs4778058 | T | C | 0.5 | -0.009 | 0.001 | 4.92E-11 | 0 | 0.006 | 0.987 | 43.18 |
| rs4787457 | A | G | 0.63 | 0.016 | 0.002 | 1.23E-28 | 0.007 | 0.006 | 0.279 | 116.64 |
| rs4793090 | A | G | 0.66 | 0.008 | 0.002 | 1.67E-08 | 0.001 | 0.006 | 0.931 | 31.36 |
| rs4810894 | A | G | 0.37 | -0.009 | 0.002 | 4.38E-09 | -0.003 | 0.006 | 0.673 | 32.11 |
| rs4839155 | T | G | 0.77 | 0.012 | 0.002 | 4.66E-12 | -0.007 | 0.007 | 0.302 | 45.76 |
| rs4846724 | A | G | 0.53 | 0.01 | 0.001 | 2.89E-12 | -0.011 | 0.006 | 0.077 | 49 |
| rs4848924 | A | C | 0.71 | -0.012 | 0.002 | 3.56E-14 | 0.006 | 0.007 | 0.347 | 60.84 |
| rs488476 | C | G | 0.62 | -0.011 | 0.002 | 8.29E-15 | 0.005 | 0.006 | 0.429 | 56.75 |
| rs4895650 | T | C | 0.55 | 0.009 | 0.001 | 5.31E-11 | -0.001 | 0.006 | 0.832 | 44.13 |
| rs4899012 | C | G | 0.6 | -0.01 | 0.001 | 8.06E-13 | 0.002 | 0.006 | 0.7 | 54.13 |
| rs4904523 | A | G | 0.52 | -0.008 | 0.001 | 6.27E-09 | -0.003 | 0.006 | 0.605 | 34.31 |
| rs4915735 | A | G | 0.14 | 0.012 | 0.002 | 9.56E-09 | -0.006 | 0.009 | 0.5 | 33.64 |
| rs4977885 | A | G | 0.4 | -0.008 | 0.001 | 6.78E-09 | 0.002 | 0.006 | 0.735 | 36 |
| rs4984541 | A | G | 0.77 | -0.014 | 0.002 | 3.40E-16 | 0.003 | 0.007 | 0.728 | 65.9 |
| rs55736314 | C | G | 0.6 | -0.015 | 0.001 | 4.59E-27 | 0.007 | 0.006 | 0.246 | 121 |
| rs56099375 | T | C | 0.24 | 0.012 | 0.002 | 3.10E-13 | -0.011 | 0.007 | 0.11 | 49.83 |
| rs56319902 | T | C | 0.22 | -0.021 | 0.002 | 5.81E-33 | -0.024 | 0.007 | 7.80E-04 | 146.84 |
| rs56391344 | A | G | 0.25 | 0.015 | 0.002 | 7.49E-20 | -0.006 | 0.007 | 0.408 | 85.56 |
| rs56794817 | A | G | 0.16 | 0.012 | 0.002 | 1.69E-10 | 0.002 | 0.008 | 0.845 | 41.23 |
| rs57349798 | A | G | 0.41 | 0.009 | 0.001 | 8.63E-11 | 0.003 | 0.006 | 0.635 | 44.13 |
| rs57352738 | A | T | 0.2 | -0.015 | 0.002 | 2.51E-18 | -0.003 | 0.008 | 0.738 | 79.94 |
| rs575113 | A | G | 0.29 | 0.013 | 0.002 | 7.52E-18 | -0.012 | 0.007 | 0.076 | 78.62 |
| rs5754753 | T | C | 0.72 | -0.013 | 0.002 | 6.15E-16 | -0.003 | 0.007 | 0.673 | 62.02 |
| rs57661533 | T | C | 0.13 | -0.011 | 0.002 | 3.24E-08 | 0.013 | 0.009 | 0.155 | 29.47 |
| rs580652 | T | C | 0.9 | -0.015 | 0.002 | 4.65E-10 | -0.024 | 0.01 | 0.02 | 37.52 |
| rs590013 | T | C | 0.68 | 0.01 | 0.002 | 2.47E-11 | 0.002 | 0.006 | 0.781 | 44.44 |
| rs6043521 | T | C | 0.6 | -0.008 | 0.001 | 9.72E-09 | 0.004 | 0.006 | 0.487 | 35.15 |
| rs6065080 | T | C | 0.36 | -0.013 | 0.002 | 7.49E-19 | -0.002 | 0.006 | 0.741 | 75.11 |
| rs6065784 | C | G | 0.7 | 0.011 | 0.002 | 5.78E-13 | 0.006 | 0.007 | 0.348 | 53.78 |
| rs61104616 | A | G | 0.52 | -0.017 | 0.001 | 1.56E-34 | 0.009 | 0.006 | 0.152 | 150.94 |
| rs6123924 | A | G | 0.84 | 0.013 | 0.002 | 3.20E-11 | 0.005 | 0.008 | 0.529 | 46.1 |
| rs61527214 | A | G | 0.41 | 0.011 | 0.001 | 1.13E-14 | -0.005 | 0.006 | 0.444 | 61.73 |
| rs61997667 | T | C | 0.15 | -0.014 | 0.002 | 2.24E-13 | -0.007 | 0.008 | 0.387 | 51.84 |
| rs62155350 | A | G | 0.02 | 0.034 | 0.005 | 1.69E-10 | -0.01 | 0.022 | 0.646 | 40.67 |
| rs62155873 | T | C | 0.12 | -0.015 | 0.002 | 1.65E-12 | 0 | 0.009 | 0.98 | 51.7 |
| rs62174974 | A | G | 0.19 | -0.011 | 0.002 | 2.76E-09 | -0.006 | 0.008 | 0.435 | 34.03 |
| rs62179650 | A | G | 0.3 | 0.011 | 0.002 | 5.66E-13 | 0.003 | 0.007 | 0.657 | 49 |
| rs62182994 | T | C | 0.69 | 0.014 | 0.002 | 2.55E-19 | -0.009 | 0.006 | 0.166 | 83.42 |
| rs62379838 | T | C | 0.7 | 0.012 | 0.002 | 1.77E-15 | -0.008 | 0.007 | 0.212 | 65.07 |
| rs628993 | A | G | 0.11 | 0.013 | 0.002 | 3.83E-08 | -0.008 | 0.011 | 0.468 | 30.97 |
| rs6449503 | A | G | 0.5 | 0.018 | 0.001 | 1.84E-39 | -0.008 | 0.006 | 0.175 | 174.62 |
| rs6490618 | T | C | 0.33 | 0.009 | 0.002 | 3.48E-09 | -0.005 | 0.006 | 0.395 | 34.42 |
| rs6493265 | T | C | 0.39 | -0.012 | 0.001 | 2.26E-17 | -0.006 | 0.006 | 0.363 | 75.94 |
| rs6557171 | T | C | 0.32 | -0.016 | 0.002 | 5.48E-25 | -0.008 | 0.006 | 0.19 | 106.78 |
| rs660001 | A | G | 0.21 | -0.016 | 0.002 | 1.66E-21 | -0.004 | 0.007 | 0.595 | 93.07 |
| rs66568921 | T | G | 0.64 | -0.018 | 0.002 | 7.60E-34 | 0.007 | 0.006 | 0.254 | 145.6 |
| rs66721975 | A | G | 0.29 | -0.009 | 0.002 | 1.51E-08 | -0.005 | 0.007 | 0.459 | 33.64 |
| rs6690195 | T | C | 0.49 | -0.011 | 0.001 | 7.89E-16 | 0.001 | 0.006 | 0.918 | 65.15 |
| rs6697584 | T | C | 0.79 | -0.013 | 0.002 | 1.43E-13 | -0.001 | 0.007 | 0.907 | 54.93 |
| rs6704768 | A | G | 0.56 | -0.012 | 0.001 | 1.84E-16 | 0.005 | 0.006 | 0.448 | 68.65 |
| rs6731373 | A | G | 0.34 | -0.012 | 0.002 | 1.39E-14 | -0.012 | 0.006 | 0.06 | 58.78 |
| rs6812533 | T | C | 0.74 | 0.011 | 0.002 | 4.85E-11 | 0.009 | 0.007 | 0.181 | 43.89 |
| rs68145588 | T | G | 0.14 | -0.013 | 0.002 | 7.53E-10 | -0.003 | 0.009 | 0.757 | 36.57 |
| rs6917154 | T | C | 0.12 | 0.013 | 0.002 | 2.55E-09 | 0.001 | 0.009 | 0.919 | 36.57 |
| rs6924023 | A | G | 0.75 | 0.011 | 0.002 | 5.17E-11 | 0.007 | 0.007 | 0.316 | 44.72 |
| rs6977237 | T | C | 0.47 | 0.008 | 0.001 | 5.59E-09 | 0.01 | 0.006 | 0.107 | 34.31 |
| rs6994287 | A | G | 0.41 | 0.01 | 0.001 | 1.11E-12 | -0.001 | 0.006 | 0.931 | 53.08 |
| rs7012546 | T | C | 0.42 | 0.009 | 0.001 | 1.25E-10 | -0.007 | 0.006 | 0.273 | 43.18 |
| rs7029718 | A | G | 0.41 | 0.024 | 0.001 | 7.82E-65 | -0.001 | 0.006 | 0.854 | 303.76 |
| rs7041702 | A | G | 0.74 | -0.011 | 0.002 | 4.53E-12 | -0.005 | 0.007 | 0.471 | 48.13 |
| rs7167688 | T | C | 0.49 | -0.009 | 0.001 | 1.71E-11 | 0.01 | 0.006 | 0.098 | 45.08 |
| rs7171405 | A | G | 0.24 | -0.01 | 0.002 | 7.53E-10 | 0.005 | 0.007 | 0.457 | 39.85 |
| rs717996 | T | C | 0.6 | -0.012 | 0.001 | 1.89E-17 | -0.004 | 0.006 | 0.52 | 75.94 |
| rs7226824 | T | C | 0.53 | -0.008 | 0.001 | 2.72E-08 | -0.001 | 0.006 | 0.901 | 31.04 |
| rs72486027 | T | C | 0.25 | -0.01 | 0.002 | 1.61E-10 | -0.008 | 0.007 | 0.271 | 42.25 |
| rs7254263 | T | C | 0.28 | -0.012 | 0.002 | 1.84E-14 | 0.003 | 0.007 | 0.624 | 55.32 |
| rs72622559 | T | C | 0.23 | -0.012 | 0.002 | 5.50E-13 | -0.005 | 0.007 | 0.52 | 49.83 |
| rs72672052 | A | T | 0.18 | -0.01 | 0.002 | 4.87E-08 | 0.001 | 0.008 | 0.917 | 31.48 |
| rs72709560 | A | G | 0.69 | 0.009 | 0.002 | 2.03E-08 | 0.013 | 0.006 | 0.049 | 32.11 |
| rs72828517 | T | C | 0.83 | -0.017 | 0.002 | 1.38E-19 | 0.003 | 0.008 | 0.741 | 78.18 |
| rs72829857 | A | G | 0.76 | -0.015 | 0.002 | 1.52E-19 | -0.004 | 0.007 | 0.624 | 86.72 |
| rs72917504 | T | C | 0.06 | -0.018 | 0.003 | 2.91E-09 | 0.003 | 0.013 | 0.84 | 35.2 |
| rs72944064 | T | C | 0.74 | -0.01 | 0.002 | 9.39E-10 | -0.01 | 0.007 | 0.156 | 37.52 |
| rs72962169 | T | C | 0.16 | -0.017 | 0.002 | 1.26E-19 | 0.009 | 0.008 | 0.298 | 83.87 |
| rs72993796 | T | C | 0.88 | 0.013 | 0.002 | 1.12E-08 | 0.011 | 0.009 | 0.241 | 33.32 |
| rs730384 | A | G | 0.44 | 0.01 | 0.001 | 9.38E-13 | -0.004 | 0.006 | 0.565 | 52.05 |
| rs73055556 | A | G | 0.14 | 0.011 | 0.002 | 4.95E-08 | -0.006 | 0.009 | 0.486 | 30.8 |
| rs73191311 | A | G | 0.34 | -0.009 | 0.002 | 2.03E-10 | 0.005 | 0.006 | 0.404 | 39.27 |
| rs7321274 | A | G | 0.8 | 0.011 | 0.002 | 8.21E-11 | 0.006 | 0.008 | 0.431 | 40.11 |
| rs7326331 | A | G | 0.28 | -0.013 | 0.002 | 3.42E-16 | 0.01 | 0.007 | 0.159 | 64 |
| rs73344830 | A | G | 0.42 | 0.017 | 0.001 | 1.00E-32 | 0 | 0.006 | 0.985 | 147.45 |
| rs736281 | T | C | 0.39 | 0.009 | 0.001 | 2.49E-09 | -0.009 | 0.006 | 0.147 | 37.73 |
| rs73648455 | T | C | 0.08 | -0.018 | 0.003 | 1.99E-12 | 0 | 0.011 | 0.978 | 49.54 |
| rs743316 | T | C | 0.79 | 0.011 | 0.002 | 8.21E-11 | -0.007 | 0.007 | 0.343 | 43.4 |
| rs74415461 | T | C | 0.08 | 0.018 | 0.003 | 6.25E-12 | 0.02 | 0.011 | 0.065 | 45.82 |
| rs7449561 | A | G | 0.23 | 0.01 | 0.002 | 9.73E-10 | -0.013 | 0.007 | 0.067 | 36 |
| rs7460106 | T | C | 0.76 | -0.012 | 0.002 | 2.06E-13 | -0.008 | 0.007 | 0.301 | 52.35 |
| rs746839 | C | G | 0.63 | 0.014 | 0.002 | 1.19E-20 | -0.002 | 0.006 | 0.708 | 81 |
| rs74787922 | A | C | 0.93 | 0.018 | 0.003 | 8.97E-11 | 0.003 | 0.012 | 0.799 | 42.25 |
| rs75033012 | C | G | 0.04 | -0.026 | 0.004 | 1.38E-11 | 0.004 | 0.015 | 0.794 | 46.1 |
| rs75177132 | T | C | 0.05 | 0.03 | 0.004 | 1.01E-16 | -0.005 | 0.014 | 0.73 | 68.52 |
| rs7575637 | A | G | 0.45 | 0.011 | 0.001 | 3.43E-14 | -0.013 | 0.006 | 0.028 | 58.41 |
| rs7603132 | A | G | 0.19 | 0.014 | 0.002 | 1.20E-14 | -0.001 | 0.008 | 0.884 | 57.93 |
| rs76076331 | T | C | 0.13 | 0.021 | 0.002 | 1.71E-23 | -0.002 | 0.009 | 0.851 | 96.23 |
| rs7617204 | A | G | 0.52 | -0.01 | 0.001 | 4.33E-12 | -0.011 | 0.006 | 0.06 | 48.01 |
| rs76235882 | A | G | 0.97 | 0.024 | 0.004 | 4.52E-10 | -0.007 | 0.018 | 0.705 | 38.19 |
| rs76246107 | A | G | 0.09 | -0.016 | 0.003 | 6.52E-10 | 0.003 | 0.012 | 0.793 | 38.82 |
| rs7650602 | T | C | 0.56 | -0.009 | 0.001 | 2.77E-10 | 0.006 | 0.006 | 0.329 | 40.41 |
| rs76577427 | C | G | 0.9 | 0.02 | 0.002 | 7.78E-18 | -0.006 | 0.01 | 0.569 | 71.54 |
| rs7672622 | A | G | 0.75 | -0.009 | 0.002 | 4.72E-08 | 0.003 | 0.007 | 0.65 | 30.25 |
| rs7683416 | T | C | 0.46 | 0.014 | 0.001 | 6.05E-23 | -0.008 | 0.006 | 0.177 | 98.58 |
| rs76878669 | C | G | 0.76 | 0.014 | 0.002 | 3.10E-17 | -0.005 | 0.007 | 0.455 | 68.79 |
| rs7692359 | T | C | 0.78 | -0.011 | 0.002 | 1.40E-10 | -0.003 | 0.007 | 0.68 | 41.11 |
| rs77025239 | A | G | 0.16 | -0.014 | 0.002 | 4.65E-13 | -0.01 | 0.008 | 0.241 | 54.29 |
| rs77609760 | A | G | 0.93 | 0.024 | 0.003 | 7.71E-17 | 0.012 | 0.012 | 0.307 | 71.04 |
| rs77719387 | A | T | 0.02 | -0.04 | 0.006 | 3.78E-12 | 0.009 | 0.026 | 0.739 | 48.28 |
| rs7849487 | T | G | 0.66 | -0.017 | 0.002 | 1.25E-29 | -0.004 | 0.006 | 0.503 | 123.95 |
| rs78648104 | T | C | 0.91 | -0.014 | 0.002 | 1.70E-08 | 0.021 | 0.011 | 0.05 | 31.81 |
| rs7875078 | A | C | 0.46 | -0.008 | 0.001 | 2.72E-08 | -0.002 | 0.006 | 0.795 | 31.04 |
| rs7894722 | T | C | 0.37 | 0.009 | 0.001 | 5.41E-09 | -0.006 | 0.006 | 0.377 | 36.86 |
| rs790647 | A | C | 0.23 | -0.015 | 0.002 | 3.81E-18 | 0.012 | 0.007 | 0.106 | 72.75 |
| rs7910403 | T | G | 0.2 | 0.012 | 0.002 | 5.08E-12 | 0.009 | 0.008 | 0.247 | 45.94 |
| rs7924036 | T | G | 0.51 | 0.013 | 0.001 | 2.55E-21 | 0.014 | 0.006 | 0.019 | 90.25 |
| rs79265434 | A | G | 0.88 | -0.02 | 0.002 | 1.98E-19 | -0.011 | 0.009 | 0.234 | 80.18 |
| rs7972246 | T | C | 0.35 | 0.011 | 0.002 | 1.01E-13 | 0.001 | 0.006 | 0.909 | 53.78 |
| rs79728014 | A | G | 0.86 | -0.013 | 0.002 | 2.06E-11 | 0.003 | 0.009 | 0.709 | 44.89 |
| rs7974852 | A | C | 0.52 | 0.012 | 0.001 | 1.14E-16 | -0.01 | 0.006 | 0.104 | 69.84 |
| rs7977614 | A | G | 0.71 | -0.013 | 0.002 | 6.84E-17 | -0.002 | 0.007 | 0.714 | 70.14 |
| rs79994730 | T | C | 0.04 | 0.024 | 0.004 | 5.88E-12 | -0.026 | 0.014 | 0.065 | 47.02 |
| rs8008382 | T | C | 0.3 | -0.011 | 0.002 | 4.07E-12 | -0.01 | 0.007 | 0.12 | 49.94 |
| rs8024 | A | C | 0.33 | -0.012 | 0.002 | 1.13E-14 | -0.013 | 0.006 | 0.049 | 58.78 |
| rs80257979 | T | G | 0.97 | -0.025 | 0.004 | 3.31E-10 | 0.008 | 0.018 | 0.643 | 39.69 |
| rs8030487 | A | G | 0.69 | 0.009 | 0.002 | 2.52E-08 | -0.006 | 0.007 | 0.393 | 32.11 |
| rs806816 | A | T | 0.75 | 0.01 | 0.002 | 4.80E-10 | 0 | 0.007 | 0.958 | 39.85 |
| rs8097125 | T | C | 0.38 | 0.009 | 0.001 | 2.31E-09 | -0.004 | 0.006 | 0.47 | 37.73 |
| rs912883 | T | C | 0.68 | 0.009 | 0.002 | 6.20E-09 | 0.008 | 0.006 | 0.227 | 33.64 |
| rs9289300 | T | C | 0.84 | -0.016 | 0.002 | 1.00E-17 | 0.003 | 0.008 | 0.756 | 74.5 |
| rs929511 | T | C | 0.13 | -0.016 | 0.002 | 3.91E-14 | 0.002 | 0.009 | 0.851 | 58.05 |
| rs9359939 | A | C | 0.24 | -0.011 | 0.002 | 1.51E-10 | -0.008 | 0.007 | 0.253 | 43.07 |
| rs936496 | A | G | 0.62 | -0.008 | 0.001 | 4.38E-08 | 0.004 | 0.006 | 0.521 | 31.84 |
| rs9373363 | A | G | 0.75 | -0.011 | 0.002 | 5.54E-12 | -0.014 | 0.007 | 0.051 | 48.13 |
| rs9375188 | T | C | 0.48 | 0.021 | 0.001 | 8.78E-52 | 0.003 | 0.006 | 0.597 | 229.31 |
| rs9411331 | A | C | 0.68 | 0.015 | 0.002 | 7.63E-22 | 0.002 | 0.006 | 0.716 | 93.44 |
| rs9492774 | C | G | 0.37 | 0.008 | 0.002 | 4.33E-08 | 0.003 | 0.006 | 0.655 | 28.44 |
| rs9529146 | T | C | 0.23 | 0.013 | 0.002 | 3.48E-14 | -0.017 | 0.007 | 0.02 | 55.81 |
| rs9556958 | T | C | 0.52 | -0.011 | 0.001 | 5.96E-15 | 0.009 | 0.006 | 0.131 | 61.73 |
| rs9616947 | T | C | 0.38 | -0.013 | 0.002 | 3.58E-20 | 0.013 | 0.006 | 0.044 | 79.8 |
| rs9633970 | T | C | 0.76 | 0.009 | 0.002 | 2.81E-08 | -0.003 | 0.007 | 0.7 | 32.35 |
| rs9655780 | A | G | 0.83 | -0.015 | 0.002 | 3.70E-16 | 0.003 | 0.008 | 0.733 | 64.84 |
| rs969512 | A | T | 0.66 | -0.011 | 0.002 | 9.84E-13 | 0.008 | 0.006 | 0.214 | 49.94 |
| rs9853928 | T | C | 0.21 | -0.013 | 0.002 | 1.59E-13 | -0.003 | 0.008 | 0.685 | 55.81 |
| rs9859556 | T | G | 0.31 | 0.029 | 0.002 | 4.61E-82 | -0.012 | 0.007 | 0.072 | 373.78 |
| rs9882532 | T | C | 0.64 | 0.012 | 0.002 | 2.15E-17 | 0.002 | 0.006 | 0.73 | 68.34 |
| rs9886703 | A | T | 0.17 | -0.014 | 0.002 | 4.46E-13 | 0.013 | 0.008 | 0.115 | 51.24 |
| rs9927137 | A | G | 0.53 | 0.008 | 0.001 | 5.11E-09 | -0.005 | 0.006 | 0.434 | 34.31 |
| rs9927842 | T | C | 0.15 | -0.013 | 0.002 | 6.22E-11 | 0.014 | 0.008 | 0.088 | 42.25 |
| rs9929556 | T | G | 0.57 | -0.01 | 0.001 | 2.31E-12 | -0.004 | 0.006 | 0.481 | 50.01 |
| rs9929762 | A | G | 0.56 | 0.01 | 0.001 | 1.03E-11 | 0.005 | 0.006 | 0.37 | 47.02 |
| rs9933256 | A | G | 0.56 | 0.012 | 0.001 | 6.06E-17 | 0 | 0.006 | 0.968 | 72.25 |

**SNPs used as instrumental variables in the MR analysis of educational attainment on orientation dispersion index**

| **SNP** | **Effect allele** | **Other allele** | **Effect allele frequency** | **beta (exposure)** | **SE (exposure)** | ***P* (exposure)** | **beta (outcome)** | **SE (outcome)** | ***P* (outcome)** | ***F*-statistic** |
| --- | --- | --- | --- | --- | --- | --- | --- | --- | --- | --- |
| rs10006235 | T | C | 0.27 | -0.009 | 0.002 | 4.48E-09 | 0.012 | 0.009 | 0.144 | 33.79 |
| rs10021733 | T | C | 0.15 | -0.016 | 0.003 | 9.45E-10 | 0.006 | 0.01 | 0.532 | 36.89 |
| rs10046069 | A | G | 0.12 | 0.013 | 0.002 | 4.40E-09 | -0.012 | 0.012 | 0.307 | 33.32 |
| rs10060023 | T | C | 0.33 | 0.011 | 0.002 | 1.88E-13 | -0.002 | 0.008 | 0.766 | 53.78 |
| rs10080647 | A | C | 0.14 | 0.012 | 0.002 | 7.05E-10 | -0.016 | 0.011 | 0.151 | 38.44 |
| rs1008078 | T | C | 0.4 | -0.018 | 0.001 | 4.50E-37 | 0.009 | 0.008 | 0.274 | 169 |
| rs10189857 | A | G | 0.57 | 0.016 | 0.001 | 4.69E-29 | 0.014 | 0.008 | 0.068 | 127.37 |
| rs10193498 | A | T | 0.76 | 0.01 | 0.002 | 3.10E-09 | -0.003 | 0.009 | 0.736 | 36.75 |
| rs10215082 | A | G | 0.47 | -0.015 | 0.001 | 7.03E-26 | 0.011 | 0.008 | 0.131 | 113.27 |
| rs10460095 | A | G | 0.57 | -0.012 | 0.001 | 2.61E-18 | -0.009 | 0.008 | 0.251 | 78.45 |
| rs10761251 | A | T | 0.66 | 0.011 | 0.002 | 2.52E-14 | -0.003 | 0.008 | 0.721 | 56.75 |
| rs10772644 | C | G | 0.89 | 0.014 | 0.002 | 1.69E-10 | -0.006 | 0.012 | 0.611 | 41.66 |
| rs10773002 | A | T | 0.25 | 0.018 | 0.002 | 2.50E-30 | 0.012 | 0.009 | 0.165 | 133.69 |
| rs10773208 | T | C | 0.25 | -0.011 | 0.002 | 8.09E-12 | 0.006 | 0.009 | 0.504 | 47.27 |
| rs10798888 | T | G | 0.17 | -0.014 | 0.002 | 5.15E-14 | 0.029 | 0.01 | 0.004 | 54.29 |
| rs10799615 | A | G | 0.25 | -0.009 | 0.002 | 4.11E-08 | 0.003 | 0.009 | 0.692 | 30.94 |
| rs10805383 | A | G | 0.49 | -0.01 | 0.001 | 2.22E-13 | -0.003 | 0.008 | 0.712 | 54.13 |
| rs10830858 | T | C | 0.51 | 0.009 | 0.001 | 5.31E-11 | 0.002 | 0.008 | 0.828 | 43.18 |
| rs10844179 | A | G | 0.23 | 0.01 | 0.002 | 5.03E-10 | -0.006 | 0.009 | 0.486 | 37.43 |
| rs10875121 | C | G | 0.84 | 0.017 | 0.002 | 4.46E-20 | 0.016 | 0.01 | 0.121 | 83.87 |
| rs10892807 | T | C | 0.57 | 0.011 | 0.001 | 1.09E-14 | -0.003 | 0.008 | 0.649 | 60.62 |
| rs10940921 | T | G | 0.42 | 0.011 | 0.001 | 1.31E-14 | -0.005 | 0.008 | 0.494 | 62.86 |
| rs10951590 | T | C | 0.33 | -0.011 | 0.002 | 3.62E-13 | -0.005 | 0.008 | 0.525 | 52.8 |
| rs10979613 | T | C | 0.64 | -0.012 | 0.002 | 1.25E-15 | -0.026 | 0.008 | 0.001 | 60.84 |
| rs10996167 | C | G | 0.65 | 0.011 | 0.002 | 7.47E-09 | 0.01 | 0.008 | 0.231 | 31.92 |
| rs11003463 | T | G | 0.61 | 0.009 | 0.002 | 5.58E-10 | -0.003 | 0.008 | 0.676 | 36.8 |
| rs11023764 | A | G | 0.64 | 0.009 | 0.002 | 5.32E-10 | -0.003 | 0.008 | 0.736 | 36.8 |
| rs11030102 | C | G | 0.75 | 0.01 | 0.002 | 4.60E-10 | 0 | 0.009 | 0.993 | 39.85 |
| rs11076962 | T | C | 0.72 | 0.009 | 0.002 | 4.29E-08 | 0.009 | 0.008 | 0.29 | 28.89 |
| rs11081529 | T | C | 0.71 | 0.013 | 0.002 | 5.22E-16 | -0.002 | 0.008 | 0.85 | 70.56 |
| rs11082011 | T | C | 0.67 | 0.019 | 0.002 | 2.06E-38 | 0.011 | 0.008 | 0.175 | 167.27 |
| rs11121177 | A | G | 0.18 | 0.015 | 0.002 | 1.44E-16 | 0.001 | 0.01 | 0.921 | 70.37 |
| rs11130380 | T | G | 0.61 | 0.009 | 0.001 | 4.17E-10 | -0.008 | 0.008 | 0.289 | 41.33 |
| rs11138947 | T | C | 0.72 | 0.011 | 0.002 | 2.36E-11 | -0.008 | 0.008 | 0.352 | 43.07 |
| rs11157931 | A | C | 0.39 | -0.014 | 0.001 | 3.86E-22 | 0.006 | 0.008 | 0.445 | 98.58 |
| rs111852224 | T | C | 0.12 | 0.016 | 0.002 | 4.85E-13 | -0.005 | 0.011 | 0.685 | 50.28 |
| rs11211123 | A | G | 0.22 | 0.01 | 0.002 | 7.62E-10 | -0.016 | 0.009 | 0.08 | 37.43 |
| rs11213482 | A | G | 0.84 | -0.011 | 0.002 | 2.68E-08 | 0.006 | 0.01 | 0.568 | 31.12 |
| rs112603734 | A | C | 0.77 | -0.013 | 0.002 | 2.72E-09 | 0.002 | 0.009 | 0.802 | 35.46 |
| rs112806496 | C | G | 0.91 | -0.016 | 0.002 | 1.72E-10 | -0.002 | 0.013 | 0.893 | 40.96 |
| rs1128956 | T | G | 0.83 | -0.014 | 0.002 | 5.34E-14 | -0.016 | 0.01 | 0.11 | 54.29 |
| rs113520408 | A | G | 0.28 | 0.016 | 0.002 | 1.03E-22 | -0.007 | 0.008 | 0.377 | 93.85 |
| rs113615161 | T | C | 0.13 | -0.012 | 0.002 | 2.98E-09 | -0.007 | 0.011 | 0.526 | 34.31 |
| rs114593137 | A | T | 0.79 | -0.012 | 0.002 | 1.65E-12 | -0.012 | 0.009 | 0.185 | 51.5 |
| rs115000530 | A | T | 0.95 | -0.025 | 0.003 | 1.09E-15 | -0.017 | 0.017 | 0.32 | 65.04 |
| rs11542663 | A | C | 0.69 | 0.011 | 0.002 | 2.44E-13 | -0.003 | 0.008 | 0.712 | 55.75 |
| rs11588857 | A | G | 0.21 | 0.02 | 0.002 | 4.45E-31 | -0.009 | 0.009 | 0.323 | 137.03 |
| rs11598765 | A | G | 0.2 | -0.011 | 0.002 | 1.73E-09 | -0.004 | 0.01 | 0.664 | 34.68 |
| rs11620355 | A | G | 0.09 | 0.018 | 0.002 | 1.09E-13 | 0.016 | 0.013 | 0.248 | 53 |
| rs11657342 | A | G | 0.37 | 0.014 | 0.002 | 2.32E-13 | 0 | 0.008 | 0.979 | 54.29 |
| rs11657979 | A | G | 0.24 | -0.009 | 0.002 | 4.57E-08 | 0.003 | 0.009 | 0.763 | 28.65 |
| rs11663602 | A | C | 0.28 | -0.013 | 0.002 | 9.27E-17 | -0.002 | 0.009 | 0.779 | 66.02 |
| rs1167827 | A | G | 0.43 | 0.01 | 0.001 | 2.93E-13 | 0.014 | 0.008 | 0.062 | 55.18 |
| rs11724690 | T | G | 0.29 | 0.009 | 0.002 | 2.08E-09 | -0.004 | 0.008 | 0.622 | 38.44 |
| rs117398064 | C | G | 0.09 | -0.014 | 0.002 | 1.16E-08 | 0.007 | 0.013 | 0.577 | 31.81 |
| rs118134876 | T | C | 0.06 | -0.022 | 0.003 | 1.50E-13 | -0.009 | 0.016 | 0.585 | 55.25 |
| rs11871429 | A | G | 0.77 | 0.013 | 0.002 | 2.52E-14 | 0.001 | 0.009 | 0.877 | 56.69 |
| rs11894424 | A | C | 0.95 | 0.021 | 0.003 | 2.14E-10 | 0.015 | 0.018 | 0.415 | 40.11 |
| rs12054166 | C | G | 0.74 | 0.01 | 0.002 | 2.30E-09 | 0.009 | 0.009 | 0.323 | 36 |
| rs12076635 | C | G | 0.78 | 0.021 | 0.002 | 3.33E-34 | -0.025 | 0.009 | 0.007 | 148.27 |
| rs12113634 | T | C | 0.65 | 0.009 | 0.002 | 2.50E-09 | -0.007 | 0.008 | 0.368 | 34.42 |
| rs12151248 | T | C | 0.11 | -0.015 | 0.002 | 3.81E-11 | 0.01 | 0.012 | 0.412 | 41.97 |
| rs1220779 | A | G | 0.47 | -0.008 | 0.001 | 1.58E-08 | -0.001 | 0.008 | 0.845 | 31.84 |
| rs12273435 | A | G | 0.21 | -0.013 | 0.002 | 4.72E-14 | -0.001 | 0.009 | 0.913 | 60.29 |
| rs12332731 | A | T | 0.19 | 0.014 | 0.002 | 5.96E-14 | 0.006 | 0.01 | 0.501 | 56.25 |
| rs12375949 | T | C | 0.43 | -0.014 | 0.001 | 1.61E-22 | -0.006 | 0.008 | 0.428 | 97.16 |
| rs12438177 | A | G | 0.37 | 0.009 | 0.002 | 1.48E-10 | -0.005 | 0.008 | 0.511 | 38.44 |
| rs12468040 | T | G | 0.38 | 0.014 | 0.001 | 3.12E-21 | -0.009 | 0.008 | 0.228 | 95.76 |
| rs12503522 | T | C | 0.28 | -0.011 | 0.002 | 3.93E-13 | -0.001 | 0.008 | 0.864 | 49.88 |
| rs12506221 | T | G | 0.44 | -0.013 | 0.001 | 4.23E-20 | -0.004 | 0.008 | 0.622 | 86.22 |
| rs12519073 | T | C | 0.23 | -0.011 | 0.002 | 1.16E-10 | 0.004 | 0.009 | 0.669 | 39.62 |
| rs12524795 | T | C | 0.44 | 0.01 | 0.001 | 5.28E-13 | -0.006 | 0.008 | 0.44 | 53.08 |
| rs12571549 | A | G | 0.14 | 0.015 | 0.002 | 1.45E-13 | 0.001 | 0.011 | 0.898 | 57 |
| rs12574281 | A | C | 0.63 | -0.008 | 0.002 | 3.04E-08 | -0.006 | 0.008 | 0.408 | 28.44 |
| rs12591647 | T | C | 0.82 | -0.015 | 0.002 | 7.78E-17 | 0.002 | 0.01 | 0.85 | 71.31 |
| rs12602286 | T | G | 0.87 | 0.016 | 0.002 | 4.62E-14 | -0.015 | 0.011 | 0.191 | 58.05 |
| rs12614263 | A | G | 0.5 | 0.008 | 0.001 | 3.38E-09 | -0.01 | 0.008 | 0.196 | 35.15 |
| rs12638072 | A | G | 0.3 | -0.011 | 0.002 | 3.13E-12 | 0.01 | 0.008 | 0.243 | 50.88 |
| rs12643771 | T | C | 0.31 | 0.013 | 0.002 | 1.32E-17 | 0.013 | 0.008 | 0.123 | 75.11 |
| rs12646523 | T | C | 0.25 | -0.013 | 0.002 | 3.79E-16 | 0.003 | 0.009 | 0.746 | 68.06 |
| rs12670376 | A | G | 0.45 | 0.009 | 0.001 | 5.99E-11 | -0.005 | 0.008 | 0.477 | 43.18 |
| rs12761761 | T | C | 0.24 | 0.015 | 0.002 | 4.79E-20 | 0.013 | 0.009 | 0.149 | 81 |
| rs12765185 | A | T | 0.28 | -0.009 | 0.002 | 2.96E-08 | 0.01 | 0.008 | 0.258 | 30.25 |
| rs12789313 | T | C | 0.49 | 0.009 | 0.001 | 6.79E-11 | 0.006 | 0.008 | 0.465 | 43.18 |
| rs12875339 | A | C | 0.64 | -0.011 | 0.002 | 1.71E-13 | 0.014 | 0.008 | 0.068 | 51.84 |
| rs12888615 | T | C | 0.2 | 0.012 | 0.002 | 9.42E-12 | 0.004 | 0.009 | 0.647 | 44.44 |
| rs12957463 | A | G | 0.8 | 0.015 | 0.002 | 9.20E-17 | 0.001 | 0.009 | 0.955 | 65.79 |
| rs12967010 | T | C | 0.77 | 0.011 | 0.002 | 1.33E-11 | 0.002 | 0.009 | 0.852 | 44.97 |
| rs12981405 | T | C | 0.17 | -0.011 | 0.002 | 2.75E-09 | 0.025 | 0.01 | 0.014 | 34.75 |
| rs1301838 | T | C | 0.32 | -0.01 | 0.002 | 1.29E-10 | 0.002 | 0.008 | 0.845 | 40.96 |
| rs13018640 | T | C | 0.6 | -0.021 | 0.001 | 1.05E-50 | 0.004 | 0.008 | 0.568 | 235.84 |
| rs13050131 | A | C | 0.66 | -0.008 | 0.002 | 4.40E-08 | -0.004 | 0.008 | 0.664 | 29.16 |
| rs13133213 | A | G | 0.5 | 0.01 | 0.001 | 7.57E-12 | 0.019 | 0.008 | 0.012 | 47.02 |
| rs13145650 | T | C | 0.92 | -0.017 | 0.002 | 8.99E-12 | 0.005 | 0.013 | 0.722 | 47.89 |
| rs13163845 | T | C | 0.85 | -0.015 | 0.002 | 4.66E-14 | -0.01 | 0.011 | 0.34 | 55.5 |
| rs13197257 | T | G | 0.28 | 0.011 | 0.002 | 2.83E-12 | 0.01 | 0.009 | 0.223 | 46.41 |
| rs13212041 | T | C | 0.8 | 0.01 | 0.002 | 7.42E-09 | 0.002 | 0.009 | 0.856 | 31.48 |
| rs13240401 | T | C | 0.78 | 0.018 | 0.002 | 2.36E-24 | 0.005 | 0.009 | 0.554 | 107.18 |
| rs13261773 | C | G | 0.16 | 0.012 | 0.002 | 3.12E-10 | 0.004 | 0.011 | 0.68 | 41.23 |
| rs13266287 | A | T | 0.67 | -0.009 | 0.002 | 1.75E-08 | 0 | 0.008 | 0.99 | 32.11 |
| rs1329125 | T | C | 0.33 | -0.01 | 0.002 | 8.37E-11 | 0.017 | 0.008 | 0.039 | 41.82 |
| rs13327482 | A | G | 0.82 | -0.012 | 0.002 | 4.47E-11 | 0.013 | 0.01 | 0.177 | 45.19 |
| rs1334297 | A | G | 0.74 | 0.026 | 0.002 | 2.53E-58 | 0.011 | 0.009 | 0.217 | 258 |
| rs13422673 | T | C | 0.47 | -0.011 | 0.001 | 6.48E-15 | -0.009 | 0.008 | 0.246 | 61.73 |
| rs137079 | T | C | 0.14 | 0.013 | 0.002 | 3.06E-10 | -0.027 | 0.011 | 0.011 | 40.96 |
| rs1426619 | T | C | 0.43 | 0.01 | 0.001 | 1.40E-11 | 0.007 | 0.008 | 0.364 | 47.02 |
| rs1434630 | T | G | 0.15 | -0.014 | 0.002 | 4.63E-12 | 0.002 | 0.011 | 0.885 | 46.92 |
| rs143743568 | A | G | 0.15 | 0.012 | 0.002 | 4.99E-09 | 0.004 | 0.01 | 0.685 | 34.81 |
| rs150537577 | A | G | 0.92 | -0.015 | 0.003 | 9.50E-09 | -0.004 | 0.014 | 0.756 | 33.73 |
| rs1505676 | C | G | 0.63 | 0.008 | 0.002 | 6.67E-09 | -0.004 | 0.008 | 0.575 | 31.36 |
| rs1542354 | A | G | 0.48 | -0.008 | 0.001 | 2.72E-09 | -0.002 | 0.008 | 0.785 | 36 |
| rs1544 | A | G | 0.26 | -0.01 | 0.002 | 1.76E-09 | 0.014 | 0.009 | 0.096 | 36 |
| rs1564347 | T | G | 0.34 | 0.009 | 0.002 | 2.39E-09 | 0.002 | 0.008 | 0.787 | 34.42 |
| rs1603460 | T | G | 0.42 | 0.009 | 0.001 | 2.20E-10 | -0.011 | 0.008 | 0.161 | 41.33 |
| rs162445 | A | G | 0.08 | 0.015 | 0.003 | 5.72E-09 | 0.002 | 0.014 | 0.889 | 34.18 |
| rs1689510 | C | G | 0.33 | 0.019 | 0.002 | 1.35E-38 | 0.01 | 0.008 | 0.202 | 167.27 |
| rs17048855 | A | G | 0.34 | 0.011 | 0.002 | 3.10E-13 | 0.003 | 0.008 | 0.71 | 51.84 |
| rs17110109 | T | C | 0.62 | -0.009 | 0.001 | 1.47E-10 | -0.013 | 0.008 | 0.097 | 44.13 |
| rs17144467 | A | C | 0.69 | 0.01 | 0.002 | 8.74E-11 | 0.005 | 0.008 | 0.561 | 42.68 |
| rs17148998 | A | G | 0.2 | 0.01 | 0.002 | 6.98E-09 | -0.003 | 0.009 | 0.736 | 35.3 |
| rs1717204 | A | C | 0.18 | -0.012 | 0.002 | 1.20E-10 | -0.013 | 0.01 | 0.189 | 42.25 |
| rs17248751 | A | G | 0.78 | -0.014 | 0.002 | 1.28E-15 | -0.001 | 0.009 | 0.87 | 64 |
| rs173003 | A | C | 0.49 | -0.008 | 0.001 | 5.50E-09 | 0.042 | 0.008 | 2.76E-08 | 34.31 |
| rs1738050 | C | G | 0.62 | -0.009 | 0.001 | 1.42E-10 | -0.009 | 0.008 | 0.253 | 44.13 |
| rs17411339 | A | G | 0.56 | 0.014 | 0.001 | 5.10E-23 | 0.007 | 0.008 | 0.383 | 100 |
| rs17428076 | C | G | 0.76 | 0.014 | 0.002 | 6.41E-17 | -0.005 | 0.009 | 0.545 | 73.32 |
| rs1747714 | T | C | 0.53 | 0.011 | 0.001 | 3.82E-14 | 0.015 | 0.008 | 0.049 | 57.33 |
| rs1747817 | T | C | 0.76 | -0.009 | 0.002 | 2.29E-08 | -0.009 | 0.009 | 0.311 | 33.06 |
| rs17489649 | A | G | 0.68 | 0.013 | 0.002 | 2.77E-17 | -0.002 | 0.008 | 0.784 | 71.68 |
| rs17502934 | T | G | 0.15 | -0.017 | 0.002 | 2.12E-17 | 0.005 | 0.011 | 0.646 | 69.72 |
| rs175325 | A | T | 0.6 | -0.01 | 0.001 | 2.24E-12 | 0.012 | 0.008 | 0.116 | 51.02 |
| rs17551064 | A | G | 0.84 | 0.014 | 0.002 | 1.53E-13 | 0 | 0.01 | 0.998 | 55.07 |
| rs17563464 | A | C | 0.22 | -0.015 | 0.002 | 1.74E-17 | 0.008 | 0.009 | 0.379 | 74.77 |
| rs17565975 | A | G | 0.56 | -0.011 | 0.001 | 5.54E-14 | -0.012 | 0.008 | 0.124 | 57.33 |
| rs17598675 | T | C | 0.51 | -0.012 | 0.001 | 1.37E-16 | -0.002 | 0.008 | 0.797 | 68.65 |
| rs17604349 | A | G | 0.19 | 0.013 | 0.002 | 2.36E-12 | 0.007 | 0.01 | 0.499 | 49.78 |
| rs176218 | T | G | 0.2 | 0.02 | 0.002 | 1.10E-29 | -0.008 | 0.01 | 0.379 | 123.46 |
| rs17732878 | T | C | 0.79 | -0.01 | 0.002 | 4.06E-08 | 0.008 | 0.009 | 0.409 | 31.23 |
| rs178183 | T | C | 0.25 | 0.015 | 0.002 | 4.81E-19 | -0.004 | 0.009 | 0.634 | 82.13 |
| rs17882802 | A | G | 0.43 | 0.008 | 0.001 | 3.00E-09 | -0.009 | 0.008 | 0.229 | 36 |
| rs182902112 | A | C | 0.02 | -0.03 | 0.005 | 1.17E-08 | 0.047 | 0.032 | 0.145 | 32.04 |
| rs1842713 | A | G | 0.79 | -0.012 | 0.002 | 9.67E-12 | 0.009 | 0.009 | 0.337 | 47.37 |
| rs1861786 | A | G | 0.38 | -0.008 | 0.001 | 2.39E-08 | -0.014 | 0.008 | 0.081 | 33.47 |
| rs1865955 | T | C | 0.82 | 0.012 | 0.002 | 4.06E-11 | 0.011 | 0.01 | 0.262 | 41.23 |
| rs1866823 | A | G | 0.54 | 0.011 | 0.001 | 3.09E-14 | -0.013 | 0.008 | 0.088 | 58.41 |
| rs1890132 | T | C | 0.71 | -0.009 | 0.002 | 6.56E-09 | -0.001 | 0.008 | 0.936 | 35.2 |
| rs192436652 | T | C | 0.03 | -0.032 | 0.004 | 2.06E-12 | 0.018 | 0.024 | 0.45 | 50.25 |
| rs1933264 | T | C | 0.75 | 0.01 | 0.002 | 5.85E-10 | -0.014 | 0.009 | 0.111 | 39.06 |
| rs1955250 | A | C | 0.91 | -0.014 | 0.002 | 2.26E-08 | 0.018 | 0.014 | 0.19 | 31.36 |
| rs1991585 | T | C | 0.71 | -0.01 | 0.002 | 6.26E-11 | -0.012 | 0.008 | 0.152 | 45.34 |
| rs2007655 | T | G | 0.5 | 0.009 | 0.001 | 8.64E-10 | 0.009 | 0.008 | 0.232 | 37.73 |
| rs2011603 | A | G | 0.74 | -0.009 | 0.002 | 1.12E-08 | 0.005 | 0.009 | 0.591 | 32.35 |
| rs2034631 | T | C | 0.65 | 0.009 | 0.002 | 2.64E-09 | 0.01 | 0.008 | 0.189 | 35.2 |
| rs2052285 | A | G | 0.6 | 0.011 | 0.001 | 7.25E-15 | 0.004 | 0.008 | 0.617 | 64 |
| rs2055940 | A | G | 0.32 | 0.008 | 0.002 | 4.95E-08 | 0.001 | 0.008 | 0.942 | 29.88 |
| rs2081652 | A | T | 0.66 | 0.012 | 0.002 | 1.09E-16 | 0.008 | 0.008 | 0.34 | 67.24 |
| rs2131167 | A | G | 0.45 | 0.008 | 0.001 | 1.52E-08 | 0.008 | 0.008 | 0.299 | 32.65 |
| rs2179152 | T | C | 0.37 | -0.013 | 0.002 | 1.68E-19 | -0.008 | 0.008 | 0.3 | 76.27 |
| rs2183271 | T | C | 0.64 | 0.008 | 0.002 | 1.91E-08 | -0.007 | 0.008 | 0.346 | 31.36 |
| rs2199409 | T | C | 0.8 | 0.01 | 0.002 | 3.14E-09 | 0.002 | 0.01 | 0.849 | 36.71 |
| rs2212430 | T | C | 0.3 | -0.01 | 0.002 | 1.32E-10 | -0.01 | 0.008 | 0.22 | 39.06 |
| rs2216144 | T | C | 0.53 | 0.01 | 0.001 | 2.58E-12 | -0.011 | 0.008 | 0.139 | 49 |
| rs2256965 | A | G | 0.42 | 0.011 | 0.001 | 2.40E-13 | 0.018 | 0.008 | 0.02 | 57.33 |
| rs2287838 | A | G | 0.55 | -0.01 | 0.001 | 1.27E-13 | 0.013 | 0.008 | 0.09 | 55.18 |
| rs2290601 | T | C | 0.77 | -0.015 | 0.002 | 4.49E-19 | 0.007 | 0.009 | 0.42 | 76.82 |
| rs2297293 | C | G | 0.31 | 0.01 | 0.002 | 8.80E-11 | 0.001 | 0.008 | 0.943 | 43.56 |
| rs232496 | T | C | 0.37 | 0.009 | 0.002 | 7.14E-10 | -0.003 | 0.008 | 0.7 | 36 |
| rs2364544 | A | G | 0.39 | -0.011 | 0.001 | 2.91E-13 | -0.01 | 0.008 | 0.189 | 56.25 |
| rs2434672 | A | C | 0.53 | 0.009 | 0.001 | 1.06E-10 | -0.002 | 0.008 | 0.788 | 42.25 |
| rs2436760 | T | C | 0.97 | 0.023 | 0.004 | 4.52E-08 | -0.031 | 0.022 | 0.172 | 30.51 |
| rs2447097 | T | G | 0.46 | 0.01 | 0.001 | 1.10E-11 | 0.012 | 0.008 | 0.107 | 47.02 |
| rs2469226 | A | T | 0.22 | 0.01 | 0.002 | 1.28E-08 | 0.004 | 0.009 | 0.628 | 31.89 |
| rs2496482 | T | C | 0.42 | 0.012 | 0.002 | 9.20E-17 | 0.005 | 0.008 | 0.498 | 65.07 |
| rs252991 | A | G | 0.37 | 0.01 | 0.002 | 6.30E-11 | -0.002 | 0.008 | 0.816 | 40.11 |
| rs2545795 | A | C | 0.44 | 0.013 | 0.001 | 1.03E-18 | 0.006 | 0.008 | 0.439 | 79.72 |
| rs2554835 | A | G | 0.4 | 0.009 | 0.001 | 8.95E-10 | -0.001 | 0.008 | 0.864 | 39.51 |
| rs2570497 | T | C | 0.64 | -0.012 | 0.002 | 1.27E-16 | 0.01 | 0.008 | 0.193 | 65.07 |
| rs2725370 | T | C | 0.3 | -0.014 | 0.002 | 4.96E-20 | -0.01 | 0.008 | 0.211 | 88.36 |
| rs2764684 | T | C | 0.83 | 0.014 | 0.002 | 9.99E-15 | 0.029 | 0.01 | 0.004 | 56.65 |
| rs2838006 | T | C | 0.36 | 0.013 | 0.002 | 1.38E-19 | -0.013 | 0.008 | 0.111 | 77.44 |
| rs28513882 | A | G | 0.18 | -0.011 | 0.002 | 2.03E-09 | -0.005 | 0.01 | 0.61 | 37.35 |
| rs2885198 | A | G | 0.53 | 0.009 | 0.001 | 4.63E-10 | -0.005 | 0.008 | 0.516 | 39.51 |
| rs2923424 | A | G | 0.61 | 0.012 | 0.001 | 6.31E-17 | 0.005 | 0.008 | 0.494 | 73.47 |
| rs2929032 | A | G | 0.47 | -0.008 | 0.001 | 8.40E-09 | -0.005 | 0.008 | 0.481 | 33.47 |
| rs2958182 | A | T | 0.34 | 0.008 | 0.002 | 3.46E-08 | 0.004 | 0.008 | 0.605 | 29.88 |
| rs2964199 | T | C | 0.31 | -0.01 | 0.002 | 4.61E-11 | 0.008 | 0.008 | 0.304 | 43.56 |
| rs2964255 | A | G | 0.31 | 0.009 | 0.002 | 6.34E-09 | -0.015 | 0.008 | 0.062 | 34.42 |
| rs2998299 | T | G | 0.21 | -0.014 | 0.002 | 1.51E-15 | -0.002 | 0.009 | 0.805 | 64 |
| rs3026996 | A | C | 0.76 | 0.014 | 0.002 | 5.42E-17 | -0.001 | 0.009 | 0.883 | 74.39 |
| rs303752 | A | G | 0.41 | -0.011 | 0.001 | 1.47E-14 | -0.001 | 0.008 | 0.899 | 61.73 |
| rs3111251 | T | C | 0.43 | -0.009 | 0.001 | 3.88E-10 | -0.001 | 0.008 | 0.862 | 41.33 |
| rs34067381 | T | G | 0.36 | -0.01 | 0.002 | 9.77E-13 | 0.003 | 0.008 | 0.662 | 48.07 |
| rs34098770 | A | G | 0.15 | -0.016 | 0.002 | 2.95E-16 | -0.004 | 0.011 | 0.695 | 64 |
| rs34305371 | A | G | 0.1 | 0.031 | 0.002 | 1.10E-39 | 0.013 | 0.013 | 0.313 | 171.17 |
| rs34394051 | A | G | 0.84 | -0.012 | 0.002 | 2.30E-09 | 0.01 | 0.01 | 0.333 | 34.22 |
| rs34720381 | T | C | 0.09 | -0.018 | 0.002 | 1.90E-13 | -0.016 | 0.013 | 0.225 | 56.25 |
| rs34807077 | A | C | 0.16 | 0.013 | 0.002 | 3.29E-11 | 0.01 | 0.01 | 0.314 | 45.39 |
| rs35417702 | T | C | 0.52 | -0.015 | 0.001 | 2.40E-27 | 0.001 | 0.008 | 0.907 | 117.88 |
| rs35606437 | A | G | 0.27 | 0.011 | 0.002 | 3.98E-11 | -0.004 | 0.009 | 0.653 | 43.07 |
| rs35929923 | A | G | 0.25 | -0.011 | 0.002 | 1.77E-12 | -0.018 | 0.009 | 0.041 | 50.77 |
| rs363096 | T | C | 0.42 | -0.014 | 0.001 | 5.77E-24 | 0.009 | 0.008 | 0.219 | 104.33 |
| rs3751331 | A | G | 0.38 | -0.01 | 0.001 | 1.75E-11 | -0.012 | 0.008 | 0.118 | 48.01 |
| rs3809634 | A | G | 0.68 | -0.011 | 0.002 | 6.82E-13 | -0.004 | 0.008 | 0.652 | 52.8 |
| rs382196 | T | G | 0.38 | -0.008 | 0.001 | 1.47E-08 | 0 | 0.008 | 0.951 | 34.31 |
| rs3859523 | T | C | 0.82 | 0.012 | 0.002 | 9.92E-12 | 0.002 | 0.01 | 0.829 | 47.46 |
| rs3897821 | A | G | 0.67 | 0.015 | 0.002 | 3.58E-25 | 0.007 | 0.008 | 0.379 | 105.4 |
| rs3948495 | T | G | 0.41 | -0.009 | 0.001 | 3.95E-10 | 0.009 | 0.008 | 0.241 | 40.41 |
| rs401526 | T | C | 0.53 | -0.01 | 0.001 | 1.15E-13 | 0.002 | 0.008 | 0.825 | 55.18 |
| rs401966 | C | G | 0.39 | -0.01 | 0.001 | 1.72E-11 | -0.019 | 0.008 | 0.018 | 48.01 |
| rs42210 | C | G | 0.71 | -0.01 | 0.002 | 2.74E-10 | 0.004 | 0.008 | 0.605 | 37.52 |
| rs42302 | A | G | 0.34 | 0.009 | 0.002 | 5.96E-09 | 0.012 | 0.008 | 0.129 | 32.87 |
| rs4298514 | T | C | 0.7 | 0.01 | 0.002 | 1.07E-11 | -0.006 | 0.008 | 0.432 | 48.07 |
| rs4320563 | A | G | 0.51 | 0.011 | 0.001 | 2.24E-14 | -0.001 | 0.008 | 0.874 | 60.62 |
| rs4328757 | T | C | 0.61 | 0.01 | 0.001 | 2.95E-11 | 0.016 | 0.008 | 0.044 | 47.02 |
| rs4352658 | T | C | 0.08 | -0.019 | 0.003 | 1.71E-13 | 0.004 | 0.014 | 0.798 | 53.4 |
| rs4358081 | A | C | 0.53 | -0.01 | 0.001 | 1.28E-11 | -0.017 | 0.008 | 0.029 | 46.05 |
| rs4369924 | A | G | 0.16 | 0.013 | 0.002 | 5.58E-11 | -0.011 | 0.01 | 0.289 | 43.28 |
| rs4384309 | A | G | 0.46 | 0.01 | 0.001 | 4.89E-13 | 0.009 | 0.008 | 0.26 | 53.08 |
| rs4467547 | T | G | 0.41 | 0.012 | 0.001 | 4.88E-17 | -0.009 | 0.008 | 0.224 | 74.7 |
| rs4500930 | T | C | 0.34 | -0.011 | 0.002 | 1.67E-13 | 0.003 | 0.008 | 0.685 | 52.8 |
| rs4663617 | A | T | 0.23 | 0.01 | 0.002 | 4.12E-10 | 0.009 | 0.009 | 0.317 | 37.43 |
| rs4673840 | T | C | 0.84 | -0.014 | 0.002 | 5.86E-13 | 0.012 | 0.01 | 0.238 | 52.75 |
| rs4675248 | A | G | 0.4 | -0.009 | 0.001 | 4.50E-11 | 0.006 | 0.008 | 0.458 | 45.08 |
| rs4719944 | T | C | 0.55 | -0.012 | 0.001 | 9.25E-18 | -0.008 | 0.008 | 0.314 | 74.7 |
| rs4726070 | A | G | 0.6 | 0.012 | 0.001 | 1.36E-17 | 0.016 | 0.008 | 0.037 | 75.94 |
| rs4757957 | C | G | 0.69 | 0.013 | 0.002 | 1.79E-18 | 0 | 0.008 | 0.978 | 78.62 |
| rs4766424 | C | G | 0.12 | 0.014 | 0.002 | 2.47E-11 | -0.002 | 0.011 | 0.884 | 45.72 |
| rs4778058 | T | C | 0.5 | -0.009 | 0.001 | 4.92E-11 | 0 | 0.008 | 0.991 | 43.18 |
| rs4787457 | A | G | 0.63 | 0.016 | 0.002 | 1.23E-28 | -0.019 | 0.008 | 0.015 | 116.64 |
| rs4793090 | A | G | 0.66 | 0.008 | 0.002 | 1.67E-08 | 0 | 0.008 | 0.953 | 31.36 |
| rs4810894 | A | G | 0.37 | -0.009 | 0.002 | 4.38E-09 | 0.003 | 0.008 | 0.69 | 32.11 |
| rs4839155 | T | G | 0.77 | 0.012 | 0.002 | 4.66E-12 | -0.005 | 0.009 | 0.569 | 45.76 |
| rs4846724 | A | G | 0.53 | 0.01 | 0.001 | 2.89E-12 | 0.004 | 0.008 | 0.564 | 49 |
| rs4848924 | A | C | 0.71 | -0.012 | 0.002 | 3.56E-14 | -0.013 | 0.008 | 0.115 | 60.84 |
| rs488476 | C | G | 0.62 | -0.011 | 0.002 | 8.29E-15 | 0.007 | 0.008 | 0.342 | 56.75 |
| rs4895650 | T | C | 0.55 | 0.009 | 0.001 | 5.31E-11 | 0.005 | 0.008 | 0.479 | 44.13 |
| rs4899012 | C | G | 0.6 | -0.01 | 0.001 | 8.06E-13 | 0.003 | 0.008 | 0.706 | 54.13 |
| rs4904523 | A | G | 0.52 | -0.008 | 0.001 | 6.27E-09 | 0.005 | 0.008 | 0.492 | 34.31 |
| rs4915735 | A | G | 0.14 | 0.012 | 0.002 | 9.56E-09 | 0.016 | 0.011 | 0.127 | 33.64 |
| rs4977885 | A | G | 0.4 | -0.008 | 0.001 | 6.78E-09 | -0.008 | 0.008 | 0.327 | 36 |
| rs4984541 | A | G | 0.77 | -0.014 | 0.002 | 3.40E-16 | -0.02 | 0.009 | 0.026 | 65.9 |
| rs55736314 | C | G | 0.6 | -0.015 | 0.001 | 4.59E-27 | -0.01 | 0.008 | 0.191 | 121 |
| rs56099375 | T | C | 0.24 | 0.012 | 0.002 | 3.10E-13 | 0.003 | 0.009 | 0.744 | 49.83 |
| rs56319902 | T | C | 0.22 | -0.021 | 0.002 | 5.81E-33 | -0.072 | 0.009 | 2.82E-15 | 146.84 |
| rs56391344 | A | G | 0.25 | 0.015 | 0.002 | 7.49E-20 | 0.017 | 0.009 | 0.052 | 85.56 |
| rs56794817 | A | G | 0.16 | 0.012 | 0.002 | 1.69E-10 | -0.012 | 0.01 | 0.261 | 41.23 |
| rs57349798 | A | G | 0.41 | 0.009 | 0.001 | 8.63E-11 | 0.005 | 0.008 | 0.558 | 44.13 |
| rs57352738 | A | T | 0.2 | -0.015 | 0.002 | 2.51E-18 | -0.02 | 0.009 | 0.036 | 79.94 |
| rs575113 | A | G | 0.29 | 0.013 | 0.002 | 7.52E-18 | 0.008 | 0.008 | 0.37 | 78.62 |
| rs5754753 | T | C | 0.72 | -0.013 | 0.002 | 6.15E-16 | -0.011 | 0.008 | 0.183 | 62.02 |
| rs57661533 | T | C | 0.13 | -0.011 | 0.002 | 3.24E-08 | -0.025 | 0.011 | 0.029 | 29.47 |
| rs580652 | T | C | 0.9 | -0.015 | 0.002 | 4.65E-10 | -0.01 | 0.013 | 0.435 | 37.52 |
| rs590013 | T | C | 0.68 | 0.01 | 0.002 | 2.47E-11 | 0.012 | 0.008 | 0.154 | 44.44 |
| rs6043521 | T | C | 0.6 | -0.008 | 0.001 | 9.72E-09 | -0.007 | 0.008 | 0.395 | 35.15 |
| rs6065080 | T | C | 0.36 | -0.013 | 0.002 | 7.49E-19 | 0.005 | 0.008 | 0.567 | 75.11 |
| rs6065784 | C | G | 0.7 | 0.011 | 0.002 | 5.78E-13 | -0.01 | 0.008 | 0.25 | 53.78 |
| rs61104616 | A | G | 0.52 | -0.017 | 0.001 | 1.56E-34 | 0.003 | 0.008 | 0.709 | 150.94 |
| rs6123924 | A | G | 0.84 | 0.013 | 0.002 | 3.20E-11 | -0.011 | 0.011 | 0.285 | 46.1 |
| rs61527214 | A | G | 0.41 | 0.011 | 0.001 | 1.13E-14 | -0.006 | 0.008 | 0.46 | 61.73 |
| rs61997667 | T | C | 0.15 | -0.014 | 0.002 | 2.24E-13 | -0.007 | 0.01 | 0.518 | 51.84 |
| rs62155350 | A | G | 0.02 | 0.034 | 0.005 | 1.69E-10 | 0.004 | 0.027 | 0.88 | 40.67 |
| rs62155873 | T | C | 0.12 | -0.015 | 0.002 | 1.65E-12 | -0.018 | 0.011 | 0.113 | 51.7 |
| rs62174974 | A | G | 0.19 | -0.011 | 0.002 | 2.76E-09 | 0.004 | 0.01 | 0.675 | 34.03 |
| rs62179650 | A | G | 0.3 | 0.011 | 0.002 | 5.66E-13 | 0.003 | 0.008 | 0.733 | 49 |
| rs62182994 | T | C | 0.69 | 0.014 | 0.002 | 2.55E-19 | 0.009 | 0.008 | 0.278 | 83.42 |
| rs62379838 | T | C | 0.7 | 0.012 | 0.002 | 1.77E-15 | 0.01 | 0.008 | 0.212 | 65.07 |
| rs628993 | A | G | 0.11 | 0.013 | 0.002 | 3.83E-08 | 0.005 | 0.013 | 0.702 | 30.97 |
| rs6449503 | A | G | 0.5 | 0.018 | 0.001 | 1.84E-39 | -0.009 | 0.008 | 0.217 | 174.62 |
| rs6490618 | T | C | 0.33 | 0.009 | 0.002 | 3.48E-09 | 0 | 0.008 | 0.974 | 34.42 |
| rs6493265 | T | C | 0.39 | -0.012 | 0.001 | 2.26E-17 | 0.003 | 0.008 | 0.741 | 75.94 |
| rs6557171 | T | C | 0.32 | -0.016 | 0.002 | 5.48E-25 | -0.001 | 0.008 | 0.876 | 106.78 |
| rs660001 | A | G | 0.21 | -0.016 | 0.002 | 1.66E-21 | 0.001 | 0.009 | 0.906 | 93.07 |
| rs66568921 | T | G | 0.64 | -0.018 | 0.002 | 7.60E-34 | -0.003 | 0.008 | 0.675 | 145.6 |
| rs66721975 | A | G | 0.29 | -0.009 | 0.002 | 1.51E-08 | -0.014 | 0.008 | 0.097 | 33.64 |
| rs6690195 | T | C | 0.49 | -0.011 | 0.001 | 7.89E-16 | 0.001 | 0.008 | 0.862 | 65.15 |
| rs6697584 | T | C | 0.79 | -0.013 | 0.002 | 1.43E-13 | -0.01 | 0.009 | 0.282 | 54.93 |
| rs6704768 | A | G | 0.56 | -0.012 | 0.001 | 1.84E-16 | 0.003 | 0.008 | 0.679 | 68.65 |
| rs6731373 | A | G | 0.34 | -0.012 | 0.002 | 1.39E-14 | 0.017 | 0.008 | 0.035 | 58.78 |
| rs6812533 | T | C | 0.74 | 0.011 | 0.002 | 4.85E-11 | 0.003 | 0.009 | 0.697 | 43.89 |
| rs68145588 | T | G | 0.14 | -0.013 | 0.002 | 7.53E-10 | -0.002 | 0.011 | 0.845 | 36.57 |
| rs6917154 | T | C | 0.12 | 0.013 | 0.002 | 2.55E-09 | -0.007 | 0.011 | 0.539 | 36.57 |
| rs6924023 | A | G | 0.75 | 0.011 | 0.002 | 5.17E-11 | 0.004 | 0.009 | 0.683 | 44.72 |
| rs6977237 | T | C | 0.47 | 0.008 | 0.001 | 5.59E-09 | -0.013 | 0.008 | 0.096 | 34.31 |
| rs6994287 | A | G | 0.41 | 0.01 | 0.001 | 1.11E-12 | 0.005 | 0.008 | 0.538 | 53.08 |
| rs7012546 | T | C | 0.42 | 0.009 | 0.001 | 1.25E-10 | 0.001 | 0.008 | 0.884 | 43.18 |
| rs7029718 | A | G | 0.41 | 0.024 | 0.001 | 7.82E-65 | 0.001 | 0.008 | 0.935 | 303.76 |
| rs7041702 | A | G | 0.74 | -0.011 | 0.002 | 4.53E-12 | 0.013 | 0.009 | 0.124 | 48.13 |
| rs7167688 | T | C | 0.49 | -0.009 | 0.001 | 1.71E-11 | -0.012 | 0.008 | 0.121 | 45.08 |
| rs7171405 | A | G | 0.24 | -0.01 | 0.002 | 7.53E-10 | -0.009 | 0.009 | 0.313 | 39.85 |
| rs717996 | T | C | 0.6 | -0.012 | 0.001 | 1.89E-17 | 0.01 | 0.008 | 0.218 | 75.94 |
| rs7226824 | T | C | 0.53 | -0.008 | 0.001 | 2.72E-08 | -0.004 | 0.008 | 0.63 | 31.04 |
| rs72486027 | T | C | 0.25 | -0.01 | 0.002 | 1.61E-10 | -0.009 | 0.009 | 0.31 | 42.25 |
| rs7254263 | T | C | 0.28 | -0.012 | 0.002 | 1.84E-14 | -0.023 | 0.008 | 0.007 | 55.32 |
| rs72622559 | T | C | 0.23 | -0.012 | 0.002 | 5.50E-13 | 0.001 | 0.009 | 0.934 | 49.83 |
| rs72672052 | A | T | 0.18 | -0.01 | 0.002 | 4.87E-08 | -0.001 | 0.01 | 0.954 | 31.48 |
| rs72709560 | A | G | 0.69 | 0.009 | 0.002 | 2.03E-08 | -0.003 | 0.008 | 0.686 | 32.11 |
| rs72828517 | T | C | 0.83 | -0.017 | 0.002 | 1.38E-19 | 0.002 | 0.01 | 0.841 | 78.18 |
| rs72829857 | A | G | 0.76 | -0.015 | 0.002 | 1.52E-19 | -0.003 | 0.009 | 0.775 | 86.72 |
| rs72917504 | T | C | 0.06 | -0.018 | 0.003 | 2.91E-09 | 0.027 | 0.016 | 0.093 | 35.2 |
| rs72944064 | T | C | 0.74 | -0.01 | 0.002 | 9.39E-10 | 0.002 | 0.009 | 0.841 | 37.52 |
| rs72962169 | T | C | 0.16 | -0.017 | 0.002 | 1.26E-19 | 0.007 | 0.011 | 0.528 | 83.87 |
| rs72993796 | T | C | 0.88 | 0.013 | 0.002 | 1.12E-08 | -0.017 | 0.012 | 0.153 | 33.32 |
| rs730384 | A | G | 0.44 | 0.01 | 0.001 | 9.38E-13 | 0.016 | 0.008 | 0.038 | 52.05 |
| rs73055556 | A | G | 0.14 | 0.011 | 0.002 | 4.95E-08 | 0.019 | 0.011 | 0.082 | 30.8 |
| rs73191311 | A | G | 0.34 | -0.009 | 0.002 | 2.03E-10 | 0.003 | 0.008 | 0.745 | 39.27 |
| rs7321274 | A | G | 0.8 | 0.011 | 0.002 | 8.21E-11 | 0.009 | 0.009 | 0.335 | 40.11 |
| rs7326331 | A | G | 0.28 | -0.013 | 0.002 | 3.42E-16 | -0.001 | 0.009 | 0.905 | 64 |
| rs73344830 | A | G | 0.42 | 0.017 | 0.001 | 1.00E-32 | 0.001 | 0.008 | 0.883 | 147.45 |
| rs736281 | T | C | 0.39 | 0.009 | 0.001 | 2.49E-09 | 0.007 | 0.008 | 0.375 | 37.73 |
| rs73648455 | T | C | 0.08 | -0.018 | 0.003 | 1.99E-12 | 0.011 | 0.014 | 0.437 | 49.54 |
| rs743316 | T | C | 0.79 | 0.011 | 0.002 | 8.21E-11 | 0.002 | 0.009 | 0.864 | 43.4 |
| rs74415461 | T | C | 0.08 | 0.018 | 0.003 | 6.25E-12 | -0.015 | 0.013 | 0.252 | 45.82 |
| rs7449561 | A | G | 0.23 | 0.01 | 0.002 | 9.73E-10 | 0.013 | 0.009 | 0.151 | 36 |
| rs7460106 | T | C | 0.76 | -0.012 | 0.002 | 2.06E-13 | -0.011 | 0.009 | 0.22 | 52.35 |
| rs746839 | C | G | 0.63 | 0.014 | 0.002 | 1.19E-20 | 0.005 | 0.008 | 0.539 | 81 |
| rs74787922 | A | C | 0.93 | 0.018 | 0.003 | 8.97E-11 | -0.013 | 0.015 | 0.383 | 42.25 |
| rs75033012 | C | G | 0.04 | -0.026 | 0.004 | 1.38E-11 | 0.018 | 0.019 | 0.35 | 46.1 |
| rs75177132 | T | C | 0.05 | 0.03 | 0.004 | 1.01E-16 | 0.007 | 0.018 | 0.699 | 68.52 |
| rs7575637 | A | G | 0.45 | 0.011 | 0.001 | 3.43E-14 | -0.006 | 0.008 | 0.416 | 58.41 |
| rs7603132 | A | G | 0.19 | 0.014 | 0.002 | 1.20E-14 | 0.004 | 0.009 | 0.675 | 57.93 |
| rs76076331 | T | C | 0.13 | 0.021 | 0.002 | 1.71E-23 | 0.008 | 0.012 | 0.481 | 96.23 |
| rs7617204 | A | G | 0.52 | -0.01 | 0.001 | 4.33E-12 | 0.029 | 0.008 | 1.14E-04 | 48.01 |
| rs76235882 | A | G | 0.97 | 0.024 | 0.004 | 4.52E-10 | 0.026 | 0.023 | 0.252 | 38.19 |
| rs76246107 | A | G | 0.09 | -0.016 | 0.003 | 6.52E-10 | 0.037 | 0.015 | 0.011 | 38.82 |
| rs7650602 | T | C | 0.56 | -0.009 | 0.001 | 2.77E-10 | -0.002 | 0.008 | 0.806 | 40.41 |
| rs76577427 | C | G | 0.9 | 0.02 | 0.002 | 7.78E-18 | 0.005 | 0.012 | 0.701 | 71.54 |
| rs7672622 | A | G | 0.75 | -0.009 | 0.002 | 4.72E-08 | 0 | 0.009 | 0.965 | 30.25 |
| rs7683416 | T | C | 0.46 | 0.014 | 0.001 | 6.05E-23 | 0.008 | 0.008 | 0.3 | 98.58 |
| rs76878669 | C | G | 0.76 | 0.014 | 0.002 | 3.10E-17 | 0.008 | 0.009 | 0.4 | 68.79 |
| rs7692359 | T | C | 0.78 | -0.011 | 0.002 | 1.40E-10 | 0.012 | 0.009 | 0.212 | 41.11 |
| rs77025239 | A | G | 0.16 | -0.014 | 0.002 | 4.65E-13 | 0.022 | 0.011 | 0.041 | 54.29 |
| rs77609760 | A | G | 0.93 | 0.024 | 0.003 | 7.71E-17 | -0.007 | 0.015 | 0.666 | 71.04 |
| rs77719387 | A | T | 0.02 | -0.04 | 0.006 | 3.78E-12 | 0.014 | 0.032 | 0.654 | 48.28 |
| rs7849487 | T | G | 0.66 | -0.017 | 0.002 | 1.25E-29 | -0.003 | 0.008 | 0.737 | 123.95 |
| rs78648104 | T | C | 0.91 | -0.014 | 0.002 | 1.70E-08 | -0.025 | 0.013 | 0.069 | 31.81 |
| rs7875078 | A | C | 0.46 | -0.008 | 0.001 | 2.72E-08 | -0.002 | 0.008 | 0.754 | 31.04 |
| rs7894722 | T | C | 0.37 | 0.009 | 0.001 | 5.41E-09 | 0.005 | 0.008 | 0.559 | 36.86 |
| rs790647 | A | C | 0.23 | -0.015 | 0.002 | 3.81E-18 | -0.012 | 0.009 | 0.166 | 72.75 |
| rs7910403 | T | G | 0.2 | 0.012 | 0.002 | 5.08E-12 | 0.001 | 0.009 | 0.895 | 45.94 |
| rs7924036 | T | G | 0.51 | 0.013 | 0.001 | 2.55E-21 | 0.013 | 0.008 | 0.08 | 90.25 |
| rs79265434 | A | G | 0.88 | -0.02 | 0.002 | 1.98E-19 | -0.016 | 0.012 | 0.164 | 80.18 |
| rs7972246 | T | C | 0.35 | 0.011 | 0.002 | 1.01E-13 | -0.011 | 0.008 | 0.154 | 53.78 |
| rs79728014 | A | G | 0.86 | -0.013 | 0.002 | 2.06E-11 | 0.005 | 0.011 | 0.661 | 44.89 |
| rs7974852 | A | C | 0.52 | 0.012 | 0.001 | 1.14E-16 | 0.021 | 0.008 | 0.007 | 69.84 |
| rs7977614 | A | G | 0.71 | -0.013 | 0.002 | 6.84E-17 | -0.007 | 0.008 | 0.405 | 70.14 |
| rs79994730 | T | C | 0.04 | 0.024 | 0.004 | 5.88E-12 | 0.016 | 0.017 | 0.37 | 47.02 |
| rs8008382 | T | C | 0.3 | -0.011 | 0.002 | 4.07E-12 | 0.006 | 0.008 | 0.469 | 49.94 |
| rs8024 | A | C | 0.33 | -0.012 | 0.002 | 1.13E-14 | 0.002 | 0.008 | 0.779 | 58.78 |
| rs80257979 | T | G | 0.97 | -0.025 | 0.004 | 3.31E-10 | -0.004 | 0.022 | 0.841 | 39.69 |
| rs8030487 | A | G | 0.69 | 0.009 | 0.002 | 2.52E-08 | 0.008 | 0.008 | 0.32 | 32.11 |
| rs806816 | A | T | 0.75 | 0.01 | 0.002 | 4.80E-10 | 0.006 | 0.009 | 0.523 | 39.85 |
| rs8097125 | T | C | 0.38 | 0.009 | 0.001 | 2.31E-09 | 0.002 | 0.008 | 0.747 | 37.73 |
| rs912883 | T | C | 0.68 | 0.009 | 0.002 | 6.20E-09 | 0.004 | 0.008 | 0.587 | 33.64 |
| rs9289300 | T | C | 0.84 | -0.016 | 0.002 | 1.00E-17 | 0.003 | 0.01 | 0.762 | 74.5 |
| rs929511 | T | C | 0.13 | -0.016 | 0.002 | 3.91E-14 | -0.025 | 0.012 | 0.031 | 58.05 |
| rs9359939 | A | C | 0.24 | -0.011 | 0.002 | 1.51E-10 | -0.006 | 0.009 | 0.532 | 43.07 |
| rs936496 | A | G | 0.62 | -0.008 | 0.001 | 4.38E-08 | -0.01 | 0.008 | 0.213 | 31.84 |
| rs9373363 | A | G | 0.75 | -0.011 | 0.002 | 5.54E-12 | -0.009 | 0.009 | 0.286 | 48.13 |
| rs9375188 | T | C | 0.48 | 0.021 | 0.001 | 8.78E-52 | 0 | 0.008 | 0.963 | 229.31 |
| rs9411331 | A | C | 0.68 | 0.015 | 0.002 | 7.63E-22 | -0.002 | 0.008 | 0.762 | 93.44 |
| rs9492774 | C | G | 0.37 | 0.008 | 0.002 | 4.33E-08 | 0.002 | 0.008 | 0.841 | 28.44 |
| rs9529146 | T | C | 0.23 | 0.013 | 0.002 | 3.48E-14 | 0.023 | 0.009 | 0.009 | 55.81 |
| rs9556958 | T | C | 0.52 | -0.011 | 0.001 | 5.96E-15 | -0.004 | 0.008 | 0.572 | 61.73 |
| rs9616947 | T | C | 0.38 | -0.013 | 0.002 | 3.58E-20 | -0.009 | 0.008 | 0.238 | 79.8 |
| rs9633970 | T | C | 0.76 | 0.009 | 0.002 | 2.81E-08 | 0.027 | 0.009 | 0.003 | 32.35 |
| rs9655780 | A | G | 0.83 | -0.015 | 0.002 | 3.70E-16 | -0.001 | 0.01 | 0.917 | 64.84 |
| rs969512 | A | T | 0.66 | -0.011 | 0.002 | 9.84E-13 | 0.009 | 0.008 | 0.261 | 49.94 |
| rs9853928 | T | C | 0.21 | -0.013 | 0.002 | 1.59E-13 | 0.004 | 0.009 | 0.675 | 55.81 |
| rs9859556 | T | G | 0.31 | 0.029 | 0.002 | 4.61E-82 | -0.01 | 0.008 | 0.223 | 373.78 |
| rs9882532 | T | C | 0.64 | 0.012 | 0.002 | 2.15E-17 | -0.003 | 0.008 | 0.702 | 68.34 |
| rs9886703 | A | T | 0.17 | -0.014 | 0.002 | 4.46E-13 | 0.019 | 0.01 | 0.06 | 51.24 |
| rs9927137 | A | G | 0.53 | 0.008 | 0.001 | 5.11E-09 | 0.006 | 0.008 | 0.443 | 34.31 |
| rs9927842 | T | C | 0.15 | -0.013 | 0.002 | 6.22E-11 | -0.013 | 0.01 | 0.23 | 42.25 |
| rs9929556 | T | G | 0.57 | -0.01 | 0.001 | 2.31E-12 | 0.004 | 0.008 | 0.62 | 50.01 |
| rs9929762 | A | G | 0.56 | 0.01 | 0.001 | 1.03E-11 | -0.013 | 0.008 | 0.082 | 47.02 |
| rs9933256 | A | G | 0.56 | 0.012 | 0.001 | 6.06E-17 | 0.012 | 0.008 | 0.116 | 72.25 |

**SNPs used as instrumental variables in the MR analysis of educational attainment on fractional anisotropy (white matter tracts)**

| **SNP** | **Effect allele** | **Other allele** | **Effect allele frequency** | **beta (exposure)** | **SE (exposure)** | ***P* (exposure)** | **beta (outcome)** | **SE (outcome)** | ***P* (outcome)** | ***F*-statistic** |
| --- | --- | --- | --- | --- | --- | --- | --- | --- | --- | --- |
| rs10006235 | T | C | 0.27 | -0.009 | 0.002 | 4.48E-09 | 0.008 | 0.008 | 0.304 | 33.79 |
| rs10021733 | T | C | 0.15 | -0.016 | 0.003 | 9.45E-10 | -0.007 | 0.01 | 0.455 | 36.89 |
| rs10046069 | A | G | 0.12 | 0.013 | 0.002 | 4.40E-09 | 0.011 | 0.011 | 0.322 | 33.32 |
| rs10060023 | T | C | 0.33 | 0.011 | 0.002 | 1.88E-13 | 0.002 | 0.008 | 0.798 | 53.78 |
| rs10080647 | A | C | 0.14 | 0.012 | 0.002 | 7.05E-10 | -0.006 | 0.01 | 0.599 | 38.44 |
| rs1008078 | T | C | 0.4 | -0.018 | 0.001 | 4.50E-37 | -0.014 | 0.008 | 0.066 | 169 |
| rs10189857 | A | G | 0.57 | 0.016 | 0.001 | 4.69E-29 | -0.007 | 0.007 | 0.368 | 127.37 |
| rs10193498 | A | T | 0.76 | 0.01 | 0.002 | 3.10E-09 | -0.002 | 0.009 | 0.778 | 36.75 |
| rs10215082 | A | G | 0.47 | -0.015 | 0.001 | 7.03E-26 | -0.017 | 0.007 | 0.017 | 113.27 |
| rs10460095 | A | G | 0.57 | -0.012 | 0.001 | 2.61E-18 | -0.019 | 0.007 | 0.009 | 78.45 |
| rs10761251 | A | T | 0.66 | 0.011 | 0.002 | 2.52E-14 | 0.006 | 0.008 | 0.438 | 56.75 |
| rs10772644 | C | G | 0.89 | 0.014 | 0.002 | 1.69E-10 | 0.027 | 0.012 | 0.02 | 41.66 |
| rs10773002 | A | T | 0.25 | 0.018 | 0.002 | 2.50E-30 | 0.004 | 0.008 | 0.595 | 133.69 |
| rs10773208 | T | C | 0.25 | -0.011 | 0.002 | 8.09E-12 | -0.004 | 0.008 | 0.65 | 47.27 |
| rs10798888 | T | G | 0.17 | -0.014 | 0.002 | 5.15E-14 | -0.006 | 0.01 | 0.548 | 54.29 |
| rs10799615 | A | G | 0.25 | -0.009 | 0.002 | 4.11E-08 | 0.013 | 0.008 | 0.115 | 30.94 |
| rs10805383 | A | G | 0.49 | -0.01 | 0.001 | 2.22E-13 | -0.007 | 0.007 | 0.314 | 54.13 |
| rs10830858 | T | C | 0.51 | 0.009 | 0.001 | 5.31E-11 | -0.006 | 0.007 | 0.379 | 43.18 |
| rs10844179 | A | G | 0.23 | 0.01 | 0.002 | 5.03E-10 | 0.014 | 0.009 | 0.107 | 37.43 |
| rs10875121 | C | G | 0.84 | 0.017 | 0.002 | 4.46E-20 | -0.019 | 0.01 | 0.06 | 83.87 |
| rs10892807 | T | C | 0.57 | 0.011 | 0.001 | 1.09E-14 | 0.005 | 0.007 | 0.461 | 60.62 |
| rs10940921 | T | G | 0.42 | 0.011 | 0.001 | 1.31E-14 | -0.004 | 0.007 | 0.552 | 62.86 |
| rs10951590 | T | C | 0.33 | -0.011 | 0.002 | 3.62E-13 | -0.003 | 0.008 | 0.655 | 52.8 |
| rs10979613 | T | C | 0.64 | -0.012 | 0.002 | 1.25E-15 | -0.025 | 0.008 | 0.001 | 60.84 |
| rs10996167 | C | G | 0.65 | 0.011 | 0.002 | 7.47E-09 | 0.001 | 0.008 | 0.869 | 31.92 |
| rs11003463 | T | G | 0.61 | 0.009 | 0.002 | 5.58E-10 | 0.017 | 0.007 | 0.022 | 36.8 |
| rs11023764 | A | G | 0.64 | 0.009 | 0.002 | 5.32E-10 | -0.003 | 0.008 | 0.677 | 36.8 |
| rs11030102 | C | G | 0.75 | 0.01 | 0.002 | 4.60E-10 | -0.023 | 0.008 | 0.006 | 39.85 |
| rs11076962 | T | C | 0.72 | 0.009 | 0.002 | 4.29E-08 | 0.001 | 0.008 | 0.949 | 28.89 |
| rs11081529 | T | C | 0.71 | 0.013 | 0.002 | 5.22E-16 | 0.004 | 0.008 | 0.622 | 70.56 |
| rs11082011 | T | C | 0.67 | 0.019 | 0.002 | 2.06E-38 | 0.016 | 0.008 | 0.035 | 167.27 |
| rs11121177 | A | G | 0.18 | 0.015 | 0.002 | 1.44E-16 | -0.013 | 0.01 | 0.187 | 70.37 |
| rs11130380 | T | G | 0.61 | 0.009 | 0.001 | 4.17E-10 | 0.002 | 0.007 | 0.738 | 41.33 |
| rs11138947 | T | C | 0.72 | 0.011 | 0.002 | 2.36E-11 | -0.002 | 0.008 | 0.76 | 43.07 |
| rs11157931 | A | C | 0.39 | -0.014 | 0.001 | 3.86E-22 | -0.011 | 0.007 | 0.132 | 98.58 |
| rs111852224 | T | C | 0.12 | 0.016 | 0.002 | 4.85E-13 | 0.007 | 0.011 | 0.531 | 50.28 |
| rs11211123 | A | G | 0.22 | 0.01 | 0.002 | 7.62E-10 | 0.007 | 0.009 | 0.391 | 37.43 |
| rs11213482 | A | G | 0.84 | -0.011 | 0.002 | 2.68E-08 | 0.016 | 0.01 | 0.087 | 31.12 |
| rs112603734 | A | C | 0.77 | -0.013 | 0.002 | 2.72E-09 | 0.003 | 0.009 | 0.724 | 35.46 |
| rs112806496 | C | G | 0.91 | -0.016 | 0.002 | 1.72E-10 | 0.025 | 0.013 | 0.049 | 40.96 |
| rs1128956 | T | G | 0.83 | -0.014 | 0.002 | 5.34E-14 | 0.004 | 0.009 | 0.638 | 54.29 |
| rs113520408 | A | G | 0.28 | 0.016 | 0.002 | 1.03E-22 | -0.004 | 0.008 | 0.594 | 93.85 |
| rs113615161 | T | C | 0.13 | -0.012 | 0.002 | 2.98E-09 | -0.011 | 0.011 | 0.297 | 34.31 |
| rs114593137 | A | T | 0.79 | -0.012 | 0.002 | 1.65E-12 | 0.003 | 0.009 | 0.743 | 51.5 |
| rs115000530 | A | T | 0.95 | -0.025 | 0.003 | 1.09E-15 | -0.016 | 0.016 | 0.322 | 65.04 |
| rs11542663 | A | C | 0.69 | 0.011 | 0.002 | 2.44E-13 | 0.015 | 0.008 | 0.054 | 55.75 |
| rs11588857 | A | G | 0.21 | 0.02 | 0.002 | 4.45E-31 | -0.015 | 0.009 | 0.086 | 137.03 |
| rs11598765 | A | G | 0.2 | -0.011 | 0.002 | 1.73E-09 | 0.005 | 0.009 | 0.607 | 34.68 |
| rs11620355 | A | G | 0.09 | 0.018 | 0.002 | 1.09E-13 | -0.021 | 0.013 | 0.099 | 53 |
| rs11657342 | A | G | 0.37 | 0.014 | 0.002 | 2.32E-13 | 0.011 | 0.007 | 0.134 | 54.29 |
| rs11657979 | A | G | 0.24 | -0.009 | 0.002 | 4.57E-08 | -0.002 | 0.009 | 0.81 | 28.65 |
| rs11663602 | A | C | 0.28 | -0.013 | 0.002 | 9.27E-17 | 0.013 | 0.008 | 0.112 | 66.02 |
| rs1167827 | A | G | 0.43 | 0.01 | 0.001 | 2.93E-13 | -0.008 | 0.007 | 0.303 | 55.18 |
| rs11724690 | T | G | 0.29 | 0.009 | 0.002 | 2.08E-09 | 0 | 0.008 | 0.964 | 38.44 |
| rs117398064 | C | G | 0.09 | -0.014 | 0.002 | 1.16E-08 | -0.008 | 0.012 | 0.525 | 31.81 |
| rs118134876 | T | C | 0.06 | -0.022 | 0.003 | 1.50E-13 | -0.004 | 0.015 | 0.809 | 55.25 |
| rs11871429 | A | G | 0.77 | 0.013 | 0.002 | 2.52E-14 | 0.001 | 0.009 | 0.904 | 56.69 |
| rs11894424 | A | C | 0.95 | 0.021 | 0.003 | 2.14E-10 | -0.004 | 0.018 | 0.833 | 40.11 |
| rs12054166 | C | G | 0.74 | 0.01 | 0.002 | 2.30E-09 | -0.002 | 0.008 | 0.771 | 36 |
| rs12076635 | C | G | 0.78 | 0.021 | 0.002 | 3.33E-34 | 0.032 | 0.009 | 2.90E-04 | 148.27 |
| rs12113634 | T | C | 0.65 | 0.009 | 0.002 | 2.50E-09 | 0.001 | 0.008 | 0.891 | 34.42 |
| rs12151248 | T | C | 0.11 | -0.015 | 0.002 | 3.81E-11 | 0.006 | 0.011 | 0.607 | 41.97 |
| rs1220779 | A | G | 0.47 | -0.008 | 0.001 | 1.58E-08 | 0.011 | 0.007 | 0.118 | 31.84 |
| rs12273435 | A | G | 0.21 | -0.013 | 0.002 | 4.72E-14 | 0.004 | 0.009 | 0.698 | 60.29 |
| rs12332731 | A | T | 0.19 | 0.014 | 0.002 | 5.96E-14 | -0.004 | 0.009 | 0.664 | 56.25 |
| rs12375949 | T | C | 0.43 | -0.014 | 0.001 | 1.61E-22 | 0.006 | 0.007 | 0.375 | 97.16 |
| rs12438177 | A | G | 0.37 | 0.009 | 0.002 | 1.48E-10 | -0.014 | 0.008 | 0.068 | 38.44 |
| rs12468040 | T | G | 0.38 | 0.014 | 0.001 | 3.12E-21 | -0.005 | 0.007 | 0.485 | 95.76 |
| rs12503522 | T | C | 0.28 | -0.011 | 0.002 | 3.93E-13 | 0.004 | 0.008 | 0.621 | 49.88 |
| rs12506221 | T | G | 0.44 | -0.013 | 0.001 | 4.23E-20 | 0.012 | 0.007 | 0.113 | 86.22 |
| rs12519073 | T | C | 0.23 | -0.011 | 0.002 | 1.16E-10 | -0.004 | 0.009 | 0.656 | 39.62 |
| rs12524795 | T | C | 0.44 | 0.01 | 0.001 | 5.28E-13 | 0.008 | 0.007 | 0.282 | 53.08 |
| rs12571549 | A | G | 0.14 | 0.015 | 0.002 | 1.45E-13 | 0.009 | 0.011 | 0.416 | 57 |
| rs12574281 | A | C | 0.63 | -0.008 | 0.002 | 3.04E-08 | 0.006 | 0.008 | 0.405 | 28.44 |
| rs12591647 | T | C | 0.82 | -0.015 | 0.002 | 7.78E-17 | -0.026 | 0.009 | 0.005 | 71.31 |
| rs12602286 | T | G | 0.87 | 0.016 | 0.002 | 4.62E-14 | 0.001 | 0.011 | 0.934 | 58.05 |
| rs12614263 | A | G | 0.5 | 0.008 | 0.001 | 3.38E-09 | -0.007 | 0.007 | 0.332 | 35.15 |
| rs12638072 | A | G | 0.3 | -0.011 | 0.002 | 3.13E-12 | -0.012 | 0.008 | 0.128 | 50.88 |
| rs12643771 | T | C | 0.31 | 0.013 | 0.002 | 1.32E-17 | -0.01 | 0.008 | 0.207 | 75.11 |
| rs12646523 | T | C | 0.25 | -0.013 | 0.002 | 3.79E-16 | -0.009 | 0.008 | 0.292 | 68.06 |
| rs12670376 | A | G | 0.45 | 0.009 | 0.001 | 5.99E-11 | 0.008 | 0.007 | 0.253 | 43.18 |
| rs12761761 | T | C | 0.24 | 0.015 | 0.002 | 4.79E-20 | -0.012 | 0.008 | 0.142 | 81 |
| rs12765185 | A | T | 0.28 | -0.009 | 0.002 | 2.96E-08 | -0.013 | 0.008 | 0.106 | 30.25 |
| rs12789313 | T | C | 0.49 | 0.009 | 0.001 | 6.79E-11 | 0.001 | 0.007 | 0.939 | 43.18 |
| rs12875339 | A | C | 0.64 | -0.011 | 0.002 | 1.71E-13 | -0.001 | 0.008 | 0.864 | 51.84 |
| rs12888615 | T | C | 0.2 | 0.012 | 0.002 | 9.42E-12 | -0.015 | 0.009 | 0.093 | 44.44 |
| rs12957463 | A | G | 0.8 | 0.015 | 0.002 | 9.20E-17 | 0.007 | 0.009 | 0.442 | 65.79 |
| rs12967010 | T | C | 0.77 | 0.011 | 0.002 | 1.33E-11 | 0.01 | 0.009 | 0.262 | 44.97 |
| rs12981405 | T | C | 0.17 | -0.011 | 0.002 | 2.75E-09 | -0.019 | 0.01 | 0.048 | 34.75 |
| rs1301838 | T | C | 0.32 | -0.01 | 0.002 | 1.29E-10 | -0.003 | 0.008 | 0.661 | 40.96 |
| rs13018640 | T | C | 0.6 | -0.021 | 0.001 | 1.05E-50 | -0.001 | 0.007 | 0.891 | 235.84 |
| rs13050131 | A | C | 0.66 | -0.008 | 0.002 | 4.40E-08 | -0.007 | 0.008 | 0.401 | 29.16 |
| rs13133213 | A | G | 0.5 | 0.01 | 0.001 | 7.57E-12 | 0.011 | 0.007 | 0.124 | 47.02 |
| rs13145650 | T | C | 0.92 | -0.017 | 0.002 | 8.99E-12 | 0.002 | 0.013 | 0.893 | 47.89 |
| rs13163845 | T | C | 0.85 | -0.015 | 0.002 | 4.66E-14 | -0.03 | 0.011 | 0.005 | 55.5 |
| rs13197257 | T | G | 0.28 | 0.011 | 0.002 | 2.83E-12 | -0.025 | 0.008 | 0.003 | 46.41 |
| rs13212041 | T | C | 0.8 | 0.01 | 0.002 | 7.42E-09 | -0.006 | 0.009 | 0.497 | 31.48 |
| rs13240401 | T | C | 0.78 | 0.018 | 0.002 | 2.36E-24 | -0.003 | 0.009 | 0.72 | 107.18 |
| rs13261773 | C | G | 0.16 | 0.012 | 0.002 | 3.12E-10 | -0.025 | 0.01 | 0.013 | 41.23 |
| rs13266287 | A | T | 0.67 | -0.009 | 0.002 | 1.75E-08 | -0.012 | 0.008 | 0.13 | 32.11 |
| rs1329125 | T | C | 0.33 | -0.01 | 0.002 | 8.37E-11 | 0 | 0.008 | 0.982 | 41.82 |
| rs13327482 | A | G | 0.82 | -0.012 | 0.002 | 4.47E-11 | -0.01 | 0.01 | 0.308 | 45.19 |
| rs1334297 | A | G | 0.74 | 0.026 | 0.002 | 2.53E-58 | 0.005 | 0.008 | 0.573 | 258 |
| rs13422673 | T | C | 0.47 | -0.011 | 0.001 | 6.48E-15 | -0.009 | 0.007 | 0.216 | 61.73 |
| rs137079 | T | C | 0.14 | 0.013 | 0.002 | 3.06E-10 | -0.015 | 0.01 | 0.14 | 40.96 |
| rs1426619 | T | C | 0.43 | 0.01 | 0.001 | 1.40E-11 | 0.009 | 0.007 | 0.218 | 47.02 |
| rs1434630 | T | G | 0.15 | -0.014 | 0.002 | 4.63E-12 | 0.013 | 0.01 | 0.188 | 46.92 |
| rs143743568 | A | G | 0.15 | 0.012 | 0.002 | 4.99E-09 | -0.011 | 0.01 | 0.268 | 34.81 |
| rs150537577 | A | G | 0.92 | -0.015 | 0.003 | 9.50E-09 | 0.019 | 0.014 | 0.169 | 33.73 |
| rs1505676 | C | G | 0.63 | 0.008 | 0.002 | 6.67E-09 | -0.003 | 0.007 | 0.641 | 31.36 |
| rs1542354 | A | G | 0.48 | -0.008 | 0.001 | 2.72E-09 | -0.005 | 0.007 | 0.476 | 36 |
| rs1544 | A | G | 0.26 | -0.01 | 0.002 | 1.76E-09 | 0.007 | 0.008 | 0.406 | 36 |
| rs1564347 | T | G | 0.34 | 0.009 | 0.002 | 2.39E-09 | 0.007 | 0.008 | 0.341 | 34.42 |
| rs1603460 | T | G | 0.42 | 0.009 | 0.001 | 2.20E-10 | -0.003 | 0.007 | 0.7 | 41.33 |
| rs162445 | A | G | 0.08 | 0.015 | 0.003 | 5.72E-09 | -0.01 | 0.014 | 0.471 | 34.18 |
| rs1689510 | C | G | 0.33 | 0.019 | 0.002 | 1.35E-38 | -0.014 | 0.008 | 0.074 | 167.27 |
| rs17048855 | A | G | 0.34 | 0.011 | 0.002 | 3.10E-13 | -0.007 | 0.008 | 0.348 | 51.84 |
| rs17110109 | T | C | 0.62 | -0.009 | 0.001 | 1.47E-10 | -0.001 | 0.007 | 0.843 | 44.13 |
| rs17144467 | A | C | 0.69 | 0.01 | 0.002 | 8.74E-11 | -0.006 | 0.008 | 0.415 | 42.68 |
| rs17148998 | A | G | 0.2 | 0.01 | 0.002 | 6.98E-09 | -0.003 | 0.009 | 0.704 | 35.3 |
| rs1717204 | A | C | 0.18 | -0.012 | 0.002 | 1.20E-10 | -0.012 | 0.009 | 0.219 | 42.25 |
| rs17248751 | A | G | 0.78 | -0.014 | 0.002 | 1.28E-15 | -0.017 | 0.009 | 0.052 | 64 |
| rs173003 | A | C | 0.49 | -0.008 | 0.001 | 5.50E-09 | -0.041 | 0.007 | 1.09E-08 | 34.31 |
| rs1738050 | C | G | 0.62 | -0.009 | 0.001 | 1.42E-10 | 0.024 | 0.007 | 0.001 | 44.13 |
| rs17411339 | A | G | 0.56 | 0.014 | 0.001 | 5.10E-23 | -0.001 | 0.007 | 0.857 | 100 |
| rs17428076 | C | G | 0.76 | 0.014 | 0.002 | 6.41E-17 | 0.013 | 0.008 | 0.134 | 73.32 |
| rs1747714 | T | C | 0.53 | 0.011 | 0.001 | 3.82E-14 | -0.005 | 0.007 | 0.507 | 57.33 |
| rs1747817 | T | C | 0.76 | -0.009 | 0.002 | 2.29E-08 | 0.001 | 0.009 | 0.951 | 33.06 |
| rs17489649 | A | G | 0.68 | 0.013 | 0.002 | 2.77E-17 | 0.013 | 0.008 | 0.084 | 71.68 |
| rs17502934 | T | G | 0.15 | -0.017 | 0.002 | 2.12E-17 | 0 | 0.01 | 0.999 | 69.72 |
| rs175325 | A | T | 0.6 | -0.01 | 0.001 | 2.24E-12 | -0.009 | 0.007 | 0.217 | 51.02 |
| rs17551064 | A | G | 0.84 | 0.014 | 0.002 | 1.53E-13 | 0.012 | 0.01 | 0.231 | 55.07 |
| rs17563464 | A | C | 0.22 | -0.015 | 0.002 | 1.74E-17 | -0.013 | 0.009 | 0.163 | 74.77 |
| rs17565975 | A | G | 0.56 | -0.011 | 0.001 | 5.54E-14 | 0.004 | 0.007 | 0.612 | 57.33 |
| rs17598675 | T | C | 0.51 | -0.012 | 0.001 | 1.37E-16 | -0.005 | 0.007 | 0.469 | 68.65 |
| rs17604349 | A | G | 0.19 | 0.013 | 0.002 | 2.36E-12 | -0.014 | 0.01 | 0.156 | 49.78 |
| rs176218 | T | G | 0.2 | 0.02 | 0.002 | 1.10E-29 | -0.013 | 0.009 | 0.149 | 123.46 |
| rs17732878 | T | C | 0.79 | -0.01 | 0.002 | 4.06E-08 | -0.006 | 0.009 | 0.485 | 31.23 |
| rs178183 | T | C | 0.25 | 0.015 | 0.002 | 4.81E-19 | -0.014 | 0.008 | 0.102 | 82.13 |
| rs17882802 | A | G | 0.43 | 0.008 | 0.001 | 3.00E-09 | 0.015 | 0.007 | 0.042 | 36 |
| rs182902112 | A | C | 0.02 | -0.03 | 0.005 | 1.17E-08 | -0.016 | 0.031 | 0.614 | 32.04 |
| rs1842713 | A | G | 0.79 | -0.012 | 0.002 | 9.67E-12 | 0.014 | 0.009 | 0.124 | 47.37 |
| rs1861786 | A | G | 0.38 | -0.008 | 0.001 | 2.39E-08 | 0.009 | 0.008 | 0.227 | 33.47 |
| rs1865955 | T | C | 0.82 | 0.012 | 0.002 | 4.06E-11 | -0.03 | 0.01 | 0.002 | 41.23 |
| rs1866823 | A | G | 0.54 | 0.011 | 0.001 | 3.09E-14 | 0.012 | 0.007 | 0.109 | 58.41 |
| rs1890132 | T | C | 0.71 | -0.009 | 0.002 | 6.56E-09 | -0.02 | 0.008 | 0.012 | 35.2 |
| rs192436652 | T | C | 0.03 | -0.032 | 0.004 | 2.06E-12 | -0.016 | 0.023 | 0.495 | 50.25 |
| rs1933264 | T | C | 0.75 | 0.01 | 0.002 | 5.85E-10 | 0.006 | 0.008 | 0.443 | 39.06 |
| rs1955250 | A | C | 0.91 | -0.014 | 0.002 | 2.26E-08 | -0.002 | 0.013 | 0.857 | 31.36 |
| rs1991585 | T | C | 0.71 | -0.01 | 0.002 | 6.26E-11 | 0.003 | 0.008 | 0.726 | 45.34 |
| rs2007655 | T | G | 0.5 | 0.009 | 0.001 | 8.64E-10 | 0.002 | 0.007 | 0.766 | 37.73 |
| rs2011603 | A | G | 0.74 | -0.009 | 0.002 | 1.12E-08 | -0.015 | 0.008 | 0.074 | 32.35 |
| rs2034631 | T | C | 0.65 | 0.009 | 0.002 | 2.64E-09 | 0.007 | 0.008 | 0.331 | 35.2 |
| rs2052285 | A | G | 0.6 | 0.011 | 0.001 | 7.25E-15 | 0.005 | 0.007 | 0.502 | 64 |
| rs2055940 | A | G | 0.32 | 0.008 | 0.002 | 4.95E-08 | 0.007 | 0.008 | 0.338 | 29.88 |
| rs2081652 | A | T | 0.66 | 0.012 | 0.002 | 1.09E-16 | -0.006 | 0.008 | 0.431 | 67.24 |
| rs2131167 | A | G | 0.45 | 0.008 | 0.001 | 1.52E-08 | 0.007 | 0.007 | 0.329 | 32.65 |
| rs2179152 | T | C | 0.37 | -0.013 | 0.002 | 1.68E-19 | 0.009 | 0.008 | 0.232 | 76.27 |
| rs2183271 | T | C | 0.64 | 0.008 | 0.002 | 1.91E-08 | 0 | 0.008 | 0.998 | 31.36 |
| rs2199409 | T | C | 0.8 | 0.01 | 0.002 | 3.14E-09 | 0.01 | 0.009 | 0.266 | 36.71 |
| rs2212430 | T | C | 0.3 | -0.01 | 0.002 | 1.32E-10 | -0.015 | 0.008 | 0.062 | 39.06 |
| rs2216144 | T | C | 0.53 | 0.01 | 0.001 | 2.58E-12 | 0.005 | 0.007 | 0.473 | 49 |
| rs2256965 | A | G | 0.42 | 0.011 | 0.001 | 2.40E-13 | 0.002 | 0.007 | 0.77 | 57.33 |
| rs2287838 | A | G | 0.55 | -0.01 | 0.001 | 1.27E-13 | 0.006 | 0.007 | 0.414 | 55.18 |
| rs2290601 | T | C | 0.77 | -0.015 | 0.002 | 4.49E-19 | -0.009 | 0.009 | 0.322 | 76.82 |
| rs2297293 | C | G | 0.31 | 0.01 | 0.002 | 8.80E-11 | 0.016 | 0.008 | 0.043 | 43.56 |
| rs232496 | T | C | 0.37 | 0.009 | 0.002 | 7.14E-10 | 0.021 | 0.008 | 0.007 | 36 |
| rs2364544 | A | G | 0.39 | -0.011 | 0.001 | 2.91E-13 | -0.001 | 0.008 | 0.907 | 56.25 |
| rs2434672 | A | C | 0.53 | 0.009 | 0.001 | 1.06E-10 | -0.013 | 0.007 | 0.092 | 42.25 |
| rs2436760 | T | C | 0.97 | 0.023 | 0.004 | 4.52E-08 | 0.016 | 0.021 | 0.455 | 30.51 |
| rs2447097 | T | G | 0.46 | 0.01 | 0.001 | 1.10E-11 | -0.003 | 0.007 | 0.683 | 47.02 |
| rs2469226 | A | T | 0.22 | 0.01 | 0.002 | 1.28E-08 | 0.006 | 0.009 | 0.529 | 31.89 |
| rs2496482 | T | C | 0.42 | 0.012 | 0.002 | 9.20E-17 | 0.008 | 0.007 | 0.31 | 65.07 |
| rs252991 | A | G | 0.37 | 0.01 | 0.002 | 6.30E-11 | -0.01 | 0.008 | 0.208 | 40.11 |
| rs2545795 | A | C | 0.44 | 0.013 | 0.001 | 1.03E-18 | 0.01 | 0.007 | 0.178 | 79.72 |
| rs2554835 | A | G | 0.4 | 0.009 | 0.001 | 8.95E-10 | -0.005 | 0.007 | 0.51 | 39.51 |
| rs2570497 | T | C | 0.64 | -0.012 | 0.002 | 1.27E-16 | 0.001 | 0.008 | 0.936 | 65.07 |
| rs2725370 | T | C | 0.3 | -0.014 | 0.002 | 4.96E-20 | 0.013 | 0.008 | 0.116 | 88.36 |
| rs2764684 | T | C | 0.83 | 0.014 | 0.002 | 9.99E-15 | 0.02 | 0.01 | 0.035 | 56.65 |
| rs2838006 | T | C | 0.36 | 0.013 | 0.002 | 1.38E-19 | 0.001 | 0.008 | 0.917 | 77.44 |
| rs28513882 | A | G | 0.18 | -0.011 | 0.002 | 2.03E-09 | -0.012 | 0.009 | 0.22 | 37.35 |
| rs2885198 | A | G | 0.53 | 0.009 | 0.001 | 4.63E-10 | -0.022 | 0.007 | 0.003 | 39.51 |
| rs2923424 | A | G | 0.61 | 0.012 | 0.001 | 6.31E-17 | 0.009 | 0.007 | 0.221 | 73.47 |
| rs2929032 | A | G | 0.47 | -0.008 | 0.001 | 8.40E-09 | -0.001 | 0.007 | 0.868 | 33.47 |
| rs2958182 | A | T | 0.34 | 0.008 | 0.002 | 3.46E-08 | 0.015 | 0.008 | 0.052 | 29.88 |
| rs2964199 | T | C | 0.31 | -0.01 | 0.002 | 4.61E-11 | -0.004 | 0.008 | 0.618 | 43.56 |
| rs2964255 | A | G | 0.31 | 0.009 | 0.002 | 6.34E-09 | -0.007 | 0.008 | 0.367 | 34.42 |
| rs2998299 | T | G | 0.21 | -0.014 | 0.002 | 1.51E-15 | 0.005 | 0.009 | 0.561 | 64 |
| rs3026996 | A | C | 0.76 | 0.014 | 0.002 | 5.42E-17 | -0.003 | 0.009 | 0.755 | 74.39 |
| rs303752 | A | G | 0.41 | -0.011 | 0.001 | 1.47E-14 | 0.007 | 0.008 | 0.365 | 61.73 |
| rs3111251 | T | C | 0.43 | -0.009 | 0.001 | 3.88E-10 | 0.006 | 0.007 | 0.417 | 41.33 |
| rs34067381 | T | G | 0.36 | -0.01 | 0.002 | 9.77E-13 | -0.004 | 0.008 | 0.625 | 48.07 |
| rs34098770 | A | G | 0.15 | -0.016 | 0.002 | 2.95E-16 | 0.014 | 0.01 | 0.173 | 64 |
| rs34305371 | A | G | 0.1 | 0.031 | 0.002 | 1.10E-39 | -0.011 | 0.012 | 0.352 | 171.17 |
| rs34394051 | A | G | 0.84 | -0.012 | 0.002 | 2.30E-09 | -0.054 | 0.01 | 5.90E-08 | 34.22 |
| rs34720381 | T | C | 0.09 | -0.018 | 0.002 | 1.90E-13 | 0.001 | 0.013 | 0.939 | 56.25 |
| rs34807077 | A | C | 0.16 | 0.013 | 0.002 | 3.29E-11 | 0.001 | 0.01 | 0.958 | 45.39 |
| rs35417702 | T | C | 0.52 | -0.015 | 0.001 | 2.40E-27 | 0.006 | 0.007 | 0.382 | 117.88 |
| rs35606437 | A | G | 0.27 | 0.011 | 0.002 | 3.98E-11 | 0 | 0.008 | 0.996 | 43.07 |
| rs35929923 | A | G | 0.25 | -0.011 | 0.002 | 1.77E-12 | 0.002 | 0.008 | 0.767 | 50.77 |
| rs363096 | T | C | 0.42 | -0.014 | 0.001 | 5.77E-24 | -0.006 | 0.007 | 0.43 | 104.33 |
| rs3751331 | A | G | 0.38 | -0.01 | 0.001 | 1.75E-11 | 0.003 | 0.008 | 0.686 | 48.01 |
| rs3809634 | A | G | 0.68 | -0.011 | 0.002 | 6.82E-13 | -0.003 | 0.008 | 0.689 | 52.8 |
| rs382196 | T | G | 0.38 | -0.008 | 0.001 | 1.47E-08 | 0.008 | 0.008 | 0.313 | 34.31 |
| rs3859523 | T | C | 0.82 | 0.012 | 0.002 | 9.92E-12 | -0.009 | 0.01 | 0.341 | 47.46 |
| rs3897821 | A | G | 0.67 | 0.015 | 0.002 | 3.58E-25 | 0.005 | 0.008 | 0.553 | 105.4 |
| rs3948495 | T | G | 0.41 | -0.009 | 0.001 | 3.95E-10 | 0.001 | 0.007 | 0.903 | 40.41 |
| rs401526 | T | C | 0.53 | -0.01 | 0.001 | 1.15E-13 | -0.023 | 0.007 | 0.002 | 55.18 |
| rs401966 | C | G | 0.39 | -0.01 | 0.001 | 1.72E-11 | 0.001 | 0.008 | 0.92 | 48.01 |
| rs42210 | C | G | 0.71 | -0.01 | 0.002 | 2.74E-10 | 0.019 | 0.008 | 0.018 | 37.52 |
| rs42302 | A | G | 0.34 | 0.009 | 0.002 | 5.96E-09 | 0.004 | 0.008 | 0.578 | 32.87 |
| rs4298514 | T | C | 0.7 | 0.01 | 0.002 | 1.07E-11 | 0.004 | 0.008 | 0.613 | 48.07 |
| rs4320563 | A | G | 0.51 | 0.011 | 0.001 | 2.24E-14 | -0.005 | 0.007 | 0.466 | 60.62 |
| rs4328757 | T | C | 0.61 | 0.01 | 0.001 | 2.95E-11 | 0.008 | 0.007 | 0.298 | 47.02 |
| rs4352658 | T | C | 0.08 | -0.019 | 0.003 | 1.71E-13 | -0.013 | 0.013 | 0.335 | 53.4 |
| rs4358081 | A | C | 0.53 | -0.01 | 0.001 | 1.28E-11 | -0.001 | 0.007 | 0.941 | 46.05 |
| rs4369924 | A | G | 0.16 | 0.013 | 0.002 | 5.58E-11 | -0.003 | 0.01 | 0.781 | 43.28 |
| rs4384309 | A | G | 0.46 | 0.01 | 0.001 | 4.89E-13 | -0.008 | 0.007 | 0.278 | 53.08 |
| rs4467547 | T | G | 0.41 | 0.012 | 0.001 | 4.88E-17 | -0.01 | 0.007 | 0.158 | 74.7 |
| rs4500930 | T | C | 0.34 | -0.011 | 0.002 | 1.67E-13 | 0.006 | 0.008 | 0.413 | 52.8 |
| rs4663617 | A | T | 0.23 | 0.01 | 0.002 | 4.12E-10 | -0.015 | 0.009 | 0.099 | 37.43 |
| rs4673840 | T | C | 0.84 | -0.014 | 0.002 | 5.86E-13 | 0.003 | 0.01 | 0.731 | 52.75 |
| rs4675248 | A | G | 0.4 | -0.009 | 0.001 | 4.50E-11 | -0.007 | 0.007 | 0.37 | 45.08 |
| rs4719944 | T | C | 0.55 | -0.012 | 0.001 | 9.25E-18 | 0.007 | 0.007 | 0.309 | 74.7 |
| rs4726070 | A | G | 0.6 | 0.012 | 0.001 | 1.36E-17 | 0.004 | 0.007 | 0.62 | 75.94 |
| rs4757957 | C | G | 0.69 | 0.013 | 0.002 | 1.79E-18 | -0.004 | 0.008 | 0.62 | 78.62 |
| rs4766424 | C | G | 0.12 | 0.014 | 0.002 | 2.47E-11 | -0.007 | 0.011 | 0.511 | 45.72 |
| rs4778058 | T | C | 0.5 | -0.009 | 0.001 | 4.92E-11 | 0.001 | 0.008 | 0.896 | 43.18 |
| rs4787457 | A | G | 0.63 | 0.016 | 0.002 | 1.23E-28 | 0.008 | 0.008 | 0.273 | 116.64 |
| rs4793090 | A | G | 0.66 | 0.008 | 0.002 | 1.67E-08 | -0.013 | 0.008 | 0.092 | 31.36 |
| rs4810894 | A | G | 0.37 | -0.009 | 0.002 | 4.38E-09 | 0.015 | 0.008 | 0.054 | 32.11 |
| rs4839155 | T | G | 0.77 | 0.012 | 0.002 | 4.66E-12 | -0.014 | 0.009 | 0.108 | 45.76 |
| rs4846724 | A | G | 0.53 | 0.01 | 0.001 | 2.89E-12 | -0.003 | 0.007 | 0.715 | 49 |
| rs4848924 | A | C | 0.71 | -0.012 | 0.002 | 3.56E-14 | 0.004 | 0.008 | 0.625 | 60.84 |
| rs488476 | C | G | 0.62 | -0.011 | 0.002 | 8.29E-15 | 0.03 | 0.007 | 7.13E-05 | 56.75 |
| rs4895650 | T | C | 0.55 | 0.009 | 0.001 | 5.31E-11 | 0.006 | 0.007 | 0.398 | 44.13 |
| rs4899012 | C | G | 0.6 | -0.01 | 0.001 | 8.06E-13 | -0.001 | 0.007 | 0.924 | 54.13 |
| rs4904523 | A | G | 0.52 | -0.008 | 0.001 | 6.27E-09 | -0.005 | 0.007 | 0.496 | 34.31 |
| rs4915735 | A | G | 0.14 | 0.012 | 0.002 | 9.56E-09 | -0.002 | 0.01 | 0.864 | 33.64 |
| rs4977885 | A | G | 0.4 | -0.008 | 0.001 | 6.78E-09 | -0.012 | 0.007 | 0.113 | 36 |
| rs4984541 | A | G | 0.77 | -0.014 | 0.002 | 3.40E-16 | 0.007 | 0.009 | 0.444 | 65.9 |
| rs55736314 | C | G | 0.6 | -0.015 | 0.001 | 4.59E-27 | -0.002 | 0.007 | 0.755 | 121 |
| rs56099375 | T | C | 0.24 | 0.012 | 0.002 | 3.10E-13 | -0.018 | 0.009 | 0.038 | 49.83 |
| rs56319902 | T | C | 0.22 | -0.021 | 0.002 | 5.81E-33 | -0.054 | 0.009 | 8.83E-10 | 146.84 |
| rs56391344 | A | G | 0.25 | 0.015 | 0.002 | 7.49E-20 | -0.013 | 0.008 | 0.12 | 85.56 |
| rs56794817 | A | G | 0.16 | 0.012 | 0.002 | 1.69E-10 | -0.001 | 0.01 | 0.925 | 41.23 |
| rs57349798 | A | G | 0.41 | 0.009 | 0.001 | 8.63E-11 | 0 | 0.008 | 0.969 | 44.13 |
| rs57352738 | A | T | 0.2 | -0.015 | 0.002 | 2.51E-18 | 0.01 | 0.009 | 0.248 | 79.94 |
| rs575113 | A | G | 0.29 | 0.013 | 0.002 | 7.52E-18 | -0.008 | 0.008 | 0.303 | 78.62 |
| rs5754753 | T | C | 0.72 | -0.013 | 0.002 | 6.15E-16 | 0.005 | 0.008 | 0.535 | 62.02 |
| rs57661533 | T | C | 0.13 | -0.011 | 0.002 | 3.24E-08 | 0.002 | 0.011 | 0.86 | 29.47 |
| rs580652 | T | C | 0.9 | -0.015 | 0.002 | 4.65E-10 | -0.023 | 0.012 | 0.066 | 37.52 |
| rs590013 | T | C | 0.68 | 0.01 | 0.002 | 2.47E-11 | -0.002 | 0.008 | 0.806 | 44.44 |
| rs6043521 | T | C | 0.6 | -0.008 | 0.001 | 9.72E-09 | 0.012 | 0.008 | 0.105 | 35.15 |
| rs6065080 | T | C | 0.36 | -0.013 | 0.002 | 7.49E-19 | 0.004 | 0.008 | 0.556 | 75.11 |
| rs6065784 | C | G | 0.7 | 0.011 | 0.002 | 5.78E-13 | -0.004 | 0.008 | 0.636 | 53.78 |
| rs61104616 | A | G | 0.52 | -0.017 | 0.001 | 1.56E-34 | 0.015 | 0.007 | 0.036 | 150.94 |
| rs6123924 | A | G | 0.84 | 0.013 | 0.002 | 3.20E-11 | 0.002 | 0.01 | 0.854 | 46.1 |
| rs61527214 | A | G | 0.41 | 0.011 | 0.001 | 1.13E-14 | -0.002 | 0.007 | 0.781 | 61.73 |
| rs61997667 | T | C | 0.15 | -0.014 | 0.002 | 2.24E-13 | 0 | 0.01 | 0.99 | 51.84 |
| rs62155350 | A | G | 0.02 | 0.034 | 0.005 | 1.69E-10 | 0.012 | 0.026 | 0.633 | 40.67 |
| rs62155873 | T | C | 0.12 | -0.015 | 0.002 | 1.65E-12 | -0.004 | 0.011 | 0.689 | 51.7 |
| rs62174974 | A | G | 0.19 | -0.011 | 0.002 | 2.76E-09 | 0 | 0.009 | 0.999 | 34.03 |
| rs62179650 | A | G | 0.3 | 0.011 | 0.002 | 5.66E-13 | 0.005 | 0.008 | 0.565 | 49 |
| rs62182994 | T | C | 0.69 | 0.014 | 0.002 | 2.55E-19 | -0.006 | 0.008 | 0.435 | 83.42 |
| rs62379838 | T | C | 0.7 | 0.012 | 0.002 | 1.77E-15 | 0 | 0.008 | 0.952 | 65.07 |
| rs628993 | A | G | 0.11 | 0.013 | 0.002 | 3.83E-08 | 0.01 | 0.013 | 0.438 | 30.97 |
| rs6449503 | A | G | 0.5 | 0.018 | 0.001 | 1.84E-39 | 0.005 | 0.007 | 0.477 | 174.62 |
| rs6490618 | T | C | 0.33 | 0.009 | 0.002 | 3.48E-09 | -0.011 | 0.008 | 0.146 | 34.42 |
| rs6493265 | T | C | 0.39 | -0.012 | 0.001 | 2.26E-17 | -0.008 | 0.007 | 0.301 | 75.94 |
| rs6557171 | T | C | 0.32 | -0.016 | 0.002 | 5.48E-25 | -0.003 | 0.008 | 0.741 | 106.78 |
| rs660001 | A | G | 0.21 | -0.016 | 0.002 | 1.66E-21 | 0.004 | 0.009 | 0.671 | 93.07 |
| rs66568921 | T | G | 0.64 | -0.018 | 0.002 | 7.60E-34 | 0.007 | 0.008 | 0.341 | 145.6 |
| rs66721975 | A | G | 0.29 | -0.009 | 0.002 | 1.51E-08 | -0.006 | 0.008 | 0.441 | 33.64 |
| rs6690195 | T | C | 0.49 | -0.011 | 0.001 | 7.89E-16 | -0.006 | 0.007 | 0.391 | 65.15 |
| rs6697584 | T | C | 0.79 | -0.013 | 0.002 | 1.43E-13 | -0.002 | 0.009 | 0.817 | 54.93 |
| rs6704768 | A | G | 0.56 | -0.012 | 0.001 | 1.84E-16 | 0.004 | 0.007 | 0.604 | 68.65 |
| rs6731373 | A | G | 0.34 | -0.012 | 0.002 | 1.39E-14 | -0.011 | 0.008 | 0.16 | 58.78 |
| rs6812533 | T | C | 0.74 | 0.011 | 0.002 | 4.85E-11 | -0.01 | 0.008 | 0.234 | 43.89 |
| rs68145588 | T | G | 0.14 | -0.013 | 0.002 | 7.53E-10 | -0.002 | 0.01 | 0.861 | 36.57 |
| rs6917154 | T | C | 0.12 | 0.013 | 0.002 | 2.55E-09 | -0.004 | 0.011 | 0.741 | 36.57 |
| rs6924023 | A | G | 0.75 | 0.011 | 0.002 | 5.17E-11 | 0.001 | 0.009 | 0.893 | 44.72 |
| rs6977237 | T | C | 0.47 | 0.008 | 0.001 | 5.59E-09 | 0.009 | 0.007 | 0.214 | 34.31 |
| rs6994287 | A | G | 0.41 | 0.01 | 0.001 | 1.11E-12 | 0.001 | 0.007 | 0.881 | 53.08 |
| rs7012546 | T | C | 0.42 | 0.009 | 0.001 | 1.25E-10 | 0.006 | 0.007 | 0.424 | 43.18 |
| rs7029718 | A | G | 0.41 | 0.024 | 0.001 | 7.82E-65 | 0.01 | 0.007 | 0.181 | 303.76 |
| rs7041702 | A | G | 0.74 | -0.011 | 0.002 | 4.53E-12 | -0.003 | 0.008 | 0.726 | 48.13 |
| rs7167688 | T | C | 0.49 | -0.009 | 0.001 | 1.71E-11 | -0.012 | 0.007 | 0.104 | 45.08 |
| rs7171405 | A | G | 0.24 | -0.01 | 0.002 | 7.53E-10 | -0.018 | 0.009 | 0.034 | 39.85 |
| rs717996 | T | C | 0.6 | -0.012 | 0.001 | 1.89E-17 | -0.004 | 0.007 | 0.618 | 75.94 |
| rs7226824 | T | C | 0.53 | -0.008 | 0.001 | 2.72E-08 | 0.01 | 0.007 | 0.164 | 31.04 |
| rs72486027 | T | C | 0.25 | -0.01 | 0.002 | 1.61E-10 | -0.001 | 0.009 | 0.917 | 42.25 |
| rs7254263 | T | C | 0.28 | -0.012 | 0.002 | 1.84E-14 | -0.014 | 0.008 | 0.08 | 55.32 |
| rs72622559 | T | C | 0.23 | -0.012 | 0.002 | 5.50E-13 | 0.005 | 0.008 | 0.523 | 49.83 |
| rs72672052 | A | T | 0.18 | -0.01 | 0.002 | 4.87E-08 | -0.005 | 0.01 | 0.593 | 31.48 |
| rs72709560 | A | G | 0.69 | 0.009 | 0.002 | 2.03E-08 | 0.021 | 0.008 | 0.007 | 32.11 |
| rs72828517 | T | C | 0.83 | -0.017 | 0.002 | 1.38E-19 | 0.029 | 0.009 | 0.002 | 78.18 |
| rs72829857 | A | G | 0.76 | -0.015 | 0.002 | 1.52E-19 | -0.01 | 0.009 | 0.237 | 86.72 |
| rs72917504 | T | C | 0.06 | -0.018 | 0.003 | 2.91E-09 | 0.009 | 0.016 | 0.546 | 35.2 |
| rs72944064 | T | C | 0.74 | -0.01 | 0.002 | 9.39E-10 | -0.002 | 0.008 | 0.781 | 37.52 |
| rs72962169 | T | C | 0.16 | -0.017 | 0.002 | 1.26E-19 | 0.002 | 0.01 | 0.83 | 83.87 |
| rs72993796 | T | C | 0.88 | 0.013 | 0.002 | 1.12E-08 | 0.018 | 0.011 | 0.106 | 33.32 |
| rs730384 | A | G | 0.44 | 0.01 | 0.001 | 9.38E-13 | 0.001 | 0.007 | 0.922 | 52.05 |
| rs73055556 | A | G | 0.14 | 0.011 | 0.002 | 4.95E-08 | 0.025 | 0.011 | 0.02 | 30.8 |
| rs73191311 | A | G | 0.34 | -0.009 | 0.002 | 2.03E-10 | 0.005 | 0.008 | 0.537 | 39.27 |
| rs7321274 | A | G | 0.8 | 0.011 | 0.002 | 8.21E-11 | 0.014 | 0.009 | 0.135 | 40.11 |
| rs7326331 | A | G | 0.28 | -0.013 | 0.002 | 3.42E-16 | 0.007 | 0.008 | 0.387 | 64 |
| rs73344830 | A | G | 0.42 | 0.017 | 0.001 | 1.00E-32 | -0.003 | 0.007 | 0.642 | 147.45 |
| rs736281 | T | C | 0.39 | 0.009 | 0.001 | 2.49E-09 | 0.004 | 0.007 | 0.562 | 37.73 |
| rs73648455 | T | C | 0.08 | -0.018 | 0.003 | 1.99E-12 | 0.022 | 0.014 | 0.108 | 49.54 |
| rs743316 | T | C | 0.79 | 0.011 | 0.002 | 8.21E-11 | -0.009 | 0.009 | 0.313 | 43.4 |
| rs74415461 | T | C | 0.08 | 0.018 | 0.003 | 6.25E-12 | 0.057 | 0.013 | 7.60E-06 | 45.82 |
| rs7449561 | A | G | 0.23 | 0.01 | 0.002 | 9.73E-10 | -0.006 | 0.009 | 0.485 | 36 |
| rs7460106 | T | C | 0.76 | -0.012 | 0.002 | 2.06E-13 | -0.019 | 0.009 | 0.028 | 52.35 |
| rs746839 | C | G | 0.63 | 0.014 | 0.002 | 1.19E-20 | -0.005 | 0.008 | 0.554 | 81 |
| rs74787922 | A | C | 0.93 | 0.018 | 0.003 | 8.97E-11 | 0.011 | 0.015 | 0.449 | 42.25 |
| rs75033012 | C | G | 0.04 | -0.026 | 0.004 | 1.38E-11 | -0.012 | 0.019 | 0.525 | 46.1 |
| rs75177132 | T | C | 0.05 | 0.03 | 0.004 | 1.01E-16 | 0.02 | 0.017 | 0.231 | 68.52 |
| rs7575637 | A | G | 0.45 | 0.011 | 0.001 | 3.43E-14 | -0.011 | 0.007 | 0.138 | 58.41 |
| rs7603132 | A | G | 0.19 | 0.014 | 0.002 | 1.20E-14 | 0.006 | 0.009 | 0.52 | 57.93 |
| rs76076331 | T | C | 0.13 | 0.021 | 0.002 | 1.71E-23 | -0.024 | 0.011 | 0.031 | 96.23 |
| rs7617204 | A | G | 0.52 | -0.01 | 0.001 | 4.33E-12 | -0.019 | 0.007 | 0.01 | 48.01 |
| rs76235882 | A | G | 0.97 | 0.024 | 0.004 | 4.52E-10 | 0.015 | 0.022 | 0.509 | 38.19 |
| rs76246107 | A | G | 0.09 | -0.016 | 0.003 | 6.52E-10 | 0.009 | 0.014 | 0.541 | 38.82 |
| rs7650602 | T | C | 0.56 | -0.009 | 0.001 | 2.77E-10 | 0.003 | 0.007 | 0.706 | 40.41 |
| rs76577427 | C | G | 0.9 | 0.02 | 0.002 | 7.78E-18 | -0.011 | 0.012 | 0.36 | 71.54 |
| rs7672622 | A | G | 0.75 | -0.009 | 0.002 | 4.72E-08 | 0.001 | 0.008 | 0.888 | 30.25 |
| rs7683416 | T | C | 0.46 | 0.014 | 0.001 | 6.05E-23 | 0.001 | 0.007 | 0.929 | 98.58 |
| rs76878669 | C | G | 0.76 | 0.014 | 0.002 | 3.10E-17 | -0.015 | 0.009 | 0.09 | 68.79 |
| rs7692359 | T | C | 0.78 | -0.011 | 0.002 | 1.40E-10 | 0.008 | 0.009 | 0.364 | 41.11 |
| rs77025239 | A | G | 0.16 | -0.014 | 0.002 | 4.65E-13 | -0.016 | 0.01 | 0.11 | 54.29 |
| rs77609760 | A | G | 0.93 | 0.024 | 0.003 | 7.71E-17 | 0.022 | 0.015 | 0.133 | 71.04 |
| rs77719387 | A | T | 0.02 | -0.04 | 0.006 | 3.78E-12 | 0.012 | 0.031 | 0.686 | 48.28 |
| rs7849487 | T | G | 0.66 | -0.017 | 0.002 | 1.25E-29 | 0.008 | 0.008 | 0.29 | 123.95 |
| rs78648104 | T | C | 0.91 | -0.014 | 0.002 | 1.70E-08 | -0.018 | 0.013 | 0.163 | 31.81 |
| rs7875078 | A | C | 0.46 | -0.008 | 0.001 | 2.72E-08 | -0.002 | 0.007 | 0.762 | 31.04 |
| rs7894722 | T | C | 0.37 | 0.009 | 0.001 | 5.41E-09 | 0.008 | 0.008 | 0.263 | 36.86 |
| rs790647 | A | C | 0.23 | -0.015 | 0.002 | 3.81E-18 | -0.015 | 0.009 | 0.082 | 72.75 |
| rs7910403 | T | G | 0.2 | 0.012 | 0.002 | 5.08E-12 | 0.002 | 0.009 | 0.825 | 45.94 |
| rs7924036 | T | G | 0.51 | 0.013 | 0.001 | 2.55E-21 | 0.02 | 0.007 | 0.006 | 90.25 |
| rs79265434 | A | G | 0.88 | -0.02 | 0.002 | 1.98E-19 | -0.006 | 0.011 | 0.627 | 80.18 |
| rs7972246 | T | C | 0.35 | 0.011 | 0.002 | 1.01E-13 | 0.008 | 0.008 | 0.299 | 53.78 |
| rs79728014 | A | G | 0.86 | -0.013 | 0.002 | 2.06E-11 | -0.005 | 0.01 | 0.652 | 44.89 |
| rs7974852 | A | C | 0.52 | 0.012 | 0.001 | 1.14E-16 | -0.013 | 0.007 | 0.075 | 69.84 |
| rs7977614 | A | G | 0.71 | -0.013 | 0.002 | 6.84E-17 | -0.01 | 0.008 | 0.196 | 70.14 |
| rs79994730 | T | C | 0.04 | 0.024 | 0.004 | 5.88E-12 | -0.026 | 0.017 | 0.122 | 47.02 |
| rs8008382 | T | C | 0.3 | -0.011 | 0.002 | 4.07E-12 | -0.009 | 0.008 | 0.244 | 49.94 |
| rs8024 | A | C | 0.33 | -0.012 | 0.002 | 1.13E-14 | -0.004 | 0.008 | 0.635 | 58.78 |
| rs80257979 | T | G | 0.97 | -0.025 | 0.004 | 3.31E-10 | 0.006 | 0.021 | 0.77 | 39.69 |
| rs8030487 | A | G | 0.69 | 0.009 | 0.002 | 2.52E-08 | -0.002 | 0.008 | 0.809 | 32.11 |
| rs806816 | A | T | 0.75 | 0.01 | 0.002 | 4.80E-10 | 0.003 | 0.008 | 0.718 | 39.85 |
| rs8097125 | T | C | 0.38 | 0.009 | 0.001 | 2.31E-09 | -0.002 | 0.007 | 0.811 | 37.73 |
| rs912883 | T | C | 0.68 | 0.009 | 0.002 | 6.20E-09 | 0.011 | 0.008 | 0.167 | 33.64 |
| rs9289300 | T | C | 0.84 | -0.016 | 0.002 | 1.00E-17 | 0.014 | 0.01 | 0.178 | 74.5 |
| rs929511 | T | C | 0.13 | -0.016 | 0.002 | 3.91E-14 | 0.001 | 0.011 | 0.901 | 58.05 |
| rs9359939 | A | C | 0.24 | -0.011 | 0.002 | 1.51E-10 | -0.005 | 0.009 | 0.539 | 43.07 |
| rs936496 | A | G | 0.62 | -0.008 | 0.001 | 4.38E-08 | -0.009 | 0.008 | 0.256 | 31.84 |
| rs9373363 | A | G | 0.75 | -0.011 | 0.002 | 5.54E-12 | -0.007 | 0.008 | 0.381 | 48.13 |
| rs9375188 | T | C | 0.48 | 0.021 | 0.001 | 8.78E-52 | 0.007 | 0.007 | 0.357 | 229.31 |
| rs9411331 | A | C | 0.68 | 0.015 | 0.002 | 7.63E-22 | 0.002 | 0.008 | 0.84 | 93.44 |
| rs9492774 | C | G | 0.37 | 0.008 | 0.002 | 4.33E-08 | -0.002 | 0.008 | 0.758 | 28.44 |
| rs9529146 | T | C | 0.23 | 0.013 | 0.002 | 3.48E-14 | 0.002 | 0.009 | 0.851 | 55.81 |
| rs9556958 | T | C | 0.52 | -0.011 | 0.001 | 5.96E-15 | 0.005 | 0.007 | 0.534 | 61.73 |
| rs9616947 | T | C | 0.38 | -0.013 | 0.002 | 3.58E-20 | 0.007 | 0.008 | 0.329 | 79.8 |
| rs9633970 | T | C | 0.76 | 0.009 | 0.002 | 2.81E-08 | 0.001 | 0.009 | 0.939 | 32.35 |
| rs9655780 | A | G | 0.83 | -0.015 | 0.002 | 3.70E-16 | 0.006 | 0.01 | 0.557 | 64.84 |
| rs969512 | A | T | 0.66 | -0.011 | 0.002 | 9.84E-13 | -0.004 | 0.008 | 0.566 | 49.94 |
| rs9853928 | T | C | 0.21 | -0.013 | 0.002 | 1.59E-13 | -0.011 | 0.009 | 0.229 | 55.81 |
| rs9859556 | T | G | 0.31 | 0.029 | 0.002 | 4.61E-82 | -0.017 | 0.008 | 0.028 | 373.78 |
| rs9882532 | T | C | 0.64 | 0.012 | 0.002 | 2.15E-17 | 0.006 | 0.008 | 0.438 | 68.34 |
| rs9886703 | A | T | 0.17 | -0.014 | 0.002 | 4.46E-13 | 0.007 | 0.01 | 0.453 | 51.24 |
| rs9927137 | A | G | 0.53 | 0.008 | 0.001 | 5.11E-09 | -0.014 | 0.007 | 0.064 | 34.31 |
| rs9927842 | T | C | 0.15 | -0.013 | 0.002 | 6.22E-11 | 0.005 | 0.01 | 0.626 | 42.25 |
| rs9929556 | T | G | 0.57 | -0.01 | 0.001 | 2.31E-12 | -0.012 | 0.007 | 0.109 | 50.01 |
| rs9929762 | A | G | 0.56 | 0.01 | 0.001 | 1.03E-11 | 0.009 | 0.007 | 0.224 | 47.02 |
| rs9933256 | A | G | 0.56 | 0.012 | 0.001 | 6.06E-17 | 0.001 | 0.007 | 0.9 | 72.25 |

**SNPs used as instrumental variables in the MR analysis of educational attainment on mean diffusivity (white matter tracts)**

| **SNP** | **Effect allele** | **Other allele** | **Effect allele frequency** | **beta (exposure)** | **SE (exposure)** | ***P* (exposure)** | **beta (outcome)** | **SE (outcome)** | ***P* (outcome)** | ***F*-statistic** |
| --- | --- | --- | --- | --- | --- | --- | --- | --- | --- | --- |
| rs10006235 | T | C | 0.27 | -0.009 | 0.002 | 4.48E-09 | -0.004 | 0.008 | 0.576 | 33.79 |
| rs10021733 | T | C | 0.15 | -0.016 | 0.003 | 9.45E-10 | 0.009 | 0.01 | 0.37 | 36.89 |
| rs10046069 | A | G | 0.12 | 0.013 | 0.002 | 4.40E-09 | -0.002 | 0.011 | 0.867 | 33.32 |
| rs10060023 | T | C | 0.33 | 0.011 | 0.002 | 1.88E-13 | -0.005 | 0.007 | 0.526 | 53.78 |
| rs10080647 | A | C | 0.14 | 0.012 | 0.002 | 7.05E-10 | 0.01 | 0.01 | 0.317 | 38.44 |
| rs1008078 | T | C | 0.4 | -0.018 | 0.001 | 4.50E-37 | 0.008 | 0.007 | 0.278 | 169 |
| rs10189857 | A | G | 0.57 | 0.016 | 0.001 | 4.69E-29 | 0.004 | 0.007 | 0.592 | 127.37 |
| rs10193498 | A | T | 0.76 | 0.01 | 0.002 | 3.10E-09 | 0.002 | 0.008 | 0.803 | 36.75 |
| rs10215082 | A | G | 0.47 | -0.015 | 0.001 | 7.03E-26 | 0.008 | 0.007 | 0.286 | 113.27 |
| rs10460095 | A | G | 0.57 | -0.012 | 0.001 | 2.61E-18 | 0.014 | 0.007 | 0.06 | 78.45 |
| rs10761251 | A | T | 0.66 | 0.011 | 0.002 | 2.52E-14 | 0.005 | 0.008 | 0.54 | 56.75 |
| rs10772644 | C | G | 0.89 | 0.014 | 0.002 | 1.69E-10 | -0.012 | 0.011 | 0.281 | 41.66 |
| rs10773002 | A | T | 0.25 | 0.018 | 0.002 | 2.50E-30 | 0.009 | 0.008 | 0.298 | 133.69 |
| rs10773208 | T | C | 0.25 | -0.011 | 0.002 | 8.09E-12 | 0.006 | 0.008 | 0.476 | 47.27 |
| rs10798888 | T | G | 0.17 | -0.014 | 0.002 | 5.15E-14 | -0.003 | 0.009 | 0.775 | 54.29 |
| rs10799615 | A | G | 0.25 | -0.009 | 0.002 | 4.11E-08 | -0.003 | 0.008 | 0.711 | 30.94 |
| rs10805383 | A | G | 0.49 | -0.01 | 0.001 | 2.22E-13 | 0.013 | 0.007 | 0.069 | 54.13 |
| rs10830858 | T | C | 0.51 | 0.009 | 0.001 | 5.31E-11 | 0.004 | 0.007 | 0.62 | 43.18 |
| rs10844179 | A | G | 0.23 | 0.01 | 0.002 | 5.03E-10 | -0.004 | 0.008 | 0.653 | 37.43 |
| rs10875121 | C | G | 0.84 | 0.017 | 0.002 | 4.46E-20 | 0.016 | 0.01 | 0.091 | 83.87 |
| rs10892807 | T | C | 0.57 | 0.011 | 0.001 | 1.09E-14 | 0.001 | 0.007 | 0.908 | 60.62 |
| rs10940921 | T | G | 0.42 | 0.011 | 0.001 | 1.31E-14 | 0.008 | 0.007 | 0.242 | 62.86 |
| rs10951590 | T | C | 0.33 | -0.011 | 0.002 | 3.62E-13 | -0.001 | 0.008 | 0.883 | 52.8 |
| rs10979613 | T | C | 0.64 | -0.012 | 0.002 | 1.25E-15 | 0.027 | 0.007 | 2.11E-04 | 60.84 |
| rs10996167 | C | G | 0.65 | 0.011 | 0.002 | 7.47E-09 | 0.002 | 0.007 | 0.772 | 31.92 |
| rs11003463 | T | G | 0.61 | 0.009 | 0.002 | 5.58E-10 | -0.008 | 0.007 | 0.267 | 36.8 |
| rs11023764 | A | G | 0.64 | 0.009 | 0.002 | 5.32E-10 | 0.004 | 0.008 | 0.61 | 36.8 |
| rs11030102 | C | G | 0.75 | 0.01 | 0.002 | 4.60E-10 | 0.009 | 0.008 | 0.267 | 39.85 |
| rs11076962 | T | C | 0.72 | 0.009 | 0.002 | 4.29E-08 | 0.006 | 0.008 | 0.442 | 28.89 |
| rs11081529 | T | C | 0.71 | 0.013 | 0.002 | 5.22E-16 | -0.002 | 0.008 | 0.829 | 70.56 |
| rs11082011 | T | C | 0.67 | 0.019 | 0.002 | 2.06E-38 | -0.01 | 0.008 | 0.197 | 167.27 |
| rs11121177 | A | G | 0.18 | 0.015 | 0.002 | 1.44E-16 | 0.006 | 0.009 | 0.488 | 70.37 |
| rs11130380 | T | G | 0.61 | 0.009 | 0.001 | 4.17E-10 | -0.002 | 0.007 | 0.797 | 41.33 |
| rs11138947 | T | C | 0.72 | 0.011 | 0.002 | 2.36E-11 | -0.008 | 0.008 | 0.285 | 43.07 |
| rs11157931 | A | C | 0.39 | -0.014 | 0.001 | 3.86E-22 | -0.01 | 0.007 | 0.151 | 98.58 |
| rs111852224 | T | C | 0.12 | 0.016 | 0.002 | 4.85E-13 | 0.001 | 0.011 | 0.902 | 50.28 |
| rs11211123 | A | G | 0.22 | 0.01 | 0.002 | 7.62E-10 | -0.002 | 0.008 | 0.842 | 37.43 |
| rs11213482 | A | G | 0.84 | -0.011 | 0.002 | 2.68E-08 | -0.004 | 0.009 | 0.701 | 31.12 |
| rs112603734 | A | C | 0.77 | -0.013 | 0.002 | 2.72E-09 | -0.002 | 0.008 | 0.813 | 35.46 |
| rs112806496 | C | G | 0.91 | -0.016 | 0.002 | 1.72E-10 | -0.02 | 0.012 | 0.105 | 40.96 |
| rs1128956 | T | G | 0.83 | -0.014 | 0.002 | 5.34E-14 | 0 | 0.009 | 0.975 | 54.29 |
| rs113520408 | A | G | 0.28 | 0.016 | 0.002 | 1.03E-22 | 0.002 | 0.008 | 0.819 | 93.85 |
| rs113615161 | T | C | 0.13 | -0.012 | 0.002 | 2.98E-09 | 0.011 | 0.01 | 0.293 | 34.31 |
| rs114593137 | A | T | 0.79 | -0.012 | 0.002 | 1.65E-12 | -0.009 | 0.009 | 0.29 | 51.5 |
| rs115000530 | A | T | 0.95 | -0.025 | 0.003 | 1.09E-15 | 0.033 | 0.016 | 0.038 | 65.04 |
| rs11542663 | A | C | 0.69 | 0.011 | 0.002 | 2.44E-13 | 0.001 | 0.008 | 0.849 | 55.75 |
| rs11588857 | A | G | 0.21 | 0.02 | 0.002 | 4.45E-31 | 0.025 | 0.009 | 0.003 | 137.03 |
| rs11598765 | A | G | 0.2 | -0.011 | 0.002 | 1.73E-09 | 0.003 | 0.009 | 0.726 | 34.68 |
| rs11620355 | A | G | 0.09 | 0.018 | 0.002 | 1.09E-13 | 0.03 | 0.013 | 0.018 | 53 |
| rs11657342 | A | G | 0.37 | 0.014 | 0.002 | 2.32E-13 | -0.02 | 0.007 | 0.007 | 54.29 |
| rs11657979 | A | G | 0.24 | -0.009 | 0.002 | 4.57E-08 | 0.005 | 0.009 | 0.601 | 28.65 |
| rs11663602 | A | C | 0.28 | -0.013 | 0.002 | 9.27E-17 | -0.01 | 0.008 | 0.192 | 66.02 |
| rs1167827 | A | G | 0.43 | 0.01 | 0.001 | 2.93E-13 | 0.009 | 0.007 | 0.219 | 55.18 |
| rs11724690 | T | G | 0.29 | 0.009 | 0.002 | 2.08E-09 | -0.003 | 0.008 | 0.722 | 38.44 |
| rs117398064 | C | G | 0.09 | -0.014 | 0.002 | 1.16E-08 | 0.006 | 0.012 | 0.597 | 31.81 |
| rs118134876 | T | C | 0.06 | -0.022 | 0.003 | 1.50E-13 | 0 | 0.015 | 0.993 | 55.25 |
| rs11871429 | A | G | 0.77 | 0.013 | 0.002 | 2.52E-14 | -0.005 | 0.008 | 0.533 | 56.69 |
| rs11894424 | A | C | 0.95 | 0.021 | 0.003 | 2.14E-10 | -0.004 | 0.017 | 0.799 | 40.11 |
| rs12054166 | C | G | 0.74 | 0.01 | 0.002 | 2.30E-09 | 0.008 | 0.008 | 0.299 | 36 |
| rs12076635 | C | G | 0.78 | 0.021 | 0.002 | 3.33E-34 | -0.013 | 0.009 | 0.144 | 148.27 |
| rs12113634 | T | C | 0.65 | 0.009 | 0.002 | 2.50E-09 | -0.001 | 0.007 | 0.864 | 34.42 |
| rs12151248 | T | C | 0.11 | -0.015 | 0.002 | 3.81E-11 | 0.001 | 0.011 | 0.952 | 41.97 |
| rs1220779 | A | G | 0.47 | -0.008 | 0.001 | 1.58E-08 | -0.004 | 0.007 | 0.584 | 31.84 |
| rs12273435 | A | G | 0.21 | -0.013 | 0.002 | 4.72E-14 | -0.001 | 0.009 | 0.898 | 60.29 |
| rs12332731 | A | T | 0.19 | 0.014 | 0.002 | 5.96E-14 | 0.011 | 0.009 | 0.205 | 56.25 |
| rs12375949 | T | C | 0.43 | -0.014 | 0.001 | 1.61E-22 | -0.011 | 0.007 | 0.129 | 97.16 |
| rs12438177 | A | G | 0.37 | 0.009 | 0.002 | 1.48E-10 | 0.015 | 0.007 | 0.041 | 38.44 |
| rs12468040 | T | G | 0.38 | 0.014 | 0.001 | 3.12E-21 | 0.008 | 0.007 | 0.286 | 95.76 |
| rs12503522 | T | C | 0.28 | -0.011 | 0.002 | 3.93E-13 | -0.001 | 0.008 | 0.941 | 49.88 |
| rs12506221 | T | G | 0.44 | -0.013 | 0.001 | 4.23E-20 | -0.008 | 0.007 | 0.259 | 86.22 |
| rs12519073 | T | C | 0.23 | -0.011 | 0.002 | 1.16E-10 | 0.005 | 0.008 | 0.544 | 39.62 |
| rs12524795 | T | C | 0.44 | 0.01 | 0.001 | 5.28E-13 | -0.014 | 0.007 | 0.056 | 53.08 |
| rs12571549 | A | G | 0.14 | 0.015 | 0.002 | 1.45E-13 | -0.001 | 0.01 | 0.933 | 57 |
| rs12574281 | A | C | 0.63 | -0.008 | 0.002 | 3.04E-08 | 0.004 | 0.007 | 0.627 | 28.44 |
| rs12591647 | T | C | 0.82 | -0.015 | 0.002 | 7.78E-17 | 0.012 | 0.009 | 0.183 | 71.31 |
| rs12602286 | T | G | 0.87 | 0.016 | 0.002 | 4.62E-14 | 0.001 | 0.011 | 0.907 | 58.05 |
| rs12614263 | A | G | 0.5 | 0.008 | 0.001 | 3.38E-09 | 0.003 | 0.007 | 0.655 | 35.15 |
| rs12638072 | A | G | 0.3 | -0.011 | 0.002 | 3.13E-12 | 0.005 | 0.008 | 0.538 | 50.88 |
| rs12643771 | T | C | 0.31 | 0.013 | 0.002 | 1.32E-17 | 0.007 | 0.008 | 0.372 | 75.11 |
| rs12646523 | T | C | 0.25 | -0.013 | 0.002 | 3.79E-16 | -0.012 | 0.008 | 0.158 | 68.06 |
| rs12670376 | A | G | 0.45 | 0.009 | 0.001 | 5.99E-11 | -0.01 | 0.007 | 0.178 | 43.18 |
| rs12761761 | T | C | 0.24 | 0.015 | 0.002 | 4.79E-20 | 0.005 | 0.008 | 0.526 | 81 |
| rs12765185 | A | T | 0.28 | -0.009 | 0.002 | 2.96E-08 | 0.009 | 0.008 | 0.264 | 30.25 |
| rs12789313 | T | C | 0.49 | 0.009 | 0.001 | 6.79E-11 | -0.001 | 0.007 | 0.862 | 43.18 |
| rs12875339 | A | C | 0.64 | -0.011 | 0.002 | 1.71E-13 | 0 | 0.007 | 0.972 | 51.84 |
| rs12888615 | T | C | 0.2 | 0.012 | 0.002 | 9.42E-12 | 0.008 | 0.009 | 0.383 | 44.44 |
| rs12957463 | A | G | 0.8 | 0.015 | 0.002 | 9.20E-17 | -0.002 | 0.009 | 0.821 | 65.79 |
| rs12967010 | T | C | 0.77 | 0.011 | 0.002 | 1.33E-11 | -0.008 | 0.009 | 0.324 | 44.97 |
| rs12981405 | T | C | 0.17 | -0.011 | 0.002 | 2.75E-09 | 0.001 | 0.01 | 0.889 | 34.75 |
| rs1301838 | T | C | 0.32 | -0.01 | 0.002 | 1.29E-10 | 0.006 | 0.008 | 0.414 | 40.96 |
| rs13018640 | T | C | 0.6 | -0.021 | 0.001 | 1.05E-50 | -0.005 | 0.007 | 0.471 | 235.84 |
| rs13050131 | A | C | 0.66 | -0.008 | 0.002 | 4.40E-08 | 0.005 | 0.008 | 0.538 | 29.16 |
| rs13133213 | A | G | 0.5 | 0.01 | 0.001 | 7.57E-12 | -0.024 | 0.007 | 6.47E-04 | 47.02 |
| rs13145650 | T | C | 0.92 | -0.017 | 0.002 | 8.99E-12 | 0.006 | 0.013 | 0.637 | 47.89 |
| rs13163845 | T | C | 0.85 | -0.015 | 0.002 | 4.66E-14 | 0.029 | 0.01 | 0.004 | 55.5 |
| rs13197257 | T | G | 0.28 | 0.011 | 0.002 | 2.83E-12 | 0.016 | 0.008 | 0.042 | 46.41 |
| rs13212041 | T | C | 0.8 | 0.01 | 0.002 | 7.42E-09 | 0.003 | 0.009 | 0.746 | 31.48 |
| rs13240401 | T | C | 0.78 | 0.018 | 0.002 | 2.36E-24 | 0.004 | 0.008 | 0.596 | 107.18 |
| rs13261773 | C | G | 0.16 | 0.012 | 0.002 | 3.12E-10 | 0.012 | 0.01 | 0.223 | 41.23 |
| rs13266287 | A | T | 0.67 | -0.009 | 0.002 | 1.75E-08 | 0.013 | 0.008 | 0.08 | 32.11 |
| rs1329125 | T | C | 0.33 | -0.01 | 0.002 | 8.37E-11 | -0.001 | 0.008 | 0.887 | 41.82 |
| rs13327482 | A | G | 0.82 | -0.012 | 0.002 | 4.47E-11 | 0.006 | 0.009 | 0.519 | 45.19 |
| rs1334297 | A | G | 0.74 | 0.026 | 0.002 | 2.53E-58 | -0.001 | 0.008 | 0.907 | 258 |
| rs13422673 | T | C | 0.47 | -0.011 | 0.001 | 6.48E-15 | 0.008 | 0.007 | 0.278 | 61.73 |
| rs137079 | T | C | 0.14 | 0.013 | 0.002 | 3.06E-10 | 0.017 | 0.01 | 0.088 | 40.96 |
| rs1426619 | T | C | 0.43 | 0.01 | 0.001 | 1.40E-11 | -0.015 | 0.007 | 0.039 | 47.02 |
| rs1434630 | T | G | 0.15 | -0.014 | 0.002 | 4.63E-12 | -0.029 | 0.01 | 0.004 | 46.92 |
| rs143743568 | A | G | 0.15 | 0.012 | 0.002 | 4.99E-09 | -0.002 | 0.01 | 0.838 | 34.81 |
| rs150537577 | A | G | 0.92 | -0.015 | 0.003 | 9.50E-09 | -0.02 | 0.013 | 0.135 | 33.73 |
| rs1505676 | C | G | 0.63 | 0.008 | 0.002 | 6.67E-09 | 0.003 | 0.007 | 0.647 | 31.36 |
| rs1542354 | A | G | 0.48 | -0.008 | 0.001 | 2.72E-09 | 0.003 | 0.007 | 0.705 | 36 |
| rs1544 | A | G | 0.26 | -0.01 | 0.002 | 1.76E-09 | -0.015 | 0.008 | 0.057 | 36 |
| rs1564347 | T | G | 0.34 | 0.009 | 0.002 | 2.39E-09 | 0.006 | 0.007 | 0.459 | 34.42 |
| rs1603460 | T | G | 0.42 | 0.009 | 0.001 | 2.20E-10 | 0.008 | 0.007 | 0.272 | 41.33 |
| rs162445 | A | G | 0.08 | 0.015 | 0.003 | 5.72E-09 | 0.006 | 0.013 | 0.641 | 34.18 |
| rs1689510 | C | G | 0.33 | 0.019 | 0.002 | 1.35E-38 | 0.029 | 0.007 | 9.39E-05 | 167.27 |
| rs17048855 | A | G | 0.34 | 0.011 | 0.002 | 3.10E-13 | 0.002 | 0.007 | 0.786 | 51.84 |
| rs17110109 | T | C | 0.62 | -0.009 | 0.001 | 1.47E-10 | 0.008 | 0.007 | 0.278 | 44.13 |
| rs17144467 | A | C | 0.69 | 0.01 | 0.002 | 8.74E-11 | 0.004 | 0.008 | 0.646 | 42.68 |
| rs17148998 | A | G | 0.2 | 0.01 | 0.002 | 6.98E-09 | 0.013 | 0.009 | 0.138 | 35.3 |
| rs1717204 | A | C | 0.18 | -0.012 | 0.002 | 1.20E-10 | 0.003 | 0.009 | 0.754 | 42.25 |
| rs17248751 | A | G | 0.78 | -0.014 | 0.002 | 1.28E-15 | 0.012 | 0.009 | 0.177 | 64 |
| rs173003 | A | C | 0.49 | -0.008 | 0.001 | 5.50E-09 | 0.015 | 0.007 | 0.038 | 34.31 |
| rs1738050 | C | G | 0.62 | -0.009 | 0.001 | 1.42E-10 | -0.021 | 0.007 | 0.004 | 44.13 |
| rs17411339 | A | G | 0.56 | 0.014 | 0.001 | 5.10E-23 | -0.003 | 0.007 | 0.652 | 100 |
| rs17428076 | C | G | 0.76 | 0.014 | 0.002 | 6.41E-17 | 0.001 | 0.008 | 0.891 | 73.32 |
| rs1747714 | T | C | 0.53 | 0.011 | 0.001 | 3.82E-14 | 0.002 | 0.007 | 0.825 | 57.33 |
| rs1747817 | T | C | 0.76 | -0.009 | 0.002 | 2.29E-08 | 0.009 | 0.008 | 0.274 | 33.06 |
| rs17489649 | A | G | 0.68 | 0.013 | 0.002 | 2.77E-17 | -0.012 | 0.007 | 0.111 | 71.68 |
| rs17502934 | T | G | 0.15 | -0.017 | 0.002 | 2.12E-17 | -0.006 | 0.01 | 0.563 | 69.72 |
| rs175325 | A | T | 0.6 | -0.01 | 0.001 | 2.24E-12 | 0.003 | 0.007 | 0.679 | 51.02 |
| rs17551064 | A | G | 0.84 | 0.014 | 0.002 | 1.53E-13 | -0.005 | 0.01 | 0.576 | 55.07 |
| rs17563464 | A | C | 0.22 | -0.015 | 0.002 | 1.74E-17 | 0.006 | 0.009 | 0.491 | 74.77 |
| rs17565975 | A | G | 0.56 | -0.011 | 0.001 | 5.54E-14 | -0.006 | 0.007 | 0.409 | 57.33 |
| rs17598675 | T | C | 0.51 | -0.012 | 0.001 | 1.37E-16 | 0.004 | 0.007 | 0.565 | 68.65 |
| rs17604349 | A | G | 0.19 | 0.013 | 0.002 | 2.36E-12 | 0.01 | 0.009 | 0.28 | 49.78 |
| rs176218 | T | G | 0.2 | 0.02 | 0.002 | 1.10E-29 | 0.016 | 0.009 | 0.069 | 123.46 |
| rs17732878 | T | C | 0.79 | -0.01 | 0.002 | 4.06E-08 | -0.002 | 0.009 | 0.857 | 31.23 |
| rs178183 | T | C | 0.25 | 0.015 | 0.002 | 4.81E-19 | 0.017 | 0.008 | 0.031 | 82.13 |
| rs17882802 | A | G | 0.43 | 0.008 | 0.001 | 3.00E-09 | -0.007 | 0.007 | 0.348 | 36 |
| rs182902112 | A | C | 0.02 | -0.03 | 0.005 | 1.17E-08 | 0.003 | 0.03 | 0.923 | 32.04 |
| rs1842713 | A | G | 0.79 | -0.012 | 0.002 | 9.67E-12 | -0.01 | 0.009 | 0.225 | 47.37 |
| rs1861786 | A | G | 0.38 | -0.008 | 0.001 | 2.39E-08 | -0.004 | 0.007 | 0.606 | 33.47 |
| rs1865955 | T | C | 0.82 | 0.012 | 0.002 | 4.06E-11 | 0.017 | 0.009 | 0.068 | 41.23 |
| rs1866823 | A | G | 0.54 | 0.011 | 0.001 | 3.09E-14 | -0.012 | 0.007 | 0.106 | 58.41 |
| rs1890132 | T | C | 0.71 | -0.009 | 0.002 | 6.56E-09 | 0.017 | 0.008 | 0.025 | 35.2 |
| rs192436652 | T | C | 0.03 | -0.032 | 0.004 | 2.06E-12 | -0.01 | 0.022 | 0.664 | 50.25 |
| rs1933264 | T | C | 0.75 | 0.01 | 0.002 | 5.85E-10 | -0.016 | 0.008 | 0.05 | 39.06 |
| rs1955250 | A | C | 0.91 | -0.014 | 0.002 | 2.26E-08 | 0.004 | 0.013 | 0.785 | 31.36 |
| rs1991585 | T | C | 0.71 | -0.01 | 0.002 | 6.26E-11 | 0.006 | 0.008 | 0.447 | 45.34 |
| rs2007655 | T | G | 0.5 | 0.009 | 0.001 | 8.64E-10 | -0.001 | 0.007 | 0.842 | 37.73 |
| rs2011603 | A | G | 0.74 | -0.009 | 0.002 | 1.12E-08 | 0.005 | 0.008 | 0.543 | 32.35 |
| rs2034631 | T | C | 0.65 | 0.009 | 0.002 | 2.64E-09 | 0.005 | 0.007 | 0.467 | 35.2 |
| rs2052285 | A | G | 0.6 | 0.011 | 0.001 | 7.25E-15 | 0 | 0.007 | 0.951 | 64 |
| rs2055940 | A | G | 0.32 | 0.008 | 0.002 | 4.95E-08 | -0.002 | 0.008 | 0.77 | 29.88 |
| rs2081652 | A | T | 0.66 | 0.012 | 0.002 | 1.09E-16 | 0.007 | 0.007 | 0.383 | 67.24 |
| rs2131167 | A | G | 0.45 | 0.008 | 0.001 | 1.52E-08 | -0.007 | 0.007 | 0.352 | 32.65 |
| rs2179152 | T | C | 0.37 | -0.013 | 0.002 | 1.68E-19 | -0.014 | 0.007 | 0.065 | 76.27 |
| rs2183271 | T | C | 0.64 | 0.008 | 0.002 | 1.91E-08 | 0.005 | 0.007 | 0.516 | 31.36 |
| rs2199409 | T | C | 0.8 | 0.01 | 0.002 | 3.14E-09 | -0.009 | 0.009 | 0.318 | 36.71 |
| rs2212430 | T | C | 0.3 | -0.01 | 0.002 | 1.32E-10 | 0.016 | 0.008 | 0.039 | 39.06 |
| rs2216144 | T | C | 0.53 | 0.01 | 0.001 | 2.58E-12 | -0.009 | 0.007 | 0.225 | 49 |
| rs2256965 | A | G | 0.42 | 0.011 | 0.001 | 2.40E-13 | -0.001 | 0.007 | 0.924 | 57.33 |
| rs2287838 | A | G | 0.55 | -0.01 | 0.001 | 1.27E-13 | -0.003 | 0.007 | 0.703 | 55.18 |
| rs2290601 | T | C | 0.77 | -0.015 | 0.002 | 4.49E-19 | 0.004 | 0.008 | 0.649 | 76.82 |
| rs2297293 | C | G | 0.31 | 0.01 | 0.002 | 8.80E-11 | -0.009 | 0.008 | 0.253 | 43.56 |
| rs232496 | T | C | 0.37 | 0.009 | 0.002 | 7.14E-10 | -0.013 | 0.007 | 0.087 | 36 |
| rs2364544 | A | G | 0.39 | -0.011 | 0.001 | 2.91E-13 | 0.007 | 0.007 | 0.34 | 56.25 |
| rs2434672 | A | C | 0.53 | 0.009 | 0.001 | 1.06E-10 | 0.011 | 0.007 | 0.132 | 42.25 |
| rs2436760 | T | C | 0.97 | 0.023 | 0.004 | 4.52E-08 | 0.01 | 0.021 | 0.622 | 30.51 |
| rs2447097 | T | G | 0.46 | 0.01 | 0.001 | 1.10E-11 | -0.007 | 0.007 | 0.328 | 47.02 |
| rs2469226 | A | T | 0.22 | 0.01 | 0.002 | 1.28E-08 | -0.005 | 0.009 | 0.529 | 31.89 |
| rs2496482 | T | C | 0.42 | 0.012 | 0.002 | 9.20E-17 | -0.015 | 0.007 | 0.045 | 65.07 |
| rs252991 | A | G | 0.37 | 0.01 | 0.002 | 6.30E-11 | 0.006 | 0.007 | 0.428 | 40.11 |
| rs2545795 | A | C | 0.44 | 0.013 | 0.001 | 1.03E-18 | -0.002 | 0.007 | 0.733 | 79.72 |
| rs2554835 | A | G | 0.4 | 0.009 | 0.001 | 8.95E-10 | 0.01 | 0.007 | 0.16 | 39.51 |
| rs2570497 | T | C | 0.64 | -0.012 | 0.002 | 1.27E-16 | 0.003 | 0.007 | 0.669 | 65.07 |
| rs2725370 | T | C | 0.3 | -0.014 | 0.002 | 4.96E-20 | -0.006 | 0.008 | 0.462 | 88.36 |
| rs2764684 | T | C | 0.83 | 0.014 | 0.002 | 9.99E-15 | -0.031 | 0.009 | 8.90E-04 | 56.65 |
| rs2838006 | T | C | 0.36 | 0.013 | 0.002 | 1.38E-19 | -0.008 | 0.007 | 0.262 | 77.44 |
| rs28513882 | A | G | 0.18 | -0.011 | 0.002 | 2.03E-09 | 0.013 | 0.009 | 0.175 | 37.35 |
| rs2885198 | A | G | 0.53 | 0.009 | 0.001 | 4.63E-10 | 0.033 | 0.007 | 4.72E-06 | 39.51 |
| rs2923424 | A | G | 0.61 | 0.012 | 0.001 | 6.31E-17 | -0.014 | 0.007 | 0.059 | 73.47 |
| rs2929032 | A | G | 0.47 | -0.008 | 0.001 | 8.40E-09 | 0.005 | 0.007 | 0.47 | 33.47 |
| rs2958182 | A | T | 0.34 | 0.008 | 0.002 | 3.46E-08 | -0.008 | 0.008 | 0.307 | 29.88 |
| rs2964199 | T | C | 0.31 | -0.01 | 0.002 | 4.61E-11 | 0.004 | 0.008 | 0.647 | 43.56 |
| rs2964255 | A | G | 0.31 | 0.009 | 0.002 | 6.34E-09 | 0.008 | 0.008 | 0.297 | 34.42 |
| rs2998299 | T | G | 0.21 | -0.014 | 0.002 | 1.51E-15 | -0.005 | 0.009 | 0.548 | 64 |
| rs3026996 | A | C | 0.76 | 0.014 | 0.002 | 5.42E-17 | -0.001 | 0.008 | 0.891 | 74.39 |
| rs303752 | A | G | 0.41 | -0.011 | 0.001 | 1.47E-14 | -0.01 | 0.007 | 0.158 | 61.73 |
| rs3111251 | T | C | 0.43 | -0.009 | 0.001 | 3.88E-10 | -0.011 | 0.007 | 0.139 | 41.33 |
| rs34067381 | T | G | 0.36 | -0.01 | 0.002 | 9.77E-13 | -0.003 | 0.007 | 0.659 | 48.07 |
| rs34098770 | A | G | 0.15 | -0.016 | 0.002 | 2.95E-16 | -0.018 | 0.01 | 0.064 | 64 |
| rs34305371 | A | G | 0.1 | 0.031 | 0.002 | 1.10E-39 | -0.006 | 0.012 | 0.618 | 171.17 |
| rs34394051 | A | G | 0.84 | -0.012 | 0.002 | 2.30E-09 | 0.041 | 0.01 | 2.75E-05 | 34.22 |
| rs34720381 | T | C | 0.09 | -0.018 | 0.002 | 1.90E-13 | -0.014 | 0.012 | 0.262 | 56.25 |
| rs34807077 | A | C | 0.16 | 0.013 | 0.002 | 3.29E-11 | -0.004 | 0.01 | 0.693 | 45.39 |
| rs35417702 | T | C | 0.52 | -0.015 | 0.001 | 2.40E-27 | -0.001 | 0.007 | 0.894 | 117.88 |
| rs35606437 | A | G | 0.27 | 0.011 | 0.002 | 3.98E-11 | 0.005 | 0.008 | 0.507 | 43.07 |
| rs35929923 | A | G | 0.25 | -0.011 | 0.002 | 1.77E-12 | -0.006 | 0.008 | 0.444 | 50.77 |
| rs363096 | T | C | 0.42 | -0.014 | 0.001 | 5.77E-24 | 0.005 | 0.007 | 0.467 | 104.33 |
| rs3751331 | A | G | 0.38 | -0.01 | 0.001 | 1.75E-11 | -0.001 | 0.007 | 0.935 | 48.01 |
| rs3809634 | A | G | 0.68 | -0.011 | 0.002 | 6.82E-13 | 0.002 | 0.008 | 0.762 | 52.8 |
| rs382196 | T | G | 0.38 | -0.008 | 0.001 | 1.47E-08 | -0.002 | 0.007 | 0.801 | 34.31 |
| rs3859523 | T | C | 0.82 | 0.012 | 0.002 | 9.92E-12 | 0.011 | 0.009 | 0.233 | 47.46 |
| rs3897821 | A | G | 0.67 | 0.015 | 0.002 | 3.58E-25 | -0.004 | 0.008 | 0.589 | 105.4 |
| rs3948495 | T | G | 0.41 | -0.009 | 0.001 | 3.95E-10 | -0.003 | 0.007 | 0.633 | 40.41 |
| rs401526 | T | C | 0.53 | -0.01 | 0.001 | 1.15E-13 | 0.01 | 0.007 | 0.142 | 55.18 |
| rs401966 | C | G | 0.39 | -0.01 | 0.001 | 1.72E-11 | -0.004 | 0.007 | 0.565 | 48.01 |
| rs42210 | C | G | 0.71 | -0.01 | 0.002 | 2.74E-10 | -0.024 | 0.008 | 0.003 | 37.52 |
| rs42302 | A | G | 0.34 | 0.009 | 0.002 | 5.96E-09 | 0.004 | 0.007 | 0.566 | 32.87 |
| rs4298514 | T | C | 0.7 | 0.01 | 0.002 | 1.07E-11 | -0.001 | 0.008 | 0.931 | 48.07 |
| rs4320563 | A | G | 0.51 | 0.011 | 0.001 | 2.24E-14 | 0.012 | 0.007 | 0.078 | 60.62 |
| rs4328757 | T | C | 0.61 | 0.01 | 0.001 | 2.95E-11 | -0.01 | 0.007 | 0.163 | 47.02 |
| rs4352658 | T | C | 0.08 | -0.019 | 0.003 | 1.71E-13 | 0.005 | 0.013 | 0.718 | 53.4 |
| rs4358081 | A | C | 0.53 | -0.01 | 0.001 | 1.28E-11 | 0.006 | 0.007 | 0.379 | 46.05 |
| rs4369924 | A | G | 0.16 | 0.013 | 0.002 | 5.58E-11 | 0.01 | 0.01 | 0.302 | 43.28 |
| rs4384309 | A | G | 0.46 | 0.01 | 0.001 | 4.89E-13 | 0.009 | 0.007 | 0.233 | 53.08 |
| rs4467547 | T | G | 0.41 | 0.012 | 0.001 | 4.88E-17 | 0.009 | 0.007 | 0.215 | 74.7 |
| rs4500930 | T | C | 0.34 | -0.011 | 0.002 | 1.67E-13 | -0.005 | 0.007 | 0.471 | 52.8 |
| rs4663617 | A | T | 0.23 | 0.01 | 0.002 | 4.12E-10 | 0.003 | 0.009 | 0.746 | 37.43 |
| rs4673840 | T | C | 0.84 | -0.014 | 0.002 | 5.86E-13 | -0.008 | 0.01 | 0.386 | 52.75 |
| rs4675248 | A | G | 0.4 | -0.009 | 0.001 | 4.50E-11 | 0.011 | 0.007 | 0.132 | 45.08 |
| rs4719944 | T | C | 0.55 | -0.012 | 0.001 | 9.25E-18 | -0.004 | 0.007 | 0.615 | 74.7 |
| rs4726070 | A | G | 0.6 | 0.012 | 0.001 | 1.36E-17 | 0.006 | 0.007 | 0.423 | 75.94 |
| rs4757957 | C | G | 0.69 | 0.013 | 0.002 | 1.79E-18 | 0.014 | 0.008 | 0.073 | 78.62 |
| rs4766424 | C | G | 0.12 | 0.014 | 0.002 | 2.47E-11 | 0.01 | 0.011 | 0.339 | 45.72 |
| rs4778058 | T | C | 0.5 | -0.009 | 0.001 | 4.92E-11 | -0.009 | 0.007 | 0.193 | 43.18 |
| rs4787457 | A | G | 0.63 | 0.016 | 0.002 | 1.23E-28 | -0.006 | 0.007 | 0.423 | 116.64 |
| rs4793090 | A | G | 0.66 | 0.008 | 0.002 | 1.67E-08 | 0.011 | 0.007 | 0.134 | 31.36 |
| rs4810894 | A | G | 0.37 | -0.009 | 0.002 | 4.38E-09 | 0.002 | 0.007 | 0.793 | 32.11 |
| rs4839155 | T | G | 0.77 | 0.012 | 0.002 | 4.66E-12 | 0.018 | 0.008 | 0.034 | 45.76 |
| rs4846724 | A | G | 0.53 | 0.01 | 0.001 | 2.89E-12 | 0.001 | 0.007 | 0.876 | 49 |
| rs4848924 | A | C | 0.71 | -0.012 | 0.002 | 3.56E-14 | -0.003 | 0.008 | 0.744 | 60.84 |
| rs488476 | C | G | 0.62 | -0.011 | 0.002 | 8.29E-15 | -0.028 | 0.007 | 9.54E-05 | 56.75 |
| rs4895650 | T | C | 0.55 | 0.009 | 0.001 | 5.31E-11 | -0.005 | 0.007 | 0.448 | 44.13 |
| rs4899012 | C | G | 0.6 | -0.01 | 0.001 | 8.06E-13 | -0.001 | 0.007 | 0.922 | 54.13 |
| rs4904523 | A | G | 0.52 | -0.008 | 0.001 | 6.27E-09 | 0.002 | 0.007 | 0.814 | 34.31 |
| rs4915735 | A | G | 0.14 | 0.012 | 0.002 | 9.56E-09 | 0.003 | 0.01 | 0.753 | 33.64 |
| rs4977885 | A | G | 0.4 | -0.008 | 0.001 | 6.78E-09 | 0.01 | 0.007 | 0.155 | 36 |
| rs4984541 | A | G | 0.77 | -0.014 | 0.002 | 3.40E-16 | 0.008 | 0.008 | 0.345 | 65.9 |
| rs55736314 | C | G | 0.6 | -0.015 | 0.001 | 4.59E-27 | -0.001 | 0.007 | 0.943 | 121 |
| rs56099375 | T | C | 0.24 | 0.012 | 0.002 | 3.10E-13 | 0.01 | 0.008 | 0.235 | 49.83 |
| rs56319902 | T | C | 0.22 | -0.021 | 0.002 | 5.81E-33 | 0.057 | 0.008 | 1.82E-11 | 146.84 |
| rs56391344 | A | G | 0.25 | 0.015 | 0.002 | 7.49E-20 | 0.009 | 0.008 | 0.292 | 85.56 |
| rs56794817 | A | G | 0.16 | 0.012 | 0.002 | 1.69E-10 | 0.002 | 0.01 | 0.85 | 41.23 |
| rs57349798 | A | G | 0.41 | 0.009 | 0.001 | 8.63E-11 | -0.003 | 0.007 | 0.728 | 44.13 |
| rs57352738 | A | T | 0.2 | -0.015 | 0.002 | 2.51E-18 | -0.005 | 0.009 | 0.586 | 79.94 |
| rs575113 | A | G | 0.29 | 0.013 | 0.002 | 7.52E-18 | 0.002 | 0.008 | 0.789 | 78.62 |
| rs5754753 | T | C | 0.72 | -0.013 | 0.002 | 6.15E-16 | 0.003 | 0.008 | 0.682 | 62.02 |
| rs57661533 | T | C | 0.13 | -0.011 | 0.002 | 3.24E-08 | 0.002 | 0.011 | 0.839 | 29.47 |
| rs580652 | T | C | 0.9 | -0.015 | 0.002 | 4.65E-10 | 0.019 | 0.012 | 0.12 | 37.52 |
| rs590013 | T | C | 0.68 | 0.01 | 0.002 | 2.47E-11 | 0 | 0.008 | 0.958 | 44.44 |
| rs6043521 | T | C | 0.6 | -0.008 | 0.001 | 9.72E-09 | -0.011 | 0.007 | 0.142 | 35.15 |
| rs6065080 | T | C | 0.36 | -0.013 | 0.002 | 7.49E-19 | -0.012 | 0.007 | 0.104 | 75.11 |
| rs6065784 | C | G | 0.7 | 0.011 | 0.002 | 5.78E-13 | 0.001 | 0.008 | 0.905 | 53.78 |
| rs61104616 | A | G | 0.52 | -0.017 | 0.001 | 1.56E-34 | -0.012 | 0.007 | 0.104 | 150.94 |
| rs6123924 | A | G | 0.84 | 0.013 | 0.002 | 3.20E-11 | 0.006 | 0.01 | 0.531 | 46.1 |
| rs61527214 | A | G | 0.41 | 0.011 | 0.001 | 1.13E-14 | 0.001 | 0.007 | 0.849 | 61.73 |
| rs61997667 | T | C | 0.15 | -0.014 | 0.002 | 2.24E-13 | 0.002 | 0.01 | 0.814 | 51.84 |
| rs62155350 | A | G | 0.02 | 0.034 | 0.005 | 1.69E-10 | 0.022 | 0.025 | 0.388 | 40.67 |
| rs62155873 | T | C | 0.12 | -0.015 | 0.002 | 1.65E-12 | 0.003 | 0.011 | 0.809 | 51.7 |
| rs62174974 | A | G | 0.19 | -0.011 | 0.002 | 2.76E-09 | 0.001 | 0.009 | 0.934 | 34.03 |
| rs62179650 | A | G | 0.3 | 0.011 | 0.002 | 5.66E-13 | -0.007 | 0.008 | 0.371 | 49 |
| rs62182994 | T | C | 0.69 | 0.014 | 0.002 | 2.55E-19 | 0.006 | 0.008 | 0.403 | 83.42 |
| rs62379838 | T | C | 0.7 | 0.012 | 0.002 | 1.77E-15 | 0.003 | 0.008 | 0.652 | 65.07 |
| rs628993 | A | G | 0.11 | 0.013 | 0.002 | 3.83E-08 | 0.004 | 0.012 | 0.775 | 30.97 |
| rs6449503 | A | G | 0.5 | 0.018 | 0.001 | 1.84E-39 | 0.005 | 0.007 | 0.486 | 174.62 |
| rs6490618 | T | C | 0.33 | 0.009 | 0.002 | 3.48E-09 | 0.014 | 0.008 | 0.061 | 34.42 |
| rs6493265 | T | C | 0.39 | -0.012 | 0.001 | 2.26E-17 | 0.008 | 0.007 | 0.284 | 75.94 |
| rs6557171 | T | C | 0.32 | -0.016 | 0.002 | 5.48E-25 | 0.001 | 0.007 | 0.894 | 106.78 |
| rs660001 | A | G | 0.21 | -0.016 | 0.002 | 1.66E-21 | -0.004 | 0.009 | 0.62 | 93.07 |
| rs66568921 | T | G | 0.64 | -0.018 | 0.002 | 7.60E-34 | -0.015 | 0.007 | 0.043 | 145.6 |
| rs66721975 | A | G | 0.29 | -0.009 | 0.002 | 1.51E-08 | 0.013 | 0.008 | 0.111 | 33.64 |
| rs6690195 | T | C | 0.49 | -0.011 | 0.001 | 7.89E-16 | 0.009 | 0.007 | 0.227 | 65.15 |
| rs6697584 | T | C | 0.79 | -0.013 | 0.002 | 1.43E-13 | 0.005 | 0.009 | 0.554 | 54.93 |
| rs6704768 | A | G | 0.56 | -0.012 | 0.001 | 1.84E-16 | -0.003 | 0.007 | 0.631 | 68.65 |
| rs6731373 | A | G | 0.34 | -0.012 | 0.002 | 1.39E-14 | 0.003 | 0.007 | 0.729 | 58.78 |
| rs6812533 | T | C | 0.74 | 0.011 | 0.002 | 4.85E-11 | 0.001 | 0.008 | 0.864 | 43.89 |
| rs68145588 | T | G | 0.14 | -0.013 | 0.002 | 7.53E-10 | 0.009 | 0.01 | 0.365 | 36.57 |
| rs6917154 | T | C | 0.12 | 0.013 | 0.002 | 2.55E-09 | 0.005 | 0.011 | 0.653 | 36.57 |
| rs6924023 | A | G | 0.75 | 0.011 | 0.002 | 5.17E-11 | -0.003 | 0.008 | 0.704 | 44.72 |
| rs6977237 | T | C | 0.47 | 0.008 | 0.001 | 5.59E-09 | -0.007 | 0.007 | 0.318 | 34.31 |
| rs6994287 | A | G | 0.41 | 0.01 | 0.001 | 1.11E-12 | 0.009 | 0.007 | 0.219 | 53.08 |
| rs7012546 | T | C | 0.42 | 0.009 | 0.001 | 1.25E-10 | -0.003 | 0.007 | 0.696 | 43.18 |
| rs7029718 | A | G | 0.41 | 0.024 | 0.001 | 7.82E-65 | -0.007 | 0.007 | 0.352 | 303.76 |
| rs7041702 | A | G | 0.74 | -0.011 | 0.002 | 4.53E-12 | 0.002 | 0.008 | 0.766 | 48.13 |
| rs7167688 | T | C | 0.49 | -0.009 | 0.001 | 1.71E-11 | 0.002 | 0.007 | 0.758 | 45.08 |
| rs7171405 | A | G | 0.24 | -0.01 | 0.002 | 7.53E-10 | 0.01 | 0.008 | 0.231 | 39.85 |
| rs717996 | T | C | 0.6 | -0.012 | 0.001 | 1.89E-17 | 0.002 | 0.007 | 0.788 | 75.94 |
| rs7226824 | T | C | 0.53 | -0.008 | 0.001 | 2.72E-08 | -0.007 | 0.007 | 0.29 | 31.04 |
| rs72486027 | T | C | 0.25 | -0.01 | 0.002 | 1.61E-10 | 0.012 | 0.008 | 0.136 | 42.25 |
| rs7254263 | T | C | 0.28 | -0.012 | 0.002 | 1.84E-14 | 0.012 | 0.008 | 0.131 | 55.32 |
| rs72622559 | T | C | 0.23 | -0.012 | 0.002 | 5.50E-13 | -0.004 | 0.008 | 0.614 | 49.83 |
| rs72672052 | A | T | 0.18 | -0.01 | 0.002 | 4.87E-08 | -0.005 | 0.009 | 0.563 | 31.48 |
| rs72709560 | A | G | 0.69 | 0.009 | 0.002 | 2.03E-08 | -0.017 | 0.008 | 0.025 | 32.11 |
| rs72828517 | T | C | 0.83 | -0.017 | 0.002 | 1.38E-19 | -0.012 | 0.009 | 0.195 | 78.18 |
| rs72829857 | A | G | 0.76 | -0.015 | 0.002 | 1.52E-19 | 0.009 | 0.008 | 0.271 | 86.72 |
| rs72917504 | T | C | 0.06 | -0.018 | 0.003 | 2.91E-09 | -0.003 | 0.015 | 0.849 | 35.2 |
| rs72944064 | T | C | 0.74 | -0.01 | 0.002 | 9.39E-10 | 0.005 | 0.008 | 0.557 | 37.52 |
| rs72962169 | T | C | 0.16 | -0.017 | 0.002 | 1.26E-19 | -0.005 | 0.01 | 0.583 | 83.87 |
| rs72993796 | T | C | 0.88 | 0.013 | 0.002 | 1.12E-08 | -0.016 | 0.011 | 0.138 | 33.32 |
| rs730384 | A | G | 0.44 | 0.01 | 0.001 | 9.38E-13 | -0.011 | 0.007 | 0.129 | 52.05 |
| rs73055556 | A | G | 0.14 | 0.011 | 0.002 | 4.95E-08 | -0.012 | 0.01 | 0.229 | 30.8 |
| rs73191311 | A | G | 0.34 | -0.009 | 0.002 | 2.03E-10 | -0.001 | 0.007 | 0.893 | 39.27 |
| rs7321274 | A | G | 0.8 | 0.011 | 0.002 | 8.21E-11 | -0.015 | 0.009 | 0.094 | 40.11 |
| rs7326331 | A | G | 0.28 | -0.013 | 0.002 | 3.42E-16 | -0.012 | 0.008 | 0.143 | 64 |
| rs73344830 | A | G | 0.42 | 0.017 | 0.001 | 1.00E-32 | -0.001 | 0.007 | 0.905 | 147.45 |
| rs736281 | T | C | 0.39 | 0.009 | 0.001 | 2.49E-09 | -0.003 | 0.007 | 0.661 | 37.73 |
| rs73648455 | T | C | 0.08 | -0.018 | 0.003 | 1.99E-12 | -0.009 | 0.013 | 0.522 | 49.54 |
| rs743316 | T | C | 0.79 | 0.011 | 0.002 | 8.21E-11 | 0.014 | 0.009 | 0.109 | 43.4 |
| rs74415461 | T | C | 0.08 | 0.018 | 0.003 | 6.25E-12 | -0.045 | 0.012 | 2.57E-04 | 45.82 |
| rs7449561 | A | G | 0.23 | 0.01 | 0.002 | 9.73E-10 | 0.011 | 0.008 | 0.184 | 36 |
| rs7460106 | T | C | 0.76 | -0.012 | 0.002 | 2.06E-13 | 0.017 | 0.009 | 0.046 | 52.35 |
| rs746839 | C | G | 0.63 | 0.014 | 0.002 | 1.19E-20 | 0.008 | 0.008 | 0.27 | 81 |
| rs74787922 | A | C | 0.93 | 0.018 | 0.003 | 8.97E-11 | -0.008 | 0.014 | 0.573 | 42.25 |
| rs75033012 | C | G | 0.04 | -0.026 | 0.004 | 1.38E-11 | 0 | 0.018 | 0.991 | 46.1 |
| rs75177132 | T | C | 0.05 | 0.03 | 0.004 | 1.01E-16 | -0.004 | 0.017 | 0.82 | 68.52 |
| rs7575637 | A | G | 0.45 | 0.011 | 0.001 | 3.43E-14 | 0.023 | 0.007 | 0.001 | 58.41 |
| rs7603132 | A | G | 0.19 | 0.014 | 0.002 | 1.20E-14 | -0.007 | 0.009 | 0.464 | 57.93 |
| rs76076331 | T | C | 0.13 | 0.021 | 0.002 | 1.71E-23 | 0.018 | 0.011 | 0.088 | 96.23 |
| rs7617204 | A | G | 0.52 | -0.01 | 0.001 | 4.33E-12 | 0.013 | 0.007 | 0.066 | 48.01 |
| rs76235882 | A | G | 0.97 | 0.024 | 0.004 | 4.52E-10 | -0.017 | 0.022 | 0.432 | 38.19 |
| rs76246107 | A | G | 0.09 | -0.016 | 0.003 | 6.52E-10 | -0.032 | 0.014 | 0.02 | 38.82 |
| rs7650602 | T | C | 0.56 | -0.009 | 0.001 | 2.77E-10 | -0.011 | 0.007 | 0.112 | 40.41 |
| rs76577427 | C | G | 0.9 | 0.02 | 0.002 | 7.78E-18 | 0.013 | 0.012 | 0.259 | 71.54 |
| rs7672622 | A | G | 0.75 | -0.009 | 0.002 | 4.72E-08 | -0.005 | 0.008 | 0.543 | 30.25 |
| rs7683416 | T | C | 0.46 | 0.014 | 0.001 | 6.05E-23 | -0.004 | 0.007 | 0.588 | 98.58 |
| rs76878669 | C | G | 0.76 | 0.014 | 0.002 | 3.10E-17 | 0.012 | 0.009 | 0.166 | 68.79 |
| rs7692359 | T | C | 0.78 | -0.011 | 0.002 | 1.40E-10 | -0.026 | 0.009 | 0.003 | 41.11 |
| rs77025239 | A | G | 0.16 | -0.014 | 0.002 | 4.65E-13 | 0.021 | 0.01 | 0.032 | 54.29 |
| rs77609760 | A | G | 0.93 | 0.024 | 0.003 | 7.71E-17 | -0.023 | 0.014 | 0.102 | 71.04 |
| rs77719387 | A | T | 0.02 | -0.04 | 0.006 | 3.78E-12 | -0.02 | 0.03 | 0.507 | 48.28 |
| rs7849487 | T | G | 0.66 | -0.017 | 0.002 | 1.25E-29 | -0.004 | 0.007 | 0.612 | 123.95 |
| rs78648104 | T | C | 0.91 | -0.014 | 0.002 | 1.70E-08 | 0.006 | 0.013 | 0.613 | 31.81 |
| rs7875078 | A | C | 0.46 | -0.008 | 0.001 | 2.72E-08 | 0.008 | 0.007 | 0.285 | 31.04 |
| rs7894722 | T | C | 0.37 | 0.009 | 0.001 | 5.41E-09 | -0.011 | 0.007 | 0.145 | 36.86 |
| rs790647 | A | C | 0.23 | -0.015 | 0.002 | 3.81E-18 | 0.002 | 0.008 | 0.832 | 72.75 |
| rs7910403 | T | G | 0.2 | 0.012 | 0.002 | 5.08E-12 | 0 | 0.009 | 0.97 | 45.94 |
| rs7924036 | T | G | 0.51 | 0.013 | 0.001 | 2.55E-21 | -0.026 | 0.007 | 2.63E-04 | 90.25 |
| rs79265434 | A | G | 0.88 | -0.02 | 0.002 | 1.98E-19 | 0.017 | 0.011 | 0.127 | 80.18 |
| rs7972246 | T | C | 0.35 | 0.011 | 0.002 | 1.01E-13 | 0.002 | 0.007 | 0.737 | 53.78 |
| rs79728014 | A | G | 0.86 | -0.013 | 0.002 | 2.06E-11 | -0.008 | 0.01 | 0.444 | 44.89 |
| rs7974852 | A | C | 0.52 | 0.012 | 0.001 | 1.14E-16 | 0.01 | 0.007 | 0.152 | 69.84 |
| rs7977614 | A | G | 0.71 | -0.013 | 0.002 | 6.84E-17 | 0.007 | 0.008 | 0.371 | 70.14 |
| rs79994730 | T | C | 0.04 | 0.024 | 0.004 | 5.88E-12 | 0.027 | 0.016 | 0.094 | 47.02 |
| rs8008382 | T | C | 0.3 | -0.011 | 0.002 | 4.07E-12 | 0.005 | 0.008 | 0.536 | 49.94 |
| rs8024 | A | C | 0.33 | -0.012 | 0.002 | 1.13E-14 | 0.004 | 0.007 | 0.637 | 58.78 |
| rs80257979 | T | G | 0.97 | -0.025 | 0.004 | 3.31E-10 | -0.039 | 0.021 | 0.06 | 39.69 |
| rs8030487 | A | G | 0.69 | 0.009 | 0.002 | 2.52E-08 | 0.004 | 0.008 | 0.613 | 32.11 |
| rs806816 | A | T | 0.75 | 0.01 | 0.002 | 4.80E-10 | 0 | 0.008 | 0.984 | 39.85 |
| rs8097125 | T | C | 0.38 | 0.009 | 0.001 | 2.31E-09 | 0.006 | 0.007 | 0.424 | 37.73 |
| rs912883 | T | C | 0.68 | 0.009 | 0.002 | 6.20E-09 | -0.019 | 0.008 | 0.012 | 33.64 |
| rs9289300 | T | C | 0.84 | -0.016 | 0.002 | 1.00E-17 | -0.018 | 0.01 | 0.061 | 74.5 |
| rs929511 | T | C | 0.13 | -0.016 | 0.002 | 3.91E-14 | -0.001 | 0.011 | 0.938 | 58.05 |
| rs9359939 | A | C | 0.24 | -0.011 | 0.002 | 1.51E-10 | 0.002 | 0.009 | 0.848 | 43.07 |
| rs936496 | A | G | 0.62 | -0.008 | 0.001 | 4.38E-08 | 0.009 | 0.007 | 0.244 | 31.84 |
| rs9373363 | A | G | 0.75 | -0.011 | 0.002 | 5.54E-12 | 0 | 0.008 | 0.954 | 48.13 |
| rs9375188 | T | C | 0.48 | 0.021 | 0.001 | 8.78E-52 | 0 | 0.007 | 0.985 | 229.31 |
| rs9411331 | A | C | 0.68 | 0.015 | 0.002 | 7.63E-22 | 0 | 0.008 | 0.991 | 93.44 |
| rs9492774 | C | G | 0.37 | 0.008 | 0.002 | 4.33E-08 | 0.003 | 0.007 | 0.635 | 28.44 |
| rs9529146 | T | C | 0.23 | 0.013 | 0.002 | 3.48E-14 | 0.015 | 0.008 | 0.077 | 55.81 |
| rs9556958 | T | C | 0.52 | -0.011 | 0.001 | 5.96E-15 | -0.011 | 0.007 | 0.121 | 61.73 |
| rs9616947 | T | C | 0.38 | -0.013 | 0.002 | 3.58E-20 | -0.007 | 0.007 | 0.336 | 79.8 |
| rs9633970 | T | C | 0.76 | 0.009 | 0.002 | 2.81E-08 | -0.002 | 0.009 | 0.846 | 32.35 |
| rs9655780 | A | G | 0.83 | -0.015 | 0.002 | 3.70E-16 | 0.003 | 0.01 | 0.755 | 64.84 |
| rs969512 | A | T | 0.66 | -0.011 | 0.002 | 9.84E-13 | 0.001 | 0.008 | 0.916 | 49.94 |
| rs9853928 | T | C | 0.21 | -0.013 | 0.002 | 1.59E-13 | 0.012 | 0.009 | 0.17 | 55.81 |
| rs9859556 | T | G | 0.31 | 0.029 | 0.002 | 4.61E-82 | 0.015 | 0.008 | 0.056 | 373.78 |
| rs9882532 | T | C | 0.64 | 0.012 | 0.002 | 2.15E-17 | -0.005 | 0.007 | 0.475 | 68.34 |
| rs9886703 | A | T | 0.17 | -0.014 | 0.002 | 4.46E-13 | -0.023 | 0.01 | 0.014 | 51.24 |
| rs9927137 | A | G | 0.53 | 0.008 | 0.001 | 5.11E-09 | 0 | 0.007 | 0.955 | 34.31 |
| rs9927842 | T | C | 0.15 | -0.013 | 0.002 | 6.22E-11 | -0.01 | 0.01 | 0.299 | 42.25 |
| rs9929556 | T | G | 0.57 | -0.01 | 0.001 | 2.31E-12 | 0.011 | 0.007 | 0.136 | 50.01 |
| rs9929762 | A | G | 0.56 | 0.01 | 0.001 | 1.03E-11 | 0 | 0.007 | 0.996 | 47.02 |
| rs9933256 | A | G | 0.56 | 0.012 | 0.001 | 6.06E-17 | -0.005 | 0.007 | 0.468 | 72.25 |

**SNPs used as instrumental variables in the MR analysis of educational attainment on volume of left hippocampus**

| **SNP** | **Effect allele** | **Other allele** | **Effect allele frequency** | **beta (exposure)** | **SE (exposure)** | ***P* (exposure)** | **beta (outcome)** | **SE (outcome)** | ***P* (outcome)** | ***F*-statistic** |
| --- | --- | --- | --- | --- | --- | --- | --- | --- | --- | --- |
| rs10006235 | T | C | 0.27 | -0.009 | 0.002 | 4.48E-09 | -0.001 | 0.009 | 0.881 | 33.79 |
| rs10021733 | T | C | 0.15 | -0.016 | 0.003 | 9.45E-10 | 0 | 0.011 | 0.975 | 36.89 |
| rs10046069 | A | G | 0.12 | 0.013 | 0.002 | 4.40E-09 | -0.013 | 0.012 | 0.285 | 33.32 |
| rs10060023 | T | C | 0.33 | 0.011 | 0.002 | 1.88E-13 | -0.002 | 0.008 | 0.807 | 53.78 |
| rs10080647 | A | C | 0.14 | 0.012 | 0.002 | 7.05E-10 | -0.013 | 0.011 | 0.237 | 38.44 |
| rs1008078 | T | C | 0.4 | -0.018 | 0.001 | 4.50E-37 | 0.002 | 0.008 | 0.814 | 169 |
| rs10189857 | A | G | 0.57 | 0.016 | 0.001 | 4.69E-29 | 0.02 | 0.008 | 0.011 | 127.37 |
| rs10193498 | A | T | 0.76 | 0.01 | 0.002 | 3.10E-09 | -0.009 | 0.009 | 0.322 | 36.75 |
| rs10215082 | A | G | 0.47 | -0.015 | 0.001 | 7.03E-26 | 0.011 | 0.008 | 0.141 | 113.27 |
| rs10460095 | A | G | 0.57 | -0.012 | 0.001 | 2.61E-18 | -0.009 | 0.008 | 0.275 | 78.45 |
| rs10761251 | A | T | 0.66 | 0.011 | 0.002 | 2.52E-14 | 0.032 | 0.008 | 1.16E-04 | 56.75 |
| rs10772644 | C | G | 0.89 | 0.014 | 0.002 | 1.69E-10 | 0.008 | 0.013 | 0.536 | 41.66 |
| rs10773002 | A | T | 0.25 | 0.018 | 0.002 | 2.50E-30 | -0.005 | 0.009 | 0.565 | 133.69 |
| rs10773208 | T | C | 0.25 | -0.011 | 0.002 | 8.09E-12 | -0.005 | 0.009 | 0.549 | 47.27 |
| rs10798888 | T | G | 0.17 | -0.014 | 0.002 | 5.15E-14 | -0.002 | 0.01 | 0.828 | 54.29 |
| rs10799615 | A | G | 0.25 | -0.009 | 0.002 | 4.11E-08 | -0.004 | 0.009 | 0.621 | 30.94 |
| rs10805383 | A | G | 0.49 | -0.01 | 0.001 | 2.22E-13 | -0.019 | 0.008 | 0.017 | 54.13 |
| rs10830858 | T | C | 0.51 | 0.009 | 0.001 | 5.31E-11 | -0.005 | 0.008 | 0.52 | 43.18 |
| rs10844179 | A | G | 0.23 | 0.01 | 0.002 | 5.03E-10 | 0.026 | 0.009 | 0.005 | 37.43 |
| rs10875121 | C | G | 0.84 | 0.017 | 0.002 | 4.46E-20 | 0.006 | 0.011 | 0.573 | 83.87 |
| rs10892807 | T | C | 0.57 | 0.011 | 0.001 | 1.09E-14 | -0.013 | 0.008 | 0.104 | 60.62 |
| rs10940921 | T | G | 0.42 | 0.011 | 0.001 | 1.31E-14 | 0.018 | 0.008 | 0.019 | 62.86 |
| rs10951590 | T | C | 0.33 | -0.011 | 0.002 | 3.62E-13 | -0.001 | 0.008 | 0.893 | 52.8 |
| rs10979613 | T | C | 0.64 | -0.012 | 0.002 | 1.25E-15 | 0.007 | 0.008 | 0.372 | 60.84 |
| rs10996167 | C | G | 0.65 | 0.011 | 0.002 | 7.47E-09 | 0.009 | 0.008 | 0.263 | 31.92 |
| rs11003463 | T | G | 0.61 | 0.009 | 0.002 | 5.58E-10 | -0.002 | 0.008 | 0.789 | 36.8 |
| rs11023764 | A | G | 0.64 | 0.009 | 0.002 | 5.32E-10 | 0.01 | 0.008 | 0.237 | 36.8 |
| rs11030102 | C | G | 0.75 | 0.01 | 0.002 | 4.60E-10 | 0.018 | 0.009 | 0.043 | 39.85 |
| rs11076962 | T | C | 0.72 | 0.009 | 0.002 | 4.29E-08 | -0.004 | 0.009 | 0.659 | 28.89 |
| rs11081529 | T | C | 0.71 | 0.013 | 0.002 | 5.22E-16 | -0.013 | 0.009 | 0.14 | 70.56 |
| rs11082011 | T | C | 0.67 | 0.019 | 0.002 | 2.06E-38 | -0.001 | 0.008 | 0.916 | 167.27 |
| rs11121177 | A | G | 0.18 | 0.015 | 0.002 | 1.44E-16 | 0.01 | 0.01 | 0.323 | 70.37 |
| rs11130380 | T | G | 0.61 | 0.009 | 0.001 | 4.17E-10 | -0.008 | 0.008 | 0.315 | 41.33 |
| rs11138947 | T | C | 0.72 | 0.011 | 0.002 | 2.36E-11 | 0.018 | 0.009 | 0.034 | 43.07 |
| rs11157931 | A | C | 0.39 | -0.014 | 0.001 | 3.86E-22 | -0.014 | 0.008 | 0.072 | 98.58 |
| rs111852224 | T | C | 0.12 | 0.016 | 0.002 | 4.85E-13 | 0.01 | 0.012 | 0.389 | 50.28 |
| rs11211123 | A | G | 0.22 | 0.01 | 0.002 | 7.62E-10 | -0.021 | 0.009 | 0.024 | 37.43 |
| rs11213482 | A | G | 0.84 | -0.011 | 0.002 | 2.68E-08 | 0.004 | 0.01 | 0.692 | 31.12 |
| rs112603734 | A | C | 0.77 | -0.013 | 0.002 | 2.72E-09 | 0.001 | 0.009 | 0.884 | 35.46 |
| rs112806496 | C | G | 0.91 | -0.016 | 0.002 | 1.72E-10 | -0.013 | 0.013 | 0.338 | 40.96 |
| rs1128956 | T | G | 0.83 | -0.014 | 0.002 | 5.34E-14 | 0.02 | 0.01 | 0.05 | 54.29 |
| rs113520408 | A | G | 0.28 | 0.016 | 0.002 | 1.03E-22 | -0.001 | 0.009 | 0.902 | 93.85 |
| rs113615161 | T | C | 0.13 | -0.012 | 0.002 | 2.98E-09 | 0.002 | 0.012 | 0.856 | 34.31 |
| rs114593137 | A | T | 0.79 | -0.012 | 0.002 | 1.65E-12 | -0.01 | 0.01 | 0.275 | 51.5 |
| rs115000530 | A | T | 0.95 | -0.025 | 0.003 | 1.09E-15 | -0.006 | 0.017 | 0.722 | 65.04 |
| rs11542663 | A | C | 0.69 | 0.011 | 0.002 | 2.44E-13 | 0.003 | 0.009 | 0.701 | 55.75 |
| rs11588857 | A | G | 0.21 | 0.02 | 0.002 | 4.45E-31 | 0.003 | 0.01 | 0.733 | 137.03 |
| rs11598765 | A | G | 0.2 | -0.011 | 0.002 | 1.73E-09 | -0.011 | 0.01 | 0.263 | 34.68 |
| rs11620355 | A | G | 0.09 | 0.018 | 0.002 | 1.09E-13 | -0.001 | 0.014 | 0.936 | 53 |
| rs11657342 | A | G | 0.37 | 0.014 | 0.002 | 2.32E-13 | 0.009 | 0.008 | 0.243 | 54.29 |
| rs11657979 | A | G | 0.24 | -0.009 | 0.002 | 4.57E-08 | 0.006 | 0.009 | 0.528 | 28.65 |
| rs11663602 | A | C | 0.28 | -0.013 | 0.002 | 9.27E-17 | -0.012 | 0.009 | 0.161 | 66.02 |
| rs1167827 | A | G | 0.43 | 0.01 | 0.001 | 2.93E-13 | -0.002 | 0.008 | 0.804 | 55.18 |
| rs11678980 | A | G | 0.45 | -0.017 | 0.001 | 1.60E-31 | 0.004 | 0.008 | 0.584 | 140.59 |
| rs11724690 | T | G | 0.29 | 0.009 | 0.002 | 2.08E-09 | -0.013 | 0.009 | 0.126 | 38.44 |
| rs117398064 | C | G | 0.09 | -0.014 | 0.002 | 1.16E-08 | -0.007 | 0.013 | 0.604 | 31.81 |
| rs118134876 | T | C | 0.06 | -0.022 | 0.003 | 1.50E-13 | 0.012 | 0.016 | 0.475 | 55.25 |
| rs11871429 | A | G | 0.77 | 0.013 | 0.002 | 2.52E-14 | -0.001 | 0.009 | 0.954 | 56.69 |
| rs11894424 | A | C | 0.95 | 0.021 | 0.003 | 2.14E-10 | 0.006 | 0.019 | 0.739 | 40.11 |
| rs12054166 | C | G | 0.74 | 0.01 | 0.002 | 2.30E-09 | 0.013 | 0.009 | 0.16 | 36 |
| rs12076635 | C | G | 0.78 | 0.021 | 0.002 | 3.33E-34 | -0.007 | 0.009 | 0.451 | 148.27 |
| rs12113634 | T | C | 0.65 | 0.009 | 0.002 | 2.50E-09 | 0.003 | 0.008 | 0.73 | 34.42 |
| rs12151248 | T | C | 0.11 | -0.015 | 0.002 | 3.81E-11 | 0.016 | 0.012 | 0.197 | 41.97 |
| rs1220779 | A | G | 0.47 | -0.008 | 0.001 | 1.58E-08 | 0.001 | 0.008 | 0.935 | 31.84 |
| rs12273435 | A | G | 0.21 | -0.013 | 0.002 | 4.72E-14 | -0.011 | 0.01 | 0.252 | 60.29 |
| rs12332731 | A | T | 0.19 | 0.014 | 0.002 | 5.96E-14 | -0.013 | 0.01 | 0.17 | 56.25 |
| rs12375949 | T | C | 0.43 | -0.014 | 0.001 | 1.61E-22 | 0 | 0.008 | 0.963 | 97.16 |
| rs12438177 | A | G | 0.37 | 0.009 | 0.002 | 1.48E-10 | -0.007 | 0.008 | 0.368 | 38.44 |
| rs12468040 | T | G | 0.38 | 0.014 | 0.001 | 3.12E-21 | -0.004 | 0.008 | 0.614 | 95.76 |
| rs12503522 | T | C | 0.28 | -0.011 | 0.002 | 3.93E-13 | 0.001 | 0.009 | 0.937 | 49.88 |
| rs12506221 | T | G | 0.44 | -0.013 | 0.001 | 4.23E-20 | -0.006 | 0.008 | 0.456 | 86.22 |
| rs12519073 | T | C | 0.23 | -0.011 | 0.002 | 1.16E-10 | 0.007 | 0.009 | 0.443 | 39.62 |
| rs12524795 | T | C | 0.44 | 0.01 | 0.001 | 5.28E-13 | 0 | 0.008 | 0.964 | 53.08 |
| rs12571549 | A | G | 0.14 | 0.015 | 0.002 | 1.45E-13 | -0.006 | 0.011 | 0.617 | 57 |
| rs12574281 | A | C | 0.63 | -0.008 | 0.002 | 3.04E-08 | 0.01 | 0.008 | 0.218 | 28.44 |
| rs12591647 | T | C | 0.82 | -0.015 | 0.002 | 7.78E-17 | -0.015 | 0.01 | 0.13 | 71.31 |
| rs12602286 | T | G | 0.87 | 0.016 | 0.002 | 4.62E-14 | -0.013 | 0.012 | 0.272 | 58.05 |
| rs12614263 | A | G | 0.5 | 0.008 | 0.001 | 3.38E-09 | 0.001 | 0.008 | 0.921 | 35.15 |
| rs12638072 | A | G | 0.3 | -0.011 | 0.002 | 3.13E-12 | -0.005 | 0.009 | 0.565 | 50.88 |
| rs12643771 | T | C | 0.31 | 0.013 | 0.002 | 1.32E-17 | -0.012 | 0.008 | 0.139 | 75.11 |
| rs12646523 | T | C | 0.25 | -0.013 | 0.002 | 3.79E-16 | 0.013 | 0.009 | 0.147 | 68.06 |
| rs12670376 | A | G | 0.45 | 0.009 | 0.001 | 5.99E-11 | -0.009 | 0.008 | 0.257 | 43.18 |
| rs12761761 | T | C | 0.24 | 0.015 | 0.002 | 4.79E-20 | 0.014 | 0.009 | 0.113 | 81 |
| rs12765185 | A | T | 0.28 | -0.009 | 0.002 | 2.96E-08 | 0.003 | 0.009 | 0.72 | 30.25 |
| rs12789313 | T | C | 0.49 | 0.009 | 0.001 | 6.79E-11 | 0.005 | 0.008 | 0.485 | 43.18 |
| rs12875339 | A | C | 0.64 | -0.011 | 0.002 | 1.71E-13 | 0.011 | 0.008 | 0.181 | 51.84 |
| rs12888615 | T | C | 0.2 | 0.012 | 0.002 | 9.42E-12 | -0.002 | 0.01 | 0.845 | 44.44 |
| rs12957463 | A | G | 0.8 | 0.015 | 0.002 | 9.20E-17 | -0.009 | 0.01 | 0.354 | 65.79 |
| rs12967010 | T | C | 0.77 | 0.011 | 0.002 | 1.33E-11 | 0.022 | 0.009 | 0.018 | 44.97 |
| rs12981405 | T | C | 0.17 | -0.011 | 0.002 | 2.75E-09 | 0.024 | 0.01 | 0.023 | 34.75 |
| rs1301838 | T | C | 0.32 | -0.01 | 0.002 | 1.29E-10 | 0.006 | 0.008 | 0.458 | 40.96 |
| rs13018640 | T | C | 0.6 | -0.021 | 0.001 | 1.05E-50 | 0.001 | 0.008 | 0.881 | 235.84 |
| rs13050131 | A | C | 0.66 | -0.008 | 0.002 | 4.40E-08 | 0.001 | 0.008 | 0.935 | 29.16 |
| rs13133213 | A | G | 0.5 | 0.01 | 0.001 | 7.57E-12 | -0.005 | 0.008 | 0.543 | 47.02 |
| rs13145650 | T | C | 0.92 | -0.017 | 0.002 | 8.99E-12 | -0.014 | 0.014 | 0.299 | 47.89 |
| rs13163845 | T | C | 0.85 | -0.015 | 0.002 | 4.66E-14 | 0.011 | 0.011 | 0.304 | 55.5 |
| rs13197257 | T | G | 0.28 | 0.011 | 0.002 | 2.83E-12 | 0.001 | 0.009 | 0.909 | 46.41 |
| rs13212041 | T | C | 0.8 | 0.01 | 0.002 | 7.42E-09 | 0.01 | 0.01 | 0.275 | 31.48 |
| rs13240401 | T | C | 0.78 | 0.018 | 0.002 | 2.36E-24 | 0.013 | 0.009 | 0.171 | 107.18 |
| rs13261773 | C | G | 0.16 | 0.012 | 0.002 | 3.12E-10 | 0.025 | 0.011 | 0.021 | 41.23 |
| rs13266287 | A | T | 0.67 | -0.009 | 0.002 | 1.75E-08 | 0.009 | 0.008 | 0.279 | 32.11 |
| rs1329125 | T | C | 0.33 | -0.01 | 0.002 | 8.37E-11 | 0.008 | 0.008 | 0.34 | 41.82 |
| rs13327482 | A | G | 0.82 | -0.012 | 0.002 | 4.47E-11 | -0.001 | 0.01 | 0.952 | 45.19 |
| rs1334297 | A | G | 0.74 | 0.026 | 0.002 | 2.53E-58 | -0.002 | 0.009 | 0.813 | 258 |
| rs13422673 | T | C | 0.47 | -0.011 | 0.001 | 6.48E-15 | 0.009 | 0.008 | 0.25 | 61.73 |
| rs137079 | T | C | 0.14 | 0.013 | 0.002 | 3.06E-10 | 0.001 | 0.011 | 0.948 | 40.96 |
| rs1426619 | T | C | 0.43 | 0.01 | 0.001 | 1.40E-11 | -0.01 | 0.008 | 0.219 | 47.02 |
| rs1434630 | T | G | 0.15 | -0.014 | 0.002 | 4.63E-12 | -0.011 | 0.011 | 0.317 | 46.92 |
| rs143743568 | A | G | 0.15 | 0.012 | 0.002 | 4.99E-09 | -0.006 | 0.011 | 0.559 | 34.81 |
| rs150537577 | A | G | 0.92 | -0.015 | 0.003 | 9.50E-09 | -0.007 | 0.015 | 0.64 | 33.73 |
| rs1505676 | C | G | 0.63 | 0.008 | 0.002 | 6.67E-09 | 0.016 | 0.008 | 0.049 | 31.36 |
| rs1542354 | A | G | 0.48 | -0.008 | 0.001 | 2.72E-09 | 0.001 | 0.008 | 0.876 | 36 |
| rs1544 | A | G | 0.26 | -0.01 | 0.002 | 1.76E-09 | 0.011 | 0.009 | 0.218 | 36 |
| rs1564347 | T | G | 0.34 | 0.009 | 0.002 | 2.39E-09 | 0.009 | 0.008 | 0.276 | 34.42 |
| rs1603460 | T | G | 0.42 | 0.009 | 0.001 | 2.20E-10 | -0.002 | 0.008 | 0.818 | 41.33 |
| rs162445 | A | G | 0.08 | 0.015 | 0.003 | 5.72E-09 | 0.003 | 0.015 | 0.838 | 34.18 |
| rs1689510 | C | G | 0.33 | 0.019 | 0.002 | 1.35E-38 | -0.012 | 0.008 | 0.151 | 167.27 |
| rs17048855 | A | G | 0.34 | 0.011 | 0.002 | 3.10E-13 | 0.006 | 0.008 | 0.491 | 51.84 |
| rs17110109 | T | C | 0.62 | -0.009 | 0.001 | 1.47E-10 | -0.006 | 0.008 | 0.47 | 44.13 |
| rs17144467 | A | C | 0.69 | 0.01 | 0.002 | 8.74E-11 | -0.002 | 0.008 | 0.779 | 42.68 |
| rs17148998 | A | G | 0.2 | 0.01 | 0.002 | 6.98E-09 | -0.005 | 0.01 | 0.601 | 35.3 |
| rs1717204 | A | C | 0.18 | -0.012 | 0.002 | 1.20E-10 | 0.008 | 0.01 | 0.425 | 42.25 |
| rs17248751 | A | G | 0.78 | -0.014 | 0.002 | 1.28E-15 | 0.007 | 0.009 | 0.471 | 64 |
| rs173003 | A | C | 0.49 | -0.008 | 0.001 | 5.50E-09 | 0.007 | 0.008 | 0.361 | 34.31 |
| rs1738050 | C | G | 0.62 | -0.009 | 0.001 | 1.42E-10 | -0.01 | 0.008 | 0.193 | 44.13 |
| rs17411339 | A | G | 0.56 | 0.014 | 0.001 | 5.10E-23 | 0.008 | 0.008 | 0.306 | 100 |
| rs17428076 | C | G | 0.76 | 0.014 | 0.002 | 6.41E-17 | 0.001 | 0.009 | 0.951 | 73.32 |
| rs1747714 | T | C | 0.53 | 0.011 | 0.001 | 3.82E-14 | 0.008 | 0.008 | 0.293 | 57.33 |
| rs1747817 | T | C | 0.76 | -0.009 | 0.002 | 2.29E-08 | 0.004 | 0.009 | 0.65 | 33.06 |
| rs17489649 | A | G | 0.68 | 0.013 | 0.002 | 2.77E-17 | -0.001 | 0.008 | 0.918 | 71.68 |
| rs17502934 | T | G | 0.15 | -0.017 | 0.002 | 2.12E-17 | 0.025 | 0.011 | 0.022 | 69.72 |
| rs175325 | A | T | 0.6 | -0.01 | 0.001 | 2.24E-12 | 0.001 | 0.008 | 0.882 | 51.02 |
| rs17551064 | A | G | 0.84 | 0.014 | 0.002 | 1.53E-13 | 0.002 | 0.011 | 0.843 | 55.07 |
| rs17563464 | A | C | 0.22 | -0.015 | 0.002 | 1.74E-17 | 0.019 | 0.009 | 0.044 | 74.77 |
| rs17565975 | A | G | 0.56 | -0.011 | 0.001 | 5.54E-14 | 0.012 | 0.008 | 0.112 | 57.33 |
| rs17598675 | T | C | 0.51 | -0.012 | 0.001 | 1.37E-16 | -0.017 | 0.008 | 0.029 | 68.65 |
| rs17604349 | A | G | 0.19 | 0.013 | 0.002 | 2.36E-12 | 0 | 0.01 | 0.973 | 49.78 |
| rs176218 | T | G | 0.2 | 0.02 | 0.002 | 1.10E-29 | -0.006 | 0.01 | 0.564 | 123.46 |
| rs17732878 | T | C | 0.79 | -0.01 | 0.002 | 4.06E-08 | -0.032 | 0.009 | 7.48E-04 | 31.23 |
| rs178183 | T | C | 0.25 | 0.015 | 0.002 | 4.81E-19 | 0.003 | 0.009 | 0.751 | 82.13 |
| rs17882802 | A | G | 0.43 | 0.008 | 0.001 | 3.00E-09 | 0.012 | 0.008 | 0.112 | 36 |
| rs182902112 | A | C | 0.02 | -0.03 | 0.005 | 1.17E-08 | 0.076 | 0.029 | 0.008 | 32.04 |
| rs1842713 | A | G | 0.79 | -0.012 | 0.002 | 9.67E-12 | 0.007 | 0.009 | 0.464 | 47.37 |
| rs1861786 | A | G | 0.38 | -0.008 | 0.001 | 2.39E-08 | -0.001 | 0.008 | 0.942 | 33.47 |
| rs1865955 | T | C | 0.82 | 0.012 | 0.002 | 4.06E-11 | 0.011 | 0.01 | 0.274 | 41.23 |
| rs1866823 | A | G | 0.54 | 0.011 | 0.001 | 3.09E-14 | 0.006 | 0.008 | 0.485 | 58.41 |
| rs1890132 | T | C | 0.71 | -0.009 | 0.002 | 6.56E-09 | -0.005 | 0.009 | 0.566 | 35.2 |
| rs192436652 | T | C | 0.03 | -0.032 | 0.004 | 2.06E-12 | 0.053 | 0.024 | 0.028 | 50.25 |
| rs1933264 | T | C | 0.75 | 0.01 | 0.002 | 5.85E-10 | -0.004 | 0.009 | 0.67 | 39.06 |
| rs1955250 | A | C | 0.91 | -0.014 | 0.002 | 2.26E-08 | -0.011 | 0.014 | 0.429 | 31.36 |
| rs1991585 | T | C | 0.71 | -0.01 | 0.002 | 6.26E-11 | 0.005 | 0.009 | 0.535 | 45.34 |
| rs2007655 | T | G | 0.5 | 0.009 | 0.001 | 8.64E-10 | -0.012 | 0.008 | 0.12 | 37.73 |
| rs2011603 | A | G | 0.74 | -0.009 | 0.002 | 1.12E-08 | 0.006 | 0.009 | 0.486 | 32.35 |
| rs2034631 | T | C | 0.65 | 0.009 | 0.002 | 2.64E-09 | -0.001 | 0.008 | 0.913 | 35.2 |
| rs2052285 | A | G | 0.6 | 0.011 | 0.001 | 7.25E-15 | -0.011 | 0.008 | 0.155 | 64 |
| rs2055940 | A | G | 0.32 | 0.008 | 0.002 | 4.95E-08 | 0.01 | 0.008 | 0.213 | 29.88 |
| rs2081652 | A | T | 0.66 | 0.012 | 0.002 | 1.09E-16 | 0.008 | 0.008 | 0.325 | 67.24 |
| rs2131167 | A | G | 0.45 | 0.008 | 0.001 | 1.52E-08 | -0.003 | 0.008 | 0.711 | 32.65 |
| rs2179152 | T | C | 0.37 | -0.013 | 0.002 | 1.68E-19 | 0 | 0.008 | 0.954 | 76.27 |
| rs2183271 | T | C | 0.64 | 0.008 | 0.002 | 1.91E-08 | -0.009 | 0.008 | 0.252 | 31.36 |
| rs2199409 | T | C | 0.8 | 0.01 | 0.002 | 3.14E-09 | -0.004 | 0.01 | 0.692 | 36.71 |
| rs2212430 | T | C | 0.3 | -0.01 | 0.002 | 1.32E-10 | -0.004 | 0.008 | 0.641 | 39.06 |
| rs2216144 | T | C | 0.53 | 0.01 | 0.001 | 2.58E-12 | 0.005 | 0.008 | 0.493 | 49 |
| rs2256965 | A | G | 0.42 | 0.011 | 0.001 | 2.40E-13 | 0.004 | 0.008 | 0.594 | 57.33 |
| rs2287838 | A | G | 0.55 | -0.01 | 0.001 | 1.27E-13 | 0.002 | 0.008 | 0.773 | 55.18 |
| rs2290601 | T | C | 0.77 | -0.015 | 0.002 | 4.49E-19 | -0.005 | 0.009 | 0.584 | 76.82 |
| rs2297293 | C | G | 0.31 | 0.01 | 0.002 | 8.80E-11 | 0.011 | 0.008 | 0.186 | 43.56 |
| rs232496 | T | C | 0.37 | 0.009 | 0.002 | 7.14E-10 | 0.002 | 0.008 | 0.764 | 36 |
| rs2364544 | A | G | 0.39 | -0.011 | 0.001 | 2.91E-13 | 0.017 | 0.008 | 0.032 | 56.25 |
| rs2434672 | A | C | 0.53 | 0.009 | 0.001 | 1.06E-10 | 0 | 0.008 | 0.989 | 42.25 |
| rs2436760 | T | C | 0.97 | 0.023 | 0.004 | 4.52E-08 | 0.043 | 0.023 | 0.058 | 30.51 |
| rs2447097 | T | G | 0.46 | 0.01 | 0.001 | 1.10E-11 | -0.002 | 0.008 | 0.847 | 47.02 |
| rs2469226 | A | T | 0.22 | 0.01 | 0.002 | 1.28E-08 | 0.035 | 0.009 | 1.49E-04 | 31.89 |
| rs2496482 | T | C | 0.42 | 0.012 | 0.002 | 9.20E-17 | -0.013 | 0.008 | 0.092 | 65.07 |
| rs252991 | A | G | 0.37 | 0.01 | 0.002 | 6.30E-11 | -0.006 | 0.008 | 0.445 | 40.11 |
| rs2545795 | A | C | 0.44 | 0.013 | 0.001 | 1.03E-18 | 0.019 | 0.008 | 0.015 | 79.72 |
| rs2554835 | A | G | 0.4 | 0.009 | 0.001 | 8.95E-10 | -0.01 | 0.008 | 0.204 | 39.51 |
| rs2570497 | T | C | 0.64 | -0.012 | 0.002 | 1.27E-16 | 0.014 | 0.008 | 0.088 | 65.07 |
| rs2725370 | T | C | 0.3 | -0.014 | 0.002 | 4.96E-20 | 0.012 | 0.008 | 0.159 | 88.36 |
| rs2764684 | T | C | 0.83 | 0.014 | 0.002 | 9.99E-15 | 0.006 | 0.01 | 0.565 | 56.65 |
| rs2838006 | T | C | 0.36 | 0.013 | 0.002 | 1.38E-19 | -0.007 | 0.008 | 0.382 | 77.44 |
| rs28513882 | A | G | 0.18 | -0.011 | 0.002 | 2.03E-09 | -0.009 | 0.01 | 0.374 | 37.35 |
| rs2885198 | A | G | 0.53 | 0.009 | 0.001 | 4.63E-10 | -0.008 | 0.008 | 0.312 | 39.51 |
| rs2923424 | A | G | 0.61 | 0.012 | 0.001 | 6.31E-17 | 0.02 | 0.008 | 0.014 | 73.47 |
| rs2929032 | A | G | 0.47 | -0.008 | 0.001 | 8.40E-09 | 0.002 | 0.008 | 0.801 | 33.47 |
| rs2958182 | A | T | 0.34 | 0.008 | 0.002 | 3.46E-08 | 0.013 | 0.008 | 0.115 | 29.88 |
| rs2964199 | T | C | 0.31 | -0.01 | 0.002 | 4.61E-11 | -0.001 | 0.008 | 0.95 | 43.56 |
| rs2964255 | A | G | 0.31 | 0.009 | 0.002 | 6.34E-09 | 0.006 | 0.008 | 0.508 | 34.42 |
| rs2998299 | T | G | 0.21 | -0.014 | 0.002 | 1.51E-15 | -0.006 | 0.01 | 0.517 | 64 |
| rs3026996 | A | C | 0.76 | 0.014 | 0.002 | 5.42E-17 | 0.009 | 0.009 | 0.332 | 74.39 |
| rs303752 | A | G | 0.41 | -0.011 | 0.001 | 1.47E-14 | 0.022 | 0.008 | 0.006 | 61.73 |
| rs3111251 | T | C | 0.43 | -0.009 | 0.001 | 3.88E-10 | 0.008 | 0.008 | 0.309 | 41.33 |
| rs34067381 | T | G | 0.36 | -0.01 | 0.002 | 9.77E-13 | -0.004 | 0.008 | 0.634 | 48.07 |
| rs34098770 | A | G | 0.15 | -0.016 | 0.002 | 2.95E-16 | -0.002 | 0.011 | 0.887 | 64 |
| rs34305371 | A | G | 0.1 | 0.031 | 0.002 | 1.10E-39 | -0.001 | 0.013 | 0.946 | 171.17 |
| rs34394051 | A | G | 0.84 | -0.012 | 0.002 | 2.30E-09 | -0.004 | 0.011 | 0.701 | 34.22 |
| rs34720381 | T | C | 0.09 | -0.018 | 0.002 | 1.90E-13 | 0.007 | 0.013 | 0.597 | 56.25 |
| rs34807077 | A | C | 0.16 | 0.013 | 0.002 | 3.29E-11 | -0.016 | 0.011 | 0.123 | 45.39 |
| rs35417702 | T | C | 0.52 | -0.015 | 0.001 | 2.40E-27 | -0.012 | 0.008 | 0.134 | 117.88 |
| rs35606437 | A | G | 0.27 | 0.011 | 0.002 | 3.98E-11 | 0.006 | 0.009 | 0.507 | 43.07 |
| rs35929923 | A | G | 0.25 | -0.011 | 0.002 | 1.77E-12 | 0.002 | 0.009 | 0.785 | 50.77 |
| rs363096 | T | C | 0.42 | -0.014 | 0.001 | 5.77E-24 | -0.006 | 0.008 | 0.452 | 104.33 |
| rs3751331 | A | G | 0.38 | -0.01 | 0.001 | 1.75E-11 | 0.008 | 0.008 | 0.31 | 48.01 |
| rs3809634 | A | G | 0.68 | -0.011 | 0.002 | 6.82E-13 | -0.01 | 0.008 | 0.244 | 52.8 |
| rs382196 | T | G | 0.38 | -0.008 | 0.001 | 1.47E-08 | -0.006 | 0.008 | 0.45 | 34.31 |
| rs3859523 | T | C | 0.82 | 0.012 | 0.002 | 9.92E-12 | 0.021 | 0.01 | 0.039 | 47.46 |
| rs3897821 | A | G | 0.67 | 0.015 | 0.002 | 3.58E-25 | -0.011 | 0.008 | 0.187 | 105.4 |
| rs3948495 | T | G | 0.41 | -0.009 | 0.001 | 3.95E-10 | -0.014 | 0.008 | 0.084 | 40.41 |
| rs401526 | T | C | 0.53 | -0.01 | 0.001 | 1.15E-13 | -0.004 | 0.008 | 0.571 | 55.18 |
| rs401966 | C | G | 0.39 | -0.01 | 0.001 | 1.72E-11 | -0.009 | 0.008 | 0.266 | 48.01 |
| rs42210 | C | G | 0.71 | -0.01 | 0.002 | 2.74E-10 | 0.008 | 0.009 | 0.352 | 37.52 |
| rs42302 | A | G | 0.34 | 0.009 | 0.002 | 5.96E-09 | 0.013 | 0.008 | 0.122 | 32.87 |
| rs4298514 | T | C | 0.7 | 0.01 | 0.002 | 1.07E-11 | -0.01 | 0.008 | 0.233 | 48.07 |
| rs4320563 | A | G | 0.51 | 0.011 | 0.001 | 2.24E-14 | 0.001 | 0.008 | 0.854 | 60.62 |
| rs4328757 | T | C | 0.61 | 0.01 | 0.001 | 2.95E-11 | -0.002 | 0.008 | 0.796 | 47.02 |
| rs4352658 | T | C | 0.08 | -0.019 | 0.003 | 1.71E-13 | 0.003 | 0.014 | 0.827 | 53.4 |
| rs4358081 | A | C | 0.53 | -0.01 | 0.001 | 1.28E-11 | 0.007 | 0.008 | 0.368 | 46.05 |
| rs4369924 | A | G | 0.16 | 0.013 | 0.002 | 5.58E-11 | 0.001 | 0.011 | 0.922 | 43.28 |
| rs4384309 | A | G | 0.46 | 0.01 | 0.001 | 4.89E-13 | -0.005 | 0.008 | 0.555 | 53.08 |
| rs4467547 | T | G | 0.41 | 0.012 | 0.001 | 4.88E-17 | 0.004 | 0.008 | 0.649 | 74.7 |
| rs4500930 | T | C | 0.34 | -0.011 | 0.002 | 1.67E-13 | 0.015 | 0.008 | 0.064 | 52.8 |
| rs4663617 | A | T | 0.23 | 0.01 | 0.002 | 4.12E-10 | -0.017 | 0.009 | 0.063 | 37.43 |
| rs4673840 | T | C | 0.84 | -0.014 | 0.002 | 5.86E-13 | -0.022 | 0.011 | 0.042 | 52.75 |
| rs4675248 | A | G | 0.4 | -0.009 | 0.001 | 4.50E-11 | 0.007 | 0.008 | 0.402 | 45.08 |
| rs4719944 | T | C | 0.55 | -0.012 | 0.001 | 9.25E-18 | 0.005 | 0.008 | 0.559 | 74.7 |
| rs4726070 | A | G | 0.6 | 0.012 | 0.001 | 1.36E-17 | -0.005 | 0.008 | 0.551 | 75.94 |
| rs4757957 | C | G | 0.69 | 0.013 | 0.002 | 1.79E-18 | -0.003 | 0.008 | 0.706 | 78.62 |
| rs4766424 | C | G | 0.12 | 0.014 | 0.002 | 2.47E-11 | 0.016 | 0.012 | 0.167 | 45.72 |
| rs4778058 | T | C | 0.5 | -0.009 | 0.001 | 4.92E-11 | 0.019 | 0.008 | 0.015 | 43.18 |
| rs4787457 | A | G | 0.63 | 0.016 | 0.002 | 1.23E-28 | 0.004 | 0.008 | 0.584 | 116.64 |
| rs4793090 | A | G | 0.66 | 0.008 | 0.002 | 1.67E-08 | 0 | 0.008 | 0.991 | 31.36 |
| rs4810894 | A | G | 0.37 | -0.009 | 0.002 | 4.38E-09 | -0.01 | 0.008 | 0.221 | 32.11 |
| rs4839155 | T | G | 0.77 | 0.012 | 0.002 | 4.66E-12 | 0.016 | 0.009 | 0.09 | 45.76 |
| rs4846724 | A | G | 0.53 | 0.01 | 0.001 | 2.89E-12 | 0.004 | 0.008 | 0.603 | 49 |
| rs4848924 | A | C | 0.71 | -0.012 | 0.002 | 3.56E-14 | 0.003 | 0.009 | 0.743 | 60.84 |
| rs488476 | C | G | 0.62 | -0.011 | 0.002 | 8.29E-15 | 0 | 0.008 | 0.952 | 56.75 |
| rs4895650 | T | C | 0.55 | 0.009 | 0.001 | 5.31E-11 | -0.001 | 0.008 | 0.925 | 44.13 |
| rs4899012 | C | G | 0.6 | -0.01 | 0.001 | 8.06E-13 | 0.011 | 0.008 | 0.161 | 54.13 |
| rs4904523 | A | G | 0.52 | -0.008 | 0.001 | 6.27E-09 | -0.011 | 0.008 | 0.172 | 34.31 |
| rs4915735 | A | G | 0.14 | 0.012 | 0.002 | 9.56E-09 | 0.015 | 0.011 | 0.188 | 33.64 |
| rs4977885 | A | G | 0.4 | -0.008 | 0.001 | 6.78E-09 | -0.016 | 0.008 | 0.04 | 36 |
| rs4984541 | A | G | 0.77 | -0.014 | 0.002 | 3.40E-16 | -0.012 | 0.009 | 0.201 | 65.9 |
| rs55736314 | C | G | 0.6 | -0.015 | 0.001 | 4.59E-27 | -0.019 | 0.008 | 0.018 | 121 |
| rs56099375 | T | C | 0.24 | 0.012 | 0.002 | 3.10E-13 | -0.001 | 0.009 | 0.956 | 49.83 |
| rs56319902 | T | C | 0.22 | -0.021 | 0.002 | 5.81E-33 | -0.021 | 0.009 | 0.027 | 146.84 |
| rs56391344 | A | G | 0.25 | 0.015 | 0.002 | 7.49E-20 | -0.011 | 0.009 | 0.227 | 85.56 |
| rs56794817 | A | G | 0.16 | 0.012 | 0.002 | 1.69E-10 | -0.017 | 0.011 | 0.113 | 41.23 |
| rs57349798 | A | G | 0.41 | 0.009 | 0.001 | 8.63E-11 | 0 | 0.008 | 0.974 | 44.13 |
| rs57352738 | A | T | 0.2 | -0.015 | 0.002 | 2.51E-18 | -0.019 | 0.01 | 0.047 | 79.94 |
| rs575113 | A | G | 0.29 | 0.013 | 0.002 | 7.52E-18 | -0.005 | 0.009 | 0.576 | 78.62 |
| rs5754753 | T | C | 0.72 | -0.013 | 0.002 | 6.15E-16 | 0.004 | 0.009 | 0.663 | 62.02 |
| rs57661533 | T | C | 0.13 | -0.011 | 0.002 | 3.24E-08 | -0.015 | 0.012 | 0.202 | 29.47 |
| rs580652 | T | C | 0.9 | -0.015 | 0.002 | 4.65E-10 | 0.034 | 0.013 | 0.009 | 37.52 |
| rs590013 | T | C | 0.68 | 0.01 | 0.002 | 2.47E-11 | 0.014 | 0.008 | 0.098 | 44.44 |
| rs6043521 | T | C | 0.6 | -0.008 | 0.001 | 9.72E-09 | 0.004 | 0.008 | 0.623 | 35.15 |
| rs6065080 | T | C | 0.36 | -0.013 | 0.002 | 7.49E-19 | 0.01 | 0.008 | 0.239 | 75.11 |
| rs6065784 | C | G | 0.7 | 0.011 | 0.002 | 5.78E-13 | 0 | 0.009 | 0.956 | 53.78 |
| rs61104616 | A | G | 0.52 | -0.017 | 0.001 | 1.56E-34 | -0.002 | 0.008 | 0.806 | 150.94 |
| rs6123924 | A | G | 0.84 | 0.013 | 0.002 | 3.20E-11 | -0.002 | 0.011 | 0.881 | 46.1 |
| rs61527214 | A | G | 0.41 | 0.011 | 0.001 | 1.13E-14 | 0.005 | 0.008 | 0.559 | 61.73 |
| rs61997667 | T | C | 0.15 | -0.014 | 0.002 | 2.24E-13 | 0.02 | 0.011 | 0.069 | 51.84 |
| rs62155350 | A | G | 0.02 | 0.034 | 0.005 | 1.69E-10 | -0.011 | 0.027 | 0.698 | 40.67 |
| rs62155873 | T | C | 0.12 | -0.015 | 0.002 | 1.65E-12 | 0.003 | 0.012 | 0.794 | 51.7 |
| rs62174974 | A | G | 0.19 | -0.011 | 0.002 | 2.76E-09 | -0.009 | 0.01 | 0.375 | 34.03 |
| rs62179650 | A | G | 0.3 | 0.011 | 0.002 | 5.66E-13 | -0.012 | 0.008 | 0.146 | 49 |
| rs62182994 | T | C | 0.69 | 0.014 | 0.002 | 2.55E-19 | -0.005 | 0.008 | 0.572 | 83.42 |
| rs62379838 | T | C | 0.7 | 0.012 | 0.002 | 1.77E-15 | 0.013 | 0.008 | 0.133 | 65.07 |
| rs628993 | A | G | 0.11 | 0.013 | 0.002 | 3.83E-08 | -0.003 | 0.013 | 0.828 | 30.97 |
| rs6449503 | A | G | 0.5 | 0.018 | 0.001 | 1.84E-39 | -0.002 | 0.008 | 0.829 | 174.62 |
| rs6490618 | T | C | 0.33 | 0.009 | 0.002 | 3.48E-09 | -0.009 | 0.008 | 0.255 | 34.42 |
| rs6493265 | T | C | 0.39 | -0.012 | 0.001 | 2.26E-17 | 0.004 | 0.008 | 0.594 | 75.94 |
| rs6557171 | T | C | 0.32 | -0.016 | 0.002 | 5.48E-25 | 0.019 | 0.008 | 0.02 | 106.78 |
| rs660001 | A | G | 0.21 | -0.016 | 0.002 | 1.66E-21 | 0.01 | 0.009 | 0.286 | 93.07 |
| rs66568921 | T | G | 0.64 | -0.018 | 0.002 | 7.60E-34 | 0 | 0.008 | 0.997 | 145.6 |
| rs66721975 | A | G | 0.29 | -0.009 | 0.002 | 1.51E-08 | 0.001 | 0.009 | 0.888 | 33.64 |
| rs6690195 | T | C | 0.49 | -0.011 | 0.001 | 7.89E-16 | 0.005 | 0.008 | 0.509 | 65.15 |
| rs6697584 | T | C | 0.79 | -0.013 | 0.002 | 1.43E-13 | -0.012 | 0.009 | 0.201 | 54.93 |
| rs6704768 | A | G | 0.56 | -0.012 | 0.001 | 1.84E-16 | 0.007 | 0.008 | 0.35 | 68.65 |
| rs6731373 | A | G | 0.34 | -0.012 | 0.002 | 1.39E-14 | 0.01 | 0.008 | 0.228 | 58.78 |
| rs6812533 | T | C | 0.74 | 0.011 | 0.002 | 4.85E-11 | -0.016 | 0.009 | 0.068 | 43.89 |
| rs68145588 | T | G | 0.14 | -0.013 | 0.002 | 7.53E-10 | -0.004 | 0.011 | 0.692 | 36.57 |
| rs6917154 | T | C | 0.12 | 0.013 | 0.002 | 2.55E-09 | 0.005 | 0.012 | 0.659 | 36.57 |
| rs6924023 | A | G | 0.75 | 0.011 | 0.002 | 5.17E-11 | -0.003 | 0.009 | 0.738 | 44.72 |
| rs6977237 | T | C | 0.47 | 0.008 | 0.001 | 5.59E-09 | -0.002 | 0.008 | 0.783 | 34.31 |
| rs6994287 | A | G | 0.41 | 0.01 | 0.001 | 1.11E-12 | -0.003 | 0.008 | 0.729 | 53.08 |
| rs7012546 | T | C | 0.42 | 0.009 | 0.001 | 1.25E-10 | 0.004 | 0.008 | 0.6 | 43.18 |
| rs7029718 | A | G | 0.41 | 0.024 | 0.001 | 7.82E-65 | 0.01 | 0.008 | 0.191 | 303.76 |
| rs7041702 | A | G | 0.74 | -0.011 | 0.002 | 4.53E-12 | -0.003 | 0.009 | 0.751 | 48.13 |
| rs7167688 | T | C | 0.49 | -0.009 | 0.001 | 1.71E-11 | -0.013 | 0.008 | 0.096 | 45.08 |
| rs7171405 | A | G | 0.24 | -0.01 | 0.002 | 7.53E-10 | -0.005 | 0.009 | 0.602 | 39.85 |
| rs717996 | T | C | 0.6 | -0.012 | 0.001 | 1.89E-17 | -0.001 | 0.008 | 0.949 | 75.94 |
| rs7226824 | T | C | 0.53 | -0.008 | 0.001 | 2.72E-08 | 0.01 | 0.008 | 0.203 | 31.04 |
| rs72486027 | T | C | 0.25 | -0.01 | 0.002 | 1.61E-10 | -0.012 | 0.009 | 0.173 | 42.25 |
| rs7254263 | T | C | 0.28 | -0.012 | 0.002 | 1.84E-14 | 0 | 0.009 | 0.96 | 55.32 |
| rs72622559 | T | C | 0.23 | -0.012 | 0.002 | 5.50E-13 | 0.012 | 0.01 | 0.228 | 49.83 |
| rs72672052 | A | T | 0.18 | -0.01 | 0.002 | 4.87E-08 | 0 | 0.01 | 0.98 | 31.48 |
| rs72709560 | A | G | 0.69 | 0.009 | 0.002 | 2.03E-08 | 0.012 | 0.008 | 0.149 | 32.11 |
| rs72828517 | T | C | 0.83 | -0.017 | 0.002 | 1.38E-19 | 0.006 | 0.01 | 0.57 | 78.18 |
| rs72829857 | A | G | 0.76 | -0.015 | 0.002 | 1.52E-19 | 0 | 0.009 | 0.989 | 86.72 |
| rs72917504 | T | C | 0.06 | -0.018 | 0.003 | 2.91E-09 | -0.008 | 0.017 | 0.639 | 35.2 |
| rs72944064 | T | C | 0.74 | -0.01 | 0.002 | 9.39E-10 | 0.003 | 0.009 | 0.765 | 37.52 |
| rs72962169 | T | C | 0.16 | -0.017 | 0.002 | 1.26E-19 | -0.001 | 0.011 | 0.958 | 83.87 |
| rs72993796 | T | C | 0.88 | 0.013 | 0.002 | 1.12E-08 | -0.011 | 0.012 | 0.368 | 33.32 |
| rs730384 | A | G | 0.44 | 0.01 | 0.001 | 9.38E-13 | 0.017 | 0.008 | 0.028 | 52.05 |
| rs73055556 | A | G | 0.14 | 0.011 | 0.002 | 4.95E-08 | -0.011 | 0.011 | 0.343 | 30.8 |
| rs73191311 | A | G | 0.34 | -0.009 | 0.002 | 2.03E-10 | -0.018 | 0.008 | 0.033 | 39.27 |
| rs7321274 | A | G | 0.8 | 0.011 | 0.002 | 8.21E-11 | 0.017 | 0.01 | 0.074 | 40.11 |
| rs7326331 | A | G | 0.28 | -0.013 | 0.002 | 3.42E-16 | -0.012 | 0.009 | 0.167 | 64 |
| rs73344830 | A | G | 0.42 | 0.017 | 0.001 | 1.00E-32 | 0.008 | 0.008 | 0.317 | 147.45 |
| rs736281 | T | C | 0.39 | 0.009 | 0.001 | 2.49E-09 | -0.001 | 0.008 | 0.922 | 37.73 |
| rs73648455 | T | C | 0.08 | -0.018 | 0.003 | 1.99E-12 | -0.012 | 0.015 | 0.401 | 49.54 |
| rs743316 | T | C | 0.79 | 0.011 | 0.002 | 8.21E-11 | 0.003 | 0.01 | 0.785 | 43.4 |
| rs74415461 | T | C | 0.08 | 0.018 | 0.003 | 6.25E-12 | 0.005 | 0.014 | 0.697 | 45.82 |
| rs7449561 | A | G | 0.23 | 0.01 | 0.002 | 9.73E-10 | -0.008 | 0.009 | 0.393 | 36 |
| rs7460106 | T | C | 0.76 | -0.012 | 0.002 | 2.06E-13 | 0.005 | 0.009 | 0.617 | 52.35 |
| rs746839 | C | G | 0.63 | 0.014 | 0.002 | 1.19E-20 | -0.002 | 0.008 | 0.821 | 81 |
| rs74787922 | A | C | 0.93 | 0.018 | 0.003 | 8.97E-11 | 0.018 | 0.016 | 0.236 | 42.25 |
| rs75033012 | C | G | 0.04 | -0.026 | 0.004 | 1.38E-11 | -0.02 | 0.02 | 0.308 | 46.1 |
| rs75177132 | T | C | 0.05 | 0.03 | 0.004 | 1.01E-16 | 0.006 | 0.018 | 0.744 | 68.52 |
| rs7575637 | A | G | 0.45 | 0.011 | 0.001 | 3.43E-14 | -0.001 | 0.008 | 0.903 | 58.41 |
| rs7603132 | A | G | 0.19 | 0.014 | 0.002 | 1.20E-14 | 0.01 | 0.01 | 0.314 | 57.93 |
| rs76076331 | T | C | 0.13 | 0.021 | 0.002 | 1.71E-23 | 0.013 | 0.012 | 0.279 | 96.23 |
| rs7617204 | A | G | 0.52 | -0.01 | 0.001 | 4.33E-12 | 0.003 | 0.008 | 0.713 | 48.01 |
| rs76235882 | A | G | 0.97 | 0.024 | 0.004 | 4.52E-10 | 0.029 | 0.024 | 0.21 | 38.19 |
| rs76246107 | A | G | 0.09 | -0.016 | 0.003 | 6.52E-10 | -0.005 | 0.015 | 0.752 | 38.82 |
| rs7650602 | T | C | 0.56 | -0.009 | 0.001 | 2.77E-10 | -0.015 | 0.008 | 0.052 | 40.41 |
| rs76577427 | C | G | 0.9 | 0.02 | 0.002 | 7.78E-18 | -0.001 | 0.013 | 0.961 | 71.54 |
| rs7672622 | A | G | 0.75 | -0.009 | 0.002 | 4.72E-08 | 0.03 | 0.009 | 7.17E-04 | 30.25 |
| rs7683416 | T | C | 0.46 | 0.014 | 0.001 | 6.05E-23 | 0.005 | 0.008 | 0.556 | 98.58 |
| rs76878669 | C | G | 0.76 | 0.014 | 0.002 | 3.10E-17 | 0.007 | 0.009 | 0.425 | 68.79 |
| rs7692359 | T | C | 0.78 | -0.011 | 0.002 | 1.40E-10 | -0.013 | 0.01 | 0.188 | 41.11 |
| rs77025239 | A | G | 0.16 | -0.014 | 0.002 | 4.65E-13 | 0.004 | 0.011 | 0.682 | 54.29 |
| rs77609760 | A | G | 0.93 | 0.024 | 0.003 | 7.71E-17 | 0.001 | 0.016 | 0.951 | 71.04 |
| rs77719387 | A | T | 0.02 | -0.04 | 0.006 | 3.78E-12 | -0.006 | 0.031 | 0.854 | 48.28 |
| rs7849487 | T | G | 0.66 | -0.017 | 0.002 | 1.25E-29 | -0.009 | 0.008 | 0.253 | 123.95 |
| rs78648104 | T | C | 0.91 | -0.014 | 0.002 | 1.70E-08 | -0.01 | 0.014 | 0.454 | 31.81 |
| rs7875078 | A | C | 0.46 | -0.008 | 0.001 | 2.72E-08 | 0.007 | 0.008 | 0.393 | 31.04 |
| rs7894722 | T | C | 0.37 | 0.009 | 0.001 | 5.41E-09 | 0.002 | 0.008 | 0.833 | 36.86 |
| rs790647 | A | C | 0.23 | -0.015 | 0.002 | 3.81E-18 | 0.002 | 0.009 | 0.853 | 72.75 |
| rs7910403 | T | G | 0.2 | 0.012 | 0.002 | 5.08E-12 | 0.003 | 0.01 | 0.746 | 45.94 |
| rs7924036 | T | G | 0.51 | 0.013 | 0.001 | 2.55E-21 | 0 | 0.008 | 0.974 | 90.25 |
| rs79265434 | A | G | 0.88 | -0.02 | 0.002 | 1.98E-19 | 0.015 | 0.012 | 0.208 | 80.18 |
| rs7972246 | T | C | 0.35 | 0.011 | 0.002 | 1.01E-13 | -0.006 | 0.008 | 0.484 | 53.78 |
| rs79728014 | A | G | 0.86 | -0.013 | 0.002 | 2.06E-11 | 0.014 | 0.011 | 0.206 | 44.89 |
| rs7974852 | A | C | 0.52 | 0.012 | 0.001 | 1.14E-16 | -0.008 | 0.008 | 0.319 | 69.84 |
| rs7977614 | A | G | 0.71 | -0.013 | 0.002 | 6.84E-17 | -0.009 | 0.009 | 0.312 | 70.14 |
| rs79994730 | T | C | 0.04 | 0.024 | 0.004 | 5.88E-12 | 0.001 | 0.018 | 0.961 | 47.02 |
| rs8008382 | T | C | 0.3 | -0.011 | 0.002 | 4.07E-12 | 0.01 | 0.009 | 0.226 | 49.94 |
| rs8024 | A | C | 0.33 | -0.012 | 0.002 | 1.13E-14 | -0.022 | 0.008 | 0.007 | 58.78 |
| rs80257979 | T | G | 0.97 | -0.025 | 0.004 | 3.31E-10 | -0.013 | 0.023 | 0.581 | 39.69 |
| rs8030487 | A | G | 0.69 | 0.009 | 0.002 | 2.52E-08 | 0.008 | 0.008 | 0.315 | 32.11 |
| rs806816 | A | T | 0.75 | 0.01 | 0.002 | 4.80E-10 | 0.005 | 0.009 | 0.61 | 39.85 |
| rs8097125 | T | C | 0.38 | 0.009 | 0.001 | 2.31E-09 | -0.021 | 0.008 | 0.008 | 37.73 |
| rs912883 | T | C | 0.68 | 0.009 | 0.002 | 6.20E-09 | 0.004 | 0.008 | 0.591 | 33.64 |
| rs9289300 | T | C | 0.84 | -0.016 | 0.002 | 1.00E-17 | 0.013 | 0.011 | 0.232 | 74.5 |
| rs929511 | T | C | 0.13 | -0.016 | 0.002 | 3.91E-14 | -0.006 | 0.012 | 0.621 | 58.05 |
| rs9359939 | A | C | 0.24 | -0.011 | 0.002 | 1.51E-10 | -0.005 | 0.009 | 0.557 | 43.07 |
| rs936496 | A | G | 0.62 | -0.008 | 0.001 | 4.38E-08 | -0.002 | 0.008 | 0.781 | 31.84 |
| rs9373363 | A | G | 0.75 | -0.011 | 0.002 | 5.54E-12 | -0.008 | 0.009 | 0.38 | 48.13 |
| rs9375188 | T | C | 0.48 | 0.021 | 0.001 | 8.78E-52 | -0.001 | 0.008 | 0.879 | 229.31 |
| rs9411331 | A | C | 0.68 | 0.015 | 0.002 | 7.63E-22 | 0.007 | 0.008 | 0.389 | 93.44 |
| rs9492774 | C | G | 0.37 | 0.008 | 0.002 | 4.33E-08 | 0.012 | 0.008 | 0.125 | 28.44 |
| rs9529146 | T | C | 0.23 | 0.013 | 0.002 | 3.48E-14 | -0.024 | 0.009 | 0.008 | 55.81 |
| rs9556958 | T | C | 0.52 | -0.011 | 0.001 | 5.96E-15 | -0.008 | 0.008 | 0.318 | 61.73 |
| rs9616947 | T | C | 0.38 | -0.013 | 0.002 | 3.58E-20 | 0.014 | 0.008 | 0.088 | 79.8 |
| rs9633970 | T | C | 0.76 | 0.009 | 0.002 | 2.81E-08 | 0.015 | 0.009 | 0.102 | 32.35 |
| rs9655780 | A | G | 0.83 | -0.015 | 0.002 | 3.70E-16 | 0.005 | 0.01 | 0.636 | 64.84 |
| rs969512 | A | T | 0.66 | -0.011 | 0.002 | 9.84E-13 | 0.011 | 0.008 | 0.167 | 49.94 |
| rs9853928 | T | C | 0.21 | -0.013 | 0.002 | 1.59E-13 | -0.001 | 0.01 | 0.954 | 55.81 |
| rs9859556 | T | G | 0.31 | 0.029 | 0.002 | 4.61E-82 | 0.016 | 0.008 | 0.058 | 373.78 |
| rs9882532 | T | C | 0.64 | 0.012 | 0.002 | 2.15E-17 | 0.006 | 0.008 | 0.458 | 68.34 |
| rs9886703 | A | T | 0.17 | -0.014 | 0.002 | 4.46E-13 | 0.003 | 0.01 | 0.796 | 51.24 |
| rs9927137 | A | G | 0.53 | 0.008 | 0.001 | 5.11E-09 | 0.005 | 0.008 | 0.483 | 34.31 |
| rs9927842 | T | C | 0.15 | -0.013 | 0.002 | 6.22E-11 | -0.022 | 0.011 | 0.037 | 42.25 |
| rs9929556 | T | G | 0.57 | -0.01 | 0.001 | 2.31E-12 | 0.009 | 0.008 | 0.259 | 50.01 |
| rs9929762 | A | G | 0.56 | 0.01 | 0.001 | 1.03E-11 | 0.01 | 0.008 | 0.218 | 47.02 |
| rs9933256 | A | G | 0.56 | 0.012 | 0.001 | 6.06E-17 | 0.001 | 0.008 | 0.865 | 72.25 |

**SNPs used as instrumental variables in the MR analysis of educational attainment on volume of right hippocampus**

| **SNP** | **Effect allele** | **Other allele** | **Effect allele frequency** | **beta (exposure)** | **SE (exposure)** | ***P* (exposure)** | **beta (outcome)** | **SE (outcome)** | ***P* (outcome)** | ***F*-statistic** |
| --- | --- | --- | --- | --- | --- | --- | --- | --- | --- | --- |
| rs10006235 | T | C | 0.27 | -0.009 | 0.002 | 4.48E-09 | -0.003 | 0.009 | 0.701 | 33.79 |
| rs10021733 | T | C | 0.15 | -0.016 | 0.003 | 9.45E-10 | 0.011 | 0.011 | 0.315 | 36.89 |
| rs10046069 | A | G | 0.12 | 0.013 | 0.002 | 4.40E-09 | 0.001 | 0.012 | 0.963 | 33.32 |
| rs10060023 | T | C | 0.33 | 0.011 | 0.002 | 1.88E-13 | -0.005 | 0.008 | 0.551 | 53.78 |
| rs10080647 | A | C | 0.14 | 0.012 | 0.002 | 7.05E-10 | -0.019 | 0.011 | 0.083 | 38.44 |
| rs1008078 | T | C | 0.4 | -0.018 | 0.001 | 4.50E-37 | -0.003 | 0.008 | 0.731 | 169 |
| rs10189857 | A | G | 0.57 | 0.016 | 0.001 | 4.69E-29 | 0.008 | 0.008 | 0.332 | 127.37 |
| rs10193498 | A | T | 0.76 | 0.01 | 0.002 | 3.10E-09 | -0.007 | 0.009 | 0.452 | 36.75 |
| rs10215082 | A | G | 0.47 | -0.015 | 0.001 | 7.03E-26 | 0.007 | 0.008 | 0.354 | 113.27 |
| rs10460095 | A | G | 0.57 | -0.012 | 0.001 | 2.61E-18 | 0.001 | 0.008 | 0.902 | 78.45 |
| rs10761251 | A | T | 0.66 | 0.011 | 0.002 | 2.52E-14 | 0.018 | 0.008 | 0.027 | 56.75 |
| rs10772644 | C | G | 0.89 | 0.014 | 0.002 | 1.69E-10 | 0.004 | 0.013 | 0.761 | 41.66 |
| rs10773002 | A | T | 0.25 | 0.018 | 0.002 | 2.50E-30 | 0.008 | 0.009 | 0.374 | 133.69 |
| rs10773208 | T | C | 0.25 | -0.011 | 0.002 | 8.09E-12 | -0.005 | 0.009 | 0.602 | 47.27 |
| rs10798888 | T | G | 0.17 | -0.014 | 0.002 | 5.15E-14 | 0.001 | 0.01 | 0.899 | 54.29 |
| rs10799615 | A | G | 0.25 | -0.009 | 0.002 | 4.11E-08 | -0.003 | 0.009 | 0.766 | 30.94 |
| rs10805383 | A | G | 0.49 | -0.01 | 0.001 | 2.22E-13 | -0.015 | 0.008 | 0.056 | 54.13 |
| rs10830858 | T | C | 0.51 | 0.009 | 0.001 | 5.31E-11 | -0.006 | 0.008 | 0.448 | 43.18 |
| rs10844179 | A | G | 0.23 | 0.01 | 0.002 | 5.03E-10 | 0.039 | 0.009 | 2.39E-05 | 37.43 |
| rs10875121 | C | G | 0.84 | 0.017 | 0.002 | 4.46E-20 | 0.012 | 0.011 | 0.253 | 83.87 |
| rs10892807 | T | C | 0.57 | 0.011 | 0.001 | 1.09E-14 | -0.02 | 0.008 | 0.013 | 60.62 |
| rs10940921 | T | G | 0.42 | 0.011 | 0.001 | 1.31E-14 | -0.007 | 0.008 | 0.401 | 62.86 |
| rs10951590 | T | C | 0.33 | -0.011 | 0.002 | 3.62E-13 | 0 | 0.008 | 0.959 | 52.8 |
| rs10979613 | T | C | 0.64 | -0.012 | 0.002 | 1.25E-15 | 0.006 | 0.008 | 0.428 | 60.84 |
| rs10996167 | C | G | 0.65 | 0.011 | 0.002 | 7.47E-09 | 0.005 | 0.008 | 0.503 | 31.92 |
| rs11003463 | T | G | 0.61 | 0.009 | 0.002 | 5.58E-10 | 0.002 | 0.008 | 0.757 | 36.8 |
| rs11023764 | A | G | 0.64 | 0.009 | 0.002 | 5.32E-10 | 0.023 | 0.008 | 0.005 | 36.8 |
| rs11030102 | C | G | 0.75 | 0.01 | 0.002 | 4.60E-10 | 0.017 | 0.009 | 0.054 | 39.85 |
| rs11076962 | T | C | 0.72 | 0.009 | 0.002 | 4.29E-08 | 0.001 | 0.009 | 0.928 | 28.89 |
| rs11081529 | T | C | 0.71 | 0.013 | 0.002 | 5.22E-16 | -0.004 | 0.009 | 0.675 | 70.56 |
| rs11082011 | T | C | 0.67 | 0.019 | 0.002 | 2.06E-38 | -0.003 | 0.008 | 0.733 | 167.27 |
| rs11121177 | A | G | 0.18 | 0.015 | 0.002 | 1.44E-16 | 0.006 | 0.01 | 0.545 | 70.37 |
| rs11130380 | T | G | 0.61 | 0.009 | 0.001 | 4.17E-10 | -0.013 | 0.008 | 0.093 | 41.33 |
| rs11138947 | T | C | 0.72 | 0.011 | 0.002 | 2.36E-11 | 0.006 | 0.009 | 0.484 | 43.07 |
| rs11157931 | A | C | 0.39 | -0.014 | 0.001 | 3.86E-22 | -0.016 | 0.008 | 0.05 | 98.58 |
| rs111852224 | T | C | 0.12 | 0.016 | 0.002 | 4.85E-13 | 0.006 | 0.012 | 0.58 | 50.28 |
| rs11211123 | A | G | 0.22 | 0.01 | 0.002 | 7.62E-10 | -0.018 | 0.009 | 0.052 | 37.43 |
| rs11213482 | A | G | 0.84 | -0.011 | 0.002 | 2.68E-08 | -0.001 | 0.01 | 0.908 | 31.12 |
| rs112603734 | A | C | 0.77 | -0.013 | 0.002 | 2.72E-09 | -0.016 | 0.009 | 0.072 | 35.46 |
| rs112806496 | C | G | 0.91 | -0.016 | 0.002 | 1.72E-10 | -0.007 | 0.013 | 0.606 | 40.96 |
| rs1128956 | T | G | 0.83 | -0.014 | 0.002 | 5.34E-14 | 0.012 | 0.01 | 0.238 | 54.29 |
| rs113520408 | A | G | 0.28 | 0.016 | 0.002 | 1.03E-22 | -0.003 | 0.009 | 0.7 | 93.85 |
| rs113615161 | T | C | 0.13 | -0.012 | 0.002 | 2.98E-09 | -0.011 | 0.012 | 0.357 | 34.31 |
| rs114593137 | A | T | 0.79 | -0.012 | 0.002 | 1.65E-12 | 0 | 0.01 | 0.985 | 51.5 |
| rs115000530 | A | T | 0.95 | -0.025 | 0.003 | 1.09E-15 | -0.036 | 0.017 | 0.033 | 65.04 |
| rs11542663 | A | C | 0.69 | 0.011 | 0.002 | 2.44E-13 | -0.007 | 0.009 | 0.392 | 55.75 |
| rs11588857 | A | G | 0.21 | 0.02 | 0.002 | 4.45E-31 | -0.002 | 0.01 | 0.803 | 137.03 |
| rs11598765 | A | G | 0.2 | -0.011 | 0.002 | 1.73E-09 | 0.003 | 0.01 | 0.793 | 34.68 |
| rs11620355 | A | G | 0.09 | 0.018 | 0.002 | 1.09E-13 | 0.001 | 0.014 | 0.958 | 53 |
| rs11657342 | A | G | 0.37 | 0.014 | 0.002 | 2.32E-13 | 0.005 | 0.008 | 0.505 | 54.29 |
| rs11657979 | A | G | 0.24 | -0.009 | 0.002 | 4.57E-08 | 0.001 | 0.009 | 0.934 | 28.65 |
| rs11663602 | A | C | 0.28 | -0.013 | 0.002 | 9.27E-17 | 0.002 | 0.009 | 0.832 | 66.02 |
| rs1167827 | A | G | 0.43 | 0.01 | 0.001 | 2.93E-13 | 0.012 | 0.008 | 0.133 | 55.18 |
| rs11678980 | A | G | 0.45 | -0.017 | 0.001 | 1.60E-31 | 0.015 | 0.008 | 0.063 | 140.59 |
| rs11724690 | T | G | 0.29 | 0.009 | 0.002 | 2.08E-09 | -0.001 | 0.009 | 0.912 | 38.44 |
| rs117398064 | C | G | 0.09 | -0.014 | 0.002 | 1.16E-08 | 0.003 | 0.013 | 0.814 | 31.81 |
| rs118134876 | T | C | 0.06 | -0.022 | 0.003 | 1.50E-13 | -0.012 | 0.016 | 0.461 | 55.25 |
| rs11871429 | A | G | 0.77 | 0.013 | 0.002 | 2.52E-14 | 0.006 | 0.009 | 0.486 | 56.69 |
| rs11894424 | A | C | 0.95 | 0.021 | 0.003 | 2.14E-10 | 0.014 | 0.019 | 0.475 | 40.11 |
| rs12054166 | C | G | 0.74 | 0.01 | 0.002 | 2.30E-09 | 0.012 | 0.009 | 0.178 | 36 |
| rs12076635 | C | G | 0.78 | 0.021 | 0.002 | 3.33E-34 | 0 | 0.009 | 0.964 | 148.27 |
| rs12113634 | T | C | 0.65 | 0.009 | 0.002 | 2.50E-09 | 0.007 | 0.008 | 0.389 | 34.42 |
| rs12151248 | T | C | 0.11 | -0.015 | 0.002 | 3.81E-11 | 0.013 | 0.012 | 0.278 | 41.97 |
| rs1220779 | A | G | 0.47 | -0.008 | 0.001 | 1.58E-08 | 0.01 | 0.008 | 0.216 | 31.84 |
| rs12273435 | A | G | 0.21 | -0.013 | 0.002 | 4.72E-14 | -0.003 | 0.01 | 0.746 | 60.29 |
| rs12332731 | A | T | 0.19 | 0.014 | 0.002 | 5.96E-14 | -0.017 | 0.01 | 0.087 | 56.25 |
| rs12375949 | T | C | 0.43 | -0.014 | 0.001 | 1.61E-22 | 0.001 | 0.008 | 0.896 | 97.16 |
| rs12438177 | A | G | 0.37 | 0.009 | 0.002 | 1.48E-10 | -0.012 | 0.008 | 0.131 | 38.44 |
| rs12468040 | T | G | 0.38 | 0.014 | 0.001 | 3.12E-21 | 0 | 0.008 | 0.982 | 95.76 |
| rs12503522 | T | C | 0.28 | -0.011 | 0.002 | 3.93E-13 | 0.003 | 0.009 | 0.738 | 49.88 |
| rs12506221 | T | G | 0.44 | -0.013 | 0.001 | 4.23E-20 | 0.004 | 0.008 | 0.605 | 86.22 |
| rs12519073 | T | C | 0.23 | -0.011 | 0.002 | 1.16E-10 | 0.002 | 0.009 | 0.844 | 39.62 |
| rs12524795 | T | C | 0.44 | 0.01 | 0.001 | 5.28E-13 | -0.002 | 0.008 | 0.826 | 53.08 |
| rs12571549 | A | G | 0.14 | 0.015 | 0.002 | 1.45E-13 | -0.015 | 0.011 | 0.173 | 57 |
| rs12574281 | A | C | 0.63 | -0.008 | 0.002 | 3.04E-08 | 0.007 | 0.008 | 0.364 | 28.44 |
| rs12591647 | T | C | 0.82 | -0.015 | 0.002 | 7.78E-17 | -0.005 | 0.01 | 0.602 | 71.31 |
| rs12602286 | T | G | 0.87 | 0.016 | 0.002 | 4.62E-14 | 0 | 0.012 | 0.978 | 58.05 |
| rs12614263 | A | G | 0.5 | 0.008 | 0.001 | 3.38E-09 | -0.001 | 0.008 | 0.848 | 35.15 |
| rs12638072 | A | G | 0.3 | -0.011 | 0.002 | 3.13E-12 | -0.016 | 0.009 | 0.071 | 50.88 |
| rs12643771 | T | C | 0.31 | 0.013 | 0.002 | 1.32E-17 | -0.002 | 0.008 | 0.824 | 75.11 |
| rs12646523 | T | C | 0.25 | -0.013 | 0.002 | 3.79E-16 | 0.003 | 0.009 | 0.704 | 68.06 |
| rs12670376 | A | G | 0.45 | 0.009 | 0.001 | 5.99E-11 | -0.011 | 0.008 | 0.163 | 43.18 |
| rs12761761 | T | C | 0.24 | 0.015 | 0.002 | 4.79E-20 | 0.006 | 0.009 | 0.506 | 81 |
| rs12765185 | A | T | 0.28 | -0.009 | 0.002 | 2.96E-08 | 0.001 | 0.009 | 0.895 | 30.25 |
| rs12789313 | T | C | 0.49 | 0.009 | 0.001 | 6.79E-11 | 0.018 | 0.008 | 0.018 | 43.18 |
| rs12875339 | A | C | 0.64 | -0.011 | 0.002 | 1.71E-13 | 0.01 | 0.008 | 0.214 | 51.84 |
| rs12888615 | T | C | 0.2 | 0.012 | 0.002 | 9.42E-12 | -0.008 | 0.01 | 0.426 | 44.44 |
| rs12957463 | A | G | 0.8 | 0.015 | 0.002 | 9.20E-17 | -0.012 | 0.01 | 0.219 | 65.79 |
| rs12967010 | T | C | 0.77 | 0.011 | 0.002 | 1.33E-11 | 0.007 | 0.009 | 0.459 | 44.97 |
| rs12981405 | T | C | 0.17 | -0.011 | 0.002 | 2.75E-09 | 0.023 | 0.01 | 0.027 | 34.75 |
| rs1301838 | T | C | 0.32 | -0.01 | 0.002 | 1.29E-10 | 0.002 | 0.008 | 0.767 | 40.96 |
| rs13018640 | T | C | 0.6 | -0.021 | 0.001 | 1.05E-50 | 0.002 | 0.008 | 0.843 | 235.84 |
| rs13050131 | A | C | 0.66 | -0.008 | 0.002 | 4.40E-08 | 0.007 | 0.008 | 0.37 | 29.16 |
| rs13133213 | A | G | 0.5 | 0.01 | 0.001 | 7.57E-12 | -0.001 | 0.008 | 0.884 | 47.02 |
| rs13145650 | T | C | 0.92 | -0.017 | 0.002 | 8.99E-12 | -0.007 | 0.014 | 0.614 | 47.89 |
| rs13163845 | T | C | 0.85 | -0.015 | 0.002 | 4.66E-14 | 0.005 | 0.011 | 0.678 | 55.5 |
| rs13197257 | T | G | 0.28 | 0.011 | 0.002 | 2.83E-12 | 0.004 | 0.009 | 0.668 | 46.41 |
| rs13212041 | T | C | 0.8 | 0.01 | 0.002 | 7.42E-09 | 0.004 | 0.01 | 0.665 | 31.48 |
| rs13240401 | T | C | 0.78 | 0.018 | 0.002 | 2.36E-24 | -0.003 | 0.009 | 0.777 | 107.18 |
| rs13261773 | C | G | 0.16 | 0.012 | 0.002 | 3.12E-10 | -0.002 | 0.011 | 0.831 | 41.23 |
| rs13266287 | A | T | 0.67 | -0.009 | 0.002 | 1.75E-08 | -0.002 | 0.008 | 0.805 | 32.11 |
| rs1329125 | T | C | 0.33 | -0.01 | 0.002 | 8.37E-11 | -0.014 | 0.008 | 0.105 | 41.82 |
| rs13327482 | A | G | 0.82 | -0.012 | 0.002 | 4.47E-11 | 0.014 | 0.01 | 0.182 | 45.19 |
| rs1334297 | A | G | 0.74 | 0.026 | 0.002 | 2.53E-58 | -0.009 | 0.009 | 0.286 | 258 |
| rs13422673 | T | C | 0.47 | -0.011 | 0.001 | 6.48E-15 | 0.001 | 0.008 | 0.854 | 61.73 |
| rs137079 | T | C | 0.14 | 0.013 | 0.002 | 3.06E-10 | 0.004 | 0.011 | 0.692 | 40.96 |
| rs1426619 | T | C | 0.43 | 0.01 | 0.001 | 1.40E-11 | -0.004 | 0.008 | 0.564 | 47.02 |
| rs1434630 | T | G | 0.15 | -0.014 | 0.002 | 4.63E-12 | -0.002 | 0.011 | 0.825 | 46.92 |
| rs143743568 | A | G | 0.15 | 0.012 | 0.002 | 4.99E-09 | -0.003 | 0.011 | 0.787 | 34.81 |
| rs150537577 | A | G | 0.92 | -0.015 | 0.003 | 9.50E-09 | -0.004 | 0.015 | 0.812 | 33.73 |
| rs1505676 | C | G | 0.63 | 0.008 | 0.002 | 6.67E-09 | 0.008 | 0.008 | 0.309 | 31.36 |
| rs1542354 | A | G | 0.48 | -0.008 | 0.001 | 2.72E-09 | 0.008 | 0.008 | 0.312 | 36 |
| rs1544 | A | G | 0.26 | -0.01 | 0.002 | 1.76E-09 | 0.009 | 0.009 | 0.321 | 36 |
| rs1564347 | T | G | 0.34 | 0.009 | 0.002 | 2.39E-09 | 0.001 | 0.008 | 0.907 | 34.42 |
| rs1603460 | T | G | 0.42 | 0.009 | 0.001 | 2.20E-10 | 0.014 | 0.008 | 0.081 | 41.33 |
| rs162445 | A | G | 0.08 | 0.015 | 0.003 | 5.72E-09 | 0.002 | 0.015 | 0.874 | 34.18 |
| rs1689510 | C | G | 0.33 | 0.019 | 0.002 | 1.35E-38 | -0.013 | 0.008 | 0.112 | 167.27 |
| rs17048855 | A | G | 0.34 | 0.011 | 0.002 | 3.10E-13 | -0.005 | 0.008 | 0.552 | 51.84 |
| rs17110109 | T | C | 0.62 | -0.009 | 0.001 | 1.47E-10 | 0.005 | 0.008 | 0.553 | 44.13 |
| rs17144467 | A | C | 0.69 | 0.01 | 0.002 | 8.74E-11 | 0 | 0.008 | 0.968 | 42.68 |
| rs17148998 | A | G | 0.2 | 0.01 | 0.002 | 6.98E-09 | -0.001 | 0.01 | 0.925 | 35.3 |
| rs1717204 | A | C | 0.18 | -0.012 | 0.002 | 1.20E-10 | -0.004 | 0.01 | 0.674 | 42.25 |
| rs17248751 | A | G | 0.78 | -0.014 | 0.002 | 1.28E-15 | 0.013 | 0.009 | 0.16 | 64 |
| rs173003 | A | C | 0.49 | -0.008 | 0.001 | 5.50E-09 | 0.008 | 0.008 | 0.317 | 34.31 |
| rs1738050 | C | G | 0.62 | -0.009 | 0.001 | 1.42E-10 | -0.007 | 0.008 | 0.386 | 44.13 |
| rs17411339 | A | G | 0.56 | 0.014 | 0.001 | 5.10E-23 | 0.005 | 0.008 | 0.546 | 100 |
| rs17428076 | C | G | 0.76 | 0.014 | 0.002 | 6.41E-17 | 0.016 | 0.009 | 0.085 | 73.32 |
| rs1747714 | T | C | 0.53 | 0.011 | 0.001 | 3.82E-14 | 0.011 | 0.008 | 0.163 | 57.33 |
| rs1747817 | T | C | 0.76 | -0.009 | 0.002 | 2.29E-08 | 0.012 | 0.009 | 0.206 | 33.06 |
| rs17489649 | A | G | 0.68 | 0.013 | 0.002 | 2.77E-17 | -0.007 | 0.008 | 0.417 | 71.68 |
| rs17502934 | T | G | 0.15 | -0.017 | 0.002 | 2.12E-17 | 0.035 | 0.011 | 0.002 | 69.72 |
| rs175325 | A | T | 0.6 | -0.01 | 0.001 | 2.24E-12 | 0.007 | 0.008 | 0.36 | 51.02 |
| rs17551064 | A | G | 0.84 | 0.014 | 0.002 | 1.53E-13 | -0.014 | 0.011 | 0.199 | 55.07 |
| rs17563464 | A | C | 0.22 | -0.015 | 0.002 | 1.74E-17 | 0.015 | 0.009 | 0.111 | 74.77 |
| rs17565975 | A | G | 0.56 | -0.011 | 0.001 | 5.54E-14 | 0.007 | 0.008 | 0.345 | 57.33 |
| rs17598675 | T | C | 0.51 | -0.012 | 0.001 | 1.37E-16 | -0.01 | 0.008 | 0.222 | 68.65 |
| rs17604349 | A | G | 0.19 | 0.013 | 0.002 | 2.36E-12 | 0.007 | 0.01 | 0.471 | 49.78 |
| rs176218 | T | G | 0.2 | 0.02 | 0.002 | 1.10E-29 | -0.018 | 0.01 | 0.062 | 123.46 |
| rs17732878 | T | C | 0.79 | -0.01 | 0.002 | 4.06E-08 | -0.015 | 0.009 | 0.125 | 31.23 |
| rs178183 | T | C | 0.25 | 0.015 | 0.002 | 4.81E-19 | -0.002 | 0.009 | 0.852 | 82.13 |
| rs17882802 | A | G | 0.43 | 0.008 | 0.001 | 3.00E-09 | 0.012 | 0.008 | 0.113 | 36 |
| rs182902112 | A | C | 0.02 | -0.03 | 0.005 | 1.17E-08 | 0.042 | 0.029 | 0.145 | 32.04 |
| rs1842713 | A | G | 0.79 | -0.012 | 0.002 | 9.67E-12 | 0.008 | 0.009 | 0.414 | 47.37 |
| rs1861786 | A | G | 0.38 | -0.008 | 0.001 | 2.39E-08 | 0.01 | 0.008 | 0.231 | 33.47 |
| rs1865955 | T | C | 0.82 | 0.012 | 0.002 | 4.06E-11 | -0.003 | 0.01 | 0.806 | 41.23 |
| rs1866823 | A | G | 0.54 | 0.011 | 0.001 | 3.09E-14 | 0.006 | 0.008 | 0.415 | 58.41 |
| rs1890132 | T | C | 0.71 | -0.009 | 0.002 | 6.56E-09 | -0.006 | 0.009 | 0.512 | 35.2 |
| rs192436652 | T | C | 0.03 | -0.032 | 0.004 | 2.06E-12 | 0 | 0.024 | 0.997 | 50.25 |
| rs1933264 | T | C | 0.75 | 0.01 | 0.002 | 5.85E-10 | -0.004 | 0.009 | 0.691 | 39.06 |
| rs1955250 | A | C | 0.91 | -0.014 | 0.002 | 2.26E-08 | -0.004 | 0.014 | 0.79 | 31.36 |
| rs1991585 | T | C | 0.71 | -0.01 | 0.002 | 6.26E-11 | 0.01 | 0.009 | 0.221 | 45.34 |
| rs2007655 | T | G | 0.5 | 0.009 | 0.001 | 8.64E-10 | -0.008 | 0.008 | 0.288 | 37.73 |
| rs2011603 | A | G | 0.74 | -0.009 | 0.002 | 1.12E-08 | 0 | 0.009 | 0.999 | 32.35 |
| rs2034631 | T | C | 0.65 | 0.009 | 0.002 | 2.64E-09 | 0.006 | 0.008 | 0.44 | 35.2 |
| rs2052285 | A | G | 0.6 | 0.011 | 0.001 | 7.25E-15 | -0.002 | 0.008 | 0.838 | 64 |
| rs2055940 | A | G | 0.32 | 0.008 | 0.002 | 4.95E-08 | 0.008 | 0.008 | 0.341 | 29.88 |
| rs2081652 | A | T | 0.66 | 0.012 | 0.002 | 1.09E-16 | 0.011 | 0.008 | 0.194 | 67.24 |
| rs2131167 | A | G | 0.45 | 0.008 | 0.001 | 1.52E-08 | -0.004 | 0.008 | 0.575 | 32.65 |
| rs2179152 | T | C | 0.37 | -0.013 | 0.002 | 1.68E-19 | 0 | 0.008 | 0.991 | 76.27 |
| rs2183271 | T | C | 0.64 | 0.008 | 0.002 | 1.91E-08 | 0 | 0.008 | 0.991 | 31.36 |
| rs2199409 | T | C | 0.8 | 0.01 | 0.002 | 3.14E-09 | -0.005 | 0.01 | 0.629 | 36.71 |
| rs2212430 | T | C | 0.3 | -0.01 | 0.002 | 1.32E-10 | -0.01 | 0.008 | 0.255 | 39.06 |
| rs2216144 | T | C | 0.53 | 0.01 | 0.001 | 2.58E-12 | 0.007 | 0.008 | 0.389 | 49 |
| rs2256965 | A | G | 0.42 | 0.011 | 0.001 | 2.40E-13 | 0.002 | 0.008 | 0.8 | 57.33 |
| rs2287838 | A | G | 0.55 | -0.01 | 0.001 | 1.27E-13 | -0.001 | 0.008 | 0.878 | 55.18 |
| rs2290601 | T | C | 0.77 | -0.015 | 0.002 | 4.49E-19 | -0.003 | 0.009 | 0.722 | 76.82 |
| rs2297293 | C | G | 0.31 | 0.01 | 0.002 | 8.80E-11 | 0.017 | 0.008 | 0.04 | 43.56 |
| rs232496 | T | C | 0.37 | 0.009 | 0.002 | 7.14E-10 | 0.004 | 0.008 | 0.647 | 36 |
| rs2364544 | A | G | 0.39 | -0.011 | 0.001 | 2.91E-13 | 0.016 | 0.008 | 0.042 | 56.25 |
| rs2434672 | A | C | 0.53 | 0.009 | 0.001 | 1.06E-10 | -0.012 | 0.008 | 0.117 | 42.25 |
| rs2436760 | T | C | 0.97 | 0.023 | 0.004 | 4.52E-08 | -0.01 | 0.023 | 0.668 | 30.51 |
| rs2447097 | T | G | 0.46 | 0.01 | 0.001 | 1.10E-11 | 0.001 | 0.008 | 0.887 | 47.02 |
| rs2469226 | A | T | 0.22 | 0.01 | 0.002 | 1.28E-08 | 0.033 | 0.009 | 3.84E-04 | 31.89 |
| rs2496482 | T | C | 0.42 | 0.012 | 0.002 | 9.20E-17 | -0.005 | 0.008 | 0.514 | 65.07 |
| rs252991 | A | G | 0.37 | 0.01 | 0.002 | 6.30E-11 | -0.014 | 0.008 | 0.074 | 40.11 |
| rs2545795 | A | C | 0.44 | 0.013 | 0.001 | 1.03E-18 | 0.021 | 0.008 | 0.007 | 79.72 |
| rs2554835 | A | G | 0.4 | 0.009 | 0.001 | 8.95E-10 | -0.01 | 0.008 | 0.23 | 39.51 |
| rs2570497 | T | C | 0.64 | -0.012 | 0.002 | 1.27E-16 | 0.01 | 0.008 | 0.197 | 65.07 |
| rs2725370 | T | C | 0.3 | -0.014 | 0.002 | 4.96E-20 | 0 | 0.008 | 0.965 | 88.36 |
| rs2764684 | T | C | 0.83 | 0.014 | 0.002 | 9.99E-15 | -0.001 | 0.01 | 0.903 | 56.65 |
| rs2838006 | T | C | 0.36 | 0.013 | 0.002 | 1.38E-19 | 0.003 | 0.008 | 0.713 | 77.44 |
| rs28513882 | A | G | 0.18 | -0.011 | 0.002 | 2.03E-09 | 0 | 0.01 | 0.975 | 37.35 |
| rs2885198 | A | G | 0.53 | 0.009 | 0.001 | 4.63E-10 | -0.008 | 0.008 | 0.323 | 39.51 |
| rs2923424 | A | G | 0.61 | 0.012 | 0.001 | 6.31E-17 | 0.015 | 0.008 | 0.056 | 73.47 |
| rs2929032 | A | G | 0.47 | -0.008 | 0.001 | 8.40E-09 | 0.007 | 0.008 | 0.352 | 33.47 |
| rs2958182 | A | T | 0.34 | 0.008 | 0.002 | 3.46E-08 | -0.002 | 0.008 | 0.835 | 29.88 |
| rs2964199 | T | C | 0.31 | -0.01 | 0.002 | 4.61E-11 | 0.005 | 0.008 | 0.524 | 43.56 |
| rs2964255 | A | G | 0.31 | 0.009 | 0.002 | 6.34E-09 | 0.014 | 0.008 | 0.101 | 34.42 |
| rs2998299 | T | G | 0.21 | -0.014 | 0.002 | 1.51E-15 | -0.004 | 0.01 | 0.654 | 64 |
| rs3026996 | A | C | 0.76 | 0.014 | 0.002 | 5.42E-17 | 0.002 | 0.009 | 0.835 | 74.39 |
| rs303752 | A | G | 0.41 | -0.011 | 0.001 | 1.47E-14 | 0.005 | 0.008 | 0.55 | 61.73 |
| rs3111251 | T | C | 0.43 | -0.009 | 0.001 | 3.88E-10 | -0.005 | 0.008 | 0.514 | 41.33 |
| rs34067381 | T | G | 0.36 | -0.01 | 0.002 | 9.77E-13 | -0.002 | 0.008 | 0.776 | 48.07 |
| rs34098770 | A | G | 0.15 | -0.016 | 0.002 | 2.95E-16 | 0.005 | 0.011 | 0.667 | 64 |
| rs34305371 | A | G | 0.1 | 0.031 | 0.002 | 1.10E-39 | 0.001 | 0.013 | 0.964 | 171.17 |
| rs34394051 | A | G | 0.84 | -0.012 | 0.002 | 2.30E-09 | 0.003 | 0.011 | 0.747 | 34.22 |
| rs34720381 | T | C | 0.09 | -0.018 | 0.002 | 1.90E-13 | 0.011 | 0.013 | 0.409 | 56.25 |
| rs34807077 | A | C | 0.16 | 0.013 | 0.002 | 3.29E-11 | -0.001 | 0.011 | 0.921 | 45.39 |
| rs35417702 | T | C | 0.52 | -0.015 | 0.001 | 2.40E-27 | -0.002 | 0.008 | 0.835 | 117.88 |
| rs35606437 | A | G | 0.27 | 0.011 | 0.002 | 3.98E-11 | 0.003 | 0.009 | 0.697 | 43.07 |
| rs35929923 | A | G | 0.25 | -0.011 | 0.002 | 1.77E-12 | 0.018 | 0.009 | 0.05 | 50.77 |
| rs363096 | T | C | 0.42 | -0.014 | 0.001 | 5.77E-24 | 0.002 | 0.008 | 0.754 | 104.33 |
| rs3751331 | A | G | 0.38 | -0.01 | 0.001 | 1.75E-11 | 0.002 | 0.008 | 0.783 | 48.01 |
| rs3809634 | A | G | 0.68 | -0.011 | 0.002 | 6.82E-13 | 0.002 | 0.008 | 0.824 | 52.8 |
| rs382196 | T | G | 0.38 | -0.008 | 0.001 | 1.47E-08 | -0.001 | 0.008 | 0.874 | 34.31 |
| rs3859523 | T | C | 0.82 | 0.012 | 0.002 | 9.92E-12 | 0.021 | 0.01 | 0.042 | 47.46 |
| rs3897821 | A | G | 0.67 | 0.015 | 0.002 | 3.58E-25 | -0.008 | 0.008 | 0.362 | 105.4 |
| rs3948495 | T | G | 0.41 | -0.009 | 0.001 | 3.95E-10 | -0.005 | 0.008 | 0.544 | 40.41 |
| rs401526 | T | C | 0.53 | -0.01 | 0.001 | 1.15E-13 | -0.011 | 0.008 | 0.154 | 55.18 |
| rs401966 | C | G | 0.39 | -0.01 | 0.001 | 1.72E-11 | 0.003 | 0.008 | 0.669 | 48.01 |
| rs42210 | C | G | 0.71 | -0.01 | 0.002 | 2.74E-10 | -0.003 | 0.009 | 0.762 | 37.52 |
| rs42302 | A | G | 0.34 | 0.009 | 0.002 | 5.96E-09 | 0.015 | 0.008 | 0.072 | 32.87 |
| rs4298514 | T | C | 0.7 | 0.01 | 0.002 | 1.07E-11 | -0.006 | 0.008 | 0.473 | 48.07 |
| rs4320563 | A | G | 0.51 | 0.011 | 0.001 | 2.24E-14 | -0.004 | 0.008 | 0.587 | 60.62 |
| rs4328757 | T | C | 0.61 | 0.01 | 0.001 | 2.95E-11 | 0.011 | 0.008 | 0.156 | 47.02 |
| rs4352658 | T | C | 0.08 | -0.019 | 0.003 | 1.71E-13 | 0.02 | 0.014 | 0.152 | 53.4 |
| rs4358081 | A | C | 0.53 | -0.01 | 0.001 | 1.28E-11 | 0.007 | 0.008 | 0.366 | 46.05 |
| rs4369924 | A | G | 0.16 | 0.013 | 0.002 | 5.58E-11 | -0.002 | 0.011 | 0.831 | 43.28 |
| rs4384309 | A | G | 0.46 | 0.01 | 0.001 | 4.89E-13 | -0.001 | 0.008 | 0.889 | 53.08 |
| rs4467547 | T | G | 0.41 | 0.012 | 0.001 | 4.88E-17 | 0.005 | 0.008 | 0.51 | 74.7 |
| rs4500930 | T | C | 0.34 | -0.011 | 0.002 | 1.67E-13 | 0.008 | 0.008 | 0.341 | 52.8 |
| rs4663617 | A | T | 0.23 | 0.01 | 0.002 | 4.12E-10 | -0.022 | 0.009 | 0.018 | 37.43 |
| rs4673840 | T | C | 0.84 | -0.014 | 0.002 | 5.86E-13 | -0.019 | 0.011 | 0.067 | 52.75 |
| rs4675248 | A | G | 0.4 | -0.009 | 0.001 | 4.50E-11 | -0.001 | 0.008 | 0.921 | 45.08 |
| rs4719944 | T | C | 0.55 | -0.012 | 0.001 | 9.25E-18 | 0 | 0.008 | 0.958 | 74.7 |
| rs4726070 | A | G | 0.6 | 0.012 | 0.001 | 1.36E-17 | -0.025 | 0.008 | 0.002 | 75.94 |
| rs4757957 | C | G | 0.69 | 0.013 | 0.002 | 1.79E-18 | -0.012 | 0.008 | 0.142 | 78.62 |
| rs4766424 | C | G | 0.12 | 0.014 | 0.002 | 2.47E-11 | 0.015 | 0.012 | 0.185 | 45.72 |
| rs4778058 | T | C | 0.5 | -0.009 | 0.001 | 4.92E-11 | 0.019 | 0.008 | 0.015 | 43.18 |
| rs4787457 | A | G | 0.63 | 0.016 | 0.002 | 1.23E-28 | 0.016 | 0.008 | 0.045 | 116.64 |
| rs4793090 | A | G | 0.66 | 0.008 | 0.002 | 1.67E-08 | -0.01 | 0.008 | 0.235 | 31.36 |
| rs4810894 | A | G | 0.37 | -0.009 | 0.002 | 4.38E-09 | -0.003 | 0.008 | 0.739 | 32.11 |
| rs4839155 | T | G | 0.77 | 0.012 | 0.002 | 4.66E-12 | 0.022 | 0.009 | 0.018 | 45.76 |
| rs4846724 | A | G | 0.53 | 0.01 | 0.001 | 2.89E-12 | 0.006 | 0.008 | 0.412 | 49 |
| rs4848924 | A | C | 0.71 | -0.012 | 0.002 | 3.56E-14 | -0.003 | 0.009 | 0.751 | 60.84 |
| rs488476 | C | G | 0.62 | -0.011 | 0.002 | 8.29E-15 | -0.002 | 0.008 | 0.802 | 56.75 |
| rs4895650 | T | C | 0.55 | 0.009 | 0.001 | 5.31E-11 | 0 | 0.008 | 0.974 | 44.13 |
| rs4899012 | C | G | 0.6 | -0.01 | 0.001 | 8.06E-13 | 0.009 | 0.008 | 0.234 | 54.13 |
| rs4904523 | A | G | 0.52 | -0.008 | 0.001 | 6.27E-09 | -0.003 | 0.008 | 0.657 | 34.31 |
| rs4915735 | A | G | 0.14 | 0.012 | 0.002 | 9.56E-09 | 0.021 | 0.011 | 0.061 | 33.64 |
| rs4977885 | A | G | 0.4 | -0.008 | 0.001 | 6.78E-09 | -0.011 | 0.008 | 0.17 | 36 |
| rs4984541 | A | G | 0.77 | -0.014 | 0.002 | 3.40E-16 | -0.008 | 0.009 | 0.415 | 65.9 |
| rs55736314 | C | G | 0.6 | -0.015 | 0.001 | 4.59E-27 | -0.025 | 0.008 | 0.002 | 121 |
| rs56099375 | T | C | 0.24 | 0.012 | 0.002 | 3.10E-13 | 0.008 | 0.009 | 0.407 | 49.83 |
| rs56319902 | T | C | 0.22 | -0.021 | 0.002 | 5.81E-33 | -0.058 | 0.009 | 6.00E-10 | 146.84 |
| rs56391344 | A | G | 0.25 | 0.015 | 0.002 | 7.49E-20 | -0.009 | 0.009 | 0.325 | 85.56 |
| rs56794817 | A | G | 0.16 | 0.012 | 0.002 | 1.69E-10 | -0.006 | 0.011 | 0.573 | 41.23 |
| rs57349798 | A | G | 0.41 | 0.009 | 0.001 | 8.63E-11 | -0.001 | 0.008 | 0.877 | 44.13 |
| rs57352738 | A | T | 0.2 | -0.015 | 0.002 | 2.51E-18 | -0.001 | 0.01 | 0.938 | 79.94 |
| rs575113 | A | G | 0.29 | 0.013 | 0.002 | 7.52E-18 | -0.008 | 0.009 | 0.37 | 78.62 |
| rs5754753 | T | C | 0.72 | -0.013 | 0.002 | 6.15E-16 | 0.014 | 0.009 | 0.104 | 62.02 |
| rs57661533 | T | C | 0.13 | -0.011 | 0.002 | 3.24E-08 | -0.014 | 0.012 | 0.226 | 29.47 |
| rs580652 | T | C | 0.9 | -0.015 | 0.002 | 4.65E-10 | 0.035 | 0.013 | 0.008 | 37.52 |
| rs590013 | T | C | 0.68 | 0.01 | 0.002 | 2.47E-11 | 0.008 | 0.008 | 0.31 | 44.44 |
| rs6043521 | T | C | 0.6 | -0.008 | 0.001 | 9.72E-09 | 0.008 | 0.008 | 0.306 | 35.15 |
| rs6065080 | T | C | 0.36 | -0.013 | 0.002 | 7.49E-19 | 0 | 0.008 | 0.975 | 75.11 |
| rs6065784 | C | G | 0.7 | 0.011 | 0.002 | 5.78E-13 | 0.006 | 0.009 | 0.481 | 53.78 |
| rs61104616 | A | G | 0.52 | -0.017 | 0.001 | 1.56E-34 | -0.011 | 0.008 | 0.162 | 150.94 |
| rs6123924 | A | G | 0.84 | 0.013 | 0.002 | 3.20E-11 | 0.009 | 0.011 | 0.383 | 46.1 |
| rs61527214 | A | G | 0.41 | 0.011 | 0.001 | 1.13E-14 | 0.005 | 0.008 | 0.488 | 61.73 |
| rs61997667 | T | C | 0.15 | -0.014 | 0.002 | 2.24E-13 | 0.015 | 0.011 | 0.163 | 51.84 |
| rs62155350 | A | G | 0.02 | 0.034 | 0.005 | 1.69E-10 | 0.007 | 0.027 | 0.808 | 40.67 |
| rs62155873 | T | C | 0.12 | -0.015 | 0.002 | 1.65E-12 | -0.009 | 0.012 | 0.43 | 51.7 |
| rs62174974 | A | G | 0.19 | -0.011 | 0.002 | 2.76E-09 | -0.016 | 0.01 | 0.099 | 34.03 |
| rs62179650 | A | G | 0.3 | 0.011 | 0.002 | 5.66E-13 | -0.003 | 0.008 | 0.758 | 49 |
| rs62182994 | T | C | 0.69 | 0.014 | 0.002 | 2.55E-19 | 0.002 | 0.008 | 0.803 | 83.42 |
| rs62379838 | T | C | 0.7 | 0.012 | 0.002 | 1.77E-15 | 0.001 | 0.008 | 0.913 | 65.07 |
| rs628993 | A | G | 0.11 | 0.013 | 0.002 | 3.83E-08 | -0.006 | 0.013 | 0.649 | 30.97 |
| rs6449503 | A | G | 0.5 | 0.018 | 0.001 | 1.84E-39 | -0.014 | 0.008 | 0.076 | 174.62 |
| rs6490618 | T | C | 0.33 | 0.009 | 0.002 | 3.48E-09 | -0.005 | 0.008 | 0.541 | 34.42 |
| rs6493265 | T | C | 0.39 | -0.012 | 0.001 | 2.26E-17 | -0.002 | 0.008 | 0.828 | 75.94 |
| rs6557171 | T | C | 0.32 | -0.016 | 0.002 | 5.48E-25 | -0.006 | 0.008 | 0.462 | 106.78 |
| rs660001 | A | G | 0.21 | -0.016 | 0.002 | 1.66E-21 | -0.005 | 0.009 | 0.588 | 93.07 |
| rs66568921 | T | G | 0.64 | -0.018 | 0.002 | 7.60E-34 | 0.008 | 0.008 | 0.344 | 145.6 |
| rs66721975 | A | G | 0.29 | -0.009 | 0.002 | 1.51E-08 | 0 | 0.009 | 0.984 | 33.64 |
| rs6690195 | T | C | 0.49 | -0.011 | 0.001 | 7.89E-16 | 0.008 | 0.008 | 0.304 | 65.15 |
| rs6697584 | T | C | 0.79 | -0.013 | 0.002 | 1.43E-13 | -0.008 | 0.009 | 0.407 | 54.93 |
| rs6704768 | A | G | 0.56 | -0.012 | 0.001 | 1.84E-16 | -0.007 | 0.008 | 0.377 | 68.65 |
| rs6731373 | A | G | 0.34 | -0.012 | 0.002 | 1.39E-14 | 0.005 | 0.008 | 0.533 | 58.78 |
| rs6812533 | T | C | 0.74 | 0.011 | 0.002 | 4.85E-11 | -0.003 | 0.009 | 0.739 | 43.89 |
| rs68145588 | T | G | 0.14 | -0.013 | 0.002 | 7.53E-10 | 0.003 | 0.011 | 0.815 | 36.57 |
| rs6917154 | T | C | 0.12 | 0.013 | 0.002 | 2.55E-09 | 0.003 | 0.012 | 0.792 | 36.57 |
| rs6924023 | A | G | 0.75 | 0.011 | 0.002 | 5.17E-11 | 0.009 | 0.009 | 0.338 | 44.72 |
| rs6977237 | T | C | 0.47 | 0.008 | 0.001 | 5.59E-09 | -0.004 | 0.008 | 0.643 | 34.31 |
| rs6994287 | A | G | 0.41 | 0.01 | 0.001 | 1.11E-12 | -0.003 | 0.008 | 0.667 | 53.08 |
| rs7012546 | T | C | 0.42 | 0.009 | 0.001 | 1.25E-10 | -0.001 | 0.008 | 0.899 | 43.18 |
| rs7029718 | A | G | 0.41 | 0.024 | 0.001 | 7.82E-65 | 0.006 | 0.008 | 0.461 | 303.76 |
| rs7041702 | A | G | 0.74 | -0.011 | 0.002 | 4.53E-12 | -0.001 | 0.009 | 0.931 | 48.13 |
| rs7167688 | T | C | 0.49 | -0.009 | 0.001 | 1.71E-11 | -0.002 | 0.008 | 0.792 | 45.08 |
| rs7171405 | A | G | 0.24 | -0.01 | 0.002 | 7.53E-10 | 0.002 | 0.009 | 0.832 | 39.85 |
| rs717996 | T | C | 0.6 | -0.012 | 0.001 | 1.89E-17 | -0.011 | 0.008 | 0.147 | 75.94 |
| rs7226824 | T | C | 0.53 | -0.008 | 0.001 | 2.72E-08 | 0.006 | 0.008 | 0.435 | 31.04 |
| rs72486027 | T | C | 0.25 | -0.01 | 0.002 | 1.61E-10 | -0.006 | 0.009 | 0.532 | 42.25 |
| rs7254263 | T | C | 0.28 | -0.012 | 0.002 | 1.84E-14 | 0.005 | 0.009 | 0.539 | 55.32 |
| rs72622559 | T | C | 0.23 | -0.012 | 0.002 | 5.50E-13 | -0.001 | 0.01 | 0.948 | 49.83 |
| rs72672052 | A | T | 0.18 | -0.01 | 0.002 | 4.87E-08 | -0.008 | 0.01 | 0.45 | 31.48 |
| rs72709560 | A | G | 0.69 | 0.009 | 0.002 | 2.03E-08 | -0.007 | 0.008 | 0.411 | 32.11 |
| rs72828517 | T | C | 0.83 | -0.017 | 0.002 | 1.38E-19 | 0.006 | 0.01 | 0.578 | 78.18 |
| rs72829857 | A | G | 0.76 | -0.015 | 0.002 | 1.52E-19 | 0.001 | 0.009 | 0.912 | 86.72 |
| rs72917504 | T | C | 0.06 | -0.018 | 0.003 | 2.91E-09 | 0.01 | 0.017 | 0.559 | 35.2 |
| rs72944064 | T | C | 0.74 | -0.01 | 0.002 | 9.39E-10 | 0.004 | 0.009 | 0.646 | 37.52 |
| rs72962169 | T | C | 0.16 | -0.017 | 0.002 | 1.26E-19 | 0.011 | 0.011 | 0.296 | 83.87 |
| rs72993796 | T | C | 0.88 | 0.013 | 0.002 | 1.12E-08 | -0.021 | 0.012 | 0.083 | 33.32 |
| rs730384 | A | G | 0.44 | 0.01 | 0.001 | 9.38E-13 | 0.015 | 0.008 | 0.06 | 52.05 |
| rs73055556 | A | G | 0.14 | 0.011 | 0.002 | 4.95E-08 | -0.012 | 0.011 | 0.269 | 30.8 |
| rs73191311 | A | G | 0.34 | -0.009 | 0.002 | 2.03E-10 | -0.006 | 0.008 | 0.453 | 39.27 |
| rs7321274 | A | G | 0.8 | 0.011 | 0.002 | 8.21E-11 | 0.013 | 0.01 | 0.164 | 40.11 |
| rs7326331 | A | G | 0.28 | -0.013 | 0.002 | 3.42E-16 | -0.009 | 0.009 | 0.313 | 64 |
| rs73344830 | A | G | 0.42 | 0.017 | 0.001 | 1.00E-32 | -0.001 | 0.008 | 0.858 | 147.45 |
| rs736281 | T | C | 0.39 | 0.009 | 0.001 | 2.49E-09 | 0.002 | 0.008 | 0.772 | 37.73 |
| rs73648455 | T | C | 0.08 | -0.018 | 0.003 | 1.99E-12 | -0.008 | 0.015 | 0.594 | 49.54 |
| rs743316 | T | C | 0.79 | 0.011 | 0.002 | 8.21E-11 | 0.005 | 0.01 | 0.593 | 43.4 |
| rs74415461 | T | C | 0.08 | 0.018 | 0.003 | 6.25E-12 | 0.022 | 0.014 | 0.102 | 45.82 |
| rs7449561 | A | G | 0.23 | 0.01 | 0.002 | 9.73E-10 | -0.002 | 0.009 | 0.789 | 36 |
| rs7460106 | T | C | 0.76 | -0.012 | 0.002 | 2.06E-13 | -0.001 | 0.009 | 0.882 | 52.35 |
| rs746839 | C | G | 0.63 | 0.014 | 0.002 | 1.19E-20 | 0.001 | 0.008 | 0.859 | 81 |
| rs74787922 | A | C | 0.93 | 0.018 | 0.003 | 8.97E-11 | 0.016 | 0.016 | 0.311 | 42.25 |
| rs75033012 | C | G | 0.04 | -0.026 | 0.004 | 1.38E-11 | -0.022 | 0.02 | 0.26 | 46.1 |
| rs75177132 | T | C | 0.05 | 0.03 | 0.004 | 1.01E-16 | 0.004 | 0.018 | 0.812 | 68.52 |
| rs7575637 | A | G | 0.45 | 0.011 | 0.001 | 3.43E-14 | 0.004 | 0.008 | 0.613 | 58.41 |
| rs7603132 | A | G | 0.19 | 0.014 | 0.002 | 1.20E-14 | 0.011 | 0.01 | 0.271 | 57.93 |
| rs76076331 | T | C | 0.13 | 0.021 | 0.002 | 1.71E-23 | -0.002 | 0.012 | 0.845 | 96.23 |
| rs7617204 | A | G | 0.52 | -0.01 | 0.001 | 4.33E-12 | 0.005 | 0.008 | 0.5 | 48.01 |
| rs76235882 | A | G | 0.97 | 0.024 | 0.004 | 4.52E-10 | 0.011 | 0.024 | 0.636 | 38.19 |
| rs76246107 | A | G | 0.09 | -0.016 | 0.003 | 6.52E-10 | -0.015 | 0.015 | 0.304 | 38.82 |
| rs7650602 | T | C | 0.56 | -0.009 | 0.001 | 2.77E-10 | 0 | 0.008 | 0.983 | 40.41 |
| rs76577427 | C | G | 0.9 | 0.02 | 0.002 | 7.78E-18 | 0.007 | 0.013 | 0.564 | 71.54 |
| rs7672622 | A | G | 0.75 | -0.009 | 0.002 | 4.72E-08 | 0.021 | 0.009 | 0.019 | 30.25 |
| rs7683416 | T | C | 0.46 | 0.014 | 0.001 | 6.05E-23 | -0.003 | 0.008 | 0.672 | 98.58 |
| rs76878669 | C | G | 0.76 | 0.014 | 0.002 | 3.10E-17 | -0.002 | 0.009 | 0.863 | 68.79 |
| rs7692359 | T | C | 0.78 | -0.011 | 0.002 | 1.40E-10 | -0.028 | 0.01 | 0.003 | 41.11 |
| rs77025239 | A | G | 0.16 | -0.014 | 0.002 | 4.65E-13 | -0.008 | 0.011 | 0.477 | 54.29 |
| rs77609760 | A | G | 0.93 | 0.024 | 0.003 | 7.71E-17 | 0.009 | 0.016 | 0.57 | 71.04 |
| rs77719387 | A | T | 0.02 | -0.04 | 0.006 | 3.78E-12 | -0.007 | 0.031 | 0.812 | 48.28 |
| rs7849487 | T | G | 0.66 | -0.017 | 0.002 | 1.25E-29 | -0.006 | 0.008 | 0.453 | 123.95 |
| rs78648104 | T | C | 0.91 | -0.014 | 0.002 | 1.70E-08 | 0.005 | 0.014 | 0.737 | 31.81 |
| rs7875078 | A | C | 0.46 | -0.008 | 0.001 | 2.72E-08 | 0 | 0.008 | 0.988 | 31.04 |
| rs7894722 | T | C | 0.37 | 0.009 | 0.001 | 5.41E-09 | 0.009 | 0.008 | 0.286 | 36.86 |
| rs790647 | A | C | 0.23 | -0.015 | 0.002 | 3.81E-18 | 0.01 | 0.009 | 0.302 | 72.75 |
| rs7910403 | T | G | 0.2 | 0.012 | 0.002 | 5.08E-12 | 0.016 | 0.01 | 0.09 | 45.94 |
| rs7924036 | T | G | 0.51 | 0.013 | 0.001 | 2.55E-21 | 0.019 | 0.008 | 0.014 | 90.25 |
| rs79265434 | A | G | 0.88 | -0.02 | 0.002 | 1.98E-19 | 0.011 | 0.012 | 0.373 | 80.18 |
| rs7972246 | T | C | 0.35 | 0.011 | 0.002 | 1.01E-13 | 0.001 | 0.008 | 0.863 | 53.78 |
| rs79728014 | A | G | 0.86 | -0.013 | 0.002 | 2.06E-11 | -0.001 | 0.011 | 0.935 | 44.89 |
| rs7974852 | A | C | 0.52 | 0.012 | 0.001 | 1.14E-16 | -0.003 | 0.008 | 0.672 | 69.84 |
| rs7977614 | A | G | 0.71 | -0.013 | 0.002 | 6.84E-17 | -0.011 | 0.009 | 0.206 | 70.14 |
| rs79994730 | T | C | 0.04 | 0.024 | 0.004 | 5.88E-12 | 0.006 | 0.018 | 0.717 | 47.02 |
| rs8008382 | T | C | 0.3 | -0.011 | 0.002 | 4.07E-12 | 0.007 | 0.009 | 0.409 | 49.94 |
| rs8024 | A | C | 0.33 | -0.012 | 0.002 | 1.13E-14 | -0.027 | 0.008 | 0.001 | 58.78 |
| rs80257979 | T | G | 0.97 | -0.025 | 0.004 | 3.31E-10 | 0.008 | 0.023 | 0.733 | 39.69 |
| rs8030487 | A | G | 0.69 | 0.009 | 0.002 | 2.52E-08 | 0.008 | 0.008 | 0.352 | 32.11 |
| rs806816 | A | T | 0.75 | 0.01 | 0.002 | 4.80E-10 | 0.001 | 0.009 | 0.944 | 39.85 |
| rs8097125 | T | C | 0.38 | 0.009 | 0.001 | 2.31E-09 | -0.004 | 0.008 | 0.629 | 37.73 |
| rs912883 | T | C | 0.68 | 0.009 | 0.002 | 6.20E-09 | -0.005 | 0.008 | 0.578 | 33.64 |
| rs9289300 | T | C | 0.84 | -0.016 | 0.002 | 1.00E-17 | 0.016 | 0.011 | 0.128 | 74.5 |
| rs929511 | T | C | 0.13 | -0.016 | 0.002 | 3.91E-14 | -0.012 | 0.012 | 0.324 | 58.05 |
| rs9359939 | A | C | 0.24 | -0.011 | 0.002 | 1.51E-10 | -0.01 | 0.009 | 0.287 | 43.07 |
| rs936496 | A | G | 0.62 | -0.008 | 0.001 | 4.38E-08 | -0.003 | 0.008 | 0.747 | 31.84 |
| rs9373363 | A | G | 0.75 | -0.011 | 0.002 | 5.54E-12 | -0.006 | 0.009 | 0.512 | 48.13 |
| rs9375188 | T | C | 0.48 | 0.021 | 0.001 | 8.78E-52 | 0.001 | 0.008 | 0.922 | 229.31 |
| rs9411331 | A | C | 0.68 | 0.015 | 0.002 | 7.63E-22 | 0.006 | 0.008 | 0.451 | 93.44 |
| rs9492774 | C | G | 0.37 | 0.008 | 0.002 | 4.33E-08 | 0.002 | 0.008 | 0.76 | 28.44 |
| rs9529146 | T | C | 0.23 | 0.013 | 0.002 | 3.48E-14 | -0.028 | 0.009 | 0.002 | 55.81 |
| rs9556958 | T | C | 0.52 | -0.011 | 0.001 | 5.96E-15 | -0.005 | 0.008 | 0.491 | 61.73 |
| rs9616947 | T | C | 0.38 | -0.013 | 0.002 | 3.58E-20 | 0.016 | 0.008 | 0.046 | 79.8 |
| rs9633970 | T | C | 0.76 | 0.009 | 0.002 | 2.81E-08 | 0.007 | 0.009 | 0.45 | 32.35 |
| rs9655780 | A | G | 0.83 | -0.015 | 0.002 | 3.70E-16 | 0.013 | 0.01 | 0.21 | 64.84 |
| rs969512 | A | T | 0.66 | -0.011 | 0.002 | 9.84E-13 | -0.01 | 0.008 | 0.205 | 49.94 |
| rs9853928 | T | C | 0.21 | -0.013 | 0.002 | 1.59E-13 | -0.008 | 0.01 | 0.405 | 55.81 |
| rs9859556 | T | G | 0.31 | 0.029 | 0.002 | 4.61E-82 | -0.004 | 0.008 | 0.618 | 373.78 |
| rs9882532 | T | C | 0.64 | 0.012 | 0.002 | 2.15E-17 | 0.008 | 0.008 | 0.35 | 68.34 |
| rs9886703 | A | T | 0.17 | -0.014 | 0.002 | 4.46E-13 | -0.004 | 0.01 | 0.674 | 51.24 |
| rs9927137 | A | G | 0.53 | 0.008 | 0.001 | 5.11E-09 | 0.005 | 0.008 | 0.496 | 34.31 |
| rs9927842 | T | C | 0.15 | -0.013 | 0.002 | 6.22E-11 | -0.01 | 0.011 | 0.348 | 42.25 |
| rs9929556 | T | G | 0.57 | -0.01 | 0.001 | 2.31E-12 | -0.004 | 0.008 | 0.637 | 50.01 |
| rs9929762 | A | G | 0.56 | 0.01 | 0.001 | 1.03E-11 | 0.011 | 0.008 | 0.146 | 47.02 |
| rs9933256 | A | G | 0.56 | 0.012 | 0.001 | 6.06E-17 | 0.004 | 0.008 | 0.571 | 72.25 |

**SNPs used as instrumental variables in the MR analysis of educational attainment on total volume of white matter hyperintensities**

| **SNP** | **Effect allele** | **Other allele** | **Effect allele frequency** | **beta (exposure)** | **SE (exposure)** | ***P* (exposure)** | **beta (outcome)** | **SE (outcome)** | ***P* (outcome)** | ***F*-statistic** |
| --- | --- | --- | --- | --- | --- | --- | --- | --- | --- | --- |
| rs10006235 | T | C | 0.27 | -0.009 | 0.002 | 4.48E-09 | 0 | 0.009 | 0.984 | 33.79 |
| rs10021733 | T | C | 0.15 | -0.016 | 0.003 | 9.45E-10 | 0.018 | 0.011 | 0.094 | 36.89 |
| rs10046069 | A | G | 0.12 | 0.013 | 0.002 | 4.40E-09 | 0.032 | 0.012 | 0.009 | 33.32 |
| rs10060023 | T | C | 0.33 | 0.011 | 0.002 | 1.88E-13 | 0 | 0.008 | 0.958 | 53.78 |
| rs10080647 | A | C | 0.14 | 0.012 | 0.002 | 7.05E-10 | 0.031 | 0.011 | 0.007 | 38.44 |
| rs1008078 | T | C | 0.4 | -0.018 | 0.001 | 4.50E-37 | 0.004 | 0.008 | 0.642 | 169 |
| rs10189857 | A | G | 0.57 | 0.016 | 0.001 | 4.69E-29 | -0.009 | 0.008 | 0.263 | 127.37 |
| rs10193498 | A | T | 0.76 | 0.01 | 0.002 | 3.10E-09 | -0.003 | 0.009 | 0.718 | 36.75 |
| rs10215082 | A | G | 0.47 | -0.015 | 0.001 | 7.03E-26 | 0.003 | 0.008 | 0.707 | 113.27 |
| rs10460095 | A | G | 0.57 | -0.012 | 0.001 | 2.61E-18 | 0.004 | 0.008 | 0.588 | 78.45 |
| rs10761251 | A | T | 0.66 | 0.011 | 0.002 | 2.52E-14 | 0.02 | 0.008 | 0.018 | 56.75 |
| rs10772644 | C | G | 0.89 | 0.014 | 0.002 | 1.69E-10 | -0.027 | 0.013 | 0.038 | 41.66 |
| rs10773002 | A | T | 0.25 | 0.018 | 0.002 | 2.50E-30 | -0.005 | 0.009 | 0.606 | 133.69 |
| rs10773208 | T | C | 0.25 | -0.011 | 0.002 | 8.09E-12 | 0.004 | 0.009 | 0.629 | 47.27 |
| rs10798888 | T | G | 0.17 | -0.014 | 0.002 | 5.15E-14 | -0.005 | 0.01 | 0.608 | 54.29 |
| rs10799615 | A | G | 0.25 | -0.009 | 0.002 | 4.11E-08 | 0.005 | 0.009 | 0.549 | 30.94 |
| rs10805383 | A | G | 0.49 | -0.01 | 0.001 | 2.22E-13 | 0.006 | 0.008 | 0.47 | 54.13 |
| rs10830858 | T | C | 0.51 | 0.009 | 0.001 | 5.31E-11 | -0.014 | 0.008 | 0.071 | 43.18 |
| rs10844179 | A | G | 0.23 | 0.01 | 0.002 | 5.03E-10 | 0.007 | 0.009 | 0.438 | 37.43 |
| rs10875121 | C | G | 0.84 | 0.017 | 0.002 | 4.46E-20 | -0.012 | 0.011 | 0.258 | 83.87 |
| rs10892807 | T | C | 0.57 | 0.011 | 0.001 | 1.09E-14 | -0.001 | 0.008 | 0.864 | 60.62 |
| rs10940921 | T | G | 0.42 | 0.011 | 0.001 | 1.31E-14 | -0.014 | 0.008 | 0.075 | 62.86 |
| rs10951590 | T | C | 0.33 | -0.011 | 0.002 | 3.62E-13 | 0.001 | 0.008 | 0.878 | 52.8 |
| rs10979613 | T | C | 0.64 | -0.012 | 0.002 | 1.25E-15 | 0.033 | 0.008 | 6.52E-05 | 60.84 |
| rs10996167 | C | G | 0.65 | 0.011 | 0.002 | 7.47E-09 | 0.005 | 0.008 | 0.569 | 31.92 |
| rs11003463 | T | G | 0.61 | 0.009 | 0.002 | 5.58E-10 | -0.003 | 0.008 | 0.717 | 36.8 |
| rs11023764 | A | G | 0.64 | 0.009 | 0.002 | 5.32E-10 | -0.005 | 0.008 | 0.529 | 36.8 |
| rs11030102 | C | G | 0.75 | 0.01 | 0.002 | 4.60E-10 | 0.01 | 0.009 | 0.264 | 39.85 |
| rs11076962 | T | C | 0.72 | 0.009 | 0.002 | 4.29E-08 | -0.008 | 0.009 | 0.346 | 28.89 |
| rs11081529 | T | C | 0.71 | 0.013 | 0.002 | 5.22E-16 | 0.008 | 0.009 | 0.332 | 70.56 |
| rs11082011 | T | C | 0.67 | 0.019 | 0.002 | 2.06E-38 | -0.014 | 0.008 | 0.105 | 167.27 |
| rs11121177 | A | G | 0.18 | 0.015 | 0.002 | 1.44E-16 | -0.005 | 0.01 | 0.657 | 70.37 |
| rs11130380 | T | G | 0.61 | 0.009 | 0.001 | 4.17E-10 | 0.001 | 0.008 | 0.892 | 41.33 |
| rs11138947 | T | C | 0.72 | 0.011 | 0.002 | 2.36E-11 | -0.024 | 0.009 | 0.007 | 43.07 |
| rs11157931 | A | C | 0.39 | -0.014 | 0.001 | 3.86E-22 | -0.018 | 0.008 | 0.025 | 98.58 |
| rs111852224 | T | C | 0.12 | 0.016 | 0.002 | 4.85E-13 | 0.002 | 0.012 | 0.862 | 50.28 |
| rs11211123 | A | G | 0.22 | 0.01 | 0.002 | 7.62E-10 | -0.031 | 0.009 | 9.11E-04 | 37.43 |
| rs11213482 | A | G | 0.84 | -0.011 | 0.002 | 2.68E-08 | -0.022 | 0.01 | 0.038 | 31.12 |
| rs112603734 | A | C | 0.77 | -0.013 | 0.002 | 2.72E-09 | 0.01 | 0.009 | 0.293 | 35.46 |
| rs112806496 | C | G | 0.91 | -0.016 | 0.002 | 1.72E-10 | 0.005 | 0.014 | 0.712 | 40.96 |
| rs1128956 | T | G | 0.83 | -0.014 | 0.002 | 5.34E-14 | -0.011 | 0.01 | 0.289 | 54.29 |
| rs113520408 | A | G | 0.28 | 0.016 | 0.002 | 1.03E-22 | -0.031 | 0.009 | 3.98E-04 | 93.85 |
| rs113615161 | T | C | 0.13 | -0.012 | 0.002 | 2.98E-09 | 0.006 | 0.012 | 0.605 | 34.31 |
| rs114593137 | A | T | 0.79 | -0.012 | 0.002 | 1.65E-12 | 0.013 | 0.01 | 0.18 | 51.5 |
| rs115000530 | A | T | 0.95 | -0.025 | 0.003 | 1.09E-15 | 0.006 | 0.017 | 0.744 | 65.04 |
| rs11542663 | A | C | 0.69 | 0.011 | 0.002 | 2.44E-13 | 0.001 | 0.009 | 0.883 | 55.75 |
| rs11588857 | A | G | 0.21 | 0.02 | 0.002 | 4.45E-31 | -0.02 | 0.01 | 0.038 | 137.03 |
| rs11598765 | A | G | 0.2 | -0.011 | 0.002 | 1.73E-09 | -0.012 | 0.01 | 0.228 | 34.68 |
| rs11620355 | A | G | 0.09 | 0.018 | 0.002 | 1.09E-13 | 0.025 | 0.014 | 0.067 | 53 |
| rs11657342 | A | G | 0.37 | 0.014 | 0.002 | 2.32E-13 | -0.012 | 0.008 | 0.136 | 54.29 |
| rs11657979 | A | G | 0.24 | -0.009 | 0.002 | 4.57E-08 | 0.03 | 0.009 | 0.002 | 28.65 |
| rs11663602 | A | C | 0.28 | -0.013 | 0.002 | 9.27E-17 | 0.011 | 0.009 | 0.233 | 66.02 |
| rs1167827 | A | G | 0.43 | 0.01 | 0.001 | 2.93E-13 | -0.019 | 0.008 | 0.018 | 55.18 |
| rs11678980 | A | G | 0.45 | -0.017 | 0.001 | 1.60E-31 | 0.013 | 0.008 | 0.098 | 140.59 |
| rs11724690 | T | G | 0.29 | 0.009 | 0.002 | 2.08E-09 | -0.007 | 0.009 | 0.413 | 38.44 |
| rs117398064 | C | G | 0.09 | -0.014 | 0.002 | 1.16E-08 | 0.008 | 0.013 | 0.535 | 31.81 |
| rs118134876 | T | C | 0.06 | -0.022 | 0.003 | 1.50E-13 | 0.01 | 0.017 | 0.535 | 55.25 |
| rs11871429 | A | G | 0.77 | 0.013 | 0.002 | 2.52E-14 | -0.031 | 0.009 | 0.001 | 56.69 |
| rs11894424 | A | C | 0.95 | 0.021 | 0.003 | 2.14E-10 | 0.003 | 0.019 | 0.856 | 40.11 |
| rs12054166 | C | G | 0.74 | 0.01 | 0.002 | 2.30E-09 | -0.007 | 0.009 | 0.425 | 36 |
| rs12076635 | C | G | 0.78 | 0.021 | 0.002 | 3.33E-34 | -0.003 | 0.01 | 0.719 | 148.27 |
| rs12113634 | T | C | 0.65 | 0.009 | 0.002 | 2.50E-09 | -0.008 | 0.008 | 0.344 | 34.42 |
| rs12151248 | T | C | 0.11 | -0.015 | 0.002 | 3.81E-11 | 0.011 | 0.012 | 0.368 | 41.97 |
| rs1220779 | A | G | 0.47 | -0.008 | 0.001 | 1.58E-08 | -0.022 | 0.008 | 0.006 | 31.84 |
| rs12273435 | A | G | 0.21 | -0.013 | 0.002 | 4.72E-14 | -0.005 | 0.01 | 0.635 | 60.29 |
| rs12332731 | A | T | 0.19 | 0.014 | 0.002 | 5.96E-14 | -0.021 | 0.01 | 0.035 | 56.25 |
| rs12375949 | T | C | 0.43 | -0.014 | 0.001 | 1.61E-22 | 0 | 0.008 | 0.957 | 97.16 |
| rs12438177 | A | G | 0.37 | 0.009 | 0.002 | 1.48E-10 | -0.012 | 0.008 | 0.144 | 38.44 |
| rs12468040 | T | G | 0.38 | 0.014 | 0.001 | 3.12E-21 | -0.007 | 0.008 | 0.419 | 95.76 |
| rs12503522 | T | C | 0.28 | -0.011 | 0.002 | 3.93E-13 | 0.001 | 0.009 | 0.951 | 49.88 |
| rs12506221 | T | G | 0.44 | -0.013 | 0.001 | 4.23E-20 | 0.001 | 0.008 | 0.923 | 86.22 |
| rs12519073 | T | C | 0.23 | -0.011 | 0.002 | 1.16E-10 | -0.003 | 0.01 | 0.761 | 39.62 |
| rs12524795 | T | C | 0.44 | 0.01 | 0.001 | 5.28E-13 | -0.009 | 0.008 | 0.26 | 53.08 |
| rs12571549 | A | G | 0.14 | 0.015 | 0.002 | 1.45E-13 | -0.014 | 0.011 | 0.235 | 57 |
| rs12574281 | A | C | 0.63 | -0.008 | 0.002 | 3.04E-08 | -0.008 | 0.008 | 0.299 | 28.44 |
| rs12591647 | T | C | 0.82 | -0.015 | 0.002 | 7.78E-17 | -0.004 | 0.01 | 0.676 | 71.31 |
| rs12602286 | T | G | 0.87 | 0.016 | 0.002 | 4.62E-14 | 0.05 | 0.012 | 3.34E-05 | 58.05 |
| rs12614263 | A | G | 0.5 | 0.008 | 0.001 | 3.38E-09 | -0.003 | 0.008 | 0.716 | 35.15 |
| rs12638072 | A | G | 0.3 | -0.011 | 0.002 | 3.13E-12 | -0.009 | 0.009 | 0.301 | 50.88 |
| rs12643771 | T | C | 0.31 | 0.013 | 0.002 | 1.32E-17 | -0.008 | 0.009 | 0.336 | 75.11 |
| rs12646523 | T | C | 0.25 | -0.013 | 0.002 | 3.79E-16 | 0.003 | 0.009 | 0.708 | 68.06 |
| rs12670376 | A | G | 0.45 | 0.009 | 0.001 | 5.99E-11 | -0.001 | 0.008 | 0.918 | 43.18 |
| rs12761761 | T | C | 0.24 | 0.015 | 0.002 | 4.79E-20 | 0.003 | 0.009 | 0.716 | 81 |
| rs12765185 | A | T | 0.28 | -0.009 | 0.002 | 2.96E-08 | -0.001 | 0.009 | 0.88 | 30.25 |
| rs12789313 | T | C | 0.49 | 0.009 | 0.001 | 6.79E-11 | 0.01 | 0.008 | 0.184 | 43.18 |
| rs12875339 | A | C | 0.64 | -0.011 | 0.002 | 1.71E-13 | 0.007 | 0.008 | 0.379 | 51.84 |
| rs12888615 | T | C | 0.2 | 0.012 | 0.002 | 9.42E-12 | -0.005 | 0.01 | 0.618 | 44.44 |
| rs12957463 | A | G | 0.8 | 0.015 | 0.002 | 9.20E-17 | 0.012 | 0.01 | 0.225 | 65.79 |
| rs12967010 | T | C | 0.77 | 0.011 | 0.002 | 1.33E-11 | 0.002 | 0.01 | 0.874 | 44.97 |
| rs12981405 | T | C | 0.17 | -0.011 | 0.002 | 2.75E-09 | 0.012 | 0.011 | 0.271 | 34.75 |
| rs1301838 | T | C | 0.32 | -0.01 | 0.002 | 1.29E-10 | 0.004 | 0.008 | 0.664 | 40.96 |
| rs13018640 | T | C | 0.6 | -0.021 | 0.001 | 1.05E-50 | 0.014 | 0.008 | 0.087 | 235.84 |
| rs13050131 | A | C | 0.66 | -0.008 | 0.002 | 4.40E-08 | 0.003 | 0.008 | 0.728 | 29.16 |
| rs13133213 | A | G | 0.5 | 0.01 | 0.001 | 7.57E-12 | -0.008 | 0.008 | 0.309 | 47.02 |
| rs13145650 | T | C | 0.92 | -0.017 | 0.002 | 8.99E-12 | -0.013 | 0.014 | 0.361 | 47.89 |
| rs13163845 | T | C | 0.85 | -0.015 | 0.002 | 4.66E-14 | 0.028 | 0.011 | 0.011 | 55.5 |
| rs13197257 | T | G | 0.28 | 0.011 | 0.002 | 2.83E-12 | -0.006 | 0.009 | 0.486 | 46.41 |
| rs13212041 | T | C | 0.8 | 0.01 | 0.002 | 7.42E-09 | -0.004 | 0.01 | 0.687 | 31.48 |
| rs13240401 | T | C | 0.78 | 0.018 | 0.002 | 2.36E-24 | -0.006 | 0.009 | 0.543 | 107.18 |
| rs13261773 | C | G | 0.16 | 0.012 | 0.002 | 3.12E-10 | 0.005 | 0.011 | 0.662 | 41.23 |
| rs13266287 | A | T | 0.67 | -0.009 | 0.002 | 1.75E-08 | 0.009 | 0.008 | 0.287 | 32.11 |
| rs1329125 | T | C | 0.33 | -0.01 | 0.002 | 8.37E-11 | 0.015 | 0.009 | 0.086 | 41.82 |
| rs13327482 | A | G | 0.82 | -0.012 | 0.002 | 4.47E-11 | -0.008 | 0.01 | 0.433 | 45.19 |
| rs1334297 | A | G | 0.74 | 0.026 | 0.002 | 2.53E-58 | 0 | 0.009 | 0.97 | 258 |
| rs13422673 | T | C | 0.47 | -0.011 | 0.001 | 6.48E-15 | 0.006 | 0.008 | 0.419 | 61.73 |
| rs137079 | T | C | 0.14 | 0.013 | 0.002 | 3.06E-10 | 0.012 | 0.011 | 0.297 | 40.96 |
| rs1426619 | T | C | 0.43 | 0.01 | 0.001 | 1.40E-11 | 0 | 0.008 | 0.999 | 47.02 |
| rs1434630 | T | G | 0.15 | -0.014 | 0.002 | 4.63E-12 | -0.015 | 0.011 | 0.189 | 46.92 |
| rs143743568 | A | G | 0.15 | 0.012 | 0.002 | 4.99E-09 | 0.001 | 0.011 | 0.955 | 34.81 |
| rs150537577 | A | G | 0.92 | -0.015 | 0.003 | 9.50E-09 | -0.002 | 0.015 | 0.869 | 33.73 |
| rs1505676 | C | G | 0.63 | 0.008 | 0.002 | 6.67E-09 | 0.012 | 0.008 | 0.141 | 31.36 |
| rs1542354 | A | G | 0.48 | -0.008 | 0.001 | 2.72E-09 | 0 | 0.008 | 0.953 | 36 |
| rs1544 | A | G | 0.26 | -0.01 | 0.002 | 1.76E-09 | -0.016 | 0.009 | 0.068 | 36 |
| rs1564347 | T | G | 0.34 | 0.009 | 0.002 | 2.39E-09 | -0.005 | 0.008 | 0.545 | 34.42 |
| rs1603460 | T | G | 0.42 | 0.009 | 0.001 | 2.20E-10 | -0.001 | 0.008 | 0.925 | 41.33 |
| rs162445 | A | G | 0.08 | 0.015 | 0.003 | 5.72E-09 | 0.02 | 0.015 | 0.177 | 34.18 |
| rs1689510 | C | G | 0.33 | 0.019 | 0.002 | 1.35E-38 | -0.011 | 0.008 | 0.192 | 167.27 |
| rs17048855 | A | G | 0.34 | 0.011 | 0.002 | 3.10E-13 | -0.002 | 0.008 | 0.811 | 51.84 |
| rs17110109 | T | C | 0.62 | -0.009 | 0.001 | 1.47E-10 | 0.004 | 0.008 | 0.648 | 44.13 |
| rs17144467 | A | C | 0.69 | 0.01 | 0.002 | 8.74E-11 | 0.005 | 0.009 | 0.526 | 42.68 |
| rs17148998 | A | G | 0.2 | 0.01 | 0.002 | 6.98E-09 | -0.011 | 0.01 | 0.279 | 35.3 |
| rs1717204 | A | C | 0.18 | -0.012 | 0.002 | 1.20E-10 | 0.008 | 0.01 | 0.438 | 42.25 |
| rs17248751 | A | G | 0.78 | -0.014 | 0.002 | 1.28E-15 | -0.003 | 0.009 | 0.738 | 64 |
| rs173003 | A | C | 0.49 | -0.008 | 0.001 | 5.50E-09 | 0.003 | 0.008 | 0.709 | 34.31 |
| rs1738050 | C | G | 0.62 | -0.009 | 0.001 | 1.42E-10 | 0.005 | 0.008 | 0.575 | 44.13 |
| rs17411339 | A | G | 0.56 | 0.014 | 0.001 | 5.10E-23 | 0.002 | 0.008 | 0.815 | 100 |
| rs17428076 | C | G | 0.76 | 0.014 | 0.002 | 6.41E-17 | -0.007 | 0.009 | 0.475 | 73.32 |
| rs1747714 | T | C | 0.53 | 0.011 | 0.001 | 3.82E-14 | -0.001 | 0.008 | 0.899 | 57.33 |
| rs1747817 | T | C | 0.76 | -0.009 | 0.002 | 2.29E-08 | 0 | 0.009 | 0.969 | 33.06 |
| rs17489649 | A | G | 0.68 | 0.013 | 0.002 | 2.77E-17 | 0.003 | 0.008 | 0.75 | 71.68 |
| rs17502934 | T | G | 0.15 | -0.017 | 0.002 | 2.12E-17 | -0.003 | 0.011 | 0.82 | 69.72 |
| rs175325 | A | T | 0.6 | -0.01 | 0.001 | 2.24E-12 | 0.002 | 0.008 | 0.796 | 51.02 |
| rs17551064 | A | G | 0.84 | 0.014 | 0.002 | 1.53E-13 | -0.013 | 0.011 | 0.241 | 55.07 |
| rs17563464 | A | C | 0.22 | -0.015 | 0.002 | 1.74E-17 | -0.004 | 0.01 | 0.67 | 74.77 |
| rs17565975 | A | G | 0.56 | -0.011 | 0.001 | 5.54E-14 | -0.005 | 0.008 | 0.497 | 57.33 |
| rs17598675 | T | C | 0.51 | -0.012 | 0.001 | 1.37E-16 | 0.009 | 0.008 | 0.258 | 68.65 |
| rs17604349 | A | G | 0.19 | 0.013 | 0.002 | 2.36E-12 | 0.007 | 0.01 | 0.517 | 49.78 |
| rs176218 | T | G | 0.2 | 0.02 | 0.002 | 1.10E-29 | 0.016 | 0.01 | 0.105 | 123.46 |
| rs17732878 | T | C | 0.79 | -0.01 | 0.002 | 4.06E-08 | 0.006 | 0.01 | 0.545 | 31.23 |
| rs178183 | T | C | 0.25 | 0.015 | 0.002 | 4.81E-19 | 0.008 | 0.009 | 0.389 | 82.13 |
| rs17882802 | A | G | 0.43 | 0.008 | 0.001 | 3.00E-09 | -0.009 | 0.008 | 0.271 | 36 |
| rs182902112 | A | C | 0.02 | -0.03 | 0.005 | 1.17E-08 | 0.03 | 0.029 | 0.307 | 32.04 |
| rs1842713 | A | G | 0.79 | -0.012 | 0.002 | 9.67E-12 | 0 | 0.01 | 0.977 | 47.37 |
| rs1861786 | A | G | 0.38 | -0.008 | 0.001 | 2.39E-08 | 0.008 | 0.008 | 0.298 | 33.47 |
| rs1865955 | T | C | 0.82 | 0.012 | 0.002 | 4.06E-11 | -0.017 | 0.01 | 0.094 | 41.23 |
| rs1866823 | A | G | 0.54 | 0.011 | 0.001 | 3.09E-14 | -0.006 | 0.008 | 0.491 | 58.41 |
| rs1890132 | T | C | 0.71 | -0.009 | 0.002 | 6.56E-09 | -0.004 | 0.009 | 0.647 | 35.2 |
| rs192436652 | T | C | 0.03 | -0.032 | 0.004 | 2.06E-12 | 0.015 | 0.025 | 0.552 | 50.25 |
| rs1933264 | T | C | 0.75 | 0.01 | 0.002 | 5.85E-10 | -0.012 | 0.009 | 0.204 | 39.06 |
| rs1955250 | A | C | 0.91 | -0.014 | 0.002 | 2.26E-08 | 0.036 | 0.014 | 0.011 | 31.36 |
| rs1991585 | T | C | 0.71 | -0.01 | 0.002 | 6.26E-11 | 0.016 | 0.009 | 0.062 | 45.34 |
| rs2007655 | T | G | 0.5 | 0.009 | 0.001 | 8.64E-10 | 0.006 | 0.008 | 0.427 | 37.73 |
| rs2011603 | A | G | 0.74 | -0.009 | 0.002 | 1.12E-08 | 0.008 | 0.009 | 0.373 | 32.35 |
| rs2034631 | T | C | 0.65 | 0.009 | 0.002 | 2.64E-09 | -0.015 | 0.008 | 0.066 | 35.2 |
| rs2052285 | A | G | 0.6 | 0.011 | 0.001 | 7.25E-15 | -0.002 | 0.008 | 0.785 | 64 |
| rs2055940 | A | G | 0.32 | 0.008 | 0.002 | 4.95E-08 | 0.007 | 0.008 | 0.391 | 29.88 |
| rs2081652 | A | T | 0.66 | 0.012 | 0.002 | 1.09E-16 | -0.015 | 0.008 | 0.065 | 67.24 |
| rs2131167 | A | G | 0.45 | 0.008 | 0.001 | 1.52E-08 | -0.013 | 0.008 | 0.11 | 32.65 |
| rs2179152 | T | C | 0.37 | -0.013 | 0.002 | 1.68E-19 | 0.009 | 0.008 | 0.253 | 76.27 |
| rs2183271 | T | C | 0.64 | 0.008 | 0.002 | 1.91E-08 | -0.005 | 0.008 | 0.518 | 31.36 |
| rs2199409 | T | C | 0.8 | 0.01 | 0.002 | 3.14E-09 | 0.009 | 0.01 | 0.371 | 36.71 |
| rs2212430 | T | C | 0.3 | -0.01 | 0.002 | 1.32E-10 | -0.007 | 0.009 | 0.401 | 39.06 |
| rs2216144 | T | C | 0.53 | 0.01 | 0.001 | 2.58E-12 | -0.019 | 0.008 | 0.018 | 49 |
| rs2256965 | A | G | 0.42 | 0.011 | 0.001 | 2.40E-13 | 0.004 | 0.008 | 0.639 | 57.33 |
| rs2287838 | A | G | 0.55 | -0.01 | 0.001 | 1.27E-13 | 0.007 | 0.008 | 0.397 | 55.18 |
| rs2290601 | T | C | 0.77 | -0.015 | 0.002 | 4.49E-19 | -0.001 | 0.009 | 0.908 | 76.82 |
| rs2297293 | C | G | 0.31 | 0.01 | 0.002 | 8.80E-11 | 0.007 | 0.009 | 0.406 | 43.56 |
| rs232496 | T | C | 0.37 | 0.009 | 0.002 | 7.14E-10 | -0.014 | 0.008 | 0.094 | 36 |
| rs2364544 | A | G | 0.39 | -0.011 | 0.001 | 2.91E-13 | -0.008 | 0.008 | 0.31 | 56.25 |
| rs2434672 | A | C | 0.53 | 0.009 | 0.001 | 1.06E-10 | -0.003 | 0.008 | 0.687 | 42.25 |
| rs2436760 | T | C | 0.97 | 0.023 | 0.004 | 4.52E-08 | -0.021 | 0.023 | 0.37 | 30.51 |
| rs2447097 | T | G | 0.46 | 0.01 | 0.001 | 1.10E-11 | -0.013 | 0.008 | 0.093 | 47.02 |
| rs2469226 | A | T | 0.22 | 0.01 | 0.002 | 1.28E-08 | -0.017 | 0.009 | 0.078 | 31.89 |
| rs2496482 | T | C | 0.42 | 0.012 | 0.002 | 9.20E-17 | -0.002 | 0.008 | 0.838 | 65.07 |
| rs252991 | A | G | 0.37 | 0.01 | 0.002 | 6.30E-11 | 0 | 0.008 | 0.952 | 40.11 |
| rs2545795 | A | C | 0.44 | 0.013 | 0.001 | 1.03E-18 | 0.003 | 0.008 | 0.716 | 79.72 |
| rs2554835 | A | G | 0.4 | 0.009 | 0.001 | 8.95E-10 | 0.007 | 0.008 | 0.399 | 39.51 |
| rs2570497 | T | C | 0.64 | -0.012 | 0.002 | 1.27E-16 | 0.012 | 0.008 | 0.157 | 65.07 |
| rs2725370 | T | C | 0.3 | -0.014 | 0.002 | 4.96E-20 | -0.013 | 0.009 | 0.127 | 88.36 |
| rs2764684 | T | C | 0.83 | 0.014 | 0.002 | 9.99E-15 | -0.021 | 0.01 | 0.044 | 56.65 |
| rs2838006 | T | C | 0.36 | 0.013 | 0.002 | 1.38E-19 | 0.001 | 0.008 | 0.924 | 77.44 |
| rs28513882 | A | G | 0.18 | -0.011 | 0.002 | 2.03E-09 | 0.028 | 0.01 | 0.007 | 37.35 |
| rs2885198 | A | G | 0.53 | 0.009 | 0.001 | 4.63E-10 | 0.017 | 0.008 | 0.033 | 39.51 |
| rs2923424 | A | G | 0.61 | 0.012 | 0.001 | 6.31E-17 | 0.014 | 0.008 | 0.091 | 73.47 |
| rs2929032 | A | G | 0.47 | -0.008 | 0.001 | 8.40E-09 | 0.001 | 0.008 | 0.901 | 33.47 |
| rs2958182 | A | T | 0.34 | 0.008 | 0.002 | 3.46E-08 | -0.001 | 0.008 | 0.896 | 29.88 |
| rs2964199 | T | C | 0.31 | -0.01 | 0.002 | 4.61E-11 | 0 | 0.009 | 0.97 | 43.56 |
| rs2964255 | A | G | 0.31 | 0.009 | 0.002 | 6.34E-09 | -0.003 | 0.009 | 0.716 | 34.42 |
| rs2998299 | T | G | 0.21 | -0.014 | 0.002 | 1.51E-15 | 0.001 | 0.01 | 0.919 | 64 |
| rs3026996 | A | C | 0.76 | 0.014 | 0.002 | 5.42E-17 | -0.006 | 0.009 | 0.493 | 74.39 |
| rs303752 | A | G | 0.41 | -0.011 | 0.001 | 1.47E-14 | 0.012 | 0.008 | 0.154 | 61.73 |
| rs3111251 | T | C | 0.43 | -0.009 | 0.001 | 3.88E-10 | -0.009 | 0.008 | 0.238 | 41.33 |
| rs34067381 | T | G | 0.36 | -0.01 | 0.002 | 9.77E-13 | -0.007 | 0.008 | 0.404 | 48.07 |
| rs34098770 | A | G | 0.15 | -0.016 | 0.002 | 2.95E-16 | -0.004 | 0.011 | 0.697 | 64 |
| rs34305371 | A | G | 0.1 | 0.031 | 0.002 | 1.10E-39 | 0.008 | 0.013 | 0.548 | 171.17 |
| rs34394051 | A | G | 0.84 | -0.012 | 0.002 | 2.30E-09 | -0.006 | 0.011 | 0.56 | 34.22 |
| rs34720381 | T | C | 0.09 | -0.018 | 0.002 | 1.90E-13 | 0.045 | 0.014 | 9.24E-04 | 56.25 |
| rs34807077 | A | C | 0.16 | 0.013 | 0.002 | 3.29E-11 | 0.004 | 0.011 | 0.73 | 45.39 |
| rs35417702 | T | C | 0.52 | -0.015 | 0.001 | 2.40E-27 | 0.001 | 0.008 | 0.906 | 117.88 |
| rs35606437 | A | G | 0.27 | 0.011 | 0.002 | 3.98E-11 | 0.006 | 0.009 | 0.536 | 43.07 |
| rs35929923 | A | G | 0.25 | -0.011 | 0.002 | 1.77E-12 | 0.01 | 0.009 | 0.293 | 50.77 |
| rs363096 | T | C | 0.42 | -0.014 | 0.001 | 5.77E-24 | -0.011 | 0.008 | 0.173 | 104.33 |
| rs3751331 | A | G | 0.38 | -0.01 | 0.001 | 1.75E-11 | 0.003 | 0.008 | 0.689 | 48.01 |
| rs3809634 | A | G | 0.68 | -0.011 | 0.002 | 6.82E-13 | 0.019 | 0.009 | 0.03 | 52.8 |
| rs382196 | T | G | 0.38 | -0.008 | 0.001 | 1.47E-08 | -0.011 | 0.008 | 0.161 | 34.31 |
| rs3859523 | T | C | 0.82 | 0.012 | 0.002 | 9.92E-12 | 0.005 | 0.01 | 0.663 | 47.46 |
| rs3897821 | A | G | 0.67 | 0.015 | 0.002 | 3.58E-25 | -0.005 | 0.008 | 0.56 | 105.4 |
| rs3948495 | T | G | 0.41 | -0.009 | 0.001 | 3.95E-10 | 0.009 | 0.008 | 0.262 | 40.41 |
| rs401526 | T | C | 0.53 | -0.01 | 0.001 | 1.15E-13 | 0.001 | 0.008 | 0.909 | 55.18 |
| rs401966 | C | G | 0.39 | -0.01 | 0.001 | 1.72E-11 | -0.002 | 0.008 | 0.787 | 48.01 |
| rs42210 | C | G | 0.71 | -0.01 | 0.002 | 2.74E-10 | -0.001 | 0.009 | 0.94 | 37.52 |
| rs42302 | A | G | 0.34 | 0.009 | 0.002 | 5.96E-09 | 0.008 | 0.008 | 0.348 | 32.87 |
| rs4298514 | T | C | 0.7 | 0.01 | 0.002 | 1.07E-11 | 0 | 0.009 | 0.957 | 48.07 |
| rs4320563 | A | G | 0.51 | 0.011 | 0.001 | 2.24E-14 | 0.008 | 0.008 | 0.323 | 60.62 |
| rs4328757 | T | C | 0.61 | 0.01 | 0.001 | 2.95E-11 | -0.001 | 0.008 | 0.854 | 47.02 |
| rs4352658 | T | C | 0.08 | -0.019 | 0.003 | 1.71E-13 | 0.006 | 0.015 | 0.666 | 53.4 |
| rs4358081 | A | C | 0.53 | -0.01 | 0.001 | 1.28E-11 | 0.007 | 0.008 | 0.39 | 46.05 |
| rs4369924 | A | G | 0.16 | 0.013 | 0.002 | 5.58E-11 | 0.002 | 0.011 | 0.883 | 43.28 |
| rs4384309 | A | G | 0.46 | 0.01 | 0.001 | 4.89E-13 | -0.007 | 0.008 | 0.388 | 53.08 |
| rs4467547 | T | G | 0.41 | 0.012 | 0.001 | 4.88E-17 | -0.001 | 0.008 | 0.915 | 74.7 |
| rs4500930 | T | C | 0.34 | -0.011 | 0.002 | 1.67E-13 | 0.01 | 0.008 | 0.235 | 52.8 |
| rs4663617 | A | T | 0.23 | 0.01 | 0.002 | 4.12E-10 | 0.012 | 0.009 | 0.22 | 37.43 |
| rs4673840 | T | C | 0.84 | -0.014 | 0.002 | 5.86E-13 | -0.015 | 0.011 | 0.152 | 52.75 |
| rs4675248 | A | G | 0.4 | -0.009 | 0.001 | 4.50E-11 | 0.015 | 0.008 | 0.064 | 45.08 |
| rs4719944 | T | C | 0.55 | -0.012 | 0.001 | 9.25E-18 | -0.012 | 0.008 | 0.134 | 74.7 |
| rs4726070 | A | G | 0.6 | 0.012 | 0.001 | 1.36E-17 | 0 | 0.008 | 0.96 | 75.94 |
| rs4757957 | C | G | 0.69 | 0.013 | 0.002 | 1.79E-18 | 0.009 | 0.008 | 0.293 | 78.62 |
| rs4766424 | C | G | 0.12 | 0.014 | 0.002 | 2.47E-11 | 0.011 | 0.012 | 0.348 | 45.72 |
| rs4778058 | T | C | 0.5 | -0.009 | 0.001 | 4.92E-11 | -0.001 | 0.008 | 0.891 | 43.18 |
| rs4787457 | A | G | 0.63 | 0.016 | 0.002 | 1.23E-28 | -0.001 | 0.008 | 0.93 | 116.64 |
| rs4793090 | A | G | 0.66 | 0.008 | 0.002 | 1.67E-08 | 0.015 | 0.008 | 0.068 | 31.36 |
| rs4810894 | A | G | 0.37 | -0.009 | 0.002 | 4.38E-09 | 0.014 | 0.008 | 0.09 | 32.11 |
| rs4839155 | T | G | 0.77 | 0.012 | 0.002 | 4.66E-12 | 0.025 | 0.009 | 0.007 | 45.76 |
| rs4846724 | A | G | 0.53 | 0.01 | 0.001 | 2.89E-12 | 0.015 | 0.008 | 0.054 | 49 |
| rs4848924 | A | C | 0.71 | -0.012 | 0.002 | 3.56E-14 | -0.003 | 0.009 | 0.719 | 60.84 |
| rs488476 | C | G | 0.62 | -0.011 | 0.002 | 8.29E-15 | -0.001 | 0.008 | 0.918 | 56.75 |
| rs4895650 | T | C | 0.55 | 0.009 | 0.001 | 5.31E-11 | 0.002 | 0.008 | 0.844 | 44.13 |
| rs4899012 | C | G | 0.6 | -0.01 | 0.001 | 8.06E-13 | 0.003 | 0.008 | 0.747 | 54.13 |
| rs4904523 | A | G | 0.52 | -0.008 | 0.001 | 6.27E-09 | -0.001 | 0.008 | 0.912 | 34.31 |
| rs4915735 | A | G | 0.14 | 0.012 | 0.002 | 9.56E-09 | -0.002 | 0.011 | 0.867 | 33.64 |
| rs4977885 | A | G | 0.4 | -0.008 | 0.001 | 6.78E-09 | -0.009 | 0.008 | 0.282 | 36 |
| rs4984541 | A | G | 0.77 | -0.014 | 0.002 | 3.40E-16 | 0.016 | 0.009 | 0.087 | 65.9 |
| rs55736314 | C | G | 0.6 | -0.015 | 0.001 | 4.59E-27 | -0.006 | 0.008 | 0.443 | 121 |
| rs56099375 | T | C | 0.24 | 0.012 | 0.002 | 3.10E-13 | 0.011 | 0.009 | 0.217 | 49.83 |
| rs56319902 | T | C | 0.22 | -0.021 | 0.002 | 5.81E-33 | 0.036 | 0.009 | 1.24E-04 | 146.84 |
| rs56391344 | A | G | 0.25 | 0.015 | 0.002 | 7.49E-20 | 0.016 | 0.009 | 0.083 | 85.56 |
| rs56794817 | A | G | 0.16 | 0.012 | 0.002 | 1.69E-10 | 0.018 | 0.011 | 0.094 | 41.23 |
| rs57349798 | A | G | 0.41 | 0.009 | 0.001 | 8.63E-11 | 0.002 | 0.008 | 0.85 | 44.13 |
| rs57352738 | A | T | 0.2 | -0.015 | 0.002 | 2.51E-18 | -0.018 | 0.01 | 0.074 | 79.94 |
| rs575113 | A | G | 0.29 | 0.013 | 0.002 | 7.52E-18 | 0.01 | 0.009 | 0.264 | 78.62 |
| rs5754753 | T | C | 0.72 | -0.013 | 0.002 | 6.15E-16 | 0.002 | 0.009 | 0.815 | 62.02 |
| rs57661533 | T | C | 0.13 | -0.011 | 0.002 | 3.24E-08 | 0.007 | 0.012 | 0.558 | 29.47 |
| rs580652 | T | C | 0.9 | -0.015 | 0.002 | 4.65E-10 | 0.022 | 0.013 | 0.102 | 37.52 |
| rs590013 | T | C | 0.68 | 0.01 | 0.002 | 2.47E-11 | -0.002 | 0.008 | 0.815 | 44.44 |
| rs6043521 | T | C | 0.6 | -0.008 | 0.001 | 9.72E-09 | -0.008 | 0.008 | 0.33 | 35.15 |
| rs6065080 | T | C | 0.36 | -0.013 | 0.002 | 7.49E-19 | -0.012 | 0.008 | 0.151 | 75.11 |
| rs6065784 | C | G | 0.7 | 0.011 | 0.002 | 5.78E-13 | 0.01 | 0.009 | 0.241 | 53.78 |
| rs61104616 | A | G | 0.52 | -0.017 | 0.001 | 1.56E-34 | 0.02 | 0.008 | 0.014 | 150.94 |
| rs6123924 | A | G | 0.84 | 0.013 | 0.002 | 3.20E-11 | 0.003 | 0.011 | 0.761 | 46.1 |
| rs61527214 | A | G | 0.41 | 0.011 | 0.001 | 1.13E-14 | -0.009 | 0.008 | 0.271 | 61.73 |
| rs61997667 | T | C | 0.15 | -0.014 | 0.002 | 2.24E-13 | 0.021 | 0.011 | 0.054 | 51.84 |
| rs62155350 | A | G | 0.02 | 0.034 | 0.005 | 1.69E-10 | -0.07 | 0.028 | 0.012 | 40.67 |
| rs62155873 | T | C | 0.12 | -0.015 | 0.002 | 1.65E-12 | 0.011 | 0.012 | 0.341 | 51.7 |
| rs62174974 | A | G | 0.19 | -0.011 | 0.002 | 2.76E-09 | 0 | 0.01 | 0.975 | 34.03 |
| rs62179650 | A | G | 0.3 | 0.011 | 0.002 | 5.66E-13 | -0.007 | 0.009 | 0.392 | 49 |
| rs62182994 | T | C | 0.69 | 0.014 | 0.002 | 2.55E-19 | -0.004 | 0.008 | 0.66 | 83.42 |
| rs62379838 | T | C | 0.7 | 0.012 | 0.002 | 1.77E-15 | 0.003 | 0.009 | 0.712 | 65.07 |
| rs628993 | A | G | 0.11 | 0.013 | 0.002 | 3.83E-08 | -0.016 | 0.013 | 0.236 | 30.97 |
| rs6449503 | A | G | 0.5 | 0.018 | 0.001 | 1.84E-39 | -0.003 | 0.008 | 0.726 | 174.62 |
| rs6490618 | T | C | 0.33 | 0.009 | 0.002 | 3.48E-09 | 0.018 | 0.008 | 0.028 | 34.42 |
| rs6493265 | T | C | 0.39 | -0.012 | 0.001 | 2.26E-17 | 0.007 | 0.008 | 0.378 | 75.94 |
| rs6557171 | T | C | 0.32 | -0.016 | 0.002 | 5.48E-25 | -0.013 | 0.008 | 0.109 | 106.78 |
| rs660001 | A | G | 0.21 | -0.016 | 0.002 | 1.66E-21 | 0.019 | 0.01 | 0.051 | 93.07 |
| rs66568921 | T | G | 0.64 | -0.018 | 0.002 | 7.60E-34 | -0.006 | 0.008 | 0.477 | 145.6 |
| rs66721975 | A | G | 0.29 | -0.009 | 0.002 | 1.51E-08 | 0.005 | 0.009 | 0.571 | 33.64 |
| rs6690195 | T | C | 0.49 | -0.011 | 0.001 | 7.89E-16 | 0.004 | 0.008 | 0.614 | 65.15 |
| rs6697584 | T | C | 0.79 | -0.013 | 0.002 | 1.43E-13 | 0 | 0.01 | 0.991 | 54.93 |
| rs6704768 | A | G | 0.56 | -0.012 | 0.001 | 1.84E-16 | 0.001 | 0.008 | 0.942 | 68.65 |
| rs6731373 | A | G | 0.34 | -0.012 | 0.002 | 1.39E-14 | 0.012 | 0.008 | 0.154 | 58.78 |
| rs6812533 | T | C | 0.74 | 0.011 | 0.002 | 4.85E-11 | 0.004 | 0.009 | 0.636 | 43.89 |
| rs68145588 | T | G | 0.14 | -0.013 | 0.002 | 7.53E-10 | -0.003 | 0.011 | 0.808 | 36.57 |
| rs6917154 | T | C | 0.12 | 0.013 | 0.002 | 2.55E-09 | 0.005 | 0.012 | 0.695 | 36.57 |
| rs6924023 | A | G | 0.75 | 0.011 | 0.002 | 5.17E-11 | -0.022 | 0.009 | 0.019 | 44.72 |
| rs6977237 | T | C | 0.47 | 0.008 | 0.001 | 5.59E-09 | 0.018 | 0.008 | 0.026 | 34.31 |
| rs6994287 | A | G | 0.41 | 0.01 | 0.001 | 1.11E-12 | 0.008 | 0.008 | 0.327 | 53.08 |
| rs7012546 | T | C | 0.42 | 0.009 | 0.001 | 1.25E-10 | 0.005 | 0.008 | 0.551 | 43.18 |
| rs7029718 | A | G | 0.41 | 0.024 | 0.001 | 7.82E-65 | -0.005 | 0.008 | 0.546 | 303.76 |
| rs7041702 | A | G | 0.74 | -0.011 | 0.002 | 4.53E-12 | -0.01 | 0.009 | 0.266 | 48.13 |
| rs7167688 | T | C | 0.49 | -0.009 | 0.001 | 1.71E-11 | 0.004 | 0.008 | 0.635 | 45.08 |
| rs7171405 | A | G | 0.24 | -0.01 | 0.002 | 7.53E-10 | 0.017 | 0.009 | 0.071 | 39.85 |
| rs717996 | T | C | 0.6 | -0.012 | 0.001 | 1.89E-17 | 0.014 | 0.008 | 0.072 | 75.94 |
| rs7226824 | T | C | 0.53 | -0.008 | 0.001 | 2.72E-08 | -0.004 | 0.008 | 0.633 | 31.04 |
| rs72486027 | T | C | 0.25 | -0.01 | 0.002 | 1.61E-10 | -0.007 | 0.009 | 0.455 | 42.25 |
| rs7254263 | T | C | 0.28 | -0.012 | 0.002 | 1.84E-14 | 0.013 | 0.009 | 0.152 | 55.32 |
| rs72622559 | T | C | 0.23 | -0.012 | 0.002 | 5.50E-13 | -0.001 | 0.01 | 0.92 | 49.83 |
| rs72672052 | A | T | 0.18 | -0.01 | 0.002 | 4.87E-08 | -0.001 | 0.01 | 0.899 | 31.48 |
| rs72709560 | A | G | 0.69 | 0.009 | 0.002 | 2.03E-08 | 0.005 | 0.008 | 0.532 | 32.11 |
| rs72828517 | T | C | 0.83 | -0.017 | 0.002 | 1.38E-19 | -0.014 | 0.01 | 0.181 | 78.18 |
| rs72829857 | A | G | 0.76 | -0.015 | 0.002 | 1.52E-19 | 0.009 | 0.009 | 0.36 | 86.72 |
| rs72917504 | T | C | 0.06 | -0.018 | 0.003 | 2.91E-09 | 0.033 | 0.017 | 0.051 | 35.2 |
| rs72944064 | T | C | 0.74 | -0.01 | 0.002 | 9.39E-10 | 0.002 | 0.009 | 0.844 | 37.52 |
| rs72962169 | T | C | 0.16 | -0.017 | 0.002 | 1.26E-19 | 0.003 | 0.011 | 0.765 | 83.87 |
| rs72993796 | T | C | 0.88 | 0.013 | 0.002 | 1.12E-08 | -0.019 | 0.012 | 0.127 | 33.32 |
| rs730384 | A | G | 0.44 | 0.01 | 0.001 | 9.38E-13 | -0.01 | 0.008 | 0.218 | 52.05 |
| rs73055556 | A | G | 0.14 | 0.011 | 0.002 | 4.95E-08 | -0.028 | 0.011 | 0.015 | 30.8 |
| rs73191311 | A | G | 0.34 | -0.009 | 0.002 | 2.03E-10 | -0.009 | 0.008 | 0.308 | 39.27 |
| rs7321274 | A | G | 0.8 | 0.011 | 0.002 | 8.21E-11 | 0.008 | 0.01 | 0.413 | 40.11 |
| rs7326331 | A | G | 0.28 | -0.013 | 0.002 | 3.42E-16 | 0.003 | 0.009 | 0.752 | 64 |
| rs73344830 | A | G | 0.42 | 0.017 | 0.001 | 1.00E-32 | -0.005 | 0.008 | 0.52 | 147.45 |
| rs736281 | T | C | 0.39 | 0.009 | 0.001 | 2.49E-09 | -0.016 | 0.008 | 0.05 | 37.73 |
| rs73648455 | T | C | 0.08 | -0.018 | 0.003 | 1.99E-12 | 0.008 | 0.015 | 0.585 | 49.54 |
| rs743316 | T | C | 0.79 | 0.011 | 0.002 | 8.21E-11 | -0.002 | 0.01 | 0.863 | 43.4 |
| rs74415461 | T | C | 0.08 | 0.018 | 0.003 | 6.25E-12 | -0.009 | 0.014 | 0.521 | 45.82 |
| rs7449561 | A | G | 0.23 | 0.01 | 0.002 | 9.73E-10 | -0.015 | 0.009 | 0.12 | 36 |
| rs7460106 | T | C | 0.76 | -0.012 | 0.002 | 2.06E-13 | 0.02 | 0.009 | 0.031 | 52.35 |
| rs746839 | C | G | 0.63 | 0.014 | 0.002 | 1.19E-20 | 0.014 | 0.008 | 0.1 | 81 |
| rs74787922 | A | C | 0.93 | 0.018 | 0.003 | 8.97E-11 | -0.011 | 0.016 | 0.482 | 42.25 |
| rs75033012 | C | G | 0.04 | -0.026 | 0.004 | 1.38E-11 | -0.016 | 0.02 | 0.414 | 46.1 |
| rs75177132 | T | C | 0.05 | 0.03 | 0.004 | 1.01E-16 | 0.02 | 0.018 | 0.269 | 68.52 |
| rs7575637 | A | G | 0.45 | 0.011 | 0.001 | 3.43E-14 | -0.016 | 0.008 | 0.038 | 58.41 |
| rs7603132 | A | G | 0.19 | 0.014 | 0.002 | 1.20E-14 | -0.007 | 0.01 | 0.455 | 57.93 |
| rs76076331 | T | C | 0.13 | 0.021 | 0.002 | 1.71E-23 | 0.025 | 0.012 | 0.039 | 96.23 |
| rs7617204 | A | G | 0.52 | -0.01 | 0.001 | 4.33E-12 | 0.005 | 0.008 | 0.489 | 48.01 |
| rs76235882 | A | G | 0.97 | 0.024 | 0.004 | 4.52E-10 | 0.034 | 0.024 | 0.153 | 38.19 |
| rs76246107 | A | G | 0.09 | -0.016 | 0.003 | 6.52E-10 | -0.009 | 0.015 | 0.533 | 38.82 |
| rs7650602 | T | C | 0.56 | -0.009 | 0.001 | 2.77E-10 | -0.006 | 0.008 | 0.478 | 40.41 |
| rs76577427 | C | G | 0.9 | 0.02 | 0.002 | 7.78E-18 | 0 | 0.013 | 0.976 | 71.54 |
| rs7672622 | A | G | 0.75 | -0.009 | 0.002 | 4.72E-08 | -0.011 | 0.009 | 0.236 | 30.25 |
| rs7683416 | T | C | 0.46 | 0.014 | 0.001 | 6.05E-23 | 0.003 | 0.008 | 0.728 | 98.58 |
| rs76878669 | C | G | 0.76 | 0.014 | 0.002 | 3.10E-17 | -0.005 | 0.009 | 0.607 | 68.79 |
| rs7692359 | T | C | 0.78 | -0.011 | 0.002 | 1.40E-10 | -0.016 | 0.01 | 0.1 | 41.11 |
| rs77025239 | A | G | 0.16 | -0.014 | 0.002 | 4.65E-13 | 0.007 | 0.011 | 0.538 | 54.29 |
| rs77609760 | A | G | 0.93 | 0.024 | 0.003 | 7.71E-17 | -0.011 | 0.016 | 0.475 | 71.04 |
| rs77719387 | A | T | 0.02 | -0.04 | 0.006 | 3.78E-12 | 0.027 | 0.032 | 0.401 | 48.28 |
| rs7849487 | T | G | 0.66 | -0.017 | 0.002 | 1.25E-29 | -0.008 | 0.008 | 0.371 | 123.95 |
| rs78648104 | T | C | 0.91 | -0.014 | 0.002 | 1.70E-08 | -0.003 | 0.014 | 0.826 | 31.81 |
| rs7875078 | A | C | 0.46 | -0.008 | 0.001 | 2.72E-08 | 0.009 | 0.008 | 0.273 | 31.04 |
| rs7894722 | T | C | 0.37 | 0.009 | 0.001 | 5.41E-09 | -0.006 | 0.008 | 0.485 | 36.86 |
| rs790647 | A | C | 0.23 | -0.015 | 0.002 | 3.81E-18 | -0.007 | 0.009 | 0.431 | 72.75 |
| rs7910403 | T | G | 0.2 | 0.012 | 0.002 | 5.08E-12 | -0.019 | 0.01 | 0.057 | 45.94 |
| rs7924036 | T | G | 0.51 | 0.013 | 0.001 | 2.55E-21 | 0.007 | 0.008 | 0.36 | 90.25 |
| rs79265434 | A | G | 0.88 | -0.02 | 0.002 | 1.98E-19 | 0.007 | 0.012 | 0.584 | 80.18 |
| rs7972246 | T | C | 0.35 | 0.011 | 0.002 | 1.01E-13 | 0.018 | 0.008 | 0.03 | 53.78 |
| rs79728014 | A | G | 0.86 | -0.013 | 0.002 | 2.06E-11 | 0.022 | 0.011 | 0.051 | 44.89 |
| rs7974852 | A | C | 0.52 | 0.012 | 0.001 | 1.14E-16 | 0.005 | 0.008 | 0.571 | 69.84 |
| rs7977614 | A | G | 0.71 | -0.013 | 0.002 | 6.84E-17 | 0.009 | 0.009 | 0.285 | 70.14 |
| rs79994730 | T | C | 0.04 | 0.024 | 0.004 | 5.88E-12 | 0.025 | 0.018 | 0.16 | 47.02 |
| rs8008382 | T | C | 0.3 | -0.011 | 0.002 | 4.07E-12 | -0.007 | 0.009 | 0.409 | 49.94 |
| rs8024 | A | C | 0.33 | -0.012 | 0.002 | 1.13E-14 | 0.001 | 0.008 | 0.9 | 58.78 |
| rs80257979 | T | G | 0.97 | -0.025 | 0.004 | 3.31E-10 | 0.014 | 0.023 | 0.541 | 39.69 |
| rs8030487 | A | G | 0.69 | 0.009 | 0.002 | 2.52E-08 | 0.001 | 0.009 | 0.927 | 32.11 |
| rs806816 | A | T | 0.75 | 0.01 | 0.002 | 4.80E-10 | 0.021 | 0.009 | 0.019 | 39.85 |
| rs8097125 | T | C | 0.38 | 0.009 | 0.001 | 2.31E-09 | 0.005 | 0.008 | 0.506 | 37.73 |
| rs912883 | T | C | 0.68 | 0.009 | 0.002 | 6.20E-09 | -0.002 | 0.008 | 0.781 | 33.64 |
| rs9289300 | T | C | 0.84 | -0.016 | 0.002 | 1.00E-17 | -0.002 | 0.011 | 0.89 | 74.5 |
| rs929511 | T | C | 0.13 | -0.016 | 0.002 | 3.91E-14 | -0.005 | 0.012 | 0.655 | 58.05 |
| rs9359939 | A | C | 0.24 | -0.011 | 0.002 | 1.51E-10 | 0.01 | 0.009 | 0.27 | 43.07 |
| rs936496 | A | G | 0.62 | -0.008 | 0.001 | 4.38E-08 | 0.006 | 0.008 | 0.434 | 31.84 |
| rs9373363 | A | G | 0.75 | -0.011 | 0.002 | 5.54E-12 | -0.009 | 0.009 | 0.329 | 48.13 |
| rs9375188 | T | C | 0.48 | 0.021 | 0.001 | 8.78E-52 | 0.001 | 0.008 | 0.923 | 229.31 |
| rs9411331 | A | C | 0.68 | 0.015 | 0.002 | 7.63E-22 | 0.003 | 0.008 | 0.697 | 93.44 |
| rs9492774 | C | G | 0.37 | 0.008 | 0.002 | 4.33E-08 | -0.016 | 0.008 | 0.049 | 28.44 |
| rs9529146 | T | C | 0.23 | 0.013 | 0.002 | 3.48E-14 | -0.007 | 0.009 | 0.481 | 55.81 |
| rs9556958 | T | C | 0.52 | -0.011 | 0.001 | 5.96E-15 | -0.008 | 0.008 | 0.316 | 61.73 |
| rs9616947 | T | C | 0.38 | -0.013 | 0.002 | 3.58E-20 | 0.001 | 0.008 | 0.865 | 79.8 |
| rs9633970 | T | C | 0.76 | 0.009 | 0.002 | 2.81E-08 | -0.002 | 0.01 | 0.802 | 32.35 |
| rs9655780 | A | G | 0.83 | -0.015 | 0.002 | 3.70E-16 | -0.018 | 0.011 | 0.095 | 64.84 |
| rs969512 | A | T | 0.66 | -0.011 | 0.002 | 9.84E-13 | -0.013 | 0.008 | 0.111 | 49.94 |
| rs9853928 | T | C | 0.21 | -0.013 | 0.002 | 1.59E-13 | -0.008 | 0.01 | 0.412 | 55.81 |
| rs9859556 | T | G | 0.31 | 0.029 | 0.002 | 4.61E-82 | -0.011 | 0.009 | 0.204 | 373.78 |
| rs9882532 | T | C | 0.64 | 0.012 | 0.002 | 2.15E-17 | 0.008 | 0.008 | 0.343 | 68.34 |
| rs9886703 | A | T | 0.17 | -0.014 | 0.002 | 4.46E-13 | -0.028 | 0.011 | 0.007 | 51.24 |
| rs9927137 | A | G | 0.53 | 0.008 | 0.001 | 5.11E-09 | -0.011 | 0.008 | 0.178 | 34.31 |
| rs9927842 | T | C | 0.15 | -0.013 | 0.002 | 6.22E-11 | -0.019 | 0.011 | 0.084 | 42.25 |
| rs9929556 | T | G | 0.57 | -0.01 | 0.001 | 2.31E-12 | -0.003 | 0.008 | 0.746 | 50.01 |
| rs9929762 | A | G | 0.56 | 0.01 | 0.001 | 1.03E-11 | -0.01 | 0.008 | 0.217 | 47.02 |
| rs9933256 | A | G | 0.56 | 0.012 | 0.001 | 6.06E-17 | -0.012 | 0.008 | 0.14 | 72.25 |

**SNPs used as instrumental variables in the MR analysis of surface area on educational attainment**

| **SNP** | **Effect allele** | **Other allele** | **Effect allele frequency** | **beta (exposure)** | **SE (exposure)** | ***P* (exposure)** | **beta (outcome)** | **SE (outcome)** | ***P* (outcome)** | ***F*-statistic** |
| --- | --- | --- | --- | --- | --- | --- | --- | --- | --- | --- |
| rs10418425 | C | G | 0.84 | -0.048 | 0.009 | 3.86E-08 | -0.003 | 0.002 | 0.149 | 30.22 |
| rs10460587 | C | T | 0.46 | -0.043 | 0.006 | 2.22E-11 | -0.003 | 0.002 | 0.079 | 44.76 |
| rs10496091 | G | A | 0.72 | 0.045 | 0.007 | 2.55E-10 | 0.014 | 0.002 | 7.47E-14 | 39.99 |
| rs111939932 | C | T | 0.82 | -0.059 | 0.008 | 1.67E-12 | -0.002 | 0.002 | 0.426 | 49.84 |
| rs11759026 | A | G | 0.77 | -0.08 | 0.008 | 8.34E-26 | -0.009 | 0.002 | 3.95E-06 | 110.32 |
| rs1712512 | A | T | 0.68 | -0.046 | 0.007 | 3.97E-11 | -0.006 | 0.002 | 0.002 | 43.63 |
| rs1802669 | G | A | 0.66 | 0.04 | 0.007 | 2.73E-09 | 0.007 | 0.002 | 1.67E-04 | 35.37 |
| rs182905872 | G | A | 0.96 | 0.102 | 0.017 | 1.84E-09 | 0.006 | 0.005 | 0.233 | 36.13 |
| rs2301718 | G | A | 0.79 | -0.05 | 0.008 | 2.74E-10 | -0.012 | 0.002 | 9.96E-09 | 39.85 |
| rs2764264 | C | T | 0.31 | -0.047 | 0.007 | 1.26E-11 | -0.009 | 0.002 | 3.93E-06 | 45.87 |
| rs35664429 | G | A | 0.95 | 0.087 | 0.015 | 5.33E-09 | 0.007 | 0.004 | 0.067 | 34.06 |
| rs3786824 | G | A | 0.67 | -0.043 | 0.007 | 1.83E-10 | -0.005 | 0.002 | 0.005 | 40.64 |
| rs3789283 | G | A | 0.42 | -0.04 | 0.006 | 6.65E-10 | -0.001 | 0.002 | 0.65 | 38.12 |
| rs3811183 | C | G | 0.6 | 0.041 | 0.007 | 3.72E-10 | 0.012 | 0.002 | 9.95E-13 | 39.25 |
| rs4685 | T | C | 0.33 | 0.043 | 0.007 | 3.31E-10 | -0.001 | 0.002 | 0.742 | 39.49 |
| rs56269136 | C | T | 0.78 | 0.112 | 0.008 | 7.72E-49 | 0.018 | 0.002 | 2.96E-17 | 215.73 |
| rs62283132 | C | T | 0.85 | -0.07 | 0.009 | 5.41E-15 | -0.01 | 0.002 | 5.25E-05 | 61.11 |
| rs7297175 | T | C | 0.43 | 0.045 | 0.006 | 2.38E-12 | 0.016 | 0.002 | 2.49E-19 | 49.14 |
| rs76169231 | T | C | 0.8 | 0.048 | 0.008 | 8.95E-10 | 0.004 | 0.002 | 0.054 | 37.54 |
| rs7671110 | C | T | 0.84 | 0.057 | 0.009 | 1.01E-10 | 0.006 | 0.002 | 0.021 | 41.79 |
| rs76928645 | C | T | 0.9 | -0.091 | 0.011 | 5.93E-18 | -0.004 | 0.003 | 0.139 | 74.54 |
| rs7715167 | T | C | 0.39 | -0.043 | 0.007 | 4.19E-11 | -0.005 | 0.002 | 0.005 | 43.52 |
| rs7797990 | A | G | 0.62 | -0.042 | 0.007 | 1.44E-10 | -0.002 | 0.002 | 0.156 | 41.11 |
| rs78852495 | G | A | 0.97 | -0.097 | 0.017 | 2.92E-08 | -0.005 | 0.005 | 0.241 | 30.76 |
| rs8756 | C | A | 0.49 | 0.054 | 0.006 | 1.51E-17 | 0.008 | 0.002 | 1.41E-06 | 72.69 |

**SNPs used as instrumental variables in the MR analysis of volume on educational attainment**

| **SNP** | **Effect allele** | **Other allele** | **Effect allele frequency** | **beta (exposure)** | **SE (exposure)** | ***P* (exposure)** | **beta (outcome)** | **SE (outcome)** | ***P* (outcome)** | ***F*-statistic** |
| --- | --- | --- | --- | --- | --- | --- | --- | --- | --- | --- |
| rs10460587 | C | T | 0.46 | -0.036 | 0.006 | 1.09E-08 | -0.003 | 0.002 | 0.079 | 32.67 |
| rs10496091 | G | A | 0.72 | 0.041 | 0.007 | 2.62E-09 | 0.014 | 0.002 | 7.47E-14 | 35.45 |
| rs111939932 | C | T | 0.82 | -0.048 | 0.008 | 3.72E-09 | -0.002 | 0.002 | 0.426 | 34.76 |
| rs1163249 | A | G | 0.2 | 0.052 | 0.008 | 3.02E-11 | 0.008 | 0.002 | 2.17E-04 | 44.16 |
| rs13097248 | A | G | 0.85 | 0.049 | 0.009 | 1.97E-08 | 0.007 | 0.002 | 0.003 | 31.53 |
| rs149549868 | G | A | 0.98 | -0.114 | 0.02 | 1.96E-08 | 0.008 | 0.006 | 0.179 | 31.53 |
| rs1979477 | A | C | 0.5 | -0.04 | 0.006 | 9.51E-11 | -0.01 | 0.002 | 1.50E-08 | 41.92 |
| rs2301718 | G | A | 0.79 | -0.05 | 0.008 | 7.28E-11 | -0.012 | 0.002 | 9.96E-09 | 42.44 |
| rs2529418 | G | A | 0.44 | 0.038 | 0.006 | 1.83E-09 | 0.003 | 0.002 | 0.094 | 36.15 |
| rs3130346 | T | C | 0.15 | 0.051 | 0.009 | 7.77E-09 | 0.014 | 0.002 | 2.30E-09 | 33.33 |
| rs3200031 | C | T | 0.92 | -0.075 | 0.012 | 7.70E-11 | -0.012 | 0.003 | 3.53E-04 | 42.33 |
| rs3789283 | G | A | 0.42 | -0.038 | 0.006 | 1.61E-09 | -0.001 | 0.002 | 0.65 | 36.39 |
| rs4259003 | G | A | 0.81 | -0.043 | 0.008 | 4.48E-08 | -0.001 | 0.002 | 0.729 | 29.93 |
| rs4273712 | A | G | 0.73 | -0.067 | 0.007 | 8.14E-22 | -0.009 | 0.002 | 2.02E-06 | 92.12 |
| rs4315664 | A | T | 0.85 | -0.07 | 0.009 | 1.03E-15 | -0.01 | 0.002 | 3.81E-05 | 64.38 |
| rs62057121 | G | A | 0.79 | 0.072 | 0.008 | 1.57E-21 | 0.018 | 0.002 | 4.02E-17 | 90.82 |
| rs7297175 | T | C | 0.43 | 0.036 | 0.006 | 1.18E-08 | 0.016 | 0.002 | 2.49E-19 | 32.52 |
| rs73100947 | C | T | 0.85 | 0.049 | 0.009 | 1.25E-08 | 0.006 | 0.002 | 0.017 | 32.41 |
| rs75745313 | G | A | 0.93 | 0.073 | 0.012 | 9.75E-10 | 0.008 | 0.004 | 0.046 | 37.37 |
| rs76169231 | T | C | 0.8 | 0.051 | 0.008 | 2.90E-11 | 0.004 | 0.002 | 0.054 | 44.24 |
| rs7715167 | T | C | 0.39 | -0.037 | 0.006 | 7.71E-09 | -0.005 | 0.002 | 0.005 | 33.35 |
| rs7986738 | A | G | 0.79 | -0.042 | 0.008 | 4.89E-08 | 0.001 | 0.002 | 0.774 | 29.76 |
| rs8756 | C | A | 0.49 | 0.052 | 0.006 | 6.00E-17 | 0.008 | 0.002 | 1.41E-06 | 69.98 |
| rs9398171 | C | T | 0.29 | -0.052 | 0.007 | 2.67E-14 | -0.008 | 0.002 | 1.00E-05 | 57.96 |
| rs9509982 | A | G | 0.28 | -0.038 | 0.007 | 2.90E-08 | 0.001 | 0.002 | 0.68 | 30.77 |

**SNPs used as instrumental variables in the MR analysis of cortical thickness on educational attainment**

| **SNP** | **Effect allele** | **Other allele** | **Effect allele frequency** | **beta (exposure)** | **SE (exposure)** | ***P* (exposure)** | **beta (outcome)** | **SE (outcome)** | ***P* (outcome)** | ***F*-statistic** |
| --- | --- | --- | --- | --- | --- | --- | --- | --- | --- | --- |
| rs12150230 | A | G | 0.78 | -0.065 | 0.008 | 3.94E-17 | 0.018 | 0.002 | 6.86E-17 | 70.81 |
| rs13079368 | C | T | 0.48 | -0.046 | 0.006 | 7.29E-13 | 0.002 | 0.002 | 0.196 | 51.46 |
| rs13107325 | C | T | 0.93 | 0.085 | 0.013 | 1.03E-11 | 0.019 | 0.003 | 4.45E-08 | 46.28 |
| rs143893586 | T | G | 0.94 | -0.076 | 0.014 | 2.76E-08 | 0.001 | 0.004 | 0.727 | 30.87 |
| rs1687225 | G | A | 0.77 | -0.045 | 0.008 | 3.54E-09 | 0.005 | 0.002 | 0.014 | 34.86 |
| rs2241722 | C | A | 0.67 | 0.064 | 0.007 | 1.06E-20 | -0.005 | 0.002 | 0.004 | 87.05 |
| rs3200031 | C | T | 0.92 | -0.091 | 0.012 | 3.19E-14 | -0.012 | 0.003 | 3.53E-04 | 57.61 |
| rs34151748 | C | G | 0.72 | -0.044 | 0.007 | 9.28E-10 | -0.004 | 0.002 | 0.043 | 37.47 |
| rs614225 | A | G | 0.53 | -0.035 | 0.006 | 4.14E-08 | -0.007 | 0.002 | 4.35E-05 | 30.08 |
| rs61753077 | G | A | 0.95 | -0.088 | 0.015 | 6.55E-09 | 0.004 | 0.004 | 0.413 | 33.66 |
| rs617577 | T | C | 0.9 | 0.08 | 0.011 | 8.78E-14 | 0.002 | 0.003 | 0.54 | 55.62 |
| rs6492273 | T | C | 0.41 | 0.041 | 0.007 | 5.21E-10 | -0.001 | 0.002 | 0.691 | 38.6 |
| rs6939 | A | C | 0.29 | 0.043 | 0.007 | 1.81E-09 | -0.009 | 0.002 | 1.30E-06 | 36.17 |
| rs7219015 | C | T | 0.78 | -0.063 | 0.008 | 6.78E-16 | 0.008 | 0.002 | 1.29E-04 | 65.2 |
| rs7224765 | G | A | 0.08 | 0.069 | 0.012 | 5.55E-09 | -0.002 | 0.003 | 0.433 | 33.98 |
| rs724265 | G | A | 0.38 | -0.039 | 0.007 | 9.06E-09 | 0 | 0.002 | 0.791 | 33.03 |
| rs72618187 | T | C | 0.76 | -0.044 | 0.008 | 5.62E-09 | 0 | 0.002 | 0.941 | 33.96 |
| rs73171590 | G | C | 0.74 | -0.043 | 0.007 | 2.98E-09 | 0.001 | 0.002 | 0.692 | 35.2 |
| rs76928645 | C | T | 0.9 | 0.06 | 0.011 | 2.06E-08 | -0.004 | 0.003 | 0.139 | 31.44 |
| rs78783493 | C | T | 0.91 | 0.074 | 0.011 | 4.80E-11 | 0.008 | 0.003 | 0.025 | 43.26 |
| rs79274348 | C | T | 0.95 | -0.084 | 0.015 | 1.87E-08 | -0.002 | 0.004 | 0.58 | 31.62 |
| rs934447 | A | G | 0.26 | 0.043 | 0.008 | 2.76E-08 | -0.001 | 0.002 | 0.66 | 30.87 |
| rs9921406 | C | T | 0.91 | -0.065 | 0.011 | 1.36E-08 | 0.006 | 0.003 | 0.051 | 32.24 |

**SNPs used as instrumental variables in the MR analysis of local gyrification index on educational attainment**

| **SNP** | **Effect allele** | **Other allele** | **Effect allele frequency** | **beta (exposure)** | **SE (exposure)** | ***P* (exposure)** | **beta (outcome)** | **SE (outcome)** | ***P* (outcome)** | ***F*-statistic** |
| --- | --- | --- | --- | --- | --- | --- | --- | --- | --- | --- |
| rs10460587 | C | T | 0.46 | -0.043 | 0.008 | 1.90E-08 | -0.003 | 0.002 | 0.079 | 31.6 |
| rs11633076 | C | T | 0.51 | 0.045 | 0.008 | 3.96E-09 | 0 | 0.002 | 0.965 | 34.64 |
| rs11759026 | A | G | 0.77 | -0.065 | 0.009 | 2.39E-13 | -0.009 | 0.002 | 3.95E-06 | 53.65 |
| rs12938775 | G | A | 0.5 | -0.056 | 0.007 | 8.46E-14 | -0.002 | 0.002 | 0.308 | 55.7 |
| rs147431626 | G | A | 0.78 | 0.081 | 0.009 | 1.94E-19 | 0.017 | 0.002 | 8.16E-16 | 81.3 |
| rs2047676 | C | T | 0.7 | -0.049 | 0.008 | 2.89E-09 | 0.004 | 0.002 | 0.027 | 35.26 |
| rs2501261 | A | G | 0.23 | 0.049 | 0.009 | 2.15E-08 | 0.005 | 0.002 | 0.012 | 31.36 |
| rs35227403 | T | A | 0.94 | 0.105 | 0.016 | 8.07E-11 | 0.004 | 0.004 | 0.217 | 42.24 |
| rs3814345 | G | A | 0.82 | -0.057 | 0.01 | 3.64E-09 | -0.008 | 0.002 | 1.62E-04 | 34.81 |
| rs6658111 | T | G | 0.64 | -0.077 | 0.008 | 7.49E-23 | -0.009 | 0.002 | 2.63E-07 | 96.85 |
| rs7146019 | A | G | 0.43 | -0.047 | 0.008 | 1.20E-09 | 0.011 | 0.002 | 5.54E-10 | 36.96 |
| rs7484851 | A | G | 0.5 | -0.041 | 0.007 | 2.96E-08 | -0.007 | 0.002 | 1.13E-04 | 30.74 |
| rs7715167 | T | C | 0.39 | -0.046 | 0.008 | 1.55E-09 | -0.005 | 0.002 | 0.005 | 36.47 |
| rs999867 | C | T | 0.92 | 0.081 | 0.014 | 1.01E-08 | 0.002 | 0.003 | 0.43 | 32.83 |

**SNPs used as instrumental variables in the MR analysis of mean curvature on educational attainment**

| **SNP** | **Effect allele** | **Other allele** | **Effect allele frequency** | **beta (exposure)** | **SE (exposure)** | ***P* (exposure)** | **beta (outcome)** | **SE (outcome)** | ***P* (outcome)** | ***F*-statistic** |
| --- | --- | --- | --- | --- | --- | --- | --- | --- | --- | --- |
| rs10197724 | C | T | 0.46 | 0.039 | 0.007 | 3.31E-08 | 0.003 | 0.002 | 0.135 | 30.52 |
| rs1048527 | T | G | 0.92 | 0.096 | 0.013 | 9.97E-14 | -0.012 | 0.003 | 3.91E-04 | 55.37 |
| rs114113354 | A | G | 0.74 | 0.046 | 0.008 | 1.17E-08 | 0 | 0.002 | 0.796 | 32.53 |
| rs116269521 | G | A | 0.97 | -0.109 | 0.019 | 1.33E-08 | -0.009 | 0.005 | 0.058 | 32.28 |
| rs11781155 | C | T | 0.43 | -0.04 | 0.007 | 1.43E-08 | 0 | 0.002 | 0.778 | 32.15 |
| rs12146713 | T | C | 0.9 | 0.073 | 0.012 | 6.83E-10 | -0.006 | 0.003 | 0.05 | 38.07 |
| rs13012151 | C | T | 0.3 | 0.043 | 0.008 | 1.51E-08 | 0 | 0.002 | 0.853 | 32.04 |
| rs13264372 | C | T | 0.63 | -0.043 | 0.007 | 2.09E-09 | 0.001 | 0.002 | 0.439 | 35.89 |
| rs17205972 | G | T | 0.81 | 0.05 | 0.009 | 1.53E-08 | 0 | 0.002 | 0.899 | 32.02 |
| rs17206779 | C | T | 0.48 | 0.041 | 0.007 | 4.05E-09 | -0.001 | 0.002 | 0.439 | 34.6 |
| rs17424787 | G | A | 0.92 | -0.135 | 0.013 | 5.92E-25 | 0.009 | 0.003 | 0.004 | 106.43 |
| rs17676457 | G | C | 0.9 | 0.088 | 0.012 | 2.27E-14 | 0.001 | 0.003 | 0.681 | 58.28 |
| rs2241722 | C | A | 0.67 | -0.041 | 0.007 | 3.01E-08 | -0.005 | 0.002 | 0.004 | 30.7 |
| rs2764686 | A | T | 0.35 | -0.051 | 0.007 | 3.07E-12 | -0.005 | 0.002 | 0.002 | 48.64 |
| rs344351 | G | C | 0.39 | 0.049 | 0.007 | 8.46E-12 | 0.002 | 0.002 | 0.194 | 46.66 |
| rs34636718 | A | G | 0.98 | -0.191 | 0.027 | 7.50E-13 | 0.012 | 0.007 | 0.094 | 51.41 |
| rs4652758 | T | C | 0.46 | 0.04 | 0.007 | 2.42E-08 | -0.002 | 0.002 | 0.182 | 31.13 |
| rs55976523 | C | T | 0.73 | -0.046 | 0.008 | 1.37E-08 | 0.001 | 0.002 | 0.505 | 32.23 |
| rs56125329 | C | T | 0.56 | -0.047 | 0.007 | 3.19E-11 | 0 | 0.002 | 0.836 | 44.05 |
| rs59682551 | T | G | 0.76 | -0.05 | 0.008 | 8.63E-10 | 0.006 | 0.002 | 0.002 | 37.61 |
| rs613511 | C | T | 0.74 | -0.055 | 0.008 | 1.88E-11 | 0.002 | 0.002 | 0.235 | 45.09 |
| rs61789085 | A | G | 0.72 | 0.048 | 0.008 | 6.93E-10 | -0.008 | 0.002 | 1.32E-05 | 38.04 |
| rs6745807 | A | C | 0.36 | -0.049 | 0.007 | 1.31E-11 | 0.005 | 0.002 | 0.011 | 45.8 |
| rs7219015 | C | T | 0.78 | 0.067 | 0.009 | 4.92E-15 | 0.008 | 0.002 | 1.29E-04 | 61.29 |
| rs748593 | C | T | 0.31 | -0.042 | 0.008 | 2.45E-08 | -0.008 | 0.002 | 4.98E-05 | 31.1 |
| rs7559029 | C | A | 0.46 | -0.041 | 0.007 | 4.06E-09 | 0.007 | 0.002 | 1.25E-04 | 34.6 |
| rs756747 | T | G | 0.48 | -0.043 | 0.007 | 1.12E-09 | 0 | 0.002 | 0.836 | 37.1 |
| rs7685018 | G | A | 0.54 | -0.041 | 0.007 | 6.51E-09 | 0.005 | 0.002 | 0.005 | 33.68 |
| rs9398172 | G | A | 0.29 | 0.052 | 0.008 | 1.07E-11 | -0.008 | 0.002 | 7.17E-06 | 46.19 |

**SNPs used as instrumental variables in the MR analysis of intrinsic curvature on educational attainment**

| **SNP** | **Effect allele** | **Other allele** | **Effect allele frequency** | **beta (exposure)** | **SE (exposure)** | ***P* (exposure)** | **beta (outcome)** | **SE (outcome)** | ***P* (outcome)** | ***F*-statistic** |
| --- | --- | --- | --- | --- | --- | --- | --- | --- | --- | --- |
| rs10498754 | T | A | 0.92 | 0.051 | 0.009 | 3.90E-08 | 0.003 | 0.003 | 0.299 | 30.2 |
| rs11134697 | G | A | 0.45 | -0.032 | 0.005 | 5.50E-11 | -0.003 | 0.002 | 0.046 | 42.99 |
| rs11157930 | T | G | 0.39 | -0.028 | 0.005 | 1.27E-08 | -0.014 | 0.002 | 4.76E-15 | 32.37 |
| rs112649229 | G | C | 0.99 | 0.121 | 0.021 | 1.29E-08 | 0.002 | 0.007 | 0.792 | 32.35 |
| rs11674612 | A | G | 0.64 | 0.028 | 0.005 | 2.48E-08 | 0.004 | 0.002 | 0.029 | 31.08 |
| rs11686881 | C | T | 0.64 | -0.031 | 0.005 | 1.65E-09 | 0.003 | 0.002 | 0.056 | 36.35 |
| rs11759026 | A | G | 0.77 | -0.045 | 0.006 | 5.38E-15 | -0.009 | 0.002 | 3.95E-06 | 61.12 |
| rs12938775 | G | A | 0.5 | -0.034 | 0.005 | 4.89E-12 | -0.002 | 0.002 | 0.308 | 47.73 |
| rs147431626 | G | A | 0.78 | 0.056 | 0.006 | 9.89E-22 | 0.017 | 0.002 | 8.16E-16 | 91.74 |
| rs1802669 | G | A | 0.66 | 0.031 | 0.005 | 1.76E-09 | 0.007 | 0.002 | 1.67E-04 | 36.23 |
| rs182905872 | G | A | 0.96 | 0.077 | 0.013 | 3.63E-09 | 0.006 | 0.005 | 0.233 | 34.81 |
| rs201872292 | G | A | 0.92 | -0.051 | 0.009 | 9.52E-09 | -0.004 | 0.003 | 0.225 | 32.94 |
| rs2241722 | C | A | 0.67 | -0.034 | 0.005 | 3.92E-11 | -0.005 | 0.002 | 0.004 | 43.65 |
| rs34216221 | T | C | 0.98 | -0.114 | 0.019 | 2.44E-09 | 0.009 | 0.006 | 0.186 | 35.58 |
| rs35518360 | A | T | 0.93 | -0.055 | 0.01 | 9.45E-09 | 0.019 | 0.003 | 9.79E-09 | 32.95 |
| rs4912174 | A | G | 0.89 | -0.044 | 0.008 | 1.33E-08 | -0.005 | 0.003 | 0.046 | 32.28 |
| rs6778085 | C | T | 0.19 | 0.034 | 0.006 | 4.20E-08 | 0.008 | 0.002 | 2.10E-04 | 30.06 |
| rs76928645 | C | T | 0.9 | -0.057 | 0.008 | 1.12E-12 | -0.004 | 0.003 | 0.139 | 50.62 |
| rs7694606 | G | A | 0.84 | 0.041 | 0.007 | 5.91E-10 | 0.005 | 0.002 | 0.034 | 38.35 |
| rs9930713 | A | G | 0.76 | -0.031 | 0.006 | 4.43E-08 | -0.006 | 0.002 | 0.005 | 29.95 |

**SNPs used as instrumental variables in the MR analysis of mean diffusivity (cortical) on educational attainment**

| **SNP** | **Effect allele** | **Other allele** | **Effect allele frequency** | **beta (exposure)** | **SE (exposure)** | ***P* (exposure)** | **beta (outcome)** | **SE (outcome)** | ***P* (outcome)** | ***F*-statistic** |
| --- | --- | --- | --- | --- | --- | --- | --- | --- | --- | --- |
| rs12048319 | G | T | 0.84 | 0.024 | 0.004 | 3.33E-08 | -0.013 | 0.002 | 3.14E-08 | 30.5 |
| rs13144836 | G | C | 0.64 | -0.025 | 0.004 | 6.16E-09 | -0.01 | 0.002 | 3.55E-08 | 33.78 |
| rs13224758 | G | A | 0.85 | 0.028 | 0.004 | 1.95E-10 | 0 | 0.002 | 0.946 | 40.51 |
| rs1452628 | A | T | 0.62 | 0.033 | 0.004 | 1.87E-14 | -0.001 | 0.002 | 0.723 | 58.66 |
| rs1500867 | C | G | 0.75 | -0.025 | 0.004 | 1.90E-08 | -0.001 | 0.002 | 0.626 | 31.59 |
| rs17279437 | G | A | 0.89 | 0.027 | 0.004 | 8.14E-10 | 0.002 | 0.003 | 0.522 | 37.73 |
| rs192856660 | G | T | 0.79 | -0.032 | 0.004 | 1.56E-12 | 0.017 | 0.002 | 8.16E-16 | 49.97 |
| rs1948948 | C | T | 0.56 | -0.033 | 0.004 | 1.90E-14 | 0.002 | 0.002 | 0.237 | 58.63 |
| rs2514536 | C | G | 0.37 | 0.027 | 0.004 | 2.47E-10 | -0.002 | 0.002 | 0.236 | 40.05 |
| rs4131861 | G | A | 0.85 | 0.025 | 0.004 | 1.16E-08 | 0.002 | 0.002 | 0.472 | 32.56 |
| rs4794895 | C | T | 0.54 | -0.026 | 0.004 | 4.03E-09 | 0.001 | 0.002 | 0.497 | 34.61 |
| rs62434144 | C | T | 0.59 | 0.024 | 0.004 | 4.40E-08 | -0.001 | 0.002 | 0.389 | 29.96 |
| rs9933149 | T | C | 0.38 | 0.029 | 0.004 | 2.53E-11 | -0.001 | 0.002 | 0.515 | 44.51 |

**SNPs used as instrumental variables in the MR analysis of intracellular volume fraction on educational attainment**

| **SNP** | **Effect allele** | **Other allele** | **Effect allele frequency** | **beta (exposure)** | **SE (exposure)** | ***P* (exposure)** | **beta (outcome)** | **SE (outcome)** | ***P* (outcome)** | ***F*-statistic** |
| --- | --- | --- | --- | --- | --- | --- | --- | --- | --- | --- |
| rs1228024 | C | A | 0.33 | 0.036 | 0.006 | 2.72E-08 | -0.003 | 0.002 | 0.058 | 30.9 |
| rs13164785 | T | G | 0.81 | 0.079 | 0.008 | 2.90E-25 | 0 | 0.002 | 0.994 | 107.85 |
| rs2863973 | A | C | 0.83 | 0.044 | 0.008 | 4.96E-08 | 0.007 | 0.002 | 9.91E-04 | 29.73 |
| rs3795736 | T | C | 0.7 | -0.039 | 0.007 | 2.28E-09 | -0.001 | 0.002 | 0.693 | 35.72 |
| rs5756922 | C | T | 0.58 | 0.034 | 0.006 | 3.05E-08 | 0.005 | 0.002 | 0.008 | 30.68 |
| rs74504435 | A | G | 0.91 | 0.078 | 0.01 | 9.33E-14 | -0.005 | 0.003 | 0.096 | 55.5 |
| rs9272456 | T | A | 0.8 | -0.043 | 0.008 | 1.47E-08 | 0.008 | 0.002 | 3.16E-04 | 32.09 |
| rs9286062 | G | A | 0.45 | 0.034 | 0.006 | 4.21E-08 | -0.001 | 0.002 | 0.596 | 30.05 |

**SNPs used as instrumental variables in the MR analysis of orientation dispersion index on educational attainment**

| **SNP** | **Effect allele** | **Other allele** | **Effect allele frequency** | **beta (exposure)** | **SE (exposure)** | ***P* (exposure)** | **beta (outcome)** | **SE (outcome)** | ***P* (outcome)** | ***F*-statistic** |
| --- | --- | --- | --- | --- | --- | --- | --- | --- | --- | --- |
| rs200261326 | C | A | 0.5 | 0.044 | 0.008 | 5.35E-09 | -0.007 | 0.002 | 3.38E-05 | 34.06 |
| rs7219015 | C | T | 0.78 | 0.054 | 0.009 | 3.78E-09 | 0.008 | 0.002 | 1.29E-04 | 34.73 |
| rs79172804 | G | A | 0.82 | 0.085 | 0.01 | 6.72E-18 | 0.018 | 0.002 | 5.58E-15 | 74.3 |
| rs9308343 | G | A | 0.42 | 0.052 | 0.008 | 8.79E-12 | -0.002 | 0.002 | 0.363 | 46.58 |

**SNPs used as instrumental variables in the MR analysis of fractional anisotropy (white matter tracts) on educational attainment**

| **SNP** | **Effect allele** | **Other allele** | **Effect allele frequency** | **beta (exposure)** | **SE (exposure)** | ***P* (exposure)** | **beta (outcome)** | **SE (outcome)** | ***P* (outcome)** | ***F*-statistic** |
| --- | --- | --- | --- | --- | --- | --- | --- | --- | --- | --- |
| rs10052710 | G | T | 0.19 | -0.137 | 0.009 | 1.34E-50 | 0 | 0.002 | 0.835 | 223.81 |
| rs11557085 | T | C | 0.47 | -0.043 | 0.007 | 6.92E-09 | -0.009 | 0.002 | 6.68E-08 | 33.56 |
| rs12735232 | C | T | 0.16 | 0.054 | 0.01 | 4.85E-08 | 0.013 | 0.002 | 1.86E-08 | 29.78 |
| rs142003765 | G | A | 0.02 | -0.15 | 0.027 | 2.00E-08 | 0.003 | 0.006 | 0.687 | 31.49 |
| rs1479569 | T | C | 0.43 | 0.043 | 0.007 | 3.98E-09 | -0.004 | 0.002 | 0.016 | 34.63 |
| rs179570 | G | A | 0.47 | 0.046 | 0.007 | 3.07E-10 | 0.008 | 0.002 | 7.43E-06 | 39.63 |
| rs186399184 | T | C | 0.12 | 0.063 | 0.011 | 2.93E-08 | 0.003 | 0.003 | 0.325 | 30.75 |
| rs1867667 | G | A | 0.26 | 0.066 | 0.008 | 2.07E-15 | -0.001 | 0.002 | 0.738 | 63 |
| rs199501 | A | G | 0.23 | -0.06 | 0.009 | 3.18E-12 | -0.014 | 0.002 | 1.55E-12 | 48.57 |
| rs2018980 | A | G | 0.37 | -0.051 | 0.008 | 1.53E-11 | -0.003 | 0.002 | 0.13 | 45.49 |
| rs2267161 | T | C | 0.31 | 0.046 | 0.008 | 6.95E-09 | 0.004 | 0.002 | 0.029 | 33.55 |
| rs2412397 | C | T | 0.37 | 0.052 | 0.008 | 5.20E-12 | 0 | 0.002 | 0.992 | 47.61 |
| rs28361101 | C | G | 0.03 | 0.13 | 0.023 | 1.15E-08 | -0.001 | 0.005 | 0.875 | 32.57 |
| rs3795736 | C | T | 0.3 | 0.044 | 0.008 | 2.97E-08 | 0.001 | 0.002 | 0.693 | 30.73 |
| rs4150221 | C | T | 0.27 | 0.056 | 0.008 | 8.62E-12 | 0 | 0.002 | 0.872 | 46.62 |
| rs4984741 | A | G | 0.26 | -0.048 | 0.008 | 8.38E-09 | -0.01 | 0.002 | 4.91E-07 | 33.19 |
| rs55705857 | G | A | 0.06 | -0.111 | 0.016 | 2.82E-12 | -0.004 | 0.004 | 0.295 | 48.81 |
| rs56362600 | A | G | 0.2 | 0.053 | 0.009 | 8.38E-09 | 0.004 | 0.002 | 0.036 | 33.19 |
| rs6062264 | T | C | 0.28 | -0.051 | 0.008 | 5.53E-10 | -0.001 | 0.002 | 0.704 | 38.48 |
| rs62086884 | C | G | 0.2 | -0.051 | 0.009 | 3.51E-08 | -0.003 | 0.002 | 0.235 | 30.4 |
| rs7590302 | C | T | 0.31 | 0.056 | 0.008 | 1.90E-12 | -0.003 | 0.002 | 0.063 | 49.59 |
| rs911547 | G | A | 0.14 | -0.066 | 0.011 | 6.44E-10 | 0.003 | 0.002 | 0.236 | 38.18 |
| rs9481189 | C | T | 0.14 | 0.059 | 0.011 | 2.09E-08 | -0.005 | 0.002 | 0.07 | 31.41 |

**SNPs used as instrumental variables in the MR analysis of mean diffusivity (white matter tracts) on educational attainment**

| **SNP** | **Effect allele** | **Other allele** | **Effect allele frequency** | **beta (exposure)** | **SE (exposure)** | ***P* (exposure)** | **beta (outcome)** | **SE (outcome)** | ***P* (outcome)** | ***F*-statistic** |
| --- | --- | --- | --- | --- | --- | --- | --- | --- | --- | --- |
| rs10052710 | G | T | 0.19 | 0.149 | 0.009 | 2.75E-63 | 0 | 0.002 | 0.835 | 282 |
| rs11128657 | C | T | 0.41 | -0.06 | 0.007 | 9.59E-17 | -0.001 | 0.002 | 0.417 | 69.05 |
| rs11133252 | A | T | 0.37 | -0.053 | 0.007 | 5.25E-13 | 0 | 0.002 | 0.873 | 52.11 |
| rs1479569 | T | C | 0.43 | -0.039 | 0.007 | 3.86E-08 | -0.004 | 0.002 | 0.016 | 30.22 |
| rs1948948 | T | C | 0.44 | 0.053 | 0.007 | 1.32E-13 | -0.002 | 0.002 | 0.237 | 54.82 |
| rs225694 | A | G | 0.23 | -0.046 | 0.008 | 4.93E-08 | -0.004 | 0.002 | 0.054 | 29.74 |
| rs2488002 | C | T | 0.15 | 0.067 | 0.01 | 7.43E-12 | 0.003 | 0.002 | 0.282 | 46.91 |
| rs2841614 | C | T | 0.07 | -0.076 | 0.014 | 3.17E-08 | 0.003 | 0.003 | 0.349 | 30.6 |
| rs33599 | A | G | 0.26 | -0.067 | 0.008 | 1.01E-16 | -0.001 | 0.002 | 0.667 | 68.95 |
| rs34399834 | A | G | 0.04 | 0.104 | 0.018 | 7.08E-09 | 0.001 | 0.004 | 0.807 | 33.51 |
| rs372295105 | C | T | 0.29 | 0.049 | 0.008 | 9.12E-10 | 0 | 0.002 | 0.941 | 37.5 |
| rs3793477 | C | G | 0.27 | 0.044 | 0.008 | 3.28E-08 | -0.002 | 0.002 | 0.207 | 30.53 |
| rs4150221 | C | T | 0.27 | -0.053 | 0.008 | 4.42E-11 | 0 | 0.002 | 0.872 | 43.42 |
| rs4862340 | A | G | 0.39 | -0.051 | 0.007 | 2.66E-12 | 0.001 | 0.002 | 0.441 | 48.93 |
| rs4894803 | G | A | 0.41 | 0.046 | 0.007 | 3.02E-10 | -0.003 | 0.002 | 0.119 | 39.66 |
| rs55705857 | G | A | 0.06 | 0.1 | 0.015 | 7.79E-11 | -0.004 | 0.004 | 0.295 | 42.31 |
| rs62071596 | C | T | 0.18 | 0.071 | 0.01 | 3.33E-13 | -0.017 | 0.002 | 4.28E-15 | 53 |
| rs62086884 | C | G | 0.2 | 0.059 | 0.009 | 3.06E-11 | -0.003 | 0.002 | 0.235 | 44.14 |
| rs77126132 | A | G | 0.09 | 0.084 | 0.012 | 9.48E-12 | 0.005 | 0.003 | 0.098 | 46.43 |
| rs7935166 | A | G | 0.13 | 0.083 | 0.01 | 1.25E-15 | -0.004 | 0.003 | 0.151 | 63.99 |
| rs8082186 | C | T | 0.32 | -0.046 | 0.008 | 2.15E-09 | -0.003 | 0.002 | 0.124 | 35.83 |
| rs9692176 | G | T | 0.41 | -0.052 | 0.007 | 2.30E-12 | 0.003 | 0.002 | 0.128 | 49.21 |

**SNPs used as instrumental variables in the MR analysis of volume of left hippocampus on educational attainment**

| **SNP** | **Effect allele** | **Other allele** | **Effect allele frequency** | **beta (exposure)** | **SE (exposure)** | ***P* (exposure)** | **beta (outcome)** | **SE (outcome)** | ***P* (outcome)** | ***F*-statistic** |
| --- | --- | --- | --- | --- | --- | --- | --- | --- | --- | --- |
| rs150434736 | T | C | 0.36 | 0.051 | 0.008 | 9.69E-10 | -0.002 | 0.002 | 0.385 | 37.41 |
| rs17178006 | G | T | 0.12 | -0.099 | 0.012 | 1.57E-16 | 0.004 | 0.003 | 0.145 | 68.16 |
| rs188402627 | A | T | 0.09 | 0.134 | 0.014 | 8.00E-22 | 0.008 | 0.003 | 0.003 | 92.28 |
| rs2970932 | G | A | 0.57 | -0.05 | 0.008 | 2.34E-10 | 0.008 | 0.002 | 8.60E-07 | 40.19 |
| rs3800230 | G | T | 0.12 | -0.067 | 0.012 | 1.95E-08 | -0.009 | 0.003 | 2.34E-04 | 31.57 |
| rs6690450 | A | C | 0.51 | -0.053 | 0.008 | 6.52E-12 | 0.005 | 0.002 | 0.007 | 47.2 |
| rs7040792 | C | T | 0.37 | 0.06 | 0.008 | 8.85E-14 | 0 | 0.002 | 0.833 | 55.66 |

**SNPs used as instrumental variables in the MR analysis of volume of right hippocampus on educational attainment**

| **SNP** | **Effect allele** | **Other allele** | **Effect allele frequency** | **beta (exposure)** | **SE (exposure)** | ***P* (exposure)** | **beta (outcome)** | **SE (outcome)** | ***P* (outcome)** | ***F*-statistic** |
| --- | --- | --- | --- | --- | --- | --- | --- | --- | --- | --- |
| rs10888849 | A | G | 0.49 | -0.044 | 0.008 | 8.80E-09 | 0.004 | 0.002 | 0.023 | 33.11 |
| rs118125918 | G | A | 0.02 | 0.139 | 0.025 | 3.76E-08 | 0.004 | 0.006 | 0.459 | 30.28 |
| rs12523793 | A | G | 0.33 | 0.048 | 0.008 | 4.51E-09 | 0 | 0.002 | 0.915 | 34.41 |
| rs150434736 | T | C | 0.36 | 0.058 | 0.008 | 5.28E-12 | -0.002 | 0.002 | 0.385 | 47.61 |
| rs2732706 | T | C | 0.22 | -0.061 | 0.009 | 9.71E-11 | -0.017 | 0.002 | 8.16E-16 | 41.91 |
| rs6432708 | T | C | 0.58 | -0.051 | 0.008 | 1.27E-10 | 0.008 | 0.002 | 8.20E-06 | 41.37 |
| rs7020341 | C | G | 0.36 | 0.05 | 0.008 | 4.65E-10 | 0.001 | 0.002 | 0.692 | 38.84 |
| rs73123652 | C | T | 0.11 | -0.085 | 0.012 | 5.62E-12 | 0.003 | 0.003 | 0.301 | 47.49 |
| rs73220476 | T | C | 0.15 | 0.08 | 0.011 | 7.29E-13 | 0.005 | 0.002 | 0.048 | 51.51 |

**SNPs used as instrumental variables in the MR analysis of total volume of white matter hyperintensities on educational attainment**

| **SNP** | **Effect allele** | **Other allele** | **Effect allele frequency** | **beta (exposure)** | **SE (exposure)** | ***P* (exposure)** | **beta (outcome)** | **SE (outcome)** | ***P* (outcome)** | ***F*-statistic** |
| --- | --- | --- | --- | --- | --- | --- | --- | --- | --- | --- |
| rs10445411 | T | C | 0.8 | 0.067 | 0.01 | 1.26E-11 | 0.011 | 0.002 | 3.83E-07 | 45.9 |
| rs11838776 | A | G | 0.28 | 0.057 | 0.009 | 1.13E-10 | 0.003 | 0.002 | 0.106 | 41.61 |
| rs12130849 | T | C | 0.03 | -0.13 | 0.023 | 3.00E-08 | 0.008 | 0.005 | 0.114 | 30.72 |
| rs12928520 | T | C | 0.44 | 0.068 | 0.008 | 3.34E-17 | -0.001 | 0.002 | 0.575 | 71.21 |
| rs17149723 | A | G | 0.29 | -0.058 | 0.009 | 3.72E-11 | 0.001 | 0.002 | 0.741 | 43.78 |
| rs17576323 | C | T | 0.2 | -0.059 | 0.01 | 2.44E-09 | 0.004 | 0.002 | 0.047 | 35.61 |
| rs17683692 | T | A | 0.17 | -0.067 | 0.011 | 2.83E-10 | 0.006 | 0.002 | 0.011 | 39.82 |
| rs1948948 | T | C | 0.44 | -0.066 | 0.008 | 9.31E-17 | -0.002 | 0.002 | 0.237 | 69.19 |
| rs3744020 | A | G | 0.19 | 0.145 | 0.01 | 4.90E-47 | 0.001 | 0.002 | 0.772 | 208.17 |
| rs4525538 | G | A | 0.63 | 0.072 | 0.008 | 2.17E-18 | 0.003 | 0.002 | 0.082 | 76.62 |
| rs4630220 | A | G | 0.28 | -0.062 | 0.009 | 2.82E-12 | 0.006 | 0.002 | 0.003 | 48.84 |
| rs554632245 | A | G | 0.24 | -0.084 | 0.009 | 6.11E-19 | 0.001 | 0.002 | 0.559 | 79.13 |
| rs62137163 | T | C | 0.18 | -0.064 | 0.01 | 7.56E-10 | 0.001 | 0.002 | 0.633 | 37.89 |
| rs6482385 | T | C | 0.51 | 0.044 | 0.008 | 2.88E-08 | 0.005 | 0.002 | 0.002 | 30.8 |
| rs6940540 | G | T | 0.4 | 0.072 | 0.008 | 4.95E-19 | 0.001 | 0.002 | 0.44 | 79.55 |
| rs7199787 | T | C | 0.22 | 0.059 | 0.01 | 1.19E-09 | 0 | 0.002 | 0.821 | 37.01 |
| rs7202450 | G | T | 0.54 | -0.046 | 0.008 | 8.02E-09 | 0 | 0.002 | 0.857 | 33.29 |
| rs7596872 | A | C | 0.1 | 0.146 | 0.013 | 3.06E-28 | -0.003 | 0.003 | 0.322 | 121.67 |
| rs76122535 | G | C | 0.14 | -0.069 | 0.012 | 2.43E-09 | 0.003 | 0.002 | 0.278 | 35.62 |
| rs7733216 | C | T | 0.2 | 0.086 | 0.01 | 2.34E-18 | 0 | 0.002 | 0.967 | 76.48 |

**SNPs used as instrumental variables in the MR analysis of surface area on late-onset Alzheimer's disease**

| **SNP** | **Effect allele** | **Other allele** | **Effect allele frequency** | **beta (exposure)** | **SE (exposure)** | ***P* (exposure)** | **beta (outcome)** | **SE (outcome)** | ***P* (outcome)** | ***F*-statistic** |
| --- | --- | --- | --- | --- | --- | --- | --- | --- | --- | --- |
| rs10460587 | C | T | 0.46 | -0.043 | 0.006 | 2.22E-11 | 0.006 | 0.014 | 0.652 | 44.76 |
| rs10496091 | G | A | 0.72 | 0.045 | 0.007 | 2.55E-10 | 0.03 | 0.016 | 0.054 | 39.99 |
| rs111939932 | C | T | 0.82 | -0.059 | 0.008 | 1.67E-12 | -0.045 | 0.019 | 0.017 | 49.84 |
| rs11759026 | A | G | 0.77 | -0.08 | 0.008 | 8.34E-26 | 0.018 | 0.017 | 0.31 | 110.32 |
| rs1802669 | G | A | 0.66 | 0.04 | 0.007 | 2.73E-09 | 0.02 | 0.015 | 0.179 | 35.37 |
| rs182905872 | G | A | 0.96 | 0.102 | 0.017 | 1.84E-09 | 0.027 | 0.044 | 0.529 | 36.13 |
| rs2301718 | G | A | 0.79 | -0.05 | 0.008 | 2.74E-10 | 0.014 | 0.017 | 0.43 | 39.85 |
| rs2764264 | C | T | 0.31 | -0.047 | 0.007 | 1.26E-11 | -0.031 | 0.015 | 0.045 | 45.87 |
| rs35664429 | G | A | 0.95 | 0.087 | 0.015 | 5.33E-09 | -0.037 | 0.038 | 0.338 | 34.06 |
| rs3786824 | G | A | 0.67 | -0.043 | 0.007 | 1.83E-10 | 0.033 | 0.016 | 0.043 | 40.64 |
| rs3789283 | G | A | 0.42 | -0.04 | 0.006 | 6.65E-10 | -0.004 | 0.015 | 0.773 | 38.12 |
| rs4685 | T | C | 0.33 | 0.043 | 0.007 | 3.31E-10 | -0.012 | 0.016 | 0.436 | 39.49 |
| rs56269136 | C | T | 0.78 | 0.112 | 0.008 | 7.72E-49 | 0.04 | 0.018 | 0.023 | 215.73 |
| rs62283132 | C | T | 0.85 | -0.07 | 0.009 | 5.41E-15 | 0.019 | 0.02 | 0.348 | 61.11 |
| rs7297175 | T | C | 0.43 | 0.045 | 0.006 | 2.38E-12 | 0.004 | 0.015 | 0.786 | 49.14 |
| rs76169231 | T | C | 0.8 | 0.048 | 0.008 | 8.95E-10 | 0.018 | 0.02 | 0.384 | 37.54 |
| rs7671110 | C | T | 0.84 | 0.057 | 0.009 | 1.01E-10 | -0.041 | 0.019 | 0.031 | 41.79 |
| rs76928645 | C | T | 0.9 | -0.091 | 0.011 | 5.93E-18 | 0.045 | 0.024 | 0.062 | 74.54 |
| rs7715167 | T | C | 0.39 | -0.043 | 0.007 | 4.19E-11 | 0.006 | 0.015 | 0.707 | 43.52 |
| rs7797990 | A | G | 0.62 | -0.042 | 0.007 | 1.44E-10 | -0.02 | 0.015 | 0.184 | 41.11 |
| rs78852495 | G | A | 0.97 | -0.097 | 0.017 | 2.92E-08 | -0.017 | 0.039 | 0.668 | 30.76 |
| rs8756 | C | A | 0.49 | 0.054 | 0.006 | 1.51E-17 | -0.011 | 0.014 | 0.437 | 72.69 |
| rs9266618 | C | A | 0.09 | 0.073 | 0.011 | 1.35E-10 | 0.038 | 0.028 | 0.178 | 41.24 |

**SNPs used as instrumental variables in the MR analysis of volume on late-onset Alzheimer's disease**

| **SNP** | **Effect allele** | **Other allele** | **Effect allele frequency** | **beta (exposure)** | **SE (exposure)** | ***P* (exposure)** | **beta (outcome)** | **SE (outcome)** | ***P* (outcome)** | ***F*-statistic** |
| --- | --- | --- | --- | --- | --- | --- | --- | --- | --- | --- |
| rs10460587 | C | T | 0.46 | -0.036 | 0.006 | 1.09E-08 | 0.006 | 0.014 | 0.652 | 32.67 |
| rs10496091 | G | A | 0.72 | 0.041 | 0.007 | 2.62E-09 | 0.03 | 0.016 | 0.054 | 35.45 |
| rs111939932 | C | T | 0.82 | -0.048 | 0.008 | 3.72E-09 | -0.045 | 0.019 | 0.017 | 34.76 |
| rs1163249 | A | G | 0.2 | 0.052 | 0.008 | 3.02E-11 | 0 | 0.018 | 1 | 44.16 |
| rs13097248 | A | G | 0.85 | 0.049 | 0.009 | 1.97E-08 | -0.002 | 0.02 | 0.936 | 31.53 |
| rs149549868 | G | A | 0.98 | -0.114 | 0.02 | 1.96E-08 | 0.047 | 0.05 | 0.349 | 31.53 |
| rs1979477 | A | C | 0.5 | -0.04 | 0.006 | 9.51E-11 | 0.02 | 0.014 | 0.172 | 41.92 |
| rs2301718 | G | A | 0.79 | -0.05 | 0.008 | 7.28E-11 | 0.014 | 0.017 | 0.43 | 42.44 |
| rs2529418 | G | A | 0.44 | 0.038 | 0.006 | 1.83E-09 | -0.002 | 0.015 | 0.904 | 36.15 |
| rs3130346 | T | C | 0.15 | 0.051 | 0.009 | 7.77E-09 | 0.074 | 0.022 | 6.99E-04 | 33.33 |
| rs3200031 | C | T | 0.92 | -0.075 | 0.012 | 7.70E-11 | 0.005 | 0.027 | 0.848 | 42.33 |
| rs3789283 | G | A | 0.42 | -0.038 | 0.006 | 1.61E-09 | -0.004 | 0.015 | 0.773 | 36.39 |
| rs4259003 | G | A | 0.81 | -0.043 | 0.008 | 4.48E-08 | 0.017 | 0.018 | 0.355 | 29.93 |
| rs4273712 | A | G | 0.73 | -0.067 | 0.007 | 8.14E-22 | 0.011 | 0.016 | 0.512 | 92.12 |
| rs62057121 | G | A | 0.79 | 0.072 | 0.008 | 1.57E-21 | 0.038 | 0.018 | 0.032 | 90.82 |
| rs7297175 | T | C | 0.43 | 0.036 | 0.006 | 1.18E-08 | 0.004 | 0.015 | 0.786 | 32.52 |
| rs73100947 | C | T | 0.85 | 0.049 | 0.009 | 1.25E-08 | -0.04 | 0.019 | 0.035 | 32.41 |
| rs75745313 | G | A | 0.93 | 0.073 | 0.012 | 9.75E-10 | 0.005 | 0.042 | 0.903 | 37.37 |
| rs76169231 | T | C | 0.8 | 0.051 | 0.008 | 2.90E-11 | 0.018 | 0.02 | 0.384 | 44.24 |
| rs7715167 | T | C | 0.39 | -0.037 | 0.006 | 7.71E-09 | 0.006 | 0.015 | 0.707 | 33.35 |
| rs7986738 | A | G | 0.79 | -0.042 | 0.008 | 4.89E-08 | 0.002 | 0.017 | 0.892 | 29.76 |
| rs8756 | C | A | 0.49 | 0.052 | 0.006 | 6.00E-17 | -0.011 | 0.014 | 0.437 | 69.98 |
| rs9398171 | C | T | 0.29 | -0.052 | 0.007 | 2.67E-14 | -0.035 | 0.016 | 0.026 | 57.96 |
| rs9509982 | A | G | 0.28 | -0.038 | 0.007 | 2.90E-08 | 0.023 | 0.016 | 0.155 | 30.77 |

**SNPs used as instrumental variables in the MR analysis of cortical thickness on late-onset Alzheimer's disease**

| **SNP** | **Effect allele** | **Other allele** | **Effect allele frequency** | **beta (exposure)** | **SE (exposure)** | ***P* (exposure)** | **beta (outcome)** | **SE (outcome)** | ***P* (outcome)** | ***F*-statistic** |
| --- | --- | --- | --- | --- | --- | --- | --- | --- | --- | --- |
| rs12150230 | A | G | 0.78 | -0.065 | 0.008 | 3.94E-17 | 0.042 | 0.018 | 0.02 | 70.81 |
| rs13079368 | C | T | 0.48 | -0.046 | 0.006 | 7.29E-13 | 0.018 | 0.014 | 0.224 | 51.46 |
| rs13107325 | C | T | 0.93 | 0.085 | 0.013 | 1.03E-11 | -0.02 | 0.027 | 0.461 | 46.28 |
| rs143893586 | T | G | 0.94 | -0.076 | 0.014 | 2.76E-08 | -0.023 | 0.036 | 0.531 | 30.87 |
| rs1687225 | G | A | 0.77 | -0.045 | 0.008 | 3.54E-09 | 0.02 | 0.018 | 0.252 | 34.86 |
| rs2241722 | C | A | 0.67 | 0.064 | 0.007 | 1.06E-20 | 0.032 | 0.016 | 0.042 | 87.05 |
| rs3200031 | C | T | 0.92 | -0.091 | 0.012 | 3.19E-14 | 0.005 | 0.027 | 0.848 | 57.61 |
| rs614225 | A | G | 0.53 | -0.035 | 0.006 | 4.14E-08 | 0.001 | 0.014 | 0.923 | 30.08 |
| rs61753077 | G | A | 0.95 | -0.088 | 0.015 | 6.55E-09 | -0.047 | 0.038 | 0.218 | 33.66 |
| rs617577 | T | C | 0.9 | 0.08 | 0.011 | 8.78E-14 | -0.034 | 0.025 | 0.174 | 55.62 |
| rs6492273 | T | C | 0.41 | 0.041 | 0.007 | 5.21E-10 | -0.001 | 0.015 | 0.936 | 38.6 |
| rs6939 | A | C | 0.29 | 0.043 | 0.007 | 1.81E-09 | -0.029 | 0.016 | 0.072 | 36.17 |
| rs7219015 | C | T | 0.78 | -0.063 | 0.008 | 6.78E-16 | -0.037 | 0.024 | 0.12 | 65.2 |
| rs7224765 | G | A | 0.08 | 0.069 | 0.012 | 5.55E-09 | -0.006 | 0.027 | 0.82 | 33.98 |
| rs724265 | G | A | 0.38 | -0.039 | 0.007 | 9.06E-09 | -0.001 | 0.015 | 0.936 | 33.03 |
| rs72618187 | T | C | 0.76 | -0.044 | 0.008 | 5.62E-09 | -0.02 | 0.018 | 0.263 | 33.96 |
| rs76928645 | C | T | 0.9 | 0.06 | 0.011 | 2.06E-08 | 0.045 | 0.024 | 0.062 | 31.44 |
| rs78783493 | C | T | 0.91 | 0.074 | 0.011 | 4.80E-11 | 0.012 | 0.035 | 0.742 | 43.26 |
| rs79274348 | C | T | 0.95 | -0.084 | 0.015 | 1.87E-08 | 0.044 | 0.038 | 0.246 | 31.62 |
| rs934447 | A | G | 0.26 | 0.043 | 0.008 | 2.76E-08 | -0.012 | 0.017 | 0.474 | 30.87 |
| rs9921406 | C | T | 0.91 | -0.065 | 0.011 | 1.36E-08 | -0.016 | 0.026 | 0.556 | 32.24 |

**SNPs used as instrumental variables in the MR analysis of local gyrification index on late-onset Alzheimer's disease**

| **SNP** | **Effect allele** | **Other allele** | **Effect allele frequency** | **beta (exposure)** | **SE (exposure)** | ***P* (exposure)** | **beta (outcome)** | **SE (outcome)** | ***P* (outcome)** | ***F*-statistic** |
| --- | --- | --- | --- | --- | --- | --- | --- | --- | --- | --- |
| rs10460587 | C | T | 0.46 | -0.043 | 0.008 | 1.90E-08 | 0.006 | 0.014 | 0.652 | 31.6 |
| rs11633076 | C | T | 0.51 | 0.045 | 0.008 | 3.96E-09 | -0.024 | 0.014 | 0.098 | 34.64 |
| rs11759026 | A | G | 0.77 | -0.065 | 0.009 | 2.39E-13 | 0.018 | 0.017 | 0.31 | 53.65 |
| rs12938775 | G | A | 0.5 | -0.056 | 0.007 | 8.46E-14 | 0.004 | 0.015 | 0.772 | 55.7 |
| rs147431626 | G | A | 0.78 | 0.081 | 0.009 | 1.94E-19 | 0.048 | 0.02 | 0.014 | 81.3 |
| rs2047676 | C | T | 0.7 | -0.049 | 0.008 | 2.89E-09 | 0.015 | 0.016 | 0.319 | 35.26 |
| rs2501261 | A | G | 0.23 | 0.049 | 0.009 | 2.15E-08 | -0.01 | 0.017 | 0.559 | 31.36 |
| rs3814345 | G | A | 0.82 | -0.057 | 0.01 | 3.64E-09 | -0.006 | 0.018 | 0.74 | 34.81 |
| rs6658111 | T | G | 0.64 | -0.077 | 0.008 | 7.49E-23 | 0.02 | 0.016 | 0.226 | 96.85 |
| rs7146019 | A | G | 0.43 | -0.047 | 0.008 | 1.20E-09 | -0.004 | 0.015 | 0.797 | 36.96 |
| rs7484851 | A | G | 0.5 | -0.041 | 0.007 | 2.96E-08 | 0.002 | 0.014 | 0.894 | 30.74 |
| rs7715167 | T | C | 0.39 | -0.046 | 0.008 | 1.55E-09 | 0.006 | 0.015 | 0.707 | 36.47 |
| rs999867 | C | T | 0.92 | 0.081 | 0.014 | 1.01E-08 | 0.008 | 0.025 | 0.738 | 32.83 |

**SNPs used as instrumental variables in the MR analysis of mean curvature on late-onset Alzheimer's disease**

| **SNP** | **Effect allele** | **Other allele** | **Effect allele frequency** | **beta (exposure)** | **SE (exposure)** | ***P* (exposure)** | **beta (outcome)** | **SE (outcome)** | ***P* (outcome)** | ***F*-statistic** |
| --- | --- | --- | --- | --- | --- | --- | --- | --- | --- | --- |
| rs10197724 | C | T | 0.46 | 0.039 | 0.007 | 3.31E-08 | 0.012 | 0.017 | 0.482 | 30.52 |
| rs1048527 | T | G | 0.92 | 0.096 | 0.013 | 9.97E-14 | 0.004 | 0.027 | 0.87 | 55.37 |
| rs114113354 | A | G | 0.74 | 0.046 | 0.008 | 1.17E-08 | -0.05 | 0.017 | 0.003 | 32.53 |
| rs116269521 | G | A | 0.97 | -0.109 | 0.019 | 1.33E-08 | -0.011 | 0.048 | 0.818 | 32.28 |
| rs11781155 | C | T | 0.43 | -0.04 | 0.007 | 1.43E-08 | 0.006 | 0.015 | 0.675 | 32.15 |
| rs12146713 | T | C | 0.9 | 0.073 | 0.012 | 6.83E-10 | -0.017 | 0.025 | 0.504 | 38.07 |
| rs13012151 | C | T | 0.3 | 0.043 | 0.008 | 1.51E-08 | -0.044 | 0.016 | 0.005 | 32.04 |
| rs13264372 | C | T | 0.63 | -0.043 | 0.007 | 2.09E-09 | -0.004 | 0.015 | 0.799 | 35.89 |
| rs17205972 | G | T | 0.81 | 0.05 | 0.009 | 1.53E-08 | -0.001 | 0.018 | 0.941 | 32.02 |
| rs17206779 | C | T | 0.48 | 0.041 | 0.007 | 4.05E-09 | 0.013 | 0.014 | 0.367 | 34.6 |
| rs17424787 | G | A | 0.92 | -0.135 | 0.013 | 5.92E-25 | -0.001 | 0.028 | 0.972 | 106.43 |
| rs2241722 | C | A | 0.67 | -0.041 | 0.007 | 3.01E-08 | 0.032 | 0.016 | 0.042 | 30.7 |
| rs34636718 | A | G | 0.98 | -0.191 | 0.027 | 7.50E-13 | 0.031 | 0.071 | 0.663 | 51.41 |
| rs4652758 | T | C | 0.46 | 0.04 | 0.007 | 2.42E-08 | 0.003 | 0.014 | 0.825 | 31.13 |
| rs55976523 | C | T | 0.73 | -0.046 | 0.008 | 1.37E-08 | -0.019 | 0.019 | 0.311 | 32.23 |
| rs56125329 | C | T | 0.56 | -0.047 | 0.007 | 3.19E-11 | -0.015 | 0.015 | 0.302 | 44.05 |
| rs59682551 | T | G | 0.76 | -0.05 | 0.008 | 8.63E-10 | -0.019 | 0.017 | 0.266 | 37.61 |
| rs613511 | C | T | 0.74 | -0.055 | 0.008 | 1.88E-11 | -0.024 | 0.017 | 0.155 | 45.09 |
| rs61789085 | A | G | 0.72 | 0.048 | 0.008 | 6.93E-10 | 0.008 | 0.016 | 0.619 | 38.04 |
| rs6745807 | A | C | 0.36 | -0.049 | 0.007 | 1.31E-11 | 0 | 0.015 | 0.992 | 45.8 |
| rs7219015 | C | T | 0.78 | 0.067 | 0.009 | 4.92E-15 | -0.037 | 0.024 | 0.12 | 61.29 |
| rs748593 | C | T | 0.31 | -0.042 | 0.008 | 2.45E-08 | 0.004 | 0.019 | 0.837 | 31.1 |
| rs7559029 | C | A | 0.46 | -0.041 | 0.007 | 4.06E-09 | -0.003 | 0.014 | 0.832 | 34.6 |
| rs756747 | T | G | 0.48 | -0.043 | 0.007 | 1.12E-09 | 0.01 | 0.015 | 0.518 | 37.1 |
| rs7685018 | G | A | 0.54 | -0.041 | 0.007 | 6.51E-09 | 0.019 | 0.015 | 0.195 | 33.68 |
| rs9398172 | G | A | 0.29 | 0.052 | 0.008 | 1.07E-11 | -0.035 | 0.016 | 0.024 | 46.19 |

**SNPs used as instrumental variables in the MR analysis of intrinsic curvature on late-onset Alzheimer's disease**

| **SNP** | **Effect allele** | **Other allele** | **Effect allele frequency** | **beta (exposure)** | **SE (exposure)** | ***P* (exposure)** | **beta (outcome)** | **SE (outcome)** | ***P* (outcome)** | ***F*-statistic** |
| --- | --- | --- | --- | --- | --- | --- | --- | --- | --- | --- |
| rs11134697 | G | A | 0.45 | -0.032 | 0.005 | 5.50E-11 | -0.002 | 0.015 | 0.907 | 42.99 |
| rs11157930 | T | G | 0.39 | -0.028 | 0.005 | 1.27E-08 | 0.014 | 0.015 | 0.335 | 32.37 |
| rs11674612 | A | G | 0.64 | 0.028 | 0.005 | 2.48E-08 | 0.002 | 0.016 | 0.907 | 31.08 |
| rs11686881 | C | T | 0.64 | -0.031 | 0.005 | 1.65E-09 | 0.01 | 0.015 | 0.501 | 36.35 |
| rs11759026 | A | G | 0.77 | -0.045 | 0.006 | 5.38E-15 | 0.018 | 0.017 | 0.31 | 61.12 |
| rs12938775 | G | A | 0.5 | -0.034 | 0.005 | 4.89E-12 | 0.004 | 0.015 | 0.772 | 47.73 |
| rs147431626 | G | A | 0.78 | 0.056 | 0.006 | 9.89E-22 | 0.048 | 0.02 | 0.014 | 91.74 |
| rs1802669 | G | A | 0.66 | 0.031 | 0.005 | 1.76E-09 | 0.02 | 0.015 | 0.179 | 36.23 |
| rs182905872 | G | A | 0.96 | 0.077 | 0.013 | 3.63E-09 | 0.027 | 0.044 | 0.529 | 34.81 |
| rs201872292 | G | A | 0.92 | -0.051 | 0.009 | 9.52E-09 | -0.012 | 0.026 | 0.656 | 32.94 |
| rs2241722 | C | A | 0.67 | -0.034 | 0.005 | 3.92E-11 | 0.032 | 0.016 | 0.042 | 43.65 |
| rs34216221 | T | C | 0.98 | -0.114 | 0.019 | 2.44E-09 | 0.109 | 0.062 | 0.076 | 35.58 |
| rs4912174 | A | G | 0.89 | -0.044 | 0.008 | 1.33E-08 | 0.008 | 0.023 | 0.73 | 32.28 |
| rs6778085 | C | T | 0.19 | 0.034 | 0.006 | 4.20E-08 | -0.01 | 0.018 | 0.597 | 30.06 |
| rs76928645 | C | T | 0.9 | -0.057 | 0.008 | 1.12E-12 | 0.045 | 0.024 | 0.062 | 50.62 |
| rs7694606 | G | A | 0.84 | 0.041 | 0.007 | 5.91E-10 | -0.041 | 0.019 | 0.035 | 38.35 |
| rs9930713 | A | G | 0.76 | -0.031 | 0.006 | 4.43E-08 | -0.017 | 0.017 | 0.308 | 29.95 |

**SNPs used as instrumental variables in the MR analysis of mean diffusivity (cortical) on late-onset Alzheimer's disease**

| **SNP** | **Effect allele** | **Other allele** | **Effect allele frequency** | **beta (exposure)** | **SE (exposure)** | ***P* (exposure)** | **beta (outcome)** | **SE (outcome)** | ***P* (outcome)** | ***F*-statistic** |
| --- | --- | --- | --- | --- | --- | --- | --- | --- | --- | --- |
| rs12048319 | G | T | 0.84 | 0.024 | 0.004 | 3.33E-08 | -0.016 | 0.02 | 0.427 | 30.5 |
| rs13224758 | G | A | 0.85 | 0.028 | 0.004 | 1.95E-10 | -0.024 | 0.021 | 0.26 | 40.51 |
| rs17279437 | G | A | 0.89 | 0.027 | 0.004 | 8.14E-10 | 0.009 | 0.025 | 0.72 | 37.73 |
| rs192856660 | G | T | 0.79 | -0.032 | 0.004 | 1.56E-12 | 0.046 | 0.02 | 0.024 | 49.97 |
| rs1948948 | C | T | 0.56 | -0.033 | 0.004 | 1.90E-14 | -0.007 | 0.015 | 0.633 | 58.63 |
| rs4131861 | G | A | 0.85 | 0.025 | 0.004 | 1.16E-08 | 0.015 | 0.02 | 0.461 | 32.56 |
| rs4794895 | C | T | 0.54 | -0.026 | 0.004 | 4.03E-09 | 0.007 | 0.015 | 0.642 | 34.61 |
| rs62434144 | C | T | 0.59 | 0.024 | 0.004 | 4.40E-08 | -0.024 | 0.015 | 0.103 | 29.96 |
| rs9933149 | T | C | 0.38 | 0.029 | 0.004 | 2.53E-11 | -0.012 | 0.015 | 0.429 | 44.51 |

**SNPs used as instrumental variables in the MR analysis of intracellular volume fraction on late-onset Alzheimer's disease**

| **SNP** | **Effect allele** | **Other allele** | **Effect allele frequency** | **beta (exposure)** | **SE (exposure)** | ***P* (exposure)** | **beta (outcome)** | **SE (outcome)** | ***P* (outcome)** | ***F*-statistic** |
| --- | --- | --- | --- | --- | --- | --- | --- | --- | --- | --- |
| rs1228024 | C | A | 0.33 | 0.036 | 0.006 | 2.72E-08 | 0.028 | 0.015 | 0.066 | 30.9 |
| rs13164785 | T | G | 0.81 | 0.079 | 0.008 | 2.90E-25 | -0.001 | 0.018 | 0.938 | 107.85 |
| rs2863973 | A | C | 0.83 | 0.044 | 0.008 | 4.96E-08 | 0.03 | 0.019 | 0.115 | 29.73 |
| rs3795736 | T | C | 0.7 | -0.039 | 0.007 | 2.28E-09 | 0.013 | 0.017 | 0.436 | 35.72 |
| rs5756922 | C | T | 0.58 | 0.034 | 0.006 | 3.05E-08 | -0.024 | 0.015 | 0.103 | 30.68 |
| rs74504435 | A | G | 0.91 | 0.078 | 0.01 | 9.33E-14 | 0.033 | 0.025 | 0.189 | 55.5 |
| rs9286062 | G | A | 0.45 | 0.034 | 0.006 | 4.21E-08 | -0.002 | 0.015 | 0.909 | 30.05 |

**SNPs used as instrumental variables in the MR analysis of orientation dispersion index on late-onset Alzheimer's disease**

| **SNP** | **Effect allele** | **Other allele** | **Effect allele frequency** | **beta (exposure)** | **SE (exposure)** | ***P* (exposure)** | **beta (outcome)** | **SE (outcome)** | ***P* (outcome)** | ***F*-statistic** |
| --- | --- | --- | --- | --- | --- | --- | --- | --- | --- | --- |
| rs200261326 | C | A | 0.5 | 0.044 | 0.008 | 5.35E-09 | -0.002 | 0.018 | 0.902 | 34.06 |
| rs7219015 | C | T | 0.78 | 0.054 | 0.009 | 3.78E-09 | -0.037 | 0.024 | 0.12 | 34.73 |
| rs79172804 | G | A | 0.82 | 0.085 | 0.01 | 6.72E-18 | 0.039 | 0.018 | 0.033 | 74.3 |
| rs9308343 | G | A | 0.42 | 0.052 | 0.008 | 8.79E-12 | -0.02 | 0.015 | 0.184 | 46.58 |

**SNPs used as instrumental variables in the MR analysis of fractional anisotropy (white matter tracts) on late-onset Alzheimer's disease**

| **SNP** | **Effect allele** | **Other allele** | **Effect allele frequency** | **beta (exposure)** | **SE (exposure)** | ***P* (exposure)** | **beta (outcome)** | **SE (outcome)** | ***P* (outcome)** | ***F*-statistic** |
| --- | --- | --- | --- | --- | --- | --- | --- | --- | --- | --- |
| rs10052710 | G | T | 0.19 | -0.137 | 0.009 | 1.34E-50 | -0.002 | 0.018 | 0.912 | 223.81 |
| rs11557085 | T | C | 0.47 | -0.043 | 0.007 | 6.92E-09 | 0.003 | 0.015 | 0.827 | 33.56 |
| rs12735232 | C | T | 0.16 | 0.054 | 0.01 | 4.85E-08 | 0.016 | 0.02 | 0.43 | 29.78 |
| rs142003765 | G | A | 0.02 | -0.15 | 0.027 | 2.00E-08 | 0.022 | 0.058 | 0.696 | 31.49 |
| rs1479569 | T | C | 0.43 | 0.043 | 0.007 | 3.98E-09 | 0 | 0.014 | 0.994 | 34.63 |
| rs179570 | G | A | 0.47 | 0.046 | 0.007 | 3.07E-10 | -0.005 | 0.014 | 0.701 | 39.63 |
| rs186399184 | T | C | 0.12 | 0.063 | 0.011 | 2.93E-08 | -0.058 | 0.021 | 0.006 | 30.75 |
| rs1867667 | G | A | 0.26 | 0.066 | 0.008 | 2.07E-15 | -0.006 | 0.016 | 0.715 | 63 |
| rs199501 | A | G | 0.23 | -0.06 | 0.009 | 3.18E-12 | -0.037 | 0.017 | 0.03 | 48.57 |
| rs2018980 | A | G | 0.37 | -0.051 | 0.008 | 1.53E-11 | -0.01 | 0.015 | 0.493 | 45.49 |
| rs2267161 | T | C | 0.31 | 0.046 | 0.008 | 6.95E-09 | 0.021 | 0.015 | 0.17 | 33.55 |
| rs2412397 | C | T | 0.37 | 0.052 | 0.008 | 5.20E-12 | 0.011 | 0.015 | 0.447 | 47.61 |
| rs3795736 | C | T | 0.3 | 0.044 | 0.008 | 2.97E-08 | -0.013 | 0.017 | 0.436 | 30.73 |
| rs4150221 | C | T | 0.27 | 0.056 | 0.008 | 8.62E-12 | 0.056 | 0.016 | 5.09E-04 | 46.62 |
| rs4984741 | A | G | 0.26 | -0.048 | 0.008 | 8.38E-09 | 0.015 | 0.017 | 0.368 | 33.19 |
| rs55705857 | G | A | 0.06 | -0.111 | 0.016 | 2.82E-12 | 0.062 | 0.042 | 0.135 | 48.81 |
| rs56362600 | A | G | 0.2 | 0.053 | 0.009 | 8.38E-09 | -0.037 | 0.019 | 0.05 | 33.19 |
| rs6062264 | T | C | 0.28 | -0.051 | 0.008 | 5.53E-10 | 0.001 | 0.018 | 0.97 | 38.48 |
| rs7590302 | C | T | 0.31 | 0.056 | 0.008 | 1.90E-12 | -0.013 | 0.015 | 0.406 | 49.59 |
| rs911547 | G | A | 0.14 | -0.066 | 0.011 | 6.44E-10 | -0.022 | 0.02 | 0.264 | 38.18 |
| rs9481189 | C | T | 0.14 | 0.059 | 0.011 | 2.09E-08 | 0.055 | 0.02 | 0.006 | 31.41 |

**SNPs used as instrumental variables in the MR analysis of mean diffusivity (white matter tracts) on late-onset Alzheimer's disease**

| **SNP** | **Effect allele** | **Other allele** | **Effect allele frequency** | **beta (exposure)** | **SE (exposure)** | ***P* (exposure)** | **beta (outcome)** | **SE (outcome)** | ***P* (outcome)** | ***F*-statistic** |
| --- | --- | --- | --- | --- | --- | --- | --- | --- | --- | --- |
| rs10052710 | G | T | 0.19 | 0.149 | 0.009 | 2.75E-63 | -0.002 | 0.018 | 0.912 | 282 |
| rs11128657 | C | T | 0.41 | -0.06 | 0.007 | 9.59E-17 | -0.014 | 0.015 | 0.329 | 69.05 |
| rs1479569 | T | C | 0.43 | -0.039 | 0.007 | 3.86E-08 | 0 | 0.014 | 0.994 | 30.22 |
| rs1948948 | T | C | 0.44 | 0.053 | 0.007 | 1.32E-13 | 0.007 | 0.015 | 0.633 | 54.82 |
| rs225694 | A | G | 0.23 | -0.046 | 0.008 | 4.93E-08 | 0.013 | 0.017 | 0.443 | 29.74 |
| rs2488002 | C | T | 0.15 | 0.067 | 0.01 | 7.43E-12 | -0.012 | 0.019 | 0.527 | 46.91 |
| rs2841614 | C | T | 0.07 | -0.076 | 0.014 | 3.17E-08 | 0.057 | 0.027 | 0.037 | 30.6 |
| rs3116836 | C | A | 0.24 | -0.057 | 0.008 | 9.12E-12 | -0.016 | 0.02 | 0.419 | 46.51 |
| rs33599 | A | G | 0.26 | -0.067 | 0.008 | 1.01E-16 | -0.004 | 0.016 | 0.809 | 68.95 |
| rs34399834 | A | G | 0.04 | 0.104 | 0.018 | 7.08E-09 | 0.021 | 0.036 | 0.573 | 33.51 |
| rs372295105 | C | T | 0.29 | 0.049 | 0.008 | 9.12E-10 | -0.022 | 0.016 | 0.16 | 37.5 |
| rs4150221 | C | T | 0.27 | -0.053 | 0.008 | 4.42E-11 | 0.056 | 0.016 | 5.09E-04 | 43.42 |
| rs4862340 | A | G | 0.39 | -0.051 | 0.007 | 2.66E-12 | 0.004 | 0.015 | 0.772 | 48.93 |
| rs4894803 | G | A | 0.41 | 0.046 | 0.007 | 3.02E-10 | 0.02 | 0.015 | 0.157 | 39.66 |
| rs55705857 | G | A | 0.06 | 0.1 | 0.015 | 7.79E-11 | 0.062 | 0.042 | 0.135 | 42.31 |
| rs62071596 | C | T | 0.18 | 0.071 | 0.01 | 3.33E-13 | -0.064 | 0.027 | 0.02 | 53 |
| rs77126132 | A | G | 0.09 | 0.084 | 0.012 | 9.48E-12 | -0.034 | 0.025 | 0.181 | 46.43 |
| rs7935166 | A | G | 0.13 | 0.083 | 0.01 | 1.25E-15 | -0.062 | 0.021 | 0.003 | 63.99 |
| rs8082186 | C | T | 0.32 | -0.046 | 0.008 | 2.15E-09 | 0.014 | 0.015 | 0.376 | 35.83 |

**SNPs used as instrumental variables in the MR analysis of volume of left hippocampus on late-onset Alzheimer's disease**

| **SNP** | **Effect allele** | **Other allele** | **Effect allele frequency** | **beta (exposure)** | **SE (exposure)** | ***P* (exposure)** | **beta (outcome)** | **SE (outcome)** | ***P* (outcome)** | ***F*-statistic** |
| --- | --- | --- | --- | --- | --- | --- | --- | --- | --- | --- |
| rs150434736 | T | C | 0.36 | 0.051 | 0.008 | 9.69E-10 | 0.077 | 0.024 | 0.001 | 37.41 |
| rs17178006 | G | T | 0.12 | -0.099 | 0.012 | 1.57E-16 | -0.049 | 0.027 | 0.072 | 68.16 |
| rs2970932 | G | A | 0.57 | -0.05 | 0.008 | 2.34E-10 | -0.016 | 0.015 | 0.271 | 40.19 |
| rs3800230 | G | T | 0.12 | -0.067 | 0.012 | 1.95E-08 | -0.042 | 0.022 | 0.055 | 31.57 |
| rs593626 | G | A | 0.81 | -0.056 | 0.01 | 2.25E-08 | 0.02 | 0.022 | 0.351 | 31.29 |
| rs6690450 | A | C | 0.51 | -0.053 | 0.008 | 6.52E-12 | 0.027 | 0.014 | 0.053 | 47.2 |
| rs7040792 | C | T | 0.37 | 0.06 | 0.008 | 8.85E-14 | 0.002 | 0.015 | 0.898 | 55.66 |

**SNPs used as instrumental variables in the MR analysis of volume of right hippocampus on late-onset Alzheimer's disease**

| **SNP** | **Effect allele** | **Other allele** | **Effect allele frequency** | **beta (exposure)** | **SE (exposure)** | ***P* (exposure)** | **beta (outcome)** | **SE (outcome)** | ***P* (outcome)** | ***F*-statistic** |
| --- | --- | --- | --- | --- | --- | --- | --- | --- | --- | --- |
| rs10888849 | A | G | 0.49 | -0.044 | 0.008 | 8.80E-09 | 0.022 | 0.014 | 0.123 | 33.11 |
| rs118125918 | G | A | 0.02 | 0.139 | 0.025 | 3.76E-08 | -0.041 | 0.064 | 0.518 | 30.28 |
| rs12523793 | A | G | 0.33 | 0.048 | 0.008 | 4.51E-09 | 0.006 | 0.015 | 0.679 | 34.41 |
| rs150434736 | T | C | 0.36 | 0.058 | 0.008 | 5.28E-12 | 0.077 | 0.024 | 0.001 | 47.61 |
| rs2732706 | T | C | 0.22 | -0.061 | 0.009 | 9.71E-11 | -0.044 | 0.02 | 0.026 | 41.91 |
| rs6432708 | T | C | 0.58 | -0.051 | 0.008 | 1.27E-10 | -0.015 | 0.015 | 0.296 | 41.37 |
| rs73123652 | C | T | 0.11 | -0.085 | 0.012 | 5.62E-12 | -0.064 | 0.028 | 0.023 | 47.49 |
| rs73220476 | T | C | 0.15 | 0.08 | 0.011 | 7.29E-13 | 0.029 | 0.022 | 0.186 | 51.51 |

**SNPs used as instrumental variables in the MR analysis of total volume of white matter hyperintensities on late-onset Alzheimer's disease**

| **SNP** | **Effect allele** | **Other allele** | **Effect allele frequency** | **beta (exposure)** | **SE (exposure)** | ***P* (exposure)** | **beta (outcome)** | **SE (outcome)** | ***P* (outcome)** | ***F*-statistic** |
| --- | --- | --- | --- | --- | --- | --- | --- | --- | --- | --- |
| rs10445411 | T | C | 0.8 | 0.067 | 0.01 | 1.26E-11 | 0.002 | 0.018 | 0.911 | 45.9 |
| rs11838776 | A | G | 0.28 | 0.057 | 0.009 | 1.13E-10 | -0.038 | 0.016 | 0.018 | 41.61 |
| rs12130849 | T | C | 0.03 | -0.13 | 0.023 | 3.00E-08 | -0.011 | 0.049 | 0.819 | 30.72 |
| rs12928520 | T | C | 0.44 | 0.068 | 0.008 | 3.34E-17 | 0.005 | 0.016 | 0.742 | 71.21 |
| rs17149723 | A | G | 0.29 | -0.058 | 0.009 | 3.72E-11 | 0.005 | 0.016 | 0.731 | 43.78 |
| rs17576323 | C | T | 0.2 | -0.059 | 0.01 | 2.44E-09 | -0.044 | 0.018 | 0.013 | 35.61 |
| rs1948948 | T | C | 0.44 | -0.066 | 0.008 | 9.31E-17 | 0.007 | 0.015 | 0.633 | 69.19 |
| rs3744020 | A | G | 0.19 | 0.145 | 0.01 | 4.90E-47 | -0.018 | 0.018 | 0.336 | 208.17 |
| rs4525538 | G | A | 0.63 | 0.072 | 0.008 | 2.17E-18 | 0.001 | 0.015 | 0.966 | 76.62 |
| rs4630220 | A | G | 0.28 | -0.062 | 0.009 | 2.82E-12 | -0.024 | 0.016 | 0.144 | 48.84 |
| rs554632245 | A | G | 0.24 | -0.084 | 0.009 | 6.11E-19 | -0.005 | 0.016 | 0.776 | 79.13 |
| rs62137163 | T | C | 0.18 | -0.064 | 0.01 | 7.56E-10 | 0.009 | 0.019 | 0.642 | 37.89 |
| rs6482385 | T | C | 0.51 | 0.044 | 0.008 | 2.88E-08 | -0.021 | 0.014 | 0.137 | 30.8 |
| rs6940540 | G | T | 0.4 | 0.072 | 0.008 | 4.95E-19 | 0.025 | 0.014 | 0.084 | 79.55 |
| rs7199787 | T | C | 0.22 | 0.059 | 0.01 | 1.19E-09 | 0.041 | 0.018 | 0.026 | 37.01 |
| rs7202450 | G | T | 0.54 | -0.046 | 0.008 | 8.02E-09 | -0.014 | 0.014 | 0.343 | 33.29 |
| rs7596872 | A | C | 0.1 | 0.146 | 0.013 | 3.06E-28 | 0.003 | 0.024 | 0.916 | 121.67 |
| rs7733216 | C | T | 0.2 | 0.086 | 0.01 | 2.34E-18 | -0.004 | 0.018 | 0.845 | 76.48 |

**SNPs used as instrumental variables in the MR analysis of late-onset Alzheimer's disease on surface area**

| **SNP** | **Effect allele** | **Other allele** | **Effect allele frequency** | **beta (exposure)** | **SE (exposure)** | ***P* (exposure)** | **beta (outcome)** | **SE (outcome)** | ***P* (outcome)** | ***F*-statistic** |
| --- | --- | --- | --- | --- | --- | --- | --- | --- | --- | --- |
| rs10808026 | A | C | 0.2 | -0.101 | 0.016 | 1.30E-10 | 0.002 | 0.008 | 0.75 | 41.06 |
| rs10933431 | G | C | 0.22 | -0.094 | 0.016 | 3.42E-09 | -0.003 | 0.008 | 0.692 | 34.88 |
| rs11218343 | C | T | 0.04 | -0.22 | 0.032 | 2.88E-12 | 0.025 | 0.017 | 0.145 | 48.91 |
| rs114812713 | C | G | 0.03 | 0.278 | 0.039 | 2.10E-13 | -0.01 | 0.021 | 0.646 | 51.4 |
| rs12539172 | T | C | 0.3 | -0.082 | 0.013 | 9.26E-10 | 0.012 | 0.007 | 0.077 | 37.72 |
| rs12721046 | A | G | NA | 1.006 | 0.02 | 1.00E-200 | 0.004 | 0.009 | 0.611 | 2606.18 |
| rs12881735 | C | T | 0.22 | -0.086 | 0.015 | 7.35E-09 | -0.016 | 0.008 | 0.028 | 33.39 |
| rs138190086 | A | G | 0.02 | 0.262 | 0.045 | 5.30E-09 | -0.012 | 0.024 | 0.607 | 33.88 |
| rs17125924 | G | A | 0.09 | 0.128 | 0.021 | 1.42E-09 | 0.013 | 0.011 | 0.242 | 36.74 |
| rs2830500 | A | C | 0.31 | -0.073 | 0.014 | 2.60E-08 | 0.007 | 0.007 | 0.325 | 28.29 |
| rs34665982 | T | C | NA | 0.097 | 0.017 | 5.80E-09 | 0.013 | 0.006 | 0.052 | 33.93 |
| rs35912475 | T | C | NA | -0.104 | 0.016 | 1.72E-10 | -0.011 | 0.006 | 0.088 | 40.63 |
| rs3740688 | G | T | 0.45 | -0.089 | 0.012 | 5.46E-13 | -0.006 | 0.006 | 0.336 | 52 |
| rs3752246 | G | C | 0.18 | 0.136 | 0.017 | 3.06E-16 | 0.003 | 0.008 | 0.762 | 66.61 |
| rs3851179 | T | C | 0.36 | -0.13 | 0.013 | 6.03E-25 | 0.025 | 0.007 | 1.11E-04 | 107.11 |
| rs4844610 | A | C | 0.19 | 0.157 | 0.016 | 3.60E-24 | 0.002 | 0.008 | 0.803 | 102.47 |
| rs57537848 | T | G | NA | 0.207 | 0.015 | 2.11E-42 | 0.004 | 0.006 | 0.493 | 187.2 |
| rs593742 | G | A | 0.3 | -0.073 | 0.011 | 6.80E-09 | 0.022 | 0.007 | 0.002 | 43.73 |
| rs6024870 | A | G | 0.09 | -0.123 | 0.022 | 3.46E-08 | 0.014 | 0.011 | 0.203 | 30.37 |
| rs62039712 | A | G | 0.12 | 0.148 | 0.028 | 3.70E-08 | 0.009 | 0.011 | 0.378 | 27.13 |
| rs6733839 | T | C | 0.41 | 0.181 | 0.013 | 2.05E-44 | -0.004 | 0.007 | 0.511 | 194.71 |
| rs7185636 | C | T | 0.18 | -0.083 | 0.017 | 2.40E-08 | 0.011 | 0.008 | 0.187 | 25.1 |
| rs73223431 | T | C | 0.37 | 0.096 | 0.013 | 6.30E-14 | -0.008 | 0.007 | 0.22 | 56.48 |
| rs77301115 | A | G | NA | 0.949 | 0.043 | 2.25E-109 | 0.015 | 0.021 | 0.458 | 493.53 |
| rs7920721 | G | A | 0.39 | 0.077 | 0.012 | 1.80E-11 | 0.009 | 0.007 | 0.18 | 42.84 |
| rs7933202 | C | A | 0.39 | -0.113 | 0.013 | 1.91E-19 | -0.014 | 0.007 | 0.032 | 81.72 |
| rs9331896 | C | T | 0.39 | -0.132 | 0.013 | 4.63E-24 | 0 | 0.006 | 0.948 | 103.1 |
| rs9473117 | C | A | 0.28 | 0.087 | 0.014 | 1.16E-10 | -0.004 | 0.007 | 0.545 | 41.82 |

**SNPs used as instrumental variables in the MR analysis of late-onset Alzheimer's disease on volume**

| **SNP** | **Effect allele** | **Other allele** | **Effect allele frequency** | **beta (exposure)** | **SE (exposure)** | ***P* (exposure)** | **beta (outcome)** | **SE (outcome)** | ***P* (outcome)** | ***F*-statistic** |
| --- | --- | --- | --- | --- | --- | --- | --- | --- | --- | --- |
| rs10808026 | A | C | 0.2 | -0.101 | 0.016 | 1.30E-10 | -0.002 | 0.008 | 0.817 | 41.06 |
| rs10933431 | G | C | 0.22 | -0.094 | 0.016 | 3.42E-09 | -0.001 | 0.007 | 0.846 | 34.88 |
| rs11218343 | C | T | 0.04 | -0.22 | 0.032 | 2.88E-12 | 0.004 | 0.016 | 0.807 | 48.91 |
| rs114812713 | C | G | 0.03 | 0.278 | 0.039 | 2.10E-13 | -0.017 | 0.02 | 0.399 | 51.4 |
| rs12539172 | T | C | 0.3 | -0.082 | 0.013 | 9.26E-10 | 0.009 | 0.007 | 0.192 | 37.72 |
| rs12721046 | A | G | NA | 1.006 | 0.02 | 1.00E-200 | 0 | 0.009 | 0.954 | 2606.18 |
| rs12881735 | C | T | 0.22 | -0.086 | 0.015 | 7.35E-09 | -0.012 | 0.007 | 0.099 | 33.39 |
| rs138190086 | A | G | 0.02 | 0.262 | 0.045 | 5.30E-09 | 0 | 0.023 | 0.993 | 33.88 |
| rs17125924 | G | A | 0.09 | 0.128 | 0.021 | 1.42E-09 | 0.012 | 0.011 | 0.28 | 36.74 |
| rs2830500 | A | C | 0.31 | -0.073 | 0.014 | 2.60E-08 | 0.012 | 0.007 | 0.082 | 28.29 |
| rs34665982 | T | C | NA | 0.097 | 0.017 | 5.80E-09 | 0.01 | 0.006 | 0.111 | 33.93 |
| rs35912475 | T | C | NA | -0.104 | 0.016 | 1.72E-10 | -0.008 | 0.006 | 0.219 | 40.63 |
| rs3740688 | G | T | 0.45 | -0.089 | 0.012 | 5.46E-13 | -0.004 | 0.006 | 0.496 | 52 |
| rs3752246 | G | C | 0.18 | 0.136 | 0.017 | 3.06E-16 | -0.004 | 0.008 | 0.655 | 66.61 |
| rs3851179 | T | C | 0.36 | -0.13 | 0.013 | 6.03E-25 | 0.021 | 0.006 | 8.47E-04 | 107.11 |
| rs4844610 | A | C | 0.19 | 0.157 | 0.016 | 3.60E-24 | 0.004 | 0.008 | 0.654 | 102.47 |
| rs57537848 | T | G | NA | 0.207 | 0.015 | 2.11E-42 | 0.005 | 0.006 | 0.429 | 187.2 |
| rs593742 | G | A | 0.3 | -0.073 | 0.011 | 6.80E-09 | 0.009 | 0.007 | 0.194 | 43.73 |
| rs6024870 | A | G | 0.09 | -0.123 | 0.022 | 3.46E-08 | 0.008 | 0.011 | 0.461 | 30.37 |
| rs62039712 | A | G | 0.12 | 0.148 | 0.028 | 3.70E-08 | 0.003 | 0.01 | 0.797 | 27.13 |
| rs6733839 | T | C | 0.41 | 0.181 | 0.013 | 2.05E-44 | 0.006 | 0.007 | 0.333 | 194.71 |
| rs7185636 | C | T | 0.18 | -0.083 | 0.017 | 2.40E-08 | 0.019 | 0.008 | 0.018 | 25.1 |
| rs73223431 | T | C | 0.37 | 0.096 | 0.013 | 6.30E-14 | -0.008 | 0.006 | 0.188 | 56.48 |
| rs77301115 | A | G | NA | 0.949 | 0.043 | 2.25E-109 | -0.002 | 0.02 | 0.91 | 493.53 |
| rs7920721 | G | A | 0.39 | 0.077 | 0.012 | 1.80E-11 | 0.012 | 0.006 | 0.066 | 42.84 |
| rs7933202 | C | A | 0.39 | -0.113 | 0.013 | 1.91E-19 | -0.004 | 0.006 | 0.514 | 81.72 |
| rs9331896 | C | T | 0.39 | -0.132 | 0.013 | 4.63E-24 | 0.005 | 0.006 | 0.438 | 103.1 |
| rs9473117 | C | A | 0.28 | 0.087 | 0.014 | 1.16E-10 | -0.004 | 0.007 | 0.562 | 41.82 |

**SNPs used as instrumental variables in the MR analysis of late-onset Alzheimer's disease on cortical thickness**

| **SNP** | **Effect allele** | **Other allele** | **Effect allele frequency** | **beta (exposure)** | **SE (exposure)** | ***P* (exposure)** | **beta (outcome)** | **SE (outcome)** | ***P* (outcome)** | ***F*-statistic** |
| --- | --- | --- | --- | --- | --- | --- | --- | --- | --- | --- |
| rs10808026 | A | C | 0.2 | -0.101 | 0.016 | 1.30E-10 | -0.008 | 0.008 | 0.333 | 41.06 |
| rs10933431 | G | C | 0.22 | -0.094 | 0.016 | 3.42E-09 | 0.001 | 0.008 | 0.948 | 34.88 |
| rs11218343 | C | T | 0.04 | -0.22 | 0.032 | 2.88E-12 | -0.033 | 0.017 | 0.051 | 48.91 |
| rs114812713 | C | G | 0.03 | 0.278 | 0.039 | 2.10E-13 | -0.008 | 0.021 | 0.697 | 51.4 |
| rs12539172 | T | C | 0.3 | -0.082 | 0.013 | 9.26E-10 | -0.001 | 0.007 | 0.858 | 37.72 |
| rs12721046 | A | G | NA | 1.006 | 0.02 | 1.00E-200 | -0.008 | 0.009 | 0.353 | 2606.18 |
| rs12881735 | C | T | 0.22 | -0.086 | 0.015 | 7.35E-09 | 0.01 | 0.008 | 0.168 | 33.39 |
| rs138190086 | A | G | 0.02 | 0.262 | 0.045 | 5.30E-09 | 0.011 | 0.024 | 0.648 | 33.88 |
| rs17125924 | G | A | 0.09 | 0.128 | 0.021 | 1.42E-09 | 0.002 | 0.011 | 0.851 | 36.74 |
| rs2830500 | A | C | 0.31 | -0.073 | 0.014 | 2.60E-08 | 0.004 | 0.007 | 0.607 | 28.29 |
| rs34665982 | T | C | NA | 0.097 | 0.017 | 5.80E-09 | 0.002 | 0.007 | 0.735 | 33.93 |
| rs35912475 | T | C | NA | -0.104 | 0.016 | 1.72E-10 | 0.004 | 0.006 | 0.553 | 40.63 |
| rs3740688 | G | T | 0.45 | -0.089 | 0.012 | 5.46E-13 | 0.004 | 0.006 | 0.564 | 52 |
| rs3752246 | G | C | 0.18 | 0.136 | 0.017 | 3.06E-16 | -0.009 | 0.008 | 0.288 | 66.61 |
| rs3851179 | T | C | 0.36 | -0.13 | 0.013 | 6.03E-25 | -0.006 | 0.007 | 0.394 | 107.11 |
| rs4844610 | A | C | 0.19 | 0.157 | 0.016 | 3.60E-24 | 0.005 | 0.008 | 0.582 | 102.47 |
| rs57537848 | T | G | NA | 0.207 | 0.015 | 2.11E-42 | 0.001 | 0.007 | 0.919 | 187.2 |
| rs593742 | G | A | 0.3 | -0.073 | 0.011 | 6.80E-09 | -0.021 | 0.007 | 0.003 | 43.73 |
| rs6024870 | A | G | 0.09 | -0.123 | 0.022 | 3.46E-08 | -0.009 | 0.011 | 0.452 | 30.37 |
| rs62039712 | A | G | 0.12 | 0.148 | 0.028 | 3.70E-08 | -0.013 | 0.011 | 0.233 | 27.13 |
| rs6733839 | T | C | 0.41 | 0.181 | 0.013 | 2.05E-44 | 0.021 | 0.007 | 0.002 | 194.71 |
| rs7185636 | C | T | 0.18 | -0.083 | 0.017 | 2.40E-08 | 0.018 | 0.009 | 0.036 | 25.1 |
| rs73223431 | T | C | 0.37 | 0.096 | 0.013 | 6.30E-14 | -0.003 | 0.007 | 0.682 | 56.48 |
| rs77301115 | A | G | NA | 0.949 | 0.043 | 2.25E-109 | -0.039 | 0.021 | 0.062 | 493.53 |
| rs7920721 | G | A | 0.39 | 0.077 | 0.012 | 1.80E-11 | 0.008 | 0.007 | 0.245 | 42.84 |
| rs7933202 | C | A | 0.39 | -0.113 | 0.013 | 1.91E-19 | 0.018 | 0.007 | 0.007 | 81.72 |
| rs9331896 | C | T | 0.39 | -0.132 | 0.013 | 4.63E-24 | 0.009 | 0.007 | 0.189 | 103.1 |
| rs9473117 | C | A | 0.28 | 0.087 | 0.014 | 1.16E-10 | -0.003 | 0.007 | 0.693 | 41.82 |

**SNPs used as instrumental variables in the MR analysis of late-onset Alzheimer's disease on local gyrification index**

| **SNP** | **Effect allele** | **Other allele** | **Effect allele frequency** | **beta (exposure)** | **SE (exposure)** | ***P* (exposure)** | **beta (outcome)** | **SE (outcome)** | ***P* (outcome)** | ***F*-statistic** |
| --- | --- | --- | --- | --- | --- | --- | --- | --- | --- | --- |
| rs10808026 | A | C | 0.2 | -0.101 | 0.016 | 1.30E-10 | -0.007 | 0.009 | 0.454 | 41.06 |
| rs10933431 | G | C | 0.22 | -0.094 | 0.016 | 3.42E-09 | 0.005 | 0.009 | 0.555 | 34.88 |
| rs11218343 | C | T | 0.04 | -0.22 | 0.032 | 2.88E-12 | 0.042 | 0.02 | 0.032 | 48.91 |
| rs114812713 | C | G | 0.03 | 0.278 | 0.039 | 2.10E-13 | -0.021 | 0.025 | 0.395 | 51.4 |
| rs12539172 | T | C | 0.3 | -0.082 | 0.013 | 9.26E-10 | 0.008 | 0.008 | 0.305 | 37.72 |
| rs12721046 | A | G | NA | 1.006 | 0.02 | 1.00E-200 | 0.005 | 0.01 | 0.624 | 2606.18 |
| rs12881735 | C | T | 0.22 | -0.086 | 0.015 | 7.35E-09 | -0.027 | 0.009 | 0.002 | 33.39 |
| rs138190086 | A | G | 0.02 | 0.262 | 0.045 | 5.30E-09 | -0.005 | 0.028 | 0.865 | 33.88 |
| rs17125924 | G | A | 0.09 | 0.128 | 0.021 | 1.42E-09 | 0.01 | 0.013 | 0.428 | 36.74 |
| rs2830500 | A | C | 0.31 | -0.073 | 0.014 | 2.60E-08 | 0.013 | 0.008 | 0.125 | 28.29 |
| rs34665982 | T | C | NA | 0.097 | 0.017 | 5.80E-09 | -0.003 | 0.008 | 0.721 | 33.93 |
| rs35912475 | T | C | NA | -0.104 | 0.016 | 1.72E-10 | -0.007 | 0.008 | 0.358 | 40.63 |
| rs3740688 | G | T | 0.45 | -0.089 | 0.012 | 5.46E-13 | -0.015 | 0.008 | 0.044 | 52 |
| rs3752246 | G | C | 0.18 | 0.136 | 0.017 | 3.06E-16 | 0.015 | 0.01 | 0.114 | 66.61 |
| rs3851179 | T | C | 0.36 | -0.13 | 0.013 | 6.03E-25 | 0.021 | 0.008 | 0.006 | 107.11 |
| rs4844610 | A | C | 0.19 | 0.157 | 0.016 | 3.60E-24 | 0.005 | 0.01 | 0.59 | 102.47 |
| rs57537848 | T | G | NA | 0.207 | 0.015 | 2.11E-42 | -0.003 | 0.008 | 0.699 | 187.2 |
| rs593742 | G | A | 0.3 | -0.073 | 0.011 | 6.80E-09 | 0.01 | 0.008 | 0.208 | 43.73 |
| rs6024870 | A | G | 0.09 | -0.123 | 0.022 | 3.46E-08 | 0.016 | 0.013 | 0.241 | 30.37 |
| rs62039712 | A | G | 0.12 | 0.148 | 0.028 | 3.70E-08 | -0.001 | 0.013 | 0.928 | 27.13 |
| rs6733839 | T | C | 0.41 | 0.181 | 0.013 | 2.05E-44 | -0.011 | 0.008 | 0.149 | 194.71 |
| rs7185636 | C | T | 0.18 | -0.083 | 0.017 | 2.40E-08 | 0 | 0.01 | 0.999 | 25.1 |
| rs73223431 | T | C | 0.37 | 0.096 | 0.013 | 6.30E-14 | -0.005 | 0.008 | 0.522 | 56.48 |
| rs77301115 | A | G | NA | 0.949 | 0.043 | 2.25E-109 | 0.012 | 0.024 | 0.629 | 493.53 |
| rs7920721 | G | A | 0.39 | 0.077 | 0.012 | 1.80E-11 | 0.001 | 0.008 | 0.85 | 42.84 |
| rs7933202 | C | A | 0.39 | -0.113 | 0.013 | 1.91E-19 | -0.014 | 0.008 | 0.065 | 81.72 |
| rs9331896 | C | T | 0.39 | -0.132 | 0.013 | 4.63E-24 | 0.003 | 0.008 | 0.733 | 103.1 |
| rs9473117 | C | A | 0.28 | 0.087 | 0.014 | 1.16E-10 | -0.001 | 0.008 | 0.925 | 41.82 |

**SNPs used as instrumental variables in the MR analysis of late-onset Alzheimer's disease on mean curvature**

| **SNP** | **Effect allele** | **Other allele** | **Effect allele frequency** | **beta (exposure)** | **SE (exposure)** | ***P* (exposure)** | **beta (outcome)** | **SE (outcome)** | ***P* (outcome)** | ***F*-statistic** |
| --- | --- | --- | --- | --- | --- | --- | --- | --- | --- | --- |
| rs10808026 | A | C | 0.2 | -0.101 | 0.016 | 1.30E-10 | 0 | 0.009 | 0.957 | 41.06 |
| rs10933431 | G | C | 0.22 | -0.094 | 0.016 | 3.42E-09 | 0.006 | 0.008 | 0.459 | 34.88 |
| rs11218343 | C | T | 0.04 | -0.22 | 0.032 | 2.88E-12 | 0.033 | 0.018 | 0.076 | 48.91 |
| rs114812713 | C | G | 0.03 | 0.278 | 0.039 | 2.10E-13 | -0.023 | 0.023 | 0.309 | 51.4 |
| rs12539172 | T | C | 0.3 | -0.082 | 0.013 | 9.26E-10 | -0.017 | 0.007 | 0.023 | 37.72 |
| rs12721046 | A | G | NA | 1.006 | 0.02 | 1.00E-200 | -0.004 | 0.01 | 0.712 | 2606.18 |
| rs12881735 | C | T | 0.22 | -0.086 | 0.015 | 7.35E-09 | -0.003 | 0.008 | 0.684 | 33.39 |
| rs138190086 | A | G | 0.02 | 0.262 | 0.045 | 5.30E-09 | -0.049 | 0.026 | 0.058 | 33.88 |
| rs17125924 | G | A | 0.09 | 0.128 | 0.021 | 1.42E-09 | -0.014 | 0.012 | 0.24 | 36.74 |
| rs2830500 | A | C | 0.31 | -0.073 | 0.014 | 2.60E-08 | -0.001 | 0.008 | 0.894 | 28.29 |
| rs34665982 | T | C | NA | 0.097 | 0.017 | 5.80E-09 | -0.01 | 0.007 | 0.175 | 33.93 |
| rs35912475 | T | C | NA | -0.104 | 0.016 | 1.72E-10 | 0.003 | 0.007 | 0.696 | 40.63 |
| rs3740688 | G | T | 0.45 | -0.089 | 0.012 | 5.46E-13 | -0.009 | 0.007 | 0.198 | 52 |
| rs3752246 | G | C | 0.18 | 0.136 | 0.017 | 3.06E-16 | 0.016 | 0.009 | 0.072 | 66.61 |
| rs3851179 | T | C | 0.36 | -0.13 | 0.013 | 6.03E-25 | 0.001 | 0.007 | 0.911 | 107.11 |
| rs4844610 | A | C | 0.19 | 0.157 | 0.016 | 3.60E-24 | -0.001 | 0.009 | 0.945 | 102.47 |
| rs57537848 | T | G | NA | 0.207 | 0.015 | 2.11E-42 | 0.008 | 0.007 | 0.257 | 187.2 |
| rs593742 | G | A | 0.3 | -0.073 | 0.011 | 6.80E-09 | -0.008 | 0.008 | 0.281 | 43.73 |
| rs6024870 | A | G | 0.09 | -0.123 | 0.022 | 3.46E-08 | 0.002 | 0.012 | 0.87 | 30.37 |
| rs62039712 | A | G | 0.12 | 0.148 | 0.028 | 3.70E-08 | 0.014 | 0.012 | 0.223 | 27.13 |
| rs6733839 | T | C | 0.41 | 0.181 | 0.013 | 2.05E-44 | -0.015 | 0.007 | 0.042 | 194.71 |
| rs7185636 | C | T | 0.18 | -0.083 | 0.017 | 2.40E-08 | -0.016 | 0.009 | 0.084 | 25.1 |
| rs73223431 | T | C | 0.37 | 0.096 | 0.013 | 6.30E-14 | -0.005 | 0.007 | 0.506 | 56.48 |
| rs77301115 | A | G | NA | 0.949 | 0.043 | 2.25E-109 | -0.006 | 0.023 | 0.791 | 493.53 |
| rs7920721 | G | A | 0.39 | 0.077 | 0.012 | 1.80E-11 | -0.012 | 0.007 | 0.106 | 42.84 |
| rs7933202 | C | A | 0.39 | -0.113 | 0.013 | 1.91E-19 | -0.005 | 0.007 | 0.447 | 81.72 |
| rs9331896 | C | T | 0.39 | -0.132 | 0.013 | 4.63E-24 | -0.005 | 0.007 | 0.473 | 103.1 |
| rs9473117 | C | A | 0.28 | 0.087 | 0.014 | 1.16E-10 | 0.014 | 0.008 | 0.082 | 41.82 |

**SNPs used as instrumental variables in the MR analysis of late-onset Alzheimer's disease on intrinsic curvature**

| **SNP** | **Effect allele** | **Other allele** | **Effect allele frequency** | **beta (exposure)** | **SE (exposure)** | ***P* (exposure)** | **beta (outcome)** | **SE (outcome)** | ***P* (outcome)** | ***F*-statistic** |
| --- | --- | --- | --- | --- | --- | --- | --- | --- | --- | --- |
| rs10808026 | A | C | 0.2 | -0.101 | 0.016 | 1.30E-10 | -0.005 | 0.006 | 0.391 | 41.06 |
| rs10933431 | G | C | 0.22 | -0.094 | 0.016 | 3.42E-09 | 0.005 | 0.006 | 0.372 | 34.88 |
| rs11218343 | C | T | 0.04 | -0.22 | 0.032 | 2.88E-12 | 0.026 | 0.013 | 0.042 | 48.91 |
| rs114812713 | C | G | 0.03 | 0.278 | 0.039 | 2.10E-13 | 0.005 | 0.016 | 0.767 | 51.4 |
| rs12539172 | T | C | 0.3 | -0.082 | 0.013 | 9.26E-10 | 0.008 | 0.005 | 0.138 | 37.72 |
| rs12721046 | A | G | NA | 1.006 | 0.02 | 1.00E-200 | 0.001 | 0.007 | 0.933 | 2606.18 |
| rs12881735 | C | T | 0.22 | -0.086 | 0.015 | 7.35E-09 | -0.007 | 0.006 | 0.245 | 33.39 |
| rs138190086 | A | G | 0.02 | 0.262 | 0.045 | 5.30E-09 | -0.027 | 0.018 | 0.129 | 33.88 |
| rs17125924 | G | A | 0.09 | 0.128 | 0.021 | 1.42E-09 | 0.012 | 0.008 | 0.154 | 36.74 |
| rs2830500 | A | C | 0.31 | -0.073 | 0.014 | 2.60E-08 | 0.002 | 0.005 | 0.78 | 28.29 |
| rs34665982 | T | C | NA | 0.097 | 0.017 | 5.80E-09 | 0.009 | 0.005 | 0.075 | 33.93 |
| rs35912475 | T | C | NA | -0.104 | 0.016 | 1.72E-10 | -0.006 | 0.005 | 0.2 | 40.63 |
| rs3740688 | G | T | 0.45 | -0.089 | 0.012 | 5.46E-13 | -0.004 | 0.005 | 0.362 | 52 |
| rs3752246 | G | C | 0.18 | 0.136 | 0.017 | 3.06E-16 | 0.008 | 0.006 | 0.184 | 66.61 |
| rs3851179 | T | C | 0.36 | -0.13 | 0.013 | 6.03E-25 | 0.011 | 0.005 | 0.03 | 107.11 |
| rs4844610 | A | C | 0.19 | 0.157 | 0.016 | 3.60E-24 | 0.005 | 0.006 | 0.437 | 102.47 |
| rs57537848 | T | G | NA | 0.207 | 0.015 | 2.11E-42 | 0.008 | 0.005 | 0.095 | 187.2 |
| rs593742 | G | A | 0.3 | -0.073 | 0.011 | 6.80E-09 | 0.018 | 0.005 | 7.06E-04 | 43.73 |
| rs6024870 | A | G | 0.09 | -0.123 | 0.022 | 3.46E-08 | 0.015 | 0.009 | 0.075 | 30.37 |
| rs62039712 | A | G | 0.12 | 0.148 | 0.028 | 3.70E-08 | 0.014 | 0.008 | 0.095 | 27.13 |
| rs6733839 | T | C | 0.41 | 0.181 | 0.013 | 2.05E-44 | -0.011 | 0.005 | 0.031 | 194.71 |
| rs7185636 | C | T | 0.18 | -0.083 | 0.017 | 2.40E-08 | -0.006 | 0.006 | 0.324 | 25.1 |
| rs73223431 | T | C | 0.37 | 0.096 | 0.013 | 6.30E-14 | -0.004 | 0.005 | 0.472 | 56.48 |
| rs77301115 | A | G | NA | 0.949 | 0.043 | 2.25E-109 | 0.005 | 0.016 | 0.755 | 493.53 |
| rs7920721 | G | A | 0.39 | 0.077 | 0.012 | 1.80E-11 | 0.005 | 0.005 | 0.321 | 42.84 |
| rs7933202 | C | A | 0.39 | -0.113 | 0.013 | 1.91E-19 | -0.014 | 0.005 | 0.006 | 81.72 |
| rs9331896 | C | T | 0.39 | -0.132 | 0.013 | 4.63E-24 | -0.004 | 0.005 | 0.361 | 103.1 |
| rs9473117 | C | A | 0.28 | 0.087 | 0.014 | 1.16E-10 | -0.004 | 0.005 | 0.439 | 41.82 |

**SNPs used as instrumental variables in the MR analysis of late-onset Alzheimer's disease on intrinsic curvature**

| **SNP** | **Effect allele** | **Other allele** | **Effect allele frequency** | **beta (exposure)** | **SE (exposure)** | ***P* (exposure)** | **beta (outcome)** | **SE (outcome)** | ***P* (outcome)** | ***F*-statistic** |
| --- | --- | --- | --- | --- | --- | --- | --- | --- | --- | --- |
| rs10808026 | A | C | 0.2 | -0.101 | 0.016 | 1.30E-10 | 0.009 | 0.009 | 0.316 | 41.06 |
| rs10933431 | G | C | 0.22 | -0.094 | 0.016 | 3.42E-09 | -0.013 | 0.009 | 0.153 | 34.88 |
| rs11218343 | C | T | 0.04 | -0.22 | 0.032 | 2.88E-12 | -0.033 | 0.02 | 0.107 | 48.91 |
| rs114812713 | C | G | 0.03 | 0.278 | 0.039 | 2.10E-13 | -0.043 | 0.025 | 0.085 | 51.4 |
| rs12539172 | T | C | 0.3 | -0.082 | 0.013 | 9.26E-10 | -0.001 | 0.008 | 0.885 | 37.72 |
| rs12721046 | A | G | NA | 1.006 | 0.02 | 1.00E-200 | 0.009 | 0.011 | 0.41 | 2606.18 |
| rs12881735 | C | T | 0.22 | -0.086 | 0.015 | 7.35E-09 | -0.003 | 0.009 | 0.733 | 33.39 |
| rs138190086 | A | G | 0.02 | 0.262 | 0.045 | 5.30E-09 | -0.003 | 0.028 | 0.914 | 33.88 |
| rs17125924 | G | A | 0.09 | 0.128 | 0.021 | 1.42E-09 | -0.014 | 0.013 | 0.3 | 36.74 |
| rs2830500 | A | C | 0.31 | -0.073 | 0.014 | 2.60E-08 | 0.012 | 0.008 | 0.149 | 28.29 |
| rs34665982 | T | C | NA | 0.097 | 0.017 | 5.80E-09 | -0.013 | 0.008 | 0.089 | 33.93 |
| rs35912475 | T | C | NA | -0.104 | 0.016 | 1.72E-10 | 0.003 | 0.008 | 0.714 | 40.63 |
| rs3740688 | G | T | 0.45 | -0.089 | 0.012 | 5.46E-13 | 0.003 | 0.008 | 0.655 | 52 |
| rs3752246 | G | C | 0.18 | 0.136 | 0.017 | 3.06E-16 | 0.004 | 0.01 | 0.669 | 66.61 |
| rs3851179 | T | C | 0.36 | -0.13 | 0.013 | 6.03E-25 | -0.003 | 0.008 | 0.689 | 107.11 |
| rs4844610 | A | C | 0.19 | 0.157 | 0.016 | 3.60E-24 | 0.008 | 0.01 | 0.439 | 102.47 |
| rs57537848 | T | G | NA | 0.207 | 0.015 | 2.11E-42 | 0.006 | 0.008 | 0.455 | 187.2 |
| rs593742 | G | A | 0.3 | -0.073 | 0.011 | 6.80E-09 | -0.017 | 0.008 | 0.041 | 43.73 |
| rs6024870 | A | G | 0.09 | -0.123 | 0.022 | 3.46E-08 | 0.002 | 0.014 | 0.912 | 30.37 |
| rs62039712 | A | G | 0.12 | 0.148 | 0.028 | 3.70E-08 | -0.015 | 0.013 | 0.232 | 27.13 |
| rs6733839 | T | C | 0.41 | 0.181 | 0.013 | 2.05E-44 | 0.009 | 0.008 | 0.246 | 194.71 |
| rs7185636 | C | T | 0.18 | -0.083 | 0.017 | 2.40E-08 | -0.013 | 0.01 | 0.188 | 25.1 |
| rs73223431 | T | C | 0.37 | 0.096 | 0.013 | 6.30E-14 | 0.002 | 0.008 | 0.765 | 56.48 |
| rs77301115 | A | G | NA | 0.949 | 0.043 | 2.25E-109 | -0.032 | 0.025 | 0.203 | 493.53 |
| rs7920721 | G | A | 0.39 | 0.077 | 0.012 | 1.80E-11 | 0.001 | 0.008 | 0.882 | 42.84 |
| rs7933202 | C | A | 0.39 | -0.113 | 0.013 | 1.91E-19 | -0.023 | 0.008 | 0.003 | 81.72 |
| rs9331896 | C | T | 0.39 | -0.132 | 0.013 | 4.63E-24 | -0.001 | 0.008 | 0.93 | 103.1 |
| rs9473117 | C | A | 0.28 | 0.087 | 0.014 | 1.16E-10 | -0.006 | 0.009 | 0.509 | 41.82 |

**SNPs used as instrumental variables in the MR analysis of late-onset Alzheimer's disease on mean diffusivity (cortical)**

| **SNP** | **Effect allele** | **Other allele** | **Effect allele frequency** | **beta (exposure)** | **SE (exposure)** | ***P* (exposure)** | **beta (outcome)** | **SE (outcome)** | ***P* (outcome)** | ***F*-statistic** |
| --- | --- | --- | --- | --- | --- | --- | --- | --- | --- | --- |
| rs10808026 | A | C | 0.2 | -0.101 | 0.016 | 1.30E-10 | 0.004 | 0.004 | 0.392 | 41.06 |
| rs10933431 | G | C | 0.22 | -0.094 | 0.016 | 3.42E-09 | 0.006 | 0.004 | 0.181 | 34.88 |
| rs11218343 | C | T | 0.04 | -0.22 | 0.032 | 2.88E-12 | -0.001 | 0.004 | 0.848 | 48.91 |
| rs114812713 | C | G | 0.03 | 0.278 | 0.039 | 2.10E-13 | 0.007 | 0.004 | 0.09 | 51.4 |
| rs12539172 | T | C | 0.3 | -0.082 | 0.013 | 9.26E-10 | 0.000 | 0.004 | 0.943 | 37.72 |
| rs12721046 | A | G | NA | 1.006 | 0.02 | 1.00E-200 | 0.007 | 0.004 | 0.131 | 2606.18 |
| rs12881735 | C | T | 0.22 | -0.086 | 0.015 | 7.35E-09 | -0.001 | 0.004 | 0.822 | 33.39 |
| rs138190086 | A | G | 0.02 | 0.262 | 0.045 | 5.30E-09 | 0.006 | 0.004 | 0.199 | 33.88 |
| rs17125924 | G | A | 0.09 | 0.128 | 0.021 | 1.42E-09 | -0.003 | 0.004 | 0.538 | 36.74 |
| rs2830500 | A | C | 0.31 | -0.073 | 0.014 | 2.60E-08 | 0.003 | 0.004 | 0.55 | 28.29 |
| rs34665982 | T | C | NA | 0.097 | 0.017 | 5.80E-09 | 0.000 | 0.004 | 0.963 | 33.93 |
| rs35912475 | T | C | NA | -0.104 | 0.016 | 1.72E-10 | -0.001 | 0.004 | 0.832 | 40.63 |
| rs3740688 | G | T | 0.45 | -0.089 | 0.012 | 5.46E-13 | 0.004 | 0.004 | 0.344 | 52 |
| rs3752246 | G | C | 0.18 | 0.136 | 0.017 | 3.06E-16 | -0.010 | 0.004 | 0.025 | 66.61 |
| rs3851179 | T | C | 0.36 | -0.13 | 0.013 | 6.03E-25 | 0.002 | 0.004 | 0.567 | 107.11 |
| rs4844610 | A | C | 0.19 | 0.157 | 0.016 | 3.60E-24 | -0.010 | 0.004 | 0.025 | 102.47 |
| rs57537848 | T | G | NA | 0.207 | 0.015 | 2.11E-42 | 0.003 | 0.004 | 0.513 | 187.2 |
| rs593742 | G | A | 0.3 | -0.073 | 0.011 | 6.80E-09 | -0.005 | 0.004 | 0.247 | 43.73 |
| rs6024870 | A | G | 0.09 | -0.123 | 0.022 | 3.46E-08 | -0.002 | 0.004 | 0.681 | 30.37 |
| rs62039712 | A | G | 0.12 | 0.148 | 0.028 | 3.70E-08 | 0.004 | 0.004 | 0.321 | 27.13 |
| rs6733839 | T | C | 0.41 | 0.181 | 0.013 | 2.05E-44 | -0.005 | 0.004 | 0.279 | 194.71 |
| rs7185636 | C | T | 0.18 | -0.083 | 0.017 | 2.40E-08 | -0.004 | 0.004 | 0.327 | 25.1 |
| rs73223431 | T | C | 0.37 | 0.096 | 0.013 | 6.30E-14 | 0.003 | 0.004 | 0.467 | 56.48 |
| rs77301115 | A | G | NA | 0.949 | 0.043 | 2.25E-109 | -0.010 | 0.004 | 0.023 | 493.53 |
| rs7920721 | G | A | 0.39 | 0.077 | 0.012 | 1.80E-11 | -0.003 | 0.004 | 0.485 | 42.84 |
| rs7933202 | C | A | 0.39 | -0.113 | 0.013 | 1.91E-19 | -0.001 | 0.004 | 0.892 | 81.72 |
| rs9331896 | C | T | 0.39 | -0.132 | 0.013 | 4.63E-24 | -0.001 | 0.004 | 0.788 | 103.1 |
| rs9473117 | C | A | 0.28 | 0.087 | 0.014 | 1.16E-10 | -0.002 | 0.004 | 0.699 | 41.82 |

**SNPs used as instrumental variables in the MR analysis of late-onset Alzheimer's disease on intracellular volume fraction**

| **SNP** | **Effect allele** | **Other allele** | **Effect allele frequency** | **beta (exposure)** | **SE (exposure)** | ***P* (exposure)** | **beta (outcome)** | **SE (outcome)** | ***P* (outcome)** | ***F*-statistic** |
| --- | --- | --- | --- | --- | --- | --- | --- | --- | --- | --- |
| rs10808026 | A | C | 0.2 | -0.101 | 0.016 | 1.30E-10 | 0.011 | 0.007 | 0.126 | 41.06 |
| rs10933431 | G | C | 0.22 | -0.094 | 0.016 | 3.42E-09 | -0.011 | 0.007 | 0.136 | 34.88 |
| rs11218343 | C | T | 0.04 | -0.22 | 0.032 | 2.88E-12 | -0.006 | 0.016 | 0.696 | 48.91 |
| rs114812713 | C | G | 0.03 | 0.278 | 0.039 | 2.10E-13 | -0.027 | 0.02 | 0.172 | 51.4 |
| rs12539172 | T | C | 0.3 | -0.082 | 0.013 | 9.26E-10 | -0.008 | 0.006 | 0.194 | 37.72 |
| rs12721046 | A | G | NA | 1.006 | 0.02 | 1.00E-200 | -0.009 | 0.008 | 0.257 | 2606.18 |
| rs12881735 | C | T | 0.22 | -0.086 | 0.015 | 7.35E-09 | -0.001 | 0.007 | 0.836 | 33.39 |
| rs138190086 | A | G | 0.02 | 0.262 | 0.045 | 5.30E-09 | -0.005 | 0.022 | 0.816 | 33.88 |
| rs17125924 | G | A | 0.09 | 0.128 | 0.021 | 1.42E-09 | -0.028 | 0.011 | 0.007 | 36.74 |
| rs2830500 | A | C | 0.31 | -0.073 | 0.014 | 2.60E-08 | 0.004 | 0.007 | 0.599 | 28.29 |
| rs34665982 | T | C | NA | 0.097 | 0.017 | 5.80E-09 | -0.016 | 0.006 | 0.01 | 33.93 |
| rs35912475 | T | C | NA | -0.104 | 0.016 | 1.72E-10 | -0.003 | 0.006 | 0.638 | 40.63 |
| rs3740688 | G | T | 0.45 | -0.089 | 0.012 | 5.46E-13 | 0.003 | 0.006 | 0.585 | 52 |
| rs3752246 | G | C | 0.18 | 0.136 | 0.017 | 3.06E-16 | 0.013 | 0.008 | 0.102 | 66.61 |
| rs3851179 | T | C | 0.36 | -0.13 | 0.013 | 6.03E-25 | -0.009 | 0.006 | 0.148 | 107.11 |
| rs4844610 | A | C | 0.19 | 0.157 | 0.016 | 3.60E-24 | 0.018 | 0.008 | 0.02 | 102.47 |
| rs57537848 | T | G | NA | 0.207 | 0.015 | 2.11E-42 | 0.002 | 0.006 | 0.767 | 187.2 |
| rs593742 | G | A | 0.3 | -0.073 | 0.011 | 6.80E-09 | -0.01 | 0.007 | 0.111 | 43.73 |
| rs6024870 | A | G | 0.09 | -0.123 | 0.022 | 3.46E-08 | -0.003 | 0.011 | 0.794 | 30.37 |
| rs62039712 | A | G | 0.12 | 0.148 | 0.028 | 3.70E-08 | -0.004 | 0.01 | 0.667 | 27.13 |
| rs6733839 | T | C | 0.41 | 0.181 | 0.013 | 2.05E-44 | 0 | 0.006 | 0.968 | 194.71 |
| rs7185636 | C | T | 0.18 | -0.083 | 0.017 | 2.40E-08 | -0.025 | 0.008 | 0.002 | 25.1 |
| rs73223431 | T | C | 0.37 | 0.096 | 0.013 | 6.30E-14 | 0.001 | 0.006 | 0.896 | 56.48 |
| rs77301115 | A | G | NA | 0.949 | 0.043 | 2.25E-109 | -0.021 | 0.02 | 0.275 | 493.53 |
| rs7920721 | G | A | 0.39 | 0.077 | 0.012 | 1.80E-11 | -0.001 | 0.006 | 0.893 | 42.84 |
| rs7933202 | C | A | 0.39 | -0.113 | 0.013 | 1.91E-19 | -0.02 | 0.006 | 0.001 | 81.72 |
| rs9331896 | C | T | 0.39 | -0.132 | 0.013 | 4.63E-24 | -0.006 | 0.006 | 0.316 | 103.1 |
| rs9473117 | C | A | 0.28 | 0.087 | 0.014 | 1.16E-10 | 0 | 0.007 | 0.973 | 41.82 |

**SNPs used as instrumental variables in the MR analysis of late-onset Alzheimer's disease on orientation dispersion index**

| **SNP** | **Effect allele** | **Other allele** | **Effect allele frequency** | **beta (exposure)** | **SE (exposure)** | ***P* (exposure)** | **beta (outcome)** | **SE (outcome)** | ***P* (outcome)** | ***F*-statistic** |
| --- | --- | --- | --- | --- | --- | --- | --- | --- | --- | --- |
| rs10808026 | A | C | 0.2 | -0.101 | 0.016 | 1.30E-10 | -0.001 | 0.009 | 0.943 | 41.06 |
| rs10933431 | G | C | 0.22 | -0.094 | 0.016 | 3.42E-09 | 0.012 | 0.009 | 0.17 | 34.88 |
| rs11218343 | C | T | 0.04 | -0.22 | 0.032 | 2.88E-12 | 0.049 | 0.02 | 0.016 | 48.91 |
| rs114812713 | C | G | 0.03 | 0.278 | 0.039 | 2.10E-13 | 0.038 | 0.025 | 0.123 | 51.4 |
| rs12539172 | T | C | 0.3 | -0.082 | 0.013 | 9.26E-10 | -0.003 | 0.008 | 0.745 | 37.72 |
| rs12721046 | A | G | NA | 1.006 | 0.02 | 1.00E-200 | -0.039 | 0.01 | 1.72E-04 | 2606.18 |
| rs12881735 | C | T | 0.22 | -0.086 | 0.015 | 7.35E-09 | -0.003 | 0.009 | 0.726 | 33.39 |
| rs138190086 | A | G | 0.02 | 0.262 | 0.045 | 5.30E-09 | 0.001 | 0.028 | 0.959 | 33.88 |
| rs17125924 | G | A | 0.09 | 0.128 | 0.021 | 1.42E-09 | 0.001 | 0.013 | 0.911 | 36.74 |
| rs2830500 | A | C | 0.31 | -0.073 | 0.014 | 2.60E-08 | -0.012 | 0.008 | 0.136 | 28.29 |
| rs34665982 | T | C | NA | 0.097 | 0.017 | 5.80E-09 | 0.008 | 0.008 | 0.278 | 33.93 |
| rs35912475 | T | C | NA | -0.104 | 0.016 | 1.72E-10 | -0.004 | 0.008 | 0.628 | 40.63 |
| rs3740688 | G | T | 0.45 | -0.089 | 0.012 | 5.46E-13 | 0.004 | 0.008 | 0.589 | 52 |
| rs3752246 | G | C | 0.18 | 0.136 | 0.017 | 3.06E-16 | 0.006 | 0.01 | 0.512 | 66.61 |
| rs3851179 | T | C | 0.36 | -0.13 | 0.013 | 6.03E-25 | 0.001 | 0.008 | 0.93 | 107.11 |
| rs4844610 | A | C | 0.19 | 0.157 | 0.016 | 3.60E-24 | -0.002 | 0.01 | 0.841 | 102.47 |
| rs57537848 | T | G | NA | 0.207 | 0.015 | 2.11E-42 | -0.005 | 0.008 | 0.532 | 187.2 |
| rs593742 | G | A | 0.3 | -0.073 | 0.011 | 6.80E-09 | 0.025 | 0.008 | 0.003 | 43.73 |
| rs6024870 | A | G | 0.09 | -0.123 | 0.022 | 3.46E-08 | -0.007 | 0.013 | 0.581 | 30.37 |
| rs62039712 | A | G | 0.12 | 0.148 | 0.028 | 3.70E-08 | 0.021 | 0.013 | 0.102 | 27.13 |
| rs6733839 | T | C | 0.41 | 0.181 | 0.013 | 2.05E-44 | -0.013 | 0.008 | 0.094 | 194.71 |
| rs7185636 | C | T | 0.18 | -0.083 | 0.017 | 2.40E-08 | 0.001 | 0.01 | 0.957 | 25.1 |
| rs73223431 | T | C | 0.37 | 0.096 | 0.013 | 6.30E-14 | -0.002 | 0.008 | 0.796 | 56.48 |
| rs77301115 | A | G | NA | 0.949 | 0.043 | 2.25E-109 | 0.002 | 0.025 | 0.929 | 493.53 |
| rs7920721 | G | A | 0.39 | 0.077 | 0.012 | 1.80E-11 | -0.004 | 0.008 | 0.569 | 42.84 |
| rs7933202 | C | A | 0.39 | -0.113 | 0.013 | 1.91E-19 | 0.016 | 0.008 | 0.039 | 81.72 |
| rs9331896 | C | T | 0.39 | -0.132 | 0.013 | 4.63E-24 | -0.005 | 0.008 | 0.487 | 103.1 |
| rs9473117 | C | A | 0.28 | 0.087 | 0.014 | 1.16E-10 | 0.008 | 0.009 | 0.372 | 41.82 |

**SNPs used as instrumental variables in the MR analysis of late-onset Alzheimer's disease on fractional anisotropy (white matter tracts)**

| **SNP** | **Effect allele** | **Other allele** | **Effect allele frequency** | **beta (exposure)** | **SE (exposure)** | ***P* (exposure)** | **beta (outcome)** | **SE (outcome)** | ***P* (outcome)** | ***F*-statistic** |
| --- | --- | --- | --- | --- | --- | --- | --- | --- | --- | --- |
| rs10808026 | A | C | 0.2 | -0.101 | 0.016 | 1.30E-10 | 0 | 0.009 | 0.975 | 41.06 |
| rs10933431 | G | C | 0.22 | -0.094 | 0.016 | 3.42E-09 | -0.004 | 0.009 | 0.662 | 34.88 |
| rs11218343 | C | T | 0.04 | -0.22 | 0.032 | 2.88E-12 | 0.037 | 0.019 | 0.055 | 48.91 |
| rs114812713 | C | G | 0.03 | 0.278 | 0.039 | 2.10E-13 | -0.032 | 0.024 | 0.188 | 51.4 |
| rs12539172 | T | C | 0.3 | -0.082 | 0.013 | 9.26E-10 | -0.016 | 0.008 | 0.04 | 37.72 |
| rs12721046 | A | G | NA | 1.006 | 0.02 | 1.00E-200 | -0.04 | 0.01 | 8.10E-05 | 2606.18 |
| rs12881735 | C | T | 0.22 | -0.086 | 0.015 | 7.35E-09 | 0.005 | 0.009 | 0.563 | 33.39 |
| rs138190086 | A | G | 0.02 | 0.262 | 0.045 | 5.30E-09 | -0.053 | 0.027 | 0.049 | 33.88 |
| rs17125924 | G | A | 0.09 | 0.128 | 0.021 | 1.42E-09 | -0.021 | 0.013 | 0.105 | 36.74 |
| rs2830500 | A | C | 0.31 | -0.073 | 0.014 | 2.60E-08 | 0.017 | 0.008 | 0.04 | 28.29 |
| rs34665982 | T | C | NA | 0.097 | 0.017 | 5.80E-09 | -0.017 | 0.007 | 0.018 | 33.93 |
| rs35912475 | T | C | NA | -0.104 | 0.016 | 1.72E-10 | -0.007 | 0.007 | 0.31 | 40.63 |
| rs3740688 | G | T | 0.45 | -0.089 | 0.012 | 5.46E-13 | 0.002 | 0.007 | 0.826 | 52 |
| rs3752246 | G | C | 0.18 | 0.136 | 0.017 | 3.06E-16 | -0.012 | 0.009 | 0.189 | 66.61 |
| rs3851179 | T | C | 0.36 | -0.13 | 0.013 | 6.03E-25 | -0.001 | 0.008 | 0.845 | 107.11 |
| rs4844610 | A | C | 0.19 | 0.157 | 0.016 | 3.60E-24 | 0.024 | 0.01 | 0.013 | 102.47 |
| rs57537848 | T | G | NA | 0.207 | 0.015 | 2.11E-42 | -0.002 | 0.007 | 0.786 | 187.2 |
| rs593742 | G | A | 0.3 | -0.073 | 0.011 | 6.80E-09 | -0.009 | 0.008 | 0.258 | 43.73 |
| rs6024870 | A | G | 0.09 | -0.123 | 0.022 | 3.46E-08 | -0.019 | 0.013 | 0.15 | 30.37 |
| rs62039712 | A | G | 0.12 | 0.148 | 0.028 | 3.70E-08 | -0.014 | 0.012 | 0.238 | 27.13 |
| rs6733839 | T | C | 0.41 | 0.181 | 0.013 | 2.05E-44 | -0.018 | 0.008 | 0.016 | 194.71 |
| rs7185636 | C | T | 0.18 | -0.083 | 0.017 | 2.40E-08 | 0.007 | 0.01 | 0.471 | 25.1 |
| rs73223431 | T | C | 0.37 | 0.096 | 0.013 | 6.30E-14 | -0.002 | 0.008 | 0.808 | 56.48 |
| rs77301115 | A | G | NA | 0.949 | 0.043 | 2.25E-109 | 0.012 | 0.024 | 0.614 | 493.53 |
| rs7920721 | G | A | 0.39 | 0.077 | 0.012 | 1.80E-11 | 0.007 | 0.008 | 0.376 | 42.84 |
| rs7933202 | C | A | 0.39 | -0.113 | 0.013 | 1.91E-19 | -0.017 | 0.007 | 0.021 | 81.72 |
| rs9331896 | C | T | 0.39 | -0.132 | 0.013 | 4.63E-24 | -0.01 | 0.007 | 0.182 | 103.1 |
| rs9473117 | C | A | 0.28 | 0.087 | 0.014 | 1.16E-10 | -0.007 | 0.008 | 0.414 | 41.82 |

**SNPs used as instrumental variables in the MR analysis of late-onset Alzheimer's disease on mean diffusivity (white matter tracts)**

| **SNP** | **Effect allele** | **Other allele** | **Effect allele frequency** | **beta (exposure)** | **SE (exposure)** | ***P* (exposure)** | **beta (outcome)** | **SE (outcome)** | ***P* (outcome)** | ***F*-statistic** |
| --- | --- | --- | --- | --- | --- | --- | --- | --- | --- | --- |
| rs10808026 | A | C | 0.2 | -0.101 | 0.016 | 1.30E-10 | 0.003 | 0.009 | 0.733 | 41.06 |
| rs10933431 | G | C | 0.22 | -0.094 | 0.016 | 3.42E-09 | 0.006 | 0.008 | 0.488 | 34.88 |
| rs11218343 | C | T | 0.04 | -0.22 | 0.032 | 2.88E-12 | -0.031 | 0.019 | 0.096 | 48.91 |
| rs114812713 | C | G | 0.03 | 0.278 | 0.039 | 2.10E-13 | 0.021 | 0.023 | 0.356 | 51.4 |
| rs12539172 | T | C | 0.3 | -0.082 | 0.013 | 9.26E-10 | 0.015 | 0.008 | 0.041 | 37.72 |
| rs12721046 | A | G | NA | 1.006 | 0.02 | 1.00E-200 | 0.053 | 0.01 | 6.77E-08 | 2606.18 |
| rs12881735 | C | T | 0.22 | -0.086 | 0.015 | 7.35E-09 | -0.001 | 0.008 | 0.929 | 33.39 |
| rs138190086 | A | G | 0.02 | 0.262 | 0.045 | 5.30E-09 | 0.06 | 0.026 | 0.022 | 33.88 |
| rs17125924 | G | A | 0.09 | 0.128 | 0.021 | 1.42E-09 | 0.028 | 0.012 | 0.025 | 36.74 |
| rs2830500 | A | C | 0.31 | -0.073 | 0.014 | 2.60E-08 | -0.008 | 0.008 | 0.281 | 28.29 |
| rs34665982 | T | C | NA | 0.097 | 0.017 | 5.80E-09 | 0.015 | 0.007 | 0.032 | 33.93 |
| rs35912475 | T | C | NA | -0.104 | 0.016 | 1.72E-10 | -0.002 | 0.007 | 0.759 | 40.63 |
| rs3740688 | G | T | 0.45 | -0.089 | 0.012 | 5.46E-13 | -0.004 | 0.007 | 0.595 | 52 |
| rs3752246 | G | C | 0.18 | 0.136 | 0.017 | 3.06E-16 | 0 | 0.009 | 0.964 | 66.61 |
| rs3851179 | T | C | 0.36 | -0.13 | 0.013 | 6.03E-25 | 0.012 | 0.007 | 0.094 | 107.11 |
| rs4844610 | A | C | 0.19 | 0.157 | 0.016 | 3.60E-24 | -0.022 | 0.009 | 0.018 | 102.47 |
| rs57537848 | T | G | NA | 0.207 | 0.015 | 2.11E-42 | 0 | 0.007 | 0.945 | 187.2 |
| rs593742 | G | A | 0.3 | -0.073 | 0.011 | 6.80E-09 | 0.006 | 0.008 | 0.471 | 43.73 |
| rs6024870 | A | G | 0.09 | -0.123 | 0.022 | 3.46E-08 | 0.018 | 0.013 | 0.154 | 30.37 |
| rs62039712 | A | G | 0.12 | 0.148 | 0.028 | 3.70E-08 | 0.012 | 0.012 | 0.302 | 27.13 |
| rs6733839 | T | C | 0.41 | 0.181 | 0.013 | 2.05E-44 | 0.01 | 0.007 | 0.165 | 194.71 |
| rs7185636 | C | T | 0.18 | -0.083 | 0.017 | 2.40E-08 | 0.01 | 0.009 | 0.296 | 25.1 |
| rs73223431 | T | C | 0.37 | 0.096 | 0.013 | 6.30E-14 | 0.006 | 0.007 | 0.422 | 56.48 |
| rs77301115 | A | G | NA | 0.949 | 0.043 | 2.25E-109 | -0.001 | 0.023 | 0.951 | 493.53 |
| rs7920721 | G | A | 0.39 | 0.077 | 0.012 | 1.80E-11 | -0.001 | 0.007 | 0.858 | 42.84 |
| rs7933202 | C | A | 0.39 | -0.113 | 0.013 | 1.91E-19 | 0.018 | 0.007 | 0.012 | 81.72 |
| rs9331896 | C | T | 0.39 | -0.132 | 0.013 | 4.63E-24 | 0.006 | 0.007 | 0.371 | 103.1 |
| rs9473117 | C | A | 0.28 | 0.087 | 0.014 | 1.16E-10 | -0.002 | 0.008 | 0.806 | 41.82 |

**SNPs used as instrumental variables in the MR analysis of late-onset Alzheimer's disease on volume of left hippocampus**

| **SNP** | **Effect allele** | **Other allele** | **Effect allele frequency** | **beta (exposure)** | **SE (exposure)** | ***P* (exposure)** | **beta (outcome)** | **SE (outcome)** | ***P* (outcome)** | ***F*-statistic** |
| --- | --- | --- | --- | --- | --- | --- | --- | --- | --- | --- |
| rs10808026 | A | C | 0.2 | -0.101 | 0.016 | 1.30E-10 | 0.009 | 0.009 | 0.319 | 41.06 |
| rs10933431 | G | C | 0.22 | -0.094 | 0.016 | 3.42E-09 | 0.003 | 0.009 | 0.721 | 34.88 |
| rs11218343 | C | T | 0.04 | -0.22 | 0.032 | 2.88E-12 | 0.019 | 0.02 | 0.346 | 48.91 |
| rs114812713 | C | G | 0.03 | 0.278 | 0.039 | 2.10E-13 | -0.023 | 0.025 | 0.349 | 51.4 |
| rs12539172 | T | C | 0.3 | -0.082 | 0.013 | 9.26E-10 | 0.014 | 0.008 | 0.104 | 37.72 |
| rs12721046 | A | G | NA | 1.006 | 0.02 | 1.00E-200 | -0.046 | 0.011 | 1.79E-05 | 2606.18 |
| rs12881735 | C | T | 0.22 | -0.086 | 0.015 | 7.35E-09 | -0.006 | 0.009 | 0.523 | 33.39 |
| rs138190086 | A | G | 0.02 | 0.262 | 0.045 | 5.30E-09 | -0.045 | 0.029 | 0.115 | 33.88 |
| rs17125924 | G | A | 0.09 | 0.128 | 0.021 | 1.42E-09 | 0.019 | 0.013 | 0.147 | 36.74 |
| rs2830500 | A | C | 0.31 | -0.073 | 0.014 | 2.60E-08 | 0.004 | 0.009 | 0.624 | 28.29 |
| rs34665982 | T | C | NA | 0.097 | 0.017 | 5.80E-09 | 0 | 0.008 | 0.972 | 33.93 |
| rs35912475 | T | C | NA | -0.104 | 0.016 | 1.72E-10 | 0.002 | 0.008 | 0.79 | 40.63 |
| rs3740688 | G | T | 0.45 | -0.089 | 0.012 | 5.46E-13 | 0.001 | 0.008 | 0.877 | 52 |
| rs3752246 | G | C | 0.18 | 0.136 | 0.017 | 3.06E-16 | -0.016 | 0.01 | 0.109 | 66.61 |
| rs3851179 | T | C | 0.36 | -0.13 | 0.013 | 6.03E-25 | 0.002 | 0.008 | 0.816 | 107.11 |
| rs4844610 | A | C | 0.19 | 0.157 | 0.016 | 3.60E-24 | -0.009 | 0.01 | 0.393 | 102.47 |
| rs57537848 | T | G | NA | 0.207 | 0.015 | 2.11E-42 | -0.01 | 0.008 | 0.213 | 187.2 |
| rs593742 | G | A | 0.3 | -0.073 | 0.011 | 6.80E-09 | 0.013 | 0.008 | 0.131 | 43.73 |
| rs6024870 | A | G | 0.09 | -0.123 | 0.022 | 3.46E-08 | 0.004 | 0.014 | 0.796 | 30.37 |
| rs62039712 | A | G | 0.12 | 0.148 | 0.028 | 3.70E-08 | 0.003 | 0.012 | 0.813 | 27.13 |
| rs6733839 | T | C | 0.41 | 0.181 | 0.013 | 2.05E-44 | 0.001 | 0.008 | 0.931 | 194.71 |
| rs7185636 | C | T | 0.18 | -0.083 | 0.017 | 2.40E-08 | -0.006 | 0.01 | 0.572 | 25.1 |
| rs73223431 | T | C | 0.37 | 0.096 | 0.013 | 6.30E-14 | 0.013 | 0.008 | 0.104 | 56.48 |
| rs77301115 | A | G | NA | 0.949 | 0.043 | 2.25E-109 | 0.019 | 0.025 | 0.452 | 493.53 |
| rs7920721 | G | A | 0.39 | 0.077 | 0.012 | 1.80E-11 | -0.005 | 0.008 | 0.541 | 42.84 |
| rs7933202 | C | A | 0.39 | -0.113 | 0.013 | 1.91E-19 | 0.008 | 0.008 | 0.342 | 81.72 |
| rs9331896 | C | T | 0.39 | -0.132 | 0.013 | 4.63E-24 | 0.002 | 0.008 | 0.768 | 103.1 |
| rs9473117 | C | A | 0.28 | 0.087 | 0.014 | 1.16E-10 | 0.012 | 0.009 | 0.156 | 41.82 |

**SNPs used as instrumental variables in the MR analysis of late-onset Alzheimer's disease on volume of right hippocampus**

| **SNP** | **Effect allele** | **Other allele** | **Effect allele frequency** | **beta (exposure)** | **SE (exposure)** | ***P* (exposure)** | **beta (outcome)** | **SE (outcome)** | ***P* (outcome)** | ***F*-statistic** |
| --- | --- | --- | --- | --- | --- | --- | --- | --- | --- | --- |
| rs10808026 | A | C | 0.2 | -0.101 | 0.016 | 1.30E-10 | 0.015 | 0.009 | 0.107 | 41.06 |
| rs10933431 | G | C | 0.22 | -0.094 | 0.016 | 3.42E-09 | 0.01 | 0.009 | 0.303 | 34.88 |
| rs11218343 | C | T | 0.04 | -0.22 | 0.032 | 2.88E-12 | 0.004 | 0.02 | 0.838 | 48.91 |
| rs114812713 | C | G | 0.03 | 0.278 | 0.039 | 2.10E-13 | -0.017 | 0.025 | 0.505 | 51.4 |
| rs12539172 | T | C | 0.3 | -0.082 | 0.013 | 9.26E-10 | 0.01 | 0.008 | 0.213 | 37.72 |
| rs12721046 | A | G | NA | 1.006 | 0.02 | 1.00E-200 | -0.042 | 0.011 | 7.18E-05 | 2606.18 |
| rs12881735 | C | T | 0.22 | -0.086 | 0.015 | 7.35E-09 | -0.001 | 0.009 | 0.874 | 33.39 |
| rs138190086 | A | G | 0.02 | 0.262 | 0.045 | 5.30E-09 | -0.04 | 0.029 | 0.157 | 33.88 |
| rs17125924 | G | A | 0.09 | 0.128 | 0.021 | 1.42E-09 | 0.006 | 0.013 | 0.665 | 36.74 |
| rs2830500 | A | C | 0.31 | -0.073 | 0.014 | 2.60E-08 | 0 | 0.009 | 0.963 | 28.29 |
| rs34665982 | T | C | NA | 0.097 | 0.017 | 5.80E-09 | 0.01 | 0.008 | 0.192 | 33.93 |
| rs35912475 | T | C | NA | -0.104 | 0.016 | 1.72E-10 | 0.013 | 0.008 | 0.099 | 40.63 |
| rs3740688 | G | T | 0.45 | -0.089 | 0.012 | 5.46E-13 | -0.015 | 0.008 | 0.055 | 52 |
| rs3752246 | G | C | 0.18 | 0.136 | 0.017 | 3.06E-16 | -0.021 | 0.01 | 0.041 | 66.61 |
| rs3851179 | T | C | 0.36 | -0.13 | 0.013 | 6.03E-25 | 0.004 | 0.008 | 0.582 | 107.11 |
| rs4844610 | A | C | 0.19 | 0.157 | 0.016 | 3.60E-24 | -0.014 | 0.01 | 0.181 | 102.47 |
| rs57537848 | T | G | NA | 0.207 | 0.015 | 2.11E-42 | -0.013 | 0.008 | 0.094 | 187.2 |
| rs593742 | G | A | 0.3 | -0.073 | 0.011 | 6.80E-09 | 0.018 | 0.008 | 0.035 | 43.73 |
| rs6024870 | A | G | 0.09 | -0.123 | 0.022 | 3.46E-08 | -0.013 | 0.014 | 0.35 | 30.37 |
| rs62039712 | A | G | 0.12 | 0.148 | 0.028 | 3.70E-08 | 0.013 | 0.012 | 0.286 | 27.13 |
| rs6733839 | T | C | 0.41 | 0.181 | 0.013 | 2.05E-44 | -0.008 | 0.008 | 0.311 | 194.71 |
| rs7185636 | C | T | 0.18 | -0.083 | 0.017 | 2.40E-08 | -0.015 | 0.01 | 0.151 | 25.1 |
| rs73223431 | T | C | 0.37 | 0.096 | 0.013 | 6.30E-14 | 0.013 | 0.008 | 0.107 | 56.48 |
| rs77301115 | A | G | NA | 0.949 | 0.043 | 2.25E-109 | 0.016 | 0.025 | 0.52 | 493.53 |
| rs7920721 | G | A | 0.39 | 0.077 | 0.012 | 1.80E-11 | 0.005 | 0.008 | 0.571 | 42.84 |
| rs7933202 | C | A | 0.39 | -0.113 | 0.013 | 1.91E-19 | 0.001 | 0.008 | 0.908 | 81.72 |
| rs9331896 | C | T | 0.39 | -0.132 | 0.013 | 4.63E-24 | -0.001 | 0.008 | 0.874 | 103.1 |
| rs9473117 | C | A | 0.28 | 0.087 | 0.014 | 1.16E-10 | -0.004 | 0.009 | 0.673 | 41.82 |

**SNPs used as instrumental variables in the MR analysis of late-onset Alzheimer's disease on total volume of white matter hyperintensities**

| **SNP** | **Effect allele** | **Other allele** | **Effect allele frequency** | **beta (exposure)** | **SE (exposure)** | ***P* (exposure)** | **beta (outcome)** | **SE (outcome)** | ***P* (outcome)** | ***F*-statistic** |
| --- | --- | --- | --- | --- | --- | --- | --- | --- | --- | --- |
| rs10808026 | A | C | 0.2 | -0.101 | 0.016 | 1.30E-10 | 0.002 | 0.01 | 0.836 | 41.06 |
| rs10933431 | G | C | 0.22 | -0.094 | 0.016 | 3.42E-09 | 0.007 | 0.01 | 0.445 | 34.88 |
| rs11218343 | C | T | 0.04 | -0.22 | 0.032 | 2.88E-12 | -0.036 | 0.021 | 0.084 | 48.91 |
| rs114812713 | C | G | 0.03 | 0.278 | 0.039 | 2.10E-13 | 0.053 | 0.025 | 0.036 | 51.4 |
| rs12539172 | T | C | 0.3 | -0.082 | 0.013 | 9.26E-10 | 0.019 | 0.008 | 0.027 | 37.72 |
| rs12721046 | A | G | NA | 1.006 | 0.02 | 1.00E-200 | 0.051 | 0.011 | 2.18E-06 | 2606.18 |
| rs12881735 | C | T | 0.22 | -0.086 | 0.015 | 7.35E-09 | -0.005 | 0.009 | 0.595 | 33.39 |
| rs138190086 | A | G | 0.02 | 0.262 | 0.045 | 5.30E-09 | 0.03 | 0.029 | 0.293 | 33.88 |
| rs17125924 | G | A | 0.09 | 0.128 | 0.021 | 1.42E-09 | -0.011 | 0.014 | 0.403 | 36.74 |
| rs2830500 | A | C | 0.31 | -0.073 | 0.014 | 2.60E-08 | -0.016 | 0.009 | 0.06 | 28.29 |
| rs34665982 | T | C | NA | 0.097 | 0.017 | 5.80E-09 | 0.023 | 0.008 | 0.003 | 33.93 |
| rs35912475 | T | C | NA | -0.104 | 0.016 | 1.72E-10 | 0.019 | 0.008 | 0.018 | 40.63 |
| rs3740688 | G | T | 0.45 | -0.089 | 0.012 | 5.46E-13 | 0.001 | 0.008 | 0.943 | 52 |
| rs3752246 | G | C | 0.18 | 0.136 | 0.017 | 3.06E-16 | 0.003 | 0.01 | 0.754 | 66.61 |
| rs3851179 | T | C | 0.36 | -0.13 | 0.013 | 6.03E-25 | -0.011 | 0.008 | 0.17 | 107.11 |
| rs4844610 | A | C | 0.19 | 0.157 | 0.016 | 3.60E-24 | -0.01 | 0.01 | 0.327 | 102.47 |
| rs57537848 | T | G | NA | 0.207 | 0.015 | 2.11E-42 | 0.004 | 0.008 | 0.635 | 187.2 |
| rs593742 | G | A | 0.3 | -0.073 | 0.011 | 6.80E-09 | 0.005 | 0.009 | 0.568 | 43.73 |
| rs6024870 | A | G | 0.09 | -0.123 | 0.022 | 3.46E-08 | 0.024 | 0.014 | 0.092 | 30.37 |
| rs62039712 | A | G | 0.12 | 0.148 | 0.028 | 3.70E-08 | -0.014 | 0.013 | 0.281 | 27.13 |
| rs6733839 | T | C | 0.41 | 0.181 | 0.013 | 2.05E-44 | -0.007 | 0.008 | 0.386 | 194.71 |
| rs7185636 | C | T | 0.18 | -0.083 | 0.017 | 2.40E-08 | -0.007 | 0.011 | 0.499 | 25.1 |
| rs73223431 | T | C | 0.37 | 0.096 | 0.013 | 6.30E-14 | -0.001 | 0.008 | 0.893 | 56.48 |
| rs77301115 | A | G | NA | 0.949 | 0.043 | 2.25E-109 | -0.01 | 0.026 | 0.708 | 493.53 |
| rs7920721 | G | A | 0.39 | 0.077 | 0.012 | 1.80E-11 | -0.01 | 0.008 | 0.212 | 42.84 |
| rs7933202 | C | A | 0.39 | -0.113 | 0.013 | 1.91E-19 | 0.005 | 0.008 | 0.497 | 81.72 |
| rs9331896 | C | T | 0.39 | -0.132 | 0.013 | 4.63E-24 | 0.004 | 0.008 | 0.641 | 103.1 |
| rs9473117 | C | A | 0.28 | 0.087 | 0.014 | 1.16E-10 | -0.015 | 0.009 | 0.092 | 41.82 |
