## Supplementary Materials for "Educational attainment, structural brain reserve, and Alzheimer’s disease: a Mendelian randomization analysis"

### **Supplementary methods**

#### **Description of brain imaging-derived phenotypes**

Local gyrification index (LGI) is a measure of gyrification, the characteristic folding of the human cerebral cortex that emerges during development.^1^ This folding arises from the tangential expansion of the cortex, a process by which an increasingly expanding surface area is enclosed in a relatively small volume.^2,3^ LGI is a surface-based method for quantifying cortical gyrification, which takes into account the three-dimensional nature of the cortical surface, and is computed by estimating gyrification at thousands of points over the entire cortical area.^4^

Mean curvature (MC) is a measure of the extrinsic curvature of the cortical surface (i.e., the folding of the cortex that arises as a result of cortical surface expansion during development). This is an extrinsic property of the cortical surface, in that it is a function of how the surface is embedded in space, rather than reflecting the shape of the surface itself.^2^

Intrinsic curvature (IC) is a mathematically fundamental property of the cortical surface that is intrinsic to the geometry of the surface (i.e., unlike extrinsic curvature, it is not a function of how the cortex is folded in space and cannot be removed without deforming or tearing the surface).^1,5^ IC arises from differential growth between cortical regions,^1^ and has been suggested to be a more sensitive marker of neurodevelopmental abnormalities compared to larger scale measures of gyrification.^2^

Fractional anisotropy (FA) and mean diffusivity (MD) are two commonly used metrics derived from diffusion tensor imaging (DTI),^6^ the standard clinical diffusion MRI technique which enables the assessment of grey and white matter integrity. FA is a measure of the overall integrity and directionality of water diffusion,^7^ which is highly sensitive to general connectivity changes.^8^ MD is a measure reflecting the rotationally invariant magnitude of water diffusion within brain tissue.^7^ Although FA and MD are the most widely used surrogate measures of microstructural tissue change, they are non-specific markers for individual tissue microstructure features.^9^

The intracellular volume fraction (ICVF) and orientation dispersion index (ODI) phenotypes are both derived from neurite orientation dispersion and density imaging (NODDI),^10^ a diffusion MRI technique which was developed specifically for in vivo estimation of the microstructural properties of dendrites and axons – collectively referred to as neurites – in both white and grey matter. NODDI can be used to compartmentalize brain tissue into three microstructural environments with distinct diffusion properties: intra-cellular, extra-cellular, and cerebrospinal fluid. The intra-cellular compartment models the space occupied by the neurites, and can be used to quantify neurite morphology both in terms of their density (ICVF) and their spatial orientation (ODI).^10,11^ ICVF and ODI have been suggested to provide more specific markers of brain tissue microstructure compared to standard DTI-derived parameters such as FA and MD.^10^

#### **Description of summary statistics data sources**

**GWAS of educational attainment by Lee *et al.*^12^**

The analysis combined summary statistics from 71 separate cohorts, all of which were restricted to individuals of European ancestry. Within each cohort, the major educational qualifications obtained by each individual were mapped onto the International Standard Classification of Education (ISCED) categories. The educational attainment phenotype was then constructed by imputing the equivalent years of education for each ISCED category. The sample-size-weighted mean (standard deviation) of the educational attainment phenotype was 16.8 (4.2) years of schooling across all cohorts. All association analyses were performed at the cohort level and adjusted for age, sex, their interaction, and genetic principal components. The sample-size weighted meta-analysis generated association estimates for approximately 10 million autosomal SNPs that passed the quality-control thresholds in all cohorts, of which 1,271 approximately independent SNPs (*r^2^* < 0.1) were genome-wide significant (*P* < 5 × 10^-8^).

A standard quality-control pipeline was applied to all cohort-level results, and genotypes were imputed using either the 1,000 Genomes Project Phase 3 European reference panel or a larger reference panel released by the Haplotype Reference Consortium. Subject-level filters included the exclusion of individuals of non-European ancestry, those with poor genotyping rates, and genetic outliers. In addition, individuals in whom the educational attainment phenotype was measured prior to age 30 were removed from the analysis. Genotype filters included the removal of INDELS and variants not located on autosomes, SNPs with known strand issues in imputation programs, SNPs with poor imputation accuracy, and those with minor allele counts < 25. Additionally, SNPs were excluded if they had invalid or duplicated chromosomal coordinates, if they had missing values for some of the variables (e.g., missing *P-*values), or if the alleles did not match those in the reference panel.

**GWAS of Late-onset Alzheimer’s disease by Kunkle *et al.*^13^**

The Stage 1 discovery sample (21,982 cases and 41,944 cognitively normal controls) was composed of 46 case-control datasets from the following four consortia: Alzheimer Disease Genetics Consortium (ADGC), Cohorts for Heart and Aging Research in Genomic Epidemiology Consortium (CHARGE), The European Alzheimer’s Disease Initiative (EADI), and Genetic and Environmental Risk in AD/Defining Genetic, Polygenic and Environmental Risk for Alzheimer’s Disease Consortium (GERAD/PERADES).

All association analyses were based on an additive genotype model and were adjusted for age (defined as age at onset of AD for cases and age at last examination for controls), sex, and principal components. The Stage 1 discovery meta-analysis was followed by Stage 2 replication in 8,362 cases and 10,483 controls, using a custom genotyping chip (iSelect) that was described in the 2013 IGAP GWAS meta-analysis (11,632 variants).^14^ Stage 3A (4,930 cases and 6,736 controls) involved replication for variants that showed suggestive association with late-onset AD (*p* < 5 × 10^-7^) in the meta-analysis of Stages 1 and 2, and variants that were genome-wide significant (*p* < 5 × 10^-8^) in the previous IGAP GWAS but not in the meta-analysis of Stages 1 and 2 (11 variants in total). Finally, variants with minor allele frequency (MAF) < 0.05 and *P* < 1 × 10^-5^ or MAF ≥ 0.05 and *P* < 5 × 10^-6^ in Stage 1 that were in regions not well-captured by the iSelect chip (33 variants in total) were selected for follow-up genotyping in Stage 3B (combined samples from Stage 2 and Stage 3A: 13,292 cases and 17,219 controls). The three stages had non-overlapping samples. The final sample consisted of 35,274 clinical and autopsy-documented cases of late-onset AD and 59,163 controls (94,437 individuals in total).

Standard quality control procedures were applied to each dataset prior to imputation, including the exclusion of individuals of non-European ancestry according to principal components analysis and those with a high degree of genetic relatedness. Individuals with low call rate and variants with low call rate were also removed from the analysis. The discovery datasets were then phased and imputed using the 1,000 Genomes Project reference panel (phase 1 integrated release 3, March 2012), using all reference population haplotypes for the imputation. Both common (MAF ≥ 0.01) and rare (MAF < 0.01) variants were sampled. The minimum imputation quality score required for inclusion was 0.4 for common variants and 0.7 for rare variants. Additionally, variants had to be present in at least 30% of cases and 30% of controls across all datasets to be retained in the analysis. A total of 9,456,058 common variants and 2,024,574 rare variants were selected for analysis after quality control.

**Multi-modal brain imaging in the UK Biobank**

Genetic association estimates with imaging-derived phenotypes were based on the largest-to-date release of combined genetic and multi-modal brain imaging data from the UK Biobank. The UK Biobank is a large prospective cohort study of approximately 500,000 individuals (aged 40–69 years at baseline recruitment), gathering extensive lifestyle, cognitive, physical, and biological measures (including genotyping), as well as data on ongoing health outcomes.^15^ An imaging extension was added to the existing UK Biobank study in 2016, aiming to acquire consistent multi-modal brain imaging data in 100,000 participants from the existing cohort by 2022.^16^ The early-2020 release of brain imaging samples from the UK Biobank contained data from approximately 40,000 participants across nearly 4,000 imaging-derived phenotypes. The imaging phenotypes span modalities that capture the anatomical and neuropathological structure of the brain (structural MRI), local microstructure of brain tissue (diffusion MRI), and brain activity (functional MRI).

**In-house GWAS of cortical macro- and micro-structure**

The macro-structure metrics were based on T1-weighted structural images obtained from the UK Biobank in minimally processed form (application number 20904), which were further pre-processed using FreeSurfer software (version 6.0.1).^17^ The processing pipeline documentation and code is available on <https://github.com/ucam-department-of-psychiatry/UKB>. The micro-structure metrics were based on structural diffusion weighted imaging data, which were obtained in processed form from the UK Biobank. Estimation of the NODDI parameters (intracellular volume fraction and orientation dispersion index) from the minimally processed diffusion images was based on the Accelerated Microstructure Imaging via Convex Optimization (AMICO) processing pipeline.^18^ Across all imaging-derived phenotypes, the maximum number of participants and variants retained after quality control was 31,977 and 10,004,255, respectively. All genetic association estimates were adjusted for age, age^2^, sex, age × sex, age^2^ × sex, the first 40 genetic principal components, and the following imaging covariates: imaging center, mean framewise displacement, maximum framewise displacement, and the Euler Index.^19^

The analysis was restricted to individuals of self-reported White European ethnicity. Subject-level exclusion criteria included excessive genetic heterozygosity, non-matching genetic and reported sex, genotyping rate of less than 95%, and a deviation of more than $\pm$5 standard deviations from the means of the first two genetic principal components. Variant quality control involved the removal of SNPs with minor allele frequency (MAF) < 0.1%, SNPs missing in more than 5% of individuals, SNPs deviating from Hardy-Weinberg equilibrium (*P* < 1 × 10^-6^) and those with an imputation *R^2^* > 0.4 (for imputed SNPs). Prior to conducting the GWAS, all phenotypes were scaled to have a mean of 0 and a standard deviation of 1. Phenotypic outliers (i.e., individuals with scores above or below 5 standard deviations from the mean) were excluded from the analysis. Outliers were further removed by visually inspecting the histograms of all phenotypes and removing values above or below 5 median absolute deviations for phenotypes that were substantially skewed, primary the mean diffusivity phenotype.

**GWAS of white matter micro-structure by Zhao *et al.*^8^**

Th GWAS of human brain white matter micro-structure was based on diffusion MRI data from 34,024 individuals of British ancestry in the UK Biobank. White matter microstructure was quantified using fractional anisotropy, mean diffusivity, and other metrics derived from diffusion MRI data using DTI models.^6^ Image processing was harmonized using the ENIGMA-DTI pipeline.^20,21^ Each of the DTI-derived metrics was estimated along 21 predefined cerebral white matter tracts (i.e., tract-averaged parameters) and across all 21 white matter tracts (i.e., global parameters). The discovery GWAS identified 151 genomic regions significantly associated with white matter microstructure (*P* < 2.3 × 10^-10^, corrected for the number of phenotypes studied), which together explained 41% of the variation in white matter microstructure.

**GWAS of hippocampal volumes and white matter hyperintensities volume by Smith *et al.*^22^**

Imaging-derived phenotypes were generated using the automated image processing pipeline developed by Alfaro-Almagro et al.^23^ on behalf of UK Biobank (<https://www.fmrib.ox.ac.uk/ukbiobank/fbp/>), which removes artifacts and aligns images across different individuals and imaging modalities. The GWAS was based on data from 33,224 participants (52.4% genetic females), with a mean (standard deviation) age of 64.28 (7.49) years. A total of 16,445,196 autosomal variants were retained in the analysis after applying quality control filters. All genetic association tests were adjusted for age, sex, 40 population genetic principal components, head size, imaging center, scanner table position, and scan date-related slow drifts.

Phenotypic outliers (i.e., greater than $\pm$ 6 median absolute deviation from the median) and individuals with missing data for 50 or more imaging-derived phenotypes were discarded from the analysis. Each imaging-derived phenotypes was quantile-normalized, resulting in a Gaussian distribution with a mean of 0 and a standard deviation of 1. Subject-level quality control filters included removing participants without recent UK ancestry (as determined by self-reported ancestry and genetic principal components threshold), as well as the exclusion of individuals based on genetic relatedness. Quality control filters for the inclusion of genetic variants included MAF ≥ 0.001, information score (INFO)^24^ ≥ 0.3, and a Hardy-Weinberg equilibrium *P* < 1 × 10^-7^.

#### **Phenotypic correlations quality control pipeline**

Individual-level data for each imaging-derived phenotype was either obtained in processed form from the UK Biobank (for left and right hippocampal volume and total volume of white matter hyperintensities), or downloaded in minimally processed form from the UK Biobank and underwent the in-house imaging processing pipeline (documented on <https://github.com/ucam-department-of-psychiatry/UKB>). Prior to calculating the pair-wise phenotypic correlations, all phenotypes were scaled to have a mean of 0 and a standard deviation of 1. We removed any individuals who scored above or below 5 standard deviations from the mean for all phenotypes, as these outliers would skew the phenotypic scores and affect the pair-wise correlations between the imaging phenotypes. Additionally, we visually inspected histograms of all phenotypes and further removed any outliers above or below 5 median absolute deviations for phenotypes with substantial skew, primarily for mean diffusivity. We then calculated pairwise Pearson’s correlations between the 15 imaging-derived phenotypes (Supplementary Fig. 2).

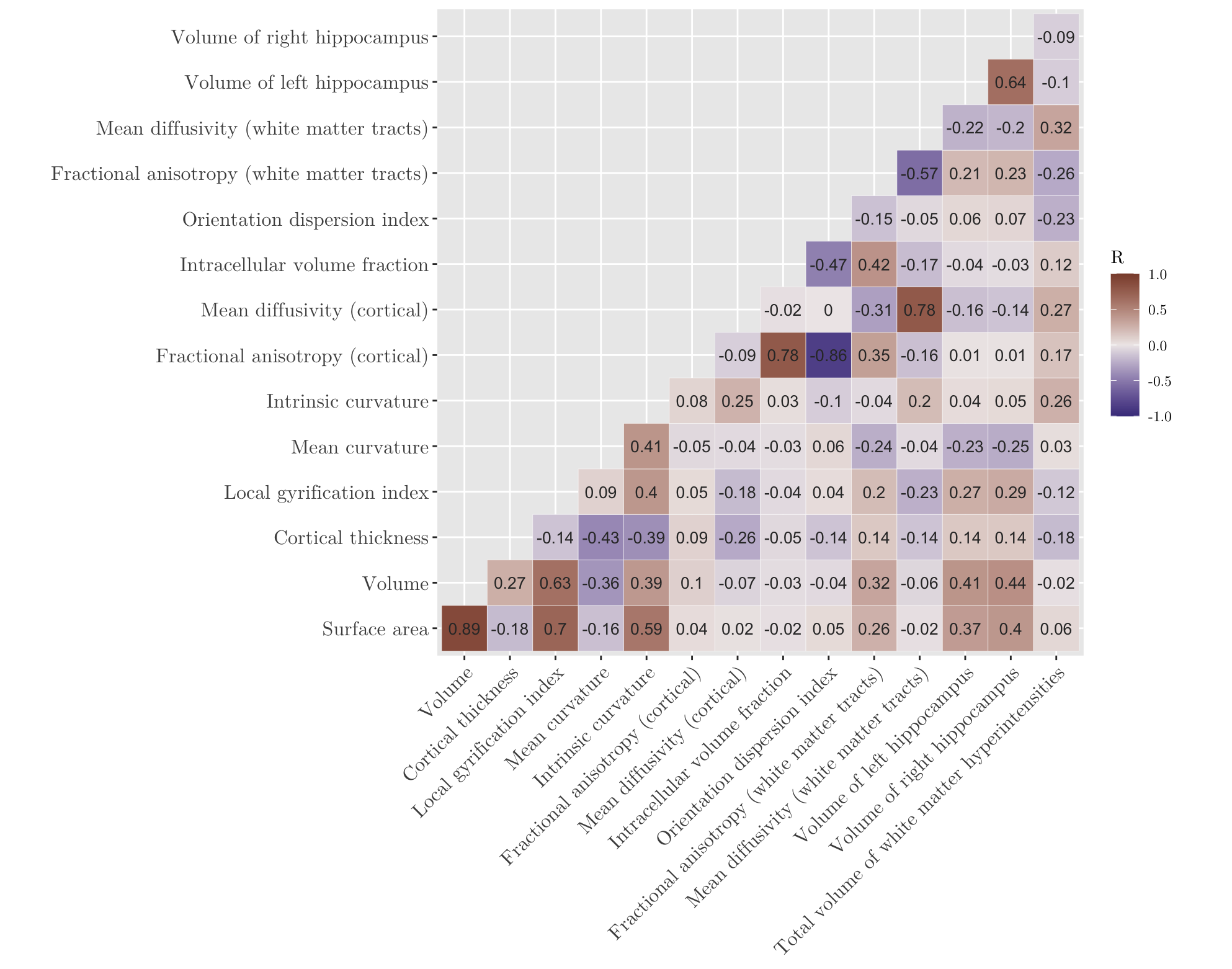

**Supplementary Fig. 1 Matrix of Pearson’s correlation coefficients between each pair of imaging-derived phenotypes.**

#### **Robust Mendelian randomization methods**

##### **Weighted Median MR**

The median method is a consensus method, in that it derives its causal effect estimate from a summarised measure of the distribution of the variant-specific ratio estimates.^143^ The simple version of the median method takes the median of the variant-specific ratio estimates as its estimate of the causal effect, with all variants receiving equal weights.^143^ The current analysis uses the weighted version of the median method,^150^ in which genetic variants receive standardised weights corresponding to the inverse of the variance of their ratio estimate, such that variants with more precise causal estimates receive higher weights. The weighted median estimator is then calculated as the median of the empirical distribution of the individual ratio estimates. Provided that at least 50% of the weights contributing to the analysis come from valid instruments, the median of the variant-specific causal estimates will tend towards the true causal parameter, and the weighted median method will provide consistent causal effect estimates (i.e., the “majority valid” assumption).^143,150^ Furthermore, since the median of a distribution is not influenced by the magnitude of extreme values, weighted median MR is naturally robust to outliers. However, the estimate from weighted median MR may be less efficient (i.e., larger standard errors and lower power) than methods that derive their causal estimate from all variants.^143^

##### **Contamination mixture method**

The contamination mixture method^151^ can provide a robust and efficient estimate of the overall causal effect under the plurality valid assumption, which states that out of all the different values taken by the variant-specific ratio estimates, the value that is taken by the largest group of variants is the true causal effect of the exposure on the outcome.^143^

The method is performed by constructing a likelihood function from the variant-specific ratio estimates. If a genetic variant is a valid instrument, then its ratio estimate is assumed to follow a normal distribution centred around the true causal effect value. For genetic variants that are invalid instruments, the method assumes that their causal estimates are normally distributed about zero with a large variance. The likelihood function is then maximised by considering different values of the causal effect parameter (*θ* ) in turn and finding the configuration of valid and invalid instruments that maximises the likelihood function for a given value of *θ*. This is a profile likelihood approach to maximisation as it is based on constructing a one-dimensional function of the causal effect parameter.^151^ The value of *θ* that maximises the profile likelihood is taken as the point estimate of the causal effect, and 95% confidence intervals are constructed using the likelihood function (i.e., by finding the range of values of *θ* for which twice the difference between the log-likelihood at that value and the log-likelihood at the maximum likelihood estimator falls below the 95^th^ percentile of a chi-squared distribution with one degree of freedom).^151^

The contamination mixture method is robust to outliers^143^ and does not make any strict modelling assumptions, and so is less likely to be affected by violations of such assumptions. However, it can be sensitive to specification of the variance parameter for the distribution of invalid instruments, as well as the addition or removal of genetic variants from the analysis^143^.

##### **MR-Egger**

The MR-Egger method is an adaptation of Egger regression, which can be used to test and correct for bias arising from directional pleiotropy (i.e., the average pleiotropic effects of genetic variants not equalling zero).^152^ MR-Egger is implemented similarly to the IVW method, except for the inclusion of an intercept term in the regression model. The slope coefficient from Egger regression provides the MR-Egger estimate of the causal effect, and the intercept term represents the average pleiotropic effect across all variants.^152^ The test of the null hypothesis that the MR-Egger estimate does not differ from zero is referred to as the MR-Egger causal test.^153^ Provided that the pleiotropic effect of each genetic variant on the outcome is independent of the association of the variant with the exposure (i.e., the ‘InSIDE’ assumption), the MR Egger method can provide consistent causal effect estimates even if all of the genetic variants included in the analysis are invalid instruments.^152^ In addition, given that the intercept term will differ from zero either when the average pleiotropic effect is not equal to zero, or when the ‘InSIDE’ assumption is violated, testing the intercept from the MR-Egger analysis can be used to assess whether the instrumental variable assumptions are violated. This test is referred to as the MR-Egger intercept test, with a non-zero intercept indicating that the IVW estimate is biased.^153^

The causal estimate from the MR-Egger method will always have a larger standard error than that from the IVW analysis, and the difference in precision can sometimes be substantial.^76^ In the IVW method, the precision of the causal estimate is dependent on the proportion of variance in the exposure that is accounted for by the genetic variants.^154^ However, the precision of the MR-Egger estimate additionally depends on the heterogeneity between genetic associations with the exposure.^155^ Therefore, if several genetic variants have similar associations with the exposure, the MR-Egger intercept and causal estimate will have low precision and wide confidence intervals.^153^

##### **MR-PRESSO**

The Mendelian randomization pleiotropy residual sum and outlier (MR-PRESSO) method^156^ is an outlier-robust method which identifies and removes horizontal pleiotropic genetic variants with outlying ratio estimates from the analysis. Provided that the assumptions of the IVW method are met for the genetic variants not identified as outliers, this method can provide consistent causal effect estimates.^76^ The MR-PRESSO method has the following three components: (1) the MR-PRESSO global test, which detects the presence of horizontal pleiotropy; (2) the MR-PRESSO outlier test, which corrects for horizontal pleiotropy by removing outliers; and (3) the MR-PRESSO distortion test, which tests for significant differences between the causal estimates before and after the removal of horizontal pleiotropic outlying variants.^156^ The MR-PRESSO method is efficient when the analysis includes a small number of outlying variants with heterogeneous estimates, as these will be removed from the analysis and the overall causal estimate will not be affected by them. However, when there are several pleiotropic genetic variants, this method can have a very high false positive rate.^143^

#### **Assumptions of univariable and multivariable Mendelian randomization**

For a genetic variant to be a valid instrumental variable in a univariable MR model^26^:

I) The variant must be associated with the exposure of interest.
II) The variant must not be associated with any measured or unmeasured confounders of the exposure-outcome association.
III) The variant must only affect the outcome indirectly via the hypothesized causal pathway through the exposure (i.e., no direct pathway from the genetic variant to the outcome).

For a genetic variant to be a valid instrumental variable in a multivariable MR model^27^:

I) The variant must be associated with at least one of the exposures.
II) The variant must not be associated with any confounders of any of the exposure-outcome associations.
III) The variant must only affect the outcome indirectly via one or more of the exposures included in the analysis model.

A directed acyclic graph comparing the assumptions of univariable and multivariable Mendelian randomization is presented in Supplementary Fig. 1.

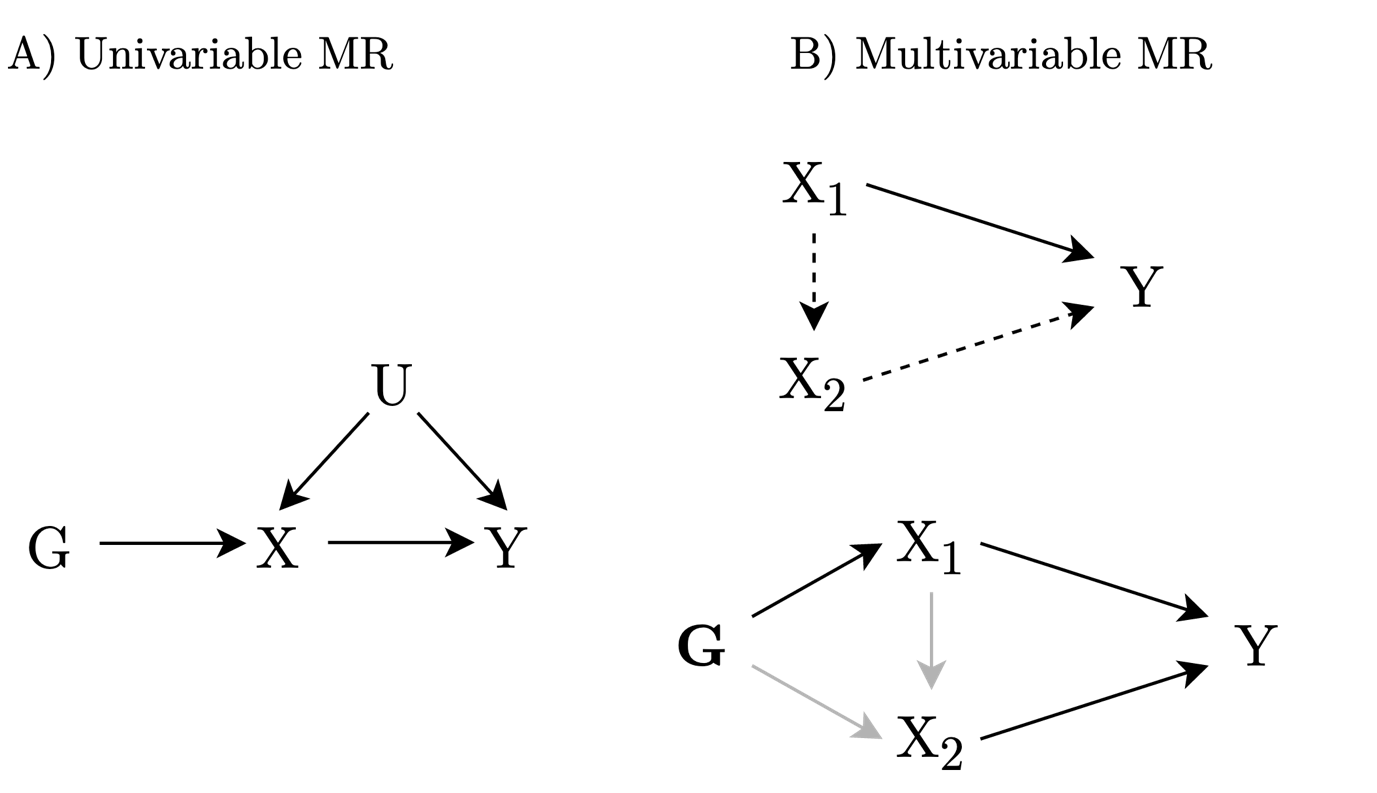

**Supplementary Fig. 2 Assumptions of univariable and multivariable Mendelian randomization.** A) directed acyclic graph illustrating univariable Mendelian randomization for genetic variant G, exposure X, and outcome Y. U represents the unknown confounders of the association between X and Y. The presence of arrows between G and X indicates that the first instrumental variable assumption is satisfied. In addition, G is independent of the confounder U (assumption II), and there are no direct pathways from G to Y apart from that passing through X (assumption III). B) directed acyclic graphs illustrating direct and indirect causal effects in a multivariable Mendelian randomization model with two exposures (X_1_ and X_2_) and one outcome (Y). As shown in the top panel, the total effect of X_1_ on Y has two components: a direct effect of X_1_ on Y (the solid arrow) and an indirect effect of X_1_ on Y via X_2_ (the dashed arrow). Assuming X_1_ is the primary exposure of interest, the estimate from a univariable MR model excluding X_2_ represents the total causal effect of X_1_ on Y. The estimates from a multivariable MR model including both X_1_ and X_2_ represent the direct effect of X_1_ on Y (i.e., not via X_2_) and the direct effect of X_2_ on Y (i.e., not via X_1_). A difference between the univariable and multivariable estimates of the effect of X_1_ on Y indicates that there is a causal effect of X_1_ on X_2_ (grey arrows), and that X_2_ is a mediator on the causal pathway from G to X_1_ to Y. Adapted from Burgess & Thompson.^26^

**Supplementary Table 1 Power calculations for two-sample Mendelian randomization analyses.**

| Exposure | Outcome | Proportion of variance in the exposure explained by the instrument (*R^2^*) | Minimum causal effect detectable with 80% power |
| --- | --- | --- | --- |
| EA | LOAD | 0.020 | OR ≤ 0.85 \| OR ≥ 1.18 |
| EA | SA | 0.022 | \|β\| ≥ 0.14 |
| EA | Vol | 0.022 | \|β\| ≥ 0.14 |
| EA | CT | 0.022 | \|β\| ≥ 0.14 |
| EA | LGI | 0.022 | \|β\| ≥ 0.14 |
| EA | MC | 0.022 | \|β\| ≥ 0.14 |
| EA | IC | 0.022 | \|β\| ≥ 0.14 |
| EA | FA (cortex) | 0.022 | \|β\| ≥ 0.14 |
| EA | MD (cortex) | 0.022 | \|β\| ≥ 0.14 |
| EA | ICVF | 0.022 | \|β\| ≥ 0.14 |
| EA | ODI | 0.022 | \|β\| ≥ 0.14 |
| EA | FA (WM tracts) | 0.022 | \|β\| ≥ 0.13 |
| EA | MD (WM tracts) | 0.022 | \|β\| ≥ 0.13 |
| EA | HC Vol (left) | 0.022 | \|β\| ≥ 0.13 |
| EA | HC Vol (right) | 0.022 | \|β\| ≥ 0.13 |
| EA | WMH Vol | 0.022 | \|β\| ≥ 0.13 |
| SA | LOAD | 0.026 | OR ≤ 0.83 \| OR ≥ 1.21 |
| Vol | LOAD | 0.020 | OR ≤ 0.81 \| OR ≥ 1.24 |
| CT | LOAD | 0.019 | OR ≤ 0.81 \| OR ≥ 1.24 |
| LGI | LOAD | 0.017 | OR ≤ 0.79 \| OR ≥ 1.26 |
| MC | LOAD | 0.026 | OR ≤ 0.83 \| OR ≥ 1.20 |
| IC | LOAD | 0.008 | OR ≤ 0.72 \| OR ≥ 1.38 |
| FA (cortex) | LOAD | – |  |
| MD (cortex) | LOAD | 0.010 | OR ≤ 0.74 \| OR ≥ 1.35 |
| ICVF | LOAD | 0.006 | OR ≤ 0.68 \| OR ≥ 1.48 |
| ODI | LOAD | 0.005 | OR ≤ 0.67 \| OR ≥ 1.50 |
| FA (WM tracts) | LOAD | 0.027 | OR ≤ 0.83 \| OR ≥ 1.20 |
| MD (WM tracts) | LOAD | 0.028 | OR ≤ 0.84 \| OR ≥ 1.20 |
| HC Vol (left) | LOAD | 0.009 | OR ≤ 0.74 \| OR ≥ 1.36 |
| HC Vol (right) | LOAD | 0.010 | OR ≤ 0.74 \| OR ≥ 1.35 |
| WMH Vol | LOAD | 0.037 | OR ≤ 0.86 \| OR ≥ 1.17 |

Power calculations were performed using the online calculator by Burgess^28^ (<http://sb452.shinyapps.io/power>). R^2^ was approximated using the following formula: $\text{R}^{\text{2}}\text{ }\text{= 2}\text{ }\text{β}_{\text{x}}^{\text{2}}\text{ }\text{MAF}\text{ }\text{(1-MAF)}$,^29^ where β_x_ is the genetic association estimate with the exposure, and MAF is the minor allele frequency. The minimum detectable causal effects are presented as odds ratios (per SD increase in genetically-predicted levels of the exposure) for the binary outcome (late-onset Alzheimer disease) and as beta coefficients (SD change in the outcome per SD increase in genetically-predicted levels of the exposure) for the continuous outcomes (imaging-derived phenotypes). EA = educational attainment; LOAD = late-onset Alzheimer’s disease; OR = odds ratio; SA = surface area; Vol = volume; CT = cortical thickness; LGI = local gyrification index; MC = mean curvature; IC = intrinsic curvature; FA = fractional anisotropy; MD = mean diffusivity; ICVF = intracellular volume fraction; ODI = orientation dispersion index; WM = white matter; HC = hippocampus; WMH = white matter hyperintensities.

**Supplementary Table 2 Cochran’s Q tests for heterogeneity from MR-IVW analyses.**

| Exposure | Outcome | *Q* | df | *P*-value |
| --- | --- | --- | --- | --- |
| EA | LOAD | 333.35 | 331 | 0.453 |
| EA | SA | 1026.59 | 375 | <.001 |
| EA | Vol | 793.55 | 375 | <.001 |
| EA | LGI | 705.43 | 375 | <.001 |
| EA | IC | 872.14 | 375 | <.001 |
| EA | ICVF | 470.19 | 375 | <.001 |
| EA | WMH Vol | 574.26 | 376 | <.001 |
| SA | EA | 127.13 | 24 | <.001 |
| Vol | EA | 148.82 | 24 | <.001 |
| IC | EA | 165.74 | 19 | <.001 |
| LOAD | ODI | 38.60 | 27 | 0.069 |
| LOAD | MD (WM tracts) | 60.06 | 27 | <.001 |
| LOAD | HC Vol (left) | 26.63 | 27 | 0.484 |
| LOAD | HC Vol (right) | 38.35 | 27 | 0.072 |

MR = Mendelian randomization; IVW = inverse-variance weighted; df = degrees of freedom; EA = educational attainment; LOAD = late-onset Alzheimer’s disease; SA = surface area; Vol = volume; LGI = local gyrification index; IC = intrinsic curvature; WMH = white matter hyperintensities; ODI = orientation dispersion index; MD = mean diffusivity; WM = white matter; HC = hippocampus.

**Supplementary Table 3	MR-Egger intercept tests for horizontal pleiotropy.**

| Exposure | Outcome | Egger intercept (95% CI) | *P*-value |
| --- | --- | --- | --- |
| EA | LOAD | 0.00 (0.00, 0.01) | 0.182 |
| EA | SA | 0.00 (-0.01, 0.00) | 0.223 |
| EA | Vol | 0.00 (-0.01, 0.00) | 0.255 |
| EA | LGI | 0.00 (0.00, 0.00) | 0.912 |
| EA | IC | 0.00 (0.00, 0.00) | 0.536 |
| EA | ICVF | 0.00 (0.00, 0.00) | 0.745 |
| EA | WMH Vol | 0.00 (0.00, 0.00) | 0.682 |
| SA | EA | 0.00 (0.00, 0.01) | 0.479 |
| Vol | EA | 0.00 (-0.01, 0.01) | 0.847 |
| IC | EA | 0.01 (0.00, 0.02) | 0.157 |
| LOAD | ODI | 0.00 (0.00, 0.01) | 0.348 |
| LOAD | MD (WM tracts) | -0.01 (-0.01, -0.00) | 0.020 |
| LOAD | HC Vol (left) | 0.00 (0.00, 0.01) | 0.442 |
| LOAD | HC Vol (right) | 0.00 (0.00, 0.01) | 0.467 |

MR = Mendelian randomization; CI = confidence interval; EA = educational attainment; LOAD = late-onset Alzheimer’s disease; SA = surface area; Vol = volume; LGI = local gyrification index; IC = intrinsic curvature; WMH = white matter hyperintensities; ODI = orientation dispersion index; MD = mean diffusivity; WM = white matter; HC = hippocampus.

**Supplementary Table 4	Results of Steiger filtering sensitivity analyses.**

| Exposure | Outcome | N _SNPs_ ^a^ | | MR analysis (all SNPs) | | MR analysis (valid SNPs) | |
| --- | --- | --- | --- | --- | --- | --- | --- |
|  |  | **Total** | **Invalid** | **β _IVW_ (95% CI)** | ***P*-value** | **β _IVW_ (95% CI)** | ***P*-value** |
| EA | LOAD | 332 | 24 | -0.36 (-0.51, -0.22) | 9.98 × 10^-7^ | -0.33 (-0.48, -0.18) | 1.40× 10^-5^ |
| EA | SA | 376 | 89 | 0.30 (0.20, 0.40) | 4.5 × 10^-9^ | 0.12 (0.05, 0.19) | 6.86 × 10^-4^ |
| EA | Vol | 376 | 87 | 0.29 (0.20, 0.37) | 3.31 × 10^-11^ | 0.14 (0.08, 0.21) | 1.92 × 10^-5^ |
| EA | LGI | 376 | 104 | 0.21 (0.11, 0.31) | 2.05 × 10^-5^ | 0.08 (0.00, 0.16) | 0.059 |
| EA | IC | 376 | 54 | 0.18 (0.11, 0.25) | 2.88 × 10^-7^ | 0.11 (0.06, 0.16) | 3.68 × 10^-5^ |
| EA | ICVF | 376 | 53 | -0.09 (-0.15, -0.03) | 0.006 | -0.07 (-0.13, -0.01) | 0.030 |
| EA | WMH Vol | 377 | 111 | -0.14 (-0.23, -0.05) | 0.003 | -0.05 (-0.13, 0.04) | 0.258 |
| SA | EA | 25 | 0 | 0.13 (0.10, 0.16) | 1.70 × 10^-14^ | 0.13 (0.10, 0.16) | 1.70 × 10^-14^ |
| Vol | EA | 25 | 0 | 0.15 (0.11, 0.19) | 2.00 × 10^-12^ | 0.15 (0.11, 0.19) | 2.00 × 10^-12^ |
| IC | EA | 20 | 0 | 0.12 (0.04, 0.19) | 0.001 | 0.12 (0.04, 0.19) | 0.001 |
| LOAD | ODI | 27 | 0 | -0.03 (-0.05, -0.01) | 0.002 | -0.03 (-0.05, -0.01) | 0.002 |
| LOAD | MD (WM tracts) | 27 | 0 | 0.03 (0.01, 0.05) | 0.008 | 0.03 (0.01, 0.05) | 0.008 |
| LOAD | HC Vol (left) | 27 | 0 | -0.03 (-0.05, -0.02) | 3.59 × 10^-5^ | -0.03 (-0.05, -0.02) | 3.59 × 10^-5^ |
| LOAD | HC Vol (right) | 27 | 0 | -0.03 (-0.05, -0.01) | 5.60 × 10^-4^ | -0.03 (-0.05, -0.01) | 5.60 × 10^-4^ |

^a^ Number of SNPs remaining after clumping for independence and data harmonization.

SNP = single nucleotide polymorphism; MR = Mendelian randomization; CI = confidence interval; EA = educational attainment; LOAD = late-onset Alzheimer’s disease; SA = surface area; Vol = volume; LGI = local gyrification index; IC = intrinsic curvature; WMH = white matter hyperintensities; ODI = orientation dispersion index; MD = mean diffusivity; WM = white matter; HC = hippocampus.

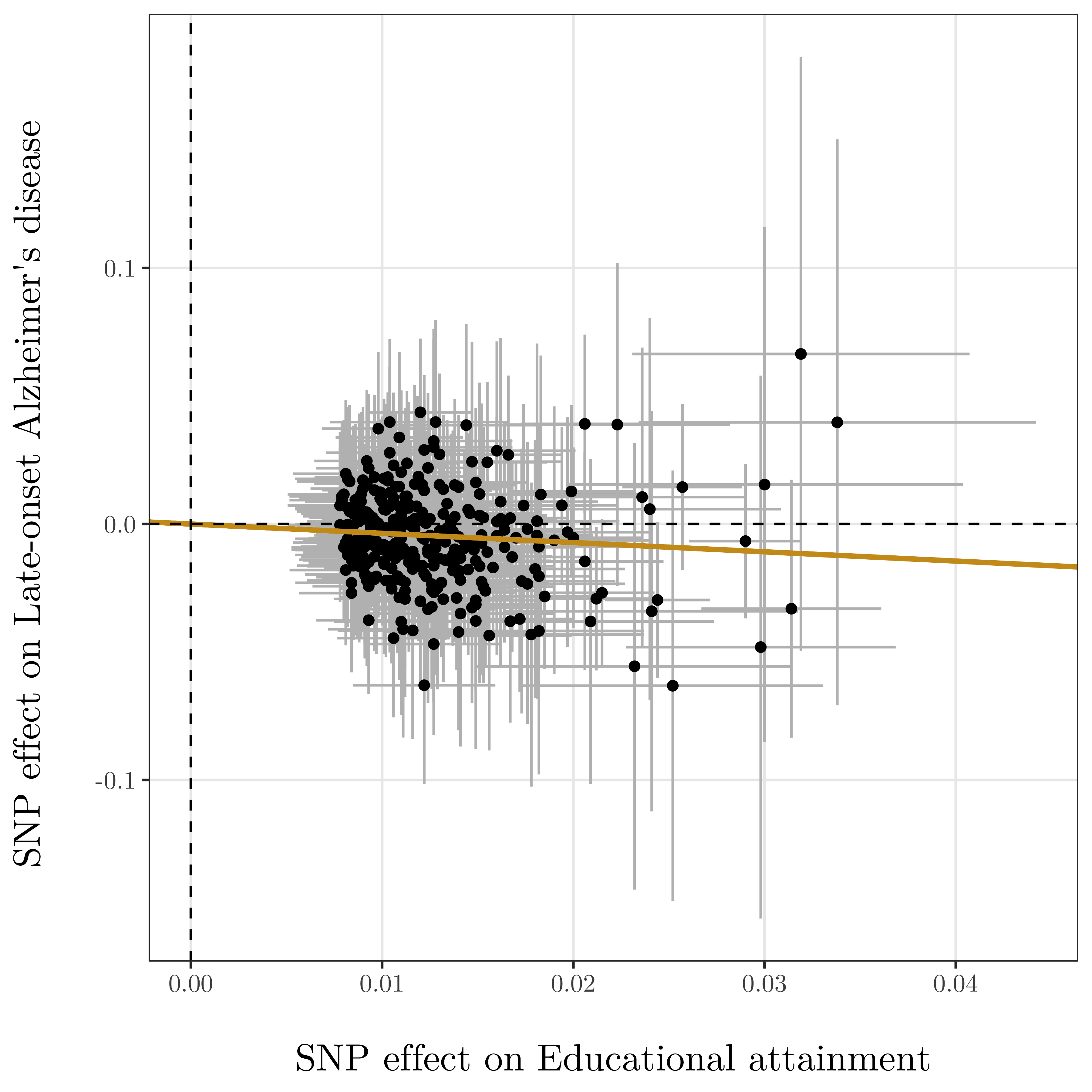

**Supplementary Fig. 3** Genetic associations with educational attainment (horizontal axis, standard deviation units) and with late-onset Alzheimer’s disease (vertical axis, log odds ratios) for 332 genetic variants associated with educational attainment at a genome-wide level of significance. Horizontal and vertical lines represent 95% confidence intervals for the genetic associations. The regression line through the origin represents the inverse-variance weighted Mendelian randomization estimate for the effect of educational attainment on late-onset Alzheimer’s disease risk.

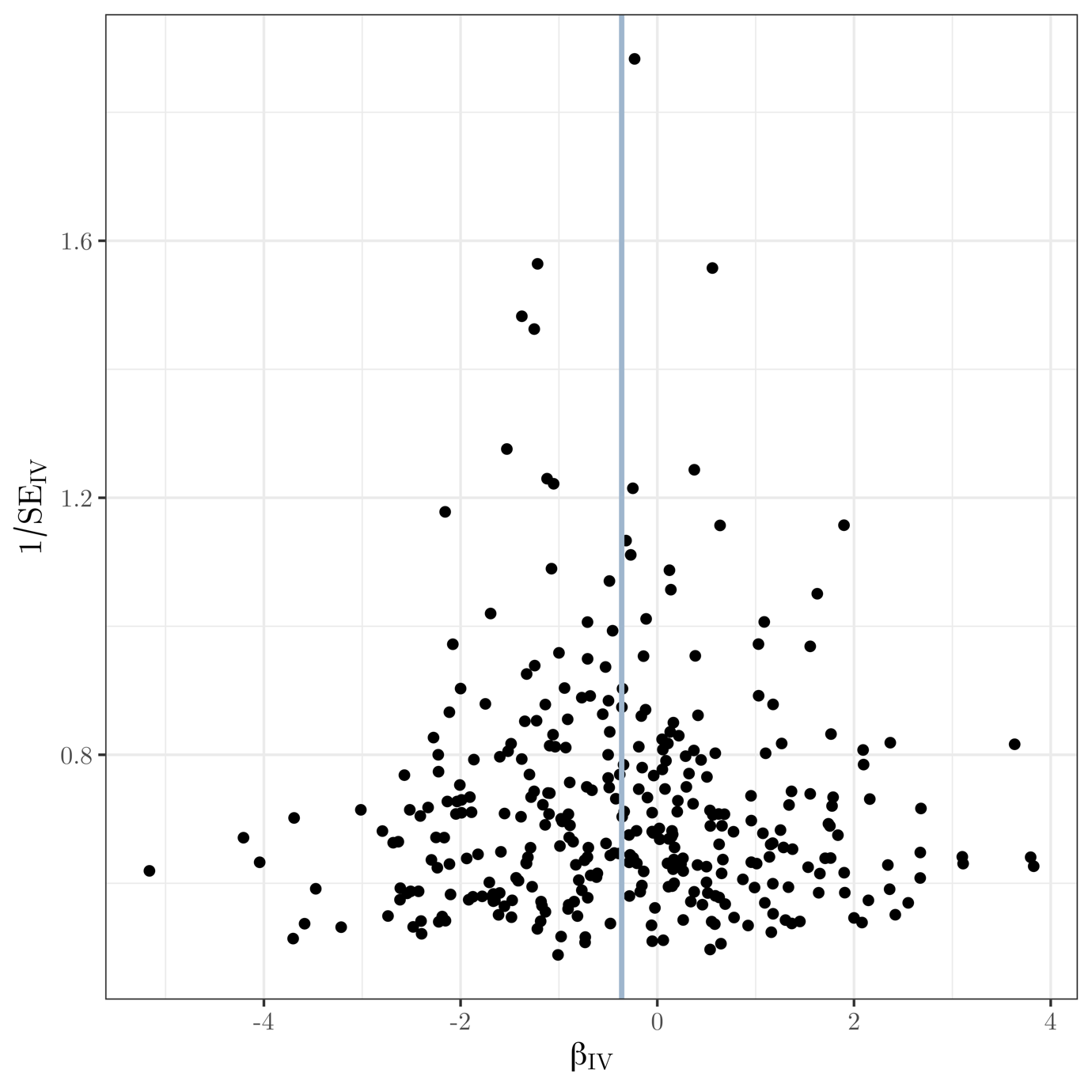

**Supplementary Fig. 4** Funnel plot of instrument precision against instrumental variable estimates for each genetic variant separately for Mendelian randomization analysis of educational attainment on late-onset AD risk. Solid vertical line is the (fixed-effect) inverse-variance weighted estimate.

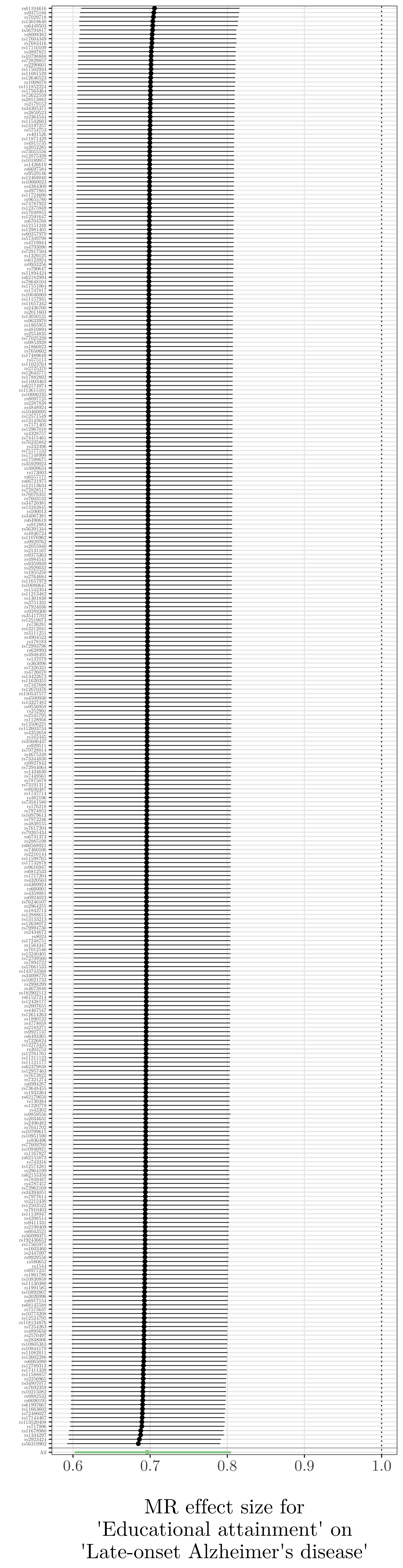

**Supplementary Fig. 5** Leave-one-out plot for MR analysis of educational attainment on late-onset AD risk.

| 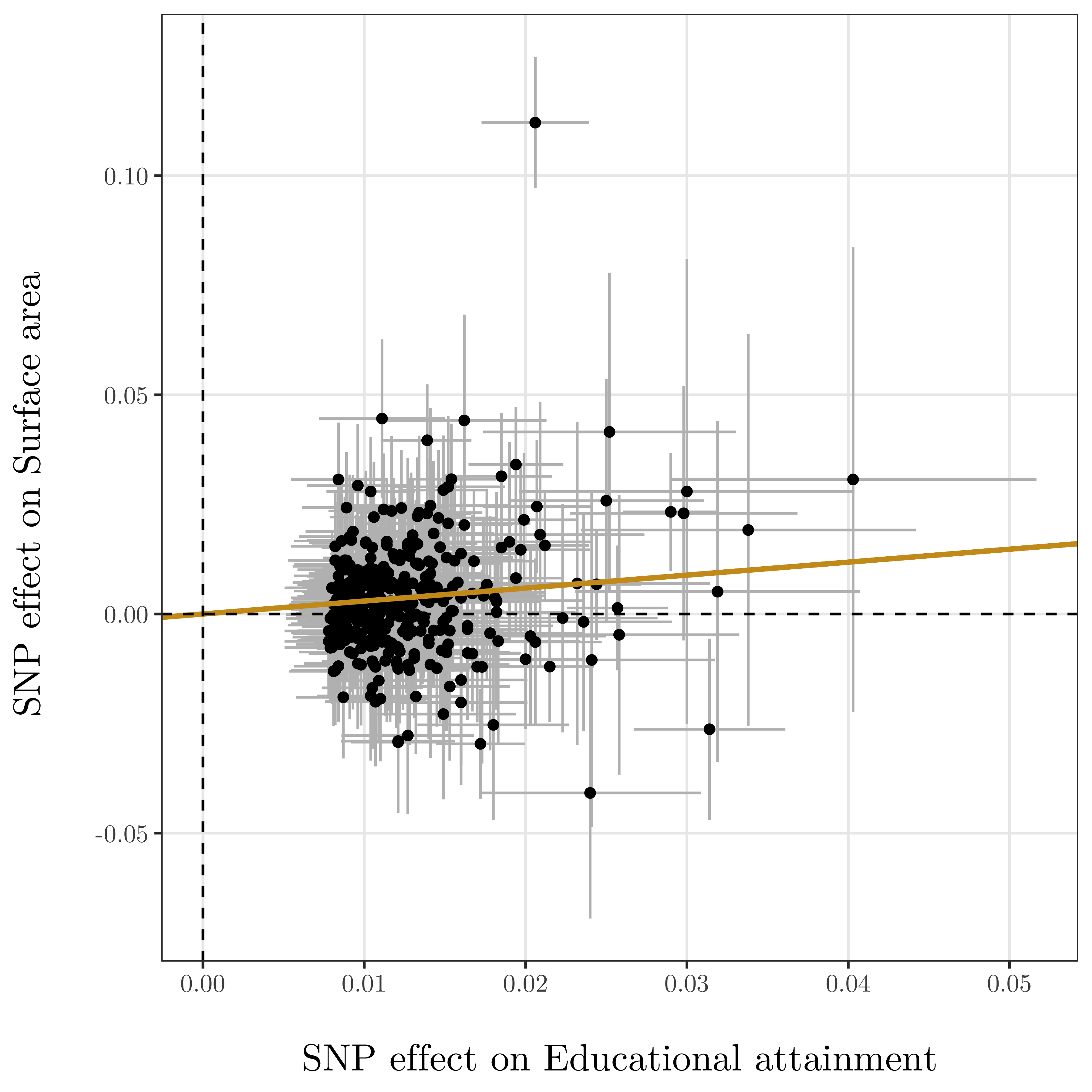 | 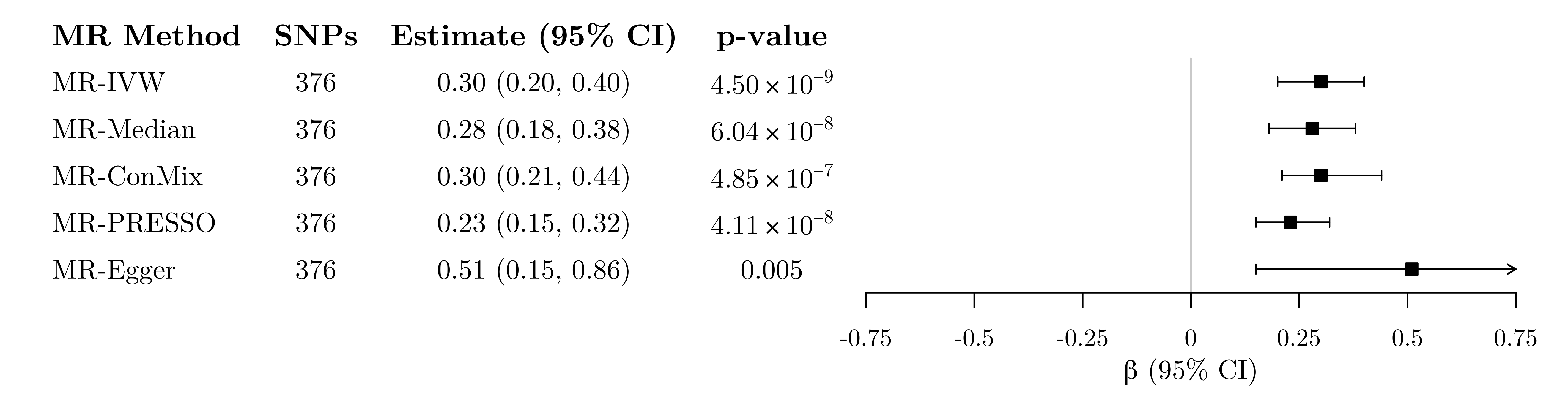 |
| --- | --- |
| 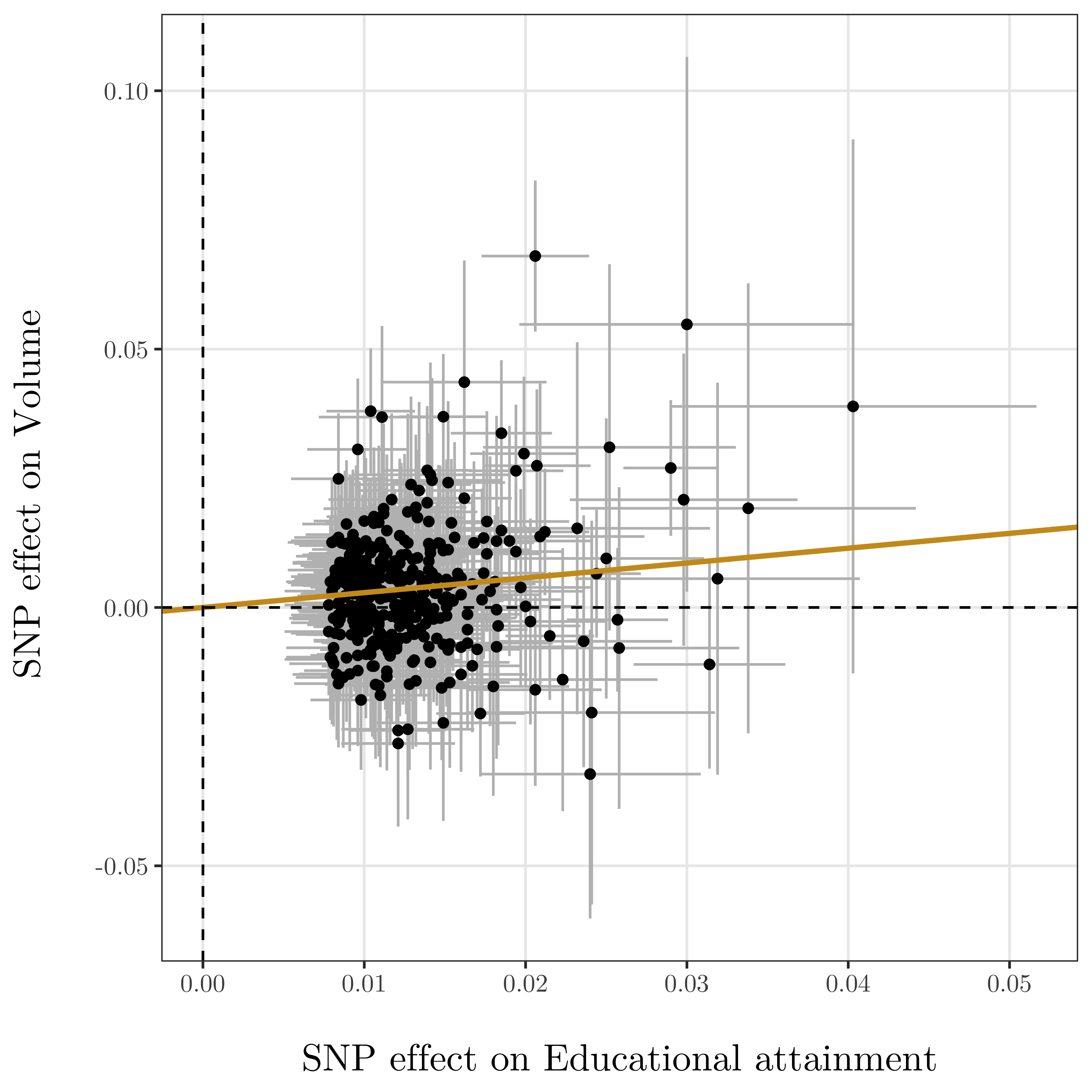 | 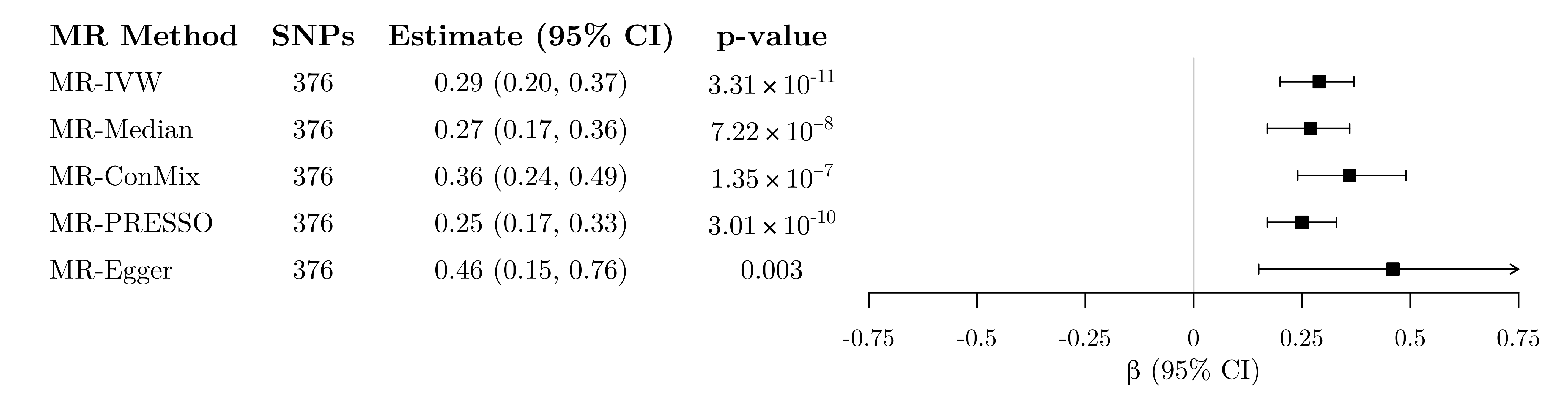 |

**Supplementary Fig. 6** Left: Genetic associations with educational attainment (horizontal axis, standard deviation units) and with imaging-derived phenotypes (vertical axis, standard deviation units) for 376 genetic variants associated with educational attainment at a genome-wide level of significance. Horizontal and vertical lines represent 95% confidence intervals for the genetic associations. The regression line through the origin represents the inverse-variance weighted Mendelian randomization estimate for the effect of educational attainment on each imaging-derived phenotype. Right: Mendelian randomization estimates of the association between genetically-proxied educational attainment and imaging-derived brain structure phenotypes. Estimates represent standard deviation change in the imaging phenotype per 1 standard deviation increase in genetically-predicted years of schooling (approximately 4.2 years).

| 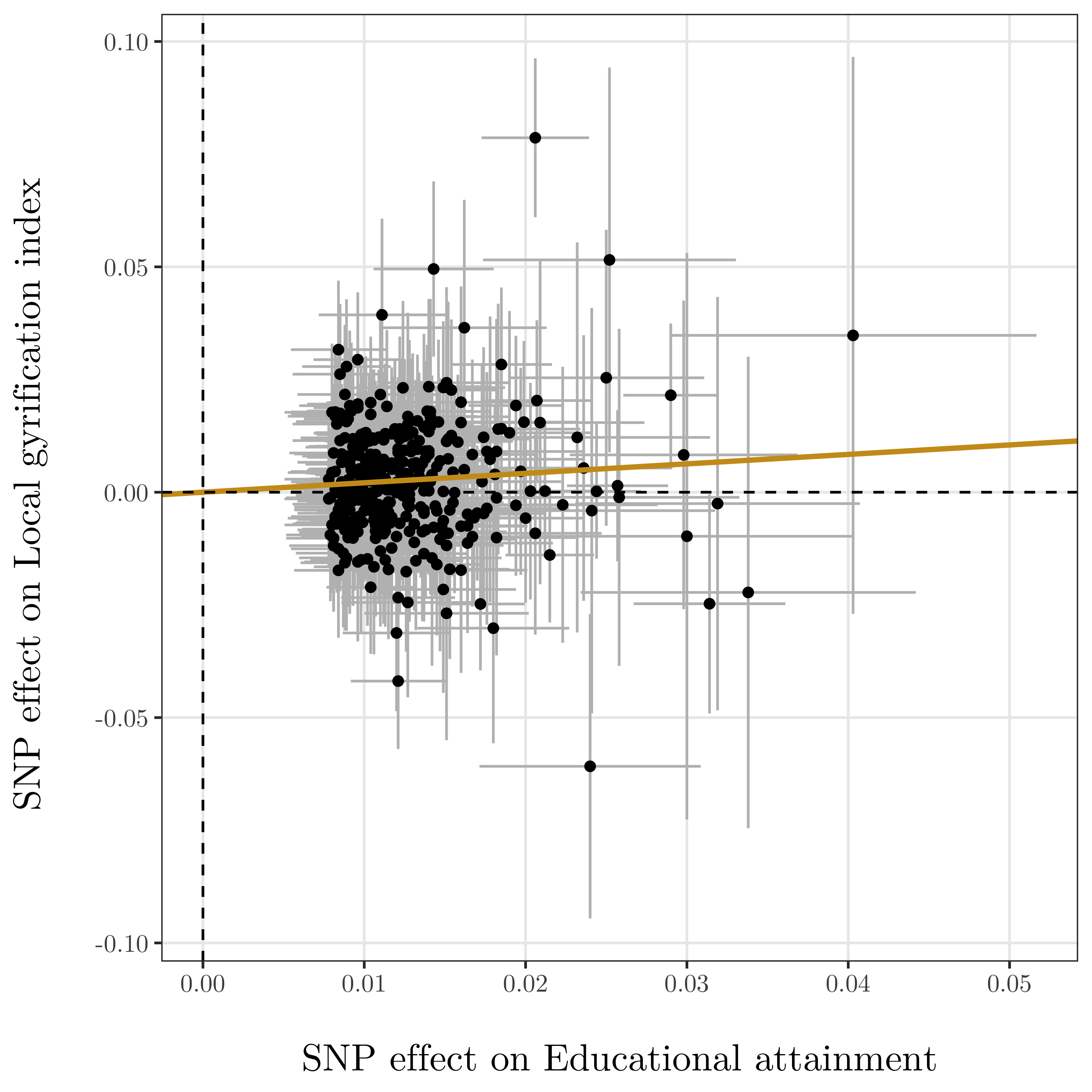 | 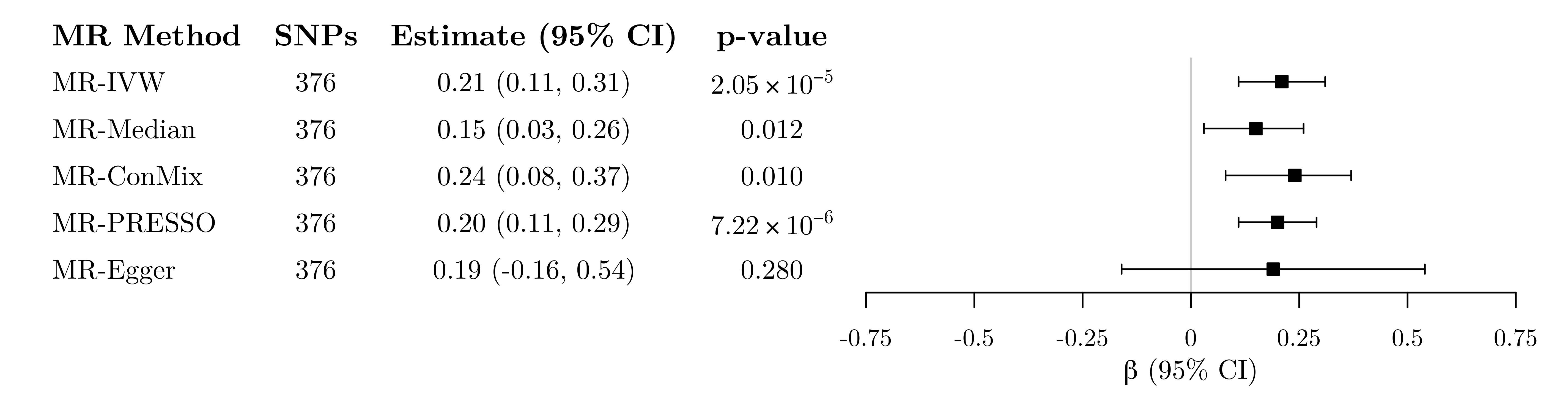 |
| --- | --- |
| 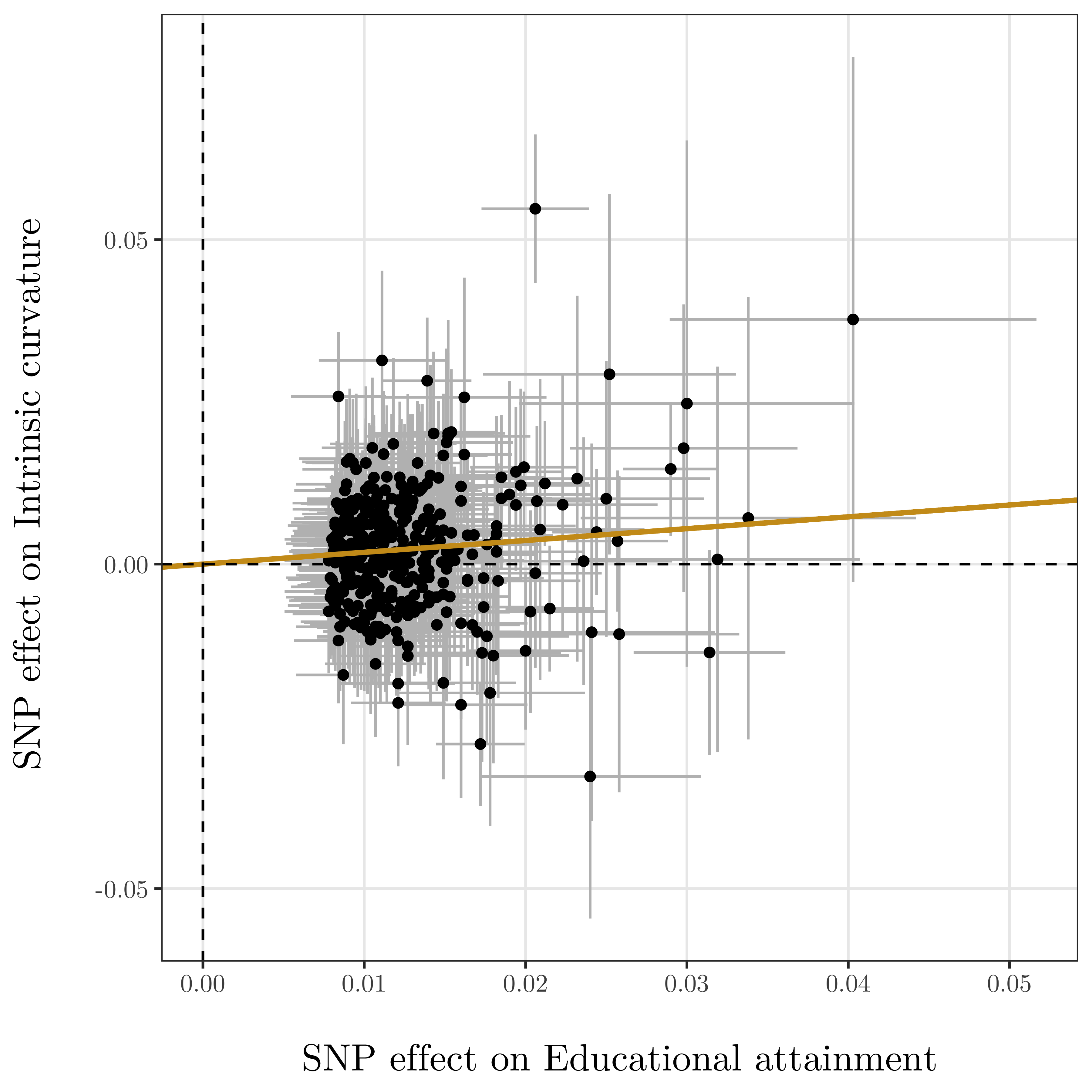 | 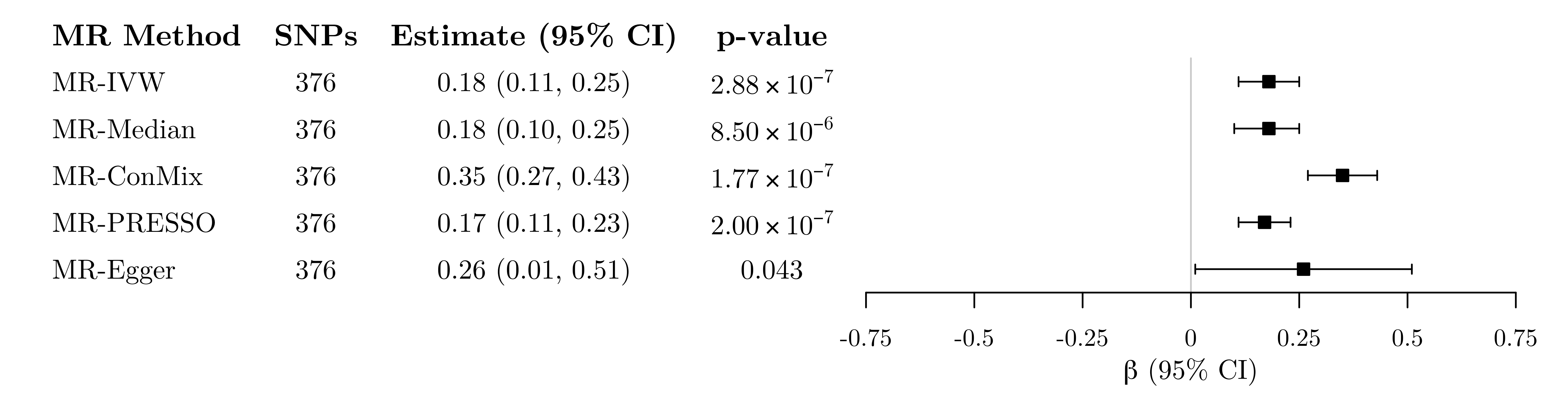 |

**Supplementary Fig. 6 (continued).** Left: Genetic associations with educational attainment (horizontal axis, standard deviation units) and with imaging-derived phenotypes (vertical axis, standard deviation units) for 376 genetic variants associated with educational attainment at a genome-wide level of significance. Horizontal and vertical lines represent 95% confidence intervals for the genetic associations. The regression line through the origin represents the inverse-variance weighted Mendelian randomization estimate for the effect of educational attainment on each imaging-derived phenotype. Right: Mendelian randomization estimates of the association between genetically-proxied educational attainment and imaging-derived brain structure phenotypes. Estimates represent standard deviation change in the imaging phenotype per 1 standard deviation increase in genetically-predicted years of schooling (approximately 4.2 years).

| 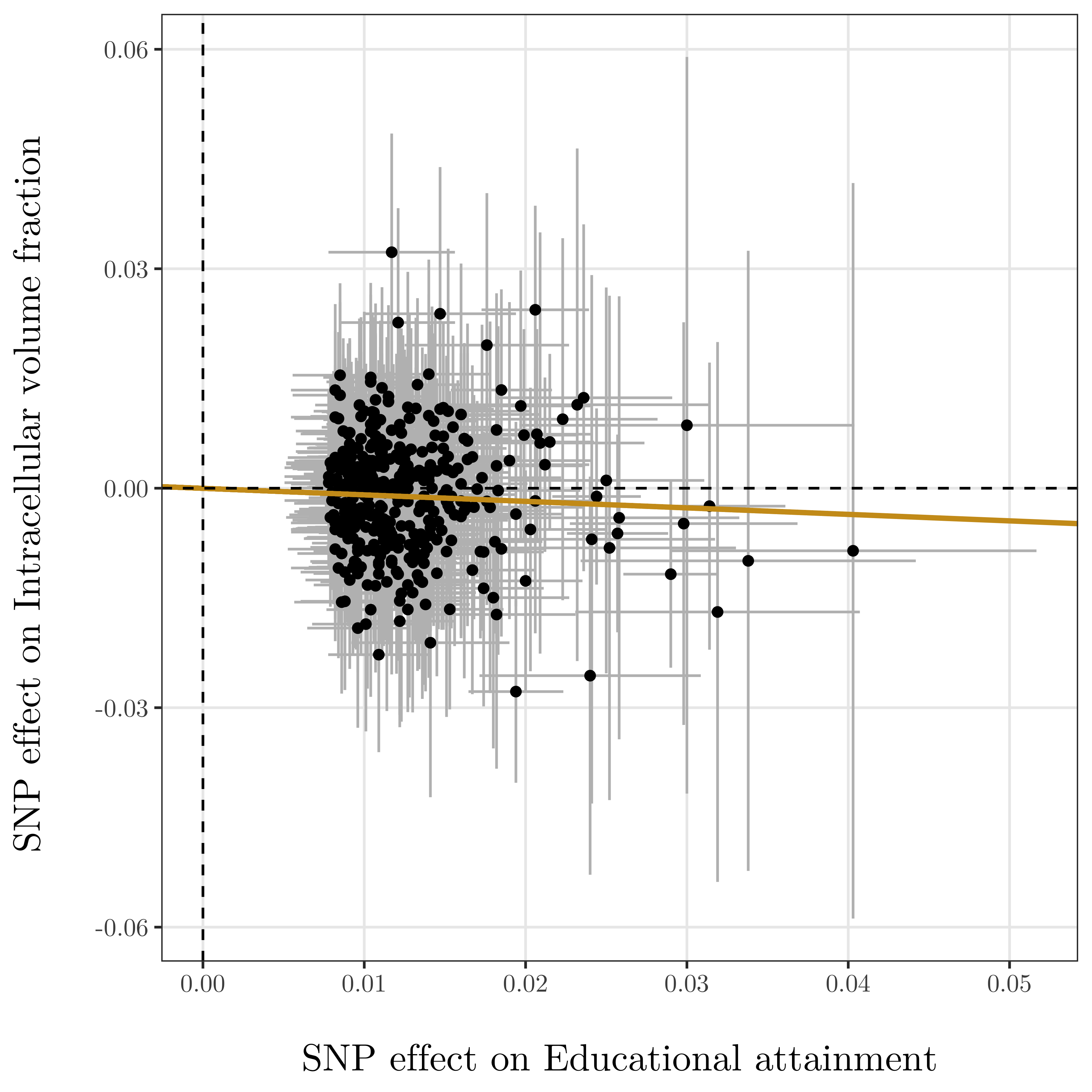 | 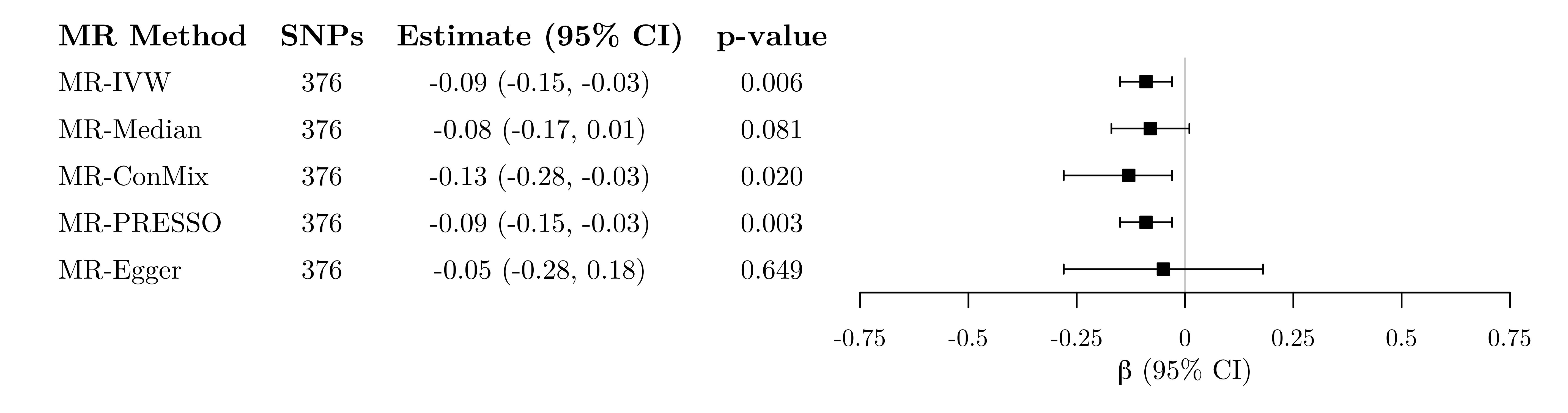 |
| --- | --- |
| 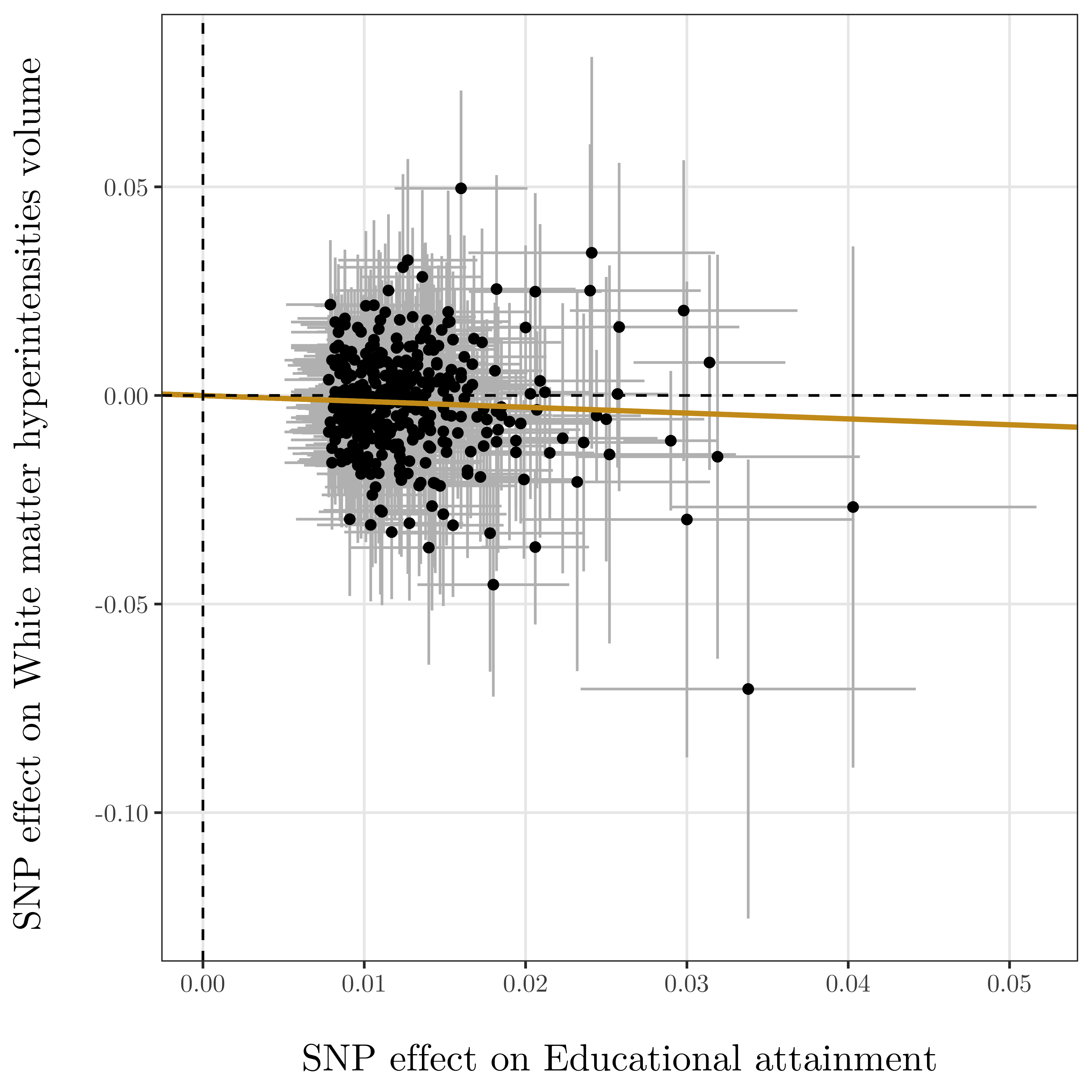 | 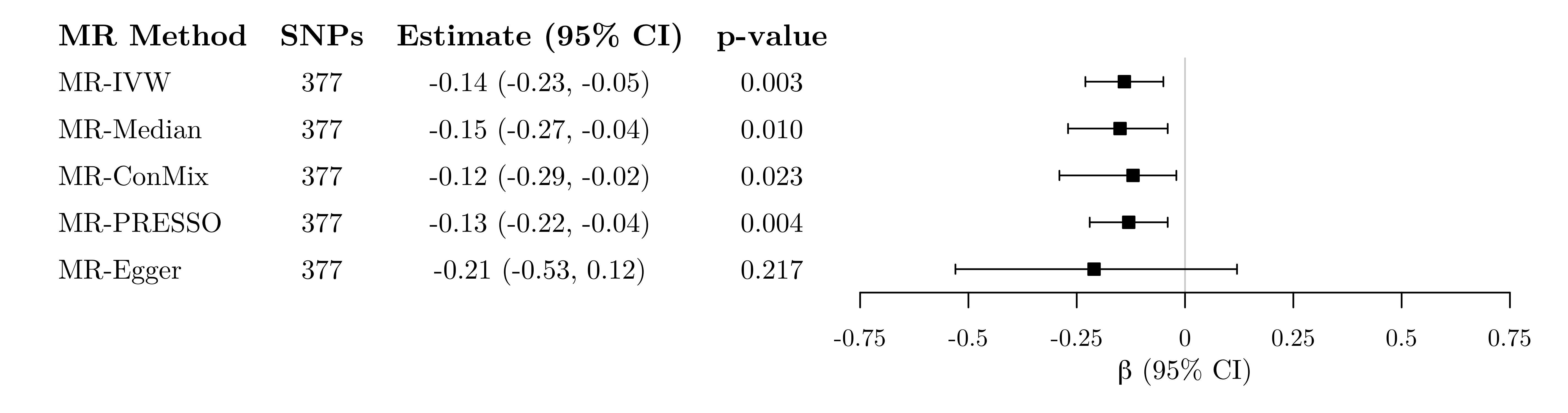 |

**Supplementary Fig. 6 (continued).** Left: Genetic associations with educational attainment (horizontal axis, standard deviation units) and with imaging-derived phenotypes (vertical axis, standard deviation units) for 376 genetic variants associated with educational attainment at a genome-wide level of significance. Horizontal and vertical lines represent 95% confidence intervals for the genetic associations. The regression line through the origin represents the inverse-variance weighted Mendelian randomization estimate for the effect of educational attainment on each imaging-derived phenotype. Right: Mendelian randomization estimates of the association between genetically-proxied educational attainment and imaging-derived brain structure phenotypes. Estimates represent standard deviation change in the imaging phenotype per 1 standard deviation increase in genetically-predicted years of schooling (approximately 4.2 years).

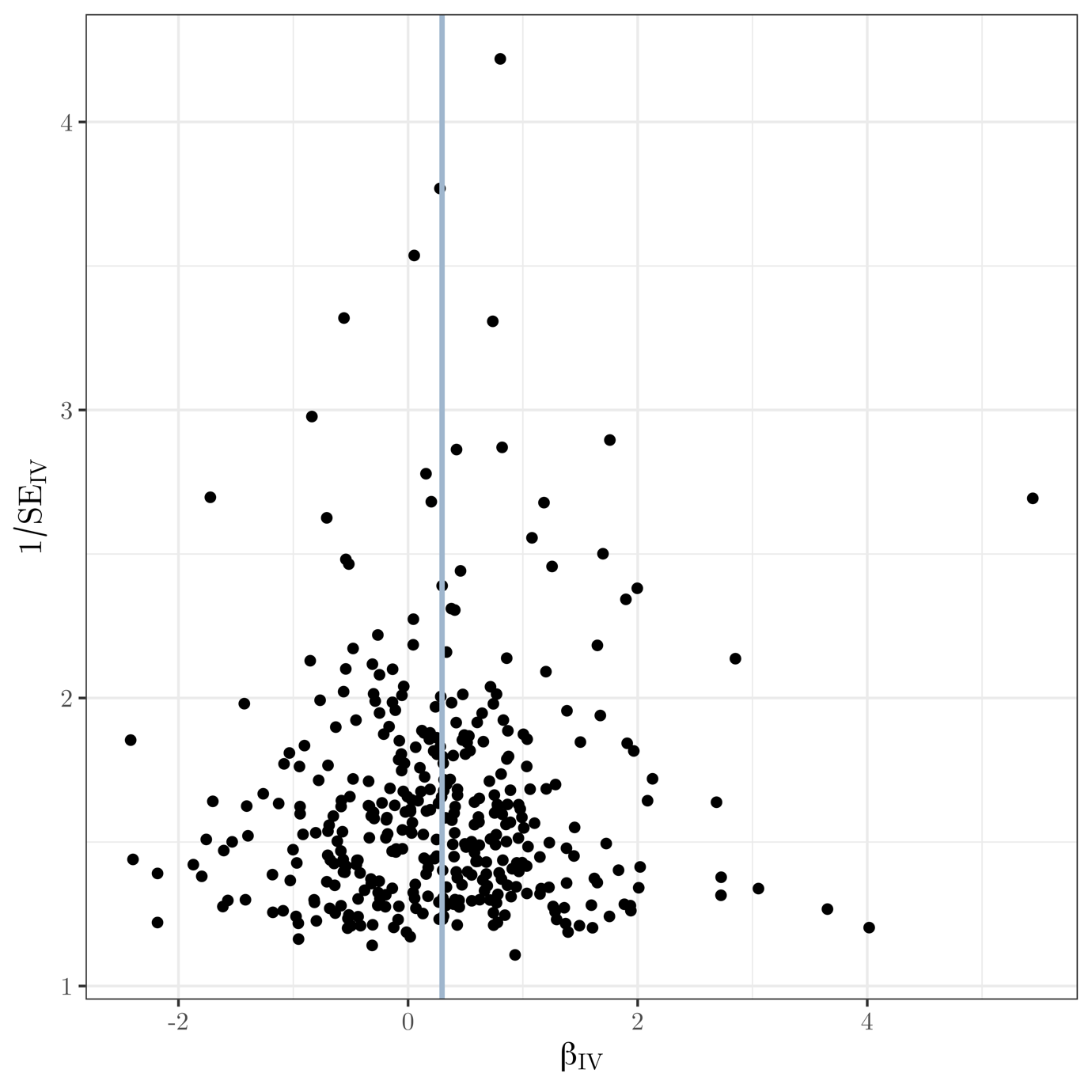

**Supplementary Fig. 7** Funnel plot of instrument precision against instrumental variable estimates for each genetic variant separately for Mendelian randomization analysis of educational attainment on surface area. Solid vertical line is the (fixed-effect) inverse-variance weighted estimate.

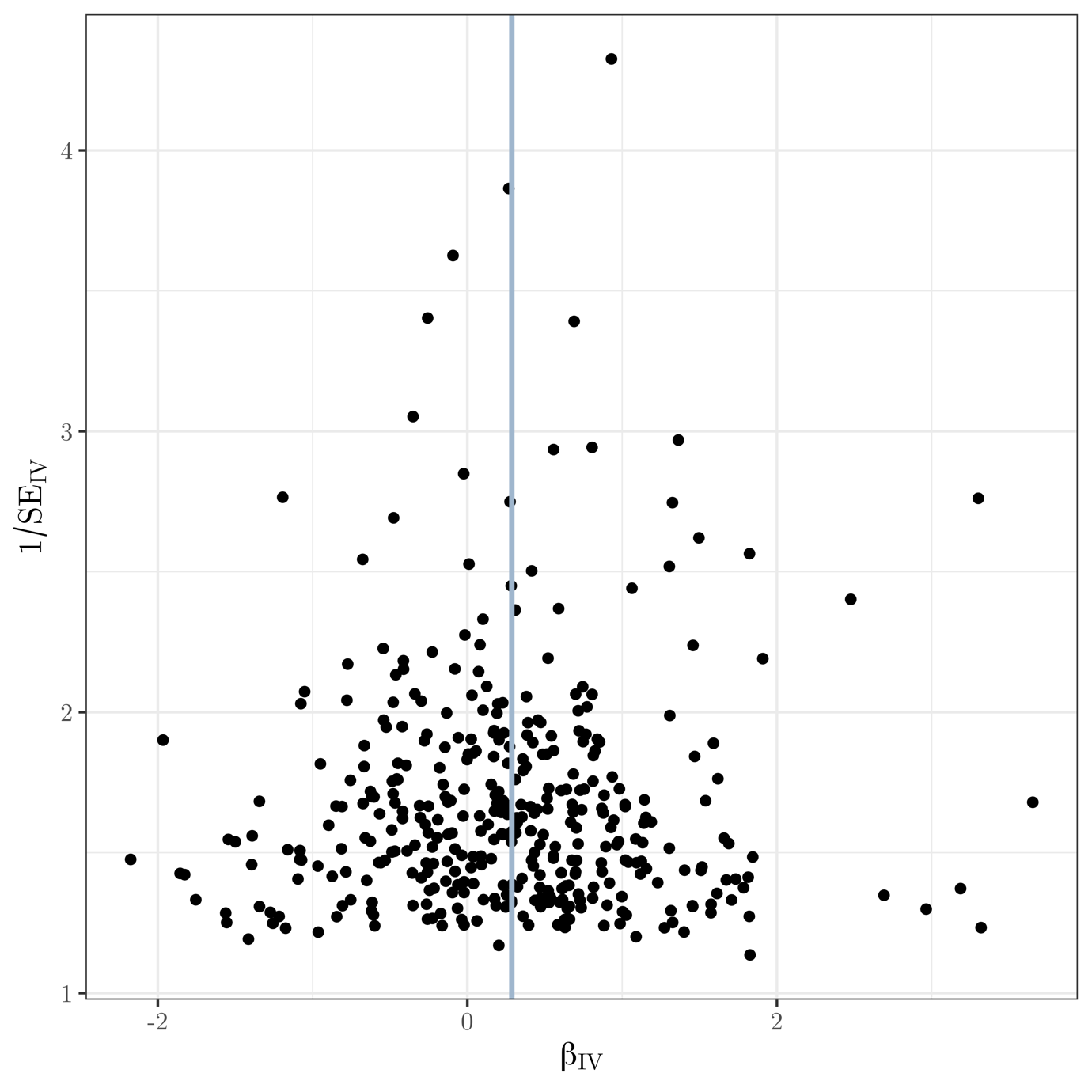

**Supplementary Fig. 8** Funnel plot of instrument precision against instrumental variable estimates for each genetic variant separately for Mendelian randomization analysis of educational attainment on volume. Solid vertical line is the (fixed-effect) inverse-variance weighted estimate.

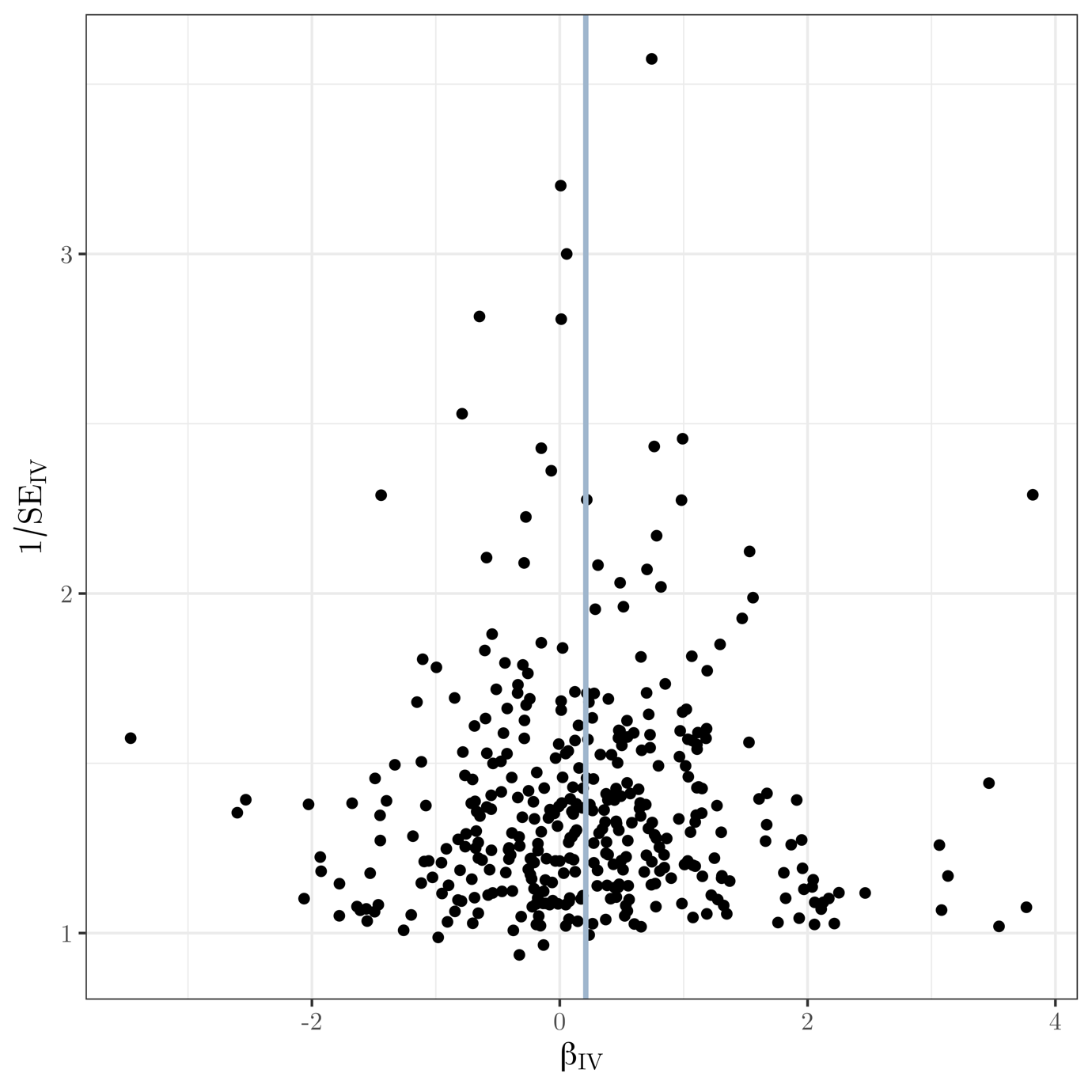

**Supplementary Fig. 9** Funnel plot of instrument precision against instrumental variable estimates for each genetic variant separately for Mendelian randomization analysis of educational attainment on local gyrification index. Solid vertical line is the (fixed-effect) inverse-variance weighted estimate.

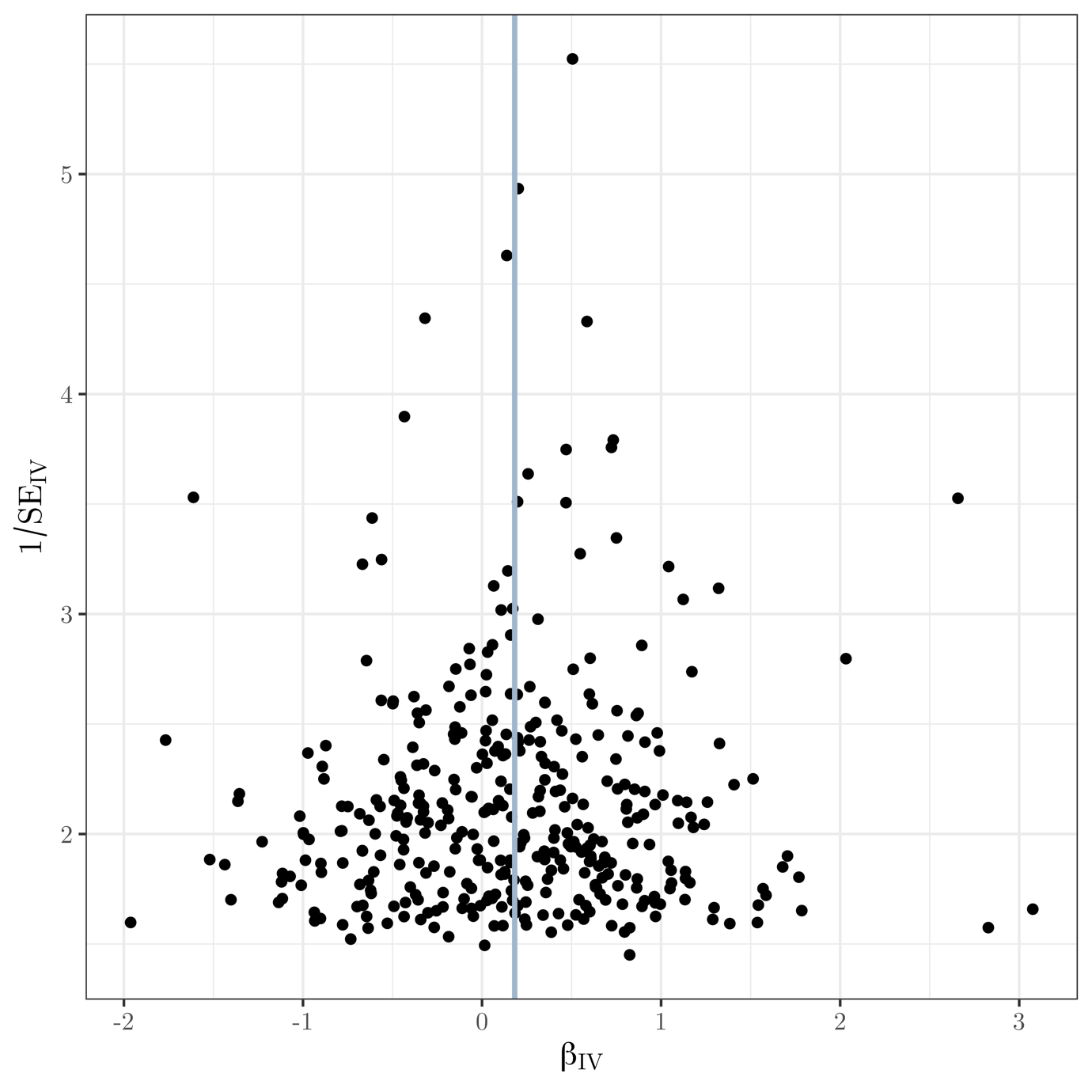

**Supplementary Fig. 10** Funnel plot of instrument precision against instrumental variable estimates for each genetic variant separately for Mendelian randomization analysis of educational attainment on intrinsic curvature. Solid vertical line is the (fixed-effect) inverse-variance weighted estimate.

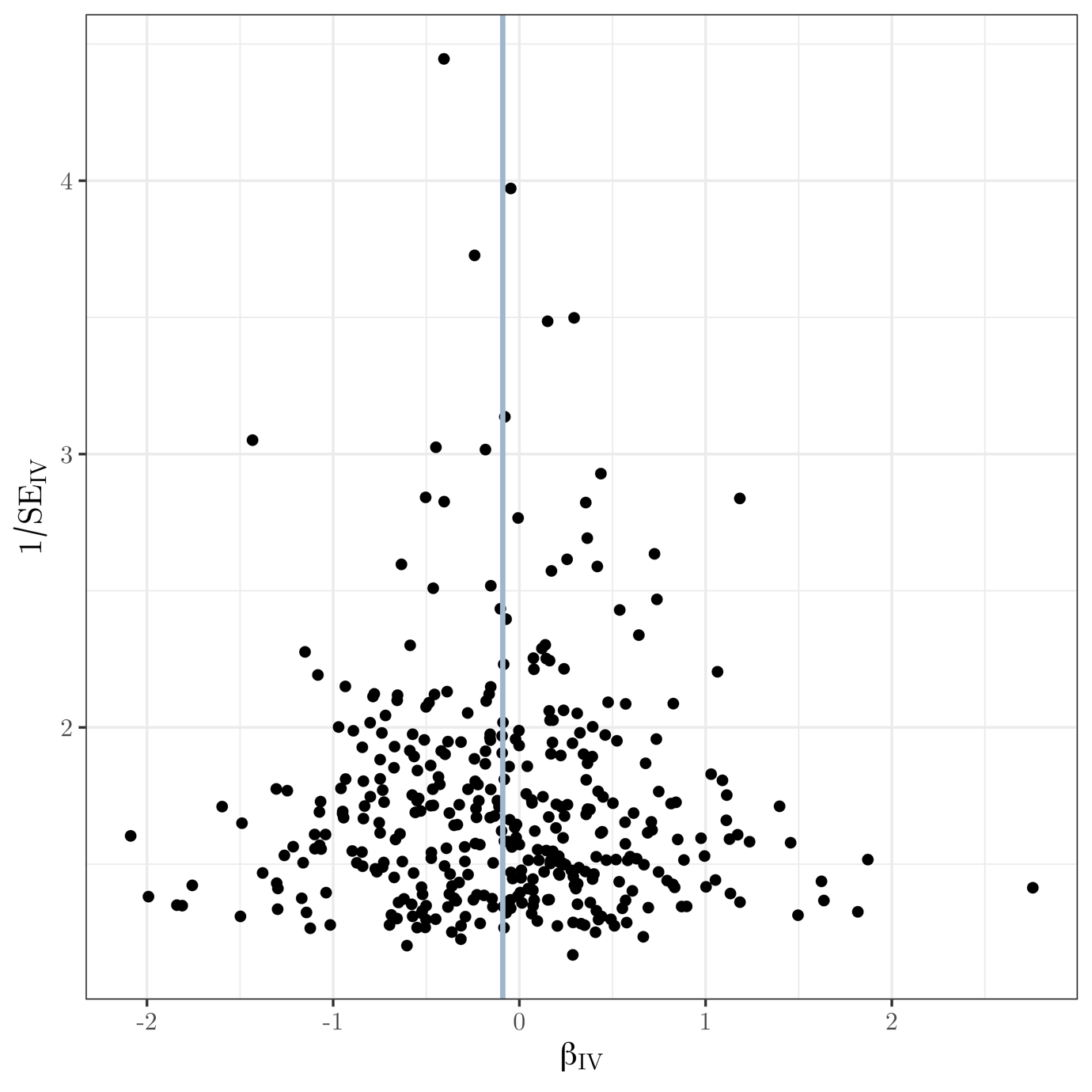

**Supplementary Fig. 11** Funnel plot of instrument precision against instrumental variable estimates for each genetic variant separately for Mendelian randomization analysis of educational attainment on intracellular volume fraction. Solid vertical line is the (fixed-effect) inverse-variance weighted estimate.

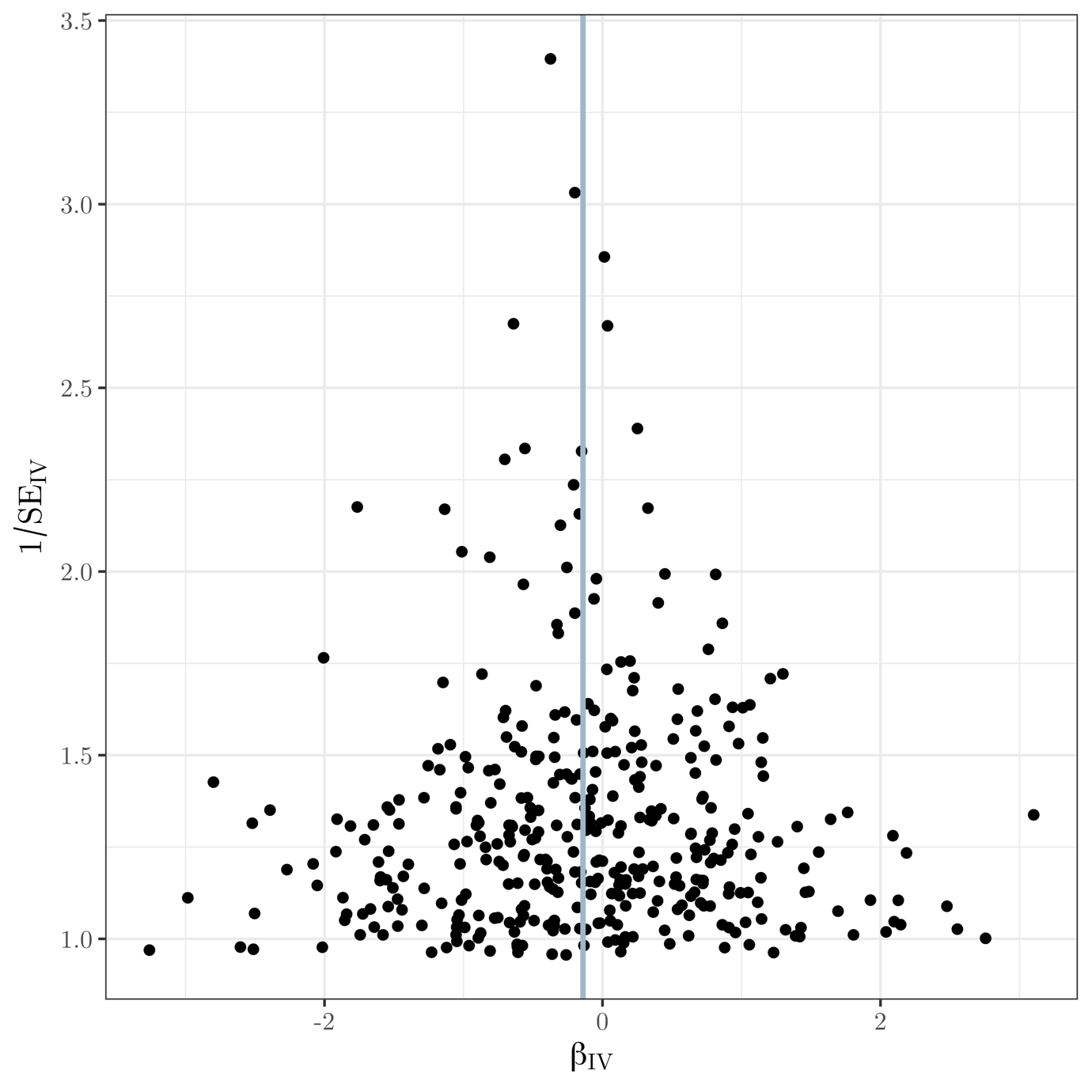

**Supplementary Fig. 12** Funnel plot of instrument precision against instrumental variable estimates for each genetic variant separately for Mendelian randomization analysis of educational attainment on total volume of white matter hyperintensities. Solid vertical line is the (fixed-effect) inverse-variance weighted estimate.

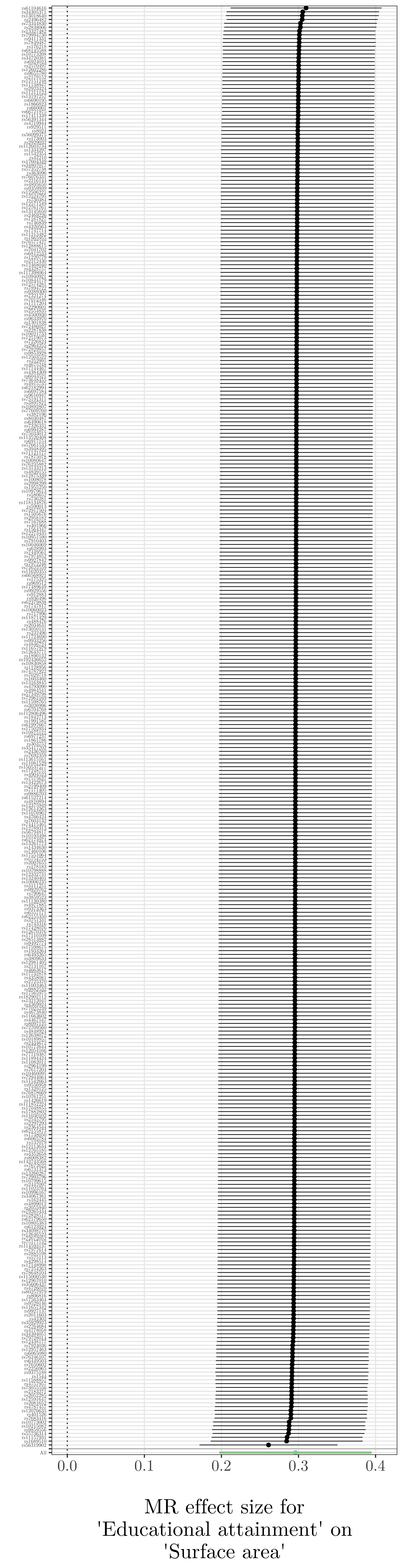

**Supplementary Fig. 13** Leave-one-out plot for MR analysis of educational attainment on surface area.

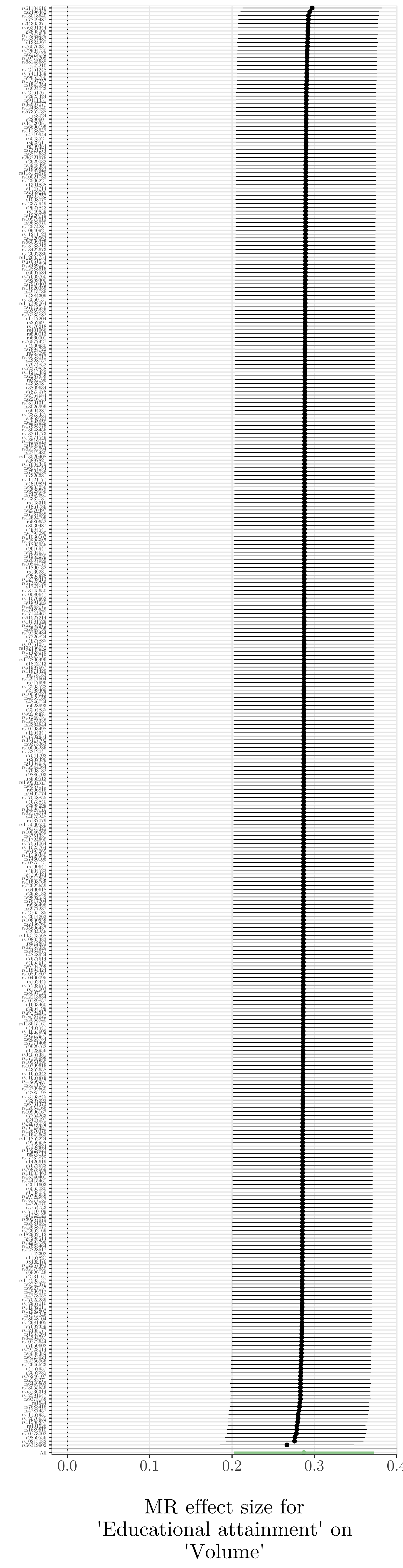

**Supplementary Fig. 14** eave-one-out plot for MR analysis of educational attainment on volume.

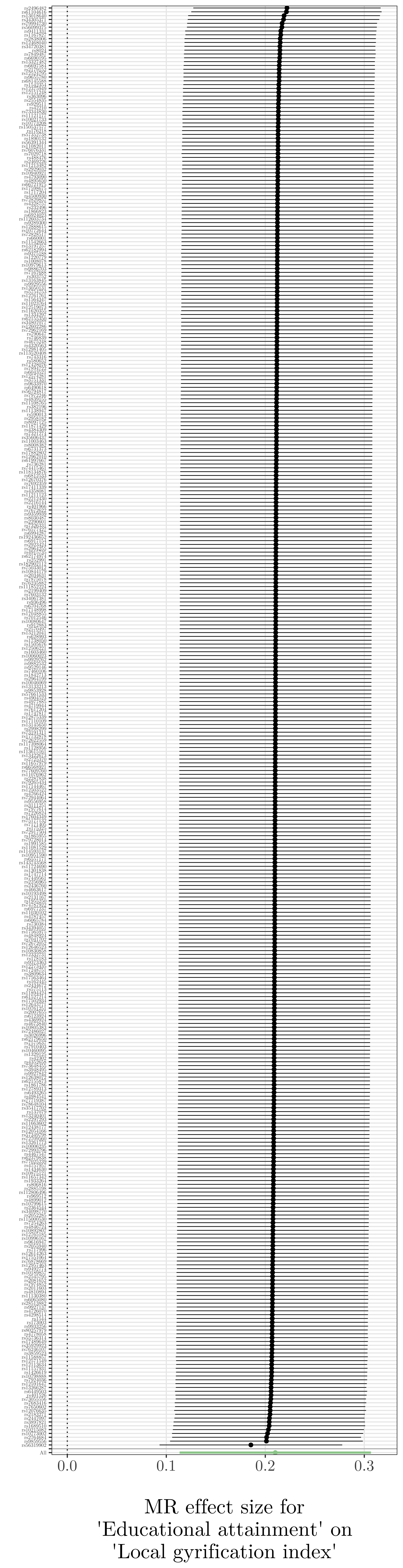

**Supplementary Fig. 15** Leave-one-out plot for MR analysis of educational attainment on local gyrification index.

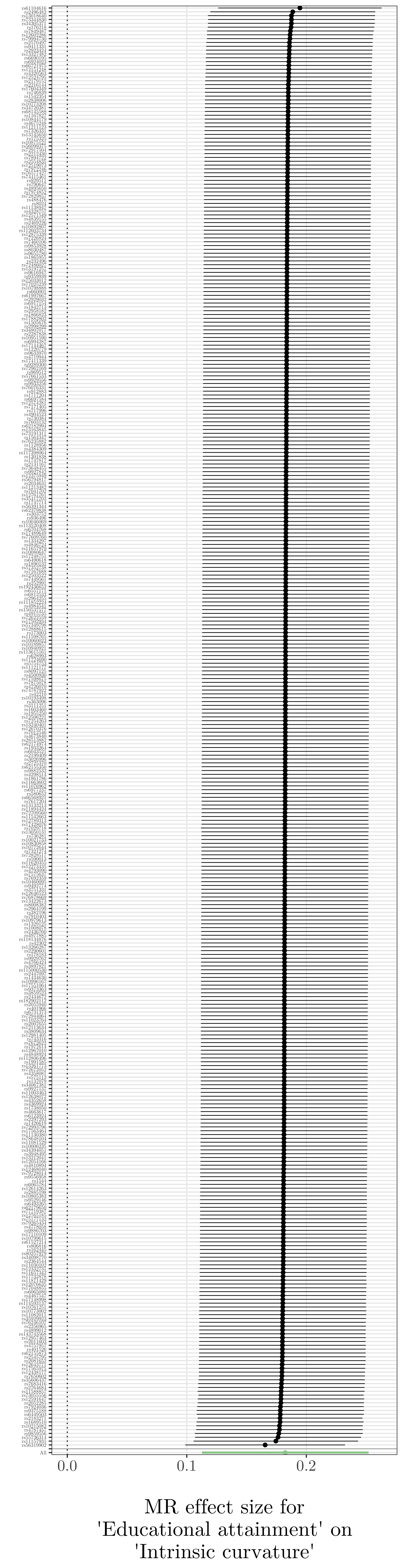

**Supplementary Fig. 16** Leave-one-out plot for MR analysis of educational attainment on intrinsic curvature.

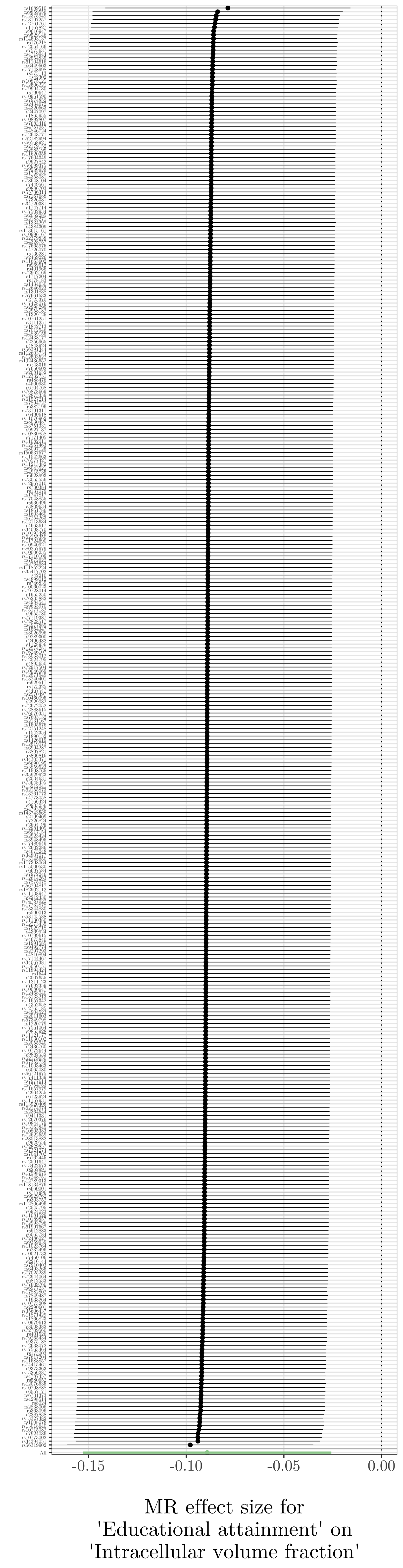

**Supplementary Fig. 17** Leave-one-out plot for MR analysis of educational attainment on intracellular volume fraction.

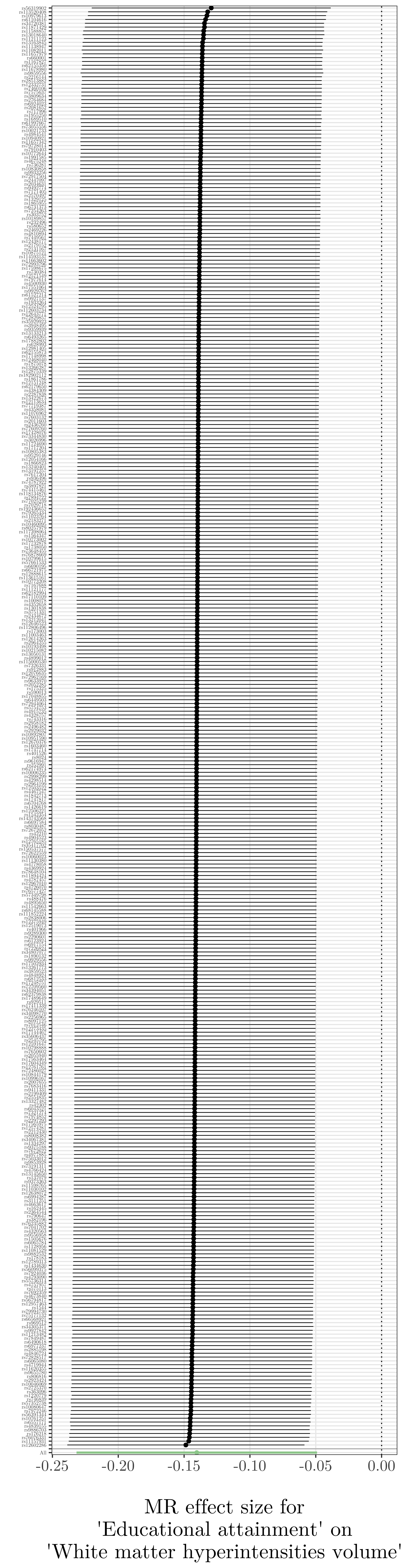

**Supplementary Fig. 18** Leave-one-out plot for MR analysis of educational attainment on total volume of white matter hyperintensities.

| 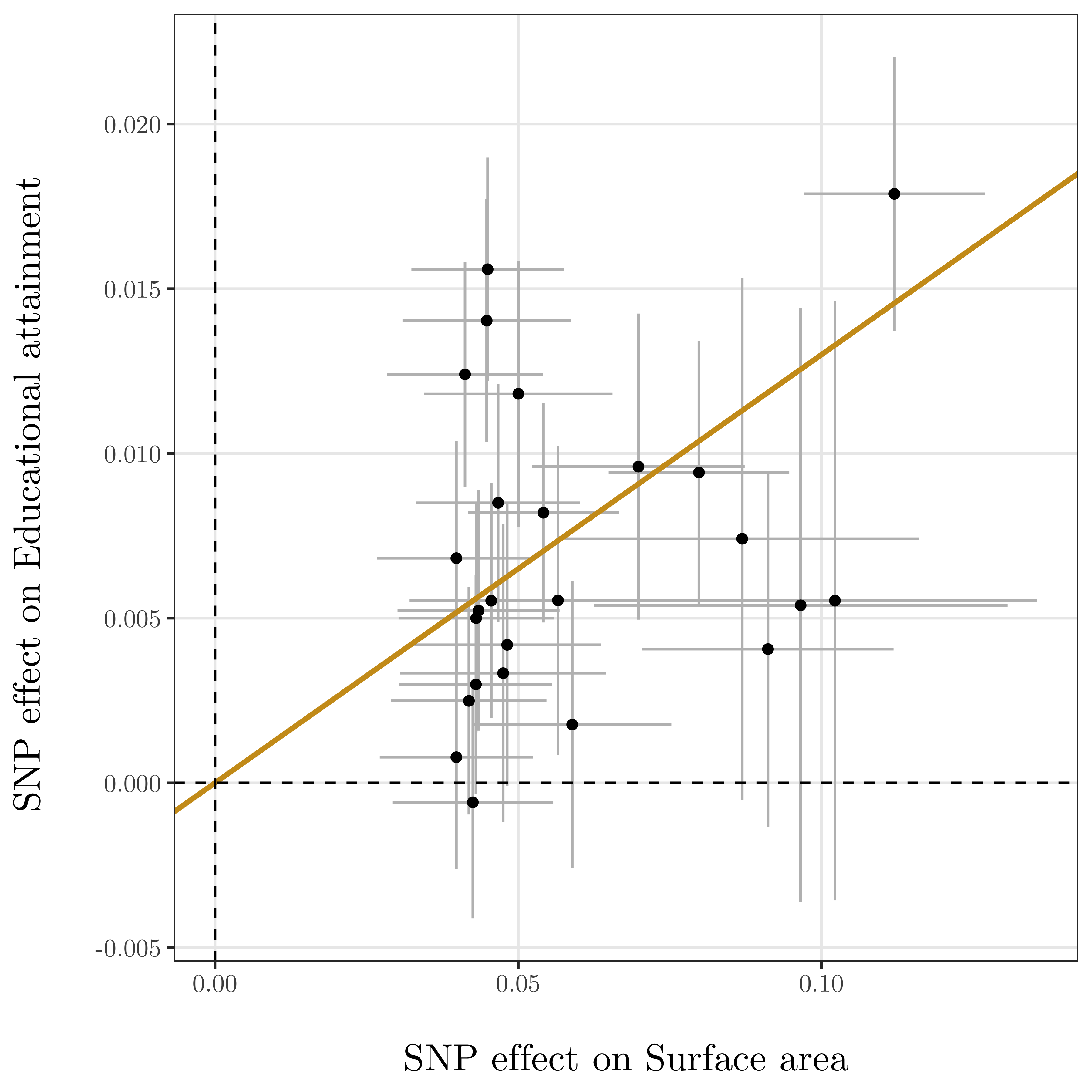 | 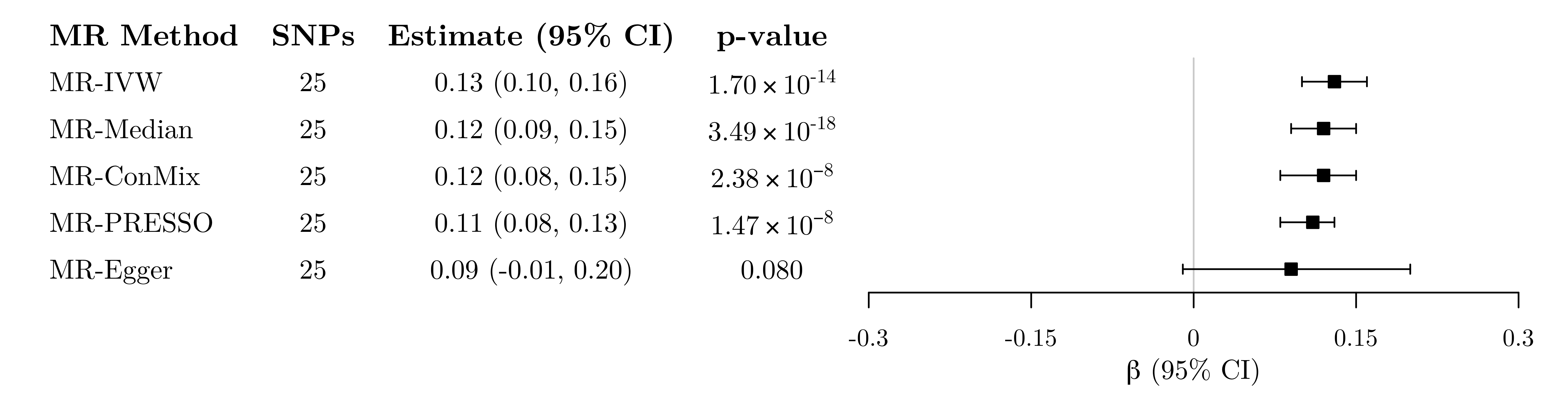 |
| --- | --- |
| 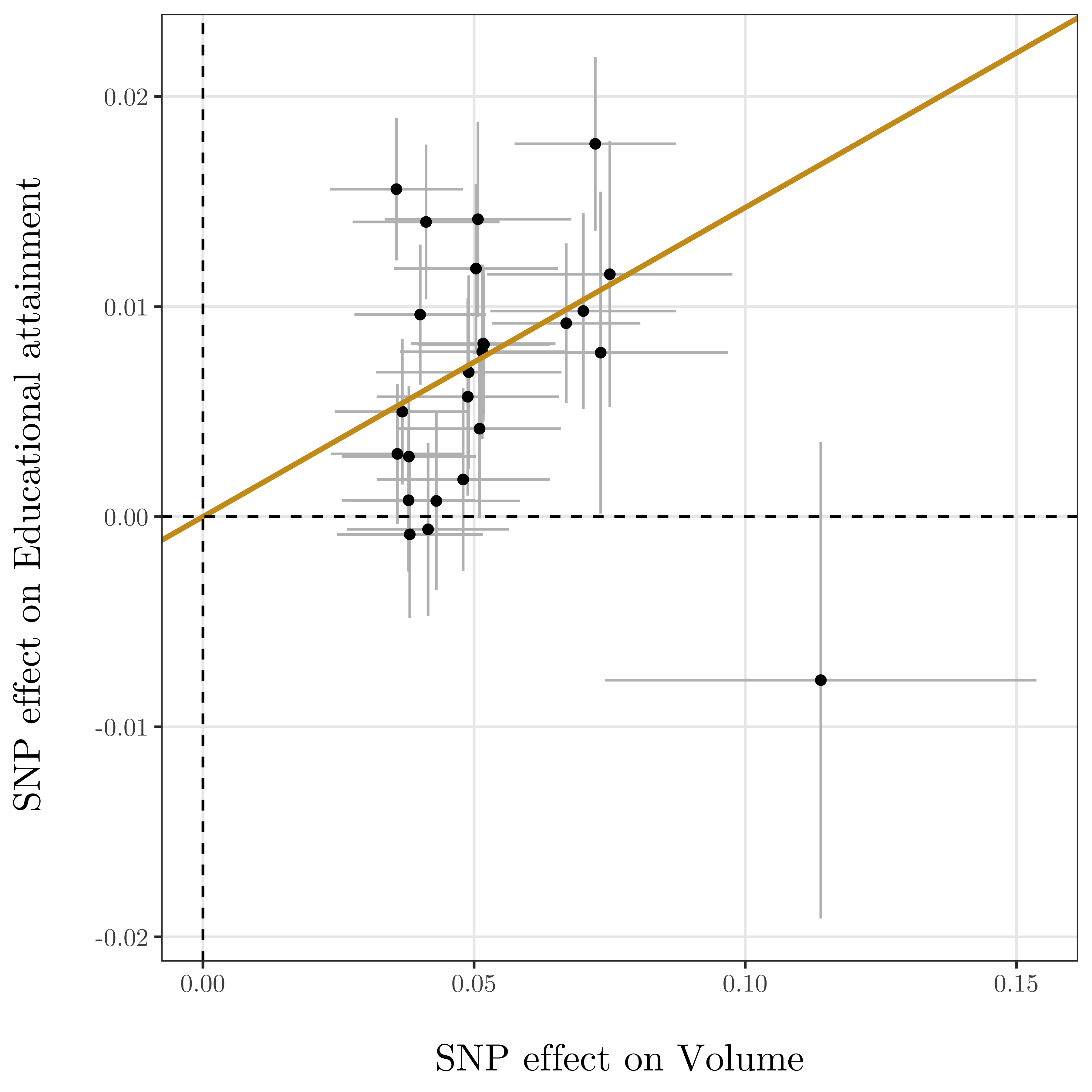 | 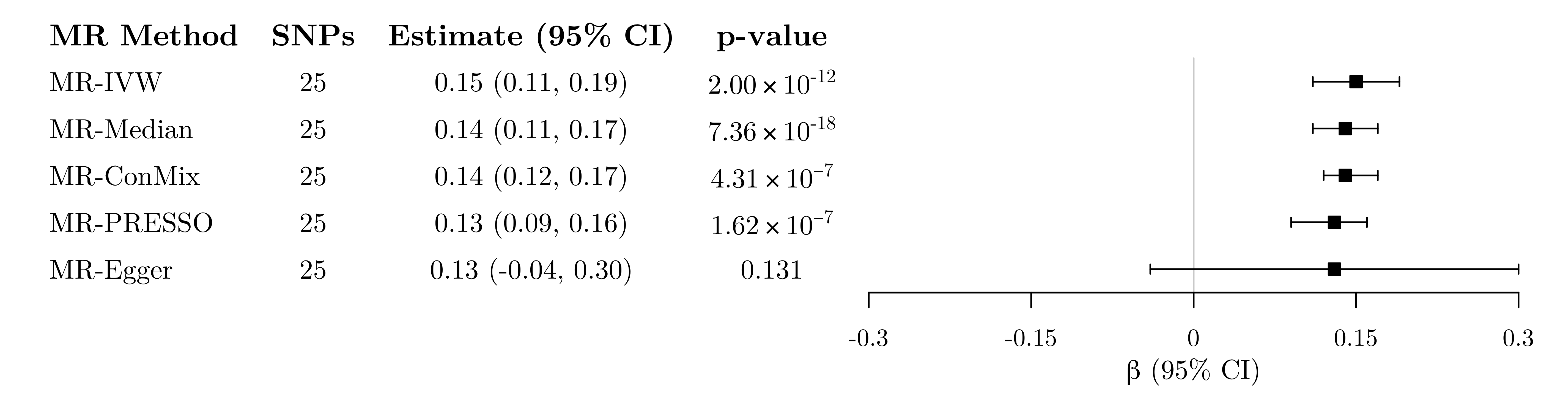 |
| 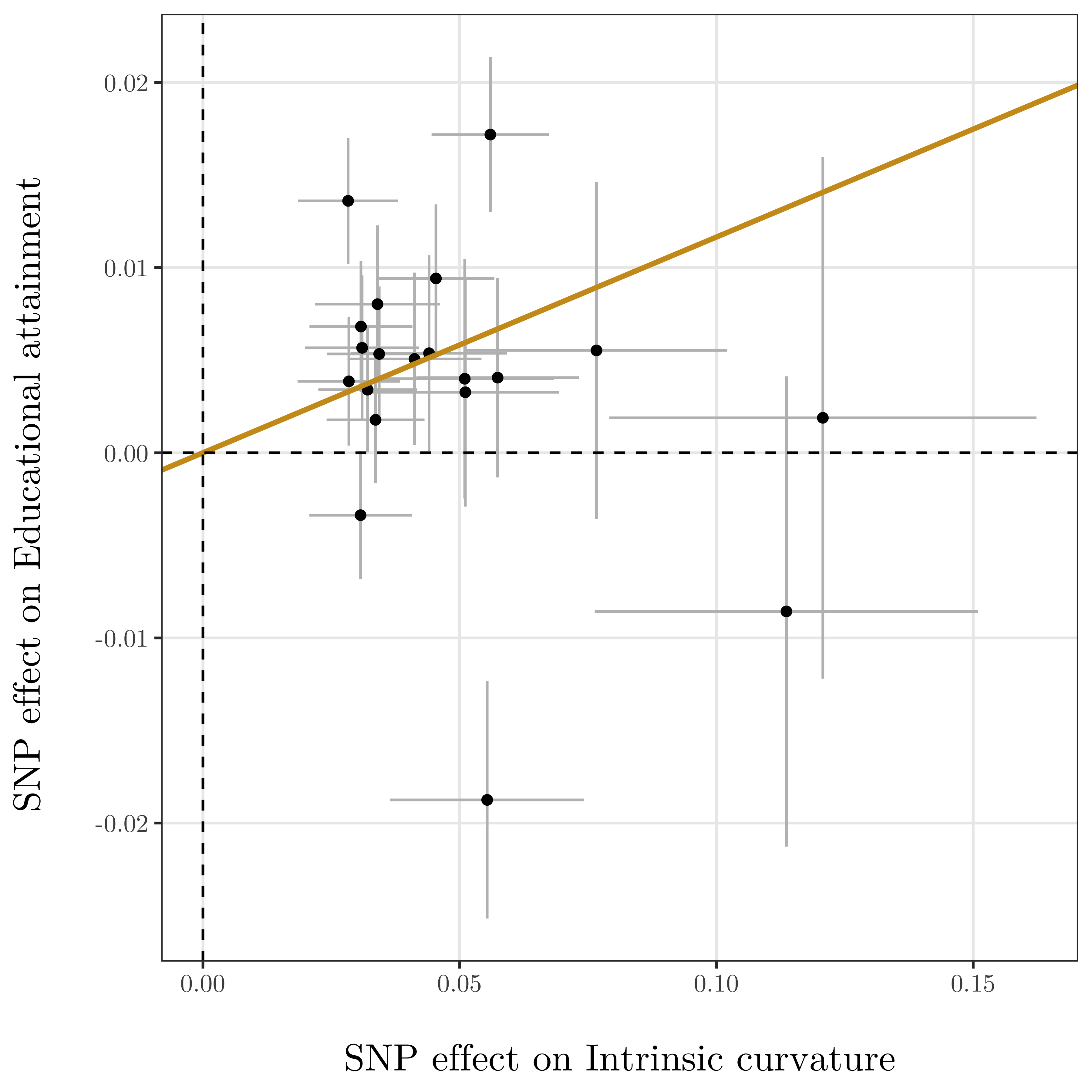 | 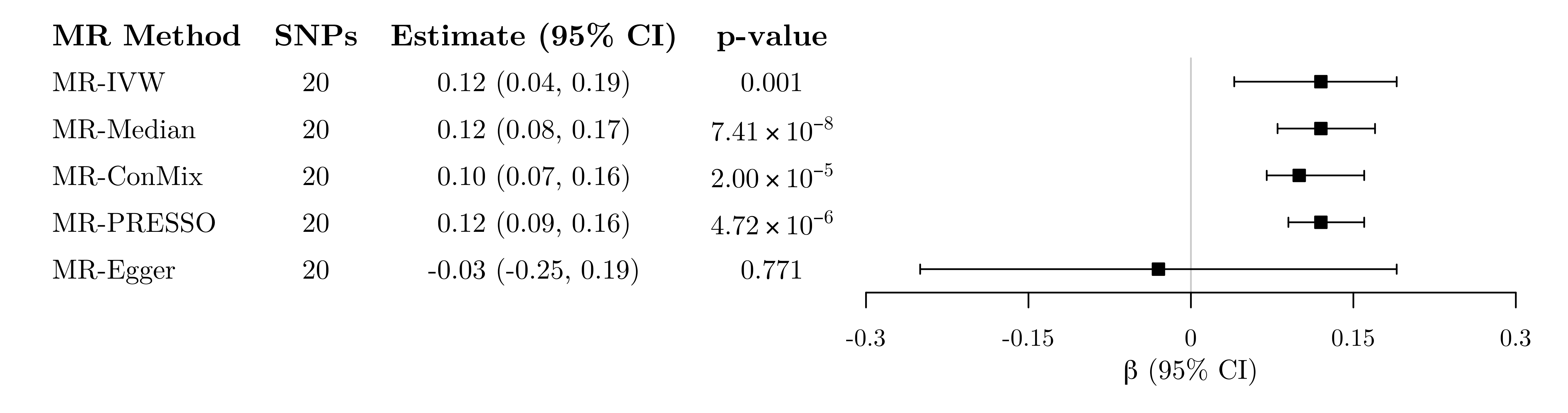 |

**Supplementary Fig. 19** Left: Genetic associations with imaging-derived phenotypes (horizontal axis, standard deviation units) and educational attainment (vertical axis, standard deviation units) for genetic variants associated with each imaging-derived phenotype at a genome-wide level of significance. Horizontal and vertical lines represent 95% confidence intervals for the genetic associations. The regression line through the origin represents the inverse-variance weighted Mendelian randomization estimate for the effect of each imaging-derived phenotype on educational attainment. Right: Mendelian randomization estimates of the association between genetically-proxied brain structure phenotypes and educational attainment. Estimates represent the standard deviation change in years of schooling (95% confidence intervals) per 1 standard deviation increase in genetically-predicted levels of each imaging-derived phenotype.

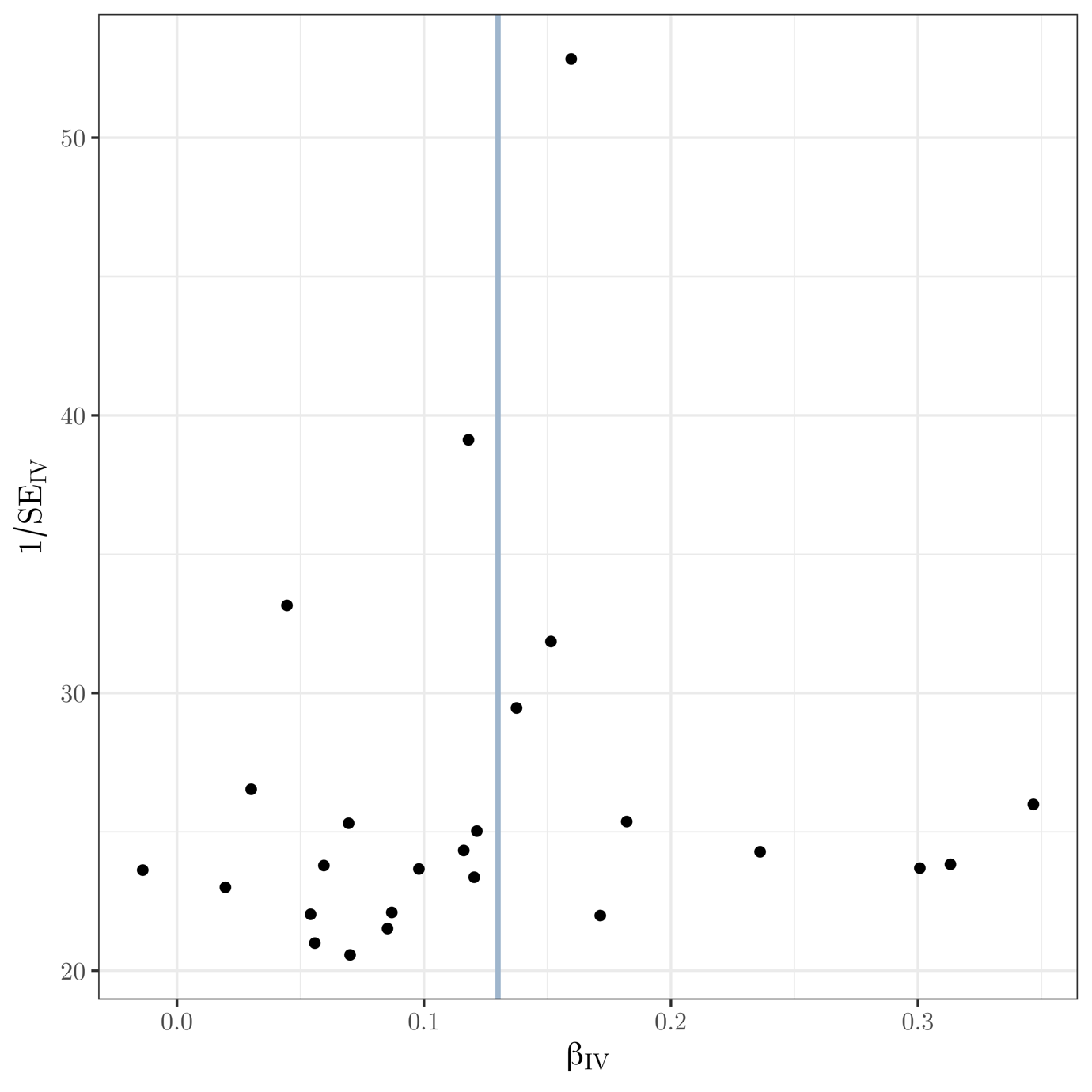

**Supplementary Fig. 20** Funnel plot of instrument precision against instrumental variable estimates for each genetic variant separately for Mendelian randomization analysis of surface area on educational attainment. Solid vertical line is the (fixed-effect) inverse-variance weighted estimate.

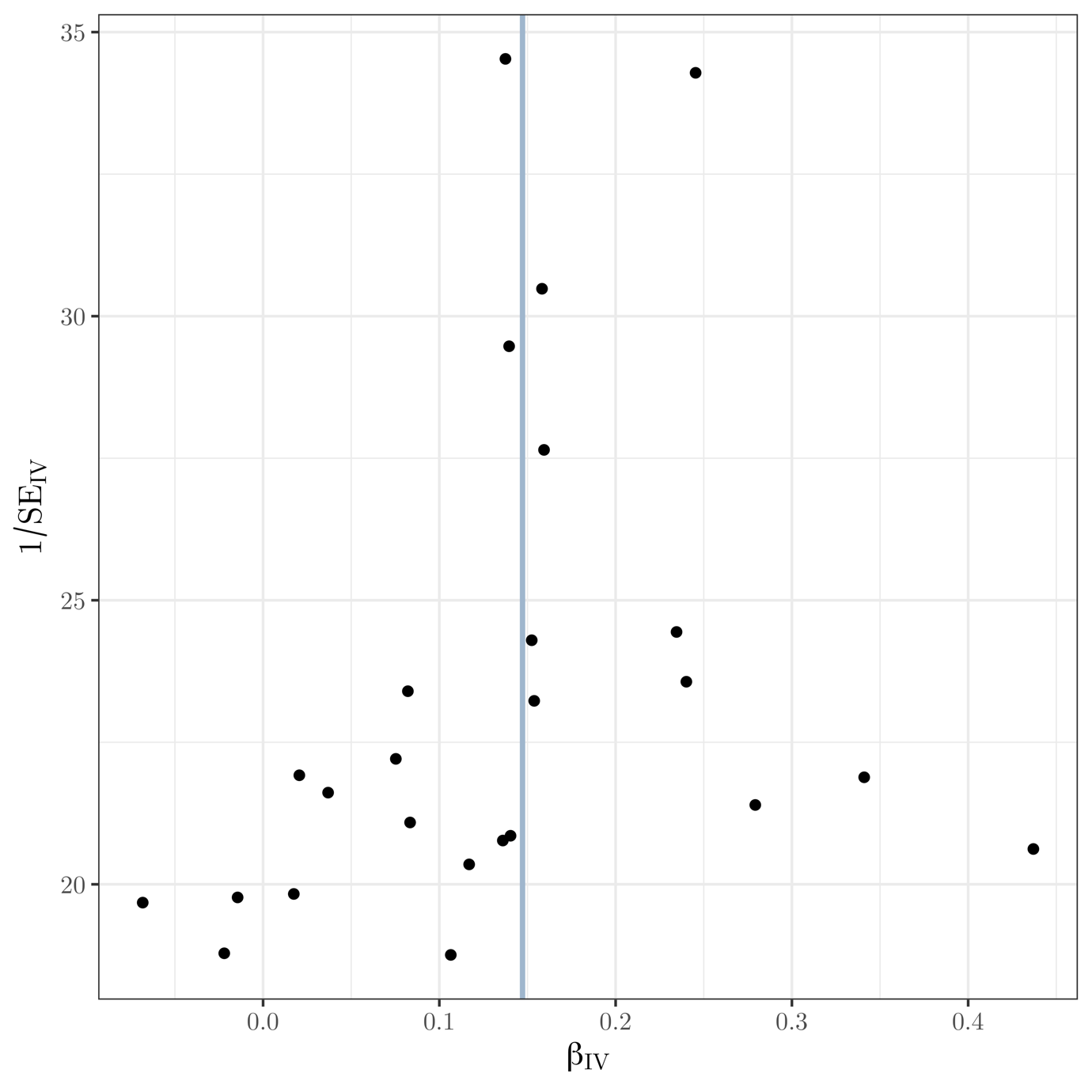

**Supplementary Fig. 21** Funnel plot of instrument precision against instrumental variable estimates for each genetic variant separately for Mendelian randomization analysis of volume on educational attainment. Solid vertical line is the (fixed-effect) inverse-variance weighted estimate.

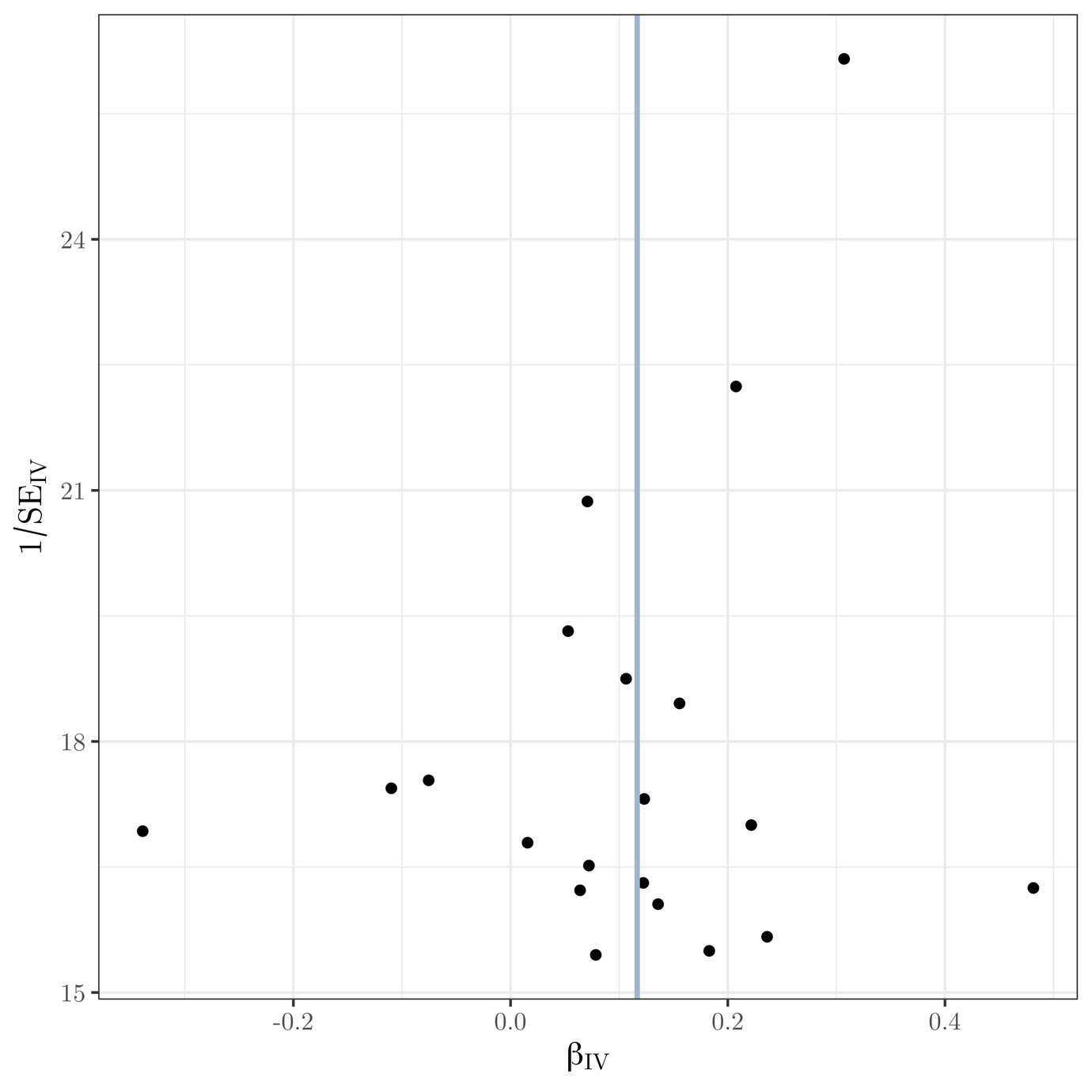

**Supplementary Fig. 22** Funnel plot of instrument precision against instrumental variable estimates for each genetic variant separately for Mendelian randomization analysis of intrinsic curvature on educational attainment. Solid vertical line is the (fixed-effect) inverse-variance weighted estimate.

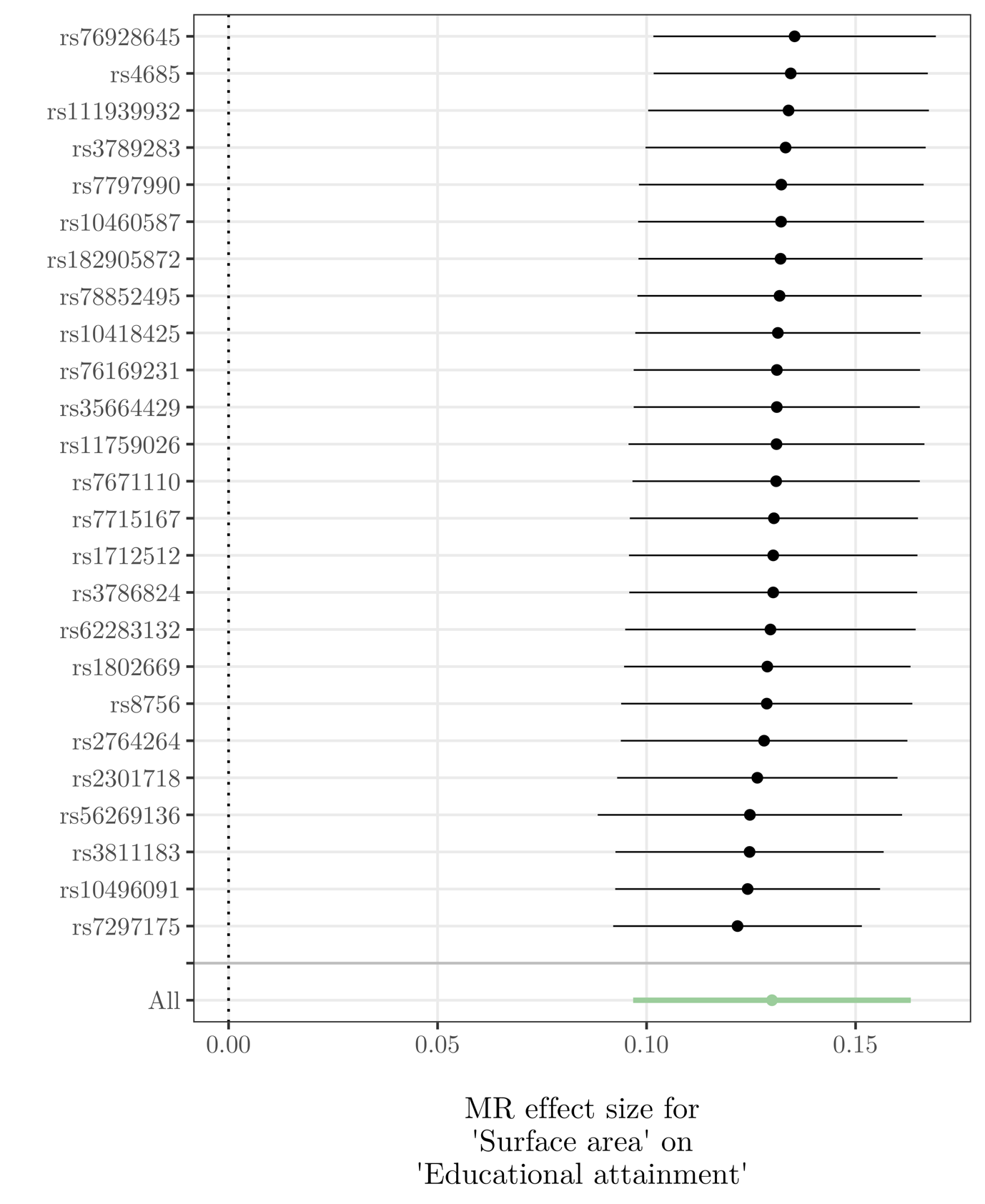

**Supplementary Fig. 23** Leave-one-out plot for MR analysis of surface area on educational attainment.

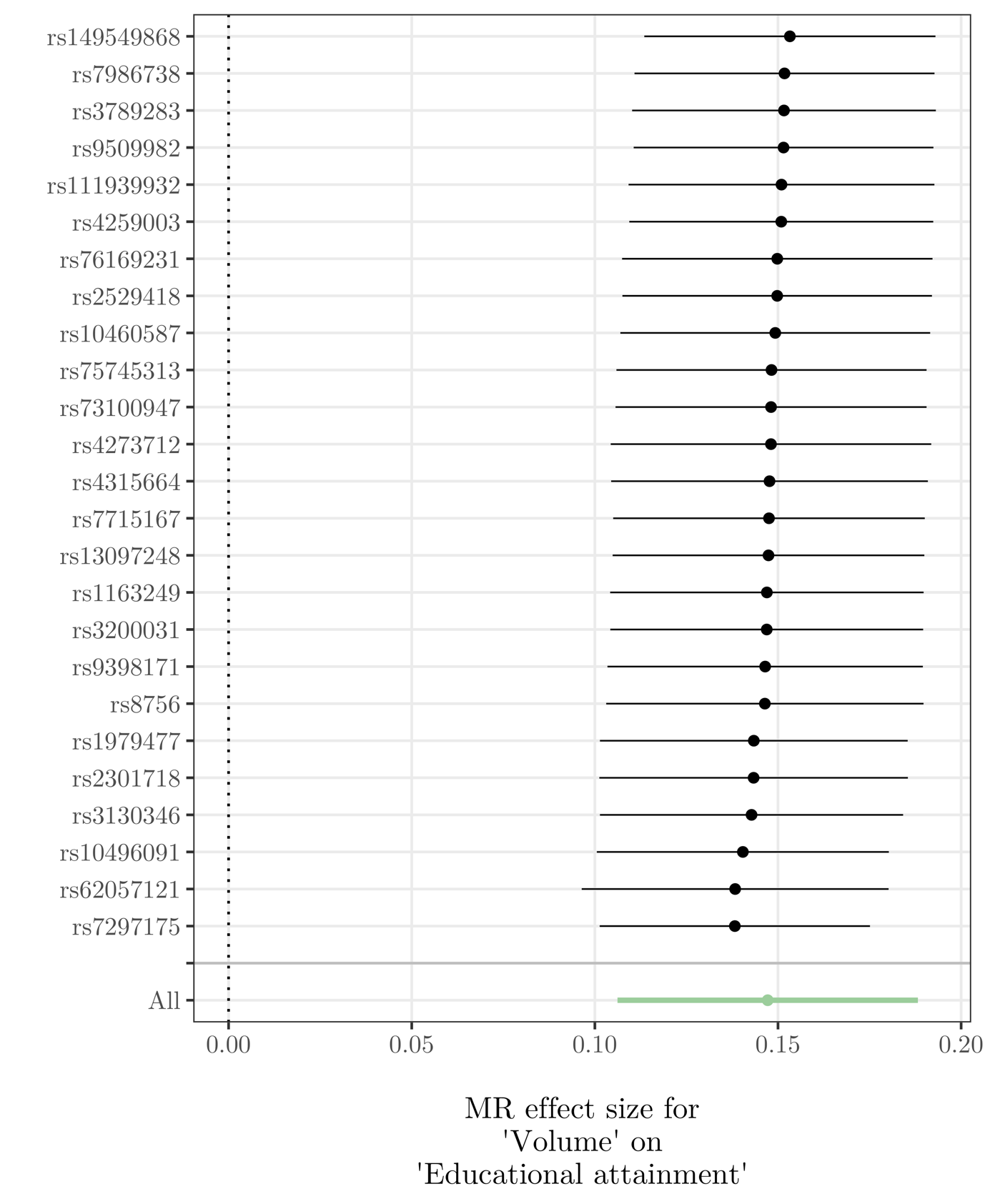

**Supplementary Fig. 24** Leave-one-out plot for MR analysis of volume on educational attainment.

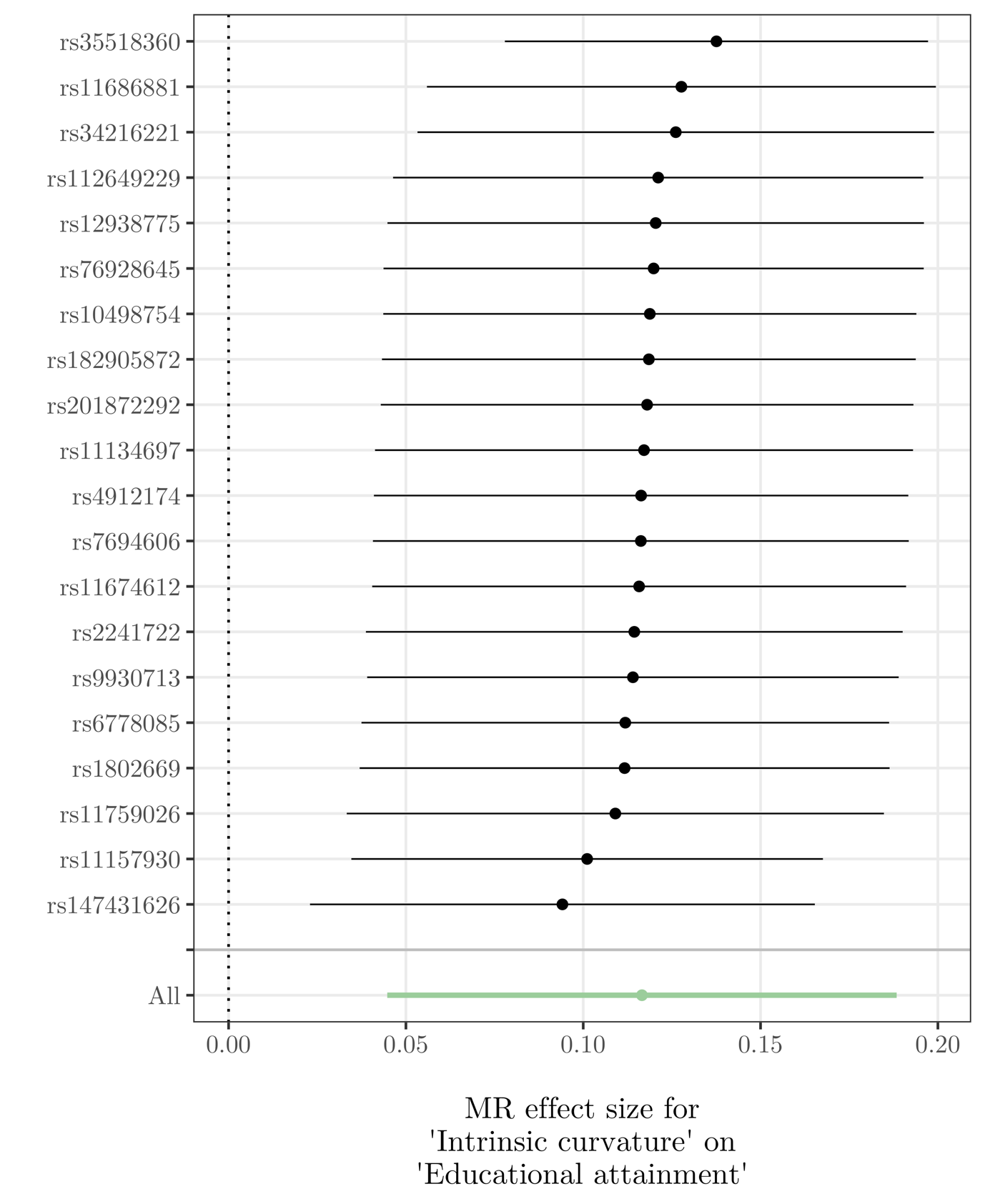

**Supplementary Fig. 25** Leave-one-out plot for MR analysis of intrinsic curvature on educational attainment.

| 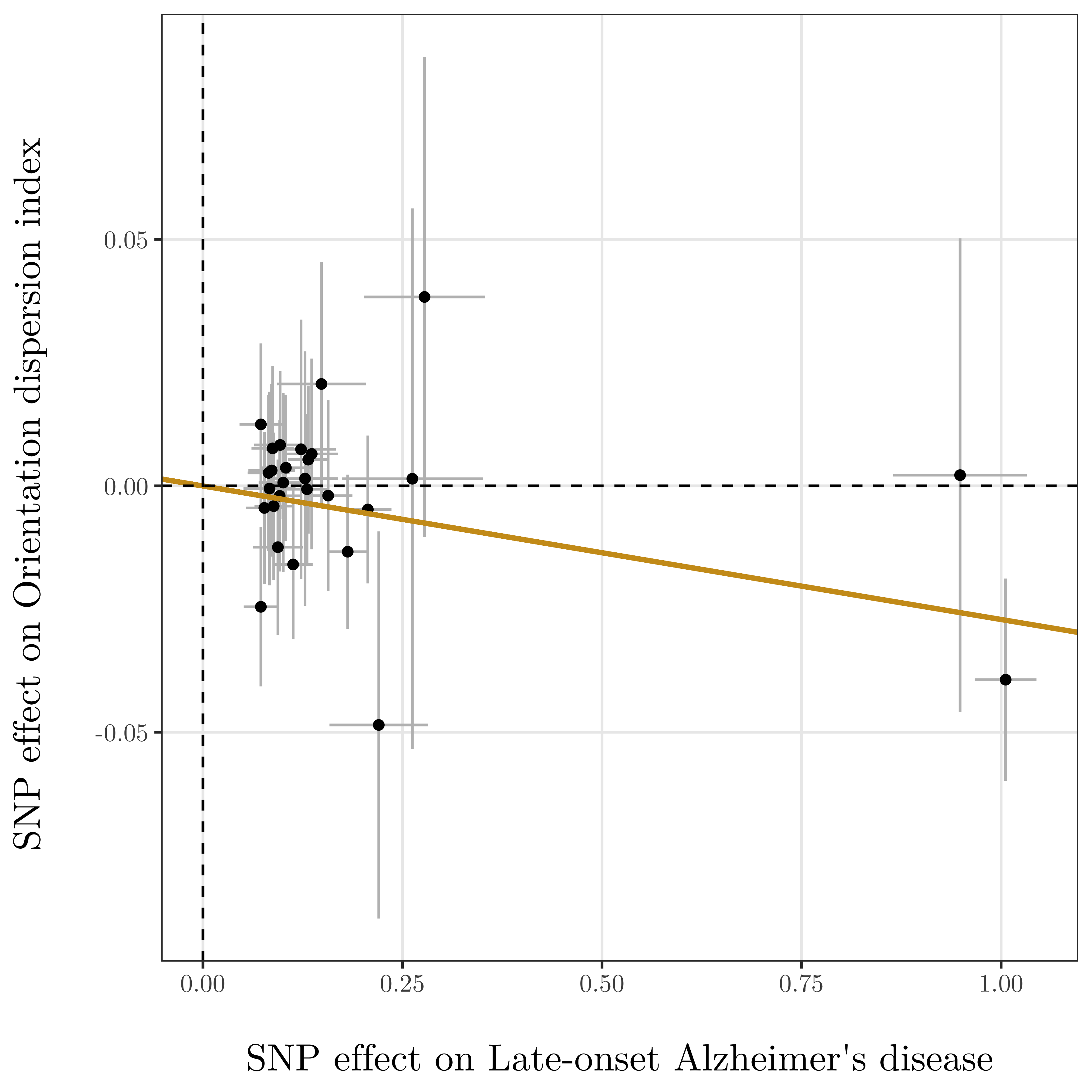 | 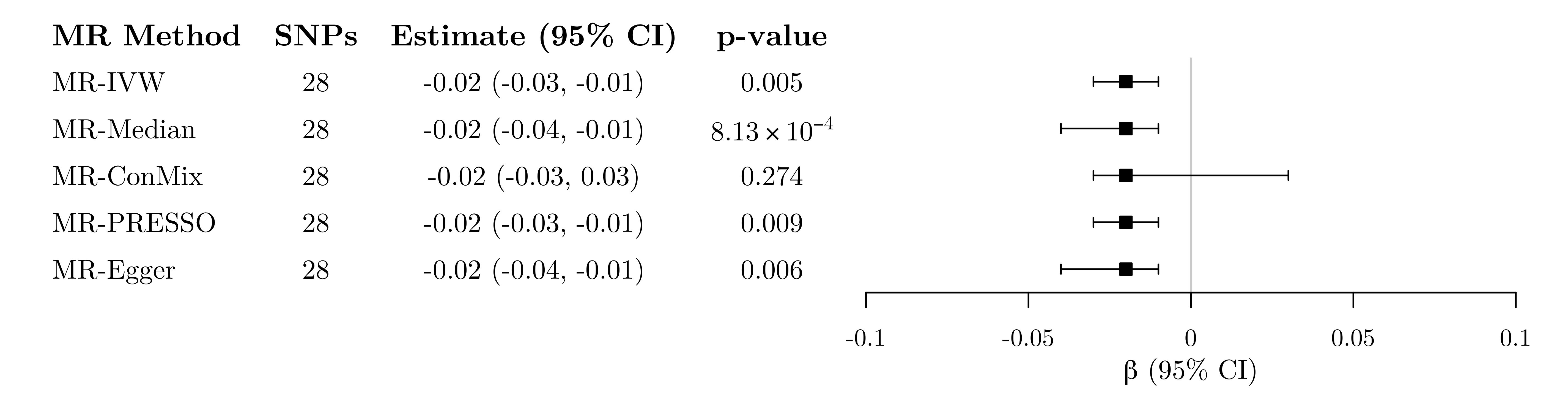 |
| --- | --- |
| 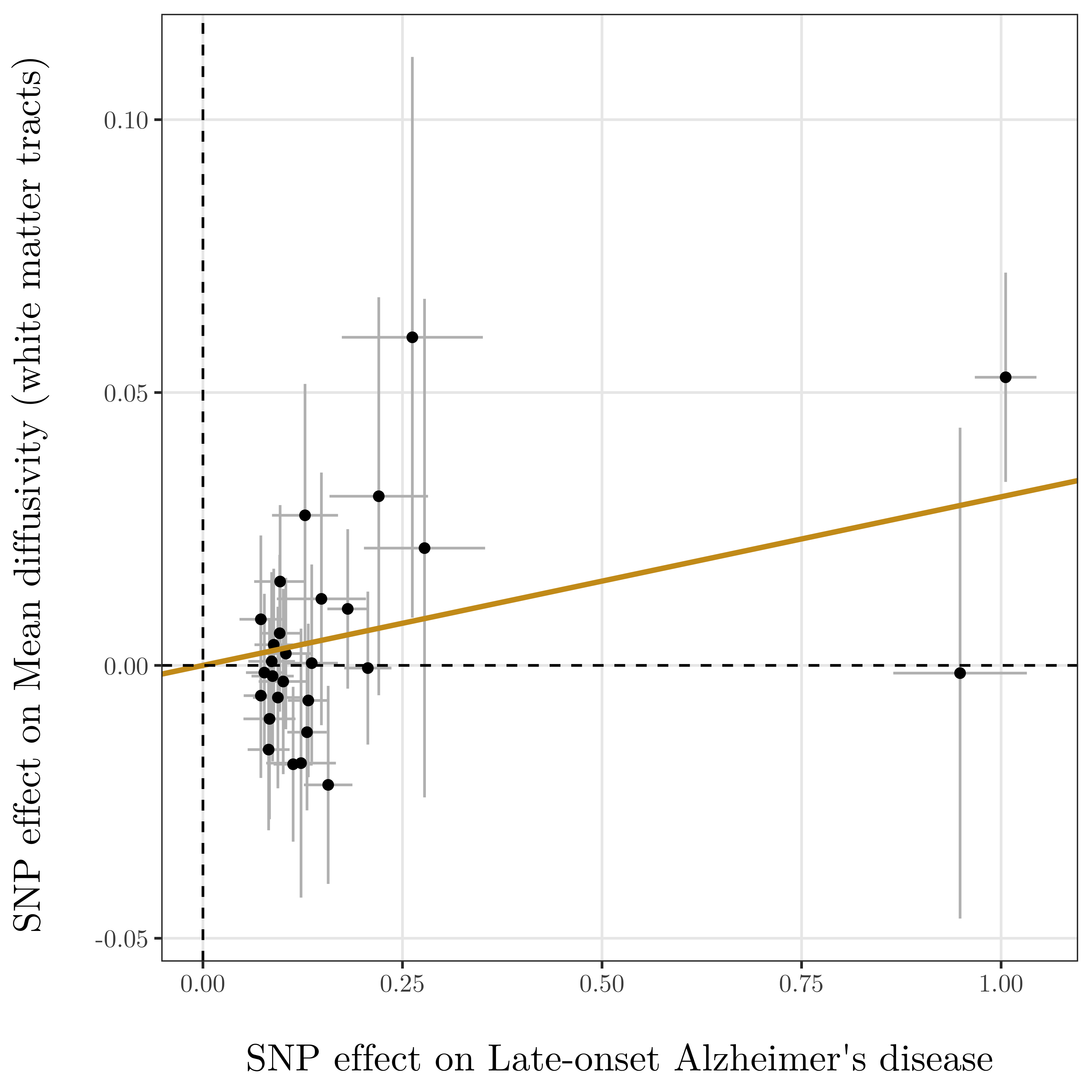 | 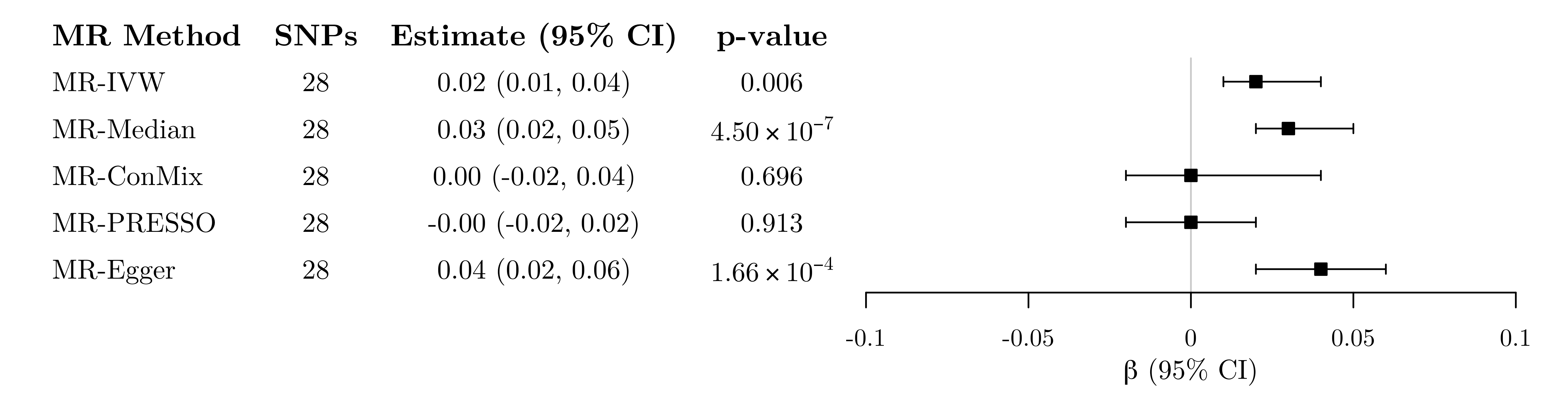 |
| 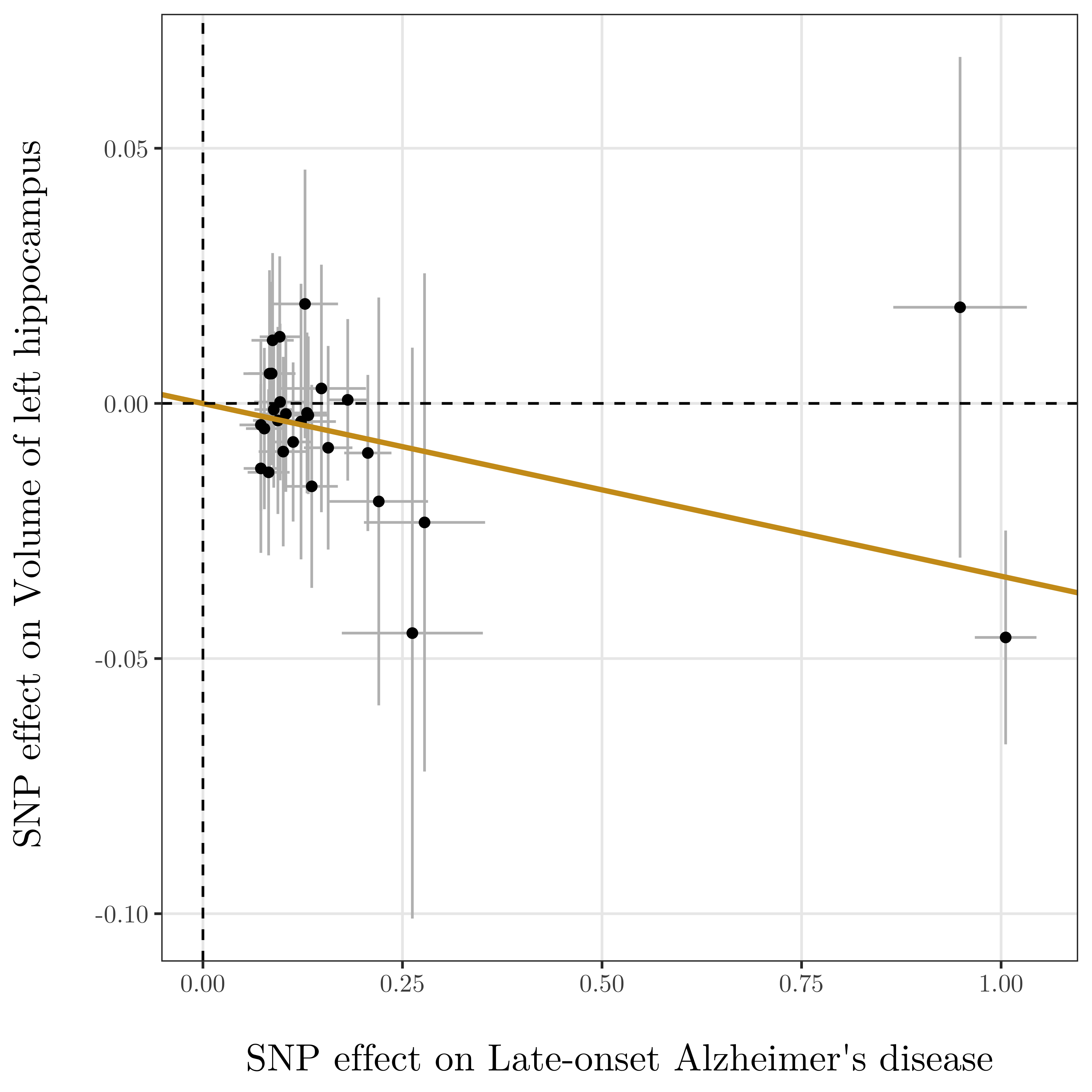 | 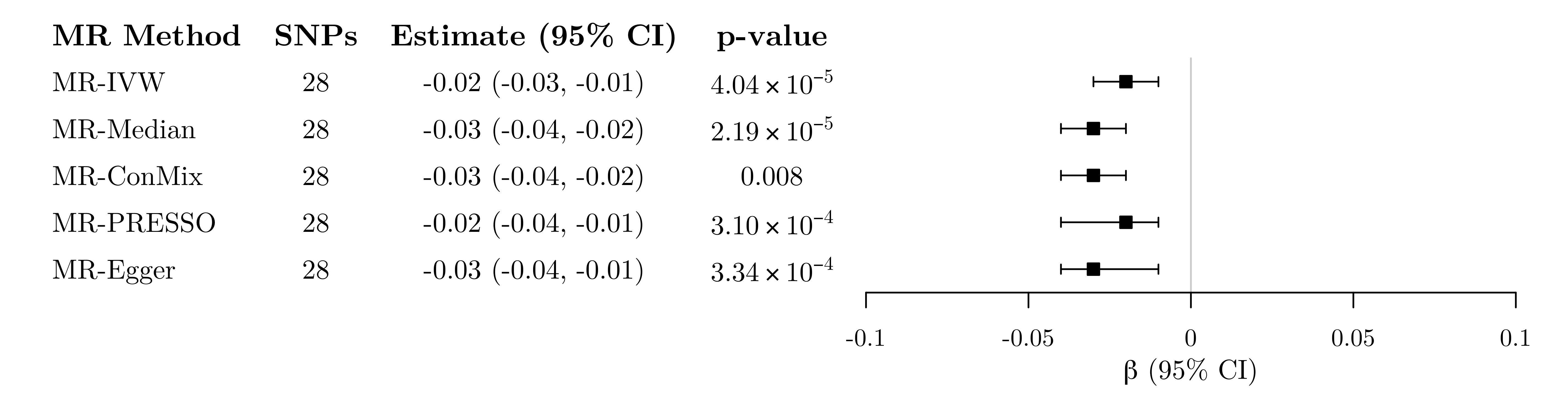 |
| 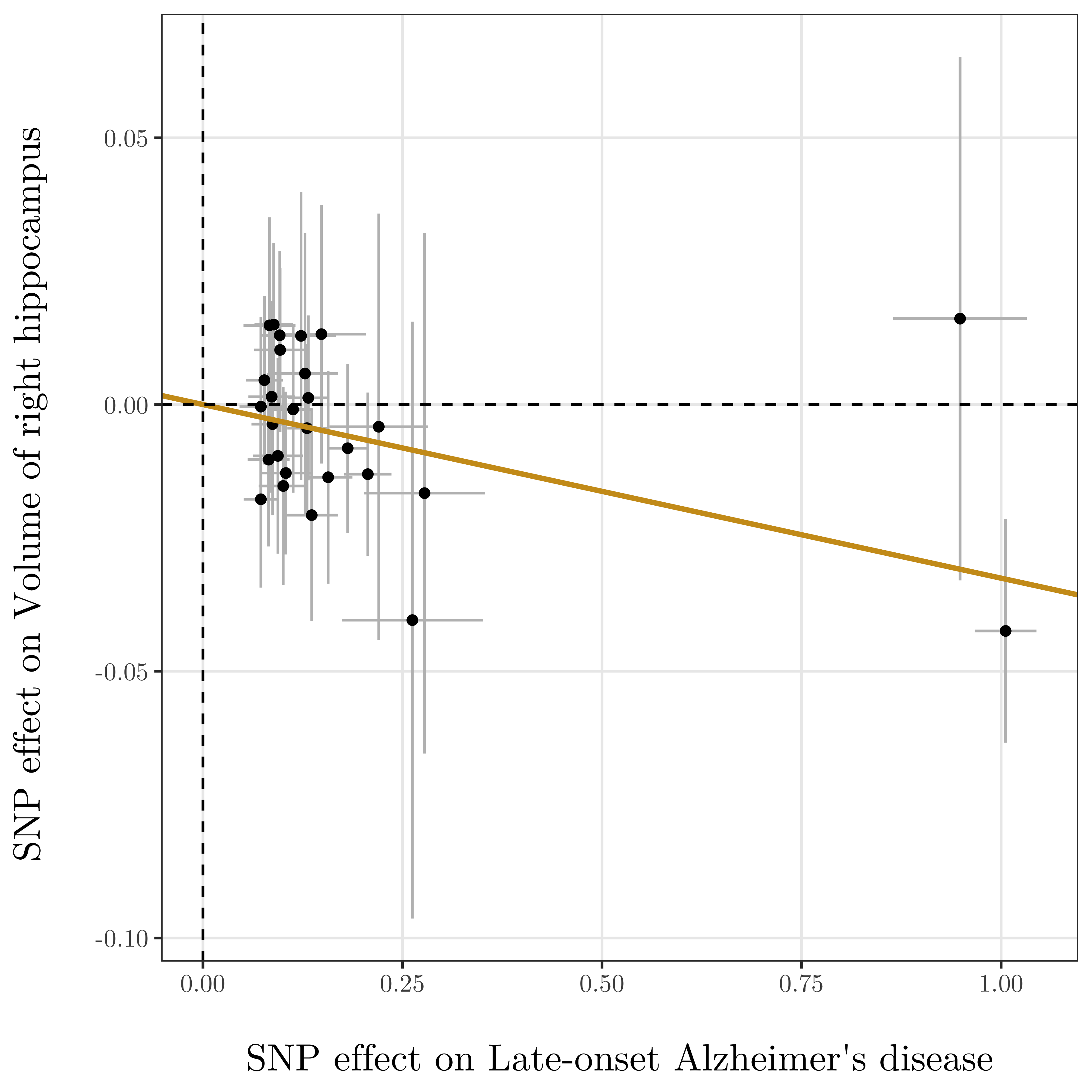 | 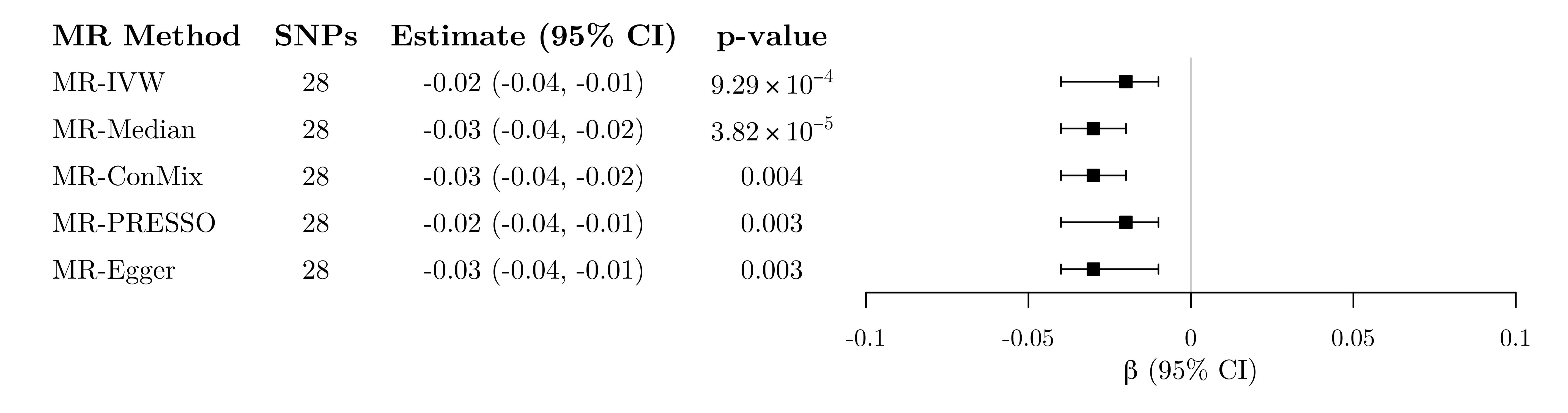 |

**Supplementary Fig. 26** Left: Genetic associations with late-onset Alzheimer’s disease (horizontal axis, log odds ratios) and with four imaging-derived phenotypes (vertical axis, standard deviation units) for 28 genetic variants associated with late-onset Alzheimer’s disease at a genome-wide level of significance. Horizontal and vertical lines represent 95% confidence intervals for the genetic associations. The regression line through the origin represents the inverse-variance weighted Mendelian randomization estimate for the effect of late-onset Alzheimer’s disease on each imaging-derived phenotype. Right: Mendelian randomization estimates of the association between genetically-proxied late-onset Alzheimer’s disease and brain structure phenotypes. Estimates represent the average change in each imaging-derived phenotype (standard deviation units) per doubling (2-fold increase) in the odds of genetically-predicted late-onset AD.

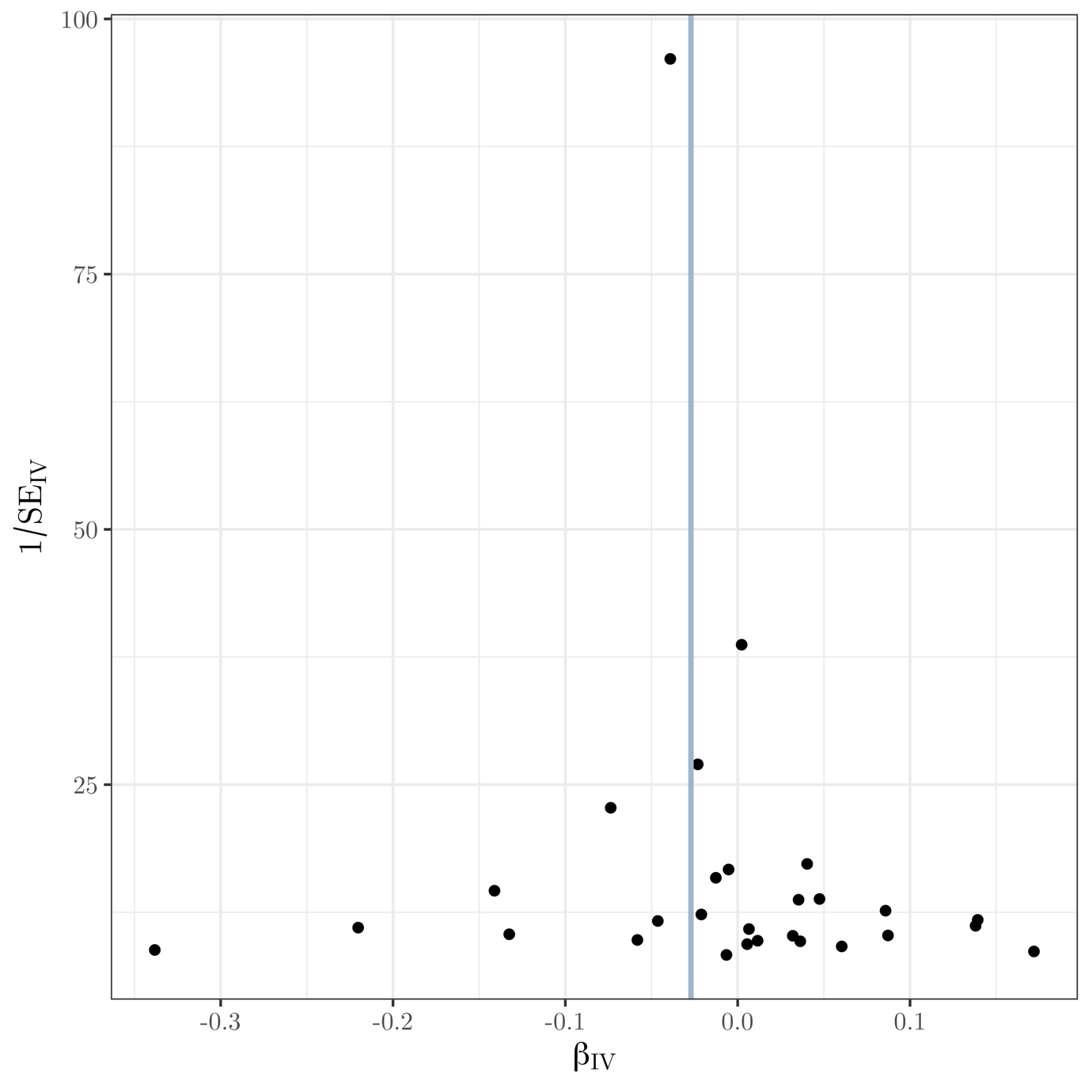

**Supplementary Fig. 27** Funnel plot of instrument precision against instrumental variable estimates for each genetic variant separately for Mendelian randomization analysis of late-onset AD on orientation dispersion index. Solid vertical line is the (fixed-effect) inverse-variance weighted estimate.

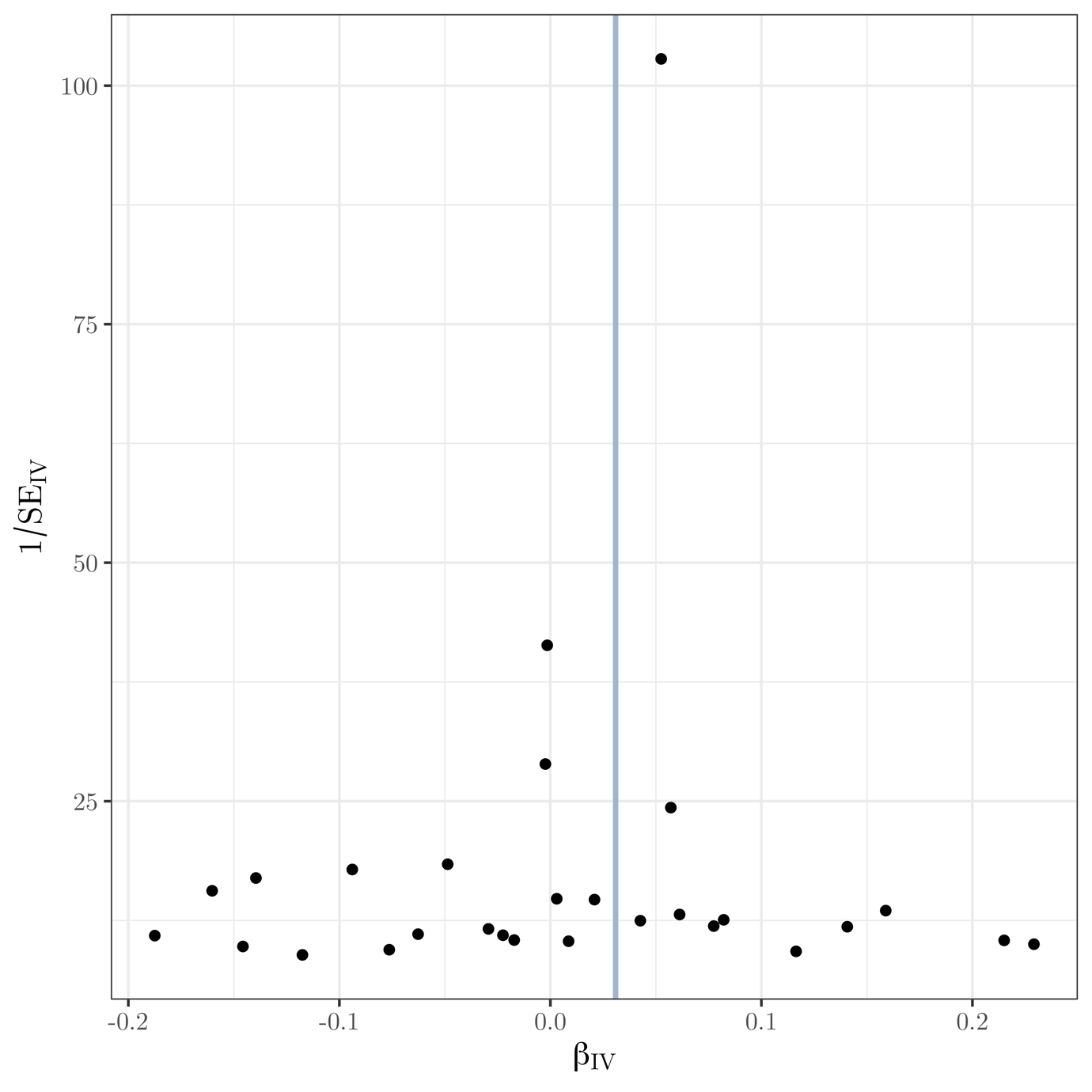

**Supplementary Fig. 28** Funnel plot of instrument precision against instrumental variable estimates for each genetic variant separately for Mendelian randomization analysis of late-onset AD on mean diffusivity (white matter tracts). Solid vertical line is the (fixed-effect) inverse-variance weighted estimate.

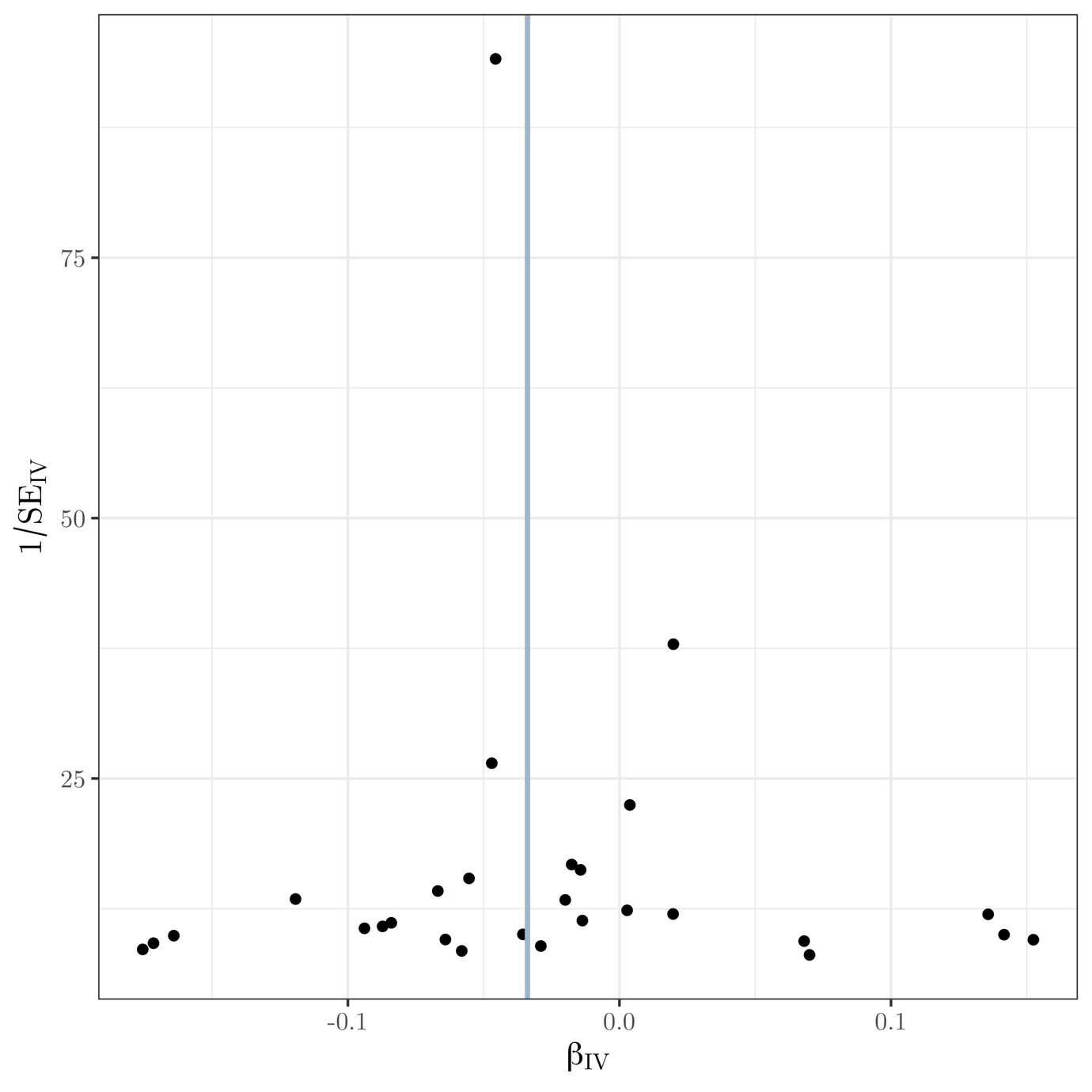

**Supplementary Fig. 29** Funnel plot of instrument precision against instrumental variable estimates for each genetic variant separately for Mendelian randomization analysis of late-onset AD on volume of left hippocampus. Solid vertical line is the (fixed-effect) inverse-variance weighted estimate.

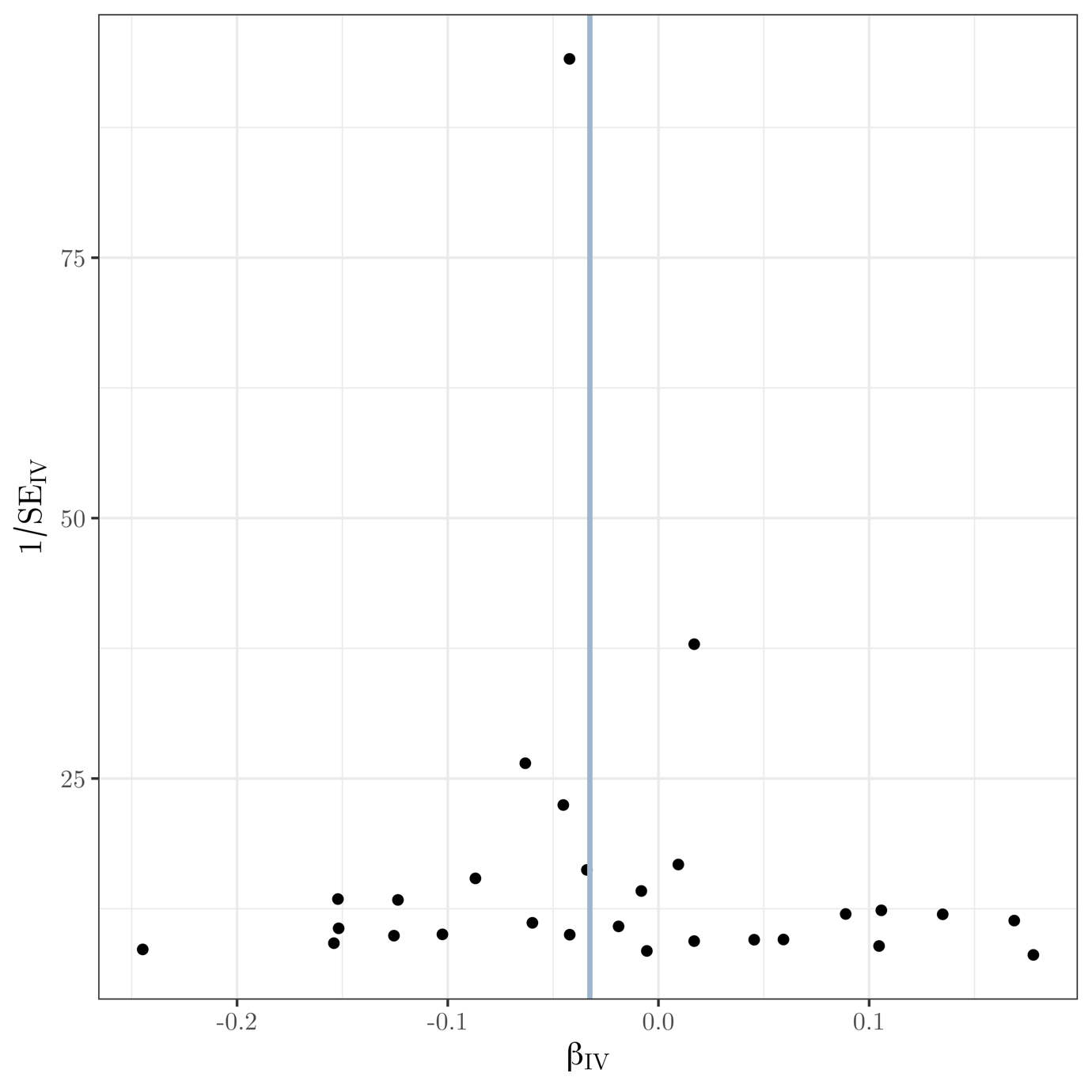

**Supplementary Fig. 30** Funnel plot of instrument precision against instrumental variable estimates for each genetic variant separately for Mendelian randomization analysis of late-onset AD on volume of right hippocampus. Solid vertical line is the (fixed-effect) inverse-variance weighted estimate.

| 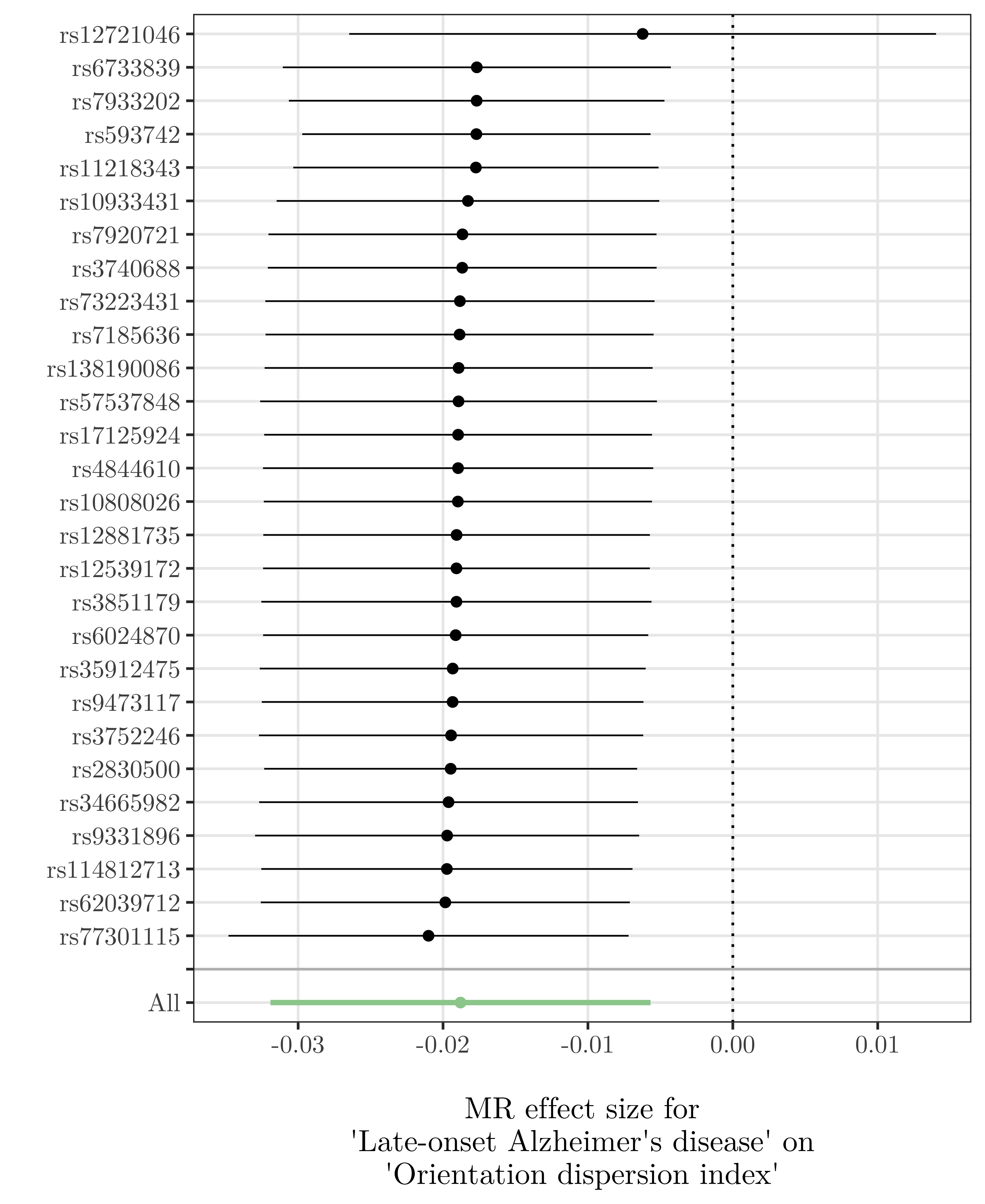 | 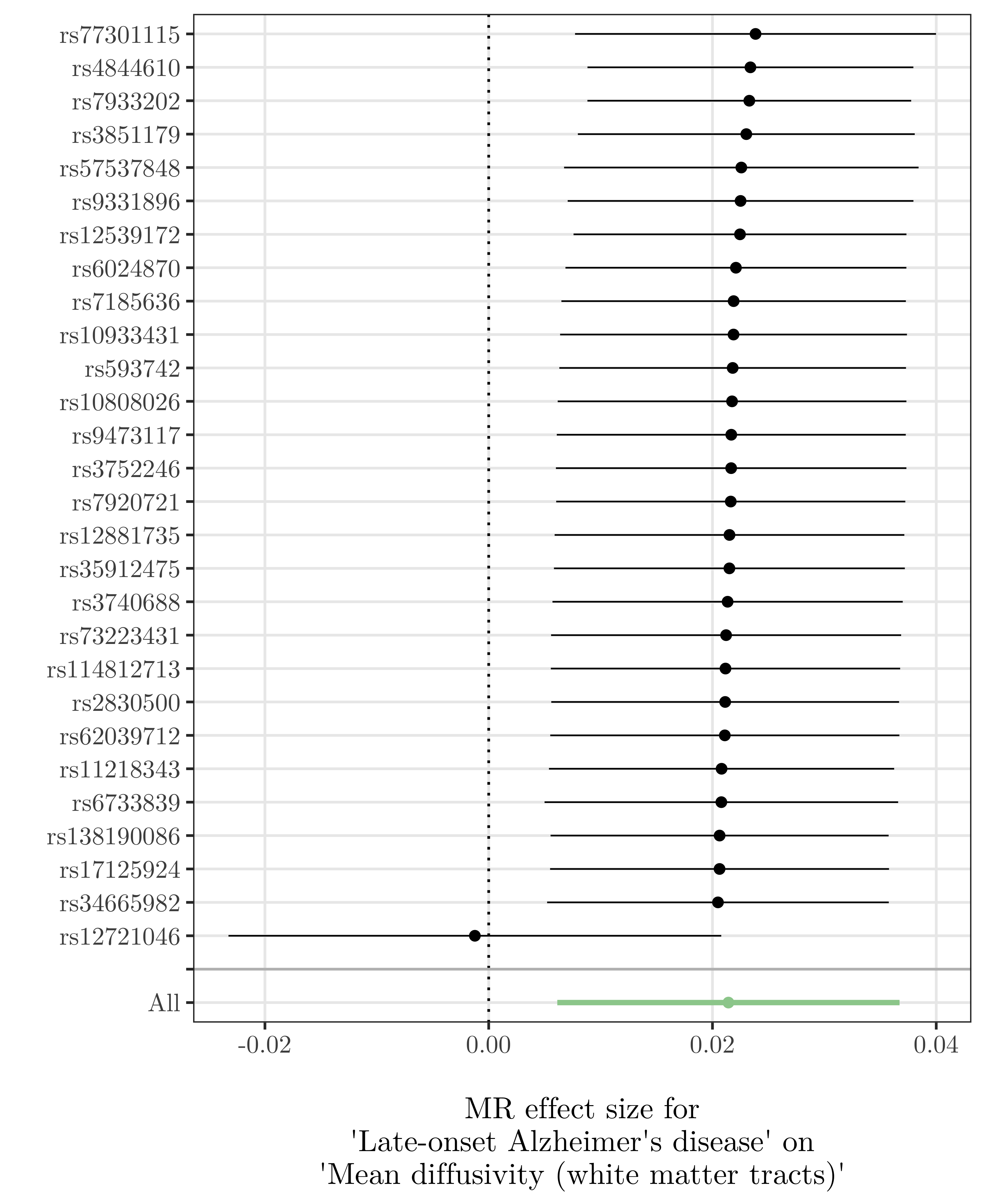 |
| --- | --- |
| 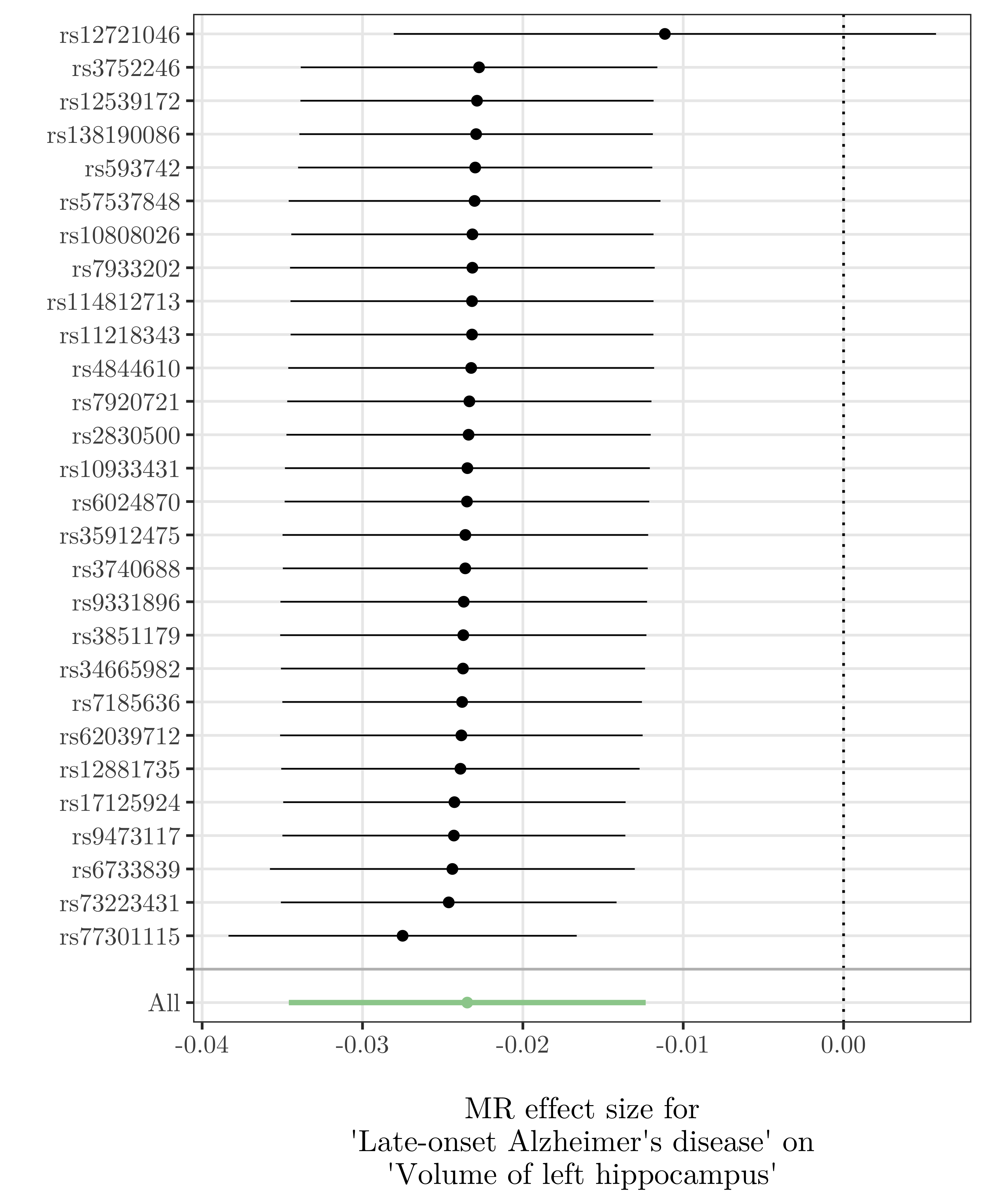 | 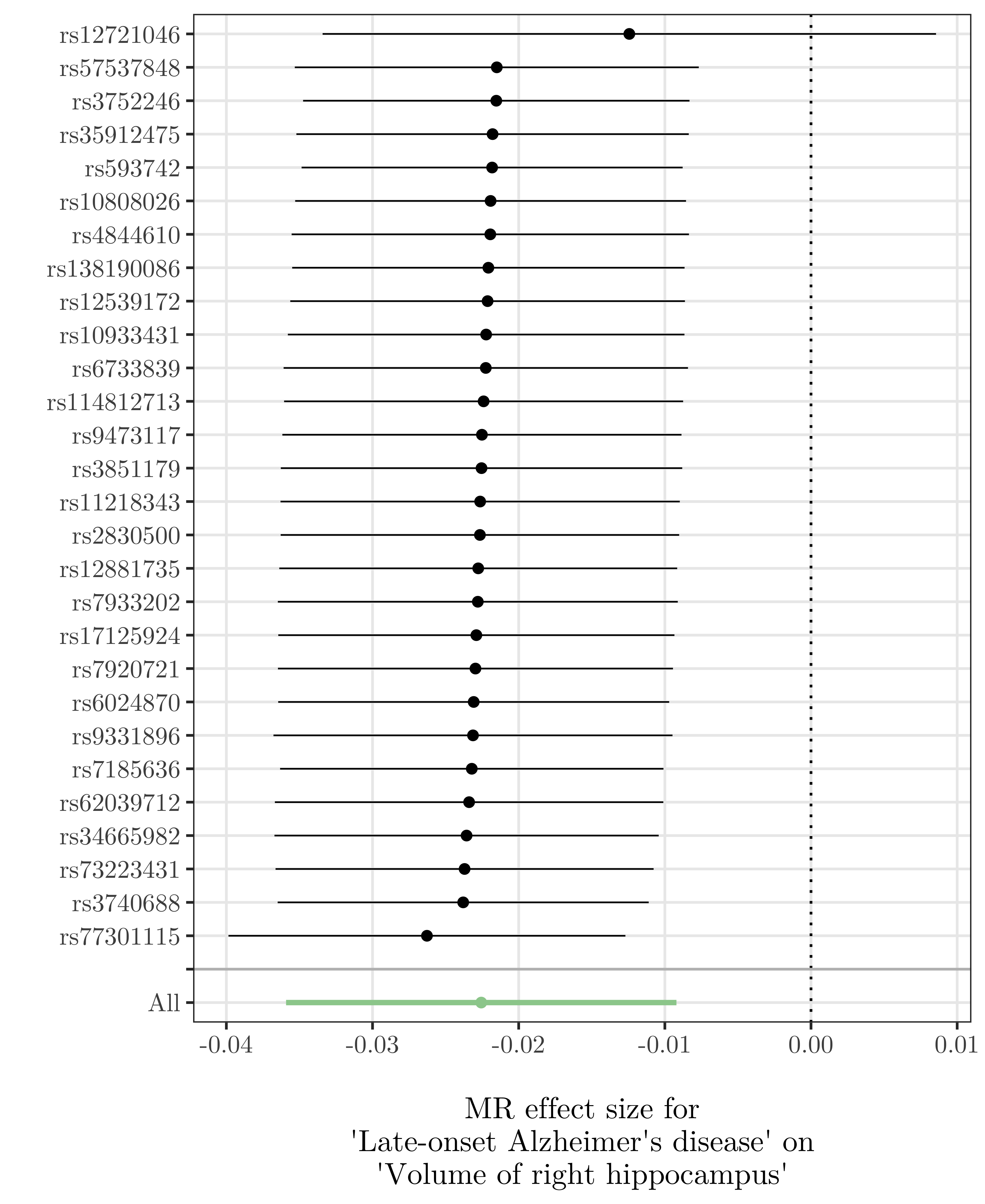 |

**Supplementary Fig. 31 Leave-one-out plots for the analysis of late-onset Alzheimer’s disease on four imaging-derived phenotypes.** Point estimates represent the inverse-variance weighted (IVW) estimate when excluding each SNP in turn. Horizontal lines represent 95% confidence intervals around the IVW estimate.

| 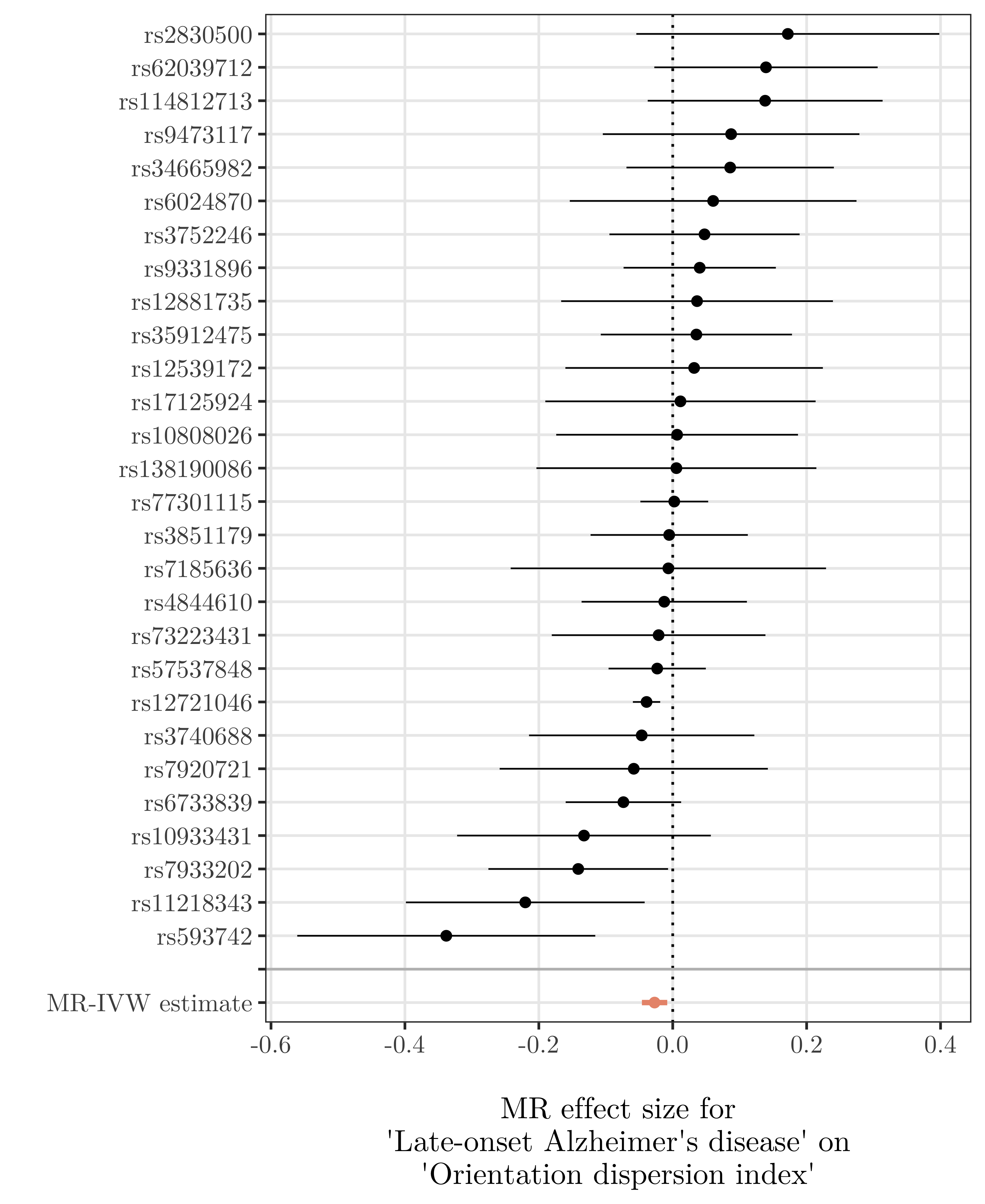 | 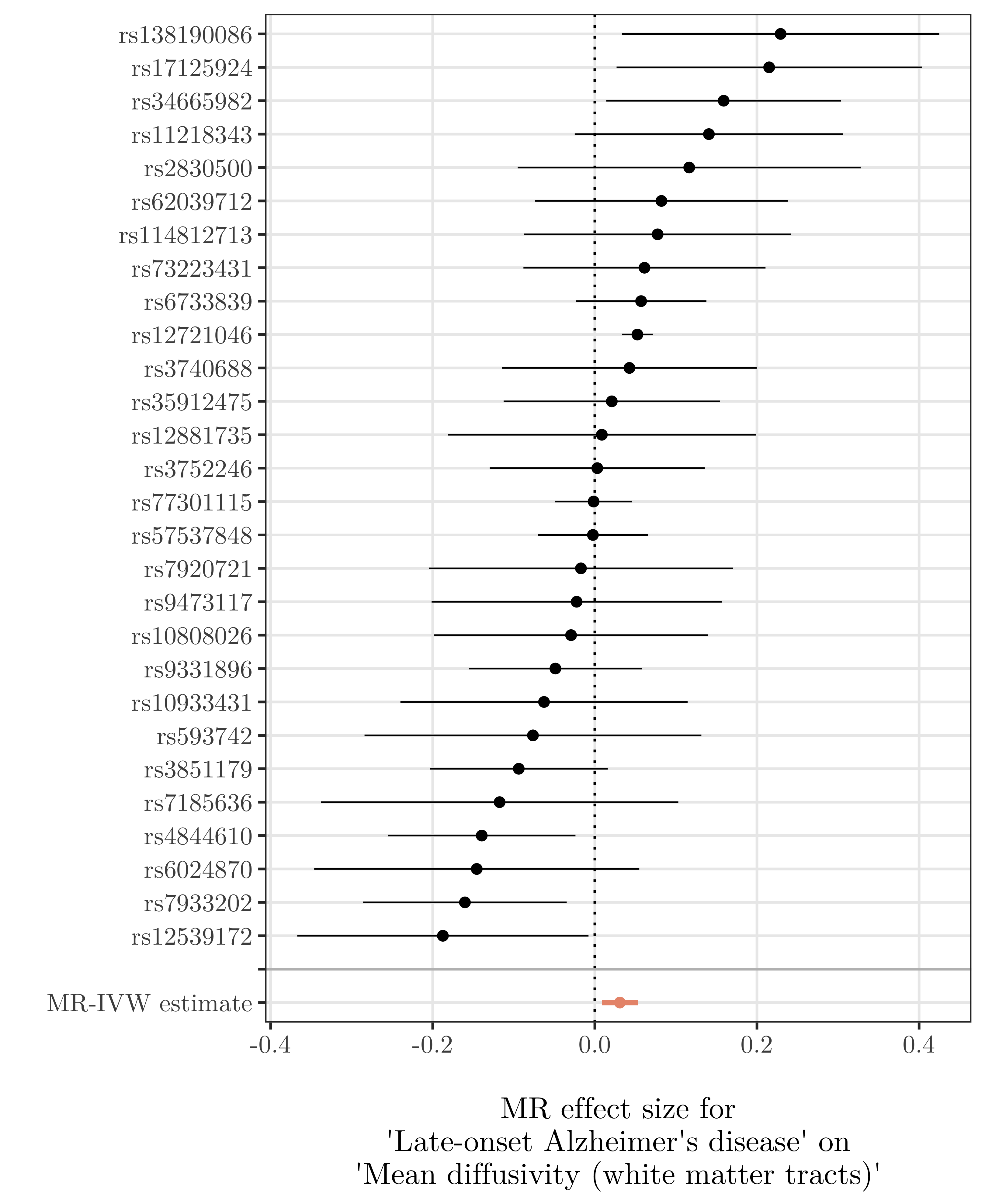 |
| --- | --- |
| 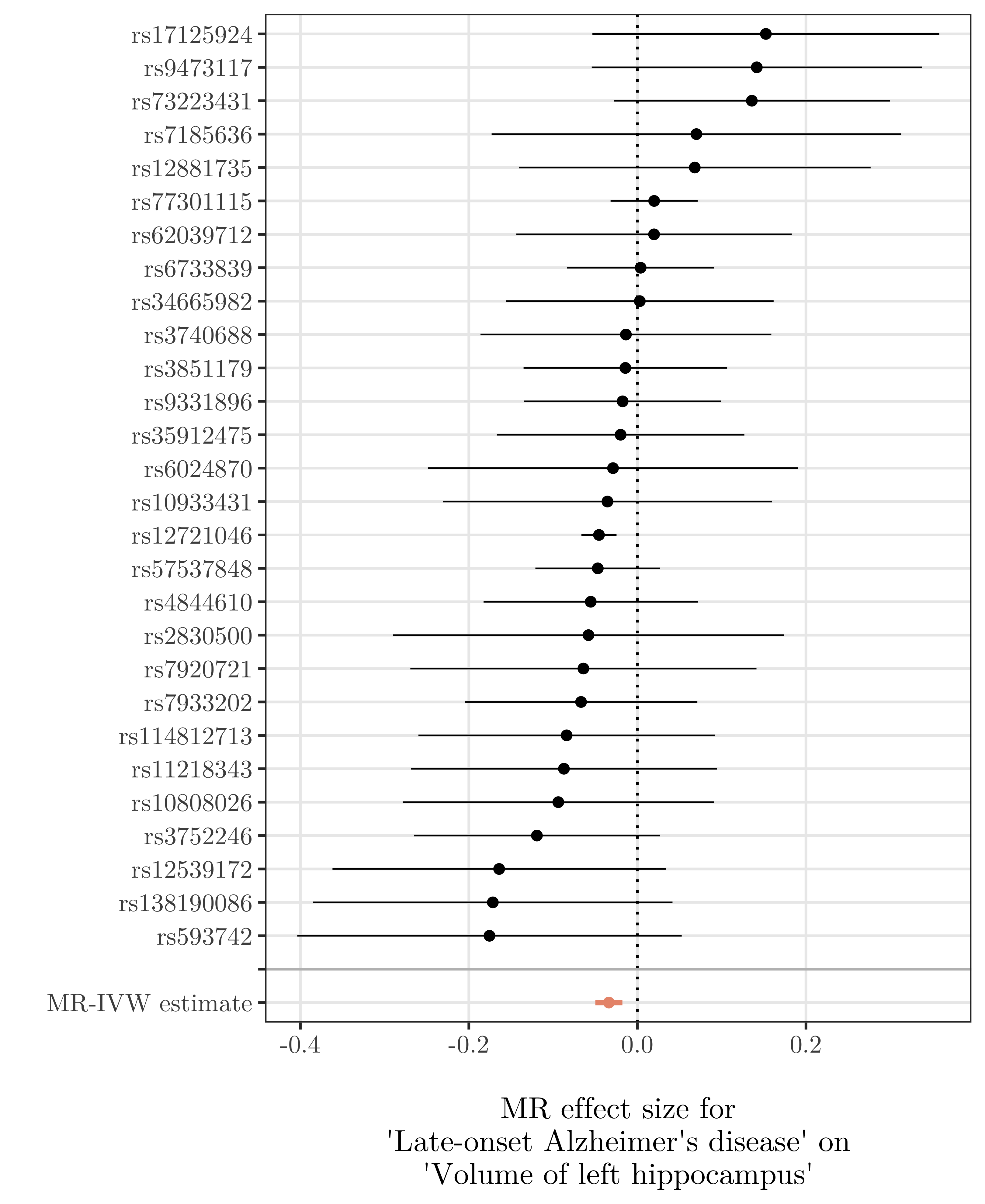 | 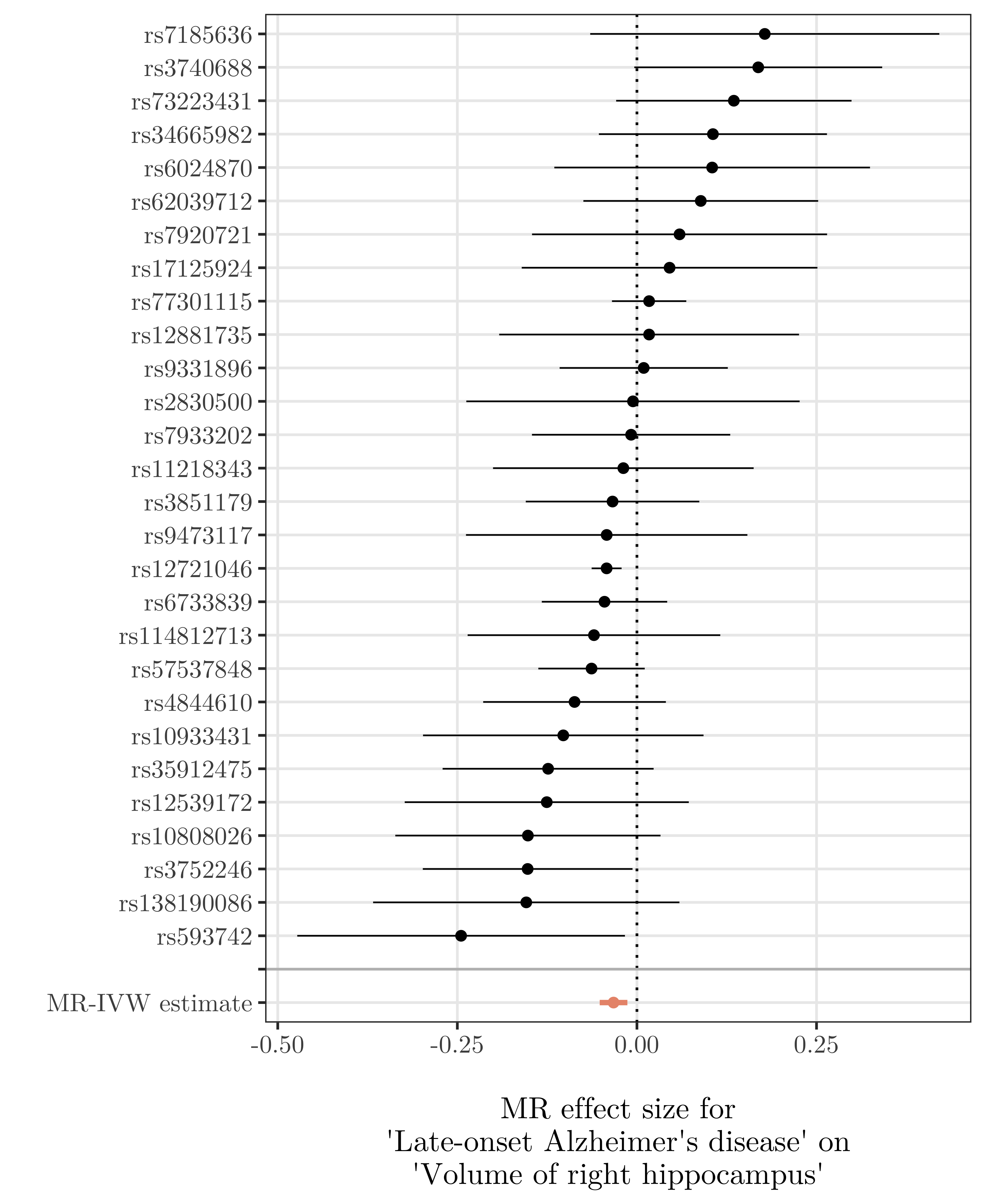 |

**Supplementary Fig. 32 Single-SNP analysis forest plots of the effect of late-onset Alzheimer’s disease on four imaging-derived phenotypes**. Point estimates represent the variant-specific ratio estimates for each SNP (in black), and the inverse-variance weighted (IVW) estimate (in orange). Horizontal lines represent 95% confidence intervals around the variant-specific ratio estimates and the IVW estimate
