## Supplementary material for "Educational attainment, structural brain reserve, and Alzheimer’s disease: a Mendelian randomization analysis": STROBE-MR Checklist

**STROBE-MR checklist of recommended items to address in reports of Mendelian randomization studies**

| **Item No.** | **Section** | **Checklist item** | **Location** |
| --- | --- | --- | --- |
| **Title and abstract** | | | |
| 1 | Title and Abstract | Indicate mendelian randomization (MR) as the study’s design in the title and/or the abstract if that is a main purpose of the study. | Title; Abstract |
| **Introduction** | | | |
| 2 | Background | Explain the scientific background and rationale for the reported study. What is the exposure? Is a potential causal relationship between exposure and outcome plausible? Justify why MR is a helpful method to address the study question. | Introduction ­– Paragraphs 1-3 |
| 3 | Objectives | State specific objectives clearly, including prespecified causal hypotheses (if any). State that MR is a method that, under specific assumptions, intends to estimate causal effects. | Introduction – Paragraph 3 |
| **Methods** | | | |
| 4 | Study design and data sources | Present key elements of the study design early in the article. Consider including a table listing sources of data for all phases of the study. For each data source contributing to the analysis, describe the following:   a) Setting: Describe the study design and the underlying population, if possible. Describe the setting, locations, and relevant dates, including periods of recruitment, exposure, follow-up, and data collection, when available. b) Participants: Report the eligibility criteria and the sources and methods of selection of participants. Report the sample size and whether any power or sample size calculations were carried out prior to the main analysis. c) Describe measurement, quality control, and selection of genetic variants. d) For each exposure, outcome, and other relevant variables, describe methods of assessment and diagnostic criteria for diseases. e) Provide details of ethics committee approval and participant informed consent, if relevant. | a) Materials and methods – Data sources; Table 1; Supplementary materials – Supplementary methods – Description of summary statistics data sources (more detailed descriptions are available in the respective GWAS publications)  b) Materials and methods – Data sources; Table 1; Supplementary materials – Supplementary methods – Description of summary statistics data sources (more detailed descriptions are available in the respective GWAS publications); Supplementary materials – Supplementary Table 1  c) Materials and methods – Selection of genetic instruments and data harmonization; Supplementary materials – Supplementary methods – Description of summary statistics data sources (more detailed descriptions are available in the respective GWAS publications)  d) Materials and methods – Data sources; Supplementary materials – Supplementary methods – Description of summary statistics data sources (more detailed descriptions are available in the respective GWAS publications)  e) Materials and methods – Ethical approval (more detailed descriptions are available in the respective GWAS publications) |
| 5 | Assumptions | Explicitly state the 3 core instrumental variable (IV) assumptions for the main analysis (relevance, independence, and exclusion restriction), as well assumptions for any additional or sensitivity analysis. | Supplementary materials – Supplementary methods – Assumptions of univariable and multivariable Mendelian randomization; Supplementary materials – Supplementary methods – Robust Mendelian randomization methods; Materials and methods ­– Statistical analyses – Sensitivity analyses – Paragraphs 1–2 |
| 6 | Statistical methods: main analysis | Describe statistical methods and statistics used.   a) Describe how quantitative variables were handled in the analyses (ie, scale, units, model). b) Describe how genetic variants were handled in the analyses and, if applicable, how their weights were selected. c) Describe the MR estimator (eg, 2-stage least squares, Wald ratio) and related statistics. Detail the included covariates and, in case of 2-sample MR, whether the same covariate set was used for adjustment in the 2 samples. d) Explain how missing data were addressed. e) If applicable, indicate how multiple testing was addressed. | a) Supplementary materials – Supplementary methods – Description of summary statistics data sources; Units are specified in the first paragraph of each sub-section in Results  b) Materials and methods – Selection of genetic instruments and data harmonization; Materials and methods – Statistical analyses – Bidirectional univariable Mendelian randomization  c) Materials and methods – Statistical analyses – Bidirectional univariable Mendelian randomization; Supplementary materials – Supplementary methods – Description of summary statistics data sources  d) NA  e) Materials and methods – Statistical analyses – Correction for multiple testing |
| 7 | Assessment of assumptions | Describe any methods or prior knowledge used to assess the assumptions or justify their validity. | Materials and methods – Statistical analyses – Sensitivity analyses |
| 8 | Sensitivity analyses and additional analyses | Describe any sensitivity analyses or additional analyses performed (eg, comparison of effect estimates from different approaches, independent replication, bias analytic techniques, validation of instruments, simulations). | Materials and methods – Statistical analyses – Sensitivity analyses |
| 9 | Software and pre-registration | a) Name statistical software and package(s), including version and settings used. b) State whether the study protocol and details were preregistered (as well as when and where). | a) Materials and methods – Statistical analyses – final paragraph  b) The analysis plan was prospectively conceived by the study team in May 2021, although the study protocol was not formally pre-registered. All analyses described here were planned a-priori, except for the single-SNP MR analyses of the effect of genetically-predicted Alzheimer’s disease on selected brain structure phenotypes, which were performed in light of findings from the primary analysis (see the ‘Association of genetically-predicted Alzheimer’s disease with brain structure’ section below). |
| **Results** | | | |
| 10 | Descriptive data | a) Report the numbers of individuals at each stage of included studies and reasons for exclusion. Consider use of a flow diagram. b) Report summary statistics for phenotypic exposure(s), outcome(s), and other relevant variables (eg, means, SDs, proportions). c) If the data sources include meta-analyses of previous studies, provide the assessments of heterogeneity across these studies. d) For 2-sample MR: i. Provide justification of the similarity of the genetic variant–exposure associations between the exposure and outcome samples. ii. Provide information on the number of individuals who overlap between the exposure and outcome studies. | a-c) Supplementary materials – Supplementary methods – Description of summary statistics data sources (more detailed descriptions are available in the respective GWAS publications)  d) Materials and methods ­­– Data sources; Discussion – Paragraph 6 |
| 11 | Main results | a) Report the associations between genetic variant and exposure and between genetic variant and outcome, preferably on an interpretable scale. b) Report MR estimates of the relationship between exposure and outcome and the measures of uncertainty from the MR analysis, on an interpretable scale, such as odds ratio or relative risk per SD difference. c) If relevant, consider translating estimates of relative risk into absolute risk for a meaningful time period. d) Consider plots to visualize results (eg, forest plot, scatterplot of associations between genetic variants and outcome vs between genetic variants and exposure). | a) Supplementary Data; Supplementary Fig. 3, 6, 19, 26  b) Fig. 3–5; Table 2; Results – first paragraph of each sub-section  c) NA  d) Fig. 3–5; Supplementary Data; Supplementary Fig. 3, 6, 19, 26 |
| 12 | Assessment of assumptions | a) Report the assessment of the validity of the assumptions. b) Report any additional statistics (eg, assessments of heterogeneity across genetic variants, such as I2, Q statistic, or E-value). | a) Results – second paragraph of each sub-section; Supplementary Tables 2–4; Supplementary Fig. 4, 7–12, 20–22, 27–30 (funnel plots)  b) Supplementary Table 2 |
| 13 | Sensitivity analyses and additional analyses | a) Report any sensitivity analyses to assess the robustness of the main results to violations of the assumptions. b) Report results from other sensitivity analyses or additional analyses. c) Report any assessment of the direction of the causal relationship (eg, bidirectional MR). d) When relevant, report and compare with estimates from non-MR analyses. e) Consider additional plots to visualize results (eg, leave-one-out analyses). | a) Results – first paragraph of each sub-section; Fig. 3, Supplementary Fig. 6, 19, 26  b) Results – second paragraph of each sub-section; Results – Mediation analysis; Table 2; Supplementary Tables 2–4; Supplementary Fig. 3–32  c) Bi-directional MR results are presented in the first paragraph of each sub-section in Results and in Fig. 4 & 5; Steiger filtering results are provided in Supplementary Table 4  d) NA  e) Supplementary Fig. 4, 7–12, 20–22, 27–30 (funnel plots); Supplementary Fig. 5, 13–18, 23–25, 31 (leave-one-out plots); Supplementary Fig. 32 (forest plots of single-SNP MR analyses) |
| **Discussion** | | | |
| 14 | Key results | Summarize key results with reference to study objectives. | Discussion – Paragraph 1 |
| 15 | Limitations | Discuss limitations of the study, taking into account the validity of the IV assumptions, other sources of potential bias, and imprecision. Discuss both direction and magnitude of any potential bias and any efforts to address them. | Discussion – Paragraph 6 |
| 16 | Interpretation | a) Meaning: Give a cautious overall interpretation of results in the context of their limitations and in comparison with other studies. b) Mechanism: Discuss underlying biological mechanisms that could drive a potential causal relationship between the investigated exposure and the outcome, and whether the gene-environment equivalence assumption is reasonable. Use causal language carefully, clarifying that IV estimates may provide causal effects only under certain assumptions. c) Clinical relevance: Discuss whether the results have clinical or public policy relevance, and to what extent they inform effect sizes of possible interventions. | a) Discussion – final paragraph  b) Discussion – Paragraphs 2–5  c) Discussion – Paragraph 2 |
| 17 | Generalisability | Discuss the generalizability of the study results (a) to other populations, (b) across other exposure periods/timings, and (c) across other levels of exposure. | Discussion – Paragraph 6 |
| **Other information** | | | |
| 18 | Funding | Describe sources of funding and the role of funders in the present study and, if applicable, sources of funding for the databases and original study or studies on which the present study is based. | Funding statement |
| 19 | Data and data sharing | Provide the data used to perform all analyses or report where and how the data can be accessed, and reference these sources in the article. Provide the statistical code needed to reproduce the results in the article or report whether the code is publicly accessible and, if so, where. | Methods – Data and code availability |
| 20 | Conflicts of interest | All authors should declare all potential conflicts of interest. | Competing interests statement |
